# Dyadic Parent/Caregiver-Infant Interventions Initiated in the First 6 Months of Life to Support Early Relational Health: A Meta-Analysis

**DOI:** 10.1101/2022.10.29.22281681

**Authors:** Andréane Lavallée, Lindsy Pang, Jennifer M. Warmingham, Ginger D. Atwood, Imaal Ahmed, Marissa R. Lanoff, Morgan A. Finkel, Ruiyang Xu, Elena Arduin, Kassidy K. Hamer, Rachel Fischman, Sharon Ettinger, Yunzhe Hu, Kaylee Fisher, Esther A. Greeman, Mia Kuromaru, Sienna S. Durr, Elizabeth Flowers, Aileen Gozali, Seonjoo Lee, David Willis, Dani Dumitriu

## Abstract

**Importance:** In 2021, the American Academy of Pediatrics published a policy statement seeking to create a paradigm shift away from a focus on childhood toxic stress and toward the emphasis on early relational health (ERH) as a buffer for childhood adversity and promoter of life-course resilience. A comprehensive appraisal of the efficacy of contemporary parent/caregiver-child interventions in – primarily – improving ERH, and – secondarily – enhancing child well-being and neurodevelopment is needed to guide widespread implementation and policy.

**Objective:** Determine the effectiveness of contemporary early dyadic parent/caregiver-infant interventions on ERH, child socio-emotional functioning and development, and parent/caregiver mental health.

**Data Sources:** PubMed, Medline, Cinhal, ERIC, and PsycInfo were searched on April 28, 2022. Additional sources: clinical trial registries (clinicaltrials.gov, ISRCTN Registry, EU Clinical Trials Register, Australian New Zealand Clinical Trials Registry), contacting authors of unpublished/ongoing studies, backward/forward reference-searching.

**Study Selection:** Studies targeting parent/caregiver-infant dyads and evaluating effectiveness of a dyadic intervention were eligible. Study selection was performed in duplicate, using Covidence.

**Data Extraction and Synthesis:** Cochrane’s methodological guidance presented per PRISMA guidelines. Data extraction and risk of bias assessment were completed in duplicate with consensuses by first author. Data were pooled using inverse-variance random effects models.

**Main Outcomes and Measures:** The primary outcome domain was ERH. Secondary outcome domains were child socio-emotional functioning and development, and parent/caregiver mental health, and were only considered in studies where at least one ERH outcome was also measured. The association between dose of intervention and effect estimates was explored.

**Results:** 93 studies (14,993 parent/caregiver-infant dyads) met inclusion criteria. Based on very low to moderate quality of evidence, we found significant non-dose-dependent intervention effects on several measures of ERH, including bonding, parent/caregiver sensitivity, attachment, and dyadic interactions, and a significant effect on parent/caregiver anxiety, but no significant effects on other child outcomes.

**Conclusion:** Current evidence does not support the notion that promoting ERH through early dyadic interventions ensures optimal child development, despite effectively promoting ERH outcomes. Given the lack of an association with dose of intervention, the field is ripe for novel, innovative, cost-effective, potent ERH intervention strategies that effectively and equitably improve meaningful long-term child outcomes.

## INTRODUCTION

In 2021, the American Academy of Pediatrics (AAP)^1^ made a paradigm-shifting statement – promoting early relational health (ERH), or the ability to form and maintain safe, stable, and nurturing parent/caregiver-child relationships, is a priority in pediatrics. Building on decades of research establishing the association between ERH and later child emotional, mental, relational, and physical health,^2-7^ the AAP policy statement postulates that bolstering ERH through early childhood interventions may be the key for universal promotion of physical health and mental well-being of both children and their parents/caregivers,^8^ by building resilience and protecting against the negative effects of adverse childhood experiences (ACEs).^1,9-12^ There is incontestable evidence that exposure to ACEs, including maltreatment and abuse, poverty, racism, and household dysfunction, conveys risk for adverse mental and physical health outcomes across the life-course.^13,14^ ACE-associated adverse outcomes are diverse, and range from biological changes,^9,15^ to lifelong impacts like mental health problems^16^ and chronic diseases,^17,18^ leading to an estimated total annual cost of $748 billion in North America.^19^ In the face of the current child mental health crisis,^20,21^ and in response to the AAP policy statement to hardwire universal ERH promotion into pediatric care, here we investigate the global effectiveness of parent/caregiver-infant dyadic interventions on ERH and associated outcomes.

ERH encompasses a variety of theoretical perspectives, often erroneously used interchangeably.^22^ Some constructs are heavily parent/caregiver- or child-driven, and others are dyadic. Bonding, for example, is characterized as a unidirectional parent/caregiver-to-infant emotional tie,^23^ that forms from pregnancy and beyond birth.^24,25^ Parent/Caregiver-driven behaviors, often described as sensitive,^26-28^ responsive,^23,28,29^ and synchronous,^28^ are the main early predictors of later child attachment. Child-to-parent/caregiver attachment develops over time, ^30^ and this tie can be classified into three categories: secure, insecure-ambivalent, or insecure-avoidant.^31^ Secure attachment is associated with available and responsive caregivers^32^ and predicts optimal social and behavioral skills.^33,34^ Insecure attachment is associated with greater risk for poor interpersonal and cognitive skills,^35,36^ depression,^37^ anxiety,^38^ as well as eating,^39^ post-traumatic,^40^ and obsessive-compulsive disorders.^41^ Insecure attachment can additionally be characterized as disorganized,^42^ a predictor of poor socio-emotional functioning.^43^ Finally, parent/caregiver-infant relationships can also be viewed through a dyadic lens, such as the lenses of emotional connection,^44^ emotional availability,^45^ dyadic synchrony,^46^ dyadic attunement^47^ or dyadic mutuality,^48^ in which strong dyadic parent/caregiver-child interactions are mutually attuned and reciprocal. While various theoretical constructs define the origins and mechanisms of healthy and impaired parent/caregiver-infant relationships differently, at their core, they are unified by their goal of measuring various components of this relationship.

A comprehensive understanding of the effectiveness of parent/caregiver-infant interventions at promoting ERH is currently lacking. Published systematic reviews have focused on specific ERH outcomes, such as attachment,^49-55^ sensitivity,^50^ and parent/caregiver-child interactions,^51,56^ or have individually looked at the effectiveness of ERH interventions on child development, leaving ERH outcomes aside.^57-61^ Using the blanket term “ERH” for all concepts describing the tie between parents/caregivers and infants, indiscriminate to their theoretical origins, and in the context of ERH by definition relying on how parent/caregiver and infant come together as a unit,^44,62,63^ this systematic review is the first to generate a landscape overview of interventions specifically targeting parent/caregiver-infant interactions. Our overarching goal was to determine the global effectiveness of these interventions in both fostering various components of ERH, as well as in promoting secondarily associated outcomes, i.e., child development, and parent/caregiver mental health.

For feasibility and due to the immediate postpartum period’s greatest potential for developmental embedding, we focus on dyadic interventions initiated within the first 6 months of life. This also expands on other public health recommendations targeting the first 6 months of life–e.g., exclusive breastfeeding^64,65^ for primary prevention of medical conditions like obesity^66,67^ and diabetes,^67,68^ and for promotion of outcomes like bonding^69^ and intelligence.^67^ Additionally, specific to the birthing parent, focusing on the early postpartum period also leverages pregnancy-related neural changes and neuroplasticity that prime the parent’s brain for optimal bonding.^70^ To account for foreseeable variability and heterogeneity, the association between the dose and timing of included dyadic interventions, as well as other child-, family- and intervention-level moderators and overall effect estimates are explored.

## METHODS

This systematic review follows Preferred Reporting Items for Systematic Reviews and Meta-Analyses (PRISMA) guidelines (eTable1),^71^ and was registered prospectively in PROSPERO (CRD42022329894). Abbreviated methods included here (detailed methods: eMethods1).

Ethical review and informed consent not applicable as only previously published data was used.

### Eligibility Criteria

Randomized controlled trials (RCT) of any type published on/after January 1, 2000, in English or French and comparing an early dyadic parent/caregiver-infant intervention to any comparator.

*<u>Population</u>*. Parent/caregiver-child dyads.

*<u>Intervention</u>*. “Dyadic” interventions, defined as targeting at least one primary caregiver and the infant together, with at least one intervention session occurring within the first 6 months postpartum. Prenatal interventions without postnatal sessions were excluded. No exclusion criteria for intervention length or intensity.

*<u>Comparator</u>*. Any type, i.e., control or active intervention.

*<u>Outcomes</u>*. Primary outcome domain was ERH, e.g., attachment, sensitivity, bonding, emotional connection. Secondary outcome domains were child socio-emotional functioning and development, and parent/caregiver mental health. Secondary outcomes were only considered in studies where at least one ERH outcome was measured.

### Information Sources and Search Strategy

PubMed, Medline, Cinhal, ERIC, and PsycInfo via EBSCO were searched on April 28, 2022 and targeted two concepts: (1) parent/caregiver-infant dyadic interventions and (2) ERH (full search strategies: eMethods2). Unpublished and ongoing studies were identified in clinical trial registries (clinicaltrials.gov, ISRCTN Registry, EU Clinical Trials Register, Australian New Zealand Clinical Trials Registry) and authors contacted. Backward/forward reference-searching of included studies was conducted.

### Selection Process

Identified studies were uploaded to EndNote V9.3.3^72^ then Covidence.^73^ Duplicates were removed. Studies were screened for eligibility independently by teams of two authors. Disagreements were resolved by the first author. The same process was followed for full-text review of potentially eligible studies. Reasons for exclusion were documented at the full-text screening stage.

### Data collection process

Data extraction was performed in duplicate and independently, using a data extraction form specifically developed for this review. Consensuses were resolved by the first author.

*<u>Study-level</u>*. Aim, study design, number of groups, target population, inclusion and exclusion criteria, group differences, unit of randomization, sample size, withdrawals.

*<u>Intervention-level</u>*. Based on TIDieR^74^: intervention name, framework or underlying theory, rationale, materials, dyadic and non-dyadic procedures, provider(s), location(s), dose, intensity.

*<u>Result-level</u>*. Outcome name, scale, procedure description for observational outcomes, timing assessment, infant age, means/medians, standard deviations (SD), standard error (SE), 95% confidence interval (95%CI)/ Inter Quartile Range (IQR)/ events (for binary outcomes).

Raw unadjusted means analyzed in intention-to-treat were preferred over adjusted means/medians. SE, 95%CI and IQR were converted to SDs following Cochrane guidelines.^75^ Numerical data only presented in figures was extracted using Engauge Digitizer^76^ v12.1 or PlotDigitizer^77^. Corresponding authors were contacted for missing data.

### Effect Measures and Synthesis Methods

Pairwise meta-analyses comparing intervention versus control (standard care, attention control, or active control) were performed separately on outcomes with available data from at least two studies. Separate analyses were conducted per follow-up (F/U) time points: end of intervention (0-1-month) F/U, 2-6-month F/U, 7-12-month F/U, 13-24-month F/U and >25-months F/U.

Unit-of-analysis was parent/caregiver-infant dyad. Summary of methodological decisions to avoid unit-of-analysis issues is presented in eTable2. Continuous outcomes were analyzed using weighted standardized mean differences (SMD) with 95%CIs. Categorical variables were analyzed using weighted odds ratios (OR) with 95%CIs. An inverse variance random-effects model was chosen because of study design and population-induced variability. Significance level was set at 0.05. Cochran’s Q test and I2 statistic were used to assess the heterogeneity and I2*>*50% or p*<*0.10 indicated statistically significant heterogeneity.

Sensitivity analyses were conducted to determine whether pooled effect estimates were robust to inclusion of cluster RCTs, active controls, and studies with a concerning/high risk-of-bias. Additionally, we tested the subset of 20 studies reporting developmental outcomes on ERH effect estimates.

In addition to the preregistered moderator ‘intervention dose’ (in provider minutes), effects of other commonly considered moderators (listed in eFigure1) were tested at parent/caregiver/family-level (target population, e.g., adolescent mothers, incarcerated mothers, and low vs high socio-economic status), infant-level (sex, age at first session, age at follow-up, and preterm/low birthweight versus term/normal birthweight), intervention-level (length of intervention, number of sessions, provider, location), and study-level (year of publication, and preregistration as an analogue of study quality). Continuous moderators were explored with a meta-regression and categorical moderators with a *Q*-test,^78^ both using a random-effects DerSimonian Laird model on each outcome pooling at least 10 studies (and with more than one study in each category in the case of categorical moderators).^79^ Analyses were conducted with Review Manager V5.4^80^ or SPSS V24.^81^

### Study Risk-of-Bias and Certainty Assessment

Two independent authors assessed risk-of-bias in individual studies using Cochrane Collaboration Risk of Bias Tool 2.^82^ Confidence in pooled outcomes was based on Grades of Recommendation, Assessment, Development and Evaluation (GRADE) guidelines.^83^ Within the GRADE assessment, risk of non-reporting bias was estimated by funnel plot inspection.

## RESULTS

### Study-Level Characteristics

A total of 93 primary studies^84-176^ (n=14,993 parent/caregiver-infant dyads) were identified (Figure 1). Most study designs were parallel-group RCTs (k=79, 85%), nine were cluster RCTs (10%), and five were pragmatic RCTs (5%). Across studies reporting demographic characteristics (k=77), the sample is comprised of 48.87±7.30% female infants and 94.79±20.92% biological mothers. The predominant population was parents and their preterm or low birthweight infants (k=31, 33.3%), followed by mothers with a confirmed diagnosis of any psychopathology (k=16, 17.2%), parents of low socio-economic status (k=12, 12.9%), parents with two or more risk categories (k=10, 10.8%), first-time mothers (k=5, 5.4%), infants with health or developmental conditions (k=3, 3.2%), foster or adoptive parents (k=3, 3.2%), adolescent mothers (k=2, 2.2%), dyads at increased risk for maltreatment (k=2, 2.2%), and mother-infant dyads in prison (k=1, 1.1%). Only one study (1.1%) specifically targeted father-infant dyads, and 7 (7.5%) targeted parent/caregiver-infant dyads without any particular risk factor. Full study-level characteristics are presented in eTable3. Twelve parent/caregiver-reported, and 41 observational assessments of ERH were identified and thoroughly described in eMethods3 and eMethods4, respectively.

**Figure 1.**
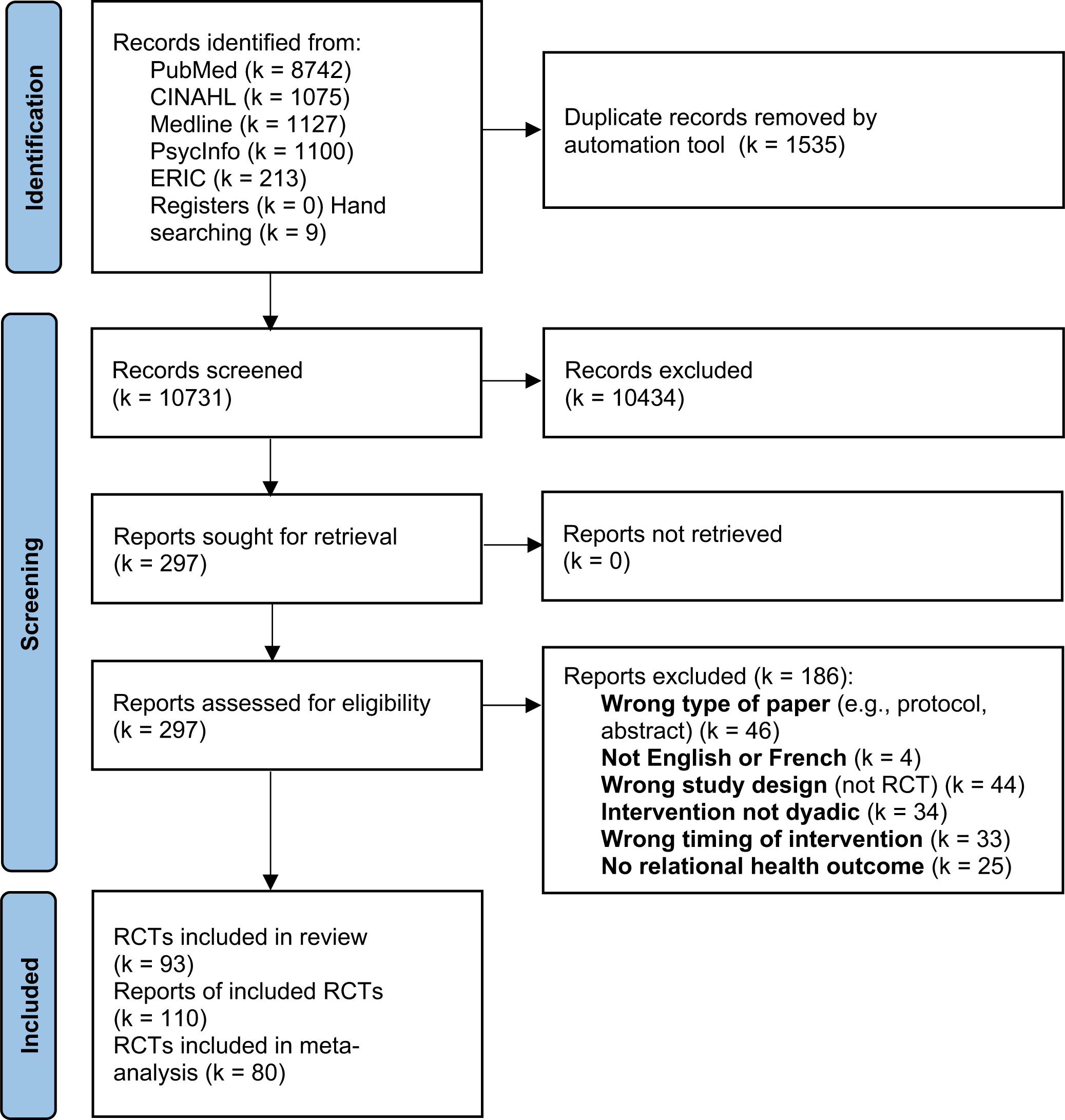
PRISMA flow diagram.

### Intervention-Level Characteristics

Main intervention characteristics are presented in Table 1 and interventions are thoroughly described in eMethods5. Some programs were evaluated in more than one study: Video Interaction Project (VIP; k=5), Playing and Learning Strategies (PALS; k=5), Parent Infant Transaction Program (MITP; k=4), Auditory-Tactile-Visual-Vestibular (ATVV; k=3), and Happiness, Understanding, Giving and Sharing (HUGS; k=2). Interventions averaged 10±12 (min-max: 0-78) sessions of 60±30 (min-max: 0-175) minutes each, for an overall mean total dose of 718±1,091 (min-max: 0-8,640) minutes provided over 161±230 (min-nax:3-1,095) days. Providers were mostly nurses (k=26, 28%). Other providers included therapists (k=10, 10.8%), mixed healthcare professionals, e.g., nurses, and/or social workers, and/or psychologists (k=9, 9.7%), PhDs or MDs (k=9, 9.7%), master’s prepared professionals (k=6, 6.5%), parents (k=6, 6.5%), or trained non-healthcare workers (k=3, 3.2%). The rest had no provider (online intervention, k=1, 1.1%) or unclassified providers (k=23, 24.7%). Interventions began prenatally (k=10, 10.8%), in-hospital perinatally (k=30, 32.3%), within the first six months postdischarge (k=24, 25.8%), or at a less specific time (e.g., “any time between birth to 3 years of age”, n=29, 31.2%).

**Table 1.**
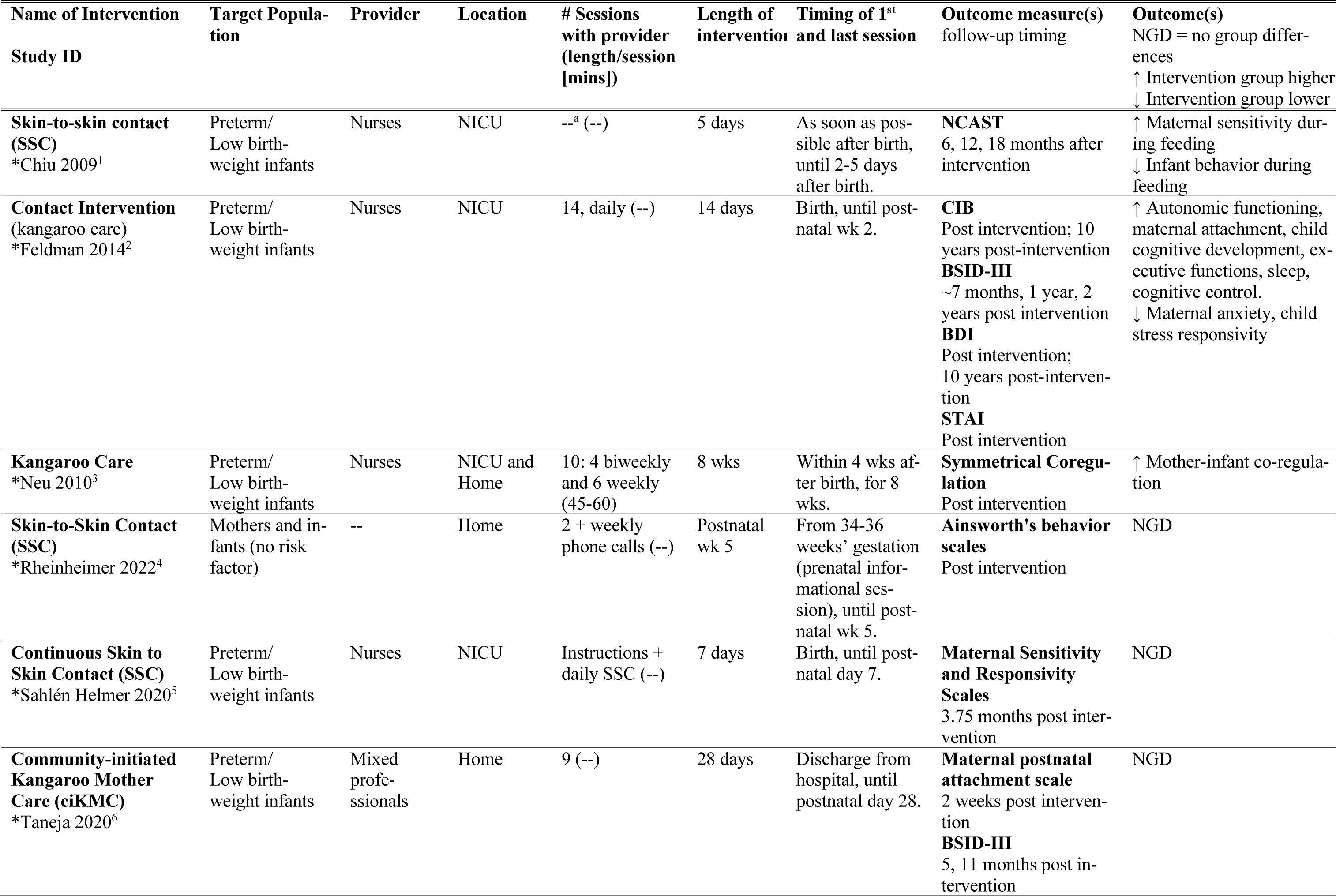
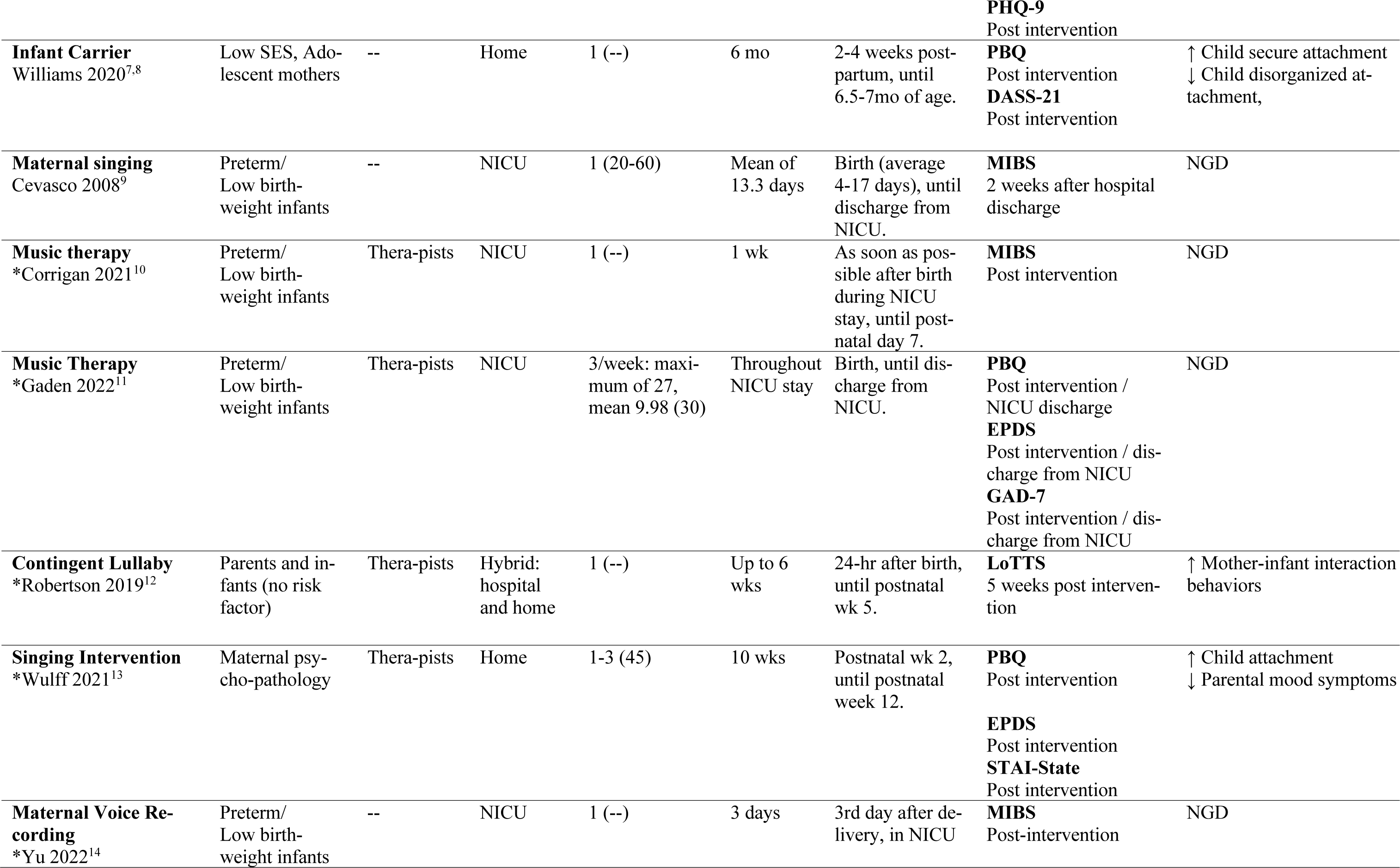
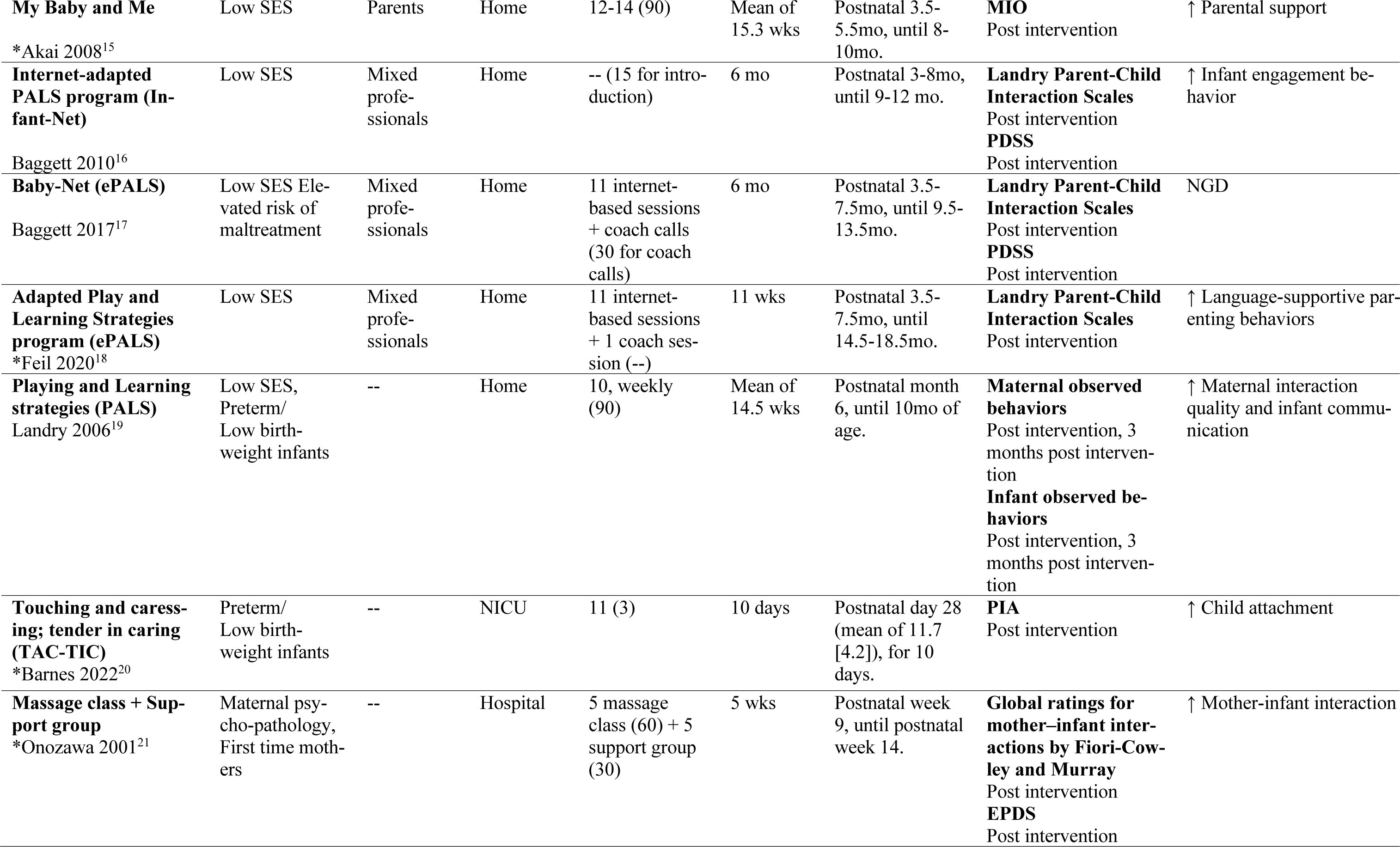
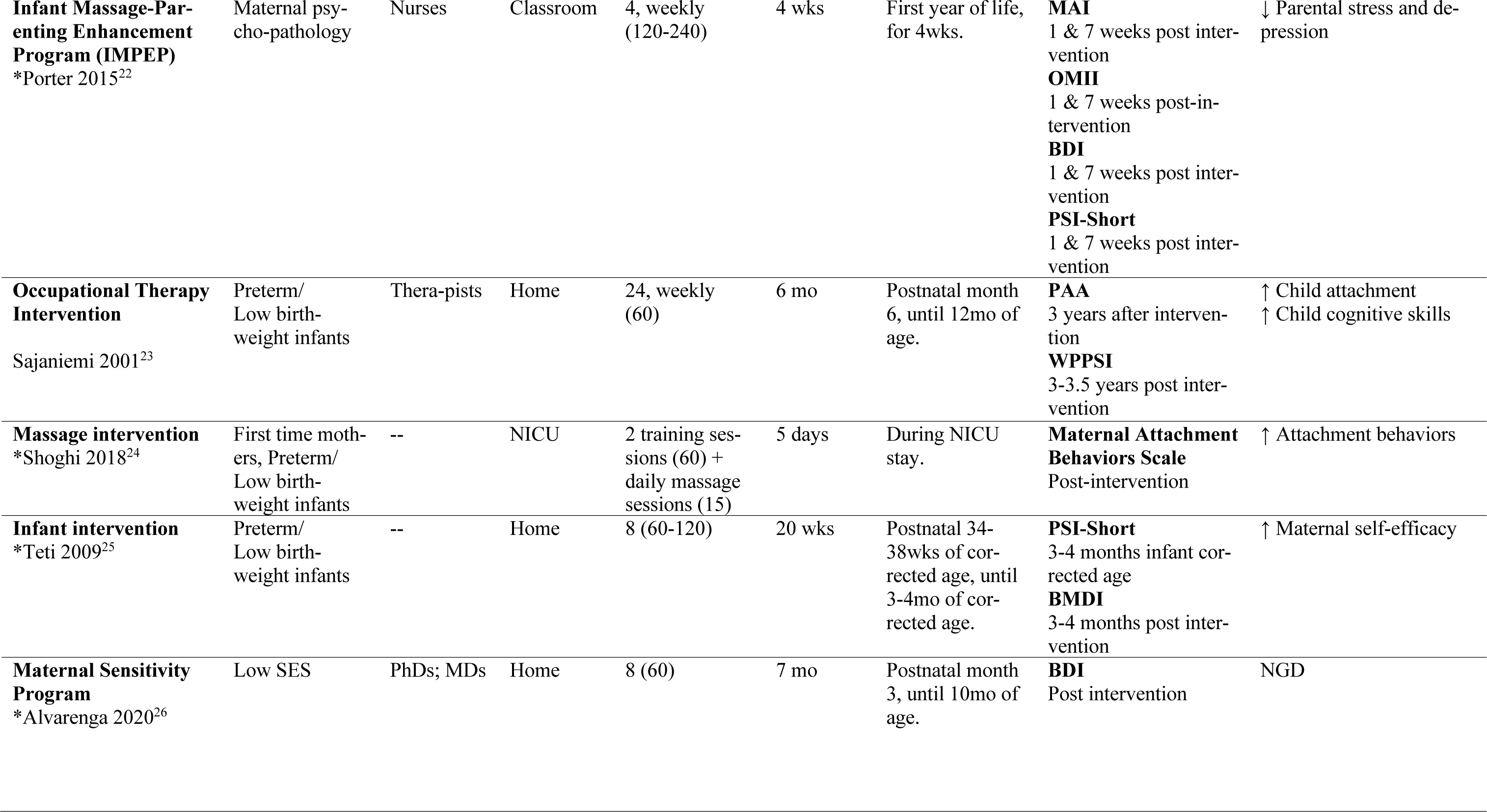
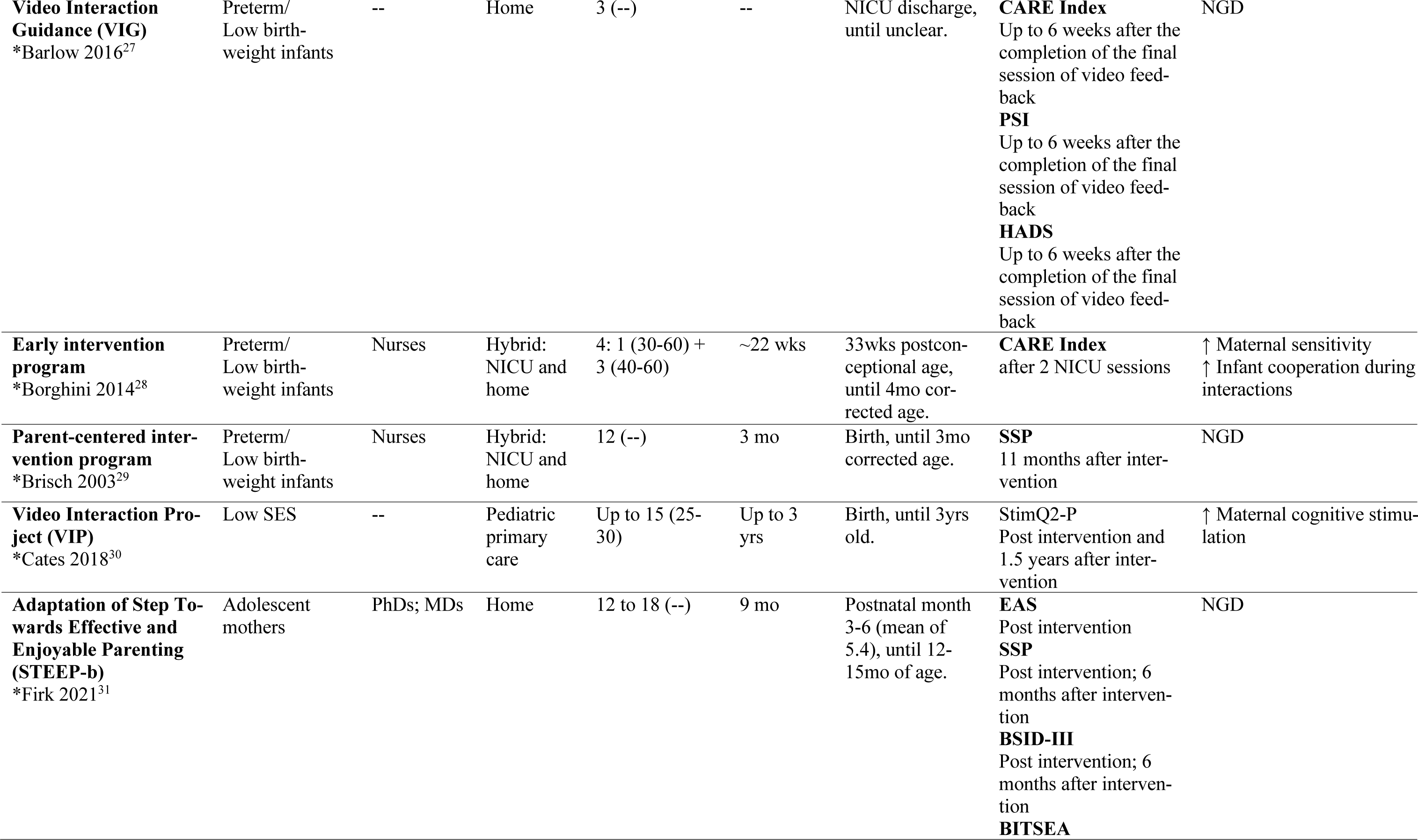
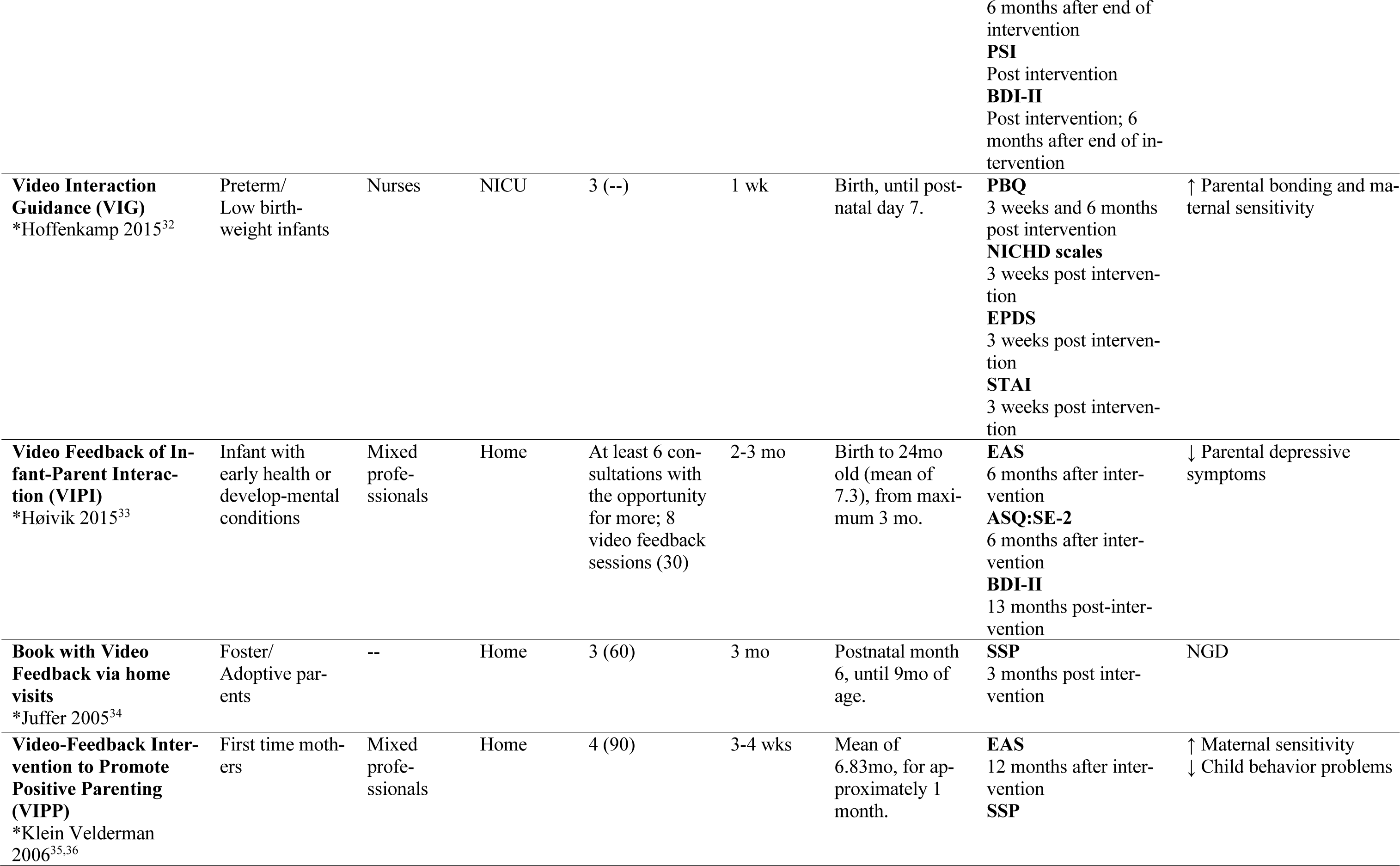
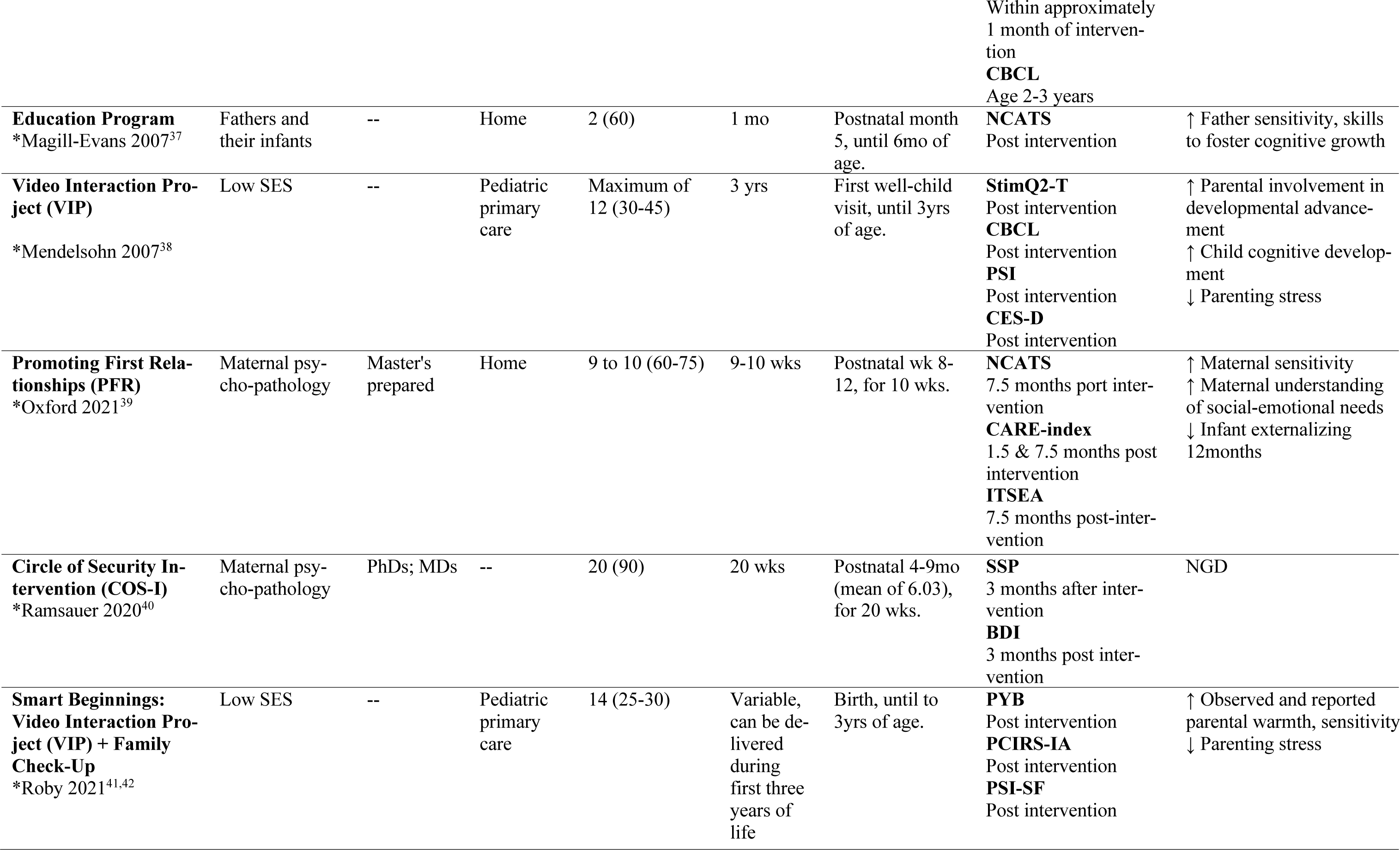
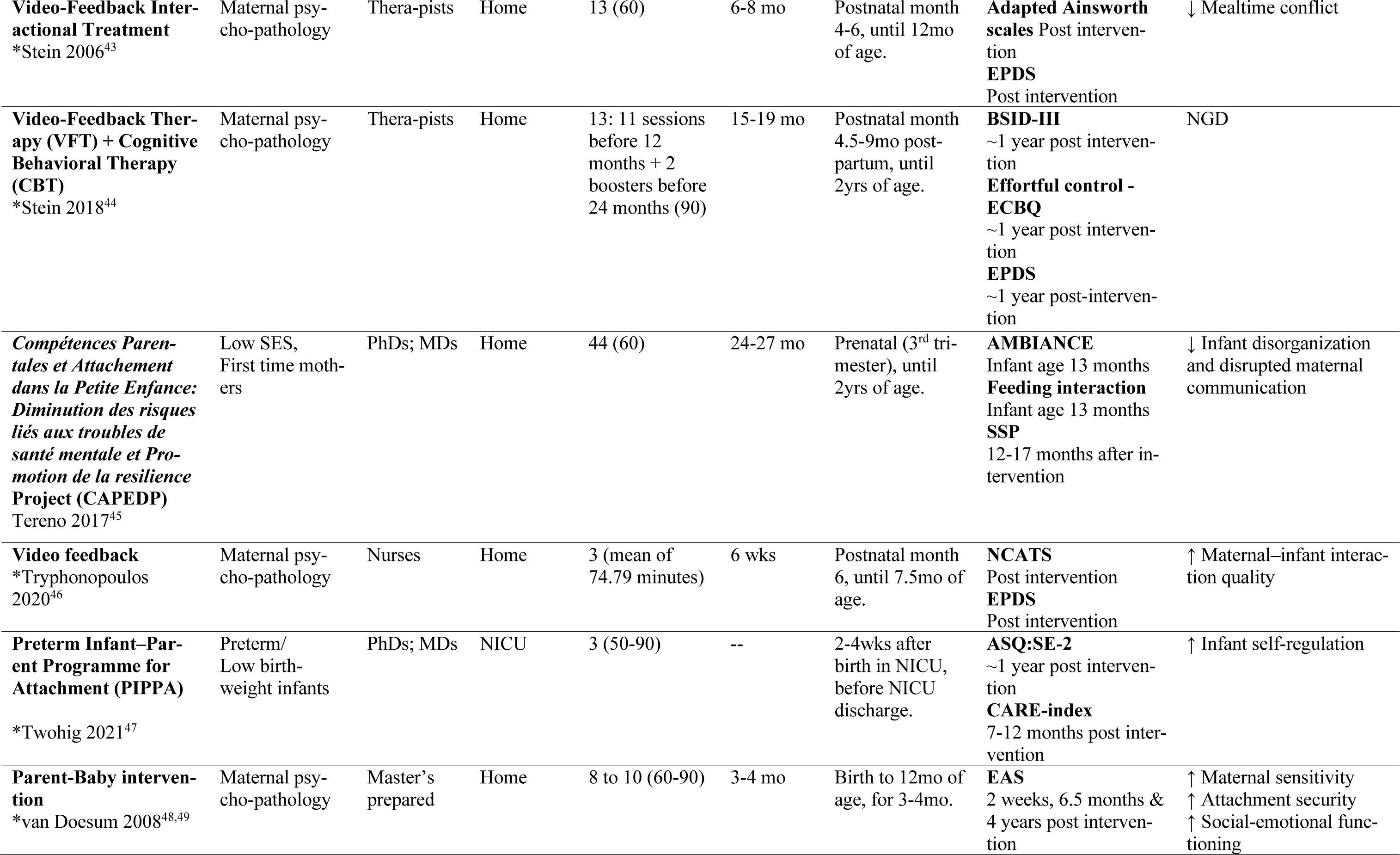
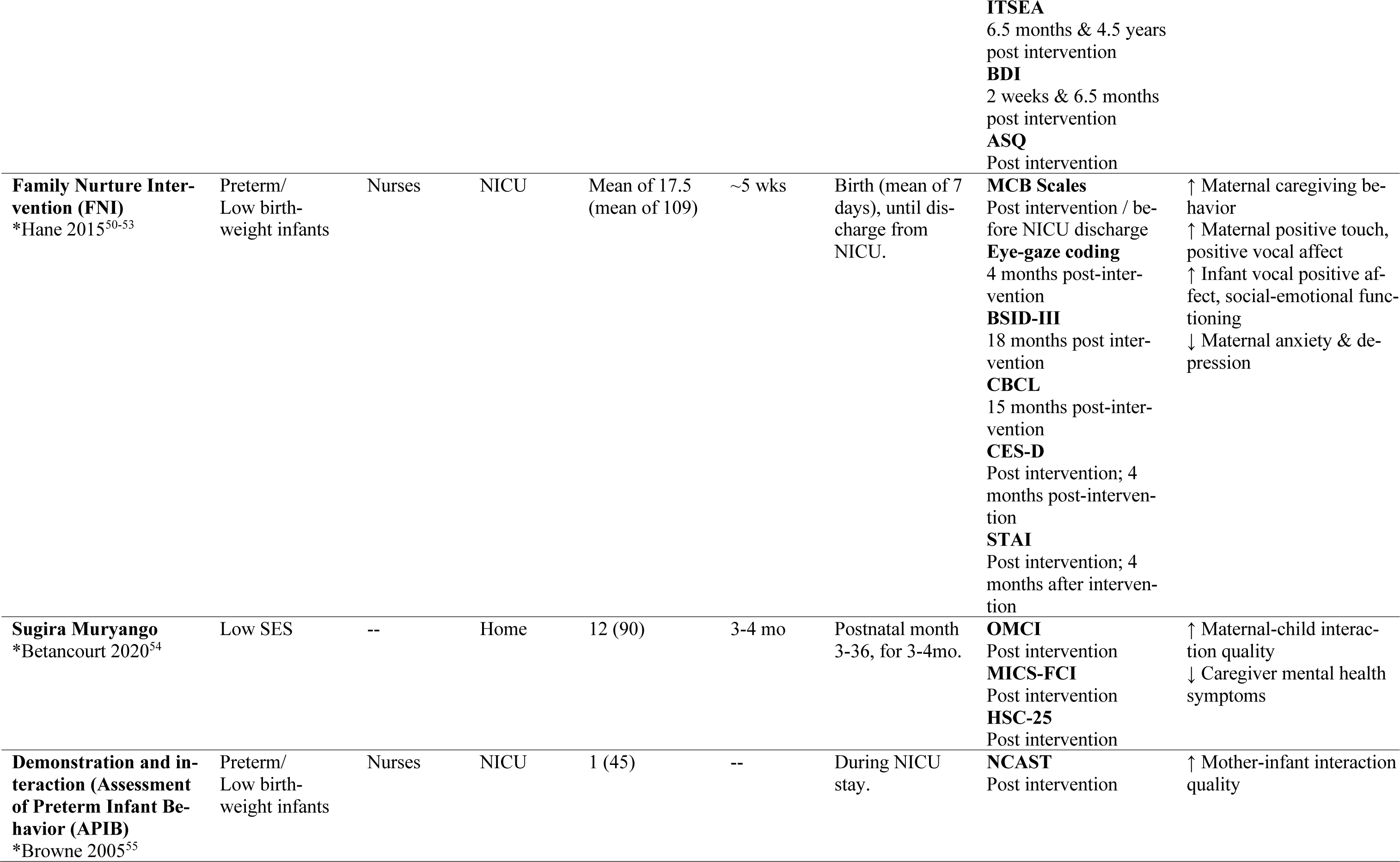
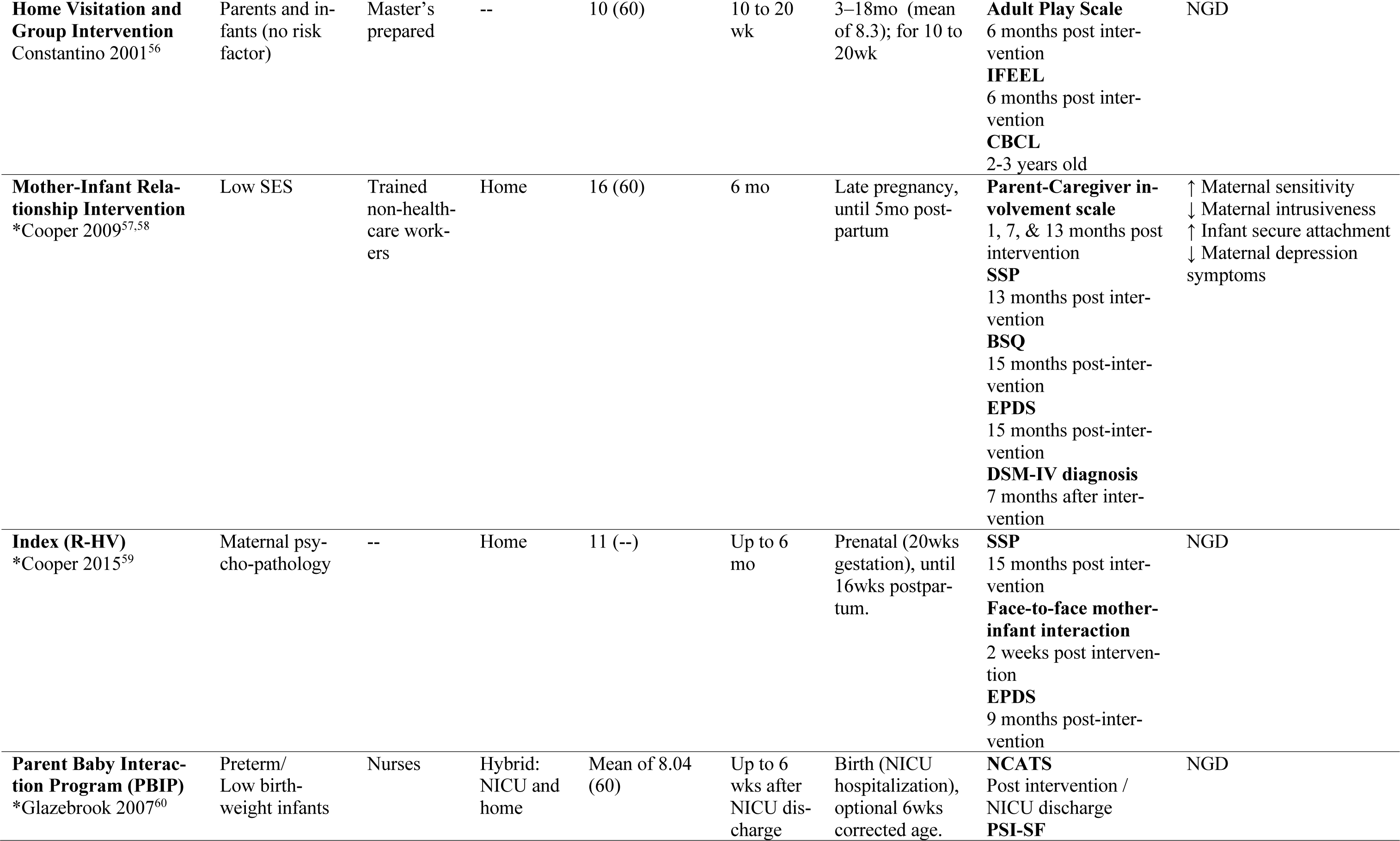
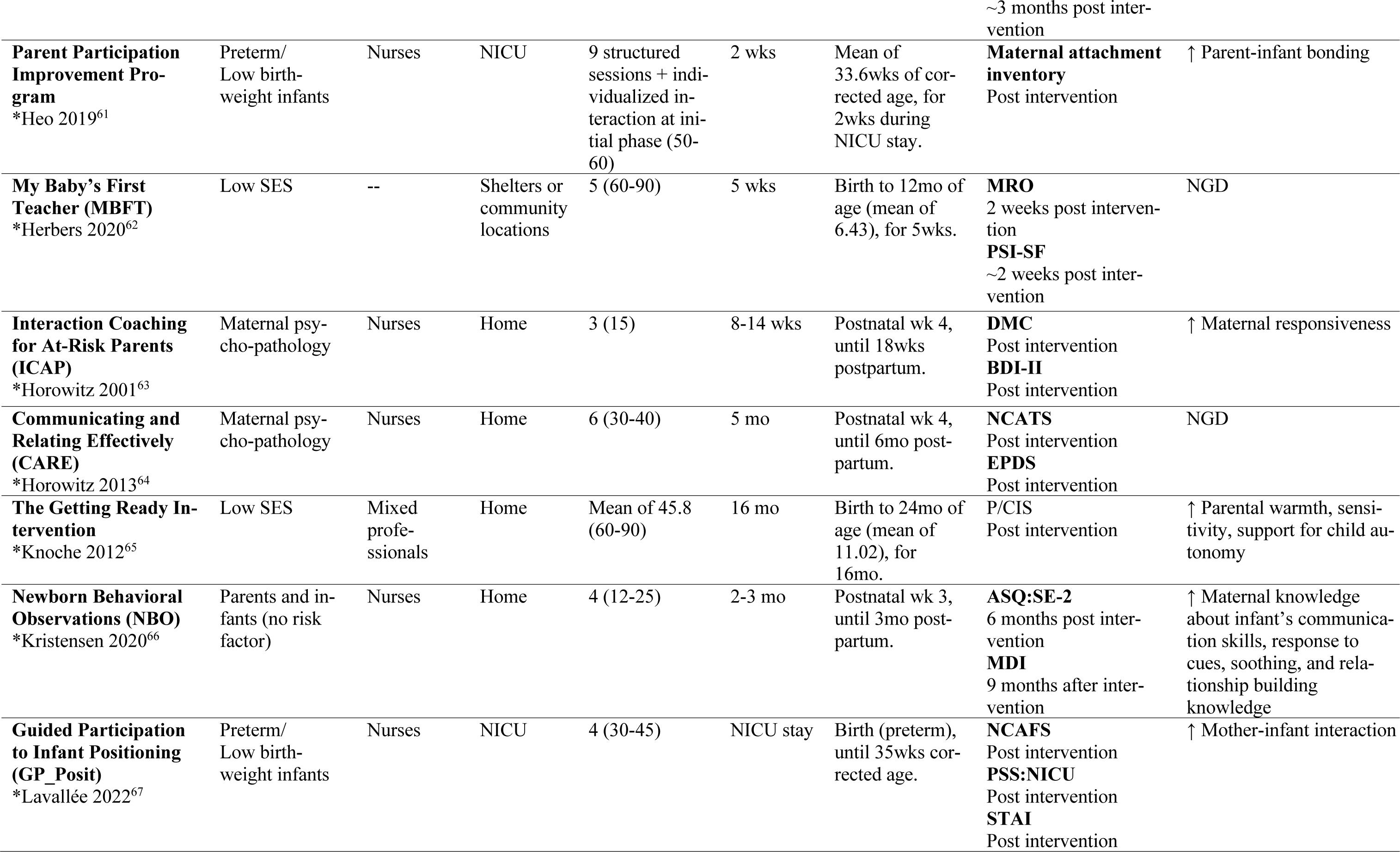
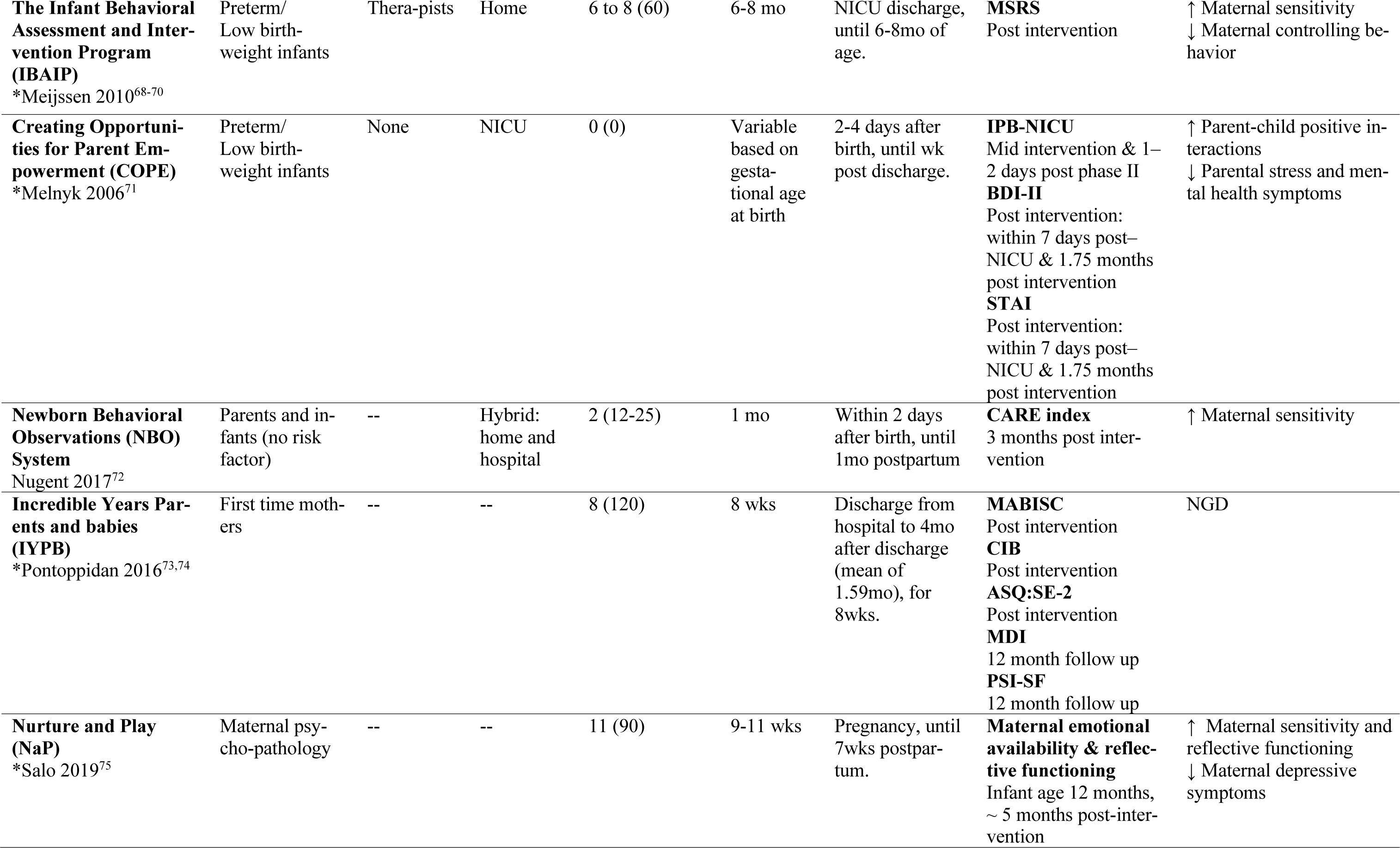
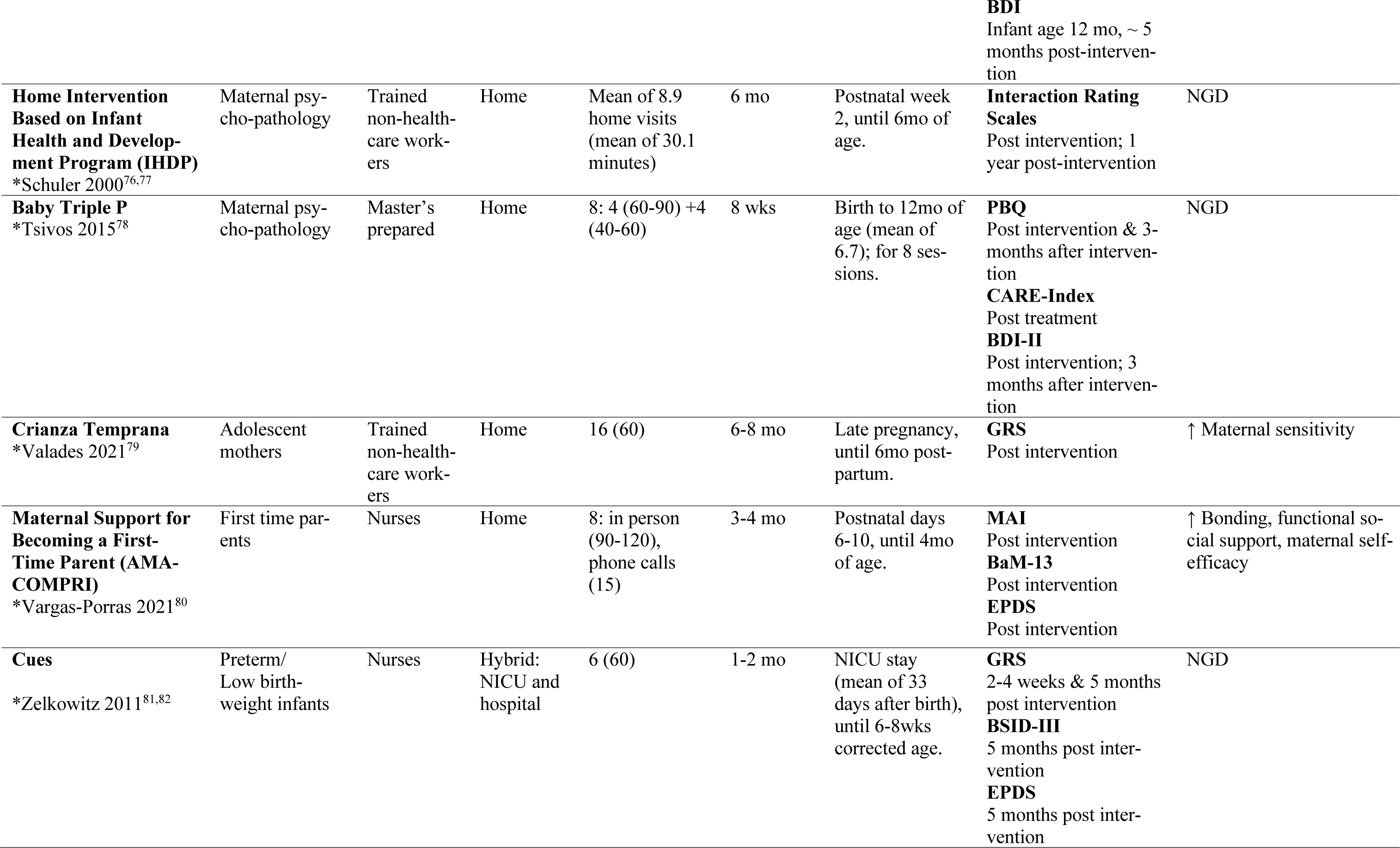
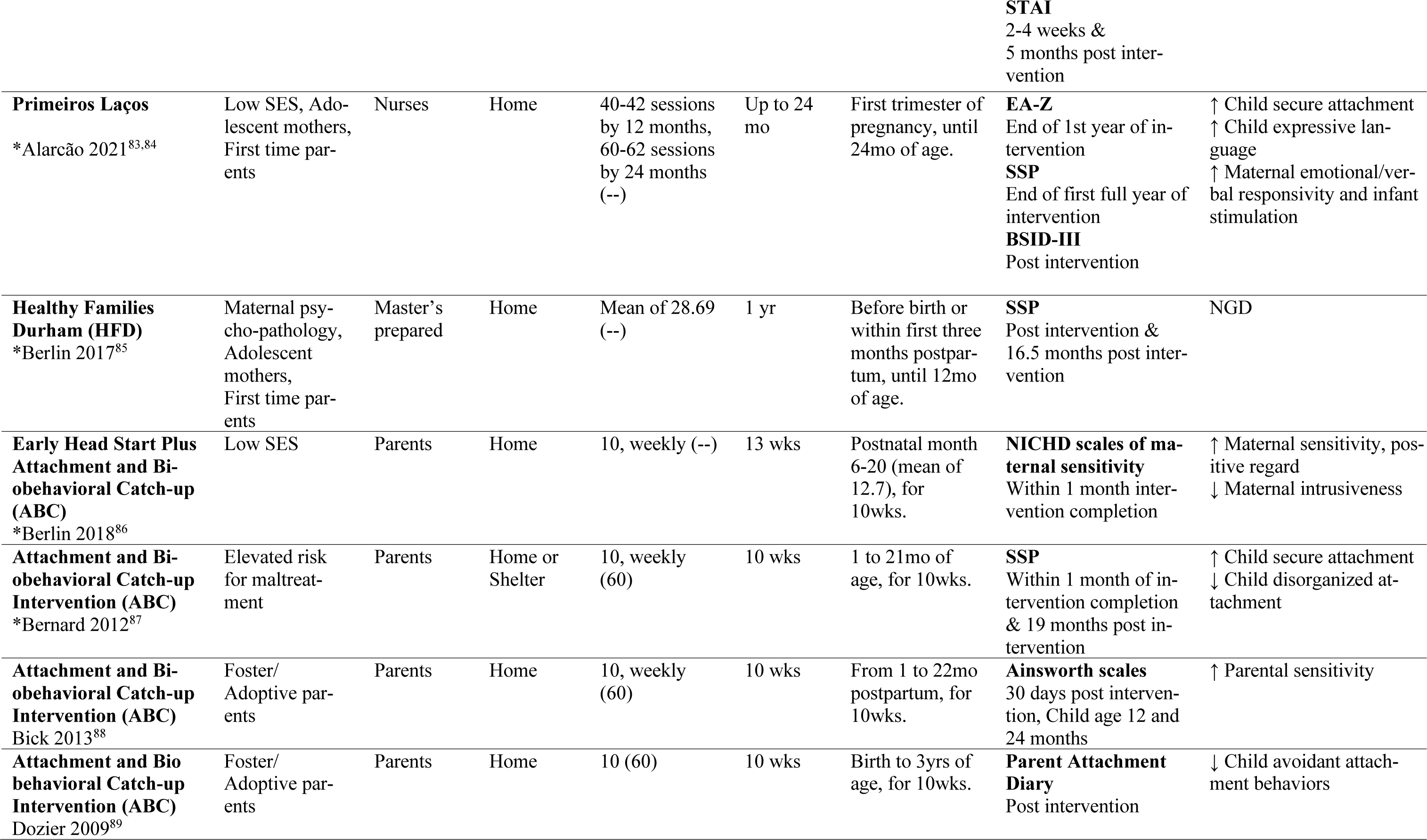
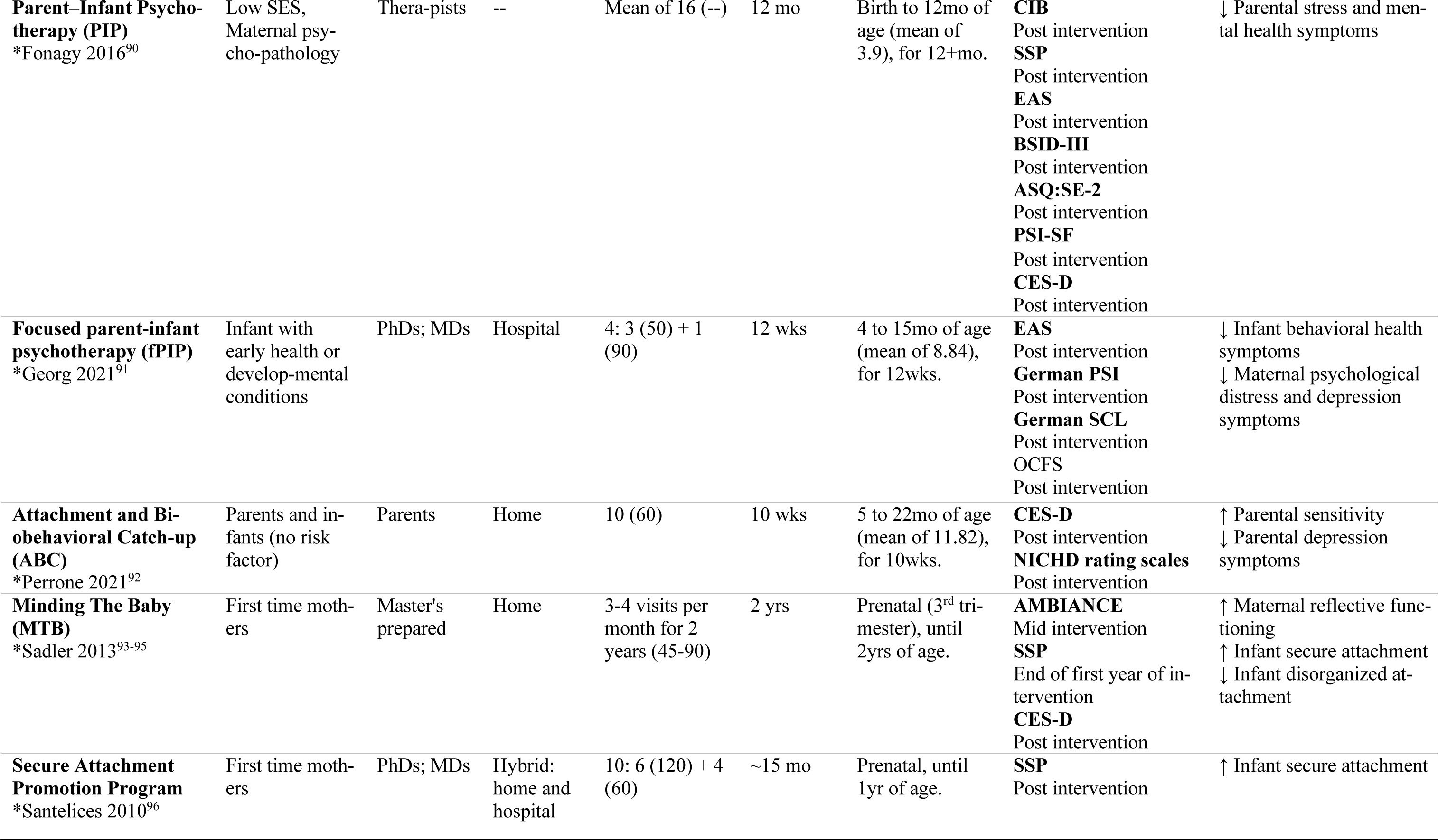
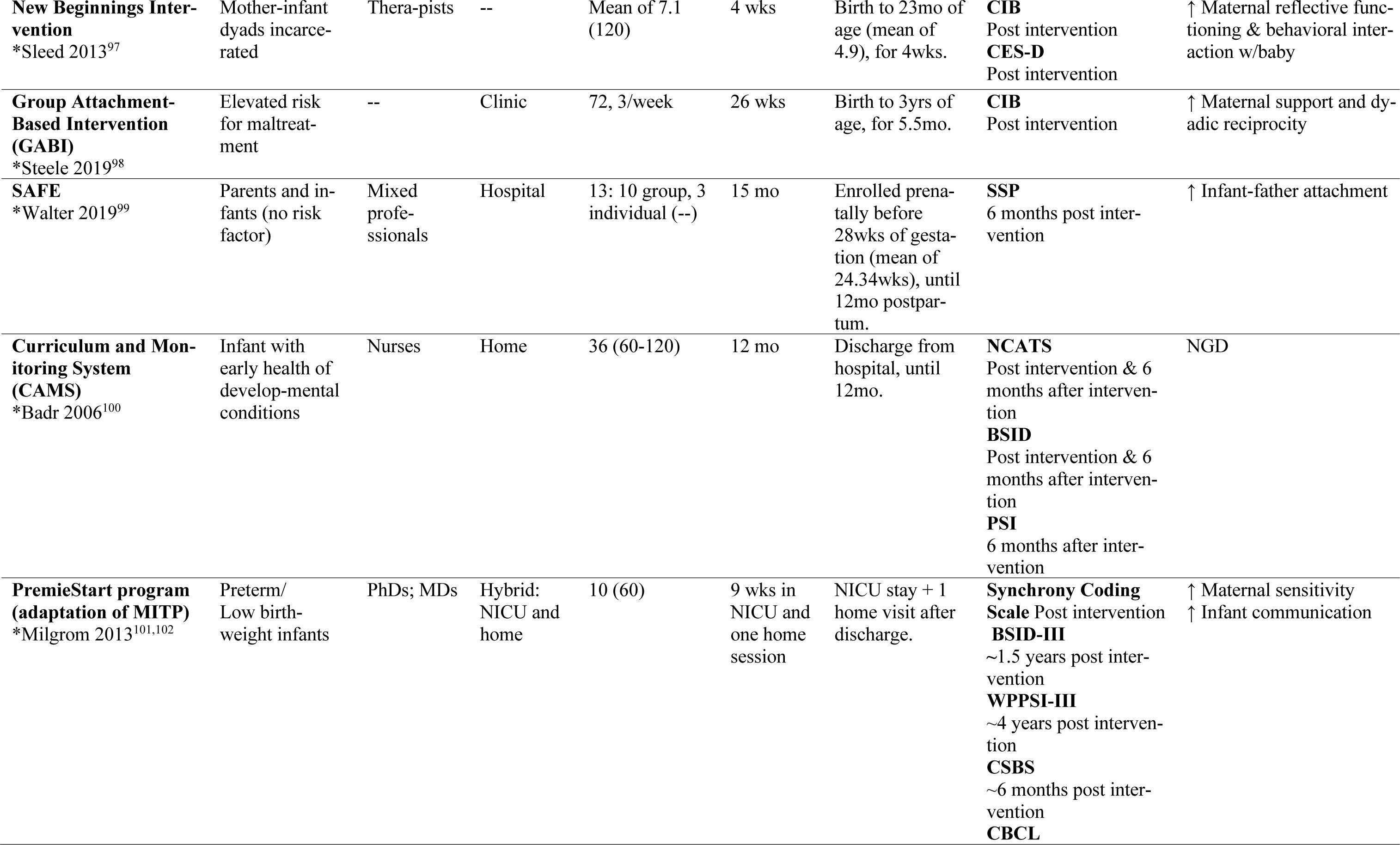
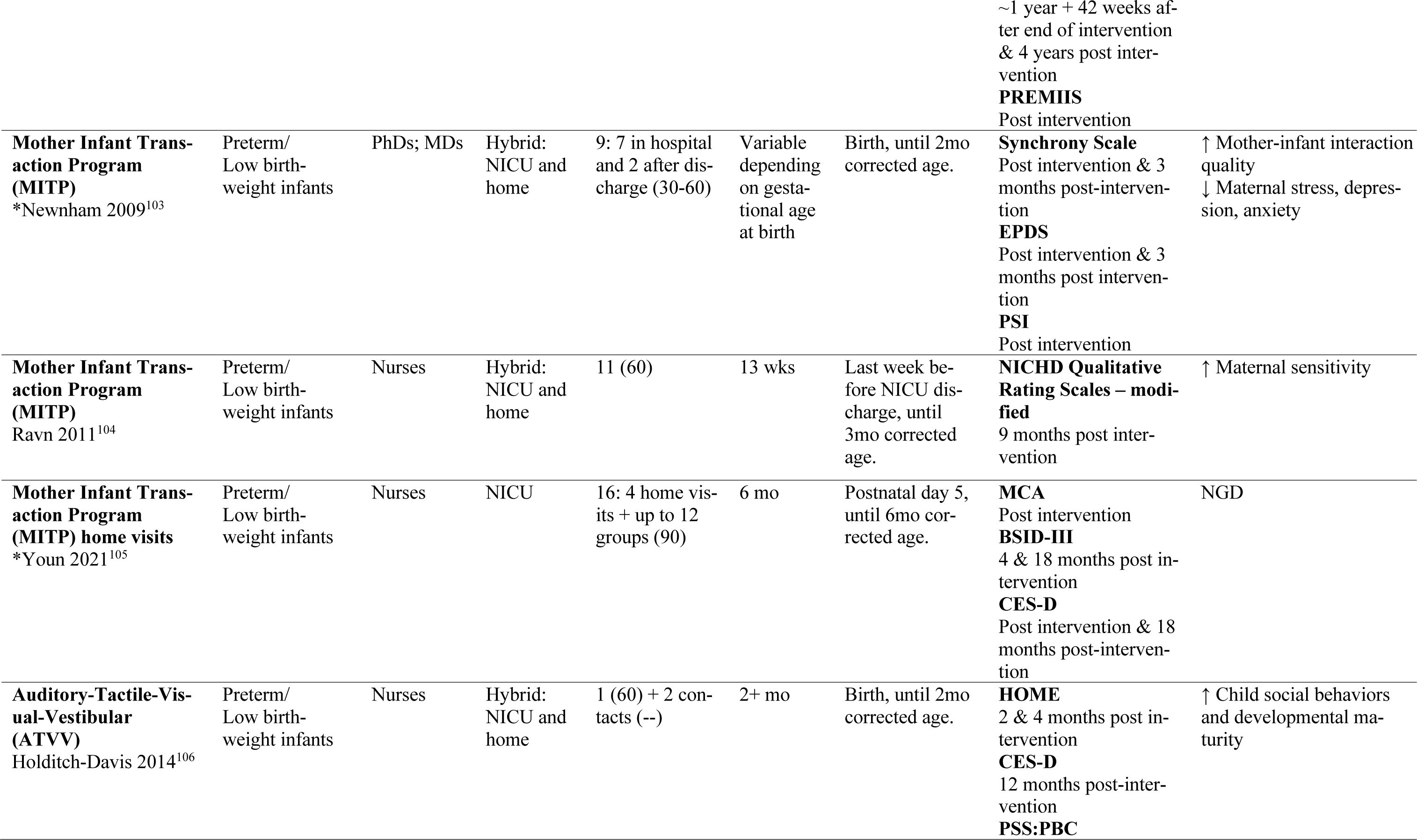
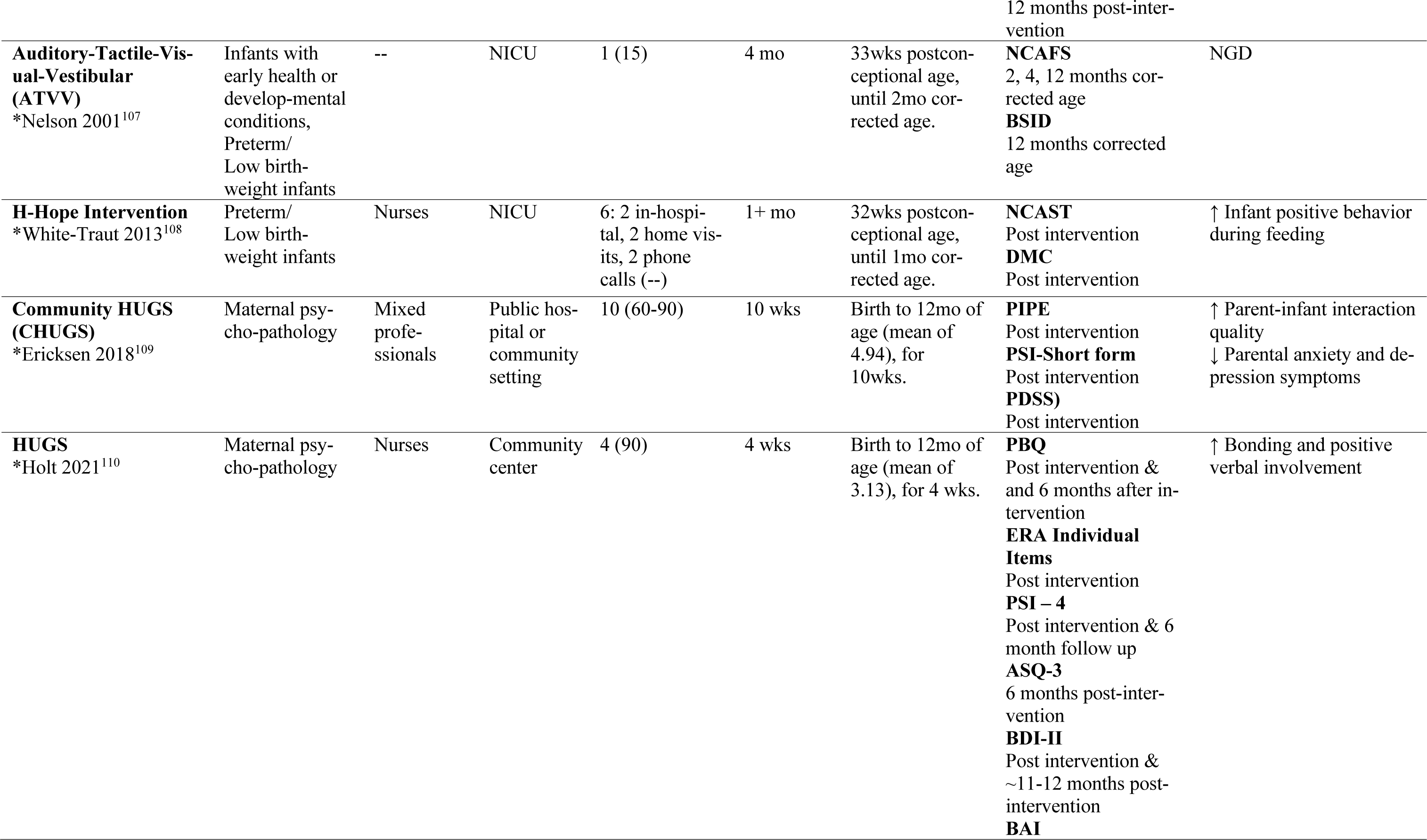
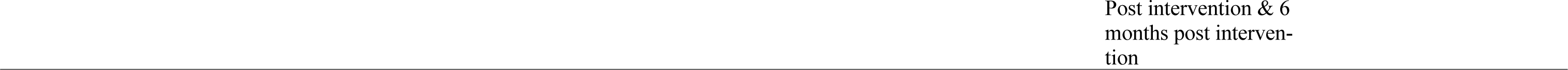
Intervention Characteristics

### Meta-Analysis Results

Meta-analytic results are shown in Figure 2 (and eFigure2 to 45). Sensitivity analyses are presented in eTable4 and moderator analyses in eFigure1.

**Figure 2.**
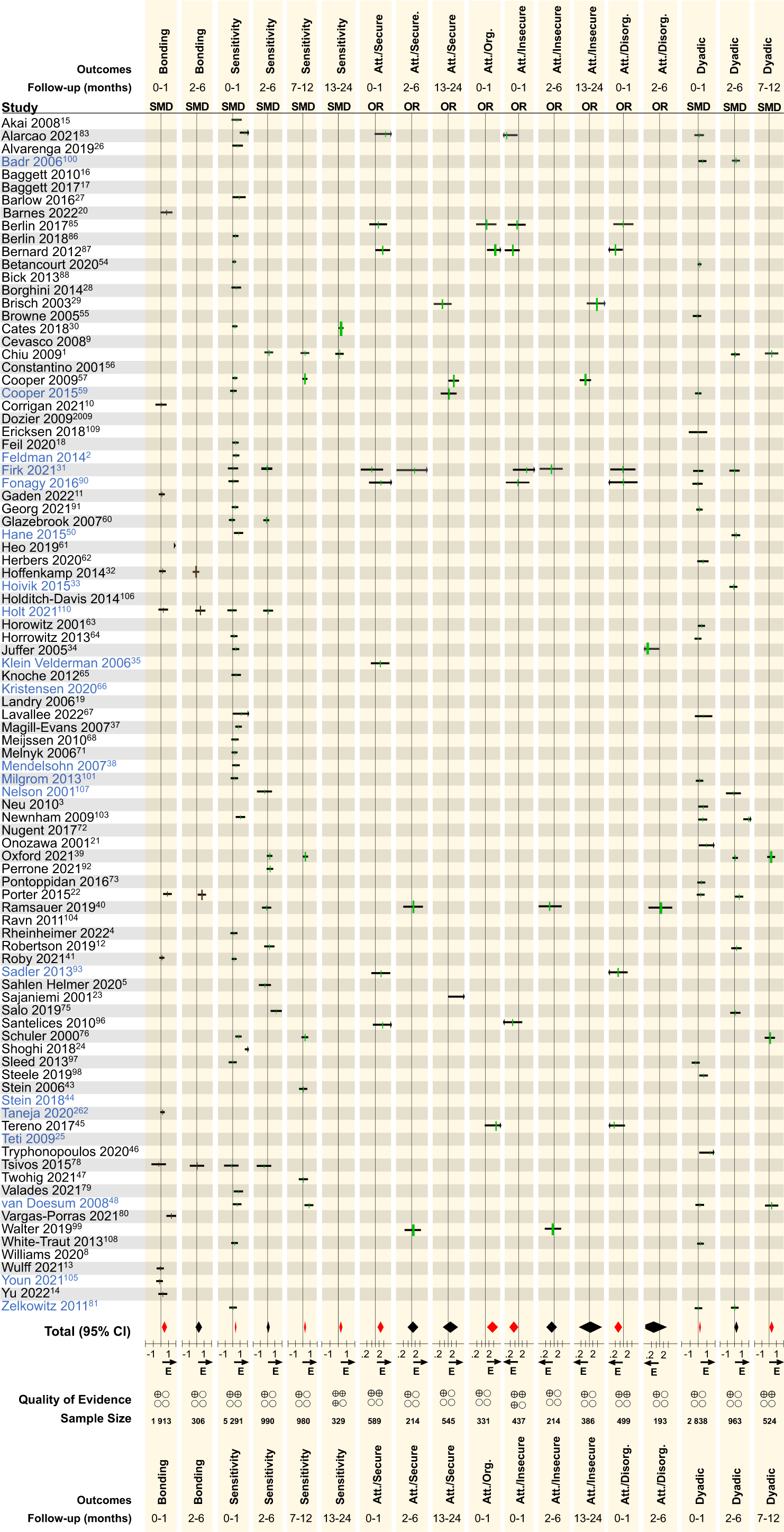
Meta-Analytic Results. Brown effect sizes: self-reported outcomes. Green effect sizes: observational outcomes. Black horizontal bars: 95% Con fidence Intervals. Red diamonds: significant effects. Blue font: study with data in at least one child development outcome. Att.: Attachment. SMD: Stand ardized Mean Differences. OR: Odds Ratios. E: Favors Experimental. Quality of Evidence: +Very Low, ++Low, +++Moderate, +++High.

#### Dyadic Interventions Promote ERH

Immediately post-intervention, parents/caregivers self-report significantly higher levels of bonding (k=14; n=1,913; SMD=0.43; 95%CI=[0.08, 0.78]; p=0.02; I^2^=91%), but not at 2-6m F/U. Observational assessments of parent/caregiver behaviors show improved sensitivity/responsivity in the first month post-intervention through 24m F/U (0-1m F/U: k=38; n=5,291; SMD=0.36; 95%CI =[0.25, 0.47]; p=<0.00001; I^2^=69%; 7-12m F/U: k=7; n=980; SMD=0.27; 95%CI=[0.13, 0.42]; p<0.00001; I^2^=15%; 13-24m F/U (k=2; n=329; SMD=0.40; 95%CI=[0.10, 0.70]; p=0.008; I^2^=74%), except at 2-6m F/U.

Dyadic interventions are also effective in increasing odds of secure attachment immediately post-intervention (k=8; n=589; OR=1.69; 95%CI=[1.20, 2.39]; p=0.003; I^2^=0%), but not at later F/U. Similarly, interventions increase odds of organized attachment (k=3; n=331; OR=2.23; 95%CI=[1.21, 4.11]; p=0.01; I^2^=21%), and decrease odds of insecure attachment (k=6; n=437; OR=0.60; 95%CI=[0.36, 0.99]; p=0.05; I^2^=30%) and disorganized attachment (k=6; n=499; OR=0.51; 95%CI=[0.33, 0.77]; p=0.002; I^2^=0%) immediately post-intervention. No other significant differences were found on insecure, disorganized, or insecure attachment.

Finally, parent/caregiver-child dyadic interactions are significantly increased immediately post-intervention (k=25; n=2,838; SMD=0.22; 95%CI=[0.10, 0.34]; p=0.0003; I^2^=46%), and 7-12m F/U (k=4; n=524; SMD=0.19; 95%CI=[0.02, 0.36]; p=0.03; I^2^=0%), but not 2-6m F/U.

Sensitivity analyses generally affirmed these results, though significant effect estimates may be inflated by high risk-of-biases, as bonding, attachment and dyadic interactions immediately post-intervention become non-significant in low risk-of-bias only studies (eTable4).

#### No Evidence of Effectiveness of Dyadic Interventions on Child Behavior, Socio-Emotional Functioning or Development

Parent/caregiver-reported child behavior (mostly on the Child Behavior CheckList) showed no improvement after participating in a dyadic intervention at any timepoint. Parent/caregiver-reported child socio-emotional functioning using the Ages and Stages Questionnaire: Socio-Emotional (ASQ:SE) was also non-significant at 2-6 months F/U. Observer-based assessments of child development (Bayley Scales of Infant Development) also did not show evidence of effectiveness of early dyadic interventions on cognitive, language, or motor development. To assess for lack of power as the main contributor of no effectiveness on developmental outcomes, pooled effect estimates of the subset of 20 studies that included a measure of development were conducted for ERH outcomes (eFigure46), showing significant differences on bonding (0-1m F/U: k=2; n=571; SMD=0.18; 95%CI=[0.01, 0.34], I^2^=0%) and sensitivity (0-1m F/U: k=10; n=888; SMD=0.23; 95%CI=[0.07, 0.40], I^2^=34%).

#### Dyadic Interventions Promote Lower Parent/Caregiver Anxiety, But Have No Effect on Parenting Stress and Depression

Pooled effect estimates showed that dyadic interventions significantly reduce parent/caregiver anxiety immediately post-intervention (k=11; n=1,168; SMD=0.16, 95%CI=[0.01, 0.30], p=0.03, I^2^=28%). No other significant effect was found on anxiety, parenting stress or depression.

### Association Between Potential Moderators and Effect Estimates

We were able to explore the association between potential moderators and nine effect estimates (eFigure1; bonding 0-1m F/U, sensitivity 0-1m and 2-6m F/U, dyadic interactions 0-1m F/U and 2-6m F/U, parenting stress 0-1m F/U, anxiety 0-1m F/U, and depression 0-1m F/U and 2-6m F/U). Unexpectedly, we did not find a significant association between our preregistered moderator, i.e., dyadic intervention dose, and any effect estimate (Figure 3). Among all other tested moderators, we only found a significant association between preregistered studies and bonding (*Q*=5.33; p=0.021), with greater effect sizes at the post-intervention follow-up in studies that were not preregistered.

**Figure 3.**
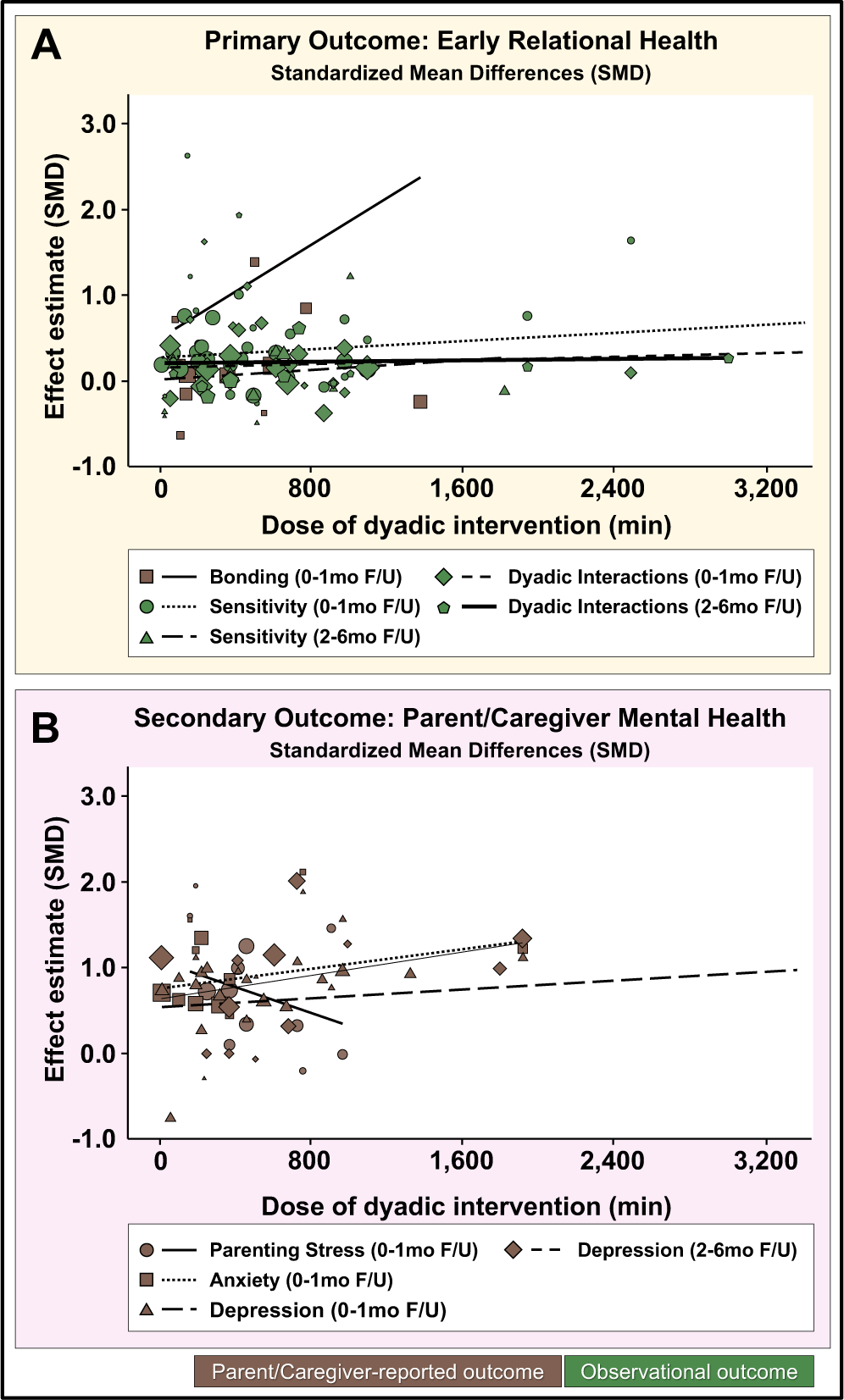
Overview of Meta-Regression Results. DerSimonian Laird model meta-regressions with minutes as predictors and Standardized Mean Differences (SMD) as outcomes. Analyses conducted only on outcomes pooling ≥10 studies. (**A**) No significant associations were identified in the primary outcomes of bonding 0-1mo F/U (β=.001, p=.39), sensitivity 0-1m F/U (β=.000, p=.45), sensitivity 2-6mo F/U (β=.000, p=.59), dyadic interactions 0-1mo F/U (β=5.36^-5^, p=.38), or dyadic interactions 2-6mo (β=3.20^-5^, p=.92). (**B**) No significant associations were identified for the secondary outcomes of parenting stress 0-1mo F/U (β=.000, p=.29), anxiety 0-1mo F/U (β=.001, p=.29), Depression 0-1mo F/U (β=7.96^-5^, p=.61) or depression 2-6mo F/U (β=.000, p=.29).

### Risk-of-Bias Within and Across Studies

Our results are mostly based on very low to moderate quality of evidence (eTable5) attributable to three factors: (1) moderate to high risk-of-biases in identified studies (eFigures48-103) driven by lack of blinding of participants, high drop-out rates, not using standardized or validated outcome assessments, selective outcome reporting, and lack of prospective registration; (2) high heterogeneity; and (3) imprecision in effect estimates.

## DISCUSSION

This systematic review identified 93 primary RCTs evaluating the effectiveness of dyadic interventions initiated within the first 6 months of life published from 2000 onwards. Our meta-analysis pooling 80 RCTs shows that dyadic interventions promote ERH, demonstrated by higher levels of bonding, higher parent/caregiver sensitivity, increased odds of secure attachment, decreased odds of disorganized attachment, and improvement of parent/caregiver-infant dyadic interactions in the first month post-intervention, though these effects are relatively small and mostly time-limited. However, contrasting the vast correlational literature associating strong ERH with better child outcomes, our meta-analysis failed to identify significant improvements on child socio-emotional, behavioral, or other developmental outcomes. While this in part could be attributed to an overall paucity of RCTs evaluating these outcomes, a lack of power could not fully explain this null finding as our sensitivity analyses including only the 20 RCT that measured developmental outcomes continued to show significant positive effects on ERH measures. Positive ERH is also strongly associated with improved parent/caregiver mental health.^8^ In this regard, our meta-analysis supports the evidence for dyadic interventions lowering anxiety, though no improvement in stress or depressive symptoms was observed.

The surprising finding that contemporary dyadic interventions bolster ERH without effectively targeting secondary child outcomes that are strongly associated with ERH should be interpreted primarily in the context of limitations in the scope of studies carried out in this field to-date rather than a definitive mechanistic lack of a causal link. Critically, significant effect estimates on ERH are of minimal to small magnitude, appear to fade over time, and represent mostly very low to moderate quality evidence. Additionally, only 20 RCTs, representing less than a quarter of the field, measured at least one child developmental outcome. Furthermore, moderator and sensitivity analyses suggest the possibility of a type I error, indicating that significant pooled effect estimates on ERH outcomes may be inflated by studies at higher risk-of-bias.

Another unique finding presented here is that, contrasting other reviews,^54^ we did not find significant associations between potential moderators and effect estimates. Nevertheless, the counterintuitive but optimistic finding that intervention effects on ERH are non-dose-dependent, affirming a prior meta-analysis result of ‘less is more’,^52^ suggests that promoting ERH is amenable to short, cost-effective interventions. Thus, investment in universal, widespread implementation of ‘light touch interventions’ in family-centered pediatric medical homes (FCPMH) has the potential to achieve public health benefits.

The AAP policy statement further emphasizes that FCPMH are integral to the universal promotion of ERH.^1^ Yet, only three identified RCTs^112,119,122^ were implemented by pediatricians or in FCPMH. Also, a majority of studies focused on biological mothers and parent/caregiver-infant dyads at high risk of impaired ERH. The ERH field has thus far neglected the development, evaluation, and implementation of more universal interventions that any parent/caregiver-infant dyad, including fathers,^177^ could benefit from.

## CONCLUSION

In the wake of AAP’s 2021 policy statement highlighting the buffering effects of the parent/caregiver-child relationship on the negative impact of toxic stress, ERH interventions are heralded to hold great promise. Meta-analyses presented here show that contemporary early dyadic interventions improve ERH non-dose-dependently, but effect sizes are currently small, time-limited and do not spill-over into other child outcomes (Figure 4). These results both offer glimmers of hope and demand us to embark on a comprehensive research agenda to develop and refine effective, scalable, equitable, evidence-based ERH interventions evaluated in high quality research.

**Figure 4.**
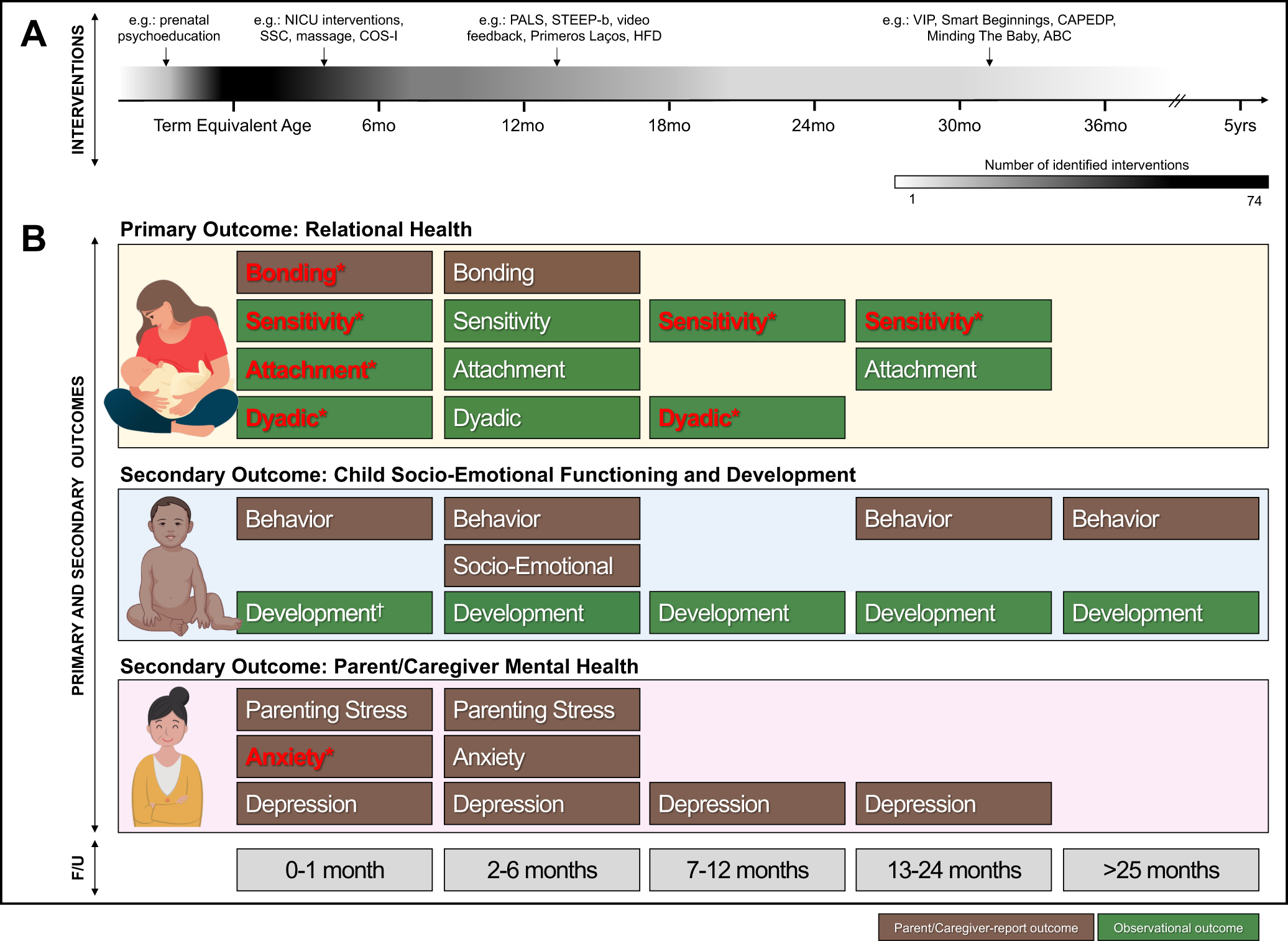
Contemporary landscape regarding efficacy of early dyadic interventions. (**A**) Number of identified interventions: cumulative number of interventions identified and active at specified age on timeline. (**B**) <u>Top:</u> Meta-analysis results indicate significant non-dose-dependent intervention effects on several measures of ERH, including bonding, parent/caregiver sensitivity, attachment, and dyadic interactions, and a significant effect on parent/caregiver anxiety, but no significant effects on secondary child outcomes. <u>Bottom</u>: Timeline indicating follow-up timing; **Bold/Red text**=significant effect; *Small effect size.

The field could benefit from a bold and iterative research agenda, done in partnership with parents/caregivers, researchers, funders, and pediatric clinicians. Upstream to driving early childhood policies, these results provide guidance for generating foundational knowledge of ERH, its life-course impact on real-world child and parent/caregiver outcomes, and in-depth mechanistic understanding of ERH promotion through universal innovations, for every family.

## AUTHOR CONTRIBUTION

Concept and design: Lavallée and Dumitriu. Acquisition, analysis, or interpretation of data: Lavallée, Warmingham, Atwood, Ahmed, Lanoff, Xu, Arduin, Hamer, Fischman, Ettinger, Hu, Fisher, Greeman, Kuromaru, Durr, Flowers, Pang, Lee and Gozali.

Drafting of the manuscript: Lavallée, Dumitriu, and Finkel. Ahmed and Lanoff (eMethods3 and eMethods4). Atwood, Hamer, Fischman, Kuromaru, Ahmed and Warmingham (eMethods5). Critical revision of the manuscript for important intellectual content: Lavallée, Warmingham, Willis and Dumitriu.

## Data Availability

All data produced in the present study are available upon reasonable request to the authors

## ACKNOWLEDGEMENTS

We thank Drs. Bernard and Badr, corresponding authors of studies identified by our systematic review, who agreed and shared unpublished data for inclusion in our meta-analysis. We also thank Drs. Bakermans-Kranenburg and White-Traut for being responsive to our request though additional data was unavailable for inclusion in our analysis. Finally, we also thank Nikki Shearman, PhD, partner in our ongoing exploration of ERH, who gave valuable input to the interpretation of our analysis. Drs. Andréane Lavallée and Dani Dumitriu had full access to all the data in the study and take responsibility for the integrity of the data and the accuracy of the data analysis.

## REFERENCES

1. Garner A, Yogman M, Committee On Psychosocial Aspects Of C, Family Health SOD, Behavioral Pediatrics COEC. Preventing Childhood Toxic Stress: Partnering With Families and Communities to Promote Relational Health. Pediatrics. 2021;148(2).

2. Cooke JE, Kochendorfer LB, Stuart-Parrigon KL, Koehn AJ, Kerns KA. Parent-child attachment and children’s experience and regulation of emotion: A meta-analytic review. Emotion. 2019;19(6):1103–1126.

3. Deneault AA, Hammond SI, Madigan S. A meta-analysis of child-parent attachment in early childhood and prosociality. Dev Psychol. 2023;59(2):236–255.

4. Le Bas GA, Youssef GJ, Macdonald JA, et al. The role of antenatal and postnatal maternal bonding in infant development: A systematic review and meta-analysis. Social Development. 2020;29(1):3–20.

5. Madigan S, Prime H, Graham SA, et al. Parenting Behavior and Child Language: A Meta-analysis. Pediatrics. 2019;144(4).

6. Pallini S, Morelli M, Chirumbolo A, Baiocco R, Laghi F, Eisenberg N. Attachment and attention problems: A meta-analysis. Clin Psychol Rev. 2019;74:101772.

7. Rodrigues M, Sokolovic N, Madigan S, et al. Paternal Sensitivity and Children’s Cognitive and Socioemotional Outcomes: A Meta-Analytic Review. Child Dev. 2021;92(2):554–577.

8. Lieberman AF. Child-Parent Psychotherapy: A Relationship-Based Approach to the Treatment of Mental Health Disorders in Infancy and Early Childhood. 2004.

9. Oh DL, Jerman P, Silverio Marques S, et al. Systematic review of pediatric health outcomes associated with childhood adversity. BMC Pediatr. 2018;18(1):83.

10. Traub F, Boynton-Jarrett R. Modifiable Resilience Factors to Childhood Adversity for Clinical Pediatric Practice. Pediatrics. 2017;139(5).

11. Felitti VJ, Anda RF, Nordenberg D, et al. Relationship of childhood abuse and household dysfunction to many of the leading causes of death in adults. The Adverse Childhood Experiences (ACE) Study. Am J Prev Med. 1998;14(4):245–258.

12. Morris AS, Robinson LR, Hays-Grudo J, Claussen AH, Hartwig SA, Treat AE. Targeting Parenting in Early Childhood: A Public Health Approach to Improve Outcomes for Children Living in Poverty. Child Dev. 2017;88(2):388–397.

13. Boyce WT, Levitt P, Martinez FD, McEwen BS, Shonkoff JP. Genes, Environments, and Time: The Biology of Adversity and Resilience. Pediatrics. 2021;147(2).

14. Short AK, Baram TZ. Early-life adversity and neurological disease: age-old questions and novel answers. Nat Rev Neurol. 2019;15(11):657–669.

15. Wesarg C, Van den Akker AL, Oei NYL, et al. Childhood adversity and vagal regulation: A systematic review and meta-analysis. Neuroscience & Biobehavioral Reviews. 2022:104920.

16. Scully C, McLaughlin J, Fitzgerald A. The relationship between adverse childhood experiences, family functioning, and mental health problems among children and adolescents: a systematic review. Journal of Family Therapy. 2020;42(2):291–316.

17. Gilbert LK, Breiding MJ, Merrick MT, et al. Childhood adversity and adult chronic disease: an update from ten states and the District of Columbia, 2010. Am J Prev Med. 2015;48(3):345-349.

18. Hughes K, Bellis MA, Hardcastle KA, et al. The effect of multiple adverse childhood experiences on health: a systematic review and meta-analysis. Lancet Public Health. 2017;2(8):e356–e366.

19. Bellis MA, Hughes K, Ford K, Ramos Rodriguez G, Sethi D, Passmore J. Life course health consequences and associated annual costs of adverse childhood experiences across Europe and North America: a systematic review and meta-analysis. Lancet Public Health. 2019;4(10):e517–e528.

20. AAP, AACAP, CHA. AAP-AACAP-CHA Declaration of a National Emergency in Child and Adolescent Mental Health. 2021. https://www.aap.org/en/advocacy/child-and-adolescent-healthy-mental-development/aap-aacap-cha-declaration-of-a-national-emergency-in-child-and-adolescent-mental-health/.

21. AAP, AACAP, CHA. Health Organizations Urge the Biden Administration to Declare a Federal National Emergency in Children’s Mental Health. 2022. https://www.aap.org/en/news-room/news-releases/aap/2022/health-organizations-urge-the-biden-administration-to-declare-a-federal-national-emergency-in-childrens-mental-health/.

22. Wittkowski A, Vatter S, Muhinyi A, Garrett C, Henderson M. Measuring bonding or attachment in the parent-infant-relationship: A systematic review of parent-report assessment measures, their psychometric properties and clinical utility. Clin Psychol Rev. 2020;82:101906.

23. Koehn AJ, Kerns KA. Parent-child attachment: meta-analysis of associations with parenting behaviors in middle childhood and adolescence. Attach Hum Dev. 2018;20(4):378–405.

24. Klaus MH, Kenell JH. Parent-Infant bonding. Vol 2: Mosby; 1982.

25. Bicking Kinsey C, Hupcey JE. State of the science of maternal-infant bonding: a principle-based concept analysis. Midwifery. 2013;29(12):1314–1320.

26. Leerkes EM, Zhou N. Maternal sensitivity to distress and attachment outcomes: Interactions with sensitivity to nondistress and infant temperament. J Fam Psychol. 2018;32(6):753–761.

27. Deans CL. Maternal sensitivity, its relationship with child outcomes, and interventions that address it: a systematic literature review. Early Child Development and Care. 2018;190(2):252–275.

28. De Wolff M, Van Ijzendoorn MH. Sensitivity and attachment: A meta-analysis on parental antecedents of infant attachment. Child Development. 1997;68:571–591.

29. Nievar M, Becker BM. Sensitivity is a priviledged predictor of attachment: A second perspective on de Wolff and van Ijzendoorn’s meta-analysis. Social Development. 2008;17:102–114.

30. Cassidy J, Shaver PR. Handbook of Attachment: Theory, Research and Clinical Applications. 3 ed: The Guilford Press; 2016.

31. Ainsworth MD, Blehar MC, Waters E, Wall SN. Pattern of Attachment: A Psychological Study of the Strange Situation. Routledge; 2015.

32. Bowlby J. Loss and Sadness and Depression. Vol 3. New York: Basic Books; 1980.

33. Gross JT, Stern JA, Brett BE, Cassidy J. The multifaceted nature of prosocial behavior in children: Links with attachment theory and research. Social Development. 2017;26(4):661–678.

34. Bowlby J. Attachment and Loss. Vol Vol. 1. 2nd ed ed. New York: Basic Books; 1988.

35. Mikulincer M, Shaver PR. An attachment perspective on psychopathology. World Psychiatry. 2012;11(1):11–15.

36. Mikulincer M, Shaver PR, Berant E. An attachment perspective on therapeutic processes and outcomes. J Pers. 2013;81(6):606–616.

37. Cantazaro A, Wei M. Adult attachment, dependence, self-criticism, and depressive symptoms: a test of a mediational model. J Pers. 2010;78(4):1135–1162.

38. Bosmans G, Braet C, Van Vlierberghe L. Attachment and symptoms of psychopathology: early maladaptive schemas as a cognitive link? Clin Psychol Psychother. 2010;17(5):374–385.

39. Illing V, Tasca GA, Balfour L, Bissada H. Attachment insecurity predicts eating disorder symptoms and treatment outcomes in a clinical sample of women. J Nerv Ment Dis. 2010;198(9):653–659.

40. Ein-Dor T, Doron G, Solomon Z, Mikulincer M, Shaver PR. Together in pain: attachment-related dyadic processes and posttraumatic stress disorder. J Couns Psychol. 2010;57(3):317–327.

41. Doron G, Moulding R, Kyrios M, Nedeljkovic M, Mikulincer M. Adult attachment insecurities are related to obsessive compulsive phenomena Journal of Social and Clinical Psychology. 2009;28(8):1022–1049.

42. Main M, Solomon J. Procedures for identifying infants as disorganized/disoriented during the Ainsworth Strange Situation. In: Attachment in the preschool years: Theory, research, and intervention. Chicago, IL, US: University of Chicago Press; 1990:121-160.

43. Granqvist P, Sroufe LA, Dozier M, et al. Disorganized attachment in infancy: a review of the phenomenon and its implications for clinicians and policy-makers. Attach Hum Dev. 2017;19(6):534–558.

44. Hane AA, LaCoursiere JN, Mitsuyama M, et al. The Welch Emotional Connection Screen: validation of a brief mother-infant relational health screen. Acta Paediatr. 2019;108(4):615–625.

45. Biringen Z, Robinson JL, Emde RN. The Emotional Availability Scale. 3 ed. Department of Human Development and Family Studies, Colorado State University: Fort Collins; 1998.

46. Crittenden PM. CARE-Index: Coding Manual. In:1979-2004.

47. Feldman R. Coding interactive behavior manual. In. Bar-Ilan University, Israel1998.

48. Censullo M, Bowler R, Lester B, Brazelton TB. An instrument for the measurement of infant-adult synchrony. Nursing Research. 1987;36(4):244–248.

49. van den Dries L, Juffer F, van Ijzendoorn MH, Bakermans-Kranenburg MJ. Fostering security? A meta-analysis of attachment in adopted children. Children and Youth Services Review. 2009;31(3):410–421.

50. Mountain G, Cahill J, Thorpe H. Sensitivity and attachment interventions in early childhood: A systematic review and meta-analysis. Infant Behav Dev. 2017;46:14–32.

51. Jeong J, Franchett EE, Ramos de Oliveira CV, Rehmani K, Yousafzai AK. Parenting interventions to promote early child development in the first three years of life: A global systematic review and meta-analysis. PLoS Med. 2021;18(5):e1003602.

52. Bakermans-Kranenburg MJ, van IMH, Juffer F. Less is more: meta-analyses of sensitivity and attachment interventions in early childhood. Psychol Bull. 2003;129(2):195–215.

53. Bakermans-Kranenburg MJ, Van IMH, Juffer F. Disorganized infant attachment and preventive interventions: A review and meta-analysis. Infant Ment Health J. 2005;26(3):191–216.

54. Facompre CR, Bernard K, Waters TEA. Effectiveness of interventions in preventing disorganized attachment: A meta-analysis. Dev Psychopathol. 2018;30(1):1–11.

55. Wright B, Fearon P, Garside M, et al. Routinely used interventions to improve attachment in infants and young children: a national survey and two systematic reviews. Health Technol Assess. 2023;27(2):1–226.

56. Shah R, Kennedy S, Clark MD, Bauer SC, Schwartz A. Primary Care-Based Interventions to Promote Positive Parenting Behaviors: A Meta-analysis. Pediatrics. 2016;137(5).

57. Regalado M, Halfon N. Primary care services promoting optimal child development from birth to age 3 years: review of the literature. Arch Pediatr Adolesc Med. 2001;155(12):1311–1322.

58. Peacock-Chambers E, Ivy K, Bair-Merritt M. Primary Care Interventions for Early Childhood Development: A Systematic Review. Pediatrics. 2017;140(6).

59. Prime H, Andrews K, Markwell A, et al. Positive Parenting and Early Childhood Cognition: A Systematic Review and Meta-Analysis of Randomized Controlled Trials. Clin Child Fam Psychol Rev. 2023.

60. Herd M, Whittingham K, Sanders M, Colditz P, Boyd RN. Efficacy of preventative parenting interventions for parents of preterm infants on later child behavior: a systematic review and meta-analysis. Infant Ment Health J. 2014;35(6):630–641.

61. Zhang L, Ssewanyana D, Martin MC, et al. Supporting Child Development Through Parenting Interventions in Low-to Middle-Income Countries: An Updated Systematic Review. Front Public Health. 2021;9:671988.

62. Frosch CA, Schoppe-Sullivan SJ, O’Banion DD. Parenting and Child Development: A Relational Health Perspective. Am J Lifestyle Med. 2021;15(1):45–59.

63. Willis DW, Eddy JM. Early relational health: Innovations in child health for promotion, screening, and research. Infant Ment Health J. 2022;43(3):361–372.

64. U.S. Department of Agriculture, U.S. Department of Health and Human Services. Dietary Guidelines for Americans, 2020-2025. In: 9 ed.2020: DietaryGuidelines.gov.

65. World Health Organization. Infant and young child feeding. https://www.who.int/news-room/fact-sheets/detail/infant-and-young-child-feeding. Published 2021. Accessed.

66. Pattison KL, Kraschnewski JL, Lehman E, et al. Breastfeeding initiation and duration and child health outcomes in the first baby study. Prev Med. 2019;118:1–6.

67. Victora CG, Bahl R, Barros AJ, et al. Breastfeeding in the 21st century: epidemiology, mechanisms, and lifelong effect. Lancet. 2016;387(10017):475–490.

68. Hu Y, Chen Y, Liu S, et al. Breastfeeding duration modified the effects of neonatal and familial risk factors on childhood asthma and allergy: a population-based study. Respir Res. 2021;22(1):41.

69. Roth MC, Humphreys KL, King LS, Gotlib IH, Robakis TK. Breastfeeding Difficulties Predict Mothers’ Bonding with Their Infants from Birth to Age Six Months. Matern Child Health J. 2021;25(5):777–785.

70. Hoekzema E, van Steenbergen H, Straathof M, et al. Mapping the effects of pregnancy on resting state brain activity, white matter microstructure, neural metabolite concentrations and grey matter architecture. Nat Commun. 2022;13(1):6931.

71. Page MJ, McKenzie JE, Bossuyt PM, et al. The PRISMA 2020 statement: an updated guideline for reporting systematic reviews. BMJ. 2021;372:n71.

72. EndNote [computer program]. Version EndNote X9. Philadelphia, PA: Clarivate; 2013.

73. *Covidence systematic review software* [computer program]. Melbourne, Australia: Veritas Health Innovation; 2022.

74. Hoffmann TC, Glasziou PP, Boutron I, et al. Better reporting of interventions: template for intervention description and replication (TIDieR) checklist and guide. BMJ. 2014;348:g1687.

75. Higgins JPT, Li T, Deeks JJ. Chapter 6: Choosing effect measures and computing estimates of effect. In: Higgins JPT, Thomas J, Chandler J, et al., eds. Cochrane Handbook for Systematic Reviews of Interventions. Vol 6.3. Cochrane; 2022.

76. *Engauge Digitizer* [computer program]. Version 12.1 2020.

77. *PlotDigitizer* [computer program]. 2022.

78. *SPSS Meta-Analysis Macro* [computer program]. 2021.

79. Deeks JJ, Higgins JPT, Altman DG. Chapter 10: Analysing data and undertaking meta-analyses. In: Higgins JPT, Thomas J, Chandler J, et al., eds. Cochrane Handbook for Systematic Reviews of Interventions version 6.3. Cochrane; 2022.

80. *Review Manager (RevMan)* [computer program]. Version 5.4 2020.

81. *IBM SPSS Statistics for Macintosh* [computer program]. Version 28. Armonk, NY: IBM Corp; 2021.

82. Higgins JPT, Savović J, Page MJ, Elbers RG, Sterne JAC. Chapter 8: Assessing risk of bias in a randomized trial. In: Higgins J, Thomas J, Chandler J, et al., eds. Cochrane Handbook for Systematic Reviews of Interventions. 6.3 ed.: Cochrane; 2022.

83. Guyatt G, Oxman AD, Akl EA, et al. GRADE guidelines: 1. Introduction-GRADE evidence profiles and summary of findings tables. J Clin Epidemiol. 2011;64(4):383–394.

84. Chiu SH, Anderson GC. Effect of early skin-to-skin contact on mother-preterm infant interaction through 18 months: randomized controlled trial. Int J Nurs Stud. 2009;46(9):1168–1180.

85. Feldman R, Rosenthal Z, Eidelman AI. Maternal-Preterm Skin-to-Skin Contact Enhances Child Physiologic Organization and Cognitive Control Across the First 10 Years of Life. Biological Psychiatry. 2014;75(1):56–64.

86. Neu M, Robinson J. Maternal holding of preterm infants during the early weeks after birth and dyad interaction at six months. J Obstet Gynecol Neonatal Nurs. 2010;39(4):401–414.

87. Rheinheimer N, Beijers R, Cooijmans KHM, Brett BE, de Weerth C. Effects of skin - to - skin contact on full - term infants’ stress reactivity and quality of mother–infant interactions. Developmental Psychobiology. 2022;64(7).

88. Sahlen Helmer C, Birberg Thornberg U, Frostell A, Ortenstrand A, Morelius E. A Randomized Trial of Continuous Versus Intermittent Skin-to-Skin Contact After Premature Birth and the Effects on Mother-Infant Interaction. Adv Neonatal Care. 2020;20(3):E48–E56.

89. Taneja S, Sinha B, Upadhyay RP, et al. Community initiated kangaroo mother care and early child development in low birth weight infants in India-a randomized controlled trial. BMC Pediatr. 2020;20(1):150.

90. Williams LR, Turner PR. Infant carrying as a tool to promote secure attachments in young mothers: Comparing intervention and control infants during the still-face paradigm. Infant Behav Dev. 2020;58:101413.

91. Cevasco AM. The Effects of Mothers’ Singing on Full-term and Preterm Infants and Maternal Emotional Responses. Journal of Music Therapy. 2008;45(3):273–306.

92. Corrigan M, Keeler J, Miller H, Naylor C, Diaz A. Music Therapy and Family-Integrated Care in the NICU: Using Heartbeat-Music Interventions to Promote Mother-Infant Bonding. Adv Neonatal Care. 2022;22(5):E159–E168.

93. Gaden TS, Ghetti C, Kvestad I, et al. Short-term Music Therapy for Families With Preterm Infants: A Randomized Trial. Pediatrics. 2022;149(2).

94. Robertson AM, Detmer MR. The Effects of Contingent Lullaby Music on Parent-Infant Interaction and Amount of Infant Crying in the First Six Weeks of Life. J Pediatr Nurs. 2019;46:33–38.

95. Wulff V, Hepp P, Wolf OT, Fehm T, Schaal NK. The influence of maternal singing on well-being, postpartum depression and bonding - a randomised, controlled trial. BMC Pregnancy Childbirth. 2021;21(1):501.

96. Yu WC, Chiang MC, Lin KC, Chang CC, Lin KH, Chen CW. Effects of maternal voice on pain and mother-Infant bonding in premature infants in Taiwan: A randomized controlled trial. J Pediatr Nurs. 2022;63:e136–e142.

97. Akai CE, Guttentag CL, Baggett KM, Noria CC, Centers for the Prevention of Child N. Enhancing parenting practices of at-risk mothers. J Prim Prev. 2008;29(3):223–242.

98. Baggett K, Davis B, Feil E, et al. A Randomized Controlled Trial Examination of a Remote Parenting Intervention: Engagement and Effects on Parenting Behavior and Child Abuse Potential. Child Maltreat. 2017;22(4):315–323.

99. Baggett KM, Davis B, Feil EG, et al. Technologies for expanding the reach of evidence-based interventions: Preliminary results for promoting social-emotional development in early childhood. Topics Early Child Spec Educ. 2010;29(4):226–238.

100. Feil EG, Baggett K, Davis B, et al. Randomized control trial of an internet-based parenting intervention for mothers of infants. Early Child Res Q. 2020;50(Pt 1):36–44.

101. Landry SH, Smith KE, Swank PR. Responsive parenting: establishing early foundations for social, communication, and independent problem-solving skills. Dev Psychol. 2006;42(4):627–642.

102. Barnes C, N. Adamson-Macedo E. Understanding the impact of newborn touch upon mothers of hospitalized preterm neonates. Journal of Human Growth and Development. 2022;32(2):294–301.

103. Onozawaa K, Gloverb V, Adamsb D, Modib N, Kumara RC. Infant massage improves mother–infant interaction for mothers with postnatal depression. Journal of Affective Disorders. 2001;63:201–207.

104. Porter LS, Porter BO, McCoy V, et al. Blended Infant Massage-Parenting Enhancement Program on Recovering Substance-Abusing Mothers’ Parenting Stress, Self-Esteem, Depression, Maternal Attachment, and Mother-Infant Interaction. Asian Nurs Res (Korean Soc Nurs Sci). 2015;9(4):318–327.

105. Sajaniemi N, Makela J, Salokorpi T, von Wendt L, Hamalainen T, Hakamies-Blomqvist L. Cognitive performance and attachment patterns at four years of age in extremely low birth weight infants after early intervention. Eur Child Adolesc Psychiatry. 2001;10(2):122–129.

106. Shoghi M, Sohrabi S, Rasouli M. The Effects of Massage by Mothers on Mother-Infant Attachment. Alternative therapies in health and medicine. 2018;24(3):34–39.

107. Teti DM, Black MM, Viscardi R, et al. Intervention With African American Premature Infants: Four-Month Results of an Early Intervention Program. Journal of Early Intervention. 2009;31(2):146–166.

108. Alvarenga P, Cerezo MA, Wiese E, Piccinini CA. Effects of a short video feedback intervention on enhancing maternal sensitivity and infant development in low-income families. Attach Hum Dev. 2020;22(5):534–554.

109. Barlow J, Sembi S, Underdown A. Pilot RCT of the use of video interactive guidance with preterm babies. Journal of Reproductive and Infant Psychology. 2016;34(5):511–524.

110. Borghini A, Habersaat S, Forcada-Guex M, et al. Effects of an early intervention on maternal post-traumatic stress symptoms and the quality of mother-infant interaction: the case of preterm birth. Infant Behav Dev. 2014;37(4):624–631.

111. Brisch KH, Bechinger D, Betzler S, Heinemann H. Early preventive attachment-oriented psychotherapeutic intervention program with parents of a very low birthweight premature infant: results of attachment and neurological development. Attach Hum Dev. 2003;5(2):120–135.

112. Cates CB, Weisleder A, Berkule Johnson S, et al. Enhancing Parent Talk, Reading, and Play in Primary Care: Sustained Impacts of the Video Interaction Project. J Pediatr. 2018;199:49–56 e41.

113. Firk C, Dahmen B, Dempfle A, et al. A mother-child intervention program for adolescent mothers: Results from a randomized controlled trial (the TeeMo study). Dev Psychopathol. 2021;33(3):992–1005.

114. Hoffenkamp HN, Tooten A, Hall RA, et al. Effectiveness of hospital-based video interaction guidance on parental interactive behavior, bonding, and stress after preterm birth: A randomized controlled trial. J Consult Clin Psychol. 2015;83(2):416–429.

115. Hoivik MS, Lydersen S, Drugli MB, Onsoien R, Hansen MB, Nielsen TS. Video feedback compared to treatment as usual in families with parent-child interactions problems: a randomized controlled trial. Child Adolesc Psychiatry Ment Health. 2015;9:3.

116. Juffer F, Bakermans-Kranenburg MJ, van Ijzendoorn MH. The importance of parenting in the development of disorganized attachment: Evidence from a preventive intervention study in adoptive families. Journal of Child Psychology and Psychiatry. 2005;46(3):263–274.

117. Klein Velderman M, Bakermans-Kranenburg MJ, Juffer F, van IMH. Effects of attachment-based interventions on maternal sensitivity and infant attachment: differential susceptibility of highly reactive infants. J Fam Psychol. 2006;20(2):266–274.

118. Magill-Evans J, Harrison M, Benzies K, Gierl M, Kimak C. Effects of Parenting Education on First-Time Fathers’ Skills in Interactions with Their Infants. Fathering: A Journal of Theory, Research, and Practice about Men as Fathers. 2007;5(1):42–57.

119. Mendelsohn AL, Valdez PT, Flynn V, et al. Use of videotaped interactions during pediatric well-child care: impact at 33 months on parenting and on child development. J Dev Behav Pediatr. 2007;28(3):206–212.

120. Oxford ML, Hash JB, Lohr MJ, et al. Randomized trial of promoting first relationships for new mothers who received community mental health services in pregnancy. Dev Psychol. 2021;57(8):1228–1241.

121. Ramsauer B, Muhlhan C, Lotzin A, et al. Randomized controlled trial of the Circle of Security-Intensive intervention for mothers with postpartum depression: maternal unresolved attachment moderates changes in sensitivity. Attach Hum Dev. 2020;22(6):705–726.

122. Roby E, Miller EB, Shaw DS, et al. Improving Parent-Child Interactions in Pediatric Health Care: A Two-Site Randomized Controlled Trial. Pediatrics. 2021;147(3):e20201799.

123. Stein A, Netsi E, Lawrence PJ, et al. Mitigating the effect of persistent postnatal depression on child outcomes through an intervention to treat depression and improve parenting: a randomised controlled trial. The Lancet Psychiatry. 2018;5(2):134–144.

124. Stein A, Woolley H, Senior R, et al. Treating Disturbances in the Relationship Between Mothers With Bulimic Eating Disorders and Their Infants: A Randomized, Controlled Trial of Video Feedback. American Journal of Psychiatry. 2006;163:899–906.

125. Tereno S, Madigan S, Lyons-Ruth K, et al. Assessing a change mechanism in a randomized home-visiting trial: Reducing disrupted maternal communication decreases infant disorganization. Development and Psychopathology. 2017;29(2):637–649.

126. Tryphonopoulos PD, Letourneau N. Promising Results From a Video-Feedback Interaction Guidance Intervention for Improving Maternal-Infant Interaction Quality of Depressed Mothers: A Feasibility Pilot Study. Can J Nurs Res. 2020;52(2):74–87.

127. Twohig A, Murphy JF, McCarthy A, et al. The preterm infant-parent programme for attachment-PIPPA Study: a randomised controlled trial. Pediatr Res. 2021;90(3):617–624.

128. van Doesum KTM, Riksen-Walraven JM, Hosman CMH, Hoefnagels C. A Randomized Controlled Trial of a Home-Visiting Intervention Aimed at PreventingRelationship Problems in Depressed Mothers and Their Infants. Child Development. 2008;79(3):547–561.

129. Hane AA, Myers MM, Hofer MA, et al. Family Nurture Intervention Improves the Quality of Maternal Caregiving in the Neonatal Intensive Care Unit: Evidence from a Randomized Controlled Trial. Journal of Developmental and Behavioral Pediatrics. 2015;36:188–196.

130. Betancourt TS, Jensen SKG, Barnhart DA, et al. Promoting parent-child relationships and preventing violence via home-visiting: a pre-post cluster randomised trial among Rwandan families linked to social protection programmes. BMC Public Health. 2020;20(1):621.

131. Browne JV, Talmi A. Family-based intervention to enhance infant-parent relationships in the neonatal intensive care unit. J Pediatr Psychol. 2005;30(8):667–677.

132. Constantino JN, Hashemib N, Solisb E, et al. Supplementation of urban home visitation with a series of group meetings for parents and infants: results of a “realworld” randomized, controlled trial. Child Abuse & Neglect. 2001;25:1571– 1581.

133. Cooper PJ, De Pascalis L, Woolgar M, Romaniuk H, Murray L. Attempting to prevent postnatal depression by targeting the mother-infant relationship: a randomised controlled trial. Prim Health Care Res Dev. 2015;16(4):383–397.

134. Cooper PJ, Tomlinson M, Swartz L, et al. Improving quality of mother-infant relationship and infant attachment in socioeconomically deprived community in South Africa: randomised controlled trial. BMJ. 2009;338:b974.

135. Glazebrook C, Marlow N, Israel C, et al. Randomised trial of a parenting intervention during neonatal intensive care. Arch Dis Child Fetal Neonatal Ed. 2007;92(6):F438-443.

136. Heo YJ, Oh WO. The effectiveness of a parent participation improvement program for parents on partnership, attachment infant growth in a neonatal intensive care unit: A randomized controlled trial. Int J Nurs Stud. 2019;95:19–27.

137. Herbers JE, Cutuli JJ, Fugo PB, Nordeen ER, Hartman MJ. Promoting parent-infant responsiveness in families experiencing homelessness. Infant Ment Health J. 2020;41(6):811–820.

138. Horowitz JA, Bell M, Trybulski J, et al. Promoting Responsiveness between Mothers with Depressive Symptoms and Their Infants. Journal of Nursing Scholarship. 2001;33(4):323–329.

139. Horowitz JA, Murphy CA, Gregory K, Wojcik J, Pulcini J, Solon L. Nurse home visits improve maternal/infant interaction and decrease severity of postpartum depression. J Obstet Gynecol Neonatal Nurs. 2013;42(3):287–300.

140. Knoche L, Sheridan SM, Clark BL, et al. GETTING READY: RESULTS OF A RANDOMIZED TRIAL OF A RELATIONSHIP-FOCUSED INTERVENTION ON THE PARENT– INFANT RELATIONSHIP IN RURAL EARLY HEAD START. Educational Psychology Papers and Publications. 2012;207.

141. Kristensen IH, Juul S, Kronborg H. What are the effects of supporting early parenting by newborn behavioral observations (NBO)? A cluster randomised trial. BMC Psychol. 2020;8(1):107.

142. Lavallée A, Côté J, Luu TM, et al. Acceptability and feasibility of a nursing intervention to promote sensitive mother-infant interactions in the NICU. Journal of Neonatal Nursing. 2022.

143. Meijssen D, Wolf MJ, Koldewijn K, et al. The effect of the Infant Behavioral Assessment and Intervention Program on mother-infant interaction after very preterm birth. J Child Psychol Psychiatry. 2010;51(11):1287–1295.

144. Melnyk BM, Feinstein NF, Alpert-Gillis L, et al. Reducing premature infants’ length of stay and improving parents’ mental health outcomes with the Creating Opportunities for Parent Empowerment (COPE) neonatal intensive care unit program: a randomized, controlled trial. Pediatrics. 2006;118(5):e1414–1427.

145. Nugent JK, Bartlett JD, Von Ende A, Valim C. The Effects of the Newborn Behavioral Observations (NBO) System on Sensitivity in Mother–Infant Interactions. Infants & Young Children. 2017;30(4):257–268.

146. Pontoppidan M, Klest SK, Sandoy TM. The Incredible Years Parents and Babies Program: A Pilot Randomized Controlled Trial. PLoS One. 2016;11(12):e0167592.

147. Salo SJ, Flykt M, Makela J, et al. The effectiveness of Nurture and Play: a mentalisation-based parenting group intervention for prenatally depressed mothers. Prim Health Care Res Dev. 2019;20:e157.

148. Schuler ME, Nair P, Black MM, Kettinger L. Mother-infant interaction: effects of a home intervention and ongoing maternal drug use. J Clin Child Psychol. 2000;29(3):424–431.

149. Tsivos ZL, Calam R, Sanders MR, Wittkowski A. A pilot randomised controlled trial to evaluate the feasibility and acceptability of the Baby Triple P Positive Parenting Programme in mothers with postnatal depression. Clin Child Psychol Psychiatry. 2015;20(4):532–554.

150. Valades J, Murray L, Bozicevic L, et al. The impact of a mother-infant intervention on parenting and infant response to challenge: A pilot randomized controlled trial with adolescent mothers in El Salvador. Infant Ment Health J. 2021;42(3):400–412.

151. Vargas-Porras C, Roa-Diaz ZM, Hernandez-Hincapie HG, Ferre-Grau C, de Molina-Fernandez MI. Efficacy of a multimodal nursing intervention strategy in the process of becoming a mother: A randomized controlled trial. Res Nurs Health. 2021;44(3):424–437.

152. Zelkowitz P, Feeley N, Shrier I, et al. The Cues and Care Randomized Controlled Trial of a Neonatal Intensive Care Unit Intervention: Effects on Maternal Psychological Distress and Mother-Infant Interaction. Journal of Developmental and Behavioral Pediatrics. 2011;32:591–599.

153. Alarcao FSP, Shephard E, Fatori D, et al. Promoting mother-infant relationships and underlying neural correlates: Results from a randomized controlled trial of a home-visiting program for adolescent mothers in Brazil. Dev Sci. 2021;24(6):e13113.

154. Berlin LJ, Martoccio TL, Appleyard Carmody K, et al. Can typical US home visits affect infant attachment? Preliminary findings from a randomized trial of Healthy Families Durham. Attach Hum Dev. 2017;19(6):559–579.

155. Berlin LJ, Martoccio TL, Jones Harden B. Improving early head start’s impacts on parenting through attachment-based intervention: A randomized controlled trial. Dev Psychol. 2018;54(12):2316–2327.

156. Bernard K, Dozier M, Bick J, Lewis-Morrarty E, Lindhiem O, Carlson E. Enhancing attachment organization among maltreated children: results of a randomized clinical trial. Child Dev. 2012;83(2):623–636.

157. Bick J, Dozier M. The Effectiveness of an Attachment-Based Intervention in Promoting Foster Mothers’ Sensitivity toward Foster Infants. Infant Ment Health J. 2013;34(2):95–103.

158. Dozier M, Lindhiem O, Lewis E, Bick J, Bernard K, Peloso E. Effects of a Foster Parent Training Program on Young Children’s Attachment Behaviors: Preliminary Evidence from a Randomized Clinical Trial. Child Adolesc Social Work J. 2009;26(4):321–332.

159. Fonagy P, Sleed M, Baradon T. Randomized Controlled Trial of Parent-Infant Psychotherapy for Parents with Mental Health Problems and Young Infants. Infant Ment Health J. 2016;37(2):97–114.

160. Georg AK, Cierpka M, Schroder-Pfeifer P, Kress S, Taubner S. The Efficacy of Brief Parent-Infant Psychotherapy for Treating Early Regulatory Disorders: A Randomized Controlled Trial. J Am Acad Child Adolesc Psychiatry. 2021;60(6):723–733.

161. Perrone L, Imrisek SD, Dash A, Rodriguez M, Monticciolo E, Bernard K. Changing parental depression and sensitivity: Randomized clinical trial of ABC’s effectiveness in the community. Dev Psychopathol. 2021;33(3):1026–1040.

162. Sadler LS, Slade A, Close N, et al. Minding the Baby: Enhancing reflectiveness to improve early health and relationship outcomes in an interdisciplinary home visiting program. Infant Ment Health J. 2013;34(5):391–405.

163. Santelices MP, Guzman GM, Aracena M, et al. Promoting secure attachment: evaluation of the effectiveness of an early intervention pilot programme with mother-infant dyads in Santiago, Chile. Child Care Health Dev. 2011;37(2):203–210.

164. Sleed M, Baradon T, Fonagy P. New beginnings for mothers and babies in prison: A cluster randomized controlled trial. Attachment & Human Development. 2013;15(4):349–367.

165. Steele H, Murphy A, Bonuck K, Meissner P, Steele M. Randomized control trial report on the effectiveness of Group Attachment-Based Intervention (GABI(c)): Improvements in the parent-child relationship not seen in the control group. Dev Psychopathol. 2019;31(1):203–217.

166. Walter I, Landers S, Quehenberger J, Carlson E, Brisch KH. *The efficacy of the attachment-based SAFE(R) prevention program: a randomized control trial including mothers and fathers. Attach Hum Dev. 2019;21(5):510–531.

167. Badr LK, Garg M, Kamath M. Intervention for infants with brain injury: results of a randomized controlled study. Infant Behav Dev. 2006;29(1):80–90.

168. Milgrom J, Newnham C, Martin PR, et al. Early communication in preterm infants following intervention in the NICU. Early Hum Dev. 2013;89(9):755–762.

169. Newnham CA, Milgrom J, Skouteris H. Effectiveness of a modified Mother-Infant Transaction Program on outcomes for preterm infants from 3 to 24 months of age. Infant Behav Dev. 2009;32(1):17–26.

170. Ravn IH, Smith L, Lindemann R, et al. Effect of early intervention on social interaction between mothers and preterm infants at 12 months of age: a randomized controlled trial. Infant Behav Dev. 2011;34(2):215–225.

171. Youn YA, Shin SH, Kim EK, et al. Preventive Intervention Program on the Outcomes of Very Preterm Infants and Caregivers: A Multicenter Randomized Controlled Trial. Brain Sci. 2021;11(5).

172. Holditch-Davis D, White-Traut RC, Levy JA, O’Shea TM, Geraldo V, David RJ. Maternally administered interventions for preterm infants in the NICU: effects on maternal psychological distress and mother-infant relationship. Infant Behav Dev. 2014;37(4):695–710.

173. Nelson MN, White-Traut RC, Vasan U, et al. One-Year Outcome of Auditory-Tactile-Visual-Vestibular Intervention in the Neonatal Intensive Care Unit: Effects of Severe Prematurity and Central Nervous System Injury. Journal of Child Neurology. 2001;16:493–498).

174. White-Traut R, Norr KF, Fabiyi C, Rankin KM, Li Z, Liu L. Mother-infant interaction improves with a developmental intervention for mother-preterm infant dyads. Infant Behav Dev. 2013;36(4):694–706.

175. Ericksen J, Loughlin E, Holt C, et al. A Therapeutic Playgroup for Depressed Mothers and Their Infants: Feasibility Study and Pilot Randomized Trial of Community Hugs. Infant Ment Health J. 2018;39(4):396–409.

176. Holt C, Gentilleau C, Gemmill AW, Milgrom J. Improving the mother-infant relationship following postnatal depression: a randomised controlled trial of a brief intervention (HUGS). Arch Womens Ment Health. 2021;24(6):913–923.

177. Sarkadi A, Kristiansson R, Oberklaid F, Bremberg S. Fathers’ involvement and children’s developmental outcomes: a systematic review of longitudinal studies. Acta Paediatr. 2008;97(2):153–158.

