## Supplemental File for "Dyadic Parent/Caregiver-Infant Interventions Initiated in the First 6 Months of Life to Support Early Relational Health: A Meta-Analysis"

#### Table of Content

eTable 1. PRISMA Checklist
eTable 2. Summary of methodological decisions to avoid unit-of-analysis issues
eTable 3. Full Study-Level Characteristics
eTable 4. Summary of Sensitivity Analysis
eTable 5. GRADE Summary Table
eMethods 1. Full Systematic Review Method
eMethods 2. Full Search Strategies
eMethods 3. Parent/Caregiver-Report Assessments of Relational Health
eMethods 4. Observational Assessments of Relational Health
eMethods 5. Description of Included Interventions
eFigure 1. Overview of Moderator Analyses
eFigure 2. Bonding 0-1-month Follow-Up Forrest Plot
eFigure 3. Bonding 2-6-month Follow-Up Forrest Plot
eFigure 4. Sensitivity 0-1-month Follow-Up Forrest Plot
eFigure 5. Sensitivity 2-6-month Follow-Up Forrest Plot
eFigure 6. Sensitivity 7-12-month Follow-Up Forrest Plot
eFigure 7. Sensitivity 15+ months Follow-Up Forrest Plot
eFigure 8. Secure Attachment 0-1-month Follow-Up Forrest Plot
eFigure 9. Secute Attachment 2-6-month Follow-Up Forrest Plot
eFigure 10. Secure Attachment 13-24-month Follow-Up Forrest Plot
eFigure 11. Organized Attachment 2-6-month Follow-Up Forrest Plot
eFigure 12. Insecure Attachment 0-1-month Follow-Up Forrest Plot
eFigure 13. Insecure Attachment 2-6-month Follow-Up Forrest Plot
eFigure 14. Bonding 13-24-month Follow-Up Forrest Plot
eFigure 15. Disorganized Attachment 0-1-month Follow-Up Forrest Plot
eFigure 16. Disorganized Attachment 2-6-month Follow-Up Forrest Plot
eFigure 17. Dyadic Interactions 0-1-month Follow-Up Forrest Plot
eFigure 18. Dyadic Interactions 2-6-month Follow-Up Forrest Plot
eFigure 19. Dyadic Interactions 7-12-month Follow-Up Forrest Plot

eFigure 20. Cognitive Development 0-1-month Follow-Up Forrest Plot
eFigure 21. Cognitive Development 2-6-month Follow-Up Forrest Plot
eFigure 22. Cognitive Development 7-12-month Follow-Up Forrest Plot
eFigure 23. Cognitive Development 13-24-month Follow-Up Forrest Plot
eFigure 24. Cognitive Development 25+-month Follow-Up Forrest Plot
eFigure 25. Language Development 0-1-month Follow-Up Forrest Plot
eFigure 26. Language Development 2-6-month Follow-Up Forrest Plot
eFigure 27. Language Development 7-12-month Follow-Up Forrest Plot
eFigure 28. Language Development 13-24-month Follow-Up Forrest Plot
eFigure 29. Montor Development 0-1-month Follow-Up Forrest Plot
eFigure 30. Motor Development 2-6-month Follow-Up Forrest Plot
eFigure 31. Motor Development 7-12-month Follow-Up Forrest Plot
eFigure 32. Motor Development 13-24-month Follow-Up Forrest Plot
eFigure 33. Socio-Emotional Functioning 2-6-month Follow-Up Forrest Plot
eFigure 34. Child Behavior 0-1-month Follow-Up Forrest Plot
eFigure 35. Child Behavior 2-6-month Follow-Up Forrest Plot
eFigure 36. Child Behavior 13-24-month Follow-Up Forrest Plot
eFigure 37. Child Behavior 25+-month Follow-Up Forrest Plot
eFigure 38. Parenting Stress 0-1-month Follow-Up Forrest Plot
eFigure 39. Parenting Stress 2-6-month Follow-Up Forrest Plot
eFigure 40. Parent/Caregiver Anxiety 0-1-month Follow-Up Forrest Plot
eFigure 41. Parent/Caregiver Anxiety 2-6-month Follow-Up Forrest Plot
eFigure 42. Parent/Caregiver Depression 0-1-month Follow-Up Forrest Plot
eFigure 43. Parent/Caregiver Depression 2-6-month Follow-Up Forrest Plot
eFigure 44. Parent/Caregiver Depression 7-12-month Follow-Up Forrest Plot
eFigure 45. Parent/Caregiver Depression 13-24-month Follow-Up Forrest Plot
eFigure 46. Overview of Meta-Analysis Results of Subset of 20 Studies
eFigure 47. RoB (RCT) – Bonding (0-1 month F/U)
eFigure 48. RoB (RCT) – Bonding (2-6 months F/U)
eFigure 49. RoB (RCT) – Sensitivity (0-1 months F/U)
eFigure 50. RoB (RCT) – Sensitivity (2-6 months F/U)
eFigure 51. RoB (cluster RCT) – Sensitivity (2-6 months F/U)
eFigure 52. RoB (RCT) – Sensitivity (7-12 months F/U)
eFigure 53. RoB (RCT) – Sensitivity (13-24 months F/U)
eFigure 54. RoB (RCT) – Secure Attachment (0-1-month F/U)
eFigure 55. RoB (cluster RCT) – Secure Attachment (0-1-month F/U)
eFigure 56. RoB (RCT) – Secure Attachment (2-6 months F/U)
eFigure 57. RoB (RCT) – Secure Attachment (13-24-month F/U)
eFigure 58. RoB (RCT) – Organized Attachment (0-1-month F/U)
eFigure 59. RoB (RCT) – Insecure Attachment (0-1-month F/U)
eFigure 60. RoB (RCT) – Insecure Attachment (2-6 months F/U)
eFigure 61. RoB (RCT) – Insecure Attachment (13-24 months F/U)
eFigure 62. RoB (RCT) – Disorganized Attachment (0-1-month F/U)
eFigure 63. RoB (cluster RCT) – Disorganized Attachment (0-1 month F/U)
eFigure 64. RoB (RCT) – Disorganized Attachment (2-6 months F/U)
eFigure 65. RoB (RCT) – Dyadic Interactions (0-1 month F/U)
eFigure 66. RoB (RCT) – Dyadic Interactions (2-6 months F/U)
eFigure 67. RoB (RCT) – Dyadic Interactions (7-12 months F/U)
eFigure 68. RoB (RCT) – Cognitive Development (0-1 month F/U)
eFigure 69. RoB (RCT) – Cognitive Development (2-6 months F/U)
eFigure 70. RoB (RCT) – Cognitive Development (7-12 months F/U)
eFigure 71. RoB (cluster RCT) – Cognitive Development (7-12 months F/U)
eFigure 72. RoB (RCT) – Cognitive Development (13-24 months F/U)
eFigure 73. RoB (RCT) – Cognitive Development (25+ months F/U)
eFigure 74. RoB (cluster RCT) – Cognitive Development (25+ months F/U)
eFigure 75. RoB (RCT) – Language Development (0-1 month F/U)

eFigure 76. RoB (RCT) – Language Development (2-6 months F/U)
eFigure 77. RoB (RCT) – Language Development (7-12 months F/U)
eFigure 78. RoB (RCT) – Language Development (13-24 months F/U)
eFigure 79. RoB (RCT) – Motor Development (0-1 month F/U)
eFigure 80. RoB (RCT) – Motor Development (2-6 months F/U)
eFigure 81. RoB (RCT) – Motor Development (7-12 months F/U)
eFigure 82. RoB (cluster RCT) – Motor Development (7-12 months F/U)
eFigure 83. RoB (RCT) – Motor Development (13-24 months F/U)
eFigure 84. RoB (cluster RCT) – Motor Development (13-24 months F/U)
eFigure 85. RoB (RCT) – Socio-Emotional Functioning (2-6 months F/U)
eFigure 86. RoB (cluster RCT) – Socio-Emotional Functioning (2-6 months F/U)
eFigure 87. RoB (RCT) – Child Behavior (0-1 month F/U)
eFigure 88. RoB (RCT) – Child Behavior (2-6 months F/U)
eFigure 89. RoB (RCT) – Child Behavior (13-24 months F/U)
eFigure 90. RoB (RCT) – Child Behavior (25+ months F/U)
eFigure 91. RoB (cluster RCT) – Child Behavior (25+ months F/U)
eFigure 92. RoB (RCT) – Parenting Stress (0-1 month F/U)
eFigure 93. RoB (cluster RCT) – Parenting Stress (0-1 month F/U)
eFigure 94. RoB (RCT) – Parenting Stress (2-6 months F/U)
eFigure 95. RoB (cluster RCT) – Parenting Stress (2-6 months F/U)
eFigure 96. RoB (RCT) – Parent/Caregiver Anxiety (0-1 month F/U)
eFigure 97. RoB (cluster RCT) – Parent/Caregiver Anxiety (0-1 month F/U)
eFigure 98. RoB (RCT) – Parent/Caregiver Anxiety (2-6 months F/U)
eFigure 99. RoB (RCT) – Parent/Caregiver Depression (0-1 month F/U)
eFigure 100. RoB (cluster RCT) – Parent/Caregiver Depression (0-1 months F/U)
eFigure 101. RoB (RCT) – Parent/Caregiver Depression (2-6 months F/U)
eFigure 102. RoB (RCT) – Parent/Caregiver Depression (7-12 months F/U)
eFigure 103. RoB (RCT) – Parent/Caregiver Depression (13-24 months F/U)

#### eTable 1. PRISMA Checklist

| Section and Topic | Item # | Checklist item | Location where item is reported |
|---|---|---|---|
| **TITLE** | | | |
| Title | 1 | Identify the report as a systematic review. | 1 |
| **ABSTRACT** | | | |
| Abstract | 2 | See the PRISMA 2020 for Abstracts checklist. | N/A |
| **INTRODUCTION** | | | |
| Rationale | 3 | Describe the rationale for the review in the context of existing knowledge. | 7-9 |
| Objectives | 4 | Provide an explicit statement of the objective(s) or question(s) the review addresses. | 9 |
| **METHODS** | | | |
| Eligibility criteria | 5 | Specify the inclusion and exclusion criteria for the review and how studies were grouped for the syntheses. | 9-10 |
| Information sources | 6 | Specify all databases, registers, websites, organisations, reference lists and other sources searched or consulted to identify studies. Specify the date when each source was last searched or consulted. | 10 |
| Search strategy | 7 | Present the full search strategies for all databases, registers and websites, including any filters and limits used. | eMethods2 |
| Selection process | 8 | Specify the methods used to decide whether a study met the inclusion criteria of the review, including how many reviewers screened each record and each report retrieved, whether they worked independently, and if applicable, details of automation tools used in the process. | 10 +eMethods1 |
| Data collection process | 9 | Specify the methods used to collect data from reports, including how many reviewers collected data from each report, whether they worked independently, any processes for obtaining or confirming data from study investigators, and if applicable, details of automation tools used in the process. | 10-11 +eMethods1 |
| Data items | 10a | List and define all outcomes for which data were sought. Specify whether all results that were compatible with each outcome domain in each study were sought (e.g. for all measures, time points, analyses), and if not, the methods used to decide which results to collect. | 11-12 +eMethods1 |
| | 10b | List and define all other variables for which data were sought (e.g. participant and intervention characteristics, funding sources). Describe any assumptions made about any missing or unclear information. | 11 +eMethods1 |
| Study risk of bias assessment | 11 | Specify the methods used to assess risk of bias in the included studies, including details of the tool(s) used, how many reviewers assessed each study and whether they worked independently, and if applicable, details of automation tools used in the process. | 13 +eMethods1 |
| Effect measures | 12 | Specify for each outcome the effect measure(s) (e.g. risk ratio, mean difference) used in the synthesis or presentation of results. | 11-12 +eMethods1 |
| Synthesis methods | 13a | Describe the processes used to decide which studies were eligible for each synthesis (e.g. tabulating the study intervention characteristics and comparing against the planned groups for each synthesis (item #5)). | 11-12 +eMethods1 |
| | 13b | Describe any methods required to prepare the data for presentation or synthesis, such as handling of missing summary statistics, or data conversions. | 11-12 +eMethods1 |
| | 13c | Describe any methods used to tabulate or visually display results of individual studies and syntheses. | 11-12 +eMethods1 |

| Section and Topic | Item # | Checklist item | Location where item is reported |
|---|---|---|---|
| | 13d | Describe any methods used to synthesize results and provide a rationale for the choice(s). If meta-analysis was performed, describe the model(s), method(s) to identify the presence and extent of statistical heterogeneity, and software package(s) used. | 11-12 +eMethods1 |
| | 13e | Describe any methods used to explore possible causes of heterogeneity among study results (e.g. subgroup analysis, meta-regression). | 11-12 +eMethods1 |
| | 13f | Describe any sensitivity analyses conducted to assess robustness of the synthesized results. | 11-12 +eMethods1 |
| Reporting bias assessment | 14 | Describe any methods used to assess risk of bias due to missing results in a synthesis (arising from reporting biases). | 12 +eMethods1 |
| Certainty assessment | 15 | Describe any methods used to assess certainty (or confidence) in the body of evidence for an outcome. | 13 +eMethods1 |
| **RESULTS** | | | |
| Study selection | 16a | Describe the results of the search and selection process, from the number of records identified in the search to the number of studies included in the review, ideally using a flow diagram. | 13 |
| | 16b | Cite studies that might appear to meet the inclusion criteria, but which were excluded, and explain why they were excluded. | N/A |
| Study characteristics | 17 | Cite each included study and present its characteristics. | Table 1 +eTable3 |
| Risk of bias in studies | 18 | Present assessments of risk of bias for each included study. | eFigure47-103 |
| Results of individual studies | 19 | For all outcomes, present, for each study: (a) summary statistics for each group (where appropriate) and (b) an effect estimate and its precision (e.g. confidence/credible interval), ideally using structured tables or plots. | Figure 2 + eFigures2-45 |
| Results of syntheses | 20a | For each synthesis, briefly summarise the characteristics and risk of bias among contributing studies. | 14-16 |
| | 20b | Present results of all statistical syntheses conducted. If meta-analysis was done, present for each the summary estimate and its precision (e.g. confidence/credible interval) and measures of statistical heterogeneity. If comparing groups, describe the direction of the effect. | 14-16 |
| | 20c | Present results of all investigations of possible causes of heterogeneity among study results. | 15 |
| | 20d | Present results of all sensitivity analyses conducted to assess the robustness of the synthesized results. | eTable4 |
| Reporting biases | 21 | Present assessments of risk of bias due to missing results (arising from reporting biases) for each synthesis assessed. | eTable4 |
| Certainty of evidence | 22 | Present assessments of certainty (or confidence) in the body of evidence for each outcome assessed. | Figure 2 +eTable5 |
| **DISCUSSION** | | | |
| Discussion | 23a | Provide a general interpretation of the results in the context of other evidence. | 17-19 |
| | 23b | Discuss any limitations of the evidence included in the review. | 17-19 |
| | 23c | Discuss any limitations of the review processes used. | 17-19 |

| Section and Topic | Item # | Checklist item | Location where item is reported |
|---|---|---|---|
| | 23d | Discuss implications of the results for practice, policy, and future research. | 17-19 |
| **OTHER INFORMATION** | | | |
| Registration and protocol | 24a | Provide registration information for the review, including register name and registration number, or state that the review was not registered. | 9 |
| | 24b | Indicate where the review protocol can be accessed, or state that a protocol was not prepared. | N/A |
| | 24c | Describe and explain any amendments to information provided at registration or in the protocol. | N/A |
| Support | 25 | Describe sources of financial or non-financial support for the review, and the role of the funders or sponsors in the review. | 19 |
| Competing interests | 26 | Declare any competing interests of review authors. | 19 |
| Availability of data, code and other materials | 27 | Report which of the following are publicly available and where they can be found: template data collection forms; data extracted from included studies; data used for all analyses; analytic code; any other materials used in the review. | N/A |

#### eTable 2. Summary of Methodological Decisions to Avoid Unit-of-Analysis Issues

| Unit-of-analysis issue | Study ID | Methodological decision |
| --- | --- | --- |
| RCTs with 3 arms | Borghini 2014 | Term control group dropped. |
| | Browne 2005 | Education and control group combined to form the comparator. |
| | Cates 2018 | Data for maternal sensitivity was only presented for the VIP and Control group. Building Blocks group was dropped. |
| | Juffer 2005 | Book only group and control group were combined to form the comparator. |
| | Klein Velderman 2006 | VIPP and VIPP-R groups were combined to form the exposed group. |
| | Neu 2010 | Blanket holding group and control group were combined to form the comparator. |
| | Porter 2015 | PEP group dropped because not dyadic and we decided to not pool another active control into the comparators. We used IMPEP and the exposed group and the standard care control group. |
| Cluster RCTs | Barnes 2022 | Sample size adjusted using the ICC reported in Kristensen 2020 and mean cluster size of 20. |
| | Betancourt 2020 | Sample size adjusted using ICC of 0.03, and a mean cluster size of 5. |
| | Feldman 2014 | Sample size not adjusted: mean cluster size not reported. |
| | Glazebrook 2007 | Sample size not adjusted: mean cluster size not reported. |
| | Herbers 2020 | Sample size adjusted using ICC reported in Betancourt 2020 and mean size 6.5. |
| | Knoche 2012 | Sample size not adjusted: mean cluster size not reported. |
| | Kristensen 2020 | Sample size adjusted using ICC of 0.001 and mean cluster size of 66. |
| | Sadler 2013 | Sample size not adjusted: mean cluster size not reported. |
| | Sleed 2013 | Sample size not adjusted: mean cluster size not reported. |

#### eTable 3. Full Study-Level Characteristics

| Study ID | Country | N | Sample characteristics | | | Design | Intervention | Control |
|---|---|---|---|---|---|---|---|---|
| | | | Mothers (%) | Female infants (%) | Population | | | |
| Akai 2008 | USA | 63 | 100.00 | N/R | Low SES | Parallel-group RCT | My Baby and Me | Standard care/Attention control |
| Alarcão 2021 | Brazil | 80 | 100.00 | 46.45 | Low SES, Adolescent mothers, First time mother | Parallel-group RCT | *Primeiros Laços* | Standard care/Attention control |
| Alvarenga 2020 | Brazil | 56 | 100.00 | 38.65 | Low SES | Parallel-group RCT | Maternal Sensitivity Program (video feedback) | Standard care/Attention control |
| Badr 2006 | USA | 62 | 100.00 | 31.25 | Infant with early health or developmental conditions | Parallel-group RCT | Curriculum and Monitoring System (CAMS) | Standard care/Attention control |
| Baggett 2010 | USA | 40 | 100.00 | 45.00 | Low SES | Parallel-group RCT | Internet adaptation of PALS program (Infant-Net) | Standard care/Attention control |
| Baggett 2017 | USA | 159 | 100.00 | N/R | Low SES, Elevated risk for maltreatment | Parallel-group RCT | Internet adaptation of PALS Program (ePALS Baby-Net) | Standard care/Attention control |
| Barlow 2016 | UK | 31 | 100.00 | 38.70 | Preterm/Low birthweight infants | Parallel-group RCT | Video interaction guidance (VIG) | Standard care/Attention control |
| Barnes 2022 | UK | 40 | 100.00 | 52.50 | Preterm/Low birthweight infants | Cluster RCT | Touching and caressing; tender in caring (TAC-TIC) | Standard care/Attention control |
| Berlin 2017 | USA | 336 | 100.00 | 50.00 | Maternal psychopathology, Adolescent mothers, First time mothers | Parallel-group RCT | Healthy Families Durham (HFD) | Standard care/Attention control |
| Berlin 2018 | USA | 208 | 100.00 | 51.00 | Low SES | Parallel-group RCT | Early Head Start Plus Attachment and Biobehavioral Catch-up (ABC) | Standard care/Attention control |

| Study ID | Country | N | Sample characteristics | | | Design | Intervention | Control |
|---|---|---|---|---|---|---|---|---|
| | | | Mothers (%) | Female infants (%) | Population | | | |
| Bernard 2012 | USA | 120 | 100.00 | 42.00 | Elevated risk for maltreatment | Parallel-group RCT | Attachment and Biobehavioral Catch-up (ABC) | Developmental Education for Families (DEF) Intervention |
| Betancourt 2020 | Rwanda | 198 | 68.23 | N/R | Low SES | Parallel-group RCT | *Sugira Muryango* | Standard care/Attention control |
| Bick 2013 | USA | 96 | 0.00 | 48.00 | Foster/Adoptive parents | Parallel-group RCT | Attachment and Biobehavioral Catch-up Intervention (ABC) | Developmental Education for Families (DEF) Intervention |
| Borghini 2014 | Switzerland | 60 | 100.00 | 49.00 | Preterm/Low birthweight infants | Parallel-group RCT | Early Intervention Program | Standard care/Attention control |
| Brisch 2003 | Germany | 87 | 100.00 | 45.77 | Preterm/Low birthweight infants | Parallel-group RCT | Parent-centered Intervention Program | Standard care/Attention control |
| Browne 2005 | USA | 99 | 100.00 | N/R | Preterm/Low birthweight infants | Parallel-group RCT | Demonstration and Interaction | Standard care/Attention control |
| Cates 2018 | USA | 675 | 100.00 | 27.30 | Low SES | Parallel-group RCT | Video Intervention Project (VIP) | Standard care/Attention control |
| Cevasco 2008 | USA | 34 | 100.00 | N/R | Preterm/Low birthweight infants | Parallel-group RCT | Maternal Singing Intervention | Standard care/Attention control |
| Chiu 2009 | USA | 100 | 100.00 | 52.00 | Preterm/Low birthweight infants | Parallel-group RCT | Skin-to-skin contact (SSC) | Standard care/Attention control |
| Constantino 2001 | USA | 148 | 100.00 | 39.00 | Mothers and infants (no risk factor) | Parallel-group RCT | Home Visitation and Group Intervention | Standard care/Attention control |
| Cooper 2009 | South Africa | 449 | 100.00 | 48.10 | Low SES | Parallel-group RCT | Mother-infant Relationship Intervention | Standard care/Attention control |

| Study ID | Country | N | Sample characteristics | | | Design | Intervention | Control |
|---|---|---|---|---|---|---|---|---|
| | | | Mothers (%) | Female infants (%) | Population | | | |
| Cooper 2015 | UK | 301 | 100.00 | 48.45 | Maternal psychopathology | Parallel-group RCT | Index (R-HV) | Standard care/Attention control |
| Corrigan 2021 | USA | 97 | 100.00 | 76.50 | Preterm/Low birthweight infants | Parallel-group RCT | Music Therapy | Standard care/Attention control |
| Dozier 2009 | USA | 46 | 0.00 | 50.00 | Foster/Adoptive parents | Parallel-group RCT | Attachment and Bio behavioral Catch-up Intervention (ABC) | Developmental Education for Families (DEF) Intervention |
| Ericksen 2018 | Australia | 31 | 100.00 | N/R | Maternal psychopathology | Parallel-group RCT | Community HUGS (CHUGS) | Standard care/Attention control |
| Feil 2020 | USA | 164 | 100.00 | 55.80 | Low SES | Parallel-group RCT | ePALS | Standard care/Attention control |
| Feldman 2014 | Israel | 146 | 100.00 | N/R | Preterm/Low birthweight infants | Parallel-group RCT | Contact Intervention (kangaroo care) | Standard care/Attention control |
| Firk 2021 | Germany | 56 | 100.00 | 48.00 | Adolescent mothers | Parallel-group RCT | Adaptation of Step Towards Effective and Enjoyable Parenting (STEEP-b) | Standard care/Attention control |
| Fonagy 2016 | UK | 76 | 100.00 | 36.50 | Low SES, Maternal psychopathology | Parallel-group RCT | Parent–Infant Psychotherapy (PIP) | Standard care/Attention control |
| Gaden 2022 | Argentina, Colombia, Israel, Norway, and Poland | 213 | 100.00 | 51.00 | Preterm/Low birthweight infants | Parallel-group RCT | Music Therapy | Standard care/Attention control |
| Georg 2021 | Germany | 154 | 100.00 | 43.50 | Infant with early health or | Parallel-group RCT | Focused Parent-Infant | Standard care/Attention control |

| Study ID | Country | N | Sample characteristics | | | Design | Intervention | Control |
|---|---|---|---|---|---|---|---|---|
| | | | Mothers (%) | Female infants (%) | Population | | | |
| | | | | | developmental conditions | | Psychotherapy (fPIP) | |
| Glazebrook 2007 | UK | 233 | 100.00 | 52.50 | Preterm/Low birthweight infants | Cluster RCT | Parent Baby Interaction Programme (PBIP) | Standard care/Attention control |
| Hane 2015 | USA | 150 | 100.00 | 47.85 | Preterm/Low birthweight infants | Parallel-group RCT | Family Nurture Intervention (FNI) | Standard care/Attention control |
| Heo 2019 | South Korea | 66 | 97.00 | 40.30 | Preterm/Low birthweight infants | Parallel-group RCT | Parent Participation Improvement Program | Standard care/Attention control |
| Herbers 2020 | USA | 45 | 100.00 | 38.00 | Low SES | Cluster RCT | My baby's first teacher (MBFT) | Standard care/Attention control |
| Hoffenkamp 2015 | The Netherlands | 150 | 100.00 | 44.00 | Preterm/Low birthweight infants | Parallel-group RCT | Video Interaction Guidance (VIG) | Standard care/Attention control |
| Høivik 2015 | Norway | 158 | 98.70 | 51.00 | Infant with early health or developmental conditions | Parallel-group RCT | Video Feedback of Infant-Parent Interaction (VIPI) | Standard care/Attention control |
| Holditch-Davis 2014 | USA | 240 | 100.00 | 53.60 | Preterm/Low birthweight infants | Parallel-group RCT | Auditory-Tactile-Visual-Vestibular (ATVV) | Standard care/Attention control |
| Holt 2021 | Australia | 77 | 100.00 | N/R | Maternal psychopathology | Parallel-group RCT | Happiness, Understanding, Giving and Sharing Intervention (HUGS) | Standard care/Attention control |
| Horowitz 2001 | USA | 122 | 100.00 | N/R | Maternal psychopathology | Parallel-group RCT | Interaction Coaching for At-Risk Parents (ICAP) | Standard care/Attention control |

| Study ID | Country | N | Sample characteristics | | | Design | Intervention | Control |
|---|---|---|---|---|---|---|---|---|
| | | | Mothers (%) | Female infants (%) | Population | | | |
| Horowitz 2013 | USA | 144 | 100.00 | N/R | Maternal psychopathology | Parallel-group RCT | Communicating and Relating Effectively (CARE) | Standard care/Attention control |
| Juffer 2005 | The Netherlands | 130 | 0.00 | 49.23 | Foster/Adoptive parents | Parallel-group RCT | Book with Video Feedback via Home Visits | Standard care/Attention control |
| Klein Velderman 2006 | The Netherlands | 84 | 100.00 | 50.62 | First time mothers | Parallel-group RCT | Video-feedback Intervention to Promote Positive Parenting (VIPP) | Standard care/Attention control |
| Knoche 2012 | USA | 234 | 94.00 | 48.30 | Low SES | Cluster RCT | The Getting Ready Intervention | Standard care/Attention control |
| Kristensen 2020 | Denmark | 2366 | 100.00 | N/R | Mothers and infants (no risk factor) | Cluster RCT | Newborn Behavioral Observations (NBO) | Standard care/Attention control |
| Landry 2006 | USA | 264 | 100.00 | 55.00 | Low SES, Preterm/Low birthweight infants | Parallel-group RCT | Playing and Learning Strategies (PALS) | Standard care/Attention control |
| Lavallée 2022 | Canada | 20 | 100.00 | 65.00 | Preterm/Low birthweight infants | Parallel-group RCT | Guided Participation to Infant Therapeutic Positioning (GP_Posit) | Standard care/Attention control |
| Magill-Evans 2007 | Canada | 183 | 0.00 | 47.00 | Fathers and infants | Parallel-group RCT | Education Program | Standard care/Attention control |
| Meijssen 2010 | The Netherlands | 176 | 100.00 | 48.33 | Preterm/Low birthweight infants | Parallel-group RCT | The Infant Behavioral Assessment and Intervention Program (IBAIP) | Standard care/Attention control |

| Study ID | Country | N | Sample characteristics | | | Design | Intervention | Control |
|---|---|---|---|---|---|---|---|---|
| | | | Mothers (%) | Female infants (%) | Population | | | |
| Melnyk 2006 | USA | 260 | 100.00 | 51.50 | Preterm/Low birthweight infants | Parallel-group RCT | Creating Opportunities for Parent Empowerment (COPE) | Standard care/Attention control |
| Mendelsohn 2007 | USA | 150 | 100.00 | 38.30 | Low SES | Parallel-group RCT | Video Intervention Project (VIP) | Standard care/Attention control |
| Milgrom 2013 | Australia | 123 | 100.00 | 51.20 | Preterm/Low birthweight infants | Parallel-group RCT | PremieStart Program | Standard care/Attention control |
| Nelson 2001 | USA | 37 | 100.00 | 50.50 | Infant with early health or developmental impairments, Preterm/low birthweight infants | Parallel-group RCT | Auditory-Tactile-Visual-Vestibular Intervention (ATVV) | Standard care/Attention control |
| Neu 2010 | USA | 87 | 100.00 | 51.50 | Preterm/Low birthweight infants | Parallel-group RCT | Kangaroo care | Standard care/Attention control |
| Newnham 2009 | Australia | 63 | 100.00 | 52.00 | Preterm/Low birthweight infants | Parallel-group RCT | Mother-Infant Transaction Program (MITP) | Standard care/Attention control |
| Nugent 2017 | USA | 40 | 100.00 | 55.00 | Mothers and infants (no risk factor) | Parallel-group RCT | Newborn Behavioral Observations (NBO) System | Standard care/Attention control |
| Onozawa 2001 | UK | 59 | 100.00 | 32.50 | Maternal psychopathology, First time mother | Parallel-group RCT | Massage class + Support group | Standard care/Attention control |
| Oxford 2021 | USA | 252 | 100.00 | 47.65 | Maternal psychopathology | Parallel-group RCT | Promoting First Relationships (PFR) Intervention | Standard care/Attention control |

| Study ID | Country | N | Sample characteristics | | | Design | Intervention | Control |
|---|---|---|---|---|---|---|---|---|
| | | | Mothers (%) | Female infants (%) | Population | | | |
| Perrone 2021 | USA | 200 | 96.00 | 51.00 | Mothers and infants (no risk factor) | Parallel-group RCT | Attachment and Biobehavioral Catch-up (ABC) | Standard care/Attention control |
| Pontoppidan 2016 | Denmark and Norway | 112 | 72.00 | 51.00 | First time mothers | Pragmatic RCT | Incredible Years Parents and Babies (IYPB) Intervention | Standard care/Attention control |
| Porter 2015 | USA | 138 | 100.00 | 53.40 | Maternal psychopathology | Pragmatic RCT | Infant Massage-Parenting Enhancement Program (IMPEP) | Standard care/Attention control |
| Ramsauer 2020 | Germany | 72 | 100.00 | 44.45 | Maternal psychopathology | Parallel-group RCT | Circle of Security Intervention (COS-I) | Standard care/Attention control |
| Ravn 2011 | Norway | 118 | 100.00 | 47.35 | Preterm/Low birthweight infants | Parallel-group RCT | Mother Infant Transaction Program (MITP) | Standard care/Attention control |
| Rheinheimer 2022 | The Netherlands | 127 | 100.00 | 51.13 | Mothers and infants (no risk factor) | Parallel-group RCT | Skin-to-skin contact (SSC) | Standard care/Attention control |
| Robertson 2019 | USA | 66 | 100.00 | 42.22 | Mothers and infants (no risk factor) | Parallel-group RCT | Contingent Lullaby | Standard care/Attention control |
| Roby 2021 | USA | 403 | N/R | 49.00 | Low SES | Parallel-group RCT | Smart Beginnings: Video Interaction Project (VIP) + Family Check-Up | Standard care/Attention control |
| Sadler 2013 | USA | 139 | 100.00 | 48.00 | First time mothers | Cluster RCT | Minding the Baby (MTB) | Standard care/Attention control |
| Sahlén Helmer 2020 | Sweden | 42 | 100.00 | 45.16 | Preterm/Low birthweight infants | Parallel-group RCT | Continuous Skin to Skin Contact (SSC) | Standard care/Attention control |
| Sajaniemi 2001 | Finland | 115 | N/R | 49.65 | Preterm/Low birthweight infants | Parallel-group RCT | Occupational Therapy Intervention | Standard care/Attention control |

| Study ID | Country | N | Sample characteristics | | | Design | Intervention | Control |
|---|---|---|---|---|---|---|---|---|
| | | | Mothers (%) | Female infants (%) | Population | | | |
| Salo 2019 | Finland | 45 | 100.00 | N/R | Maternal psychopathology | Parallel-group RCT | Nurture and Play (NaP) | Standard care/Attention control |
| Santelices 2011 | Chile | 100 | 100.00 | N/R | First time mothers | Parallel-group RCT | Secure Attachment Promotion Program | Standard care/Attention control |
| Schuler 2000 | USA | 192 | 100.00 | 53.55 | Maternal psychopathology | Parallel-group RCT | Home Intervention: Based on the program used by the infant health and development program (IHDP) | Standard care/Attention control |
| Shoghi 2018 | Iran | 40 | 100.00 | 60.00 | First time mothers, Preterm/Low birthweight infants | Parallel-group RCT | Massage Intervention | Standard care/Attention control |
| Sleed 2013 | UK | 163 | 100.00 | 60.68 | Mother-infant dyads incarcerated | Cluster RCT | New Beginnings Intervention | Standard care/Attention control |
| Steele 2019 | USA | 228 | 100.00 | N/R | Elevated risk for maltreatment | Parallel-group RCT | Group Attachment-Based Intervention (GABI) | Steps Toward Effective, Enjoyable Parenting (STEP) Intervenion |
| Stein 2006 | UK | 80 | 100.00 | 53.75 | Maternal psychopathology | Parallel-group RCT | Video-Feedback Interactional Treatment | Standard care/Attention control |
| Stein 2018 | UK | 144 | 100.00 | 51.39 | Maternal psychopathology | Parallel-group RCT | Video-Feedback Therapy (VFT) + Cognitive Behavioral Therapy (CBT) | Standard care/Attention control |
| Taneja 2020 | India | 552 | 100.00 | 58.50 | Preterm/Low birthweight infants | Parallel-group RCT | Community-initiated Kangaroo Mother Care (ciKMC) | Standard care/Attention control |

| Study ID | Country | N | Sample characteristics | | | Design | Intervention | Control |
|---|---|---|---|---|---|---|---|---|
| | | | Mothers (%) | Female infants (%) | Population | | | |
| Tereno 2017 | France | 120 | 100.00 | 52.50 | Low SES, First time mothers | Parallel-group RCT | *"Compétences Parentales et Attachement dans la Petite Enfance: Diminution des risques liés aux troubles de santé mentale et Promotion de la resilience*" project (CAPEDP) | Standard care/Attention control |
| Teti 2009 | USA | 173 | 100.00 | 53.00 | Preterm/Low birthweight infants | Parallel-group RCT | Infant Intervention | Standard care/Attention control |
| Tryphonopoulos 2020 | Canada | 12 | 100.00 | 58.35 | Maternal psychopathology | Parallel-group RCT | Video Feedback | Standard care/Attention control |
| Tsivos 2015 | UK | 27 | 100.00 | 55.25 | Maternal psychopathology | Parallel-group RCT | Baby Triple P | Standard care/Attention control |
| Twohig 2021 | Republic of Ireland | 98 | 100.00 | 54.05 | Preterm/Low birthweight infants | Pragmatic RCT | Preterm Infant–Parent Program for Attachment (PIPPA) | Standard care/Attention control |
| Valades 2021 | El Slavador | 64 | 100.00 | N/R | Adolescent mothers | Parallel-group RCT | *Crianza Temprana* (Early Parenting) | Standard care/Attention control |
| van Doesum 2008 | The Netherlands | 85 | 100.00 | 40.50 | Maternal psychopathology | Parallel-group RCT | Mother-Baby Intervention | Standard care/Attention control |
| Vargas-Porras 2021 | Columbia | 76 | 100.00 | 53.00 | First time mothers | Parallel-group RCT | Maternal Support for Becoming a First-time Mother (AMACOMPRI) | Standard care/Attention control |

| Study ID | Country | N | Sample characteristics | | | Design | Intervention | Control |
|---|---|---|---|---|---|---|---|---|
| | | | Mothers (%) | Female infants (%) | Population | | | |
| Walter 2019 | US and Germany | 169 | 52.16* | 49.70 | Parents and infants (no risk factor) | Parallel-group RCT | Secure Attachment Family Education (SAFE) | Standard care/Attention control |
| White-Traut 2013 | USA | 198 | 100.00 | 47.00 | Preterm/Low birthweight infants | Parallel-group RCT | H-Hope Intervention | Standard care/Attention control |
| Williams 2020 | USA | 74 | 100.00 | 42.35 | Low SES, Adolescent mothers | Parallel-group RCT | Infant Carrier | Standard care/Attention control |
| Wulff 2021 | Germany | 120 | 100.00 | N/R | Maternal psychopathology | Parallel-group RCT | Singing Intervention | Standard care/Attention control |
| Youn 2021 | South Korea | 151 | 100.00 | 50.80 | Preterm/Low birthweight infants | Parallel-group RCT | Mother–Infant Transaction Program (MITP) home visits | Standard care/Attention control |
| Yu 2022 | Taiwan | 64 | 100.00 | 51.56 | Preterm/Low birthweight infants | Parallel-group RCT | Maternal Voice Recording | Standard care/Attention control |
| Zelkowitz 2011 | Canada | 122 | 100.00 | 50.00 | Preterm/Low birthweight infants | Parallel-group RCT | Cues | Standard care/Attention control |

**eTable 4. Summary of Sensitivity Analysis**

| | | Removed: High Risk of Bias | | Removed: High Risk and Some Concern Risk of Bias | | Removed: Cluster RCT | | Removed: Active Control Group | |
|---|---|---|---|---|---|---|---|---|---|
| **Outcome** | **Statistic** | **Number of studies (participants)** | **Effect estimate** | **Number of studies (participants)** | **Effect estimate** | **Number of studies (participants)** | **Effect estimate** | **Number of studies (participants)** | **Effect estimate** |
| **Outcome Domain: Relational Health** | | | | | | | | | |
| Bonding (0-1m F/U) | SMD (95%CI) | k=7 (n=1 588) | 0.64 (.19, 1.10) | k=4 (n=843) | 0.41 (-.01, .83) | N/A | N/A | N/A | N/A |
| Bonding (2-6m F/U) | SMD (95%CI) | N/A | N/A | N/A | N/A | N/A | N/A | N/A | N/A |
| Sensitivity (0-1m F/U) | SMD (95%CI) | k=32 (n=4 334) | .33 (.21, .45) | k=15 (n=2 448) | 0.24 (.07, .40) | k=34 (n=4 800) | .39 (.27, .50) | N/A | N/A |
| Sensitivity (2-6m F/U) | SMD (95%CI) | k=11 (n=971) | .12 (-.10, .34) | k=5 (n=578) | .00 (-.26, .26) | k=11 (n=791) | .14 (-.08, .37) | N/A | N/A |
| Sensitivity (7-12m F/U) | SMD (95%CI) | k=4 (n=454) | .35 (0.08, 0.62) | k=2 (n=313) | .26 (.04, .49) | N/A | N/A | N/A | N/A |
| Sensitivity (13-24m F/U) | SMD (95%CI) | N/A | N/A | N/A | N/A | N/A | N/A | N/A | N/A |
| Secure Attachment (0-1m F/U) | OR (95%CI) | k=6 (n=436) | 1.65 (1.11, 2.46) | k=3 (11) | 1.47 (0.59, 3.67) | k=7 (n=395) | 1.69 (1.17, 2.44) | k=7 (n=469) | 1.59 (1.08, 2.34) |
| Secure Attachment (2-6m F/U) | OR (95% CI) | k=2 (n=110) | 1.21 (.50, 2.91) | N/A | N/A | N/A | N/A | N/A | N/A |
| Secure Attachment (14-24m F/U) | OR (95% CI) | k=3 (n=497) | .00 (.46, 2.12) | N/A | N/A | N/A | N/A | N/A | N/A |
| Organized Attachment (0-1m F/U) | OR (95%CI) | k=3 (n=331) | 2.23 (1.21, 4.11) | N/A | N/A | N/A | N/A | k=2 (n=211) | 1.84 (.65, 5.21) |
| Insecure Attachment (0-1m F/U) | OR (95%CI) | k=5 (n=365) | .64 (.35, 1.19) | k=3 (n=151) | .76 (.21, 2.75) | N/A | N/A | k=5 (n=317) | .66 (.35, 1.26) |
| Insecure Attachment (2-6m F/U) | OR (95%CI) | k=2 (n=110) | .66 (.28, 1.57) | N/A | N/A | N/A | N/A | N/A | N/A |
| Insecure Attachment | OR (95%CI) | N/A | N/A | N/A | N/A | N/A | N/A | N/A | N/A |

| | | Removed: High Risk of Bias | | Removed: High Risk and Some Concern Risk of Bias | | Removed: Cluster RCT | | Removed: Active Control Group | |
|---|---|---|---|---|---|---|---|---|---|
| **Outcome** | **Statistic** | **Number of studies (participants)** | **Effect estimate** | **Number of studies (participants)** | **Effect estimate** | **Number of studies (participants)** | **Effect estimate** | **Number of studies (participants)** | **Effect estimate** |
| (12-24m F/U) | | | | | | | | | |
| Disorganized Attachment (0-1m F/U) | OR (95%CI) | N/A | N/A | k=3 (n=214) | .55 (.26, 1.16) | k=5 (n=428) | .51 (.32, .82) | k=5 (n=379) | .60 (.36, 1.01) |
| Disorganized Attachment (2-6m F/U) | OR (95%CI) | N/A | N/A | N/A | N/A | N/A | N/A | N/A | N/A |
| Dyadic (0-1m F/U) | SMD (95%CI) | k=20 (n=2 478) | .18 (.06, .30) | k=10 (n=1 603) | .10 (-.01, .20) | k=24 (n=2 750) | .24 (.13, .35) | k=24 (n=2 760) | .20 (.08, .31) |
| Dyadic (2-6 m F/U) | SMD (95%CI) | k=9 (n=795) | .09 (-.05, .23) | k=5 (n=550) | .01 (-.16, .18) | N/A | N/A | N/A | N/A |
| Dyadic (7-12m F/U) | SMD (95%CI) | k=3 (n=393) | .22 (.02, .42) | N/A | N/A | N/A | N/A | N/A | N/A |
| Behavior (0-1m F/U) | SMD (95%CI) | N/A | N/A | N/A | N/A | N/A | N/A | N/A | N/A |
| Behavior (2-6m F/U) | SMD (95%CI) | N/A | N/A | N/A | N/A | N/A | N/A | N/A | N/A |
| Behavior (13-24m F/U) | SMD (95%CI) | k=5 (n=533) | .43 (-.01, .87) | N/A | N/A | N/A | N/A | k=5 (n=478) | .40 (-.08, .89) |
| Behavior (25+m F/U) | SMD (95%CI) | N/A | N/A | N/A | N/A | k=2 (n=140) | -.34 (-.87, .18) | N/A | N/A |
| Socio-Emotional (2-6m F/U) | SMD (95%CI) | N/A | N/A | N/A | N/A | N/A | N/A | N/A | N/A |
| Cognitive (0-1m F/U) | SMD (95%CI) | k=2 (n=100) | -.38 (-.78, .02) | N/A | N/A | N/A | N/A | N/A | N/A |
| Cognitive (2-6m F/U) | SMD (95%CI) | k=5 (n=954) | .09 (-.04, .21) | k=3 (n=693) | .07 (-.08, .22) | N/A | N/A | N/A | N/A |
| Cognitive (7-12m F/U) | SMD (95%CI) | N/A | N/A | N/A | N/A | N/A | N/A | N/A | N/A |
| Cognitive (13-24m F/U) | SMD (95%CI) | k=4 (n=394) | .08 (-.12, .28) | k=3 (n=312) | .07 (-.15, .30) | k=5 (n=421) | .10 (-.09, .29) | k=5 (n=438) | .32 (.06, .57) |

| | | Removed: High Risk of Bias | | Removed: High Risk and Some Concern Risk of Bias | | Removed: Cluster RCT | | Removed: Active Control Group | |
|---|---|---|---|---|---|---|---|---|---|
| **Outcome** | **Statistic** | **Number of studies (participants)** | **Effect estimate** | **Number of studies (participants)** | **Effect estimate** | **Number of studies (participants)** | **Effect estimate** | **Number of studies (participants)** | **Effect estimate** |
| Cognitive (25+m F/U) | SMD (95%CI) | N/A | N/A | N/A | N/A | N/A | N/A | N/A | N/A |
| Language (5-12m) | SMD (95%CI) | N/A | N/A | N/A | N/A | N/A | N/A | N/A | N/A |
| Language (0-1m F/U) | SMD (95%CI) | N/A | N/A | N/A | N/A | N/A | N/A | N/A | N/A |
| Language (2-6m F/U) | SMD (95%CI) | N/A | N/A | k=4 (n=784) | .04 (-.17, .24) | N/A | N/A | N/A | N/A |
| Language (7-12m F/U) | SMD (95%CI) | N/A | N/A | N/A | N/A | N/A | N/A | N/A | N/A |
| Language (13-24m F/U) | SMD (95%CI) | N/A | N/A | k=2 (n=177) | -0.03 (-.47, .40) | N/A | N/A | N/A | N/A |
| Motor (0-1m F/U) | SMD (95%CI) | k=2 (n=100) | -.04 (-.43, .36) | N/A | N/A | N/A | N/A | N/A | N/A |
| Motor (2-6m F/U) | SMD (95%CI) | k=4 (n=820) | .02 (-.12, .16) | k=3 (n=693( | .02 (-.12, .17) | N/A | N/A | N/A | N/A |
| Motor (7-12m F/U) | SMD (95%CI) | N/A | N/A | N/A | N/A | k=2 (n=543) | -.04 (-.21, .12) | N/A | N/A |
| Motor (13-24m F/U) | SMD (95%CI) | k=3 (n=262) | -.03 (-.27, .22) | k=2 (n=180) | 0.00 (-.34, .33) | k=3 (n=262) | -.03 (-.27, .22) | N/A | N/A |
| Parenting Stress (0-1m F/U) | SMD (95%CI) | k=11 (n=681) | .08 (-.12, .28) | k=4 (n=267) | .08 (-.31, .19) | N/A | N/A | N/A | N/A |
| Parenting Stress (2-6m F/U) | SMD (95%CI) | k=3 (n=315) | -.19 (-.42, .04) | N/A | N/A | k=4 (n=270) | 0.00 (-.40, .40) | N/A | N/A |
| Anxiety (0-1m F/U) | SMD (95%CI) | k=9 (n=909) | .12 (-.03, .26) | k=2 (n=154) | .06 (-.25, .38) | k=10 (n=1 022) | .10 (-.03, .23) | N/A | N/A |
| Anxiety (2-6m F/U) | SMD (95%CI) | N/A | N/A | k=2 (n=145) | .03 (-.56, .61) | N/A | N/A | N/A | N/A |
| Depression (0-1m F/U) | SMD (95%CI) | k=22 (n=2 586) | -.04 (-.22, .14) | k=4 (n=781) | -.64 (-1.46, .18) | k=25 (n=3 125) | .01 (-.14, .17) | N/A | N/A |
| Depression (2-6m F/U) | SMD (95%CI) | k=10 (n=847) | .10 (-.10, .30) | k=3 (n=194) | -.21 (-.49, .07) | N/A | N/A | N/A | N/A |

| | | Removed: High Risk of Bias | | Removed: High Risk and Some Concern Risk of Bias | | Removed: Cluster RCT | | Removed: Active Control Group | |
|---|---|---|---|---|---|---|---|---|---|
| **Outcome** | **Statistic** | **Number of studies (participants)** | **Effect estimate** | **Number of studies (participants)** | **Effect estimate** | **Number of studies (participants)** | **Effect estimate** | **Number of studies (participants)** | **Effect estimate** |
| Depression (7-12m F/U) | SMD (95%CI) | k=2 (n=282) | -.15 (-.50, .20) | N/A | N/A | k=3 (n=631) | -.03 (-.30, .24) | k=2 (n=505) | .11 (-.07, 28) |
| Depression (13-24m F/U) | SMD (95%CI) | N/A | N/A | N/A | N/A | k=2 (n=283) | -.05 (-.29, .18) | N/A | N/A |

**Note**:

| No change on effect estimate | Effect estimate becomes significant | Effect estimate becomes non-significant |
|---|---|---|

**eTable 5. GRADE Summary Table**

| Outcomes | № of participants (studies) Follow-up | Certainty of the evidence (GRADE) | Relative effect (95% CI) | Anticipated absolute effects | |
|---|---|---|---|---|---|
| | | | | Risk with Control | Risk difference with Dyadic interventions |
| Bonding (0-1m F/U) | 1913 (14 RCTs) | ⊕◯◯◯ Very low[a,b,c] | - | - | SMD **0.43 SD higher** (0.08 higher to 0.78 higher) |
| Bonding (2-6m F/U) | 557 (4 RCTs) | ⊕◯◯◯ Very low[d,e,f] | - | - | SMD **0.27 SD higher** (0.19 lower to 0.73 higher) |
| Sensitivity (0-1m F/U) | 5291 (38 RCTs) | ⊕⊕◯◯ Low[d,e] | - | - | SMD **0.36 SD higher** (0.25 higher to 0.47 higher) |
| Sensitivity (2-6m F/U) | 990 (12 RCTs) | ⊕◯◯◯ Very low[c,d,e] | - | - | SMD **0.1 SD higher** (0.11 lower to 0.31 higher) |
| Sensitivity (7-12m F/U) | 980 (7 RCTs) | ⊕◯◯◯ Very low[a,e,f] | - | - | SMD **0.27 SD higher** (0.13 higher to 0.42 higher) |
| Sensitivity (13-24m F/U) | 329 (2 RCTs) | ⊕⊕⊕◯ Moderate[f] | - | - | SMD **0.25 SD higher** (0.03 higher to 0.47 higher) |

| Outcomes | № of participants (studies) Follow-up | Certainty of the evidence (GRADE) | Relative effect (95% CI) | Anticipated absolute effects | |
|---|---|---|---|---|---|
| | | | | Risk with Control | Risk difference with Dyadic interventions |
| Secure attachment (0-1m F/U) | 589 (8 RCTs) | ⊕⊕◯◯ Low[d,f] | OR 1.69 (1.20 to 2.39) | 473 per 1,000 | **130 more per 1,000** (46 more to 209 more) |
| Secure Attachment (2-6m F/U) | 214 (3 RCTs) | ⊕◯◯◯ Very low[c,d] | OR 1.16 (0.64 to 2.11) | 510 per 1,000 | **37 more per 1,000** (110 fewer to 177 more) |
| Secure Attachment (13-24m F/U) | 545 (4 RCTs) | ⊕◯◯◯ Very low[a,c,e] | OR 1.18 (0.51 to 2.75) | 599 per 1,000 | **39 more per 1,000** (167 fewer to 205 more) |
| Organized Attachment (0-1m F/U) | 331 (3 RCTs) | ⊕◯◯◯ Very low[c,d] | OR 2.23 (1.21 to 4.11) | 626 per 1,000 | **163 more per 1,000** (43 more to 247 more) |
| Insecure Attachment (0-1m F/U) | 437 (6 RCTs) | ⊕⊕⊕◯ Moderate[f] | OR 0.60 (0.36 to 0.99) | 516 per 1,000 | **126 fewer per 1,000** (239 fewer to 3 fewer) |
| Insecure Attachment (2-6m F/U) | 214 (3 RCTs) | ⊕◯◯◯ Very low[b,c,d] | OR 0.77 (0.43 to 1.40) | 373 per 1,000 | **59 fewer per 1,000** (169 fewer to 81 more) |

| Outcomes | № of participants (studies) Follow-up | Certainty of the evidence (GRADE) | Relative effect (95% CI) | Anticipated absolute effects | |
|---|---|---|---|---|---|
| | | | | Risk with Control | Risk difference with Dyadic interventions |
| Insecure Attachment (13-24m F/U) | 386 (2 RCTs) | ⊕◯◯◯ Very low[b,c,d] | OR 1.14 (0.31 to 4.12) | 263 per 1,000 | **26 more per 1,000** (163 fewer to 332 more) |
| Disorganized Attachment (0-1m F/U) | 499 (6 RCTs) | ⊕⊕◯◯ Low[c] | OR 5.51 (0.33 to 0.77) | 352 per 1,000 | **398 more per 1,000** (200 fewer to 57 fewer) |
| Disorganized Attachment (2-6 F/U) | 193 (2 RCTs) | ⊕◯◯◯ Very low[c,d,e] | OR 0.51 (0.12 to 2.27) | 209 per 1,000 | **90 fewer per 1,000** (178 fewer to 166 more) |
| Dyadic Interactions (0-1m F/U) | 2838 (25 RCTs) | ⊕◯◯◯ Very low[d,e,f] | - | - | SMD **0.22 SD higher** (0.10 higher to 0.34 higher) |
| Dyadic Interactions (2-6m F/U) | 963 (12 RCTs) | ⊕◯◯◯ Very low[b,c,d] | - | - | SMD **0.23 SD higher** (0.03 lower to 0.50 higher) |
| Dyadic Interactions (7-12m F/U) | 393 (4 RCTs) | ⊕⊕◯◯ Low[d,f] | - | - | SMD **0.19 SD higher** (0.02 higher to 0.36 higher) |

| Outcomes | № of participants (studies) Follow-up | Certainty of the evidence (GRADE) | Relative effect (95% CI) | Anticipated absolute effects | |
|---|---|---|---|---|---|
| | | | | Risk with Control | Risk difference with Dyadic interventions |
| Cognitive Development (0-1m F/U) | 188 (3 RCTs) | ⨁⨁◯◯ Low[f] | - | - | SMD **0.37 SD lower** (0.66 lower to 0.08 lower) |
| Cognitive Development (2-6m F/U) | 1036 (6 RCTs) | ⨁⨁⨁◯ Moderate[f] | - | - | SMD **0.09 SD higher** (0.03 lower to 0.21 higher) |
| Cognitive Development (7-12m F/U) | 662 (2 RCTs) | ⨁◯◯◯ Very low[c,e] | - | - | SMD **0.17 SD higher** (0.18 lower to 0.51 higher) |
| Cognitive Development (13-24m F/U) | 567 (6 RCTs) | ⨁⨁◯◯ Low[e,f] | - | - | SMD **0.24 SD higher** (0.01 lower to 0.49 higher) |
| Cognitive Development (25+m F/U) | 265 (2 RCTs) | ⨁◯◯◯ Very low[b,c] | - | - | SMD **0.19 SD higher** (0.44 lower to 0.82 higher) |
| Language Development (0-1m F/U) | 100 (2 RCTs) | ⨁⨁◯◯ Low[c] | - | - | SMD **0.08 SD lower** (0.48 lower to 0.31 higher) |

| Outcomes | № of participants (studies) Follow-up | Certainty of the evidence (GRADE) | Relative effect (95% CI) | Anticipated absolute effects | |
|---|---|---|---|---|---|
| | | | | Risk with Control | Risk difference with Dyadic interventions |
| Language Development (2-6m F/U) | 911 (5 RCTs) | ⊕⊕⊕◯ Moderate[f] | - | - | SMD **0.02 SD higher** (0.12 lower to 0.16 higher) |
| Language Development (7-12m F/U) | 645 (2 RCTs) | ⊕⊕◯◯ Low[c] | - | - | SMD **0.06 SD lower** (0.22 lower to 0.09 higher) |
| Language Development (13-24m F/U) | 259 (3 RCTs) | ⊕⊕◯◯ Low[c] | - | - | SMD **0.02 SD lower** (0.28 lower to 0.24 higher) |
| Motor Development (0-1m F/U) | 188 (3 RCTs) | ⊕⊕◯◯ Low[c] | - | - | SMD **0.23 SD lower** (0.52 lower to 0.06 higher) |
| Motor Development (2-6m F/U) | 902 (5 RCTs) | ⊕◯◯◯ Very low[b,c] | - | - | SMD **0.26 SD lower** (0.69 lower to 0.16 higher) |
| Motor Development (7-12m F/U) | 689 (3 RCTs) | ⊕◯◯◯ Very low[a,c] | - | - | SMD **0.06 SD lower** (0.21 lower to 0.09 higher) |

| Outcomes | № of participants (studies) Follow-up | Certainty of the evidence (GRADE) | Relative effect (95% CI) | Anticipated absolute effects | |
|---|---|---|---|---|---|
| | | | | Risk with Control | Risk difference with Dyadic interventions |
| Motor Development (13-24m F/U) | 408 (4 RCTs) | ⊕⊕◯◯ Low[c] | - | - | SMD **0.06 SD lower** (0.25 lower to 0.14 higher) |
| Socio-Emotional Development (2-6m F/U) | 1428 (3 RCTs) | ⊕⊕◯◯ Low[c] | - | - | SMD **0.1 SD higher** (0.17 lower to 0.37 higher) |
| Child Behavior (0-1m F/U) | 146 (2 RCTs) | ⊕◯◯◯ Very low[c,d] | - | - | SMD **0.23 SD lower** (0.55 lower to 0.1 higher) |
| Child Behavior (2-6m F/U) | 116 (2 RCTs) | ⊕◯◯◯ Very low[c,d] | - | - | SMD **0.02 SD lower** (0.39 lower to 0.34 higher) |
| Child Behavior (13-24m F/U) | 610 (6 RCTs) | ⊕◯◯◯ Very low[b,c,d] | - | - | SMD **0.35 SD higher** (0.03 lower to 0.74 higher) |
| Child Behavior (25+m F/U) | 237 (3 RCTs) | ⊕◯◯◯ Very low[b,c,d] | - | - | SMD **0.09 SD lower** (0.65 lower to 0.46 higher) |

| Outcomes | № of participants (studies) Follow-up | Certainty of the evidence (GRADE) | Relative effect (95% CI) | Anticipated absolute effects | |
|---|---|---|---|---|---|
| | | | | Risk with Control | Risk difference with Dyadic interventions |
| Parenting Stress (0-1m F/U) | 799 (13 RCTs) | ⊕◯◯◯ Very low[c,d] | - | - | SMD **0.06 SD higher** (0.12 lower to 0.24 higher) |
| Parenting Stress (2-6m F/U) | 469 (5 RCTs) | ⊕◯◯◯ Very low[a,c,e] | - | - | SMD **0.07 SD lower** (0.37 lower to 0.24 higher) |
| Parent/Caregiver Anxiety (0-1m F/U) | 1168 (11 RCTs) | ⊕⊕◯◯ Low[d,f] | - | - | SMD **0.16 SD higher** (0.01 higher to 0.3 higher) |
| Parent/Caregiver Anxiety (2-6m F/U) | 452 (4 RCTs) | ⊕◯◯◯ Very low[b,c] | - | - | SMD **0.34 SD higher** (0.11 lower to 0.8 higher) |
| Parent/Caregiver Depression (0-1m F/U) | 3271 (27 RCTs) | ⊕◯◯◯ Very low[d,e,f] | - | - | SMD **0.02 SD higher** (0.13 lower to 0.16 higher) |
| Parent/Caregiver Depression (2-6m F/U) | 996 (12 RCTs) | ⊕◯◯◯ Very low[c,d,e] | - | - | SMD **0.18 SD higher** (0.03 lower to 0.38 higher) |

| Outcomes | № of participants (studies) Follow-up | Certainty of the evidence (GRADE) | Relative effect (95% CI) | Anticipated absolute effects | |
|---|---|---|---|---|---|
| | | | | Risk with Control | Risk difference with Dyadic interventions |
| Parent/Caregiver Depression (7-12m F/U) | 505 (3 RCTs) | ⊕◯◯◯ Very low[c,d,e] | - | - | SMD **0.03 SD lower** (0.3 lower to 0.24 higher) |
| Parent/Caregiver Depression (13-24m F/U) | 283 (2 RCTs) | ⊕◯◯◯ Very low[c,d] | - | - | SMD **0.05 SD lower** (0.29 lower to 0.18 higher) |

***The risk in the intervention group** (and its 95% confidence interval) is based on the assumed risk in the comparison group and the **relative effect** of the intervention (and its 95% CI).

**CI:** confidence interval; **OR:** odds ratio; **SMD:** standardised mean difference

**GRADE Working Group grades of evidence**
**High certainty:** we are very confident that the true effect lies close to that of the estimate of the effect.
**Moderate certainty:** we are moderately confident in the effect estimate: the true effect is likely to be close to the estimate of the effect, but there is a possibility that it is substantially different.
**Low certainty:** our confidence in the effect estimate is limited: the true effect may be substantially different from the estimate of the effect.
**Very low certainty:** we have very little confidence in the effect estimate: the true effect is likely to be substantially different from the estimate of effect.

Summary of findings tables were generated using GRADE profiler Guideline Development Tool software and GRADE criteria (2015, McMaster University and Evidence Prime Inc.).

#### eMethods1. Full Systematic Review Method

**Eligibility Criteria**

We considered randomized controlled trials (RCT) of any type that compared an early dyadic parent/caregiver-infant intervention to any comparator. Only English or French-language papers, as well as published and unpublished study results retrieved from corresponding authors, were eligible. In order to focus on contemporary interventions with the potential for further development and widespread implementation, we limited the review to studies published on or after January 1, 2000.

*Population*. Any parent/caregiver-child dyad.

*Intervention*. We defined as "dyadic" any intervention that targeted at least one primary caregiver and the infant together, as a dyad, by promoting physical, emotional, or face-to-face interactions, e.g., skin-to-skin, massage, video interaction guidance. Interventions were eligible for inclusion if at least one intervention session occurred within the first 6 months of the infant's life. Prenatal interventions without postnatal sessions were excluded. No exclusion criteria were considered for the length or intensity of the intervention.

*Comparator*. Any type of comparator was considered, i.e., control or active intervention.

*Outcomes*. The primary outcome domain was ERH. Specific outcomes within this domain included established and validated proxy measures of ERH, e.g., attachment, sensitivity, bonding, emotional connection. Secondary outcome domains were child socio-emotional functioning and development, and parent/caregiver mental health. Secondary outcomes were only considered in studies where at least one ERH outcome was also measured.

**Information Sources and Search Strategy**

Systematic literature searches were conducted in five databases: PubMed, Medline, Cinhal, ERIC, and PsycInfo via EBSCO. The search strategy was designed to focus on two main concepts: (1) parent-infant dyadic interventions and (2) ERH (see eMethods 2 for full search strategies). Searches were conducted on April 28, 2022. To identify unpublished and ongoing studies, we searched clinical trial registries (clinicaltrials.gov, ISRCTN Registry, EU Clinical Trials Register, Australian New Zealand Clinical Trials Registry). Authors of unpublished and ongoing studies were contacted. After completing the selection process described below, we conducted backward, and forward reference searching of included studies.

**Selection Process**

Identified study records were uploaded to EndNote V9.3.3[111] and subsequently to Covidence.[112] Duplicates were removed and study records were screened for eligibility independently by teams of two review authors. Disagreements were resolved by the first author for consistency. The same process was followed for the full-text review of potentially eligible studies. Reasons for exclusion were documented at the full-text screening stage.

**Data collection process**

Data extraction was performed in duplicate and independently, using a data extraction form specifically developed for this review. Consensuses were done by the first author. We extracted data at the study-, intervention-, and outcome-level.

- Study-level: aim, study design, number of groups, target population, inclusion and exclusion criteria, group differences, unit of randomization, sample size and withdrawals.
- Intervention-level, based on the Template for Intervention Description and Replication (TIDieR)[113] checklist: name of the intervention, framework or underlying theory, rationale, materials, dyadic and non-dyadic procedures, provider(s), location(s), dose, and intensity.
- Result-level: name of the outcome, scale, description of the procedure if outcome was observational, timing and age of infant at assessment, means or medians, standard deviations (SD), standard error (SE), 95% confidence interval (CI) or Inter Quartile Range (IQR), or events for binary outcomes.

Raw unadjusted means analyzed in intention to treat were preferred over adjusted means and medians for consistency across studies. Only post-test measures were extracted, except for studies where outcomes were measured after one full year of a much longer intervention.[93,114] SE, 95% CI and IQR were converted to SDs following the Cochrane guidelines.[115] If numerical data were only presented in figures, we extracted data from the figures using software such as Engauge Digitizer[116] v12.1 and PlotDigitizer[117]. When necessary, we contacted corresponding authors to retrieve missing data.

**Data Items**
The primary outcome of this systematic review was parent/caregiver-child ERH measured at any time point. Established child social and developmental outcomes and caregiver mental health were considered as secondary outcomes.

**Effect Measures and Synthesis Methods**
We performed pairwise meta-analyses on three outcome domains where at data from at least two studies was available, looking at specific outcomes separately:

i) Primary Outcome Domain – ERH:
   a. self-reported maternal bonding,
   b. observed parent/caregiver sensitivity,
   c. observed child attachment,
   d. observed dyadic interactions.

ii) Secondary Outcome Domain - Child Socio-Emotional Functioning and Development:
   a. parent/caregiver reported child behaviors,
   b. parent/caregiver-reported child socio-emotional development,
   c. observed cognitive development,
   d. observed language development,
   e. observed motor development.

iii) Secondary Outcome Domain – Parent/Caregiver Mental Health:
   a. self-reported parenting stress,
   b. self-reported anxiety,
   c. self-reported depression.

As only four studies used an active intervention as a comparator, our pairwise comparison was early dyadic intervention vs. control, i.e., standard care, attention-control, or active control. Sensitivity analyses were conducted to determine whether pooled effect estimates were robust to the inclusion of the active controls. To facilitate pooling of studies, we also conducted separate analyses per follow-up time point, follow-up (F/U) time points, i.e., end of intervention (0) to 1-month F/U, 2- to 6-month F/U, 7- to 12-month F/U, 13- to 24-month F/U and 25 months and up. To account for the different child ages at assessment, we did a moderation analysis to test whether effect estimates differed by child age at assessment. When studies had more than one outcome within a domain for the same time frame, or multiple time points within a time frame, we chose the outcome that provided the most comprehensive measure of the domain, and the latest time point within a time frame.

The unit of analysis was parent/caregiver-infant dyads. To avoid unit-of-analysis issues, we combined cluster-RCTs with individually randomized trials by using a direct estimate of the required effect measure, e.g., odds ratio with 95% CI, from analyses that properly accounted for the cluster design. Alternatively, based on Cochrane guidelines,[118] we used the intra-cluster correlation coefficient (ICC) to correct the sample size to account for the design effect. ICCs were ideally extracted from the study itself, or from a similar trial. However, when neither adjusted analyses, nor the mean cluster size were reported, we couldn't adjust for the design effect and accounted for it in our sensitivity analyses. For studies with three arms, we combined intervention groups together, combined two control groups together, or dropped one of the three groups, depending on what was most appropriate. A summary of our methodological decisions to avoid unit-of-analysis issues is presented in eTable2.

Continuous outcomes were analyzed using weighted standardized mean differences (SMD) with 95% CIs. Categorical variables were analyzed using weighted odds ratios (OR) with 95% CIs. An inverse variance random-effects model to account for study design and patient population induced variability was chosen. Significance level was set at 0.05 for all analyses. Cochran's Q test and I2 statistic were used to assess the heterogeneity and I2 values >50% or p-value <0.10 indicated statistically significant heterogeneity. We also conducted a sensitivity analysis to examine if our results were robust to the inclusion of studies with an overall concerning or high risk of bias. Additionally, we tested the subset of 20 studies reporting developmental outcomes on ERH effect estimates.

In addition to our preregistered moderator 'intervention dose' (in provider minutes), we tested the effects of several other commonly considered parent/caregiver/family-level moderators (target population, e.g., adolescent mothers, incarcerated mothers, and low vs high socio-economic status), infant-level moderators (infant sex, infant age at first session, age at follow-up, and preterm/low birthweight versus term/normal birthweight), intervention-level moderators (length of intervention in days, number of sessions, provider type, location), and study-level moderators (year of publication, and pre-registration as an analogue of study quality). Continuous moderators were explored with a meta-regression and categorical moderators with a *Q*-

test,[119] both using a random-effects DerSimonian Laird model. Moderator analyses were conducted on each outcome pooling at least 10 studies, and with more than one study in each category in the case of categorical moderators.All analyses were conducted with Review Manager V5.4[120] and SPSS V24.[121]

**Study Risk of Bias and Certainty Assessment**

Two independent review authors assessed risk of bias in individual studies using the Cochrane Collaboration Risk of Bias Tool 2.[122] Confidence in pooled outcomes was based on the Grades of Recommendation, Assessment, Development and Evaluation (GRADE) guidelines.[123] Summary of findings tables were generated using the GRADE profiler Guideline Development Tool software and the GRADE criteria (2015, McMaster University and Evidence Prime Inc.). Within the GRADE assessment, we estimated the risk of publication bias by funnel plot inspection.

#### eMethods2. Full Search Strategies

Run date: 04/28/2022

| Database | Total |
|---|---|
| PubMed | 8 742 |
| CINAHL | 1 075 |
| ERIC | 213 |
| PsycInfo | 1 100 |
| Medline | 1 127 |
| **Total** | **12 257** |

#### PubMed

| | | Results |
|---|---|---|
| 1 | "treatment outcome"[MeSH Terms] OR "early intervention, educational"[MeSH Terms] OR (Program*[Title/Abstract]) OR (Intervention*[Title/Abstract]) | 3 058 072 |
| 2 | ((((Infant[MeSH Terms]) OR (infant, newborn[MeSH Terms])) OR (infant, premature[MeSH Terms])) OR (child, preschool[MeSH Terms])) OR (((((Babies[Title/Abstract]) OR (Infant*[Title/Abstract])) OR (Baby[Title/Abstract])) OR (Newborn*[Title/Abstract])) OR (Child*[Title/Abstract])) | 2 714 044 |
| 3 | (((((parenting[MeSH Terms]) OR (maternal behavior[MeSH Terms])) OR (mother child relations[MeSH Terms])) OR (parent child relations[MeSH Terms])) OR (object attachment[MeSH Terms])) OR ((Connect*[Title/Abstract]) OR (Nurtur*[Title/Abstract]) OR (Sensitiv*[Title/Abstract]) OR (Responsiv*[Title/Abstract]) OR (Attach*[Title/Abstract]) OR (Bond*[Title/Abstract]) OR (Interact*[Title/Abstract]) OR (Relation*[Title/Abstract])) | 5 942 408 |
| | 1 AND 2 AND 3 | 111 815 |
| 8 | ((clinicaltrial[Filter] OR randomizedcontrolledtrial[Filter]) AND (english[Filter] OR french[Filter]) AND (2000:2022[pdat])) | 8 742 |

#### Cinhal via EBSCO

| | | **Results** |
|---|---|---|
| 1 | TI ( (infant* OR bab* OR newborn* OR birth*) ) OR AB ((infant* OR bab* OR newborn* OR birth*)) | 243 685 |
| 2 | TI ( (parent* OR mother* OR father* OR maternal OR paternal OR famil*) N5 (attach* OR bond* OR interact* OR relation* OR sensitiv* OR respons* OR nurtur*) ) OR AB ( (parent* OR mother* OR father* OR maternal OR paternal OR famil*) N5 (attach* OR bond* OR interact* OR relation* OR sensitiv* OR respons* OR nurtur*) ) | 56 291 |
| 3 | (MH "Parent-Child Relations+") OR (MH "Parenting") OR (MH "Maternal Behavior") OR (MH "Parental Behavior") OR (MH "Family Relations") OR (MH "Attachment Behavior") OR (MH "Nurturing Behavior") OR (MH "Family | 115 171 |

| | | |
|---|---|---|
| | Attitudes+") OR (MH "Parental Attitudes+") OR (MH "Mothers/PF") OR (MH "Fathers/PF") OR (MH "Parents/PF") | |
| 4 | 2 OR 3 | 145 916 |
| 5 | (TI attachment N3 intervention OR AB attachment N3 intervention) OR (TI parent* N3 training OR AB parent* N3 training) OR (TI parent* N3 intervention OR AB parent* N3 intervention) OR (TI family N3 intervention OR AB family N3 intervention) OR (TI relationship N3 intervention OR AB relationship N3 intervention) OR (TI bonding N3 intervention OR AB bonding N3 intervention) OR (TI family N3 training OR AB family N3 training) | 18 192 |
| 6 | 1 AND 4 AND 5<br>Limited to 2000 to 2022 | 1 075 |

#### ERIC via EBSCO

| | | **Results** |
|---|---|---|
| 1 | TI ( (infant* OR bab* OR newborn* OR birth*) ) OR AB ((infant* OR bab* OR newborn* OR birth*)) | 28 876 |
| 2 | TI ( (parent* OR mother* OR father* OR maternal OR paternal OR famil*) N5 (attach* OR bond* OR interact* OR relation* OR sensitiv* OR respons* OR nurtur*) ) OR AB ( (parent* OR mother* OR father* OR maternal OR paternal OR famil*) N5 (attach* OR bond* OR interact* OR relation* OR sensitiv* OR respons* OR nurtur*) ) | 33 811 |
| 3 | (SU "Parent Child Relationship") OR (SU "Parenting skills") OR (SU "Parenting styles") OR (SU "Parents") OR (SU "Family Relationship") OR (SU "Attachment Behavior") OR (SU "Family Attitudes+") OR (SU "Mothers attitudes") OR (SU "Fathers attitudes") | 48 876 |
| 4 | 2 OR 3 | 66 552 |
| 5 | (TI attachment N3 intervention OR AB attachment N3 intervention) OR (TI parent* N3 training OR AB parent* N3 training) OR (TI parent* N3 intervention OR AB parent* N3 intervention) OR (TI family N3 intervention OR AB family N3 intervention) OR (TI relationship N3 intervention OR AB relationship N3 intervention) OR (TI bonding N3 intervention OR AB bonding N3 intervention) OR (TI family N3 training OR AB family N3 training) | 8 710 |
| 6 | 1 AND 4 AND 5<br>Limited to 2000 to 2022 | 213 |

#### PsychInfo via EBSCO

| | | **Results** |
|---|---|---|
| 1 | TI ( (infant* OR bab* OR newborn* OR birth*) ) OR AB ((infant* OR bab* OR newborn* OR birth*)) | 156 874 |
| 2 | TI ( (parent* OR mother* OR father* OR maternal OR paternal OR famil*) N5 (attach* OR bond* OR interact* OR relation* OR sensitiv* OR respons* OR nurtur*) ) OR AB ( (parent* OR mother* OR father* OR maternal OR paternal OR famil*) N5 (attach* OR bond* OR interact* OR relation* OR sensitiv* OR respons* OR nurtur*) ) | 148 661 |

| | | |
|---|---|---|
| 3 | (MA "Parent Child Relations") OR (MA "Parenting") OR (MA "Family Relations") OR (MA "Attachment Behavior") OR (MA "Family Attitudes+") OR (MA "Parental Attitudes+") OR (MA "Mothers Child Relations") OR (MA "Fathers Child Relations") OR (MA "Parents/PF") | 29 549 |
| 4 | 2 OR 3 | 167 851 |
| 5 | (TI attachment N3 intervention OR AB attachment N3 intervention) OR (TI parent* N3 training OR AB parent* N3 training) OR (TI parent* N3 intervention OR AB parent* N3 intervention) OR (TI family N3 intervention OR AB family N3 intervention) OR (TI relationship N3 intervention OR AB relationship N3 intervention) OR (TI bonding N3 intervention OR AB bonding N3 intervention) OR (TI family N3 training OR AB family N3 training) | 32 454 |
| 6 | 1 AND 4 AND 5<br>Limited to 2000 to 2022 | 1 100 |

#### MEDLINE via EBSCO

| | | **Results** |
|---|---|---|
| 1 | TI ( (infant* OR bab* OR newborn* OR birth*) ) OR AB ((infant* OR bab* OR newborn* OR birth*)) | 849 745 |
| 2 | TI ( (parent* OR mother* OR father* OR maternal OR paternal OR famil*) N5 (attach* OR bond* OR interact* OR relation* OR sensitiv* OR respons* OR nurtur*) ) OR AB ( (parent* OR mother* OR father* OR maternal OR paternal OR famil*) N5 (attach* OR bond* OR interact* OR relation* OR sensitiv* OR respons* OR nurtur*) ) | 131 479 |
| 3 | (MH “Parenting”) OR (MH “Maternal Behavior”) OR (MH “Mother-Child Relations”) OR (MH “Paren-Child Relations”) OR (MH “Object Attachment”) OR (MH "Family Relations") | 68 310 |
| 4 | 2 OR 3 | 178 506 |
| 5 | (TI attachment N3 intervention OR AB attachment N3 intervention) OR (TI parent* N3 training OR AB parent* N3 training) OR (TI parent* N3 intervention OR AB parent* N3 intervention) OR (TI family N3 intervention OR AB family N3 intervention) OR (TI relationship N3 intervention OR AB relationship N3 intervention) OR (TI bonding N3 intervention OR AB bonding N3 intervention) OR (TI family N3 training OR AB family N3 training) | 25 552 |
| 6 | 1 AND 4 AND 5<br>Limited to 2000 to 2022 | 1 127 |

#### eMethods 3. Parent/Caregiver-Report Assessments of Relational Health

Among the 93 studies included in the systematic review, we identified 12 distinct Parent/Caregiver-reported assessments of relational health. An overview of the main characteristics of these scales is presented in the table below and an in-depth description of these scales is subsequently provided. Based on information provided in the reviewed literature, a narrative description of each observational assessment is also provided after the table (links auto-direct to a description of each observation assessment). Note that summaries of each instrument are drawn only from the reviewed studies; detailed information about instrument development, and validity are not included in scale descriptions.

##### Main Characteristics of Parent/Caregiver-reported Assessments of relational health

| Scale | Acronym | Construct measured | Main reference(s) | Studies using this scale |
|---|---|---|---|---|
| Attachment Diaries | -- | Attachment behaviors during distressing situations | Stovall and Dozier 2000[124] | Dozier 2009[89] |
| Maternal Attachment Inventory | -- | Caregiver attachment and emotional bonding within the dyad | Muller 1994[125] | Heo 2019[61]<br>Vargas-Porras 2021[80] |
| Mother-Infant Bonding Scale | -- | Maternal rating of mother-infant bond | -- | Cevasco 2008[9] |
| Mother-to-Infant Bonding Scale | MIBS | Mother's feelings toward her infant | -- | Corrigan 2021[10] |
| Mother-to-Child Attachment | MCA | -- | -- | Youn 2021[105] |
| Mother-Infant Bonding Inventory | -- | Mother's thoughts, feelings and commitment toward her baby | Sheih, Ying, Li, and Hsieh 2015[126] | Yu 2022[14] |
| My Baby and I Questionnaire | MBI | Parent-infant relationship | Furman and O'Riordan[127] | Hoffenkamp 2015[32] |
| Parent-to-Infant Attachment Questionnaire | PIA | Parent-child relationship quality | -- | Barnes 2022[20] |
| Parenting Your Baby Questionnaire | PYB | Parental warmth | -- | Canfield 2020[42]<br>Roby 2021[41] |
| Postpartum Bonding Questionnaire | PBQ | Mother-infant bonding | Brockington, Oates, and George 2001; Brockington, Fraser, and Wilson 2006 [128,129] | Gaden 2022[11]<br>Hoffenkamp 2015[32]<br>Holt 2021[110]<br>Tsivos 2015[78]<br>Williams 2020[8]<br>Wulff 2021[13] |
| Working Model of the Child Interview | WMCI | Maternal feelings about their relationship with their child | Zeanah and Benoit 1995[130] | Meijssen 2010[68]<br>Meijssen 2011[69] |
| Yale Inventory of Parental Thoughts and Actions | YIPTA | Parental bonding and postpartum distress | Feldman, Weller, Leckman, Kuint, and Eidelman, 1999[131] | Hoffenkamp 2015[32] |

#### Attachment Diaries

The Attachment Diaries allow parents to self-report infants' behaviors during distressing situations.[124]

**Populations.** Not specified.

**Scoring.** Parents document infants' initial attachment-seeking behaviors as well as their own behavioral responses and the infants' reactive responses during distressing situations. They also provide a narrative of the incident. Parents keep track of these incidents over the course of three days. Scores are determined by summing the instances of proximity seeking by the child, successful calming by the parent, avoidance, or resistance. Proximity seeking and successful calming scores are summed to achieve a secure score for behavior. Behaviors on behalf of both the child and parent can be classified as secure, avoidant, or resistant. Secure behaviors include moving toward the parent or being soothed by the parent without expressions of anger or ambivalence. Avoidant behaviors include suppressing hurt or fear or moving away from the parent. Resistant behaviors include outward displays of angry behavior such as kicking, screaming, biting, or continual fussiness. Documented situations that are deemed not sufficiently distressing (i.e., child left with a familiar caregiver) are considered not relevant and missing.

**Psychometric Properties.** Not specified.

#### Attachment Inventory

The Maternal Attachment Inventory measures caregiver attachment and emotional bonding to their infants.[125]

**Populations.** This scale has been culturally adapted and translated into Korean[132] and Spanish.[133] It has been validated for use in Colombian first-time mothers of term infants and for parents with preterm infants.

**Scoring.** This self-report questionnaire is comprised of 24[80] to 26[61] items rated on a 4-point Likert scale. Higher scores correlate with better attachment to infants. Scores are rated as low (85 or less), moderate (86 to 89 points), or high (90 or more).

**Psychometric properties.** Cronbach's alpha ranges from 0.85[61] to 0.90.[80]

#### Mother-Infant Bonding Scale

The Mother-Infant bonding scale is a self-report tool used to assess the quality of maternal-infant bond.

**Populations.** Not specified.

**Scoring.** Bonding is assessed using a 5-point Likert scale for four indices relating to how the mother rates her relationship to her infant. A low score of 4 indicates low mother-infant bonding, and a high score of 20 indicates a positive mother-infant bond.

**Psychometric properties.** Not specified.

#### Mother-to-Infant Bonding Scale (MIBS)

The Mother-Infant bonding scale is used to measure mother-infant bonding by assessing the degree of emotional connectedness mothers have toward their infants.[134]

**Populations.** Not specified.

**Scoring.** This self-report tool directs mothers to rank a series of 8 different feelings they may experience in relation to their infant, such as loving, neutral or resentful on a scale from very much like them, to not at all like them. This scale is scored 0-24, with higher scores indicating worse mother-infant bonding, and scores of 2 or more indicating clinically compromised maternal-infant bonding.

**Psychometric properties.** The MIBS has been found to have acceptable criterion-related validity and construct validity.[135]

#### Mother-to-Child Attachment (MCA)

The Mother-to-Child Attachment (MCA) scale is used to measure attachment in mother-child dyads.[105]

**Populations.** Not specified.

**Scoring.** Not specified.

**Psychometric properties.** Not specified.

#### Mother-Infant Bonding Inventory

Mother-Infant Bonding Inventory is used to measure maternal thoughts, feelings and commitment in relation to her infant.

**Populations**. Not specified.

**Scoring.** The scale consists of four domains: proximity, parental adjustment, commitment, and confidence of reciprocity. Each item was scored using a 6-point Likert scale with scores of 1-6 corresponding to strongly disagree, disagree, slightly disagree, slightly agree, agree, and strongly agree, respectively. Higher scores indicate better bonding.

**Psychometric properties.** The Cronbach's α of the inventory was 0.88–0.89.[126]

#### My Baby and I Questionnaire (MBI)

The My Baby and I Questionnaire is used to assess the parent's feelings about the parent-child relationship in terms of responsiveness, enjoyment, and separation anxiety.

**Populations**. Not specified.

**Scoring.** MBI-W measures worry in three items, producing scores ranging from 3-15. Greater scores on the MBI-W indicate greater infant-related concerns. MBI-ER measures Enjoyment and Responsiveness in seven items, producing scores from 7-31. Higher scores on the MBI-ER indicate more positive feelings about the infant and higher responsiveness to the infant. MBI-SA measures separation anxiety in four items, producing scores of 4-20. Greater scores on the MBI-SA indicate greater parental anxiety when leaving the infant.

**Psychometric properties**. Internal consistency ranged from acceptable to very good across the dimensions MBI-W (mothers: 0.90 and 0.77, fathers: 0.88 and 0.75), MBI-ER (mothers: 0.83 and 0.60, fathers: 0.83 and 0.67), and MBI-SA (mothers: 0.70 and 0.82, fathers: 0.70 and 0.75).[32]

#### Parent-to-Infant Attachment Questionnaire (PIA)

The Parent-to-Infant Attachment Questionnaire (PIA) is a self-report tool that is used to assess the emotional bond or tie of affection experienced by the parent toward the infant. The Parent-to-infant attachment questionnaire (PIA56) is a 19-item scale with 3 subscales including quality of attachment, absence of hostility, and pleasure in interaction.

**Populations**. Not specified.

**Scoring.** Not specified.

**Psychometric properties**. The test-retest reliability and internal consistency of the instrument are acceptably high.[136] All 19 items have high face validity in terms of reflecting various facets of the emotional experience of the parent toward their infant.[136] The criterion validity of the instrument has yet to be established.[136]

#### Parenting Your Baby Questionnaire (PYB)

The Parenting your baby Questionnaire (PYB) is a self-report questionnaire regarding parental experience. In our study, two authors utilized the PYB's Supporting and Enjoying subscale to measure parental warmth.

**Populations**. This scale has shown high construct, convergent, and predictive validity in children at school entry[137] and has been widely used with toddlers and young children.[42]

**Scoring.** This eight-item subscale asks parents about their parenting activities and feelings in the past month, including items like "play with your child in a way that was fun for him/her" and "feel confident in reading your child's cues." Items are rated on a 7-point scale from 1 (not at all) to 7 (most of the time).

**Psychometric properties**. This scale has shown high construct, convergent, and predictive validity.[41] Cronbach's alpha in Roby (2021)'s sample ranged from 0.67 to 0.73.[41]

#### Postpartum Bonding Questionnaire (PBQ)

The Postpartum Bonding Questionnaire (PBQ) is a self-report tool that measures mother-infant bonding in the period following birth.[128] The PBQ targets the mother's feelings, experiences and attitudes toward her infant.[11]

**Populations**. The PBQ has been validated in the UK,[128,129,138] Germany,[139] Japan,[140,141] Spain,[142] Italy,[143] India,[144] and France,[145] in general and clinical populations.

**Scoring.** The PBQ contains four subscales: general impaired bonding, rejection and pathologic anger, anxiety about the infant, and incipient abuse.[11] The questionnaire consists of statements about the mother's feelings toward her infant, rated on a 6-point Likert scale with responses ranging from "always" to "never." The total score ranges from 0 to 125, with higher scores indicating an impaired bond.[11,32]

**Psychometric properties**. The PBQ has good internal consistency and high test-retest reliability.[78] The PBQ has also been found to have acceptable criterion and construct validity.[135]

#### Working Model of the Child Interview (WMCI)

The Working Model of the Child Interview aims to elicit parents' feelings about their relationships with their children.[130] Developed in reference to the Adult Attachment Interview (AAI), the WMCI focuses on the parent's emotional reactions during pregnancy, perception of the infant's personality and development, characteristics of the bond with the infant, reactions to infant behavior, and anticipated difficulties in later development.[68] Additionally, parents are asked to provide anecdotes about their infant that would help illustrate the child's personality and behavior.

**Populations**. Not specified.

**Scoring.** The interviews are videotaped and take about 60 minutes to complete. The children are not present for the interview. The interviews are rated with a 5-point Likert scale in six scales: richness of perception, openness to change, intensity of involvement, coherence, caregiving sensitivity and acceptance. Based on the caregiver's narrative answers to the questions, their children are then classified by score into one of three attachment representations: balanced (secure), disengaged (insecure), or distorted (insecure).

**Psychometric properties**. Not specified.

#### Yale Inventory of Parental Thoughts and Actions (YIPTA)

The Yale Inventory of Parental Thoughts and Actions (YIPTA) measures parental bonding and distress postpartum.

**Populations.** The YIPTA has been validated for use in parents with term or preterm infants.

**Scoring.** The self-report questionnaire is comprised of 27-items falling into five subscales:
Frequency of Thoughts and Worries (YIPTA-FTW; 9-items), Distress Caused by Thoughts and Worries (YIPTA-DTW; 5-items), Compulsive Checking (YIPTA-CC; 4-items), Affiliative Behavior (YIPTA-AB; 5-items), and

Attachment Representations (YIPTA-AR; 4-items). Each item is scored on a 4-point scale, for a maximum overall score of 108. Higher scores correlate to more infant-related worries and distress, or greater bonding and caregiver behaviors depending on the subscale.

**Psychometric properties.** The scale exhibited strong internal consistency for YIPTA-FTW (mothers: 0.92; fathers: 0.90), YIPTA-DTW (mothers: 0.87; fathers: 0.86), and YIPTA-CC (mothers: 0.81; fathers: 0.77). The scale demonstrated moderate consistency for YIPTA-AB (mothers: 0.55; fathers: 0.66) and YIPTA-AR (mothers: 0.55; fathers: 0.59).[32]

#### eMethods 4. Observational Assessments of Relational Health

Among the 93 studies included in the systematic review, we identified 41 observational assessments of relational health. An overview of the main characteristics of these scales is presented in the table below. Based on information provided in the reviewed literature, a narrative description of each observational assessment is also provided after the table (links auto-direct to a description of each observation assessment). Note that summaries of each instrument are drawn only from the reviewed studies; detailed information about instrument development, validity, and an exhaustive list of paradigms are not included in scale descriptions. For a systematic review of observational coding instruments for use in infancy, see Lotzin et al., 2015.[146]

##### Observational Assessments of Relational Health

| Scale | Acronym | Construct measured | Main reference(s) | Paradigm(s) | Studies using this assessment |
|---|---|---|---|---|---|
| Adult Play Scale | -- | Physical and emotional child-caregiver connection | Howes and Stewart 1987[147] | Free play | Constantino 2001[56] |
| Ainsworth's Maternal Sensitivity Scales | -- | Maternal sensitivity towards the infant | Ainsworth, Bell, and Slayton 1971, 1974[148,149] | Free play, Bathing | Klein Velderman 2006[36] Rheinheimer 2022[4] Sahlen-Helmer 2019[5] |
| Attachment Q-Sort | AQS | Infant attachment behavior | Waters 1995[150] | Naturalistic dyadic home observation | Klein Velderman 2006[36] Stein 2018[44] van Doesum 2008[48] |
| Attachment Story Completion Task | ASCT | Child attachment representations | Verschueren and Marcoen 1994[151] Bretherton, Ridgeway, and Cassidy 1990[152] | Child-led story completions | van Doesum 2008[48] |
| Atypical Maternal Behavior Instrument for Assessment and Classification Scale | AMBIANCE | Affective communication and behavior between caregivers and infants | Bronfman, Madigan, and Lyons-Ruth 1992-2009[153] Bronfman, Parsons, and Lyons-Ruth 1992-2008[154] | Face-to-face dyadic interactions | Sadler 2013[93] Tereno 2017[45] |
| Avant Maternal Attachment Behaviors Scale | -- | Dyadic attachment | Avant 1978[155] | Face-to-face interaction | Shoghi 2018[24] |
| Barrier Task | -- | Regulation and dysregulation of child behaviors | Goldsmith and Rothbart 1993[156] | Barrier Task | Valades 2021[79] |
| Child Adult Relationship Evaluation Index | CARE-Index | Quality and patterns of parent-infant interaction | Crittenden 1979-2004[157] Crittenden 1979-2010[158] | Free play, Semi-structured play | Barlow 2016[27] Borghini 2014[28] Nugent 2017[72] Oxford 2021[39] |

| Scale | Acronym | Construct measured | Main reference(s) | Paradigm(s) | Studies using this assessment |
|---|---|---|---|---|---|
| | | | | | Tryphonopoulos 2020[46] Tsivos 2015[78] Twohig 2021[47] |
| Coding Interactive Behavior | CIB | Maternal and infant behavior and quality of mother-infant interactions | Feldman 1998[159] | Free play | Fonagy 2016[90] Sleed 2013[97] Steele 2019[98] |
| Coding system by Piccinini, Alvarenga and Frizzo (2007) | -- | Maternal sensitivity in maternal-infant interactions | Piccinini, Alvarenga and Frizzo, 2007[160] | Free play | Alvarenga 2020[26] |
| Dyadic Mutuality Code | DMC | Responsivity, mutuality, and synchrony in the dyadic relationship | Censullo, 1991; Censullo, Bowler, Lester, & Brazelton, 1987 | Free play | Horowitz 2001[63] Nelson 2001[107] White-Traut 2013[108] |
| Emotional Availability Scales and Emotional Attachment Zones Evaluation | EAS EA-Z | Quality of dyadic interactions and the global dyadic relationship | Biringen, 2008; EA-Z, Wurster, Sarche, Trucksess, Morse, & Biringen, 2019; Biringen, Robinson, & Emde, 1998 | Free play | Firk 2021[31] Fonagy 2016[90] Georg 2021[91] Hoivik 2015[33] Klein Velderman 2006[36] Salo 2019[75] van Doesum 2008[48] |
| Eye gaze | -- | Quality of mother-infant interactions | Beebe, Jaffe, Markese, Buck, Chen, Cohen, Bahrick, Andrews and Feldstein [161] | Face-to-face interaction, Free play | Hane 2015[50] |
| Global Rating Scale | GRS | Dyadic interaction patterns and overall quality of dyadic interactions | Murray, 1996[162] | Still Face (reunion phase) | Valades 2021[79] Williams 2020[8] Zelkowitz 2011[81] |
| Holditch-Davis Coding System | -- | Mother-infant interaction behavior patterns | Holditch-Davis et al., 2007[163]; Holditch-Davis, Miles, Burchinal, & Goldman, 2011[164] | Naturalistic dyadic interaction | Holditch-Davis 2014[106] |

| Scale | Acronym | Construct measured | Main reference(s) | Paradigm(s) | Studies using this assessment |
|---|---|---|---|---|---|
| The Home Observation for Measurement of the Environment Inventory | HOME-Inventory | Quality of the home environment in supporting child development | Caldwell and Bradley, 1978[165] | Observations of the home environment | Alarcao 2021[83] Badr 2016[100] Betancourt 2020[54] Glazebrook 2007[60] Holditch-Davis 2014[106] |
| Index of Parental Behavior | IPB | Quality of dyadic interactions | Melnyk et al., 1998[166] | Face-to-face dyadic interactions | Melnyk 2006[71] |
| Infant and Caregiver Engagement Phases | ICEP | Infant and caregiver affect, engagement, and the quality of the engagement | Weinberg & Tronick, 1999[167] | Still Face | Meijssen 2010[68] |
| Landry Parent-Child Interaction Scales | -- | Quality of maternal-infant interactions | Landry et al., 1996[168]; Landry, Smith, Miller-Loncar, & Swank, 1998[169] | Freeplay, Book reading activities | Baggett 2010[16] Baggett 2017[17] Feil 2020[18] |
| The Looking, Touching, Talking, Smiling Parent-Infant Interaction Coding Scale | LoTTS | Frequency of interaction behaviors | Beatty et al., 2011[170] | Free play | Robertson 2019[12] |
| Maternal Behavioral Q-set | MBQ | Maternal sensitivity toward the infant | Pederson & Moran, 1995[171] | Naturalistic observation | Teti 2009[25] |
| Maternal Caregiving Behaviors | -- | Quality of feeding, holding, and other maternal caregiving behaviors | Hane & Fox, 2006[172]; Hane & Philbrook, 2012[173] | Feeding and holding interactions | Hane 2015[50] |
| Maternal Interactive Behaviors Likert Scales | -- | Quality of maternal interactive behavior | Erickson, Sroufe & Egeland, 1985[174]; Smeekens, Riksen-Walraven, & Van Bakel, 2008[175] | Free play, Structured discourse task | Kersten-Alvarez 2010[49] van Doesum 2008[48] |
| Maternal Sensitivity and Responsivity Scales | MSRS | Maternal attunement to infant behaviors during interaction | Cenciotti, Tronick, & Reck, 2004[176] | Still Face | Sahlen-Helmer 2020[5] Meijssen 2010[68] |

| Scale | Acronym | Construct measured | Main reference(s) | Paradigm(s) | Studies using this assessment |
|---|---|---|---|---|---|
| Mini-Maternal Behavior Q-Sort for Video Coding | Mini-MBQS-V | Maternal sensitivity | Moran 2009[177] | Free-play, Diaper change, Book sharing | Ramsauer 2019[178] |
| Mother-Infant Interaction Rating Scales | -- | Quality of parent-child interaction | Cowan & Cowan, 1992[179]; Cowan & Cowan 1992[180] | Feeding interaction | Schuler 2000[76] Schuler 2002[181] |
| Mutually Responsive Orientation | MRO | Responsiveness of parent-child relationship defined as close, mutually binding, cooperative, and affectively positive interactions | Kochanska, 2002[182] | Free play | Herbers 2020[62] |
| National Institute of Child Health and Human Development Scales (Qualitative Scales of the Observational Record of the Caregiving Environment) | NICHD Scales | Maternal sensitivity and responsiveness | NICHD, 1999[183,184] | Play | Berlin 2018[86] Hoffenkamp 2015[32] Ravn 2011[104] Peronne, 2021[92] |
| Nursing Child Assessment Feeding Scale & Teaching Scales | NCAFS NCATS | Parental sensitivity and relational effectiveness | Sumner & Spietz, 1994[185] | Feeding, Teaching | Badr 2006[100] Chiu 2009[1] Glazebrook 2007[60] Horowitz 2013[64] Lavallée 2022[67] Nelson 2001[107] Oxford 2021[39] Tryphonopoulos 2020[46] White-Traut 2013[108] |
| Observation Checklist on Mother-Infant Interaction | OMII | Infant and maternal interactive behaviors, secure/approach and insecure/ avoidant measures as well as maternal warmth and negative affect, | Ukeje, Bendersky, and Lewis (2001)[186] | Playtime and temporary separation | Porter 2015[22] |

| Scale | Acronym | Construct measured | Main reference(s) | Paradigm(s) | Studies using this assessment |
|---|---|---|---|---|---|
| | | and infant positive affect and anger | | | |
| Observation of Mother-Child Interaction | OMCI | Mother-child interaction for responsive caregiving skills | Landry et al., 2006[19] | Dyadic interactions | Betancourt 2020[54] |
| Parent-Caregiver Involvement Scale | P/CIS | Quality of mother-child interaction, maternal sensitivity, and maternal intrusiveness | Knoche et al.2012 [65] | Free play | Cooper 2009[57] |
| Parent Child Early Relational Assessment | ERA | Quality of mother-child relationships | Clark, 2015[187] | Free play | Holt 2021[110] |
| Parent-Child Interaction Rating Scales – Infant Adaptation | PCIRS-IA | Parental sensitivity and quality of parent-child interaction | Clark, 1999[188] | Free play | Canfield 2020[42]<br>Roby 2021[41] |
| Pediatric Infant Parent Exam | PIPE | Quality of parent-infant interactions | Fiese, Poehlmann, Irwin, Gordon, & Curry-Bleggi, 2001[189] | Dyadic interactions while playing a game | Ericksen 2018[109] |
| Preterm Mother-Infant Interaction Scale | PREMIIS | Mother-infant relationship for sensitive and responsive caregiving | Newnham, Milgrom, and Skouteris 2009[103] | Undressing, bathing, and dressing | Milgrom 2013[101] |
| Sensitivity and Cooperation | -- | Maternal sensitivity and responsiveness | Ainsworth et al., 1974[149] Ainsworth, Bell, and Slayton 1971[148] | Free play | Juffer 2005[34] |
| Still Face Procedure | SFP | Infant response to social challenge | Ainsworth 1978[190] | Still face | Meijssen 2010[68]<br>Valades 2021[79] |
| StimQ2 | -- | Caregiver cognitive stimulation | Dreyer, Mendelsohn, & Tamis-LeMonda, 1996[191] | Structured interview | Cates 2018[30]<br>Mendelsohn 2007[38]<br>Mendelsohn 2011[192]<br>Roby 2021[41] |
| Strange Situation Procedure | SSP | Infant Attachment | Ainsworth et al., 1978[190] | Strange situation | Berlin 2017[85]<br>Bernard 2012[87]<br>Brisch 2003[29]<br>Cooper 2009[57] |

| Scale | Acronym | Construct measured | Main reference(s) | Paradigm(s) | Studies using this assessment |
|---|---|---|---|---|---|
| | | | | | Klein Velderman 2006[36]<br>Sadler 2013[93]<br>Santelcies 2011[96]<br>Tereno 2017[45] |
| Synchrony Scale | -- | Mother-infant interaction for reciprocity and synchrony | Milgrom & Meitz, 1988[193] | Free play | Newnham 2009[103] |

#### Adult Play Scale

The Adult Play Scale is used to rate parent-infant interactions.[147]

**Paradigms.** Mothers and infants engage in a 20-minute videotaped free-play session. This provides an opportunity to assess the appropriateness and extent of parental physical and emotional engagement with the child.

**Populations.** Not specified.

**Scoring.** Interactions for which mutual eye gaze was sustained for more than a moment or in which parents were actively involved with the child are rated according to clearly defined criteria. Child behavior or dyadic engagement is not scored. Caregiver behavior was rated: (0)Ignores, if the caregiver ignored the child; (1) Routine, if the caregiver touches the child for routine care, but made no verbal responses; (2) Minimal, if the adult touched the child for discipline or in response to requests for help; (3) Simple, if the caregiver verbally responds to the child, but did not elaborate; (4) Elaborated, if the caregiver made positive physical gestures or stayed in close proximity to the infant; or (5) Intense, if the caregiver hugged the child, engaged in conversation with the child, repeated child statements, or played with the child[147].

The scale point for each interaction is multiplied by the duration of exchange, summed, and then divided by the total session time (20 minutes) to determine the mean level of play.

**Psychometric properties.** Test-retest reliability (0.85) was determined by assessing four children a week after the initial assessment.[147] There was a significant correlation between scores on the Adult Play Scale and overall scores for quality of care in 55 family daycare homes, $r = 0.52$, $p < 0.01$.[147]

**Coding Manual/Training.** Not specified.

#### Ainsworth's Maternal Sensitivity Scales

The Ainsworth's Maternal Sensitivity Scales are used to assess maternal sensitivity towards the infant[148,149,190].

**Paradigms**. Ainsworth's Maternal Sensitivity Scales have been used to code maternal behavior in free-play paradigms. For example, in free-play paradigms, mothers could be instructed to play with their infants as they normally would for ten minutes with a standardized collection of toys (i.e. squeaking duck, mirror, rattles) provided by the researchers (as done in Klein-Velderman, 2006). Behavior during the bathing routine has also been coded using the same scales[4].

**Populations**. This scale has been validated for use in infants 3-24 months of age[5]

**Scoring**. Maternal behavior is assessed on the following subscales: sensitivity, interference-cooperation, availability, acceptance-rejection, and positive and negative regard. Infant behavior is assessed on responsivity, involvement, and negative mood. Negative mood, positive regard, and negative regard are scored on a 7-point scale while the remaining subscales range from 1-9, with lower scores corresponding to less secure behaviors (i.e. 1 corresponds to highly insensitive or highly interfering mothers). The infant responsivity and involvement scales are averaged to determine a composite score. Maternal sensitivity and cooperation scales were also averaged to create a composite score[4,194]. For double-scored videos (n = 52%), published research uses a final score that is determined by averaging the individual scores of the two coders. If the scores differed by one point, the score that deviated more from the mean was chosen. If the scores differed by more than a point, a third coder rated the video, and the scores of the two coders with the most agreement were used to determine the final score[4].

**Psychometric properties**. The scales have been tested for validity and reliability. Infant responsivity and involvement were highly correlated (r = 0.90) as well as maternal sensitivity and cooperation (r = 0.93). T1 pre-test scores strongly correlated with T2 post-test scores (r = 0.45, p < 0.001; see Klein-Velderman, 2006).

**Coding Manual/Training.** Not specified.

#### Attachment Q-Sort (AQS)

The AQS is used to measure infants' attachment behavior in a home setting (AQS[150,195]).

**Paradigms.** As described in Klein-Velderman and colleagues (2006): Trained observers watch a 1.5 to 2.5-hour videotaped observation of the mother and infant interacting naturally in their home setting. The first segment of the visit is approximately two hours of unstructured interactions, including lunch and other daily activities. This is followed by the child playing a hammering game, the dyad playing with clay for 15 minutes, and dyadic play without toys for five minutes.

**Populations.** The AQS has been validated for use in children aged 12-48 months[48].

**Scoring.** The AQS consists of 90 descriptive statements of attachment behaviors. Statements are sorted into a nine-category, ten-card distribution from "most characteristic of the child" to "least characteristic of the child." Security scores were derived by correlating the Q-sort description to the criterion sort. Security scores range from –1.00 for an extremely insecure infant to +1.00 for an extremely secure infant.

**Psychometric properties.** The AQS has been shown to be reliable and has good discriminant, convergent, and predictive validity of attachment security in all domains except the avoidance scale (α = 0.15;[196].

**Coding Manual/Training.** A coding manual has been published by Waters & Deane (1985)[195].

#### Attachment Story Completion Task (ASCT)

The Attachment Story Completion Task is used to measure child attachment representations[151,152,197].

**Paradigms.** Children are given a mother and a child doll. They are instructed to complete five story stems that activate their attachment representations. Two of the story beginnings deal with emotionally charged and relationship-acknowledging interactions between the dyad: (1) the child gives the mother a handmade present and (2) the child says, "I'm sorry, Mom." The other three stories deal with distress or external conflict: (1) the child's bike is stolen by an unfamiliar child; (2) there is a monster in the child's bedroom; (3) the child is crying because they argued with another child at school[151]. This paradigm provides the opportunity to observe the extent to which the child views themselves in a secure relationship with the mother as well as maternal responsiveness to positive social signals from the child, when the child feels threatened, and to signals of distress from the child[197].

**Populations.** In addition to the task described above that is typically used among preschool-aged children, the ASCT has been adapted for use in 5-year-olds[198].

**Scoring.** Videotaped transcriptions of the story completions are rated on a 5-point security scale. Stories scored as a 4 or 5 were classified as secure if the child completed the story with minimal hesitation and displayed positive interactions with a responsive mother. Stories scored as a 1 or 2 were categorized as ambivalent/bizarre or avoidant, both of which were considered insecure categories. Ambivalent/bizarre stories had negative and hostile interactions between the child and mother mixed with brief, positive interactions. Avoidant stories had minimal child-mother interactions, or the child hesitated to complete the story. Stories with a score of 3 were not clearly secure or insecure and were categorized as secure/insecure. The scores for all the story completions were summed to achieve an overall attachment security score. Higher scores indicate a more secure attachment with the attachment figure.

**Psychometric properties.** The scale was also tested for reliability and consistency of the stories, yielding a Cronbach's alpha of 0.68 in a study by Kersten-Alvarez and colleagues (2010)[49].

**Coding Manual/Training.** For an unpublished coding manual, see Verschueren and Marcoen (1994)[151].

#### Atypical Maternal Behavior Instrument for Assessment and Classification (AMBIANCE)

The Atypical Maternal Behavior Instrument for Assessment and Classification is used to measure affective communication and behavior between caregivers and infants[153,199].

**Paradigms.** This scale is used to assess videotaped face-to-face interactions between the mother and infant.

**Populations.** The AMBIANCE scale was originally developed to code maternal behavior during the 12-month Strange Situation[199] and has been validated for use with 4-month-old infants and their mothers[200].

**Scoring.** The six dimensions of the scale include affective communication errors, role confusion, fearfulness/disorientation, intrusiveness/negativity, withdrawal, and controlling behavior. A frequency score for each of the dimensions is calculated by coders masked for any participant status characteristics. Overall disrupted communication is also scored on a 7-point scale: 1 (sensitive communication), 3 (mostly positive with some disrupted communication), 5 (repeated disruptive communication), or 7 (disruptive communication with little or no amelioration). Higher scores indicate greater levels of disrupted communication. Scores of 5 or above are considered "disrupted" while scores less than 5 are considered "not-disrupted" [153,201].

**Psychometric properties.** This assessment has been correlated with the Strange Situation as well as dyadic behavior in the home[199,202]. The AMBIANCE scale also demonstrates strong stability over time[203] and is predictive of child behavioral problems from toddlerhood to 20 years old[204-206].

**Coding Manual/Training.** Training information is provided online at: https://www.challiance.org/academics/research/family-studies-lab/ambiance-training

#### Avant Maternal Attachment Behaviors Scale

The Avant Maternal Attachment Behaviors Scale is used to assess emotional attachment between dyads[155].

**Paradigms.** In Shoghi and colleagues' (2018) intervention study, dyadic attachment was measured before the intervention started as well as on the first, third, and fifth days after the intervention commenced. Dyads engaged in naturalistic interactions for 15 minutes. This session was observed directly by a member of the research team who was masked to intervention group assignment. Attachment assessment was performed an hour after the intervention session (i.e., massage) but before breastfeeding.

**Populations.** Used with mothers and newborn infants in Shoghi and colleagues' (2018) study[24] and also published in research with infants conducted by the scale developer[155,207].

**Scoring.** The Avant Scales[155] are comprised of 14 observable behaviors that assess emotional behaviors (kissing, looking, cuddling, talking, smiling, etc.), proximity behaviors (hugging with no contact with the mother's body, hugging with little contact to the mother's body, hugging by wrapping arms around the neonate), and caring behaviors (changing the diaper or clothes, burping the neonate, organizing the neonate's clothes). These attachment behaviors are scored in 15 one-minute sections: the researcher observes the mother's behavior for the first 30 seconds and records them in the latter 30 seconds. If a behavior was observed, it received a score of 1; otherwise, a zero was recorded. The maximum score for each behavior is 15, and the maximum overall score is 210. Higher scores correlate with better dyadic attachment.

**Psychometric properties.** No specified.

**Coding Manual/Training.** Not specified.

#### Barrier Task

This measure was developed to assess infants' reactions to non-social challenges[156].

**Paradigms.** The infant is presented with a novel toy and allowed to play with it for 30 seconds. The toy is removed from the infant's possession and placed behind a transparent barrier in their line of sight for one minute. During this paradigm, the mother is near the infant, but she is instructed to have a still, neutral face. This task presents the opportunity to determine infants' capacity to face challenges in a regulated way and to assess infants' use of communication in distressing circumstances[79].

**Populations.** Not specified.

**Scoring.** A coding scheme developed by Bozicevic and colleagues (2016)[208] was used to code videos in Valades and colleagues' (2021)[79] intervention study. Interactions were coded for the presence or absence of various regulated and dysregulated behaviors on a 1-s time basis. Regulated behaviors include (a) Social: either through gaze or referencing (looking toward mother or researcher and/or pointing or vocalizing for the toy); (b) Non-social goal-directed: gazing or reaching for the toy; and (c) Non-social not goal-directed: distraction or play/exploration. Dysregulated behaviors include escape/aversion and tantrums.

**Psychometric properties.** Not specified.

**Coding Manual/Training.** Not specified.

#### Child Adult Relationship Evaluation (CARE) Index

The CARE index is used to assess the quality and patterns of parent-infant interaction in regard to affect and cognition, and it is a robust indicator for future attachment behaviors (CARE-Index; Crittenden, 1979–2004[157]; ICI; Crittenden,1979–2010[158]).

**Paradigms.** Dyads are typically instructed to engage in free play[27,47] or semi-structured play[72] for 3-5 minutes. Mothers have the option to use toys, either their own or those provided by the experimenters[39].

**Populations.** This measure has been validated for use in infants from birth[28,46] to 15 months of age[46,47] and has also been used at 30 months[28].

**Scoring.** The CARE Index has several subscales to assess both mother and infant relational measures. Mothers are rated on sensitivity, control, and unresponsiveness. Infants are rated on their levels of cooperativity, compulsivity, difficultness, and passivity. Behaviors of interest include facial and verbal expression, positional and body contact, affection, turn-taking, control, and choice of activity[46]. Each scale is scored from 0-7 points[28] or 0-14 points[27,39,46,47,72,78]. Higher scores indicate more of that behavior (i.e., 0 being highly insensitive and 14 being highly sensitive); lower scores on the infant scales reflect more positive interaction. Maternal sensitivity scores ranging from 0–4 indicate the need for child protection, psychotherapy, or other forms of intervention. Scores from 5 to 6 also indicate the need for intervention, but no outward hostility or lack of empathy towards the child is exhibited. Scores of 7–8 indicate adequate play with brief periods of dyssynchrony. Scores of 9–14 reflect satisfactory or excellent parental sensitivity. Infant cooperativity is categorized as such: 0-4 (difficult/passive), 5-9 (mixed cooperative), or 9-14 (cooperative). A binary variable can be created to categorize scores greater than 9 as satisfactory maternal sensitivity or infant cooperativeness and scores less than 9 as maternal insensitivity or infant uncooperativeness[72]. The CARE index can also yield a score for dyadic synchrony (the global quality of the interaction). Lower scores on the dyadic synchrony scale reflect lower dyadic synchrony.

**Psychometric properties.** The CARE-Index can be used in families across different socioeconomic backgrounds and has been well validated for assessing parent-infant interaction, attachment quality, child interactive behavior, childrearing problems like abuse or neglect, and the effectiveness of interventions[72,209,210]. It is strongly correlated with the Strange Situation, especially at infants' one year[47]. Previous studies have also shown the CARE-Index to be highly stable[211]. Maternal and infant scales are typically correlated with each other (i.e., maternal sensitivity is related to infant cooperativity).

**Coding Manual/Training.** P. M. Crittenden, 1979–2004, CARE-Index: Coding Manual, unpublished. The author of the coding scheme (Crittenden) oversaw the coders' work in Oxford and colleagues' investigation using this coding system (2021)[39].

#### Coding Interactive Behavior (CIB)

The Coding Interactive Behavior (CIB) system measures the quality of mother-child interactions as well as maternal and infant behaviors (Feldman, 1998[212]).

**Paradigms.** Mothers are typically instructed to play with their infant as they normally would in a 5[74] to 10-minute[97,98] free-play episode. This provides an opportunity to assess maternal and child behavior in a naturalistic setting.

**Populations.** The CIB system has been validated for use in both normative and high-risk populations[74].

**Scoring.** The CIB is comprised of 43[98] to 45[97] discrete items, rated on a 5-point scale describing the frequency and intensity of maternal behaviors, child behaviors, and overall quality of the dyadic interaction. Higher scores indicate higher frequency or intensity. Videos were coded in 10-second intervals for categories of interest, and each category is comprised of mutually exclusive codes. CIB behavioral categories can be aggregated into various subscales or composites using factor analysis including, but not limited to, (a) maternal hostility, dyadic constriction, supportive maternal presence, and dyadic reciprocity (used in Steele, 2019); (b) dyadic attunement, parental positive engagement, and child involvement (used in Sleed, 2016; Fonagy, 2018; Sleed, Baradon & Fonagy, 2013); (c) maternal affiliative behavior, maternal stimulatory touch, and infant alert (used in Feldman, 2007); or (d) sensitivity, intrusiveness, limit setting, involvement, withdrawal, compliance, dyadic reciprocity, and dyadic negative states (used in Pontoppidan, 2022).

**Psychometric properties.** This system has been well-validated, demonstrating concurrent and discriminative validity, and is sensitive to change and various risk factors such as maternal substance use, delivery pain, infant prematurity, and massage therapy for preterm infants[213-217]. Internal consistency among previously published studies are as follows: Dyadic Attunement ($\alpha = 0.40$), Parent Positive Engagement ($\alpha = 0.833$), and Child Involvement ($\alpha = 0.857$[97]); total score ($\alpha = 0.92$), sensitivity ($\alpha = 0.92$), intrusiveness ($\alpha = 0.53$), involvement ($\alpha = 0.71$), withdrawal ($\alpha = 0.26$), reciprocity ($\alpha = 0.84$), and negative states ($\alpha = 0.41$).[74]

**Coding Manual/Training.** Mother-Newborn Coding System of the Coding Interaction Behavior Manual (CIB, Feldman, 1998).[212]

#### Coding system by Piccinini, Alvarenga and Frizzo (2007)

This coding system is used to assess maternal sensitivity during maternal-infant interactions[160].

**Paradigms.** Dyads typically engage in a 10-minute free play episode during which the mother was asked to play with the infant with novel toys provided by the experimenter (a rattle, rubber puppy, and small rubber ball). In an intervention study conducted by Alvarenga and colleagues (2019), two different toys (a turtle-shaped toy with geometric puzzle pieces and a medium ball) were used to better match the infants' current developmental stage. This enables researchers to assess verbal and non-verbal maternal behaviors.

**Populations.** Not specified. In Alvarenga and colleagues (2019) study, infants were assessed at 3 and 11 months.

**Scoring.** The initial 5-minutes of the free-play session is usually coded at intervals of 12 seconds. In Alvarenga (2019), coding procedures included (a) infant behavior: smiles, emits sounds (stammers or attempts to vocalize), or cries; (b) maternal responses to infant behaviors: interprets, speaks, smiles, holds, touches/stimulates, or acts intrusively. The latter included maternal non-sensitive behaviors. All behaviors were coded as mutually exclusive, except for pacifier sucking, sneezing, coughing, and yawning. Infant behaviors are coded for the first 6 seconds of the 12-second interval while maternal behaviors were coded for the full 12-second interval.

**Psychometric properties.** Not specified

**Coding Manual/Training.** More details about the coding system can be found in Piccinini, Alvarenga, and Frizzo (2007).[160]

#### Dyadic Mutuality Code (DMC)

The Dyadic Mutuality Code measures mutuality, responsivity, and synchrony in dyadic interactions[218,219].

**Paradigms.** The mother is instructed to have the infant in her lap or in an infant seat so that the pair can engage in face-to-face interaction. The pair engage in normal play for 5 minutes without a toy or pacifier. Sessions are either observed live or video-recorded for later scoring.

**Populations.** This scale has been validated for use in healthy and preterm infants as well as high-risk infants[220-222].

**Scoring.** The DMC consists of six scales: mutual attention, positive affect, mutual turn-taking, maternal pauses, infant clarity of cues, and maternal sensitivity[218]. Items are scored as 1 (none or very brief) or 2 (more than half the time) and are summed to yield a total score (ranging from 6-12). Scores are categorized as low (6-8), moderate (9-10), or high responsiveness (11-12[63]).

**Psychometric properties.** This scale shows evidence of concurrent and construct validity[219]. Cronbach's alpha was 0.66, 0.63, and 0.70 across evaluation windows in a study done by Horowitz and colleagues (2001)[63].

**Coding Manual/Training.** Not specified.

#### Emotional Availability Scales (EAS) and Emotional Attachment Zones Evaluation (EA-Z)

The Emotional Availability Scales and Emotional Attachment Zones Evaluation are used to measure the quality of dyadic interactions and the overall dyadic relationship, with a particular focus on attachment-related behaviors[223-225].

**Paradigms.** Typically, dyads are videotaped for 15[75,91] to 20[36,75] or 30-minute[33] interactions in which the mothers are instructed to play with their child as they normally would, with or without toys. If the assessment is given prenatally, mothers are instructed to perform playful activities with the fetus for the duration of the session such as playing an instrument to the fetus[226].

**Populations.** The EAS/EA-Z has been validated for use prenatally or in children from birth up to 4 years[223,225].

**Scoring.** The EAS consists of six subscales: (a) adult (sensitivity, structuring, non-intrusiveness, and non-hostility) and (b) child (responsivity and involvement). Higher scores are more optimal for all the subscales. Scales can be assessed on a 3-, 5-, 7-, or 9-point system. A Likert 7-point scale (1-low, 7-high) was used for all subscales in most studies research reviewed that applied this coding system[31,75,91]. Van Doesum and colleagues (2008[48]) used the following scoring: parental sensitivity (9-point), structuring, non-intrusiveness, and non-hostility (5-point), and child responsivity and involvement (7-point). EA-composite scores for the caretaker (EA-P) and the infant (EA-C) are calculated by summing the corresponding scores. An EAS total score can be calculated by summing individual subscale scores[90,91]. A binary variable can be created to categorize the proportion of dyads rated as sensitive or responsive, which was defined by EA scores of 5.5 (low end of neutral-moderate sensitivity or responsivity) to 7 (high/optimal sensitivity or responsivity)[31].

Emotional availability can be assessed prenatally by combining the sensitivity and non-hostility scales[226,227]. The EA-Z is scored on a 100-point scale that is divided into four categories: (a) emotionally available, (b) complicated, (c) detached, and (d) problematic/disturbed (Wurster et al., 2019). These categories correlate with the four attachment styles: secure, insecure-anxious, insecure-avoidant, and insecure-disorganized, respectively[31,224,228]. Higher scores indicate better emotional availability.

**Psychometric properties.** The EAS and EA-Z have been well-validated and strongly correlate with assessments of attachment/infant attachment security[229,230]. Cronbach's alpha was 0.97 across three different time points across infancy in a prior publication using this measure[33].

**Coding Manual/Training**. In prior studies that use these scales, raters were trained by the scale developer, Z. Biringen, and certified for reliability (e.g., as done in Georg, 2021[91]; Hoivik, 2015[33]).

#### Eye gaze

This is used to assess the quality of mother-infant interactions[161].

**Paradigms.** Dyads are seated opposite each other, and mothers were told to play with their babies as they normally would, without toys, for a 10-minute episode. Synchronized cameras mounted on opposite walls allowed researchers to achieve a split-screen view of mothers and infants during the interaction.

**Populations.** In one study reviewed, this paradigm was used among infants at 4 months old[51].

**Scoring.** Coders score the first 2.5 minutes of the 10-minute interaction episode on a 1-s time basis. For multiple behaviors occurring in one second, the behavior occurring during the latter half is recorded. Behaviors of interest included eye gaze (on/off partner's face), infant vocal affect (high positive, neutral/positive, none, fuss/whimper, angry-protest, or cry), mother touch (affectionate, static, playful, none, caregiver jiggle, infant-directed oral touch, object-mediated, centripetal, rough, or high intensity-intrusive). For mother touch, type of touch, location, and intensity are all accounted for. Mother-infant modality pairings are developed to analyze self- and interactive contingency: (1) infant gaze-mother gaze; (2) infant gaze-mother touch; (3) infant vocal affect-mother touch; and (4) infant vocal affect-mother gaze[51].

**Psychometric properties.** Not specified.

**Coding Manual/Training.** Not specified.

#### Global Rating Scale (GRS)

The Global Rating Scale is used to assess the quality of dyadic interactions[162].

**Paradigms.** Reference the reunion phase of the Still Face paradigm.

**Populations.** Not specified.

**Scoring.** Maternal behavior is rated on four subscales: sensitivity, intrusiveness, remoteness, and depression. Infant behaviors are rated on three continuum subscales: happy vs. distressed, fretful vs. non-fretful, and attentive vs. avoidant. The overall quality of the interaction is also scored. Individual items on the maternal subscales behaviors were coded on a 5-point scoring system ranging from less optimal to more optimal. These are then summed and averaged to calculate the subscale score, ranging from 1 to 5. Higher scores reflect more positive behavior. For infant behavior, the happy-distressed scale is coded by determining the amount of time the infant engaged in happy (smiling or positive vocalizations), neutral (neutral affect and no signs of distress), or distressed behaviors (crying, shouting, withdrawing, frowning, etc.). The fretful-non-fretful scale is coded by counting the number of seconds infants engaged in crying, shouting, pushing the caregiver, or arching their back. The attentive-avoidant scale is coded by determining the amount of time the infant was attending to the caregiver's face.

Raw scores for the happy-distressed scale were calculated by adding the amount of time the infant was happy and half of the neutral time together. The resulting scores could range from 0 (100% distressed) to 1 (100% happy). For example, if the infant was happy 30% of the time and neutral 50% of the time, the raw score would be 0.30 + (0.50/2) = 0.55. Raw scores can be further categorized into quintiles: 1 (<0.20), 2 (0.21-0.40), 3 (0.41-0.60), 4 (0.61-0.80), and 5 (0.81-1.00). Infants with GRS scores of 3 were assigned secondary ratings (3N, 3(1), 3(2), or 3(3)) to differentiate between infants who were neutral most of the time versus infants who had a broad range of emotions and scored a 3. If the infant is neutral more than 74% of the time, they were assigned 3N. If the infant is distressed most of the time, they were assigned 3(1) or 3(2). If most of the distressing noises (>66%) occurred in the first part of the paradigm, they were assigned 3(1). If most occurred in the second part of the paradigm, they were assigned

3(2). Infants with a happy/distressed score of 3 that don't fall into the above categories were assigned 3(3). The fretful-non-fretful and attentive-avoidant scales are also categorized into quintiles on 5-point scales[7,8].

**Psychometric properties.** This scale has been validated cross-culturally, for use in both low- and high-risk groups, including among mothers with elevated depression symptoms[231]. The infant subscales have been validated and correlate with attachment categories[232].

**Coding Manual/Training.** Not specified

#### Holditch-Davis Coding System

The Holditch-Davis Coding System is used to assess the quality of the dyadic relationship and dyadic interactions[163,164].

**Paradigms.** Mother-infant interactions are typically assessed through a 45-minute videotaped session, at 2 and 6 months corrected age, when the infant was alert and due for a feeding. Mothers are instructed to care for their children as they normally would. This setup provided the opportunity to record naturalistic mother and child behaviors.

**Populations.** This scale was used in a population of pre-term infants and their mothers[106].

**Scoring.** Maternal and infant behaviors were coded in 10-second intervals; if a behavior occurred repeatedly in that interval, the behavior was only counted once. Maternal behaviors of interest included touch, talk, interaction, involvement with the child, positive affect, teaching, mother touch, and playing with the child. Infant behaviors of interest included expression of positive or negative affect, locomotion, gesturing, vocalizations, and independent play with objects. Coded behaviors were recorded as a percentage of the total videotape. Child activity level (asleep, sedentary, moderate activity, or very active) was also noted.

**Psychometric properties.** Not specified.

**Coding Manual/Training.** Not specified.

#### The Home Observation for Measurement of the Environment (HOME) Inventory

The HOME Inventory is used to assess the appropriateness of the home environment (i.e., quality of support and stimulation) for promoting infant development[165].

**Paradigms.** Assessments are made from videotaped or live observations of the home environment and parenting behaviors as well as interview questions with the caregiver.

**Populations.** This scale has been validated for use in children aged 0-3 years (Caldwell & Bradley, 1980).

**Scoring.** The HOME Inventory is a 43-item[54] or 45-item[83,106] instrument. Scores can be broken down into six dimensions: emotional and verbal responsivity, avoidance of restriction and punishment, organization of the physical/temporal environment, provision of appropriate play materials, parental involvement with the child, and opportunities for variety in daily stimulation. The responsivity subscale and maternal involvement subscales were of particular interest in research reviewed[60,106].

Two maternal dimensions (maternal positive involvement and developmental stimulation) can be generated from 8 maternal behaviors and the parental involvement subscale. Behaviors comprised of positive mood, touch, hold, interaction, uninvolved with child (reversed), play with the child, mother talking, and teaching. Similarly, three child dimensions (child social behaviors, developmental maturity, and child irritability) can be generated from 7 child behaviors. Behaviors consisted of expression of positive affect, gesturing, vocalizations, independent play with objects, locomoting, negative mood, and fuss as a percent of time with the mother[163,233]. Dimension scores were calculated by converting each variable to Z-scores and averaging them.

**Psychometric properties.** The HOME inventory is strongly correlated with child cognitive development[106], especially at 3 years[100], as well as maternal and child behaviors[234-236]. The assessment has been translated into Brazilian Portuguese[83]. Cronbach's alpha for the HOME total was 0.84 at 2 months and 0.83 at an assessment conducted at 6 months[106] and 0.76 in another sample[54] ; for the maternal involvement subscale, alpha was 0.70 in a prior study[106]. Internal consistency for the derived maternal and child dimensions were 0.78 (maternal positive involvement), 0.69 (developmental stimulation), 0.68 (child social behaviors), 0.58 (developmental maturity), and 0.91 (child irritability) as reported in Holditch-Davis and colleagues (2014)[106].

**Coding Manual/Training.** Not specified.

#### Index of Parental Behavior (IPB)

The Index of Parental Behavior (IPB) is used to assess the quality of dyadic interactions[166].

**Paradigms.** Dyads are typically observed for 30 minutes in the NICU which provided the opportunity to assess parent-infant interactions.

**Populations.** Infants in the NICU.

**Scoring.** The IPB consists of 15 statements measuring parental behaviors (i.e., parent seeks to interact face-to-face with infant) related to the infant. Parental behaviors are rated on a dichotomous scale (1-behavior present or 0-behavior not present). Higher scores indicate more sensitive maternal-infant interactions. Two subscales were derived through factor analysis: positive interaction with the infant in a quiet, alert state (7 items) and altering environment and interaction with a stressed infant (8 items; see Melnyk, 2006[71]).

**Psychometric properties.** Eight maternal-child experts and a cultural competence expert established content validity for the IPB (Melnyk, 2006).

**Coding Manual/Training.** Not specified.

#### Infant and Caregiver Engagement Phases (ICEP)

The Infant and Caregiver Engagement Phases (ICEP) measures infant and caregiver affect and engagement as well as the quality of the engagement[167].

**Paradigms.** The Still Face Procedure is typically used. This paradigm is usually conducted at the hospital, clinic, or in the home, with all distractors (i.e., television, music, phones) removed. The paradigm is typically videotaped using two cameras, one aimed at the infant, and one aimed at the mother.

**Scoring.** The ICEP assesses several different infant and caregiver behavioral phases. Infant behavioral phases include negative engagement (fussiness, protest, crying, negative facial expressions), object/environment engagement, social monitoring or mother-focused behavior, social positive engagement (positive facial expressions, smiles, or coos), sleep, oral self-comforting, self-clasp (infant's hands are touching), distancing, and infant autonomic stress indicators (hiccupping or spitting up).

Caregiver behaviors include negative engagement (stern, sad, angry, etc.), intrusiveness, withdrawn, non-infant-focused engagement, social monitoring without speech or with neutral speech, social monitoring with positive speech, social positive engagement, exaggerated positive engagement (laughter, play, surprise, etc.), rough touches, and procedural violations. All infant and maternal behaviors are mutually exclusive. Infant and maternal behavior were coded on separate runs. The duration of each behavior was recorded and converted to a percentage for each episode of the Still Face paradigm (normal play, still-face, reunion).

**Psychometric properties.** Not specified.

**Coding Manual/Training:** See Weinberg & Tronick, (1999)[167]

#### Landry Parent-Child Interaction Scales

The Landry Parent-Child Interaction Scales are used to assess naturalistic dyadic interactions between mother and infant[168,169].

**Paradigms.** Mothers are instructed to engage with their infants as they normally would during a 30-minute videotaped session. Session breakdown are typically either 30 minutes of free-play[16], 20 minutes of free-play and 10 minutes of a book read activity[17].

**Populations.** Not specified, used mostly with parents and their infants in the reviewed literature.

**Scoring.** The Landry scales are scored on a 5- to 7-point Likert scale. Maternal behaviors of interest include positive affect, warmth, flexible responding, positive verbal content, intrusive behavior, verbal and affective negativity, and book-sharing techniques. Infant behaviors of interest include attention/arousal, warmth-seeking, positive behavior, behavioral regulation, expressive and receptive language, and negative affect.

**Psychometric properties.** The Landry Scales have been widely used and adequate reliability and predictive validity have been established, specifically in relation to child socio-emotional outcomes[237,238]. Higher scores are also correlated with more positive child behavioral and developmental outcomes[17]. In one study, factor analysis of the warm/positive and flexible responding observational codes for parents yielded a single parenting factor (factor loadings of > .3), explaining 44-47% of variance in coded behavior ratings[17].

**Coding Manual/Training.** Not specified.

#### The Looking, Touching, Talking, Smiling (LoTTS) Parent-Infant Interaction Coding Scale

The Looking, Touching, Talking, Smiling Parent-Infant Interaction Scale is used to measure the frequency of interaction behaviors between dyads[170].

**Paradigms.** Mothers are instructed to play with their infants as they normally would for a four-minute videotaped session.

**Populations.** Not specified.

**Scoring.** Behaviors of interest consisted of touching, looking, talking, smiling[170], and singing[12]; these behaviors are recorded every 20 seconds. Maternal responsiveness and warmth were globally rated on a 5-point scale, with higher scores representing more positive interactive behaviors.

**Psychometric properties.** Not specified.

**Coding Manual/Training.** Not specified.

#### Maternal Behavioral Q-Set (MBQ)

This assessment measures maternal sensitivity towards the infant[171].

**Paradigms.** In the reviewed literature, dyads are typically assessed during a 1–2-hour naturalistic home observation in which mothers were told to interact with their children as they normally would.

**Populations.** Not specified.

**Scoring.** The Q-set is comprised of 90 items related to maternal behavior including interaction style, sensitivity to infant state, feeding interactions, etc. Experimenters organize these items into a rectangular distribution with 10 items in each of nine piles, ranging from least characteristic of the mother's behavior (pile 1) to most characteristic (pile 9). Items that are not directly observed during the session (i.e., diaper change) must go in the middle piles[171,239].

**Psychometric properties.** The MBQ has well-established criterion and construct validity as well as test-retest reliability[240].

**Coding Manual/Training.** A coding manual for the MBQ can be found in Appendix B of Pederson & Moran (1995).[171]

#### Maternal Caregiving Behaviors

The Maternal Caregiving Behaviors scale is used to assess observations of feeding, holding, and other maternal caregiving behaviors [172,173].

**Paradigms.** Dyads are typically scheduled for a videotaped caregiving observation when the infants reached 36 weeks gestational age. Recordings have taken place at the infant crib in the NICU. Mothers were instructed to hold their child in a chair adjacent to the crib. This was followed by bottle feeding. The entire session lasts 15 minutes, and mothers were told to soothe their babies as they normally would. The tripod is set up in the curtained area by a research assistant, but the dyad was alone for the videotaped portion that is subsequently coded.

**Populations.** This scale has been used in previous research with full-term 9-month-olds and full-term neonates during feeding, bathing, and changing episodes[172,173].

**Scoring.** The MCB holding score is determined using the acceptance-rejection, consideration-intrusiveness, psychological availability, quality of physical and vocal contact, and expressed joy-delight subscales of Ainsworth's original scales. An MCB feeding score was derived using the sensitivity to the infant's nursing pace, quality of feeding transitions, psychological availability, quality of physical and vocal contact, and amount of visual contact subscales. All scales were scored on a 9-point system, with 9 being the most sensitive. Higher scores indicate highly sensitive and attentive feeding or holding sessions with more positive affect, skin-to-skin contact, face-to-face gaze, etc. Overall MCB composite scores were derived by averaging the feeding and holding scores.

**Psychometric properties.** In investigations conducted by Hane and Fox (2006)[172] and Hane and Philbrook (2012)[173], MCB holding and feeding scores have been shown to be strongly correlated, r (63) = 0.57, $p < 0.001$. Composite MCB scores were normally distributed (mean = 4.74, SD = 0.91). The scores and normality of the distribution resemble those from previous research with neonates and 9-month-olds.

**Coding Manual/Training.** Coders were trained using the Maternal Caregiving Coding Manual for use in High-Risk Population of Infants in the NICU developed by Amie Hane, PhD[50].

#### Maternal Interactive Behaviors Likert Scales

The Maternal Interactive Behaviors Likert Scales assesses the quality of maternal interactive behaviors[174,175].

**Paradigms.** Two episodes of dyadic interactions were videotaped during a home visit. The first segment was 10-15 minutes of free play. The second segment involved a structured discourse task in which dyads were given an image of a child displaying a particular emotion (i.e., happy, anxious, sad, or angry). The child was then asked to label how the child in the picture was feeling and was subsequently asked, "Have you ever felt like this?" and, if yes: "What made you feel like this?"

**Scoring.** The discourse task assessed maternal behavior using five 7-point scales including supportive presence, respect for child autonomy, structure and limit setting, quality of instruction, and hostility (Erickson, Sroufe, & Egeland, 1985). The free play segment was scored using a different 7-point scale that measures scaffolding (i.e., the provision of developmentally appropriate guidance[175]). The two composite scores were summed to determine the overall score for the quality of maternal interactive behavior.

**Psychometric properties.** These scales have been well-validated[241]. Composite scores for the discourse task and free-play task correlated with an r = 0.52, p < 0.001 in a study conducted by Kersten-Alvarez and colleagues (2010)[49].

#### Maternal Sensitivity and Responsivity Scales (MSRS)

The Maternal Sensitivity and Responsivity Scales measures maternal attunement to the infant during interaction behaviors[176].

**Paradigms.** Reference the Still Face Paradigm. This scale is used to code behavior during phase 1 (face-to-face normal interaction) and 3 (reunion face-to-face interaction) of the Still-Face Paradigm.

**Populations.** Not specified.

**Scoring.** The MSRS has three subscales scored on a 5-point rating. The first scale measures sensitivity/responsivity, from 1 (mothers with ignoring behavior) to 5 (responsive behavior). Sensitive and responsive behavior includes detecting the infant's behavioral cues and appropriate reactions to them. The second scale assesses undercontrol/withdrawal, from 1 (an engaged response) to 5 (extremely withdrawn response). Undercontrolling behavior includes disengagement in the interactions as well as flat affect. The last subscale measures overcontrol/intrusiveness, from 1 (aware, but nonintrusive) to 5 (strongly overcontrolling). Intrusive behavior is defined as the mother privileging her own behavior and disregarding infant cues and behaviors[68].

**Psychometric properties.** There is limited information available about the validity of the MSRS[5].

**Coding Manual/Training.** See Cenciotti, Tronick, and Reck, 2004[176].

#### Mini-Maternal Behavior Q-Sort for Video Coding (Mini-MBQS-V)

The Mini-Maternal Behavior Q-Sort for Video Coding is used to measure maternal sensitivity[177].

**Paradigms**. Mothers and their infants are recorded in free-play episodes, a diaper-changing session, and a book-sharing episode.

**Scoring.** This assessment is a short 25-item version of the Maternal Behavior Q-Set. Maternal behavior is described by matching the description of generic maternal behavior and observed maternal behavior into five categories (very like, like, neither, unlike, and very unlike mom). The sensitivity score is based on the correlation between the descriptive sort and a criterion sort of a characteristically sensitive mother. These scores range from least sensitive at -1.0 to most prototypically sensitive at 1.0.

**Psychometric properties.** Reliability was confirmed in a 10-minute play interaction in infants aged 10 months, and convergent validity was confirmed with the 90-item MBQS completed at infant age six months. This study yielded Cronbach's alpha of .95[178].

**Coding Manual/Training.** See Moran (2009)[177].

#### Mother-Infant Interaction Rating Scales

The Mother-Infant Interaction Rating Scales are used to assess maternal and infant behavior[179,180]

**Paradigms**. Scales can be used to assess either a play or a feeding paradigm.

**Populations**. These scales are not tied to specific age-related behavior, so they can be used on parents and children of any age (see Schuler, 2002[181]).

**Scoring.** There are two scales, one for parent behavior and one for infant behavior, with each item representing a global rating of the behavior during the interaction. Each item was scored on a 5-point scale ranging from one (very low) to five (very high).

**Psychometric properties**. Internal consistency was measured for both maternal behaviors and infant behaviors with alpha coefficients of .95 and .90 respectively in a study conducted by Schuler and colleagues (2000[76]).

**Coding Manual/Training:** Not provided.

#### Mutually Responsive Orientation (MRO)

The Mutually Responsive Orientation assessment is used to measure responsiveness within the parent-child relationship.

**Paradigms**. Mothers and their infants are recorded in free-play episodes.

**Populations**. There is support for extensive structural stability in the MRO in children ages seven to 15 months[242]. MRO has positive implications for multiple aspects of children's moral development, both concurrently and longitudinally. Those positive links hold across multiple ages, multiple assessment types, and diverse contexts. **Scoring.** Scores (1-5) show the degree of responsiveness within the relationship, namely, coordinated routines, communication, cooperation, and emotional ambiance. Higher scores indicate higher levels of responsiveness.

**Psychometric properties**. This scale was deemed to have good inter-rater reliability, test-retest reliability and sensitivity to developmental fluctuation[146]. See above for validity across populations.

**Coding Manual/Training:** Not provided.

#### National Institute of Child Health and Human Development Scales (NICHD)

The National Institute of Child Health and Human Development Scales (also called the Qualitative Scales of the Observational Record of the Caregiving Environment) is an observational tool used to measure maternal behavior and affect toward her infant.

**Paradigms**. Play interactions were videotaped and coded by raters who were blind to group status.

**Populations**. Minor adaptations were made to the original tool to accommodate preterm infants in Hoffenkamp and colleagues (2015[32]).

**Scoring.** This set of scales includes scoring for maternal sensitivity and responsiveness, maternal intrusiveness, and maternal positive regard. Maternal sensitivity/responsiveness represents the degree to which the mother showcases emotionally supportive behaviors that are appropriate and in sync with the infant's cues. Maternal intrusiveness is the degree to which the mother exhibited verbal or physical interference with the infant's needs, interests or behavior. Lastly, maternal positive regard represents the mother's ability to express warmth, enthusiasm, and praise to her infant. In studies by Berlin and colleagues (2018[86]) and Ravn and colleagues (2011[104]), a predetermined scale from 1-5 was utilized to measure the domains of maternal and infant behavior. This five-point scale indicates the degree to which the displayed behaviors match the behaviors specified in the coding manual based on both the quality and quantity of the observed interaction. Scales were adapted for use in specific samples in studies by Hoffenkamp and colleagues (2015[32]; preterm infants). Perrone and colleagues (2021) also stated that scales were adapted for use[92].

**Psychometric properties**. There has been strong evidence pointing to the validity of these scales from many studies that link maternal characteristics and child outcomes to various sociodemographic factors such as family cumulative risk[243].

**Coding manual/training.** "Qualitative Ratings for Parent-Child Interaction at 3–15 months of age[244,245]" is a modification of the coding system used by the National Institute of Child Health and Human Development (NICHD) Study of Early Child Care[183].

#### Nursing Child Assessment Feeding Scale (CAFS) & Teaching Scales (NCATS)

The Nursing Child Assessment Satellite Training Program (NCAST) Feeding and Teaching scales[185] was developed to assess the quality of interactional behaviors between infants and parents[246]. More specifically, the Nursing Child Assessment Feeding and Teaching Scales measure maternal-infant relational effectiveness and parental responsiveness to their infants[64].

**Paradigms**. The Nursing Child Assessment Satellite Training Program (NCAST) Feeding and Teaching scales either involves a feeding or teaching paradigm, respectively. In the teaching session, the mother is videotaped as she teaches her infant a task that she believes the infant will not know how to do[1]. First, the caregiver is shown a list of sensorimotor skills in increasing order of difficulty and is asked to select the first skill on the list that the child has not yet acquired[60]. Examples include following a toy with eye gaze, grasping a toy, or transferring an object from one hand to another[39,60]. This scale can be used to code feeding sessions, where mothers are videotaped while they feed their infants[67].

**Populations**. The feeding scale can be used with infants from birth to one year of age, while the teaching scale can be used with infants from birth to 3 years of age[1,60].

**Scoring.** Specific behaviors during the interaction are scored present or absent. Dyadic interactive disturbance is suggested by positive scores for less than 43 items (1–6 months), 46 items (9–12 months), 52 items (13–24 months), or 53 items (25–36 months)[64]. An overall summary score is then yielded[100]. Both the teaching and feeding scales consist of six subscales that each assesses mother-infant interaction, including: sensitivity to cues, response to infant distress, social-emotional growth fostering, cognitive growth fostering, clarity of cues, and responsiveness of the child to the caregiver. Each subscale lists a multitude of caregiver and infant behaviors that the evaluator observes during a routine feeding episode and a teaching episode. Higher scores indicate an interaction that is richer in positive affect, sensitivity, synchronicity, vocalization, and reciprocity[1].

**Psychometric properties**. Both the teaching and feeding scales have established validity and reliability. Internal consistency reliability was reported as .87 for the Overall Total Scale[185].

**Coding Manual/Training:** Not provided.

#### Observational Checklist on Mother-Infant Interaction (OMII)

The Observational Checklist on Mother-Infant Interaction (OMII) scale is used to measure mother-infant interactive behaviors[186].

**Paradigms**. The OMII utilizes a 5-minute free play period, of which the last three minutes are coded by trained researchers, as well as the first 2 minutes of the mother-infant reunion period following a planned 2-minute separation[22].

**Populations**. Not specified.

**Scoring.** Verbal and educational interaction, visual monitoring, warmth, negative affect, and the structuring of toys by the mothers were coded using a one-way mirror. Infant positive affect and anger were coded during the play phase, while attachment style can be coded during the reunion phase. The coding system scored for both frequency and duration of the interaction. Additionally, a scoring methodology was applied in which points were given for infant behaviors reflecting secure attachment and subtracted for avoidant, resistant, or insecure behaviors. Higher scores reflected more positive interactions[22,186].

**Psychometric properties**. Not specified.

**Coding Manual/Training:** Not provided.

#### Observation of Mother-Child Interaction (OMCI)

The Observation of Mother-Child Interaction (OMCI) is used to assess the mother-child interaction for responsive caregiving skills. The conceptual model for the OMCI was developed by Landry and colleagues (2006[19]).

**Paradigms**. Mothers and their children are videotaped in an interaction surrounding a picture book. The same picture book, compromised of colorful, familiar, and culturally acceptable pictures, was given to the participants by the lab for the duration of the episode[247].

**Populations**. The OMCI was developed as part of a battery of maternal and child assessments used in a cluster randomized controlled trial evaluating interventions on a cohort of mothers and their children aged 0-2[248,249].

**Scoring.** The OMCI contains thirteen items for maternal behaviors, six for child behaviors, and one for mutual enjoyment behaviors. Scores follow a yes or no binary format.[247]

**Psychometric properties**. A positive association was found between OMCI, Responsiveness and Involvement subscales of the HOME, parenting knowledge and practice, maternal depressive symptoms, child development, and child growth, suggesting that the scale shows good validity[247]. Inter-rater reliability was also high in the study conducted by Rasheed and colleagues (2015)[247].

**Coding manual/training.** See Rasheed and Yousafzai (2015)[247].

#### Parent-Caregiver Involvement Scale (P/CIS)

The Parent-Caregiver Involvement Scale (P/CIS) is used to assess the amount and quality of involvement between a caregiver and child[250].

**Paradigms**. Caregivers and their children can be videotaped in either free or structured play episodes[57].

**Populations**. The P/CIS is designed for use with children ages 0-3[250]. The P/CIS was designed to operate independently of socioeconomic status and can be used with children across the full range of abilities[251].

**Scoring.** This scale measures 11 parental behaviors: (a) physical involvement; (b) verbal involvement; (c) responsiveness of caregiver to child; (d) play interaction; (e) teaching behavior; (f)control of activities; (g) directives, demands; (h) relationship among activities; (i) positive statements/regard; (j) negative statements/regard; and (k) goal setting. P/CIS behaviors are rated on a scale of 1 (low) to 5 (high), anchored with specific descriptions of the behaviors that characterize each rating. For analyses, mean scores on each scale were computed. Mean values, as opposed to sum scores, were used to avoid deflation of scores due to nonapplicable items[250]. Each of the 11 behaviors is then rated across three distinct dimensions including quality, appropriateness, and amount, for a total of 33 codes. Quality ratings refer to aspects of each of the 11 caregiver behaviors that promote optimal development of the child such as intensity, sensitivity, fluidity, flexibility, and consistency. Appropriateness is the degree of match between the caregiver's behavior and the child's developmental level and interest during the play episode.

**Psychometric properties**. Not specified.

**Coding Manual/Training:** Not provided.

#### Parent Child Early Relational Assessment (ERA)

The Parent Child Early Relational Assessment (ERA) is used to capture the child's experience of the parent, the parent's experience of the child, the affective and behavioral characteristics that each brings to the interaction as well as the overall quality of the relationship[187].

**Paradigms**. Mothers are instructed to play with their child as they normally would. A standardized set of toys is provided. This design facilitates the probability that parents and children will behave as they typically do.

**Populations**. Not specified.

**Scoring.** Sixty-five individual items are rated on 5-point Likert-type scales, including 29 parental items, 28 child items, and 8 dyadic items. Each item is rated in terms of strength and concern. Higher scores indicate more positive interactions.

**Psychometric properties**. The ERA is a validated tool that has been used in over 400 programs and projects[252].

**Coding Manual/Training:** Not provided.

#### Parent-Child Interaction Rating Scales – Infant Adaptation (PCIRS-IA)

The Parent-Child Interaction Rating Scales-Infant Adaptation is an observational scale that aims to assess the quality of parent-child interaction[188].

**Paradigms**. The parent and child interact in a free-play episode. Mothers play with their infant by using toys typically available in homes[41].

**Populations**. Not specified.

**Scoring.** Five domains are coded: parental sensitivity, parental intrusiveness, parental support for cognitive development, parental support for language quantity, and parental support for language quality. Parental sensitivity was scored on a scale from 1-7, low to high, with seven representing a mother that characteristically demonstrated the described behavior[42].

**Psychometric properties**. Not specified.

**Coding Manual/Training:** Not provided.

#### Pediatric Infant Parent Exam (PIPE)

The Pediatric Infant Parent Exam (PIPE) is an observational tool that can be used by community workers to ascertain the quality of parent-infant interactions[189].

**Paradigms**. Mothers and their infants are observed playing a game. This interaction is evaluated for the degree of interactional reciprocity and positive affect at the beginning, middle, and end of the episode[109].

**Populations**. Not specified.

**Scoring.** At each time point, the interaction is scored on a scale of 1 (more favorable interaction patterns) to 6 (less favorable interaction patterns). A total score for the interaction is then calculated by adding the scores from the beginning, middle and end.

**Psychometric properties**. Not specified.

**Coding Manual/Training:** Not provided.

#### Preterm Mother–Infant Interaction Scale (PREMIIS)

The Preterm Mother Infant Interaction Scale (PREMIIS) is a behavioral observation tool that assesses the mother-infant relationship for sensitive and responsive caregiving and awareness of infant cues. The PREMIIS was adapted from the Synchrony Coding Scales for use in preterm infants[101].

**Paradigms**. Mothers can be videotaped before undressing infants, bathing infants, and then dressing infants at term equivalent age. Each segment (undressing, bathing, and dressing) lasts approximately 3 min in a study done by Milgrom and colleagues (2013)[101].

**Populations**. The PREMIIS was specifically adapted from the Synchrony Coding Scales for use in preterm infants.

**Scoring.** 10 maternal items covering responsiveness and affect on 4-point scales, 7 infant items covering stress cues and unstressed behaviors, and 2 dyadic measures of overall synchrony on 5-point scales are measured by an observer.

**Psychometric properties**. Not specified.

**Coding Manual/Training:** Not provided.

#### Sensitivity and Cooperation

The Sensitivity and Cooperation scale is used to assess parental sensitive responsiveness[149].

**Paradigms**. Mothers and their infants are observed in an 8-minute free-play episode. The infant is placed in a seat in front of a low table with the mother sitting adjacent to them. The researcher then presents a transparent box containing 10 toys. The mother is instructed to play with her child as she typically would[34].

**Populations**. Not specified.

**Scoring.** Sensitive responsiveness is rated with two 9-point rating scales for Sensitivity and Cooperation. Lower scores are indicative of insensitivity and interference, and higher scores are indicative of cooperation and sensitivity[34].

**Psychometric properties**. Not specified.

**Coding Manual/Training:** Not provided.

#### Still-Face Procedure (SFP)

The Still-Face Procedure is a widely used observational tool used to assess infant response to a social challenge[190].

**Paradigms**. The Still-Face Procedure introduces an age-appropriate developmental task (face-to-face social interaction) and an age-appropriate episode of mild stress (the mother portraying a still-face and remaining unresponsive), and a reunion episode in which the infant and mother can reorient with one another after the disruption of the still-face (as described in Meijjsen, 2010[68]). The mother sits opposite the infant at eye level. The procedure starts with a 2-minute face-to-face social interaction episode in which the parent is instructed to interact with the infant as they normally would. The parent is told not to use toys in this part of the procedure. Then the parent will turn their back to the infant for fifteen seconds, before turning back with a still-face. The parent is instructed to maintain a still-face and not make any contact with the infant for two minutes. Afterward, the reunion episode can begin, and the parent can begin to interact with the infant normally again.

**Populations**. The still-face effect can be found regardless of most sample variations such as gender of infant or risk status[253].

**Scoring.** Many different coding systems can be used for the Still Face Procedure. In Valades and colleagues study (2021)[79], the coding was based on an established rating scheme[254]. Videos were coded second by second scanning for behaviors and marking them as present or absent. These comprised regulated and non-regulated behaviors. Regulated behaviors include socially positive vocalizations such as coos or laughter. Dysregulated behaviors were comprised of negative vocalizations such as fussing or crying.

In Meijjsen and colleagues' study (2010)[68], the procedure was observed using two cameras, one facing the infant and one facing the parent. The two images were combined using a split-screen approach. For scoring, the Observer 5.0 (Noldus) program was used. In this program, all behaviors are scored on a second-by-second basis using the ICEP coding system.

**Psychometric properties**. Longitudinal research has shown that regulated infant behavior during the Still Face procedure is a reliable predictor of later attachment security[232,255]. Additional meta-analyses also confirmed that higher maternal sensitivity predicted more positive infant affect during the paradigm. Consequently, infants' higher positive affect and lower negative affect during the still-face were found to be predictive of secure attachment at one year of age[253].

**Coding Manual/Training:** Not provided.

#### StimQ2

The StimQ2 measures caregiver cognitive stimulation and interactions in the home using a structured interview[191,256].

**Populations.** This scale has been validated for use primarily in English or Spanish-speaking low-SES populations[191,192].

**Scoring.** The StimQ2 Infant/Toddler has three subscales: (1) Parent Verbal Responsivity (PVR); (2) Parental Involvement in Developmental Advance (PIDA); (3) Reading Activities (READ); and (4) Availability of Learning Materials (ALM). The four subscales are summed for a total score ranging from 0 to 39 (Mendelsohn, 2007) or 43 (Mendelsohn, 2011). PVR assesses verbal interactions between dyads in Everyday Routines and Play and Pretend (Cates, 2018[30]; scored 0-11, Mendelsohn, 2011[192]; or 0-4, Mendelsohn, 2007[38]). PIDA measures caregiver teaching and play activities including naming objects and playing pretend with the infant (scored 0-7, Mendelsohn, 2011[192]; or 0-10, Mendelsohn, 2007[38]). READ measures the number, diversity, and frequency of books read to the child as well as associated interactions across 3 subdomains (Quantity, Quality, Diversity of Concepts, Cates, 2018[30]; scored 0-19, Mendelsohn, 2011[192]; or 0-18, Mendelsohn, 2007[38]). ALM assesses learning materials provided by the caregiver (scored 0-6, Mendelsohn, 2011; or 0-7, Mendelsohn, 2007).

**Psychometric Properties.** The StimQ2 has strong internal consistency; Cronbach's $\alpha = 0.76$[30]- 0.88[38,192]. The assessment also shows good test-retest reliability (ICC = 0.93) as well as criterion-related validity with the HOME Inventory ($r = 0.5$-$0.6$, $p < 0.001$[38,192]). It also demonstrates strong concurrent validity with developmental, cognitive, and language measures ($r = 0.3$-$0.5$) and is correlated with the Bayley Scales of Infant Development Mental Development Index (semi-partial regression (sr) = 0.45, $p < 0.001$) and the One Word Picture Vocabulary Tests (Receptive sr = 0.38, $p = 0.01$; Expressive sr = 0.33, $p = 0.03$) in a study done by Mendelsohn and colleagues (2007)[38].

**Coding Manual/Training.** Not specified.

#### Strange Situation Procedure (SSP)

The Strange Situation Procedure is a widely used observational assessment that measures infant attachment to a caregiver[190].

**Paradigms**. The strange situation procedure consists of eight 3-minute episodes of separation and reunion with a caregiver, specifically designed to induce enough stress in the child to activate attachment behavior[178]. In the first separation, the child is left with a female stranger while the mother exits the playroom, and in the second separation, the child is left fully alone in the playroom. The stranger and mother enter the room in subsequent episodes.

**Populations**. This laboratory paradigm was intended for 12–18-month-old children. This procedure has been used successfully with low-income mothers and mothers from various cultural backgrounds[93].

**Scoring.** Infant behavior during the strange situation procedure is indicative of the infant's attachment pattern. Infant behavior toward the caregiver can be categorized as secure, insecure-avoidant, insecure-resistant, and disorganized/disoriented. The Strange Situation Procedure is videotaped and coded using four 7-point scales for proximity seeking, contact maintaining, avoidance, and resistance and one 9-point scale for disorganization coded through the Main and Solomon (1990[257]) system[45,114,178]. Children are classified as secure if they sought out contact with and were then soothed by their parent. Children are classified as avoidant if they failed to look to their parent for reassurance upon returning to the playroom and/or engage in active avoidant behavior during reunions. Children were classified as resistant if they were not soothed by the parent despite seeking out the parent's comfort upon return. Lastly, children were classified as disorganized, as specified by Main and Solomon (1990) if they displayed contradictory behaviors such as approaching the stranger when distressed, fearful or apprehensive behavior toward or around the parent, disoriented wandering, or rapid changes in affect (descriptions drawn from Bernard et al., 2012[87]).

**Psychometric properties**. This assessment has good reliability and predictive validity[57]. Interrater reliability has been found to be high in numerous studies[45,85,96,178].

**Coding Manual/Training:** Information not provided in reviewed studies.

#### Synchrony Scale

The Synchrony Scale measures mother-infant interaction in terms of behaviors of the mother, the infant, and the dyad[193]. The scale was developed based on the rating protocols described by Censullo and colleagues[258].

**Paradigms**. Mothers are asked to play with their infants for 9 minutes as they normally would and then are instructed to elicit “talking” from their infant for the next 6 minutes.

**Populations**. Not specified.

**Scoring.** Mother, infant, and dyadic behaviors are coded in three-minute increments by a researcher masked to the condition/characteristics of the dyad. Scores range from 0 (lowest) to 3 (highest). The higher the score, the more frequently behaviors occur.

**Psychometric properties**. Construct validity has been deemed acceptable by testing associations with the Global Ratings Scales[259]. The Synchrony Scale has been used in numerous published reports[260].

**Coding Manual/Training:** Not provided.

#### eMethods 5. Description of Included Interventions

Each individual intervention tested in an RCT included in this review is described. Note that very similar models of intervention are often described separately in this appendix to provide more details about the intervention variations (e.g., length, interventionist, location, sample) in the studies included in this review.

##### Skin-to-skin contact (SSC)

All information about the intervention was obtained from Chiu and Anderson (2009)[1] unless otherwise stated.

**Intervention Description**
Dyads were encouraged to begin SSC as early as possible after birth, and to maintain SSC for as long as possible. For mothers initiating SSC immediately after birth, infants were placed between their mother's breasts and covered with a warming blanket that was tucked under the mother on both sides, covering the infant's back. Infants wore a cap that was replaced should it become damp. Mothers who did not begin SSC immediately after birth would hold their infants between their breasts with either their hospital gown or their own clothing and a blanket folded across their infant's back for warmth. Infants wore small diapers and, usually, a cap.
Interventionists were nurse researchers.

**Target Population**
Preterm mother-infant dyads.

**Goal (and framework when applicable)**
Skin-to-skin contact (SSC) is synonymous with kangaroo care (KC) and connected kangaroo mother care (KCM), which usually refers to nearly continuous skin-to-skin contact to facilitate lactation and breastfeeding[1].

**Physical and Informational Materials**
Dyads used large warming blankets. Infants wore small diapers and caps.

**Setting**
SSC was performed face-to-face in the hospital [1].

**Dose and Intensity**
SSC began as soon as possible after birth and was performed for 2 to 5 days.
Because the interventionist had little to no role in this intervention, there may only be one intervention session associated with this intervention (instructional). Most sessions do not involve direct intervention time (i.e., 0 minutes of staff time).

**Studies that Employed this intervention:** Chiu and Anderson [1]

##### Contact Intervention (Kangaroo Care)

All information about the intervention was obtained from Feldman et al. (2014)[2] unless otherwise stated.

**Intervention Description**
Nurses took infants out of incubators, undressed them, and placed them between the mother's breasts while the mother sat in a standard rocking chair. Infants remained attached to their cardiorespiratory monitors during the SSC. Nurses recorded dyadic interactions and would record the exact time of contact. Mothers used a bedside screen for privacy.

**Target population**
Preterm mother-infant dyads.

**Goal (and framework when applicable)**
This intervention draws on Kangaroo Care (KC), initially developed in Bogota, Colombia, as a strategy to cope with a lack of incubators. KC helps premature infants to maintain body heat and contributes to the neuromaturation,

autonomic maturity, electroencephalogram complexity, pain response, and physiologic stability of premature infants. KC has also demonstrated beneficial effects on maternal-infant bonding and maternal mood in early infancy.

**Physical and Informational Materials**
Physical materials included a cardiorespiratory monitor, a standard rocking chair, and a bedside screen.

**Setting**
Face-to-face individually, in the NICU

**Dose and Intensity**
The intervention began at birth and lasted for 14 days.

**Studies that Employed this intervention:** Feldman, Rosenthal and Eidelman [2]

#### Kangaroo Care

All information about the intervention was obtained from Neu and Robinson (2010)[3] unless otherwise stated.

**Intervention Description**
Nurses provided encouragement in holding infants, promoting relaxation during holding infants, information about early infant development, and education about recognizing and responding to infant cues. Nurses also discussed the benefits of holding infants with the mothers. Mothers were instructed to use the kangaroo method to hold their infants for 60 consecutive minutes at least once daily.

**Target population:**
Preterm mother-infant dyads.

**Goal (and framework when applicable)**
The Kangaroo Intervention was derived from the kangaroo holding approach, a method that originated in Bogota, Colombia to support infant survival and care.

**Physical and Informational Materials**
Mothers were offered a holding diary and were asked to record daily quantities of infant holding, including who held the infant and the type of holding.

**Setting**
Dyads were visited in the hospital and in the home. The majority (81.82%) of dyads received 8 of the visits in the home, and the remaining 18.18% of dyads received 7 of the nurse home visits.

**Dose and Intensity**
The intervention was provided in 10, 45–60-minute sessions with interventionists. The first 4 sessions occurred biweekly over the span of 2 weeks, and the remaining 6 sessions occurred weekly.
The intervention began within 4 weeks after birth and lasted for 8 weeks.

**Studies that Employed this intervention:** Neu and Robinson [3]

#### Skin-to-skin contact (SSC)

All information about the intervention was obtained from Rheinheimer et al., (2022)[4] unless otherwise stated.

**Intervention Description**
Mothers were encouraged to undress their infants and to place them in an upright position on the mother's bare chest. Mothers were asked to perform one daily uninterrupted hour of SSC from birth until postnatal week 5.
The intervention was delivered by a researcher.

**Target Population**
Full-term mother-infant dyads.

**Goal (and framework when applicable)**
This Skin-to-Skin contact intervention was built on Feldman's biobehavioral theory of parent-infant interactions, which postulates that repeated mother–infant contact and the resulting exchange of biobehavioral cues in the first postnatal months, facilitate infants' maturation of their ability to regulate autonomous stress reactions [261].

**Physical and Informational Materials**
Mothers used a logbook to record periods of contact (SSC, holding, breastfeeding) The SSC condition received detailed verbal and written instructions on the intervention by the researcher.

**Setting**
Face-to-face in person.

**Dose and Intensity**
The interventionist had little to no role in this intervention after instructions were given outside of phone calls from research staff. Mothers were asked to perform one uninterrupted hour of SSC per day for 35 days (from birth until postnatal week 5). Mothers engaged in 21-35 hours of SSC over the course of the intervention.
Mothers also received 2+ weekly phone calls from research staff.

**Studies that Employed this intervention:** Rheinheimer, Beijers, Cooijmans, Brett and de Weerth [4]

#### Continuous Skin-to-skin contact (SSC)

All information about the intervention was obtained from Sahlen-Helmer et al. (2020)[5] unless otherwise stated.

**Intervention Description**
Infants were placed on their mother's chest in an upright position as soon as possible after birth, where they remained as the mother was transferred to the NICU. Mothers were also instructed to record who provided SSC and whether they paused SSC at any time, for any reason. Additionally, mothers also received a structured education about identifying and responding to preterm infants' signals. This educational program was disseminated by a nurse certified in the Newborn Individualized Developmental Care and Assessment Program (NIDCAP).

**Target population**
Mothers of infants in the NICU

**Goal (and framework when applicable)**
The infant education program was influenced by and aligned with Newborn Individualized Developmental Care and Assessment Program (NIDCAP), a relationship-based model that considers the competence of the infant and the family and promotes infant comfort and optimizing long-term outcomes.

**Physical and Informational Materials**
Not reported

**Setting**
In the NICU (either level 3 NICU at Linköping University Hospital and 1 level 2 NICU at Sachs' Children's Hospital in Stockholm)

**Dose and Intensity**
1 instruction session (as the nurse places the baby on the mother).
Mothers maintained SSC for 7 days, for almost 24 hours a day, but the active time with the interventionist was not explicitly stated (and likely only the initial, 15-minute session).

**Studies that Employed this intervention:** Sahlen Helmer, Birberg Thornberg, Frostell, Ortenstrand and Morelius [5]

#### Community-Initiated Kangaroo Mother Care (ciKMC)

All information about the intervention was obtained from Taneja, Sinha, et al. (2020)[262] unless otherwise stated.

**Intervention Description**
During home visits, a pair of intervention workers visited dyads to explain, initiate, and support the practice of KMC. Interventionists advised mothers to perform skin-skin contact for as long as possible during the day and night. The team observed the mom practicing KMC, enquired about SSC and breastfeeding in the previous 24-hour period, and supported the mother and the family to solve any problems or overcome barriers to effective KMC.
Mothers in both groups also received standard home-based postnatal care (HBNPC) visits by government health workers (Accredited Social Health Activists; ASHAs)

**Target population**
Mothers and low birth weight babies

**Goal (and framework when applicable)**
Draws on "survive and thrive," skin-to-skin contact, and kangaroo mother care (KMC) theories. The intervention sought to improve survival, prevent infections, and promote nutrition and neurodevelopment.

**Physical and Informational Materials**
Not reported.

**Setting**
In the home.

**Dose and Intensity**
The intervention began as soon as possible after birth and lasted 28 days or until the baby wriggled out and no longer accepted SSC, whichever happened first.
The pair of interventionists visited mothers daily for the first 3 days, then on days 5 and 7. Then, interventionists visited dyads twice in the second week and then once a week for the 3rd and 4th week.
The length of each home visit was not reported.

**Studies that Employed this intervention** Taneja, Sinha, Upadhyay, Mazumder, Sommerfelt, Martines, Dalpath, Gupta, Kariger, Bahl, Bhandari, Dua and group [262]

#### Baby Wearing Intervention
All information about the intervention was obtained from Williams and Turner (2020)[7,8] unless otherwise stated.

**Intervention Description**
Mothers assigned to the infant carrier condition were provided with infant carriers and instructed on how to use the carrier by a certified babywearing educator. They were asked to wear their babies in the carrier for at least 1 hour every day. Mothers responded to weekly text messages for three months to report on how often they were using the carriers.

**Target Population**
Young, low-income mothers who were enrolled in Healthy Families or Teen Outreach Pregnancy Services.

**Goal (and framework when applicable)**
The baby wearing intervention was predicated on the theory that the proximity of mothers and infants increases maternal awareness of the child’s needs and subsequent responsive maternal behavior and promotes secure attachment between mothers and their infants

**Physical and Informational Materials**
Infant carrier

**Setting**
The training session took place at home.

**Dose and Intensity**

The intervention included 1 formal training session at 2-4 weeks postpartum, where mothers were instructed on how to use the infant carrier. The length of this initial session with the interventionist was not reported.
Mothers were asked to use the infant carrier for at least one hour a day for the duration of the intervention (6 months). Mothers self-reported baby wearing an average of 4.2 days a week (SD=1.4) for approximately 77 minutes a day (range of 16-200 minutes), for a total of 5.9 hours per week (range of 1.3 - 16 hours).

**Studies that Employed this intervention:** Williams and Turner [7,]Williams and Turner [8]

#### Maternal Singing

All information about the intervention was obtained from Cevasco et al., (2008)[9] unless otherwise stated.

**Intervention Description**
The researcher helped each mother design a song, drawing on a list of songs as inspiration during the hospital stay after birth. The songs were recorded onto a CD that mothers would take home from the hospital with them. The researcher also kept a copy of the CD. Infants were not present for the recording of the CD, and mothers were not present while the CD was subsequently played for the infants.
The CD of the mother's singing was played for each infant 20 minutes per day, 3 to 5 times per week, until the time of discharge. The time of day for music listening was determined based on the infant, visiting mother, and nurse's schedule. These sessions were often scheduled when the infants were awake, sleeping, receiving feedings, or receiving routine medical procedures.

**Target Population**
Preterm mother-infant dyads.

**Goal (and framework when applicable)**
The goal of this intervention was to determine the effects of mothers' recorded singing on mother-infant bonding. An additional goal was to understand the effects of using music in the home in the first two weeks after the infant's birth.

**Physical and Informational Materials**
Mothers were given a notebook containing a list of song titles, including lullabies, children's songs, and popular songs (see Table 3 in Cevasco et al., 2008), as well as the lyrics for each song.
An Olympus WS-100 digital voice recorder and Audio-technical ATR35s omni directional microphone were used to record the singing of mothers of both term and preterm infants. DBpower-AMP Music Converter transferred the Windows Media Audio files to wave files, and Audacity was used to edit wave files. Nero burning room software was used to compile and make CDs. Music was played for preterm infants on RadioShack Moisture Resistant Speaker System MX 1 (Cat No. 40-1400), which was connected to a RadioShack AM/FM Stereo Portable CD player (Cat. No. 42-6013). A RadioShack Sound Level meter (Cat. No. 33-2055) was used to determine if the decibel level for each infant's isolate and open crib was within the guidelines provided by the American Academy of Pediatrics Committee on Environmental Health

**Setting**
Face-to-face individually.
Mothers resided in a NICU 1 setting in a regional medical center. The NICU 1 setting provides both basic care and well care for newborns.

**Dose and Intensity**
The recording process took approximately 20 –60 minutes, depending on the number of songs the mother chose.
The CD of the mother's singing was played for each infant 20 minutes per day, 3 to 5 times per week until time of discharge. Recordings ranged from 20 min to an hour, and total listening time ranged from 240 min - 400 min.
The intervention began soon after birth (between 4 and 17 days) and lasted for 13.3 days (average).

**Studies that Employed this intervention:** Cevasco [9]

#### Music Therapy

All information about the intervention was obtained from Corrigan et al. (2022)[10] unless otherwise stated.

**Intervention Description**
Mothers met with a board-certified music therapist for music therapy sessions. The therapist assessed the mother's ability to cope with hospitalization. The infant's heartbeat was recorded, and the mother chose a song that was meaningful to them that was paired with the heartbeat. The mothers were told to listen to this recording as often as they would like for one week. The number of times the mothers listened was self-reported after one week.

**Target Population**
Preterm mother-infant dyads.

**Goal (and framework when applicable)**
This intervention was based on a theory that personalizing music evokes emotion and promotes comfort, and that heartbeat music improves bereavement bonding.

**Physical and Informational Materials**
Mothers were provided with a CD or digital copy of music containing an infant heartbeat and the mother's music preference.

**Setting**
Face to face, individually in the NICU.

**Dose and Intensity**
The mothers chose how much they listened to their customized CD for one week. The intervention began as soon as possible after birth and continued until postnatal day 7.
The interventionist was only involved in the initial session.

**Studies that Employed this intervention:** Corrigan, Keeler, Miller, Naylor and Diaz [10]

#### Music Therapy (MT)

All information about the intervention was obtained from Gaden et al. (2022) [11] unless otherwise stated.

**Intervention Description**
The intervention was delivered by 11 music therapists who had training and/or prior clinical experience with music therapy in the NICU. Sessions took place while the mother was engaged in skin-skin contact, feeding, or while infants were lying in the incubator or cot. Sessions consisted of parent-led, infant-directed singing, which the music therapist would facilitate support. The quantity of time per session actively making music varied between sessions, depending on the infant's tolerance. During the parts of the session when mothers were not making music, therapists would engage mothers in a discussion about the family's needs.

**Target population**
Preterm mother-infant dyads.

**Goal (and framework when applicable)**
The hypothesis was that this musical therapy intervention, combined with standard care, would improve mother-infant bonding and parental mental health relative to dyads who received standard are alone.

**Physical and Informational Materials**
Melodies were adapted to suit the infant's state, engagement, and age. For infants aged PMA approximately 26-32 weeks, musical therapy would employ, "single notes, simple melodies, and short musical phrases adapted from children's songs or parent-preferred music" [11]. After infants were aged PMA 32 weeks, the music increased in complexity. Throughout the intervention, accompanying instruments were used sparingly to accentuate the parent's voice.

**Setting**
Sessions took place in the NICU, at the mother's bedside or in the family's hospital room.

**Dose and Intensity**
Mothers were offered 3 individual music therapy sessions per week throughout their hospitalization. The maximum number of sessions was 27, and the average was 9.98.
Sessions lasted approximately 30 minutes.
The intervention began in the NICU and lasted until discharge.

**Studies that Employed this intervention:** Gaden, Ghetti, Kvestad, Bieleninik, Stordal, Assmus, Arnon, Elefant, Epstein, Ettenberger, Lichtensztejn, Lindvall, Mangersnes, Roed, Vederhus and Gold [11]

#### Contingent Lullaby

All information about the intervention was obtained from Robertson and Detmer (2019)[12] unless otherwise stated.

**Intervention Description**
The researcher helped mothers to write an original lullaby, that would also be recorded as a reference for the mothers. The researcher would teach each mother how to choose moments to sing the original song to the baby. In line with their contingent music strategy, mothers were instructed to use the lullaby at moments when their baby was quiet and alert. In contrast, mothers were taught not to use their lullabies during periods of infant distress (for example, fussing or crying).

**Target population**
Full-term mother-infant dyads.

**Goal (and framework when applicable)**
This intervention uses contingent music, a behaviorism technique where music either starts or stops when an infant behavior is exhibited. Within the context of this study, this means that when the infant is in a calm state, music is provided, and that music is not provided when the infant is in a state of distress (ex: fussy, crying).

**Physical and Informational Materials**
The researcher helped mothers to write an original lullaby that they would use during the intervention. The lullaby was set to a tune that the mother was familiar with, and the lyrics reflected how mothers felt toward their babies.

**Setting**
Face-to-face, individually. The study took place in regional medical centers (in Florida and Kentucky, USA).

**Dose and Intensity**
There was one session with the interventionist and the length of this session was not reported.
The intervention began soon after birth and continued for up to 6 weeks.

**Studies that Employed this intervention:** Robertson and Detmer [12]

#### Singing

All information about the intervention was obtained from Wulff et al., (2021)[13] unless otherwise stated.

**Intervention Description**
This intervention employed a singing and music-based interaction between the mother and her infant. Songs and games were taught to the mothers during group sessions, facilitated by a music therapist and a member of the study team. If participants requested, the music therapist would incorporate mothers' requests for specific musical content.

**Target Population**
Mothers with postpartum depression and their infants.

**Goal (and framework when applicable)**
Not reported.

**Physical and Informational Materials**
Intervention sessions involved games, songs, and movements.

**Setting**
Sessions were conducted in groups of five to ten women, in person at the gymnastic room of the Clinic for Gynecology and Obstetrics at the University Hospital Duesseldorf.

**Dose and Intensity**
This intervention consisted of 1-3 45-minute sessions
Infants were between 3-10 weeks old at baseline. The intervention took approximately 2 and a half hours in total.

**Studies that Employed this intervention:** Wulff, Hepp, Wolf, Fehm and Schaal [13]

#### Maternal Voice Recording

All information about the intervention was obtained from Yu et al. (2022)[14] unless otherwise stated.

**Intervention Description**
During this intervention, maternal voice recordings were obtained in the NICU and subsequently played for infants in the intervention group starting at 3 min before a heel stick procedure and lasting until the procedure completion, 13 min in total. The infants received containment and nonnutritive suckling after the procedure if they cried for >1 min. The intervention was recorded using a camera, and the videos were privately shared with the mothers to facilitate mother–infant bonding and to offer the mothers a visualization of the study process.

**Target Population**
Preterm mother-infant dyads.

**Goal (and framework when applicable)**
Not reported.

**Physical and Informational Materials**
Maternal voice recordings of the mother's reading of a children's book presenting the reflections of a premature infant's mother[263]. In this book, premature infants were likened unto "immature persimmons who are nurtured by their family through patience and affection while undergoing self-exploration and passing through all difficulties to eventually become sweet and mature fruits" [14,263]. Mothers also included words that they wanted their infant to hear. The recordings were edited into a 13-minute audio files using the music editing software Sound Organizer 2, per study methods.

**Setting**
Pre-recording of mother's voice + infant in person, in the NICU.

**Dose and Intensity**
The intervention began on postnatal day 4.
There were 3 sessions that took place on 3 consecutive days, each lasting approximately 39 minutes.

**Studies that Employed this intervention:** Yu, Chiang, Lin, Chang, Lin and Chen [14]

#### My Baby and Me

All information about the intervention was obtained from Akai et al. (2008)[15] unless otherwise stated.

**Intervention Description**
My Baby and Me is an intervention protocol designed to improve early parenting by enabling parents to understand their baby's developmental needs. There are three modules: responsiveness training (from the PALS curriculum[264]), developmental knowledge training, and loving touch training.

**Target population**

Mothers who are at risk for problematic parenting, and their 3.5-5.5-month-old infants.

**Goal (and framework when applicable)**
The main goal of the My Baby and Me program is to improve parenting during a child's first year. The aim is to decrease psychological control and increase behavioral control. This is achieved by addressing harsh and intrusive thoughts for parents, facilitating parental responsiveness to infants' developmental needs, and encouraging warm behavior and positive emotional expression.

**Physical and Informational Materials**
Mothers are given a scripted protocol to aid their responsiveness to infants' needs. They are given a package of My Baby and Me training modules. They are also given a "Take Time for Kids" booklet set, which contains one booklet for each month of the child's first year.

**Setting**
Intervention sessions occur face-to-face individually, in the participant's home.

**Dose and Intensity**
There are 12 to 14 sessions, each session is 1.5 hours.
The intervention starts when the infant is 3.5-5.5 months old and continues for 12-14 weeks (average of 15.3 weeks).

**Studies that Employed this intervention**: Akai, Guttentag, Baggett, Noria and Centers for the Prevention of Child [15]

#### Infant-Net (Internet-Adapted PALS Program)

All information about the intervention was obtained from Baggett et al. (2010, 2017)[16,17] unless otherwise stated.

**Intervention Description**
Mothers are given a computer, 6 months of internet connection, and access to the Infant-Net program, an Internet-adapted version of the PALS program. The intervention content includes video models and multi-media information on concepts, behaviors, and skills that featured caregivers and infants of diverse in terms of races, ethnicities, and ages [265,266]. The intervention includes self-directed aspects and ongoing contact with a coach. Throughout each session, there are questions with answers that could be reviewed by the caregivers and the coach after each self-directed learning session. The program includes summaries of the key concepts and homework based on the various skills taught during each session. Mother-infant videos were collected throughout the program and revised by the parent and coach to see how mothers were implementing each of the skills they learned. Coaches provided parents with individualized support via weekly phone calls.
Through the intervention, mothers were given tools and strategies to maintain the infant's focus during routine care activities. The intervention also used therapeutic and educational strategies such as coaching behavioral skills, personal behavior reflection, and time for parents to practice new skills.

**Target population**
Mother-infant dyads in low-SES environments [16,17]; mother-infant dyads at a higher risk of maltreatment [17].

**Goal (and framework when applicable)**
The PALS intervention was based on attachment theory. This internet version was derived from the traditional PALS program, which had the goal of supporting the caregiver-infant relationship. The intervention employed videos and video-feedback to demonstrate infant signals and promote warm, sensitive caregiver responses.

**Physical and Informational Materials**
A computer with a manual was provided to the participants as well as instructions for how to use the Infant-Net program.

**Setting**
This intervention was given at home on the computer with access to a remote coach.

**Dose and Intensity**
Each session was approximately 90 minutes (about 1 and a half hours) long, with the program consisting of 11 sessions, including an introductory session in which mothers learned how to use the laptop and interface which lasted approximately 15 minutes. The self-directed part of each session took participants 25-35 minutes on average and the participants were given a laptop for 6 months [161]Mothers also participated in 30-minute weekly coach calls. In total, participants spent 22.7 hours on this intervention (ranging from 11.6 to 46.1 hours). This total time also includes the time that mothers optionally spent reviewing completed videos and materials on their own.
The time that participants spent interacting with parenting coaches directly was not reported.

**Studies that Employed this intervention:** Baggett, Davis, Feil, Sheeber, Landry, Carta and Leve [16,]Baggett, Davis, Feil, Sheeber, Landry, Leve and Johnson [17]

#### Adapted Play and Learning Strategies Program (ePALS)

All information about the intervention was obtained from Feil et al. (2020)[18] unless otherwise stated.

**Intervention Description**
This bilingual, 11-session adaptation of the Play and Learning Strategies (PALS) program consisted of individualized, online coaching sessions. Mothers could select their preferred language (either English or Spanish) at the beginning or during the intervention. These sessions consisted of:

1. Self-directed skill-learning through video-based teaching, with check-in questions, using immediate individualized feedback
2. Constructing an action plan that outlines daily homework based on the skills taught
3. Video-recorded practice that is uploaded to a project server
4. Coach calls to co-view weekly videos with mothers, and to provide mothers with individualized support

Sessions were facilitated by coaches, who held at minimum a bachelor's degree in counseling, psychology, social work, or early childhood education. Counselors partook in extensive training, including a training course with a national PALS counselor, meetings with a coach supervisor, and completion of the program from the parent's perspective.

**Target population**
Mother-infant dyads in low-income environments

**Goal (and framework when applicable)**
The ePALS program draws on the theory that caregivers' language and responses influence an infant's social-emotional, cognitive, and language development and that promoting sensitive and responsive parenting during infancy supports these developmental outcomes.

**Physical and Informational Materials**
Materials included a laptop computer, video modeling of skills, skills practice homework, a self-guided instructional component that resented key concepts, and example videos that included narration with questions to aid the mother's completion of the course. Mothers were reminded that they could complete the course at their own pace.

**Setting**
The intervention was performed online, in the home, using a video platform.

**Dose and Intensity**
11 sessions and one instructional session. The sessions began when infants were between 3.5 and 7.5 months, with an average baseline age of 4.41 months. The program took 4-6 months to complete.
The estimated time of each session, specifically the time during which participants were meeting actively with a coach, was not reported.

**Studies that Employed this intervention:** Feil, Baggett, Davis, Landry, Sheeber, Leve and Johnson [18]

#### Playing and Learning Strategies (PALS)

All information about the intervention was obtained from Landry et al. (2006)[19] unless otherwise stated.

**Intervention Description**
This in-person intervention used a combination of video-recording and immediate feedback. The intervention was led by PALS facilitators, who were instructed to follow a detailed PALS curriculum, but to disseminate it in a flexible, individually adapted way.
The format of the intervention included:

1. Asking mothers to review their experiences across the last week related to their efforts to try the targeted behaviors
2. Describing the current visit's targeted behavior
3. Watching and discussing with mothers the educational videotape of mothers from similar backgrounds
4. Videotaping mother-infant interactions in situations selected by the mothers (ex. Toy play, feeding, or bathing) with coaching
5. Supporting mothers to critique their behaviors and the infants' responses during the videotaped practice
6. Planning with mothers as to how to integrate responsive behaviors into everyday activities with laminated cards that define the behavior and its importance

**Target population**
Low-income mothers with low birth weight or term infant

**Goal (and framework when applicable)**
The PALS intervention builds on attachment theory. The goals of the intervention are to incorporate each family's social context into the intervention and to improve infant development by teaching caregivers how to engage in responsive parenting behaviors.
The goal of each of the ten home visits is provided in Table 1 of the Appendix of Landry et al. (2006)[19].

**Physical and Informational Materials**
PALS facilitators followed a curriculum that included behaviors linked to four aspects of responsiveness. Mothers were also provided with developmental handouts covering issues such as sleep, feeding, and pacifiers.

**Setting**
The intervention was performed face-to-face individually, in the home.

**Dose and Intensity**
10 weekly home visits, each visit was 90 minutes.
The first session was performed when infants were approximately 6-months old. Each family was seen for 14 visits, including assessments. Therefore, the intervention took 13-14 weeks to complete, with a mean of 14.5 weeks.

**Studies that Employed this intervention:** Landry, Smith and Swank [19]

#### TAC-TIC (Touching and caressing; tender in caring)

All information about the intervention was obtained from Barnes et al. (2022)[20] unless otherwise stated.

**Intervention Description**
In the TAC-TIC intervention group, mothers were taught a specific protocol of systematic touch known as TAC-TIC (Touching and Caressing; Tender In Caring). This version of TAC-TIC was developed for use with relatively healthy preterm infants. The program consists of 14 individual stroking movements that are repeated continuously 3 times during a 3-minute session without undressing the baby. This version of TAC-TIC was designed to be carried out by the mother, in contrast to prior versions of TAC-TIC that were designed to be carried out by a researcher. Mothers were taught how to perform TAC-TIC on a doll first before touching their own baby. The information provided to mothers was only procedural and involved describing and demonstrating how the baby should be touched. Once mothers understood the instructions, they proceeded with the first session of the 10-day study period (1 session of 3 minutes per day) under supervision by a trained psychologist associated with the study.

**Target population**
Preterm mother-infant dyads.

**Goal (and framework when applicable)**
Not reported.

**Physical and Informational Materials**
Mothers were taught the TAC-TIC protocol of systematic touch. The information provided to mothers was only procedural and involved describing and demonstrating how the baby should be touched.

**Setting**
Not reported. It is not clear who taught the initial session, but the first supervised session was overseen by the principal investigator (a trained psychologist).

**Dose and Intensity**
The intervention began on postnatal day 28 (mean of 11.7; SD = 4.2) and lasted for 10 days.
The initial introduction session was followed by 1 session of 3 minutes per day. The total intervention length was approximately 33 minutes.

**Studies that Employed this intervention:** Barnes and N. Adamson-Macedo [20]

#### Massage Class + Support Group

All information about the intervention was obtained from Onozawaa et al. (2001)[21] unless otherwise stated.

**Intervention Description**
Mothers were taught to recognize and quickly respond to infant self-regulation cues. Mothers attended informal group discussion sessions led by study staff that covered practical problems and coping strategies. The intervention also included infant massage sessions. These classes began with a relaxation period where mothers could unwind. Massage sessions were also taught by instructors trained according to procedures approved by the International Association of Infant Massage disseminated a curriculum of massage techniques. During massage sessions, the instructor demonstrated the massage strokes on a doll, while the mother massaged her own infant.
Mothers were taught that massage is most effective when babies are in a quiet, alert state. Engagement cues include “bright-eyed focused expression, still/calm attentiveness, relaxed arms, shoulders and palms. Disengagement cues include gaze aversion, yawning, arching, grimacing, anxious tongue poking and legs/arms held stiffly.”
The massage curriculum proceeded as follows:
“The class begins with a short period of relaxation which allows the parents to unwind. Then they place a resting hand on the infant, and for some very young or sensitive infants this will be as far as they go. If infants indicate that they are not in the right mood, the massage is not begun. The massage begins with slow rhythmic strokes, the mother’s speed and timing being guided by the infant’s body signals. Each part of the body is treated in a different way, e.g. legs: milking strokes from hip to foot, gentle squeezes and twists in a wringing motion; foot: gentle pressing on a sole of foot, stroking from toes to ankle on the top of the foot; abdomen: hand over hand strokes in a paddle wheel fashion, circular clockwise direction strokes avoiding the ribs.” [21]

**Target population**
Mothers with postpartum depression and their infants.

**Goal (and framework when applicable)**
Not reported.

**Physical and Informational Materials**
Mothers were taught to observe and respond to their infants’ body language and cues.

**Setting**
Hospital (Queen Charlotte’s and Chelsea Hospital)

**Dose and Intensity**
The intervention consisted of 5 dyadic massage classes and 5 non-dyadic support classes, each offered weekly for 5 weeks. Massage classes were 60 minutes, and support group sessions were 30 minutes.
The total combined timing of the intervention was 450 minutes.

Infants were 9 weeks (on average) at baseline.

**Studies that Employed this intervention:** Onozawaa, Gloverb, Adamsb, Modib and Kumara [21]

#### Infant-Massage-Parenting Enhancement Program (IMPEP)

All information about the intervention was obtained from Porter et al. (2015)[22] unless otherwise stated.

**Intervention Description**
Groups of 4-6 mother-infant dyads were taught infant massage techniques by certified nurse practitioners. The sessions were psychoeducational, incorporated both demonstrations and supervised practice, and included massage practice, interactive group discussions, and question-and-answer sessions focused on childcare. Mothers were taught simple games and interactive action-based songs to stimulate the infant and facilitate mother-infant communication while performing infant massage.
The classes included a 10-minute break, which included food and beverages. The last class was a celebration of completion, where mothers received an appreciation certificate and a gift basket of age-appropriate childcare items. IMPEP was built off of the parenting enhancement program (PEP) which only consisted of psychoeducational group sessions.

**Target population**
Recovering substance-abusing mothers (SAMs) and their infants.

**Goal (and framework when applicable)**
This intervention was influenced by Bandura's Social learning theory, which posits that a person's expectations and incentives influence a person's behavior. The aim of the intervention was to empower substance-abusing mothers to make cognitive behavioral changes through learning and applying new parenting knowledge and skills. Through doing so, Porter (2015) aimed to improve a mother's confidence in her knowledge of infant massage and understanding of her infant's body signals, and thereby to empower mothers with the knowledge that they can help relieve their infant's discomfort.

**Physical and Informational Materials**
Psychoeducational group sessions incorporated demonstration and supervised practice of infant massage techniques, interactive group discussions, and question-and-answer periods focused on childcare practices.
Mothers were taught simple games and interactive songs to employ during infant massage.
Sessions included demonstration and practice assessing infant temperature, pulse, respirations, and cardiopulmonary resuscitation.

**Setting**
The intervention was conducted in person, in groups of 4-6 dyads. The sessions took place in a dedicated meeting space at each study site.

**Dose and Intensity**
4, weekly 120–240-minute sessions. The baseline age of infants in this study was not reported

**Studies that Employed this intervention**: Porter, Porter, McCoy, Bango-Sanchez, Kissel, Williams and Nunnewar [22]

#### Occupational Therapy Intervention

All information about the intervention was obtained from Sajaniemi et al. (2001)[23] unless otherwise stated.

**Intervention Description**
The intervention combined several occupational therapy methods that focused on the child but achieved this through interaction with the parents. During the sessions, the interventionists instructed parents to provide the right amount and combination of stimulation at the right time to help the child cope with the sensory demands of the social and physical world. Therapists also worked with parents to help interpret their infant's messages.

**Target population**
Extremely low birth weight infants and their mothers

**Goal (and framework when applicable)**
The general goals of this intervention were to promote normal sensorimotor development, the development of play, and social emotional development through supporting the parent-infant relationship. Specific goals were individually tailored to the specific needs of families.

**Physical and Informational Materials**
The primary topics covered during sessions included parenting a premature baby, general child development, risk factors of premature infants, and the development of play in children.

**Setting**
Face-to-face individually, in the home + clinic visits.

**Dose and Intensity**
The intervention consisted of 60 minutes of occupational therapy per week, at home, starting when infants were age 6 months to 12 months. The average number of sessions was 20 (approximately 1200 minutes total).

**Studies that Employed this intervention**: Sajaniemi, Makela, Salokorpi, von Wendt, Hamalainen and Hakamies-Blomqvist [23]

#### Massage Intervention

All information about the intervention was obtained from Shoghi et al., (2018)[24] unless otherwise stated.

**Intervention Description**
The massage intervention consisted of 2, 1-hour massage training sessions with a researcher. Mothers were also provided with an educational booklet and compact disc published by the Neonatal Health Department of the Ministry of Health of the Islamic Republic of Iran and were shown a training video. The researcher provided a 24-hour call service to answer any of the mothers' questions. The researcher was present for the mother's first 15-minute session massaging her infant. The mothers continued to massage their infants for 15 minutes, 3 times per day for 5 days.
During training sessions, mothers were taught massage techniques using infant mannequins.

**Target population**
First-time mothers and their premature infants. Infants were born in the late preterm birth stage (gestational ages between 34-37 weeks).

**Goal (and framework when applicable)**
The intervention integrates attachment theory and the literature on skin-skin contact. This cost-effective intervention is intended to improve mother-infant interaction, and thereby promote the formation of a secure attachment between mothers and infants.

**Physical and Informational Materials**
Mothers were provided with an education booklet and a compact disk published by the Neonatal Health Department of the Ministry of Health of the Republic of Iran. Mothers also watched a training video on infant massage.

**Setting**
Face-to-face in a hospital setting.

**Dose and Intensity**
2, 1-hour training sessions provided by the researcher.
Mothers continued to massage their infants for 15 minutes, 3 times a day for 5 days. A researcher was present for the initial massage sessions, and all subsequent sessions were performed independently.
Infants were 7.20 days old (average) at the start of the intervention. The intervention lasted for 5 days.

**Studies that Employed this intervention:** Shoghi, Sohrabi and Rasouli [24]

#### Infant Intervention

All information about the intervention was obtained from Teti et al. (2009)[25] unless otherwise stated.

**Intervention Description**

The intervention contained several components– infant tactile-kinesthetic stimulation, a psychoeducational aspect, and employment of the Brazelton Neonatal Behavioral Assessment Scale (NBAS).
Interventionists were certified NBAS examiners.
NBAS:The interventionist administered the Brazelton NBAS seven times throughout the intervention, beginning when the infant was 34-38 weeks PCA [267]. This administration was followed by a discussion between the observers about the infant's strengths and areas of special need. Parents became increasingly involved in the administration of the NBAS as the intervention progressed. The NBAS was administered at 36 to 40, 38 to 42, 40 to 44, 44 to 48, 48 to 52, and 52 to 56 weeks PCA. Parents were encouraged to repeat the NBAS with their infants with the goal of completing the exam, with guidance, by 44 to 48 weeks.
Massage Therapy: Massage therapy was introduced when the infant was 36-40 weeks PCA and was included in the remaining five intervention sessions, with NBAS administration following the massage session. The massage protocol was adapted from previous protocols[268]. The interventionist demonstrated the massage techniques and gave parents a copy of the massage protocol.
When the infant was between 36-40 weeks PCA, the massage protocol involved alternating 5-minute phases of tactile and kinesthetic stimulation. During tactile stimulation, parents were taught to use both hands to massage the infant's head, neck, shoulder, back, waist, thigh, foot, and arm regions with 10-12 strokes. During the kinesthetic stimulation phase, the parent administered 5, 1-minute segments of gentle, passive flexing and extending of the infant's arms, and legs while the infant was in a supine position.
The interventionist demonstrated the massage technique at 38 to 42, 40 to 44, 44 to 48, 48 to 52, and 52 to 56 weeks' PCA, followed each time by NBAS administration.
The massage protocol was practiced and discussed during intervention sessions. Parents were also encouraged to practice massage sessions 2 to 3 times a day, and to record how often they massaged their babies.
Psychoeducation: A 20-minute video titled "Premie Talk: Understanding Your Premature Baby's Behavior" was shown to parents when their infants were 32 to 36 and 34 to 38 weeks' PCA in a private room or, if the infant had been discharged, in the home [269]. The interventionist then facilitated a discussion about premature infants that had several points:

(a) premature infants are sensitive to their environments and use behavior to communicate
(b) the NICU is a noisy, over-stimulating environment
(c) preterm infants are not as responsive as full-term babies
(d) preterm babies respond best to gentle stimulation
(e) preterm babies have three basic states of consciousness: sleep, awake and alert, and fussy
(f) the awake and alert state is best for social interaction. Two weeks later, at 34 to 38 weeks' PCA, the video was shown again to parents and the interventionist reviewed the points covered at 32 to 36 weeks' PCA, responded to questions, and covered the following additional points
(g) preterm babies use engagement and disengagement cues to indicate when they are ready to interact and when they "need a break,"
(h) what to do when the baby shows disengagement cues during interaction
(i) how to awaken a drowsy baby for a feeding
(j) how to calm a fussy baby.

This was followed with a review and recap of all major points.

**Target population**

African American mothers and their low birth weight and / or premature infants.

**Goal (and framework when applicable)**

The goal of the intervention is to improve outcomes by targeting both parents and infants, rather than to target one or the other alone [270]. The intervention included parentally administered infant massage (infant tactile-kinesthetic stimulation) to promote infant development and improve parental knowledge of infant cues. The intervention included a psychoeducational component that targeted foster parents' knowledge about preterm infants' needs and cues and how to, "read, respond to, and facilitate infant social behavior.[25]"

**Physical and Informational Materials**
Interventionists used the Brazelton Neonatal Behavioral Assessment Scale (NBAS), as well as a 20-minute video entitled "Premie Talk: Understanding Your Premature Baby's Behavior, that highlights preterm infants' perceptual and interactive capacities that can inform caregiving practices [269].

**Setting**
The intervention was delivered face to face, individually in a private room or, if the infant had been discharged, in the home.

**Dose and Intensity**
8 sessions that ranged from 60 to 120 minutes in length (720 minutes total). The intervention began at 32-38 weeks PCA and lasted for 20 weeks.

**Studies that Employed this intervention:** Teti, Black, Viscardi, Glass, O'Connell, Baker, Cusson and Hess [25]

#### Maternal Sensitivity Program (MSP)

All information about the intervention was obtained from Alvarenga et al. (2020)[26] unless otherwise stated.

**Intervention Description**
The Maternal Sensitivity Program (MSP) is a short video feedback intervention protocol that focuses on mother-infant interactions to enhance maternal sensitivity. The intervention is conducted over the course of 8 visits, each with 2 parts. In the first part, the mother is recorded playing with her infant. During the second part, an intervener selects scenes from the video recording to watch with the mother and discusses ways to facilitate the infant's development.

**Target population**
Low-income parents with infants under 3 months old.

**Goal (and framework when applicable)**
The aim of MSP is to improve maternal sensitivity and promote positive infant development among low-income families.

**Physical and Informational Materials**
The intervention includes a manual, which describes the intervener's role in the visit and specifies optimal mother and infant behaviors in the videos. The intervention also used the Coding of Early Maternal and Child Interaction (CITMI-R), a structured coding system, to analyze mother-infant interactions. Interveners were supervised and given recommendations for theoretical and practical training.

**Setting**
The intervention occurred in the mother's home.

**Dose and Intensity**
MSP consists of 8, 1-hour home visits when the infant was 3 months old and continued until infants were 10 months old.

**Studies that Employed this intervention:** Alvarenga, Cerezo, Wiese and Piccinini [26]

#### Video-Interaction Guidance (VIG)

All information about the intervention was obtained from Hoffenkamp et al. (2015) or Barlow et al. (2016)[27,32] unless otherwise stated.

**Intervention Description**

VIG is a short video-feedback intervention that focuses on behavior to encourage parents' self-reflection on successful parent-child interactions. Parent-infant interactions were recorded and a VIG professional guided the parent in observation, analysis, and discussion of the infant's behavior [32].
The core aspects of the VIG model include the following: 1) video recording the parent-infant interaction during play or daily caregiving; 2) editing the recording to select moments of optimal parent-infant interaction where the parent attends to their infant's signals; and 3) joint viewing of these recordings with the VIG guide and the parent. VIG provides a parent with the opportunity to view their own interactions to evoke feelings of empowerment and self-efficacy[27].

**Target population**
Preterm infants and their caregivers

**Goal (and framework when applicable)**
VIG sought to improve parental sensitivity after leaving the NICU and returning home [27]. The intervention also aims to foster parental bonding, improve the quality of parent-infant interactions, and promote parental well-being [32]. VIG is based on the assumption that newborns seek parental contact [32]. It is also based on two concepts: intersubjectivity and mediated learning. Intersubjectivity is modeled by the VIG guide through their interactions with the parent and is also pointed out in the recorded parent-infant interactions. Mediated learning occurs as the VIG guide supports the parent's learning by reviewing the video recordings[27].

**Physical and Informational Materials**
Not reported.

**Setting**
VIG occurs in both the NICU and the parent's own home.

**Dose and Intensity**
VIG occurs in 3 sessions. The duration of these sessions and the total duration of the intervention are not reported [27]. The intervention began at birth and continued until postnatal day 7 [32].

**Studies that Employed this intervention**
Barlow, Sembi and Underdown [27,]Hoffenkamp, Tooten, Hall, Braeken, Eliens, Vingerhoets and van Bakel [32]

#### Early Intervention Program
All information about the intervention was obtained from Borghini et al. (2014)[28] unless otherwise stated.

**Intervention Description**
The early intervention program is a transactional preventative intervention. The intervention occurs in 3 phases: (1) 33 weeks after conception, (2) 42 weeks after conception, and (3) 4 months after 40 weeks of conception. During phase 1, the mother, NICU nurse, and intervener jointly observe the infant's behavior in the NICU. During phase 2, behavioral assessments of the infant are videotaped to identify the infant's stress reactions and self-regulation strategies. Mothers address their own emotions and explore symptoms of post-traumatic stress disorder. During phase 3, video recordings of mother-infant free play are produced to observe the infant and promote the mother's caregiving qualities.

**Target population**
Infants born preterm (less than 33 weeks gestational age)

**Goal (and framework when applicable)**
This program follows a therapeutic treatment model which incorporates family system theory and preventive intervention measures.
The goal is to improve parents' understanding of their infants' characteristics and competencies in addition to promoting parental sensitivity and responsiveness.

**Physical and Informational Materials**

The Neonatal Behavioral Assessment Scale (NBAS) is used as a framework to record and review a behavioral assessment of the infant.

**Setting**
The intervention takes place in the NICU.

**Dose and Intensity**
This intervention takes place over 4 sessions. The first session lasted between 30-60 minutes.is one session that lasts between 30-60 minutes. During phase 3, there are 3 different sessions, each one week apart. Each session is 40-60 minutes long. During these sessions, the mother and infant take part in 10 minutes of free play. The intervention begins 33 weeks after conception and ends at 4 months (corrected age).

**Studies that Employed this intervention**: Borghini, Habersaat, Forcada-Guex, Nessi, Pierrehumbert, Ansermet and Muller-Nix [28]

#### Parent-Centered Intervention Program

All information about the intervention was obtained from Brisch et al. (2003)[29] unless otherwise stated.

**Intervention Description**
This intervention is made up of group psychotherapy, attachment-oriented individual therapy, a home visit, and video-based sensitivity training. It is intended to help parents cope after their baby's premature birth, specifically focusing on their emotions and their experiences with neonatal intensive care. This parent-centered intervention program deals with the experiences of loss, separation, and positive attachment from the parent's perspective.

**Target population**
Parents and their premature infants

**Goal (and framework when applicable)**
The goal of this intervention is to improve parental coping, optimize attachment, and enhance parent-infant interaction. This intervention is based on attachment theory.

**Physical and Informational Materials**
Group therapy sessions, individual therapy sessions, a home visit, and video sensitivity training.

**Setting**
Hybrid, some sessions took place in the NICU and others took place in the home.

**Dose and Intensity**
There are approximately 12 sessions in total, although the actual number of sessions varied. Five sessions took place in a group setting (range of 1-8 group sessions in the study reviewed); 5 were individual sessions (range of 1-10 in the study reviewed). Families additionally took part in 1 home visit and 1 sensitivity training. The length of the sessions was not reported. The intervention begins when the infant is In the NICU and continues until they are 3 months (corrected age).

**Studies that Employed this intervention:** Brisch, Bechinger, Betzler and Heinemann [29]

#### Step Towards Effective and Enjoyable Parenting b (STEEP-b)

All information about the intervention was obtained from Firk et al. (2021)[31] unless otherwise stated.

**Intervention Description**
STEEP-b is an adaptation of the original STEEP, which is an attachment-based program for adolescent mothers. STEEP-b is designed to be a shorter version of this program and specifically focuses on parental sensitivity. This intervention uses video feedback to improve material sensitivity. Each session focuses on one of four modules: child development, maternal sensitivity, frightening and intrusive behaviors of the mother, and sensitive parental discipline practices. Each module is worked on twice throughout the intervention.

**Target population**
High-risk adolescent mothers and their infants

**Goal (and framework when applicable)**
The main goals of STEEP-b are to further secure maternal-infant attachment by advancing parental sensitivity using video feedback. Video feedback is utilized to capture maternal attachment and its influence on parenting behavior. The framework behind this intervention is attachment theory as STEEP-b focuses on maternal sensitivity to enhance attachment security.

**Physical and Informational Materials**
There is no manual for STEEP-b. All participants receive publicly funded healthcare and social services when appropriate. All STEEP-b trainers are child and adolescent psychiatrists, psychotherapists, or clinical social workers.

**Setting**
The intervention takes place in the mother's home.

**Dose and Intensity**
STEEP-b occurs in 12–18 sessions over a 9-month period, starting at postnatal month 3-6 (5.41 months average). Adolescent mothers are visited at home every 2–3 weeks by the same trainer for a duration of 9 months.

**Studies that Employed this intervention**: Firk, Dahmen, Dempfle, Niessen, Baumann, Schwarte, Koslowski, Kelberlau, Konrad and Herpertz-Dahlmann [31]

#### Video Feedback of Infant-Parent Interaction (VIPI)

All information about the intervention was obtained from Noivik et al. (2015)[33] unless otherwise stated.

**Intervention Description**
VIPI is a home-based intervention program that targets parental sensitivity by recording and reviewing parent-child interactions.

**Target population**
Families with children under 24 months old who had early health or developmental conditions.

**Goal (and framework when applicable)**
VIPI is based on the Marte Meo method, a home-based intervention that integrates interactional and behavioral approaches with the representational approach. The intervention was designed to treat families with many different types of parent-child interactional problems.

**Physical and Informational Materials**
A VIPI manual was used to standardize the intervention. The manual detailed the order of the intervention's sequences, the number of meetings, and homework between sessions. Participants also received videotapes of the therapists' feedback.

**Setting**
The visits took place in the participants' homes.

**Dose and Intensity**
There are at least 6 sessions, with the opportunity for additional sessions if necessary. It is recommended that the sessions occur weekly. In the present study, the average number of sessions was 8, 30 minutes each, for a total of approximately 4 hours.
The intervention begins when the child is between 0-24 months old, with an average of 7.3 months. The maximum intervention length is 3 months.

**Studies that Employed this intervention**: Hoivik, Lydersen, Drugli, Onsoien, Hansen and Nielsen [33]

#### Book + Video-Feedback (VF) Home Visits

All information about the intervention was obtained from Juffer et al. (2005)[34] unless otherwise stated.

**Intervention Description**
This intervention consisted of 3 sessions of video feedback along with a personal book, which contains information focused on sensitive parenting. The book has the infant's name integrated into the text and includes suggestions for sensitive and playful mother-child interactions. The mother-child interactions were videotaped and then reviewed with an intervener.

**Target population**
Families with an adopted child, with or without birth children.

**Goal (and framework when applicable)**
The intervention is based on attachment theory. The goal of the book + VF intervention was to promote secure attachment in mother-child relationships along with infant competence.

**Physical and Informational Materials**
Mothers were given a personal book with their infant's name integrated into the text. This book provided information about sensitive parenting and suggestions for how to practice it.

**Setting**
Visits occurred in the parent's home.

**Dose and Intensity**
The VF intervention occurs in three, 1-hour sessions. The estimated total length of the intervention is 3 hours. The intervention began when the child was 6 months old and continued for 3 months, ending when the child was 9 months old.

**Studies that Employed this intervention**: Juffer, Bakermans-Kranenburg and van Ijzendoorn [34]

#### Video-feedback interaction to promote positive parenting (VIPP); VIPP with Representational focus (VIPP-R)

All information about the intervention was obtained from Klein Velderman et al. (2006)[36] unless otherwise stated.

**Intervention Description**
VIPP is an early and brief video-feedback intervention that focuses on improving maternal sensitivity during the infant's first year of life. VIPP-R is a modified version of the VIPP intervention that promotes positive parenting but has an additional representational focus intended to influence the mother's representation of attachment.

**Target population**
First-time mothers with insecure attachment, and their young children (between 7 and 10 months old)

**Goal (and framework when applicable)**
Both VIPP and VIPP-R are based on attachment theory and Patterson's coercion theory, which posits that repeated coercive parent-child interactions establish a behavioral system that reinforces negative and antisocial behaviors.

**Physical and Informational Materials**
Mothers are asked to complete a "baby's diary," to chronicle their baby's behavior and the activities they do with their baby. They are also given questionnaires on social support and behavioral problems.

**Setting**
The intervention sessions occurred in the participant's home.

**Dose and Intensity**

VIPP takes place over 4 sessions that are 90 minutes each, for a total of 6 hours. The intervention begins when infants are between 7-10 months old (average of 6.83 months) and continues for 3-4 weeks.
VIPP-R takes place over 4 sessions that are 3 hours each, for a total of 12 hours. The intervention begins when infants are between 7-10 months old and continues for 3-4 weeks.

**Studies that Employed this intervention**: Klein Velderman, Bakermans-Kranenburg, Juffer and van [36]

#### Education Program

All information about the intervention was obtained from Magill-Evans et al. (2007)[37] unless otherwise stated.

**Intervention Description**
The intervention consisted of two sessions with identical protocols. Intervention sessions consisted of videotaped self-modeling and positive feedback regarding father-infant interactions.
Fathers were videotaped while teaching their 5-month-old infant to play with a novel toy (rattle, squeak toy, blocks, etc.). Fathers selected the toy from a list from the Nursing Child Assessment Teaching Scale [271].
Immediately after the videotape was recorded, the father and home visitor reviewed and discussed the videotape together. The home visitor would identify, explain, and praise moments when the father recognized and responded to their infant's cues. Fathers received a copy of their interactive videotape after the first home visit. Fathers were provided with handouts that described aspects of parent-infant interaction.
The intervention was delivered by home visitors.

**Target Population**
Fathers and their infants.

**Goal (and framework when applicable)**
Videotaped self-modeling sought to reinforce positive behaviors. Literature has found that adults focus their attention and become emotionally aroused when viewing them-selves on screen [272]. Thus, videotaped self-modeling is hypothesized to be an especially effective route to optimizing parenting behaviors.

**Physical and Informational Materials**
Fathers selected novel toys (including a rattle, squeak toy, blocks, and other items) to use during the videotaped play interaction.
Fathers received two handouts that covered various aspects of parent-infant interaction. One handout described infant behavioral cues while the other outlined "the teaching loop," which consisted of a series of steps to engage infants in positive interactions: alerting the baby, showing and explaining, giving the baby time to try, and praising and providing suggestions.

**Setting**
Face to face, in the home

**Dose and Intensity**
Two 60-minute sessions with a home visitor

**Studies that Employed this intervention:** Magill-Evans, Harrison, Benzies, Gierl and Kimak [37]

#### Video Intervention Project (VIP)

All information about the intervention was obtained from Mendelsohn et al. (2007) and Cates et al. (2018)[30,38] unless otherwise stated.

**Intervention Description**
At each session, parent-child dyads were video-recorded for approximately 5 minutes while they interacted with a developmentally appropriate toy and/or book provided by the program. These recorded interactions were reviewed by the interventionist and the parent together, while the interventionist indicated instances of positive parenting behaviors during the interaction (ex. responding to vocalizations, engaging in conversation), to reinforce these behaviors and promote self-reflection on the part of the parent. Parents are also given pamphlets that provide

suggestions for interactions in the contexts of play, shared reading, and everyday routines, and also encouraged to develop plans for interactions to promote their child's development. The video and learning material used in the interaction were both given to the parents to take home.
Sessions are facilitated by an interventionist, who meets one on one with families, providing an individualized, relationship-based intervention.

**Target Population**
Low SES mother-infant dyads.

**Goal (and framework when applicable)**
The goal of the intervention was to enhance parent-child interaction in order to improve language, cognitive and social-emotional development and ultimately promote school readiness and school performance. The intervention was built on the principles of Reach Out and Read.

**Physical and Informational Materials**
To promote the generalization of positive parenting behaviors in the home, the video is given to the parent to take home, along with the learning material used in the interaction. Parents are also given pamphlets that provide suggestions for interactions in the contexts of play, shared reading, and everyday routines

**Setting**
Face to face, individually, in a pediatric primary care setting.

**Dose and Intensity**
Up to 12, 30- to 45-minute sessions that take place primarily on the day of primary care visits [38] or up to 15, 25- to 30-minute sessions [30]. The intervention began at birth [30] or at the first well-child visit [38] and continued until the child was 3 years of age.

**Studies that Employed this intervention:** Cates, Weisleder, Berkule Johnson, Seery, Canfield, Huberman, Dreyer and Mendelsohn [30,]Mendelsohn, Valdez, Flynn, Foley, Berkule, Tomopoulos, Fierman, Tineo and Dreyer [38]

#### Promoting First Relationships (PFR) Intervention

All information about the intervention was obtained from Oxford et al. (2021)[39] unless otherwise stated.

**Intervention Description**
Interventionists (master's-prepared mental health professionals) delivered instructional content and activities based on the Promoting First Relationships (PFR) manual. Each individual mother had the pace of delivery of components specifically tailored for them, and the content was adapted for infants. One component was a videotaped interaction between the caregiver and the infant. Every other week, the intervention session would focus on video-feedback of a mother-infant interaction. This feedback mainly focused on identifying the mother's strengths as a parent and her interpretation of the infant's cues.

**Target Population**
Mothers with psychopathology and their infants.

**Goal (and framework when applicable)**
This intervention was related normalization of stress response systems in children [273,274] as well as reduced child sleep problems reported by caregivers, mediated in part by reduced separation distress between child and caregiver [275] or by adversity buffering [276].

**Physical and Informational Materials**
Informational materials and a videotape of parent-infant interactions

**Setting**
Face to face, individually, at home.

**Dose and Intensity**

9-10 sessions, lasting between 60 and 75 minutes. Most intervention group participants received a full dose of the intervention, 82% completed 9–10 sessions, 9% completed 5–8 sessions, and 8% completed 1–4 sessions. A "full dose" took approximately 9-12.5 hours in total.
The intervention began at postnatal week 8-12 and continued for 10 weeks.

**Studies that Employed this intervention:** Oxford, Hash, Lohr, Bleil, Fleming, Unutzer and Spieker [39]

#### Circle of Security-Intensive (COS-I)

All information about the intervention was obtained from Ramsauer et al. (2019)[178] unless otherwise stated.

**Intervention Description**
The Circle of Security-Intensive (COS-I) intervention involved preparing individualized protocols for each mother-infant dyad. Each unique protocol was based on their previously recorded interactions and took the infant's attachment and autonomy into consideration. COS-I occurred in 3 phases. In the first phase, participants were presented with information on attachment and human development. The second phase was about self-reflection and discussion based on video recordings. In the third phase, mothers were shown the positive changes they made and the difficulties that continued to persist. At the end of the intervention, participants were awarded a COS-I certificate.

**Target population**
Mothers with postpartum depression and their infants.

**Goal (and framework when applicable)**
The purpose of COS-I was to promote secure infant attachment and maternal sensitivity in mothers with postpartum depression. The goal was to determine whether a mother's unresolved attachment status affects the intervention. COS-I is based on attachment theory.

**Physical and Informational Materials**
A COS DVD and handouts were used to present theoretical and empirical findings about attachment and human development. Interventionists introduced the "shark music" concept, a metaphor used to make the mother aware of potentially negative or threatening emotions and cognitions.

**Setting**
COS-I took place in a video laboratory.

**Dose and Intensity**
The COS-I intervention took place over 20 sessions. Each session was 90 minutes long.
COS-I began when infants were aged 4-9 months (6.03 months, average) and continued for 20 weeks.

**Studies that Employed this intervention**: Ramsauer, Muhlhan, Lotzin, Achtergarde, Mueller, Krink, Tharner, Becker-Stoll, Nolte and Romer [178]

#### Smart Beginnings

All information about the intervention was obtained from Roby et al. (2021)[41] unless otherwise stated.

**Intervention Description**
The Smart Beginnings (SB) intervention combines the Video Intervention Project (VIP) and the Family Check-Up (FCU). Families begin receiving the VIP portion of the intervention at birth, but they do not start receiving the FCU portion until 6 months postpartum.
VIP sessions consisted of a discussion of the child's development, using developmentally appropriate learning materials (books, toys) chosen to engage and improve parent-child interactions. The intervention coach would record the parent-child interaction and immediately review the video with the parent to identify and reinforce strength in the interaction and promote self-reflection. The parent is given a copy of the video as well as a pamphlet with information about developmental milestones, suggestions for engaging the child, and parental goals for interacting with the child at home.

The Family Check-Up (FCU) is an evidence-based home-visiting model that seeks to reduce the development of early disruptive behavior and to help parents engage with services that improve parenting practices.

**Goal (and framework when applicable)**
The general goal of the intervention is to positively influence parent-child activities and interactions. The specific goal of the VIP aspect of the SB intervention is to maximize maternal identification, engagement, and retention while minimizing costs.

**Physical and Informational Materials**
Mothers were given book or toy as a part of the VIP intervention. Mothers also received a personalized pamphlet that includes information about developmental milestones, age- specific suggestions for engaging with their child, and the parent's goals for interacting with their child at home.

**Setting**
Sessions were conducted individually.
FCU was provided in the home and/or in a pediatric primary care setting.

**Dose and Intensity**
14 sessions of 25-30 minutes (5.83-7 hours total).
The intervention spanned from birth to 3 years of life.

**Studies that Employed this intervention:** Roby, Miller, Shaw, Morris, Gill, Bogen, Rosas, Canfield, Hails, Wippick, Honoroff, Cates, Weisleder, Chadwick, Raak and Mendelsohn [41]

#### Video-Feedback Interactional Treatment

All information about the intervention was obtained from Stein et al. (2006)[43] unless otherwise stated.

**Intervention Description**
This intervention used mother- and infant- targeted sessions as well as video-feedback to attempt to improve mother-infant interaction, specifically surrounding mealtime. Treatment consisted of three stages. The first stage concentrated on the infant's signals, the second stage focused on the mother's perspective, and the third stage integrated videotapes to help the mother identify and address potential triggers of mealtime conflict. During this stage, the therapist videotaped the mother and infant in the home during mealtimes. Then, therapist and mother would watch and discuss clips of the video to identify the infant's signals and exploration. The goal of this is to refine mothers' observational skills. Seven videotapes are used in total.
In the current study, this intervention was combined with guided cognitive behavior self-help for eating disorders, which prior research has shown to be useful in a primary care setting. It was adapted to the postnatal period and administered to mothers during half of the first eight treatment sessions.
Interventionists were therapists who had prior experience in child and family mental health care and undertook project and intervention model-specific training.
Weekly team meetings, supervised by psychiatrists were held to discuss the progress of therapy and to re-view videotapes and samples of tape recordings of the treatment sessions to ensure consistency and compliance with the protocol.

**Target Population**
The target population was women who met the DSM-IV diagnostic criteria for an eating disorder, either bulimia nervosa or another eating disorder with similar clinical severity.

**Goal (and framework when applicable)**
The goal of the video-feedback treatment was to prevent or reduce mother-infant conflict and enhance mother-child interaction, specifically during mealtimes, by facilitating maternal recognition of and responsiveness to her infant's cues and by improving her awareness of the infant's developing skills and needs.
The goal of the self-help treatment was to help mothers regain control over their eating, reduce vomiting and laxative use, and reduce extreme concerns about shape and weight.

**Physical and Informational Materials**

As a part of the self-help treatment, mothers in both groups received a self-help manual that contained information about eating programs that were tailored to the postnatal period.

**Setting**
In the mother's home.

**Dose and Intensity**
Mothers completed thirteen 1-hour treatment sessions, beginning when the infants were between 4 and 6-months old, and completed by the time the infants were 12 months old.

**Studies that Employed this intervention:** Stein, Woolley, Senior, Hertzmann, Lovel, Lee, Cooper, Wheatcroft, Challacombe, Patel, Nicol-Harper, Menzes, Schmidt, Juszczak and Fairburn [43]

#### Video-Feedback Therapy (VFT) + Cognitive Behavioral Therapy (CBT)

All information about the intervention was obtained from Stein et al. (2018)[44] unless otherwise stated.

**Intervention Description**
This intervention addresses how providing Video-Feedback Therapy (VFT) in conjunction with Cognitive Behavioral Therapy (CBT) can lead to improved cognitive, language, behavioral, and attachment outcomes in children of mothers with postnatal depression. Mothers partake in therapy sessions with a single therapist. Each session alternates between CBT and VFT.

**Target population**
Women with postnatal depression and their infants.

**Goal (and framework when applicable)**
The aim of this intervention is to target parental behaviors associated with postnatal depression that may lead to adverse child outcomes. The aim of the video-feedback therapy portion of the intervention is to improve mother-child interaction by enhancing maternal attention to the child's cues, maternal warmth and support, and maternal sensitivity in the context of the child's attachment needs.
This intervention is based on the CBT model, specifically focusing on behavioral interaction. This intervention uses CBT to attend to common symptoms of postnatal depression.

**Physical and Informational Materials**
Not reported.

**Setting**
Therapy sessions take place in the mother's home (or at a mother's request, may take place at an alternative location like the Children's Centre).

**Dose and Intensity**
Mothers participated in 13 therapy sessions throughout the duration of the intervention. Each session is approximately 90 minutes. The intervention begins when the child is between 6-8 months old. The first session is CBT, the second is VFT. The following sessions are equally divided between CBT and VFT, donating 45 minutes to each therapy per session. The first 6 sessions occur weekly. The next 5 sessions occur every 2 weeks. There are 2 final booster sessions between 6-10 months after the end of the therapy.

**Studies that Employed this intervention:** Stein, Netsi, Lawrence, Granger, Kempton, Craske, Nickless, Mollison, Stewart, Rapa, West, Scerif, Cooper and Murray [44]

#### Compétences parentales et Attachement dans la Petite Enfance: Diminutiondes risques liés aux troubles de santé mentale et Promotion de la resilience project (CAPEDP); CAPEDP Attachment Study (CAPEDP-A)

All information about the intervention was obtained from Tereno et al. (2017)[45] unless otherwise stated.

**Intervention Description**
CAPEDP uses a blended intervention model that combines home visits of maternal sensitivity training through video-feedback techniques. Home visits take place during pregnancy and continue through the child's second year of life. These visits pertain to routine prenatal and well-baby care, familial involvement, and health education counseling. Video-feedback is used to address disrupted parenting behaviors. CAPEDP-A uses the same intervention model but is specifically designed to investigate attachment outcomes. In these interventions, everyday mother-child interactions are filmed, then the videos are watched and discussed with a psychologist.

**Target population**
Low SES and first-time mothers and their infants.

**Goal (and framework when applicable)**
The aim of CAPEDP is to improve developmental and attachment outcomes in impoverished families. The goal is to promote the development of secure attachment while simultaneously reducing family stress, disorganized infant attachment, and infant mental health issues. The aim of CAPEDP-A is to further assess the impact of the intervention on infant attachment and maternal communication. This intervention is based on attachment theory.

**Physical and Informational Materials**
Families were given information brochures. They are also provided with a series of six DVDs that include short films on pregnancy, childcare, and child development. A comprehensive document is shared that details infant emotional development and mother–child attachment quality.

**Setting**
Visits took place at the participant's home.

**Dose and Intensity**
CAPEDP(-A) occurs in 44 sessions, with each session lasting approximately 60 minutes. The intervention begins during the third trimester, when the mother is about 27 weeks pregnant, and continues until the child is 2 years old.

**Studies that Employed this intervention:** Tereno, Madigan, Lyons-Ruth, Plamondon, Atkinson, Guedeney, Greacen, Dugravier, Saias and Guedeney [45]

#### Video-Feedback Intervention

All information about the intervention was obtained from Tryphonopoulos and Letourneau (2020)[46] unless otherwise stated.

**Intervention Description**
This program follows a seven-step protocol that focuses on maternal sensitivity and responsiveness to promote "serve and return" interactions between mothers and infants (e.g., baby smiles, mom smiles back), which are foundational to an infant's healthy brain development.
The intervention uses the Nursing Child Assessment Teaching Scale, a structure that provided developmentally appropriate teaching tasks for mothers to complete with their infants [271].
The intervention also contained a video-feedback protocol [277]. Before the intervention, mothers were provided with a "Children's Activities Card," which was a list of activities that increased in difficulty (ex: holding a rattle, tying a shoelace). Mothers were asked to select the first task on the list that the infant could not perform.
Then, mother-infant interactions were observed and video-recorded in a 5-minute, uninterrupted episode. Interventionists reviewed the video with the mother multiple times (2-3 times), with opportunities for replaying and slowly reviewing selected portions. The interventionist used praise to reinforce desired maternal behaviors. A specific example of feedback from the interventionist is: "That's great the way you play and talk with him until he turns his head away to let you know he needs a break."
Interventionists were nurses. An intervention check list was used for each session to ensure implementation fidelity.

**Target population**
Mothers with postpartum depression and their infants.

**Goal (and framework when applicable)**

This Video Feedback intervention uses the Barnard Model as a theoretical framework for understanding how caregiver sensitivity and other elements are needed for optimal maternal-infant interactions to occur [278].
The Barnard Model characterizes optimal maternal–infant interaction as mutually adaptive and dependent upon both caregivers and infants fulfilling certain responsibilities. For example, parents must demonstrate affectionate caregiving by being sensitive and responsive to the infant's needs, and infants must provide clear cues so that caregivers can respond appropriately. Contingency (i.e., responding to infant's smile with a smile) is also central to Barnard's Model since infants develop a sense of self-efficacy when caregivers respond reciprocally to their behavior.

**Physical and Informational Materials**
Mothers were provided with a Child Activities Card, which is a list of developmentally appropriate teaching tasks that increase in difficulty. They were also given an intervention checklist to ensure they remain consistent with the implementation of the intervention.

**Setting**
Home visits that were offered face-to-face individually with nurse-interventionist.

**Dose and Intensity**
The intervention consists of 3 video-feedback sessions conducted at three-week intervals. Each session is between 60-90 minutes (mean = 74.79, SD = 7.52), for a total of 3-4.5 hours. The first session occurs when the infant is between 4-9 months old. The intervention lasts for 6 weeks.

**Studies that Employed this intervention**: Tryphonopoulos and Letourneau [46]

#### Preterm Infant–Parent Program for Attachment (PIPPA)

All information about the intervention was obtained from Twohig et al. (2021)[47] unless otherwise stated.

**Intervention Description**
The intervention was delivered by a child psychiatrist clinician trained in VIG.
Session One: Semi-Structured, Reflective Interview of the mother.
The questions integrated the Working Model of the Child Interview (WMCI), asking about pregnancy, birth, the mother's sense the infant's personality, as well as the developing mother-infant relationship.
Session Two: Infant Observation and Parent Discussion
A video of approximately five minutes of the caregivers interacting with the infant was made. After watching the video, there was a supervised discussion between the caregivers and the facilitator, focusing on the parent's observation of the baby.
Session Three: Shared Review of Edited and Prepared Video
During this session, the video was edited to exhibit optimal parent–infant interactions and shown to the parents.

**Goal (and framework when applicable)**
This intervention followed attachment and VIG theories. The goal of this intervention was to enhance caregiver sensitivity and reduce infant socioemotional issues.

**Physical and Informational Materials**
Not reported.

**Setting**
Face to face, individually.

**Dose and Intensity**
This intervention took approximately 3 total hours, first session: 45 minutes-1.5 hours, second session: ~50 minutes, third session: 50 minutes-1.5 hours.
This intervention began the third or fourth week after birth, with the timing of the sessions varying based on parent availability and infant well-being.

**Studies that Employed this intervention:** Twohig, Murphy, McCarthy, Segurado, Underdown, Smyke, McNicholas and Molloy [47]

#### Parent-Baby Intervention

All information about the intervention was obtained from van Doesum et al. (2008) and Kersten-Alvarez et al. (2010)[48,49] unless otherwise stated.

**Intervention Description**
This home-based intervention was used to prevent adverse outcomes by improving the quality of maternal-infant interactions using video-feedback is used.

**Target population**
Mothers with postpartum depression and their infants.

**Goal (and framework when applicable)**
The intervention aims to enhance the quality of mother-child interactions by using video-feedback methods to enhance maternal sensitivity. The intention is that this intervention will show lasting effects by the time the child is 5 years old.

**Physical and Informational Materials**
Not reported.

**Setting**
Visits occur at the mother's home.

**Dose and Intensity**
The intervention occurs over a 3–4-month period during which mothers receive 8-10 home visits from a prevention specialist. Each visit lasts 60-90 minutes. The intervention begins when the child is between 1-12 months, with an average of 6 months old. Visits initially occur weekly, but then taper to once every other week. There is a follow-up visit 3 months after the program is complete.

**Studies that Employed this intervention**: van Doesum, Riksen-Walraven, Hosman and Hoefnagels [48,]Kersten-Alvarez, Hosman, Riksen-Walraven, Van Doesum and Hoefnagels [49]

#### Family Nurture Intervention

All information about the intervention was obtained from Hane et al. (2018) and Beebe et al. (2018)[50,51] unless otherwise stated.

**Intervention Description**
The Family Nurture Intervention (FNI) focused on improving the emotional connection between mothers and infants through regular calming sessions while infants were in the NICU. The intervention was delivered by nurture specialists, who were former NICU registered nurses. Sessions involved scent cloth exchange, maternal vocalizations, sustained eye gaze, frequent skin-to-skin and clothed holding, and family-based support sessions to develop emotional connection. Mothers also had access to full-time feeding specialists to learn about infant nutrition and how to use a breast pump. Intervention activities, length of sessions, and number of sessions were recorded by nurture specialists. Mothers used self-report logs to self-report intervention activities practiced outside of nurture specialist sessions.

**Target population**
Preterm mother-infant dyads.

**Goal (and framework when applicable)**
The goal of FNI is to improve emotional connection, caregiving behaviors, and family support by focusing on family cooperation and function. It also aims to establish a Calming Cycle routine.

**Physical and Informational Materials**
Mothers were given two, 5x7 scent-cloths that they exchanged with their infants [51]. Mothers were also given nurture logs to keep track of activities that occurred outside of nurture specialist sessions [50].

**Setting**
Face-to-face, individually in the NICU

**Dose and Intensity**
Visits began as soon as possible after delivery (mean = 7 days) and continued until discharge from the NICU [50]. Intervention length and quantity varied between families depending on their individual needs and availability. Both treatment and control mothers met with researchers at least 4 times a week to fill out questionnaires. Mothers participated in calming sessions an average of 3.5 times/week (median = 3.7; IQR = 2.7 - 4.1). The mean length of each session was 1.6 hours, for a total average of 6.4 hours/week. Intervention and control mothers visited the NICU an average of 4.0 ± .3 and 3.7 ± .3times per week, respectively.  Prior to discharge, FNI families met with the PI (a family psychiatrist) for one session to discuss a post-discharge support plan.

**Studies that Employed this intervention**: Hane, Myers, Hofer, Ludwig, Halperin, Austin, Glickstein and Welch [50,]Beebe, Myers, Lee, Lange, Ewing, Rubinchik, Andrews, Austin, Hane, Margolis, Hofer, Ludwig and Welch [51]

#### Sugira Muryango + Classic Public Works (cPW) and/or expanded public works (ePW)

All information about the intervention was obtained from Betancourt et al. (2020)[54] unless otherwise stated.

**Intervention Description**
The intervention was organized into 12 modules. Sessions included a 15-minute "active play and communication" session where caregivers received live feedback while interacting with their child. Throughout the intervention, CBCs offered active coaching aimed at promoting early stimulation, play, nutrition, hygiene, responsive parenting, nonviolent interactions, and engagement of female and male caregivers.
Both the Sugira Muryango group and the control group received access to a classic public works (cPW) and / or expanded public works (ePW) program. This program, entitled Vision 2020 Umurenge Program (VUP), targets nutrition and early child development through cash transfers, nutrition-sensitive direct support, and public works program.
The intervention was delivered by community-based coaches (CBCs) who were selected from the community.

**Target population**
Vulnerable families with children ages birth-36 months classified as Ubudehe 1 (most extreme level of poverty in the Rwandan government's household-ranking system)

**Goal (and framework when applicable)**
The goal of the Sugira Muryango (strengthen the family) program was to reduce family violence and increase paternal engagement in play and caregiving.

**Physical and Informational Materials**
Not reported.

**Setting**
The intervention was offered face-to-face, individually, in the home

**Dose and Intensity**
12 weekly 90-minute sessions. The estimated total time is 1080 minutes.
Babies were between 6 and 36 months old at baseline. The intervention lasted 3-4 months.

**Studies that Employed this intervention**: Betancourt, Jensen, Barnhart, Brennan, Murray, Yousafzai, Farrar, Godfroid, Bazubagira, Rawlings, Wilson, Sezibera and Kamurase [54]

#### Demonstration and Interaction (Assessment of Preterm Infant Behavior (APIB)

All information about the intervention was obtained from Browne and Talmi (2005)[55] unless otherwise stated.

**Intervention Description**
During the Assessment of Preterm Infant Behavior (APIB) intervention, an examiner observes and analyzes infant behavioral response to environmental stimuli, and then works with the mother to discuss techniques to help the infant develop self-regulatory behaviors. The infant's behaviors were quantified for the following systems: physiologic, motor, regulatory, attention-interaction, sleep and wake states, and amount of examiner facilitation. The examiner discussed the infant's behavioral response with the mother both during and after the examination. After each session, each mother took the Mother's Assessment of the Behavior of her Infant (MABI), which was encouraged to be used to observe and elicit specific behaviors from their infant. The intervention occurred 1 week prior to NICU discharge, and only infants who were expected to stay at least 2 weeks in the NICU were included in the study.

**Target population**
Parents from economically disadvantaged households with preterm infants

**Goal (and framework when applicable)**
The Assessment of Preterm Infant Behavior (APIB) is a refined version of the Brazelton Neonatal Behavioral Assessment Scale (BNBAS), and the goal is to help mothers observe and elicit specific optimal behaviors from their infant.

**Physical and Informational Materials**
Mother's Assessment of the Behavior of her Infant (MABI) was conducted after the APIB demonstration.

**Setting**
Face-to-face, one-on-one, in hospital

**Dose and Intensity**
The intervention was one 45-min session that occurred 1 week prior to NICU discharge.

**Studies that Employed this intervention**: Browne and Talmi [55]

#### Home Visitation and Group Intervention

All information about the intervention was obtained from Constantino et al. (2001)[56] unless otherwise stated.

**Intervention Description**
The intervention was a 10-session curriculum that aimed to increase parental engagement in home visiting programs and improve parent-infant interactions. Each session focused on a theme related to relational health development. Each session consisted of 1) 5-10 minutes free-play and an introduction of the session's topic, 2) 40-minutes of guided mentorship of play techniques, 3) viewing a parenting videotape followed by discussion, and 4) a 20-minute journaling session. During the last intervention session, parents were encouraged to join home visitation programs. The intervention was delivered by "mentors" who were parents with master's experience in early childhood education.

**Target population**
Parents and infants (no risk factor).

**Goal (and framework when applicable)**
Home visitation programs are public health interventions intended to reduce child abuse and neglect. The intervention on Cohort 1 aimed to increase enrollment in home visitation programs and consisted of a curriculum that primarily aimed to educate parents their role in their infant's development, guide parent-infant play, and provide participants a support network of other parents.

**Physical and Informational Materials**
A short reading and a parenting videotape were shared and discussed with participants.

**Setting**
Each session consisted of 8-10 parent-child dyads and two mentors who conducted the session.

**Dose and Intensity**
The 10-session curriculum occurred over 10 to 20 weeks, and each session was approximately 60 minutes. Infants were 3-18 months old (8.3 average, SD=5.7) at baseline.

**Studies that Employed this intervention**: Constantino, Hashemib, Solisb, Alonb, Haleyb, McClureb, Nordlichtb, Constantinob, Elmenb and Carlsonc [56]

#### Improving mother–infant interaction during infant feeding

All information about the intervention was obtained from Cooper et al. (2009) and Tomlinson et al. (2020)[57,58] unless otherwise stated.

**Intervention Description**
This intervention provided women with parenting guidance and support from a community worker. The intervention used items from the neonatal behavioral assessment schedule to sensitize mothers to infant's capacities and needs. Interventionists were four residents (all women) of Khayelitsha, selected by the local community counsel. Interventionists were supervised by an experienced community clinical psychologist.

**Target Population**
Low SES mother-infant dyads.

**Goal (and framework when applicable)**
The aim of the intervention was to encourage the mother to improve maternal sensitivity during mother-infant interactions.

**Physical and Informational Materials**
Items from the neonatal behavioral assessment scale were used to sensitize the mothers to their infants' needs. The intervention was adapted from the Social Baby manual (available at www.reading.ac.uk/psychology/research/child-development/clinical-subgroup.asp )

**Setting**
Face to face, individually, at home.

**Dose and Intensity**
Mothers were visited twice prenatally and then once a week for the first eight weeks postpartum. Mothers were then visited every two weeks for the following two months. Mothers were then visited monthly for two more months.

There were 16 sessions total, spanning from the prenatal period to five months postpartum.
Each session was approximately 1 hour long.

**Studies that Employed this intervention** Cooper, Tomlinson, Swartz, Landman, Molteno, Stein, McPherson and Murray [57,]Tomlinson, Rabie, Skeen, Hunt, Murray and Cooper [58]

#### Index (R-HV)

All information about the intervention was obtained from Cooper et al. (2015)[59] unless otherwise stated.

**Intervention Description**
The intervention consisted of three components.
1. Supportive counseling [279]. The goal of this element of the intervention was to encourage the mothers to express their feelings in a non-judgmental and supportive environment.
2. Specific strategies were employed to make mothers more sensitive to their infants' characteristics. This element used items from the Brazelton Neonatal Behavioral Assessment Scale (NBAS), [267])

3. Interventionists provided specific, targeted assistance to mothers to help them manage infant behavioral problems (ex: sleeping feeding crying, outlined in The Social Baby) [280].

Therapists were NHS-employed health visitors who received training in the administration of the NBAS.

**Target population**
Mothers at risk of postnatal depression and their infants.

**Goal (and framework when applicable)**
Not reported

**Physical and Informational Materials**
Interventionists worked with an interactive neonatal assessment, based on the Brazelton Neonatal Behavioral Assessment Scale, which focused on the infant's responsiveness to the social and non-social environment (visual tracking, responding to the mother's voice), as well as individual differences in infant capacities for regulating their state and behavioral responses (ex: via habituation and covering the infant's eyes briefly with a soft cloth).

**Setting**
Face-to-face individually, in the home

**Dose and Intensity**
The intervention consisted of 11 home visits; 2 prenatal visits and 9 in the first 16 weeks postpartum.

**Studies that Employed this intervention**: Cooper, De Pascalis, Woolgar, Romaniuk and Murray [59]

#### Parent Baby Interaction Program (PBIP)

All information about the intervention was obtained from Glazebrook et al. (2007)[60] unless otherwise stated.

**Intervention Description**
The Parent Baby Interaction Program consisted of four types of activities, as well as principles of developmental care, such as using incubator covers to shield infants.

1. Tactile (ex: stroking infant)
2. Discussion (ex: infant development)
3. Verbal (ex: greeting infant)
4. Observation (ex: identifying different infant states)

The intervention is delivered by neonatal nurses who are overseen by a senior neonatal nurse.

**Target population**
Very premature infants (<32 weeks) and their parents.

**Goal (and framework when applicable)**
The goal of the PBIP program is to enhance parents' understanding of and sensitivity to infant cues.

**Physical and Informational Materials**
A PBIP program manual, written by a senior neonatal nurse.

**Setting**
Face to face, individually. They started in the neonatal care unit and there was an option to continue intervention in the home for up to 6 weeks after discharge.

**Dose and Intensity**
The intervention began in the NICU and consisted of weekly, 60-minute sessions that could continue for up to 6 weeks after discharge.
The total number of sessions was determined by the length of time needed to deliver the program and the mother's availability.
The mean number of sessions was 8.04 (SD = 4.34). The estimated average total number of minutes is 482.

**Studies that Employed this intervention**: Glazebrook, Marlow, Israel, Croudace, Johnson, White and Whitelaw [60]

#### Parent Participation Improvement Program

All information about the intervention was obtained from Heo and Oh (2019)[61] unless otherwise stated.

**Intervention Description**
This was a multi-stage intervention delivered by NICU nurses. The stages progressed as follows:

1. Individualized Interaction: parents identified the factors that impeded their experience of parenting in the NICU
    a. Answers were used to establish mutual goals and promote parent participation in areas such as feeding, bathing, clothing, holding, and developing an awareness of infant signals.
    b. There were individual variations in the quantity and content of goals
    c. Mothers and fathers established separate goals
2. Pre-Participation stage: provided parents with information on selected topics based on interviews from the individualized interaction stage and practical exercises
    a. Also involved learning about the ecology of the NICU, infants' signals, and preterm infants' sleep cycles
3. Active Participation stage: premature infants' parents engaged in nursing care a total of 6 times
    a. Took place during regular handling time for premature infants
    b. Based on education and practice in pre-participation stage
    c. Activities included diaper changes, breastfeeding, soothing, kangaroo care, bathing, clothing, developmental positioning, singing, and talking.

**Target population**
Parents and their preterm infants.

**Goal (and framework when applicable)**
The intervention was based on King's goal attainment theory and interpersonal system of action, reaction, perception, and judgment. In-depth interviews with parents that aimed to understand their participation needs and to select possible activities for their participation in neonatal care were used to design the intervention.

**Physical and Informational Materials**
Informational materials were printed in a booklet and provided to parents.

**Setting**
The intervention takes place face to face, individually, in an education room in the NICU

**Dose and Intensity**
The first visit was an individualized interaction stage; the duration of this session was not reported. 9 intervention sessions followed this initial interaction. The first 3 sessions constituted the pre-participation stage. Each of these sessions lasted 50-60 minutes. There were 6 sessions in the active stage, each lasted 50-60 minutes. The estimated total time is 495 minutes.
At baseline, infants were (on average), 33.6 weeks corrected gestational age.
The intervention was delivered over the course of 2 weeks.

**Studies that Employed this intervention**: Heo and Oh [61]

#### My Baby's First Teacher (MBFT)

All information about the intervention was obtained from Herbers et al. (2020)[62] unless otherwise stated.

**Intervention Description**
The intervention was guided by video modeling that showed pre-recorded videos of families in shelters demonstrating skills and appropriate parent-infant interactions. Specific skills included sensitively responding to infant signals and cues, placing infants on their stomachs for active play to develop their core muscles, and infant

massage to encourage soothing touch and affection. These videos included pauses during which facilitators could break for coaching and practice. The staff would coach parents, and would encourage parents to coach and support one another in the skills that were taught in the videos

**Target population**
Parents with infants staying in emergency homeless shelters.

**Goal (and framework when applicable)**
The goal of the course is to provide parents with the skills to enhance their relationships with their infants. Specifically, the intervention seeks to emphasize the links between sensory experiences, brain development, attachment relationships, developmental milestones, and the importance of healthy early development to prevent later problems.

**Physical and Informational Materials**
Participating parents received gifts at each session, including blankets, age-appropriate toys, and infant carriers.

**Setting**
The intervention was conducted face-to-face in groups. Sessions took place in playrooms in an onsite emergency shelter.

**Dose and Intensity**
There were 5, weekly group sessions that lasted for 60-90 minutes each (total estimated time was approximately 375 minutes).
Infants were 6.43 months old, on average, at baseline.
The intervention lasted for 5 weeks.

**Studies that Employed this intervention**: Herbers, Cutuli, Fugo, Nordeen and Hartman [62]

#### Interaction Coaching for At-Risk Parents (ICAP)

All information about the intervention was obtained from Horowitz et al. (2001)[63] unless otherwise stated.

**Intervention Description**
The ICAP intervention focuses on mother-infant face-to-face interaction. During each visit, the interventionist asks the mother to engage in a 5-minute face-to-face play interaction, after which the interventionist coachs the mother in infant behaviors. The key elements of the intervention include:

1. Teaching the mother to identify the infant's behavioral cues and to tailor her responses to her infant's preferences
2. Guiding the mother to physically position the infant in her line of vision
3. Demonstrating maternal strategies, including the use of pauses, imitation, sequences, and affect, voice, and touch
4. Encouraging practice of suggestions and learning through trial-and-error
5. Using positive reinforcement of sensitive responsiveness
6. Praising success

Elements of the intervention were individually tailored, repeated, and varied, depending on the needs of each dyad. Interventionists were four advanced practice nurses.

**Target population**
Mothers experiencing postpartum depressive symptoms and their infants.

**Goal (and framework when applicable)**
Designed to strengthen the mother-infant relationship.

**Physical and Informational Materials**
Not reported

**Setting**

Face to face, individually, in the patient's home.

**Dose and Intensity**
All women received three home visits when their babies were 4-8 weeks (Time 1), 10-14 weeks (Time 2), and 14-18 weeks old (Time 3). Each interactive coaching session took approximately 15 minutes. The total time was approximately 45 minutes.

**Studies that Employed this intervention**: Horowitz, Bell, Trybulski, Munro, Moser, Hartz, McCordic and Sokol [63]

#### Communicating and Relating Effectively (CARE)

All information about the intervention was obtained from Horowitz et al. (2013)[64] unless otherwise stated.

**Intervention Description**
The Communicating and Relating Effectively (CARE) intervention uses a combination of behavioral coaching and teaching to improve maternal awareness and understanding of infant cues. The intervention is divided into 6 sessions, outlined below.
Session 1: Focuses on infant's cues about their readiness to interact with their environment and their states (including disengagement cues).
Sessions 2-3: Identifies cues demonstrated by the infant during play observation, having mothers observe infant cues, and assigning homework to observe baby's cues and to use Mother's CARE and Child Communication Cue forms.
Sessions 2-5: Reviewing homework, discussing observations, reinforcing knowledge about infant's cues, returning to session 1, and teaching mothers about new, developmentally relevant cues.
The intervention was delivered by a nurse.

**Target population**
Mothers with postpartum depression and their infants.

**Goal (and framework when applicable)**
The goal of the intervention was to promote responsive maternal interaction between depressed mothers and their infants. Specifically, CARE sought to improve mother's abilities to interpret infants' behavioral cues and to respond sensitivity.

**Physical and Informational Materials**
Not reported

**Setting**
Face to face, individually. The study visits were conducted in participants' residences (although women had the option to select an alternate site for sessions).

**Dose and Intensity**
All study participants received a home visit at 6 weeks and 3, 6, and 9 months postpartum.
Dyads in the CARE intervention group also received visits at 2 and 4 months postpartum. These visits lasted for approximately 30-40 minutes.
The intervention began when infants were approximately 6 weeks postpartum. The duration of the intervention was approximately 5 months.

**Studies that Employed this intervention**: Horowitz, Murphy, Gregory, Wojcik, Pulcini and Solon [64]

#### The Getting Ready Intervention + Rural Early Head Start

All information about the intervention was obtained from Knoche et al., (2012)[65] unless otherwise stated.

**Intervention Description**
The intervention was conducted by 64 Early Childhood Professionals (ECPs) who received extensive training before delivering the intervention.

During sessions, ECPs guide moms to interact with their children in warm and responsive ways, to support their children's autonomy, and to participate in children's learning. ECPs also engage parents in collaborative interactions to support learning and development in the home. ECPs used strategies such as modeling, observation, and engaging in mutual goal setting and activity and event planning.
The Getting Ready Intervention was designed to support and enhance the rural Early Head Start program.

**Target population**
Families enrolled in rural Early Head Start

**Goal (and framework when applicable)**
The intervention draws on triadic strategies[281], an "early childhood consultation approach that has been validated with young children with disabilities" [282]. The intervention was also based on collaborative consultation models [283,284]
The goal of this intervention is to improve parental warmth, sensitivity, and engagement in learning interactions with their children. Ultimately, the intervention was designed to create a shared responsibility between parents and interventionists to improve children's developmental success.

**Physical and Informational Materials**
None

**Setting**
Face to face, individually. Monthly group activities were held at the community agency, while individual sessions were held in the home.

**Dose and Intensity**
Weekly home visits lasted for 60-90 minutes as a part of EHS. Families received an average of 45.8 (SD = 28.45) visits over the course of 16 months.
Infants were 11.02 months on average at baseline (SD = 7).

**Studies that Employed this intervention**: Knoche, Sheridan, Clark, Edwards, Marvin, Cline, Cline and Kupzyk [65]

#### Newborn Behavioral Observations (NBO) System

All information about the intervention was obtained from Nugent et al. (2017) and Kristensen et al. (2020)[66,72] unless otherwise stated.

**Intervention Description**
The NBO is an infant-focused, family-centric intervention that consists of 18 behavioral items that describe the newborn's physiological, motor, state, and social capacities over the first 3 months of life. During the intervention, clinicians guide mothers to understand their infant's behavioral responses and to formulate caregiving strategies and handling techniques based upon observations of infant behavior. Types of behavior discussed included motor behaviors, infant sleep behaviors, feeding cues, and skin-to-skin contact. The intervention is "baby-led," meaning that it was shaped by the infant's behavior.
The intervention is led by a clinician.

**Target population**
Mothers of full-term newborns [72].
Mothers and their infants in a community setting [66].

**Goal (and framework when applicable)**
The NBO was based on research with the Neonatal Behavioral Assessment Scale [267,285]. The goal of the intervention was to provide clinicians with a tool that shifted the focus from assessment and diagnosis to observation and relationship building. Another goal of the intervention was to enhance parents' understanding of their infant's cues.

**Physical and Informational Materials**
Sessions begin with a shared observation of the baby's initial state.
Infant sleep was used as an opportunity to discuss the infant's state and caregiving opportunities.

The clinician guides the parents to elicit motor behaviors (ex: grasp, sucking and rooting, and crawling), and once again, they discuss the implication of the responses for touch and skin-to-skin contact, feeding cues, and sleep positions.
The intervention also provides opportunities for face-to-face interaction.
There is an emphasis on the infant's threshold levels of stimulation, and therefore provides opportunities to observe and monitor the infant's autonomic, motor, and (state) behavioral self-regulation.

**Setting**
In the hospital and in the home [72].
Mainly home visits [66].

**Dose and Intensity**
Mothers received routine hospital care and participated in the NBO within 2 days of delivery. Mother-infant dyads participated in a second NBO session in the home at 1 month postpartum. The length of the session ranged from 12-25 minutes, depending on the infant's state [72].
Mothers received routine hospital care and participated in the NBO 3 weeks after delivery. Mother-infant dyads participated in one to three more NBO sessions in the home over the first 3 months of the infant's life. The length of the session ranged from 12-25 minutes, depending on the infant's state [66].

**Studies that Employed this intervention**
Kristensen, Juul and Kronborg [66,]Nugent, Bartlett, Von Ende and Valim [72]

#### Guided participation to infant therapeutic positioning (GP_Posit)

All information about the intervention was obtained from Lavallée et al. (2022)[67] unless otherwise stated.

**Intervention Description**
The intervention used Guided participation theory for mothers to participate in their preterm infant's therapeutic positioning (GP_Posit). The intervention was delivered by neonatal nurses who guided mothers in detecting their infant's behavioral stress and stability cues during caregiving activities. These care activities include diaper changes followed by supine, side-lying, and prone positioning. Activities were introduced in a progressive manner, depending on each mother's confidence level and abilities. The interventionist would introduce additional caregiving activities, such as bottle-feeding or breastfeeding, in accordance with each mother's comfort level. The intervention also included didactic information on how to position preterm infants and infant development.

**Target Population**
Preterm mother-infant dyads.

**Goal (and framework when applicable)**
The intervention was based on Attachment theory, guided participation theory, and Als's Synactive theory of infant development. The goal of the intervention was to refine mothers' sensitivity to their infant's behavioral cues through positioning as a caregiving activity.

**Physical and Informational Materials**
Booklet online modules.

**Setting**
The intervention was conducted face-to-face, individually, in the NICU.

**Dose and Intensity**
Weekly, 30- to 45-minute sessions were conducted from birth to discharge from the NICU. On average, dyads received 4 sessions (2 hours total).

**Studies that Employed this Intervention:** Lavallée, Côté, Luu, Bell, Grou, Blondin and Aita [67]

#### The Infant Behavioral Assessment and Intervention Program (IBAIP)

All information about the intervention was obtained from Meijssen et al. (2010a, 2010b) and Meijssen et al., (2011)[68-70] unless otherwise stated.

**Intervention Description**
The Infant Behavioral Assessment and Intervention Program (IBAIP) provides guidance to parents of pre-term infants to help them support their infant's development of self-regulatory competence via centering parental availability and responsiveness. Interventionists help parents support their infant's self-regulation by helping the infant stabilize themselves when they are focused on an object or altering the brightness of a room when the infant shows behaviors such as turning away or excessive blinking.
The Infant Behavioral Assessment (IBA) is used during intervention sessions as a tool to sensitize parents to their infant's self-regulatory behavior to external stimuli.

**Target population**
Preterm mother-infant dyads.

**Goal (and framework when applicable)**
The IBAIP, based on the Newborn Individual Developmental Care and Assessment Program (NIDCAP), aims to improve infant self-regulatory competence and parenting confidence through positive parent-infant interactions. It relies on ideas that parent availability, attentiveness, and responsiveness strengthen infant competence and development and parenting confidence. It is based on the Newborn Individual Developmental Care and Assessment Program (NIDCAP).

**Physical and Informational Materials**
The mother receives a report of each infant's neurobehavioral and developmental progress and parenting advice after each session.
The Infant Behavioral Assessment (IBA) is used during the intervention as a tool to sensitize parents to their infant's self-regulatory behavior to external stimuli.

**Setting**
NICU (level III) in two hospitals and five city hospitals participated in the study. The intervention consisted of face-to-face (home visits) by pediatric physical therapists trained in IBAIP.

**Dose and Intensity**
The number of visits was individualized and ranged from 6-8 home visits, and each visit lasted approximately one hour. The program ran post-discharge up to 6-8 months of age.

**Studies that Employed this intervention**: Meijssen, Wolf, Koldewijn, Houtzager, van Wassenaer, Tronick, Kok and van Baar [68,]Meijssen, Wolf, van Bakel, Koldewijn, Kok and van Baar [69,]Meijssen, Wolf, Koldewijn, van Wassenaer, Kok and van Baar [70]

#### Creating Opportunities for Parent Empowerment (COPE)

All information about the intervention was obtained from Melnyk et al. (2006)[71] unless otherwise stated.

**Intervention Description**
The COPE intervention is a 4-phase educational-behavioral intervention. Each phase uses audiotapes and written recordings to provide information on the appearance and behavioral characteristics of premature infants and the parent-infant relationship.
Phase I: Parents are provided with a set of parenting activities and were instructed to keep a log of the milestones they experienced in the NICU.
Phase II: Reinforce content of phase I and provided supplemental information on premature infants' behaviors and development. This phase also included activities aimed to help parents identify infants' cues (specifically for stress and alertness).
Phase III: Provide developmentally appropriate information about infant states and information about how to interpret and respond to infant stress cues. This phase was also designed to help prepare mothers for discharge from the NICU.

Phase IV: Provided information about preterm infant development and included activities that foster infants' cognitive development.

**Target population**
Preterm mother-infant dyads.

**Goal (and framework when applicable)**
COPE intervention is based on theories of self-regulation and control. The approach of the intervention involves reinforcing information with behavioral activities. of the intervention include helping parents meet their infants' needs, enhancing the quality of parent-infant interaction, and optimizing their infants' development.

**Physical and Informational Materials**
The intervention was delivered as a series of audiotapes and written information, along with prescribed activities.

**Setting**
The intervention took place individually online and in the NICU.

**Dose and Intensity**
Phases I-III were delivered in the NICU, while Phase IV was delivered in the parents' home 1 week after discharge. Phase I occurred 2 to 4 days after the infants' admission to the NICU. Phase II occurred 2-4 days after the first intervention. Phase III occurred 1 to 4 days before discharge.
The number and length of sessions were not reported.

**Studies that Employed this intervention**: Melnyk, Feinstein, Alpert-Gillis, Fairbanks, Crean, Sinkin, Stone, Small, Tu and Gross [71]

#### Incredible Years Parents and Babies (IYPB)

All information about the intervention was obtained from Pontoppidan et al. (2016)[73] unless otherwise stated.

**Intervention Description**
The Incredible Years Parents and Babies (IYPB) is a therapy intervention performed in groups of 8 families. In each session, group leaders showed American video vignettes of real-life parent-child interactions with Danish subtitles and facilitated group discussion following the video. Group leaders also presented information about infant neural development and parental roles via the Incredible Years baby poster. Participants also received *The Incredible Babies* book, which was translated into Danish and included ways for parents to support infant development.

**Target population**
Mothers and fathers with infants in Ikaste-Brande or Herning local area in Denmark.

**Goal (and framework when applicable)**
The goal of IYPB is to improve physical, language, and emotional development of infants via guiding parents how to respond to their child's needs in a shared learning space and peer-support networks. The course consisted of six sections: "1) Getting to Know Your Baby; 2) Babies as Intelligent Learners; 3) Providing Physical, Tactile and Visual Stimulation; 4) Parents Learning to Read Babies' Minds; 5) Gaining Support; and 6) Babies Emerging Sense of Self."

**Physical and Informational Materials**
Parents watched American IY videos with Danish subtitles that introduced real-life vignette situations of parents and infants. Parents also received a Danish translation of *The Incredible Babies* book which provided parenting advice and a journaling section.

**Setting**
The intervention occurred in a group setting of 8 families and two trained group leaders. The location of group sessions was not specified.

**Dose and Intensity**

Each session was approximately 120 minutes long. Sessions were offered weekly for 8 weeks. For some groups, the weeks were interrupted by holidays or breaks. Other groups finished within 8 continuous weeks. The intervention began after discharge from hospital but before 4 months of age (mean age of 1.59 months) and lasted for 8 weeks.

**Studies that Employed this intervention**: Pontoppidan, Klest and Sandoy [73]

#### Nurture and Play (NaP)

All information about the intervention was obtained from Salo et al. (2019)[75] unless otherwise stated.

**Intervention Description**
Theraplay is a form of child-interactive therapy that is active, adult-led, and playful. The intervention used Theraplay-based activities, including singing, playing musical instruments, and interaction activities such as infant massage and peek-a-boo.
During pregnancy NaP sessions, mothers are encouraged to stimulate the fetus by stroking their bellies while singing lullabies. Pregnancy NaP sessions are also centered around reflection and discussion about maternal representation of childhood, pregnancy, and motherhood.
Baby NaP sessions (postpartum) focused on infant massage, songs that used rhythmic movements, and holding and rocking strategies. Additionally, sessions focused on understanding infants' mental states. Each session includes cognitive and affect regulation techniques with direct attention on handling depressive mood and dysregulated sleep and eating patterns.
Discussions can also focus on coping strategies for handling stress
Homework and diaries are provided to stimulate child-directed thought. Mothers are offered relaxation techniques and massage practices to decrease prenatal depression.
The NaP intervention is delivered by trained instructors.

**Target population**
Prenatally depressed mothers and their infants

**Goal (and framework when applicable)**
The Nurture and Play intervention was based on Cognitive Behavioral Therapy and visualization/mentalization strategies. All Theraplay-based activities were designed to promote affectionate mother-infant contact, using physical touch, reciprocity, synchrony, and joint attention.

**Physical and Informational Materials**
Not reported

**Setting**
The first meeting was individual, followed by group sessions. All sessions were in-person.

**Dose and Intensity**
Pregnancy group sessions were held biweekly, while baby groups were held weekly. Sessions were 1.5 hours. The Baby Group was invited to arrive 15-20 minutes before the session to settle in, feed, and change diapers. Mothers were offered tea and coffee after the sessions for an additional 30-45 minutes.
There were 11 sessions, each approximately 90 minutes.
The intervention began prenatally and spanned 9 weeks.

**Studies that Employed this intervention**: Salo, Flykt, Makela, Biringen, Kalland, Pajulo and Punamaki [75]

#### Home Intervention Adapted from the Infant Health and Development Program (IHDP)

All information about the intervention was obtained from Schuler et al. (2000, 2002)[76,286] unless otherwise stated.

**Intervention Description**
This home-based program adapted from the IHDP was designed to optimize infant development and consisted of both a parent and an infant component. Specifically, this study amended the home-visiting protocol of the IHDP by adding information about drug use and treatment.

Mothers in the intervention group received weekly home visits during the first 6 months postpartum from one of two full-time lay visitors. The visitors were two-middle-aged African American women with previous experience making home visits and knowledge of the community where the mothers lived. These visitors met with a pediatrician and a psychologist weekly to track the progress of the families and to discuss concerns about the families.
The intervention covers a range of topics, from housing, public assistance programs, partner abuse, and the effects of drug use and drug treatment. Home visits are monitored with personal contract forms that contained information about the content and quality of the visit, including the relationship between the mother and home visitor, the caretaker's concern about the mother, the infant's development, and health education.
The infant-component sought to enhance mother-infant communication and provide a stimulating play environment through games and activities. Home visitors used the HELP at Home: Hawaii Early Learning Profile, a curriculum consisting of 650 developmental skills for children from birth to 35 months [287]. Visitors would teach mothers appropriate ways to play with their infants in order to enhance communication between mothers and infants.

**Target Population**
Mothers with substance use histories and their infants.

**Goal (and framework when applicable)**
The home intervention was developmentally oriented and was based on the program used by the IHDP [288]. The goal of the parent component was to increase maternal empowerment by enhancing the mothers' ability to manage self-identified problems.
The goal of the infant component was to promote infant development by using a program of games and activities. The goal of the parent component was to help mothers use family and social support to manage self-identified problems.

**Physical and Informational Materials**
Not reported.

**Setting**
Visits are conducted in the home.

**Dose and Intensity**
The mean number of visits made during the first 6 months was 8.9 (SD = 5.6, range = 0–23), and the mean length was 30.1 min (SD = 5.8). The estimated total time is 4.46 hours [76].
The average number of visits made to mothers in the intervention group in the first 18 months was 19.9 (SD=13.0, range = 0-57), and the mean length was 28.5 minutes (SD=4.6) [286].

**Studies that Employed this intervention:** Schuler, Nair, Black and Kettinger [76;]Schuler, Nair and Black [286]

#### Baby Triple P

All information about the intervention was obtained from Tsivos et al. (2015)[78] unless otherwise stated.

**Intervention Description**
The 'implementing parenting routines' sessions involve active skills practice between the mother–infant dyads. The interventionist was a triple P-accredited doctoral student who was overseen by a clinical psychologist. The practitioner provided feedback and prompted maternal self-evaluation.
Baby Triple P consists of eight sessions:

1. Positive parenting
2. Responding to your baby
3. Survival skills
4. Partner support
5. Implementing parenting routines (1)
6. Implementing parenting routines (2)
7. Implementing parenting routines (3)
8. Implementing parenting routines (4) and maintenance and closure (see Table 1).

**Target population**
Mothers with postnatal depression and their infants

**Goal (and framework when applicable)**
The Triple P intervention has 3 specific goals[289]:

1. To enhance parental knowledge and resourcefulness
2. To promote nurturing, low-conflict environments for children
3. To promote children's social, emotional, and intellectual competencies through positive parenting practices

The Triple P framework is adaptable to individual parental needs, regardless of age, gender, and socio-cultural differences [290].

**Physical and Informational Materials**
The interventionist used a checklist and workbook that detailed session content [291].

**Setting**
Face-to-face in person, in the home.

**Dose and Intensity**
The intervention consists of 8 weekly sessions. The first 4 sessions lasted 60-90 minutes and the remaining 4 sessions lasted 40-60 minutes.
Infants were 6.7 months on average at baseline, and the intervention lasted 8 weeks.

**Studies that Employed this intervention**: Tsivos, Calam, Sanders and Wittkowski [78]

#### Crianza Temprana

All information about the intervention was obtained from Valades et al. (2021)[79] unless otherwise stated.

**Intervention Description**
This intervention provides basic guidance and support on topics such as mother's support network, birth process, care of the infant, and early childhood development. Mothers are given support in caring for their infants. Mothers are made aware of the infants' social engagement and advised while talking and engaging in play with their infants. The mother's behaviors are reinforced by the facilitator when the infant exhibited a positive reaction, and the facilitators noted how the mother's behaviors enhanced the infant's social engagement. The facilitator also models and guides some behaviors for the mothers when appropriate. Other caregivers are encouraged to be present for the sessions.

**Goal (and framework when applicable)**
The Thula Sana intervention was renamed 'Crianza Temprana,' which means 'Early Parenting' [292]. This intervention was based on parenting principles and early child development strategies outlined in 'The Social Baby' [280]. This intervention also incorporated the key ideas from the World Health Organization's document 'Improving the Psychosocial Development of Children' [293].

**Physical and Informational Materials**
The Neonatal Behavioral Assessment Scale (NBAS) Intervention manual was provided, as well as guidance and support on topics such as creating a support network, birth process, care of the infant, and early childhood development. Mothers were given support caring for their infant and behaviors were sometimes modeled for the caregivers by the facilitator.

**Setting**
Face to face, individual home visits.

**Dose and Intensity**
This intervention included 16 sessions, approximately one hour each. Two of the visits are conducted in the late antenatal period, and the remainder 4 of the visits are conducted weekly for the first eight weeks postnatal. After that, every two weeks for two more months, as well as monthly visits for two more months postpartum (16 sessions, approximately 16 total hours).

**Studies that Employed this intervention:** Valades, Murray, Bozicevic, De Pascalis, Barindelli, Meglioli and Cooper [79]

#### Maternal Support for Becoming a First-time Mother (AMACOMPRI)

All information about the intervention was obtained from Vargas-Porras et al. (2021)[80] unless otherwise stated.

##### Intervention Description

This intervention employs usual postnatal healthcare and the multimodal intervention, "Maternal Support for Becoming a First-time Mother," (the Spanish acronym is AMACOMPRI).
The multimodal intervention aspect includes eight alternating home and telephone sessions, delivered by an AMACOMPRI trained nurse (to ensure consistency and compliancy). The sessions focus on functional social support, the mother-infant bond, perceived maternal self-efficacy, and becoming a mother. The information is provided on software that is downloaded on the participant's phone. The participants could also access the information on their own, the participants could write in the software and ask the nurse for advice and the nurse could respond. A nursing care plan is then developed and implemented, according to the needs of the mother, based on the visits.

##### Goal (and framework when applicable)

This intervention is based on Mercer's "Becoming a Mother Theory." This theory encompasses a new mother's personal growth through the required lifetime commitment of motherhood and the challenges that this brings [294]. The four stages of be-coming a mother according to Mercer (2004) are: "(1) commitment, attachment, and preparation (during pregnancy); (2) acquaintance, learning, and physical restoration (first 2–6 weeks postpartum); (3) moving toward a new normal (2 weeks–4 months); and (4) achievement of maternal identity (around 4 months)"(p. 231). Mercer also acknowledges Bronfenbrenner's Theory (1986), accounting for the three main environments of development: family and friends, community, and society [294,295]. The goal of this intervention was to focus on the family-and-friends environment as well as supporting the mother during the postpartum stages, to enhance early development and parent–infant interaction [296].

##### Physical and Informational Materials

This intervention includes audio-visual educational and support software that was downloaded on the participant's smartphones at the beginning of the study. Included in this software are twenty-six audio guides, conducted by an expert, about specific postpartum issues, as well as four videos of a first-time mother sharing her personal experiences.

##### Setting

This intervention took place individually, face to face, in the participants' homes and over the phone.

##### Dose and Intensity

A total of eight nurse-delivered visits, alternating between 90- to 120-minute in-person visits and 15-minute telephone encounters (four of each type of visit). This intervention took approximately 7 total hours.

**Studies that Employed this intervention:** Vargas-Porras, Roa-Diaz, Hernandez-Hincapie, Ferre-Grau and de Molina-Fernandez [80]

#### Cues

All information about the intervention was obtained from Zelkowitz et al. (2021)[81] unless otherwise stated.

##### Intervention Description

The intervention consists of observational exercises that helped mothers learn about their infant's behavior and apply the knowledge they acquired during the intervention. During each session, the mother and interventionist discuss the content of each booklet and its applications.
Sessions 1 and 2 focus on managing maternal anxiety.

Sessions 3-6 focus on infant behavioral cues, infant states, and sensitive mother-infant interaction. Session 3 focuses on the infant's state, session 4 focuses on infant cues, and session 5 focuses on feeding interactions. Session 6 integrates all earlier sessions.
The intervention was delivered by a nurse, a psychologist, or a nursing or psychology graduate student.

**Target population**
Mothers of VLBW (preterm) infants.

**Goal (and framework when applicable)**
The goals of the intervention were:

1. To reach mothers to recognize signs of their own anxiety and distress
2. To teach mothers to use various strategies to alleviate their distress
3. To teach mothers to read their infant's communication cues
4. To teach mothers to respond sensitively to their infant's cues and distress

**Physical and Informational Materials**
The information disseminated in the anxiety reduction and sensitivity components of the intervention was contained in a series of five "The Keys to Caregiving" booklets.
Each booklet addressed different topics related to parent-infant interaction.

**Setting**
Face-to-face, individually, in the NICU and in the home

**Dose and Intensity**
The intervention consists of 6, 60-minute sessions. The first 5 sessions took place in the NICU, while the last session took place in the home 2-4 weeks after discharge.
The estimated total length of the intervention was 600 minutes.
Infants were 33 days old at baseline (SD = 12). The intervention duration was 9-23 days (until the infant was 6-8 weeks corrected age).

**Studies that Employed this intervention**: Zelkowitz, Feeley, Shrier, Stremler, Westreich, Dunkley, Steele, Rosberger, Lefebvre and Papageorgiou [81]

#### Primeiros Laços

All information about the intervention was obtained from Alarcao et al. (2021)[83] unless otherwise stated.

**Intervention Description**
Primeiros Laços is an individualized home visiting program. During visits, interventionists help parents develop child-centered interactions to improve the parent-infant bond, reflect on their own attachment history and their relationship with their parents, and help parents consider their infants' feelings and thoughts. Interventionists used modeling to encourage attuned, attentive, and sensitive parenting.
Interventionists are nurses specializing in maternal or mental health and were supervised weekly by senior nurses and psychologists.

**Target Population**
Adolescent first-time mothers and their infants.

**Goal (and framework when applicable)**
Primeiros Laços is based on three theoretical frameworks, Attachment Theory, Self-Efficacy Theory, and Bioecological Development Theory. [295,297,298]The intervention was structured in five axes (healthcare, health environment, parenting and attachment, social and family network, life project).
The goal of the intervention is to strengthen maternal capacity for warm and responsive care and to establish positive relationships between home-visitors and the family.

**Physical and Informational Materials**
Not reported.

**Setting**
The intervention was delivered face-to-face, individually, in the mother's home.

**Dose and Intensity**
The intervention began during the first 16 weeks of pregnancy and continued until the child was 24 months.
Visits were weekly (first/last month of pregnancy/puerperium), biweekly (gestation/2-20 months of child's age), and monthly (21-24 months of child's age).
Mothers were expected to receive 40–42 visits by the time their infants were aged 12 months and 60–62 visits by the time their infants were 24 months of age.

**Studies that Employed this intervention:** Alarcao, Shephard, Fatori, Amavel, Chiesa, Fracolli, Matijasevich, Brentani, Nelson, Leckman, Miguel and Polanczyk [83]

#### Healthy Families Durham (HFD)

All information about the intervention was obtained from Berlin et al. (2017)[85] unless otherwise stated.

**Intervention Description**
Interventionists facilitate discussions on topics related to the infant-parent relationship and promoting children's socio-emotional development. Visits are tailored to the child's age and developmental level as well as to the parent's needs. During home visits, parents are provided with handouts related to socio-emotional development and toys and activities designed to promote parent-child interaction.
Interventionists were Healthy Families America home visitors who were mostly social workers and counseling professionals with master's degrees.

**Target population**
High-risk mother–infant dyads.

**Goal (and framework when applicable)**
The intervention is based on attachment and bio-ecological systems theories (ABC) and trauma-informed care. The goal of the intervention was to promote healthy attachment. The intervention is credentialed as a Healthy Families America program and drew from the Parents as Teachers curriculum, a parent education tool that seeks to promote all aspects of child development [299,300].

**Physical and Informational Materials**
Home visitors brought simple toys and activities to each session to try to encourage parent-child interaction. Home visitors also provided parents with handouts (topics included the importance of infant-parent relationships to children's socioemotional development).
The HFD intervention used the Parents as Teachers curriculum.

**Setting**
Face to face, individually, in the home.

**Dose and Intensity**
Visits began prenatally or during the first 3 months of birth. The interventionist continued to visit parents weekly for 1 year, and then continued to meet with families on an as needed basis. The mean number of sessions was 28.69. The length of each session was not reported.

**Studies that Employed this intervention**: Berlin, Martoccio, Appleyard Carmody, Goodman, O'Donnell, Williams, Murphy and Dodge [85]

#### Early Head Start Plus Attachment and Biobehavioral Catch-Up (ABC)

All information about the intervention was obtained from Berlin et al. (2018)[86] unless otherwise stated.

**Intervention Description**

The intervention consists of discussion, guided practice, and watching video clips from previous sessions. Sessions included observation and live feedback from ABC coaches on mother-infant interactions. Mothers are provided homework outside of intervention sessions to reflect on their parenting behaviors, but this homework was not required nor formally evaluated. Families also engaged in Early Head Start interventions.
Interventionists were two female fully bilingual (English and native Spanish) parent coaches who were accredited by ABC/Infant-Caregiver Project staff.

**Target population**
Mothers and infants who were already receiving the early head start program. This included specifically low-income Latino and Spanish speaking mothers.

**Goal (and framework when applicable)**
The goal of the ABC-component of the intervention was to promote the development of a secure mother-infant attachment and improve early childhood behavioral physiological regulation. The intervention targeted three specific aspects of sensitive caregiving behavior:

1. Providing nurturance
2. Following the child's lead with delight
3. Avoiding intrusive and frightening behaviors

**Physical and Informational Materials**
The ABC manual, which was also translated into Spanish.

**Setting**
Face-to-face, individually, in the mother's homes.

**Dose and Intensity**
10 sessions, although the length of each session was not reported. At baseline, infants were within 6-20 months of age (average of 12.7 months, SD=4.2). The intervention was delivered approximately weekly, but the reported average was 13 weeks (SD=6.9).

**Studies that Employed this intervention**: Berlin, Martoccio and Jones Harden [86]

#### Attachment and Biobehavioral Catchup (ABC)

All information about the intervention was obtained from Dozier et al. (2009), Bernard et al., (2012), Bick and Dozier (2013), and Perrone et al., (2021)[87-89,92] unless otherwise stated.

**Intervention Description**
ABC is an attachment-based intervention that seeks to help caregivers interpret children's behaviors and provide nurturing care. The intervention uses interactive parent discussion, videotaped feedback, and dyadic practice to teach and solidify concepts.
Interventionists varied between studies. The intervention was delivered by professional or social workers or psychologists with at least 5 years of clinical experience [89], parent trainers with experience with children and strong interpersonal skills [87], and parent coaches with varying racial, ethnic, professional and educational backgrounds [92].
The intervention is standardized so that the same issues are introduced across the ten sessions, although the age and target population vary among studies. While these themes remained constant, specific activities were tailored to the children's ages and family circumstances.
The focus of each session is outlined below:

1. Sessions 1 and 2: Providing nurturance when the child pushes away
2. Sessions 3, 4, and 5: Practice following the child's lead
3. Sections 6 and 7: providing nurturance at difficult moments for the parent
4. Session 8: physical touch (holding and touching the child)
5. Session 9: reducing parent's frightening behavior
6. Session 10: Learning to respond to the child's negative emotions

**Target population**

This intervention has been used to target foster parents and their infants [88,89], infants and parents in problematic or neglectful environments [87], and parent-infant dyads from urban community settings [92].

**Goal (and framework when applicable)**
ABC is based on Ainsworth's 1976 model of attachment and construct of maternal sensitivity. It uses a multi-pronged approach to improve parental nurturance, particularly in moments when it doesn't come naturally. As a result, the intervention seeks to help children develop regulatory capacities and help attachment organization (to promote secure attachment formation).

**Physical and Informational Materials**
The intervention employed video feedback and manualized information[89]. Mothers were asked to keep an "attachment diary" for 3 days, recording moments when the child pushes away. Video feedback was used to consolidate skills obtained in the sessions, support maternal competence, highlight parental strengths, challenge weaknesses, and celebrate changes in parental behaviors.

**Setting**
The intervention was conducted face-to-face, individually, in the home. Intervention setting included shelters [87] and foster parent homes [88,89].

**Dose and Intensity**
10 sessions, 60 minutes each, 600 minutes total.
Duration of intervention varied: 11.82 months (average), SD=4.36 [92]; 19.2 months (average), SD=5.2 [87]; 18.9 months (average), SE=1.8 [89].
Age at Baseline varied: 5.64-21.48 months (m=11.82, SD=4.36) [92]19.2 (SD=5.2) [87]; not reported [89]; 10 months (average); SD=7.3 [88].

**Studies that Employed this intervention**: Bernard, Dozier, Bick, Lewis-Morrarty, Lindhiem and Carlson [87,]Bick and Dozier [88,]Dozier, Lindhiem, Lewis, Bick, Bernard and Peloso [89,]Perrone, Imrisek, Dash, Rodriguez, Monticciolo and Bernard [92]

#### Parent-Infant Psychotherapy (PIP)

All information about the intervention was obtained from Fonagy et al. (2016) [90] unless otherwise stated.

**Intervention Description**
The intervention consists of parent-infant psychotherapy sessions. Parents, infants, and therapists would sit on the floor and discuss issues such as parental mental feelings and state, the parent-infant relationship, and issues centered around the infant. During these sessions, therapists work to observe interactions in the room, nonverbal communication, and communication errors. The infant is present in each session. The relationship with the therapist was viewed as an important agent of change through the sessions.
Interventionists were 6 parent-infant psychotherapists who had received 2 weeks of group supervision to discuss the intervention and ensure model adherence before the intervention.

**Target population**
Parents with mental health problems and their infants.

**Goal (and framework when applicable)**
It was hypothesized that PIP would improve infant development, parent-infant relation outcomes, and maternal mental health relative to the standard care group [90].

**Physical and Informational Materials**
The intervention model was described in a manual [301].

**Setting**
Face to face, individually. The PIP intervention was offered at three locations that were local and accessible to participating families.

**Dose and Intensity**
The intervention consisted of 16 sessions, offered on a weekly or bimonthly basis. Psychotherapy sessions were continued until therapists and parents mutually agreed upon an end date. The mean number of PIP sessions attended during the 1-year study period was 16 (range= 1-49), 41% of families had completed therapy by the 6-month follow-up. Some families continued to attend PIP sessions after the final 12-month follow-up. The length of each session was not reported. The baseline age of infants in this study was 3.9 months (SD=3.2).

**Studies that Employed this intervention**: Fonagy, Sleed and Baradon [90]

#### Focused Parent-Infant Psychotherapy (fPIP)

All information about the intervention was obtained from Georg et al. (2021)[91] unless otherwise stated.

**Intervention Description**
This intervention focused on strengthening the relationship between the parent and the infant, to have a positive impact on infant development through psychodynamic interventions. The fPIP intervention explored the mother's own attachment experiences and related that to her current relationship with her infant. This was done using a supportive strategy and an expressive strategy during psychotherapy sessions. Both strategies focused on strengthening the ability of the parent to mentalize the child, as the key aspect for change.
Either one or both primary caregivers and their infant attended psychotherapy sessions. Interventionists were postgraduate students with psychoanalytic training.

**Target Population**
Infants with early developmental or health impairments and their caregivers.

**Goal (and framework when applicable)**
The intervention has several goals. Infant-related goals are to reduce sleeping, crying, and feeding problems for infants. Caregiver-related goals include reducing caregiver's depression and psychological distress and to increase caregiver's emotional availability, self-efficacy, and reflective functioning.

**Physical and Informational Materials**
Not reported.

**Setting**
Face to face, individually, at the Department for Family Therapy at Heidelberg University Hospital.

**Dose and Intensity**
This intervention consisted of one double, 90-minute, session and three single, 50-minute sessions. This intervention took approximately 4 total hours.
Infants were 4 to 15 months of age at baseline (mean= 8.84), and the intervention continued for 12 weeks.

**Studies that Employed this intervention:** Georg, Cierpka, Schroder-Pfeifer, Kress and Taubner [91]

#### Minding the Baby

All information about the intervention was obtained from Sadler et al. (2013) and Londono et al. (2022)[93,94] unless otherwise stated.

**Intervention Description**
Minding the Baby is an interdisciplinary intervention program that combines two early intervention approaches: The Nurse-Family Partnership and Infant-Parent Psychotherapy. This program works to promote sensitive and reflective parenting and improve the overall quality of the dyadic attachment relationship. The parent-clinician relationship was the primary means of integrating this change. MTB elaborated on these two existing models, focusing on the following topics:

(1) maternal mentalization (the mother's reflective functioning on the baby's and her own internal experiences)
(2) prenatal and childcare, health, and development (delivered by the NP)

(3) contraceptive counseling (NP)
(4) ongoing assessment of the dyad and their mental health (delivered by the CSW).

Other activities include hands-on play between the dyad, recording and watching videos of play/care routines, and maternal journaling or scrapbooking. The administration of the program is individualized to each family's needs. Families were recruited from prenatal groups at their primary care clinic, allowing for the clinician-parent relationship to strengthen.

Interventionists are nurse practitioners and social workers who alternated visits such that the NP would attend one week and the CSW would lead the following week.

**Target population**
First-time, young mothers and their infants

**Goal (and framework when applicable)**
The goal of MTB is to promote developmental, relational, and health outcomes in first-time, multiethnic young mothers and children. In order to accomplish this, MTB used an interdisciplinary approach by combining Nurse-Family Partnership and Infant-Parent Psychotherapy early intervention models. It is based on Fraiberg's 1980 IPP model as well as Ainsworth's 1976 model of Maternal Sensitivity and Attachment [190,302].

**Physical and Informational Materials**
Control families received monthly informational brochures from Healthy Steps[303] regarding child rearing and health. They were also sent birthday and holiday cards.

**Setting**
Face-to-face home visits by trained social workers and nurse practitioners

**Dose and Intensity**
MTB consists of weekly visits beginning in the late second or early third trimester until the child's first birthday and biweekly visits from the child's first birthday until they were 24 months old. For the child's first birthday, there was a celebratory transition visit; at two years, there was a graduation visit in which both home visitors attended.

Mean home visits per month = 3.4 (SD = 1.5); session length ranged from 45 min to 90 min, and generally averaged one hour.

**Studies that Employed this intervention:** Sadler, Slade, Close, Webb, Simpson, Fennie and Mayes [93], Londono Tobon, Condon, Sadler, Holland, Mayes and Slade [94]

#### Secure Attachment Promotion Program

All information about the intervention was obtained from Santelices et al. (2011)[96] unless otherwise stated.

**Intervention Description**
The intervention consists of prenatal lectures, group discussions, observation, and psychoeducation designed to promote secure mother-infant attachment. During the first phase of the intervention, groups of mothers meet with an interventionist and discussed mother-infant attachment, feelings about pregnancy, representations about motherhood and their own experience as daughters, imaginary vs. real babies, maternity and maternal functions, and the promotion of secure attachment.

The second, postnatal phase of the intervention are comprised of individual sessions. During these sections, interventionists observe mother-infant interactions and provide psychoeducational feedback, including suggestions to improve maternal sensitivity. The interventionists aim to promote secure attachment through these sessions.

Interventionists were psychology graduates.

**Target population**
Pregnant primipara women in Chile

**Goal (and framework when applicable)**
The intervention is grounded in Ainsworth's theory of attachment. The primary goals of the intervention were to promote maternal sensitivity, to change maternal representations of the infant, and to aid in the formation of a secure and healthy bond between mother and infant.

**Physical and Informational Materials**
Researchers worked from an intervention manual that sought to observe the process by which a secure attachment is formed.

**Setting**
All sessions were conducted face-to-face. Phase one consisted of prenatal group sessions of 6 women. Phase two consisted of four individual sessions.
Most sessions were conducted in a health center, but the rest were conducted in the mother's home.

**Dose and Intensity**
There were 10 sessions in total. Phase one of the intervention (conducted during pregnancy) consisted of 6, 2-hour sessions. Phase two, conducted postpartum, consisted of 4, 1-hour sessions. These four postnatal sessions were offered four times during the child's first year of life, when the baby was 1, 3, 6, and 12 months old.

**Studies that Employed this intervention**: Santelices, Guzman, Aracena, Farkas, Armijo, Perez-Salas and Borghini [96]

#### New Beginnings Intervention
All information about the intervention was obtained from Sleed et al. (2013)[97] unless otherwise stated.

**Intervention Description**
The intervention is delivered by two parent-infant psychotherapists and consisted of group discussions that center around potential triggers to the attachment relationship. Specifically, topics include the links between past and present patterns of relating, observation, and reflection on non-conscious behaviors between mothers and babies. During sessions, facilitators and mothers in the group observe and note mother-infant communications and mothers' responses to infants' signals.
Mothers and babies in both groups had access to standard health and social care provision as provided by the prison service. But the New Beginnings Intervention was compared to a group who only received standard care.

**Target population**
Incarcerated mothers and their infants

**Goal (and framework when applicable)**
This intervention drew on attachment theory. The goal of the intervention was to help mothers understand the relationship between their babies' behavior, the internal emotional world of their babies, as well as their own state of mind.

**Physical and Informational Materials**
The intervention employed group discussion, handouts, individual worksheets, and homework tasks outside of sessions.

**Setting**
The intervention was offered face-to-face in a group of 6 mother-baby dyads. Sessions took place In Mother and Baby Units in the hospital. During sessions, mothers sat on the floor while babies were placed on baby mats to facilitate active participation in intervention activities.

**Dose and Intensity**
There were eight, 2-hour sessions over four weeks, with two sessions per day, one day a week. The mean number of sessions attended was 7.1 (SD = 1.6). The intended total time was 16 hours, while the average total time was approximately 14.2 hours.
At baseline, infants ranged between birth and 23 months of age (mean of 4.9 months) and lasted for 4 weeks.

**Studies that Employed this intervention**: Sleed, Baradon and Fonagy [97]

#### Group Attachment-Based Intervention (GABI)

All information about the intervention was obtained from Steele et al. (2019)[98] unless otherwise stated.

**Intervention Description**
Each intervention session has three components. Sessions include time for parents and children to interact with one another, a period of video filming and feedback, and a "reunion" episode between parents and children after a period of separation while mothers are reviewing video footage of their interactions.
The intervention was delivered by trained GABI clinicians.

**Target population**
Families who are at risk for maltreatment and their biological infants.

**Goal (and framework when applicable)**
GABI is a clinical, trauma-informed approach to improving parent-child relationships. The intervention builds off of strategies intrinsic in other attachment-based interventions, described above (Dozier 2009, Bernard 2012).
The specific goals of the video-feedback portion of the program include:

1. The goal of training clinicians
2. Therapeutic goal of reviewing the videos with the parents to discuss and reflect on dyadic interactions

**Physical and Informational Materials**
The contents of the intervention were outlined in a GABI manual. The intervention also included video recordings of mother-child interactions.

**Setting**
The intervention was conducted face-to-face in multi-family groups at the New School's Center for Attachment Research (in a clinical context).

**Dose and Intensity**
Tripartite sessions were offered three times weekly for 26 weeks. Each session lasted 120 minutes. Each session began with a 45-minute dyadic, parent-child group psychotherapy session. Next, parents and children separate into concurrently run, hour-long group sessions. Each session ended with a 15-minute parent-child reunion. Mothers were offered a maximum of 72 sessions (156 hours total)
Infants and toddlers ranged in age from birth to three years old at baseline. The intervention was offered over 6 months.
A 24/7 hotline was also available to families partaking in the intervention.

**Studies that Employed this intervention**: Steele, Murphy, Bonuck, Meissner and Steele [98]

#### Secure Attachment Family Education (SAFE)

All information about the intervention was obtained from Walter et al. (2019)[99] unless otherwise stated.

**Intervention Description**
Secure Attachment Family Education (SAFE) is a multifaceted program aimed to promote secure infant-parent attachment. The intervention involves both mothers and fathers. The program consists of both group and individual sessions, beginning prenatally and spanning the postnatal period. Group sessions are discussion-based, with an special focus on facilitating communication. The program consists of four elements:

1. Video-based sensitivity training to enhance parental sensitivity

The video-based component begins prenatally, is an aspect of nearly every group session. In group sessions, participants watch video clips that show positive infant-parent interactions and are encouraged to reflect on the inner states of the parent and infant in the video. In individual sessions, facilitators provided feedback on the mothers and fathers (individual) interactions with the infant. Facilitators chose a single frame from the video and used it to deliver positive reinforcement on sensitive parental behavior

2. Reflection on parents' early and later attachment experiences and trauma

Parental psychological distress is discussed during group sessions. This included a discussion of social supports and stabilizing exercises (such as Luise Reddemann's "the imaginary journeys' exercise [304].

3. Psychoeducation about attachment theory and child development

Included discussions about the benefits of secure attachment, the key factors to promote attachment security, and the intergenerational transmission of attachment.

4. Social support from the facilitator and a group of other parents

The intervention was delivered by SAFE facilitators who were predominantly female (90.1%) professionals in social-service fields (including physicians, midwives, psychologists, nurses, and social workers). SAFE facilitators also received a 4-day training program prior to the intervention.

**Target population**
A non-clinical, low-risk, self-selected sample of mothers, fathers, and infants.

**Goal (and framework when applicable)**
The goal of the SAFE program is to promote parent-infant attachment and to prevent the intergenerational transmission of trauma.

**Physical and Informational Materials**
The information delivered was summarized in an intervention manual, but the contents of each session were individually tailored and deviated from the manual.

**Setting**
Not reported.

**Dose and Intensity**
The intervention began prenatally (the mean week of pregnancy at the beginning of the course was 24.34 (SD = 6.25) and spanned until the child was 1 year old.
10 group sessions and three individual sessions (13 sessions total).
Four sessions were conducted prenatally (1 individual, 4 as a group). The remaining 6 sessions were conducted postnatally, (2 individually, 6 as a group). Postnatal sessions were dyadic and lasted for a full day (10 am to 5pm; 7 hours). The estimated total length of the intervention was
Facilitators also established an emergency hotline that was available throughout the intervention.

**Studies that Employed this intervention:** Walter, Landers, Quehenberger, Carlson and Brisch [99]

#### Curriculum and Monitoring System

All information about the intervention was obtained from Badr et al. (2006)[100] unless otherwise stated.

**Intervention Description**
The Curriculum and Monitoring System (CAMS) is a 12-month long home-based stimulation program aimed to improve cognitive and sensorimotor skills in infants diagnosed with brain injury before discharge. Mothers are encouraged to provide a daily 20-minute stimulation activity, and researchers logged how many total minutes the intervention is provided to the infant. Outcome measures consisted of the following: motor and mental development, MRI examinations, home environment assessment, parent-infant interactions, parenting stress, and satisfaction with social support.

**Target population**
Infants with suspected brain injury

**Goal (and framework when applicable)**
CAMS is a program with over 100 activities that aim to improve motor and mental development via daily cognitive and sensorimotor stimulation. It consists of weekly home visits by public health nurses and daily stimulation activities. The curriculum has five sections: Cognitive, Language, Motor, Self-Help, and Social Skills. It has previously been used for research at Utah State University. It targets children from birth to 5 years of age.

**Physical and Informational Materials**
Motor and mental development were measured via Bayley Scales, neurological examinations were performed at 6, 12, and 18 months via MRI, the home environment was assessed via the Caldwell HOME inventory, parent-infant

interactions were coded via the Nursing Child Assessment Feeding Scale (NCAFS) and Nursing Child Assessment Teaching Scale (NCATS), parent stress was self-reported via the Parenting Stress Index (PSI), and social support was assessed da the Perceived Social Support Scale (PSS).

**Setting**
Home visits led by public health nurses at the pediatric clinics at UCLA.

**Dose and Intensity**
Home visits by public health nurses occurred twice a week for the first month (following hospital discharge), then weekly until the infant was 12 months old. The total number of visits was 36. Each home visit ranged from 60 minutes to 120 minutes. Outside of home visits, mothers were coached to provide daily stimulation activities for 20 minutes.
In total, the intervention is estimated to involve 3240 minutes of face-to-face interventionist time.

**Studies that Employed this intervention**: Badr, Garg and Kamath [100]

#### PremieStart Program

All information about the intervention was obtained from Milgrom et al. (2013)[101] unless otherwise stated.

**Intervention Description**
This parent-training program was adapted from the MITP. As a part of the intervention, mothers are encouraged to implement stress-reduction strategies as early as possible. The content of the PremieStart program is based on the MITP, with the addition of certain topics (described below). Each session focused on a specific topic, but previous topics were revisited briefly in each session.
In addition to the content of the original MITP curriculum, mothers are trained to increase their sensitivity in recognizing signs of infant stress (shutting down mechanisms, alert-available behavior, facial expressions, quality of motor behaviors, posture and muscle tone, graded stimulation, avoiding overwhelming infants
Four sessions included topics centered around touch, movement, and massage (kangaroo care, multi-sensory stimulation, debriefing and normalizing parental feelings, challenging dysfunctional thoughts, and parental diary keeping
Interventionists were psychologists (a neuropsychologist and clinical psychologist, each with extensive experience with premature infants).

**Target population**
Preterm mother-infant dyads (less than 30 weeks gestational age).

**Goal (and framework when applicable)**
To teach mothers psychoeducational signs of infant stress.

**Physical and Informational Materials**
The PremieStart program adhered to a manualized protocol.

**Setting**
In the NICU and in the home.

**Dose and Intensity**
Begins in the NICU. There are 9 weekly sessions in the NICU, plus one session at home (10 sessions total; approximately 10 weeks). Sessions lasted approximately 1 hour. The estimated total time of the intervention is 600 minutes.

**Studies that Employed this intervention**: Milgrom, Newnham, Martin, Anderson, Doyle, Hunt, Achenbach, Ferretti, Holt, Inder and Gemmill [101]

#### Mother-Infant Transaction Program (MITP)

All information about the intervention was obtained from Newnham et al. (2009)[103] unless otherwise stated.

**Intervention Description**
The intervention followed previously described sequence, outlined below [305]:
Session 1: The focus of session one is to become acquainted. Mothers are asked to describe their birthing, hospitalization experience, and perceptions of their infant.
Sessions 2-7: These sessions focus on recognizing infant disorganization, stress, and availability, and then working with mothers to apply that knowledge during care and play.

(i) Recognizing stress cues through homeostatic systems (respiration, skin circulation, visceral movements and activities), the motor system (posture, tone, movement) and infant states
(ii) Engaging and sustaining infant attention without allowing disorganization
(iii) Recognizing and responding to infant cues in daily care, including massage and kangaroo care
(iv) Initiating activity (combining the earlier principles with mothers' own ideas for alerting, responding to, regulating the infant's behavior and play.

Session 8: This session takes place in the home and centers on mutual enjoyment through play. Additional aspects of the session include exploring new play ideas within sensory modalities, adjustment to home, revisiting the program to help mothers regulate infant responses.
This iteration of the MITP differed from the original in that the hospital-based sessions were given over a 2-week period, 2 post-discharge sessions were given, twins and single mothers were included, and a bath session was used. This version of the intervention also included information about kangaroo dare and massage.
The intervention was delivered by a psychologist.

**Target population**
Preterm mother-infant dyads.

**Goal (and framework when applicable)**
Not reported.

**Physical and Informational Materials**
The intervention employed verbal instruction, infant observation, practical experience in handling infants and modeling, as well as written materials. The informational content was described in prior literature [305]. Newnham (2009) amended the intervention to include information about kangaroo care, massage, and an infant bath session.

**Setting**
Face to face, individually, In the NICU or nursery and in the home.

**Dose and Intensity**
There are 9 sessions in total, ranging from 30-60 minutes. The first 7 sessions were in the hospital, while the last two sessions took place in the home, post-discharge. The duration of the intervention was variable and depended on the infant's gestational age at birth.

**Studies that Employed this intervention**: Newnham, Milgrom and Skouteris [103]

#### Mother-Infant Transaction Program (MITP)

All information about the intervention was obtained from Ravn et al. (2011)[104] unless otherwise stated.

**Intervention Description**
The first session is used to become acquainted with to explain the intervention, and to demonstrate the infant's uniqueness and potential for self-regulation and interaction.
On the second session, the nurse introduces the mother to the behavioral indices of the homeostatic reflex systems. During the third session the nurse introduces the mother to the concept that posture, tone, and movement can signal disorganization. The nurse teaches the mother that her infant's progressive levels of consciousness could indicate whether the infant is socially accessible or whether social stimulation would be disruptive.
On the fifth day, the mother learns how to engage the infant and sustain social interaction with him or her. Familiar now with her baby's capabilities, the mother is ready to provide daily care in a more effective manner.
The 3 subsequent home visits focus on consolidation, mutual enjoyment through play, and temperament patterns.
The intervention was delivered by 10 trained neonatal nurses.

**Target population**
Mothers and moderately and late preterm infants.

**Goal (and framework when applicable)**
The intervention was influenced by Als' study of underweight newborn fragile organization and Bromwich's six-stage behavior progression model to sensitize mothers to infant cues and enable mothers to adjust to relatively poorly regulated preterm infants [305-307].
The goal of the intervention was to help parents appreciate their infant's unique characteristics, temperament, and developmental potential, to make the parents more sensitive and responsive to their infants' physiological and social cues, particularly that signal an overload of stimulus.

**Physical and Informational Materials**
Not listed in manuscript, a detailed description of the program has been previously reported [305].

**Setting**
Face to face, individually, in the NICU and in the home.

**Dose and Intensity**
Seven sessions were carried out during the last week before NICU discharge; four sessions were given at home during the first 3 months with the infant, with the mother and father (if possible) present. Each session lasted for 1 hour. The total estimated time is 11 hours. The intervention began as soon as possible after birth and lasted until 3 months postpartum.

**Studies that Employed this intervention**: Ravn, Smith, Lindemann, Smeby, Kyno, Bunch and Sandvik [104]

#### Mother–Infant Transaction Program (MITP) home visits

All information about the intervention was obtained from Youn et al. (2021)[105] unless otherwise stated.

**Intervention Description**
In this intervention RCT, the group intervention was introduced five days earlier than the original IHDP. However, the intervention is based on the IHDP framework. Up to 12 group sessions—usually 4-5 family members attended—occur while the infant is between 3-6 months of age. The group sessions are facilitated by a physiotherapist specialist—specializing in infant neurodevelopment—assisted by an experienced pediatric physiotherapist.
The timing of the home visits was based on MITP framework. The families received four home visits by an experienced NICU nurse staff member when the infant was five-days, two-weeks, one month and finally at two-months corrected age. Education and evaluation of the of caregivers were standardized using checklists.

**Goal (and framework when applicable)**
This intervention was influenced by the Mother–Infant Transaction Program (MITP) as well as IHDP. The goal was to provide the parent with a fuller understanding of their infant's behavioral cues, temperament, satiety, sleeping patterns, developmental milestones.

**Physical and Informational Materials**
Group Sessions – Information was provided to the infant's caregiver by a physiotherapist specializing in infant neurodevelopment, assisted by an experienced pediatric physiotherapist. The information included the following topics: parent-infant bonding, infant's growth and neurodevelopment, developmental milestones and various activities to promote sensory stimulation and gross motor development. The physiotherapists also offered emotional support to the caregivers to encourage attachment between baby and caregiver. During the visit, the caregivers and providers observed the infant's behaviors, and the environment was modified based on the needs of the infant. This was to ensure that the caregivers were understanding the material that was being taught. After the meetings, caregivers shared their experience and ideas.
Home Visits – Information was provided regarding the infant's behavioral cues (crying, temperament, satiety, hygiene, defecation, sleep patterns and position, feeding support, as well as what to do in case of an emergency).

**Setting**
Face to face, individually, in the NICU and in the home.

**Dose and Intensity**
Up to 12 group sessions were allowed while the infant was between 3-6 months of age. After discharge from the NICU, the families had four home visits by a nurse when the infant was five-days, two-weeks, one month and finally at two-months corrected age. Each home visit took approximately 90 minutes (about 1 and a half hours). Altogether, this intervention took approximately 18+ total hours.

**Studies that Employed this intervention:** Youn, Shin, Kim, Jin, Jung, Heo, Jeon, Park and Sung [105]

#### Auditory-Tactile-Visual-Vestibular (ATVV)

All information about the intervention was obtained from Nelson et al. (2001) and Holditch-Davis et al. (2014)[106,107] unless otherwise stated.

**Intervention Description**
During the ATVV intervention, interventionists teach mothers how to perform a 15-minute sequence of multimodal stimulation on their infants.
The session begins with auditory stimulation, progresses to bimodal auditory and tactile stimulation, and eventually visual, eye-to-eye stimulation is added as the infant becomes alert[308]. This combination of voice, moderate stroking or massage, and eye contact is sustained for the first 10 minutes of the intervention. During the final 5 minutes, the tactile component is withdrawn, and horizontal rocking is added. Tactile stimuli preceded from the least to the most sensitive areas and included moderate stroking of the top and back of the infant's head, back, chest, abdomen, arms, legs, and forehead.
Mothers were that ATVV increases feeding readiness, and therefore suggested they perform ATVV before a feed. Mothers were instructed not to perform the intervention while infants were on mechanical ventilation or continuous positive airway pressure [106].
Mothers were also videotaped while administering the ATVV session, which was reviewed by the NICU nurse and supervisor to guide and correct mothers [106]. The interventionist was a NICU nurse [106], or a research assistant trained to criterion for this intervention [107].

**Target population**
Preterm mother-infant dyads [106] Mothers and preterm infants with extreme prematurity or intraventricular hemorrhage and/or periventricular leukomalacia [107].

**Goal (and framework when applicable)**
The goal of the ATVV intervention is to positively affect mothers and the mother-infant relationship.
The three modes of stimulation were chosen because the tactile and vestibular sensory pathways are well-developed by 33 weeks postconceptional age, and are most relevant to mother-infant interaction and learning.

**Physical and Informational Materials**
Not reported

**Setting**
Hybrid; in the NICU and in the home [106] or solely in the NICU [107].

**Dose and Intensity**
Mothers were trained by a NICU nurse in approximately 1 hour. Mothers had an additional two contacts with NICU nurses to videotape and review the intervention, although the timing of these contacts was not reported. Mothers were asked to visit the NICU to perform ATVV independently as often as possible (preferably daily), but at least 3 times a week for at least 15 minutes. The intervention continued until the infants were 2 months corrected age [106].
One 15-minute training session at 33 weeks postconceptional age. The intervention continued until the infants were 2 months corrected age [107].

**Studies that Employed this intervention**: Holditch-Davis, White-Traut, Levy, O'Shea, Geraldo and David [106,]Nelson, White-Traut, Vasan, Silvestri, Comiskey, Meleedy-Rey, Littau, Gu and Patel [107]

#### Hospital to Home: Optimizing the Infants Environment (H-Hope) Intervention

All information about the intervention was obtained from White-Traut et al. (2013)[108] unless otherwise stated.

**Intervention Description**

The H-Hope intervention combines an infant remediation intervention (ATVV) with maternal education. The goal is to improve maternal understanding of and response to infant cues. The intervention consists of 6 sessions with interventionists– 2 in the hospital, 2 at home, and 2 phone calls.
The intervention was delivered by a Nurse-Advocate team (NAT).
During the first hospital visit, the NAT taught the mother how to perform the ATVV component, which consisted of 10 minutes of auditory, tactile, and visual stimulation, followed by 5 minutes of vestibular stimulation (horizontal rocking). The ATVV began when the infant reached 32 weeks PMA, or at entry to the study at 33-34 weeks. After this training, mothers were asked to perform ATVV on their own twice a day until the infant was 1 month corrected age. Prior to discharge, if mothers were unable to come into the hospital to perform the ATVV, they would coordinate with their NAT team to perform the ATVV for them.
During the mother-directed component of the intervention (offered twice in the hospital, twice at home, and twice over the phone), the NAT asked about and responded to the mother's concerns about herself and her infant. Interventionists referred mothers to appropriate support services as needed.
During visit 2, the NAT taught mothers how to identify and respond to infant behavioral cues, in part by using an instructional pamphlet titled "How to Soothe a Fussy Baby."
During the home visits, the NAT discussed feeding, weighing, and the mother's stressors, supports, or signs of depression. The mothers also practiced the ATVV during home visits. The NAT also observed an infant feeding and offered participatory guidance.
The 2 phone call sessions provided mothers another opportunity to express any questions or concerns.

**Target population**

Preterm mother-infant dyads

**Goal (and framework when applicable)**

The H-HOPE intervention was based on the transactional model and integrates infant remediation intervention (ATVV) with maternal re-education and re-definition through participatory guidance.
The goal of the H-HOPE intervention was to establish positive interaction patterns, improve the quality of mother-infant interactions, and target the needs of both mothers and premature infants.

**Physical and Informational Materials**

NATs used a pamphlet titled "How to Soothe a Fussy Baby," which provided instruction about how to identify and respond to infant behavioral cues. The pamphlet was adapted from materials in the medical poster parent handbook [309]. Information about how to modulate infant state and use of soothing behaviors (from "Keys to Caregiving" curriculum) was provided to mothers. Topics covered include describing the age of the newborn, what infant behaviors mean in the context of feeding and mother-infant interaction, and teaching caregivers how to support infant sleep and wakefulness states.

**Setting**

In-person before hospital discharge, then at home (in person and over the phone).

**Dose and Intensity**

The NAT component was provided during 2 in-hospital visits, 2 home visits and 2 phone calls after discharge.
The length of each session was not reported (although each individual ATVV session was 15 minutes).
Infants were between 32-34 weeks PMA at baseline, and the intervention lasted from enrollment to 1 month corrected age.

**Studies that Employed this intervention**: White-Traut, Norr, Fabiyi, Rankin, Li and Liu [108]

#### Community HUGS (CHUGS)

All information about the intervention was obtained from Ericksen et al. (2018)[109] unless otherwise stated.

**Intervention Description**
This group intervention was divided into four modules.
Module 1: 3 sessions, "Let's Play"

- Focus: group formation, play, and anxiety management
- Content: play activities, moving together with the baby in space, stress busters, and infant massage

Module 2: 3 sessions "Getting to Know Each Other"

- Focus: parenting and attachment
- Content: observation of baby in play, practice in face-to-face playful communication and experiences in mother-infant bodily holding in seated play and movement together that interactively engage with baby. Includes information on temperament (infant individual differences)

Module 3: 3 sessions "Know Yourself"

- Focus: family or origin and cognitive behavioral therapy
- Content: movement for mothers and babies in group circles and individual pathways while listening to music, with a focus on mother-infant energy levels. These sessions also included live percussion. Further content included a discussion of parenting behaviors, hopes and expectations for infants, interpreting thoughts and feelings, and addressing balance in your life.

Module 4: 1 session "Happiness, Understanding, Giving, and Sharing"

- Focus: consolidation in thinking and movement, as well as a celebration of intervention progress.
- Content: brainstorming and sharing takeaways from CHUGS.

Interventionists were the program designers as well as newly trained facilitators. Facilitator competence was achieved through a role play-based method.

**Target population**
Mothers vulnerable to postpartum depression and their infants

**Goal (and framework when applicable)**
Eriksen (2018) hypothesized that CHUGS would reduce stress on the infant-parent relationship while improving mother-infant interactions, parenting skills, maternal self-efficacy, and maternal health and well-being.

**Physical and Informational Materials**
Not reported.

**Setting**
Face to face in a group of 4-8 women and their infants. Sessions took place in a comfortable play space in a public hospital. During the intervention, mothers were encouraged to sit on the floor on a blanket.

**Dose and Intensity**
Participants completed four modules over 10 weekly sessions that lasted between 60-90 minutes (approximately). The total estimated time is 10 to 15 hours. Infants were 4.94 months old at baseline (on average). The intervention lasted for 10 weeks.

**Studies that Employed this intervention**: Ericksen, Loughlin, Holt, Rose, Hartley, Buultjens, Gemmill and Milgrom [109]

#### HUGS

All information about the intervention was obtained from Holt et al. (2021)[110] unless otherwise stated.

**Intervention Description**
This intervention consists of four mother-infant group sessions. Although HUGS was developed as a three-session program, the suggested content was used to expand this intervention into a four-session program [310], as the developers intended if time allowed [311]. The HUGS facilitator was either a clinical psychologist or a maternal child health nurse, trained specifically to deliver the HUGS intervention.
Sessions included psychoeducation and behavioral exercises, including interaction coaching and infant massage.

Session One: Play and Physical Contact. During this session, facilitators allowed the caregivers to interact with their infants and alternative responses were modelled.
Session Two: Observing and Understanding Baby's Signals. During this session, the facilitator used guided exercises to reinforce 'good enough' interactions.
Session Three: Parental Responses to Infant Cues. This session worked to build up the caregiver's cognitive strategies that they had learned during the PND program. Specifically, separating past experiences and challenging cognitions that had been distorted so that the caregiver could focus on the reality of the infant.
Session Four (Booster Session): Consolidating Gains. This session was a reinforcing session. The facilitator did this by noting positive interaction cognitions and behaviors that the caregiver had toward the infant.

**Target population**
Mothers with postnatal depression and their infants.

**Goal (and framework when applicable)**
HUGS aims to ameliorate mother-infant interaction by targeting maternal communication, observation, and responsiveness and increase maternal emotional availability by improving distorted maternal internal representations and cognitions.
HUGS draws on existing interventions [312], as well as theories and developmental research describing a 'good enough' parent interaction. HUGS targets two key elements of mother-infant relationships: (1) maternal skills in communication, observation, and responsiveness and (2) distorted maternal internal representations/cognitions.
By treating maternal depression first, this design aimed to allow for increased maternal emotional availability, a basis for optimal mother-infant interactions

**Physical and Informational Materials**
Not reported.

**Setting**
Sessions took place face-to-face in a group setting at a parent-infant research institute or local community center.

**Dose and Intensity**
The intervention had 4, weekly sessions, each lasting 90 minutes (360 minutes total).
Infants were 3.13 months on average at baseline. The intervention lasted for 4 weeks.

**Studies that Employed this intervention**: Holt, Gentilleau, Gemmill and Milgrom [110]

#### eFigure 1. Overview of Moderator Analyses

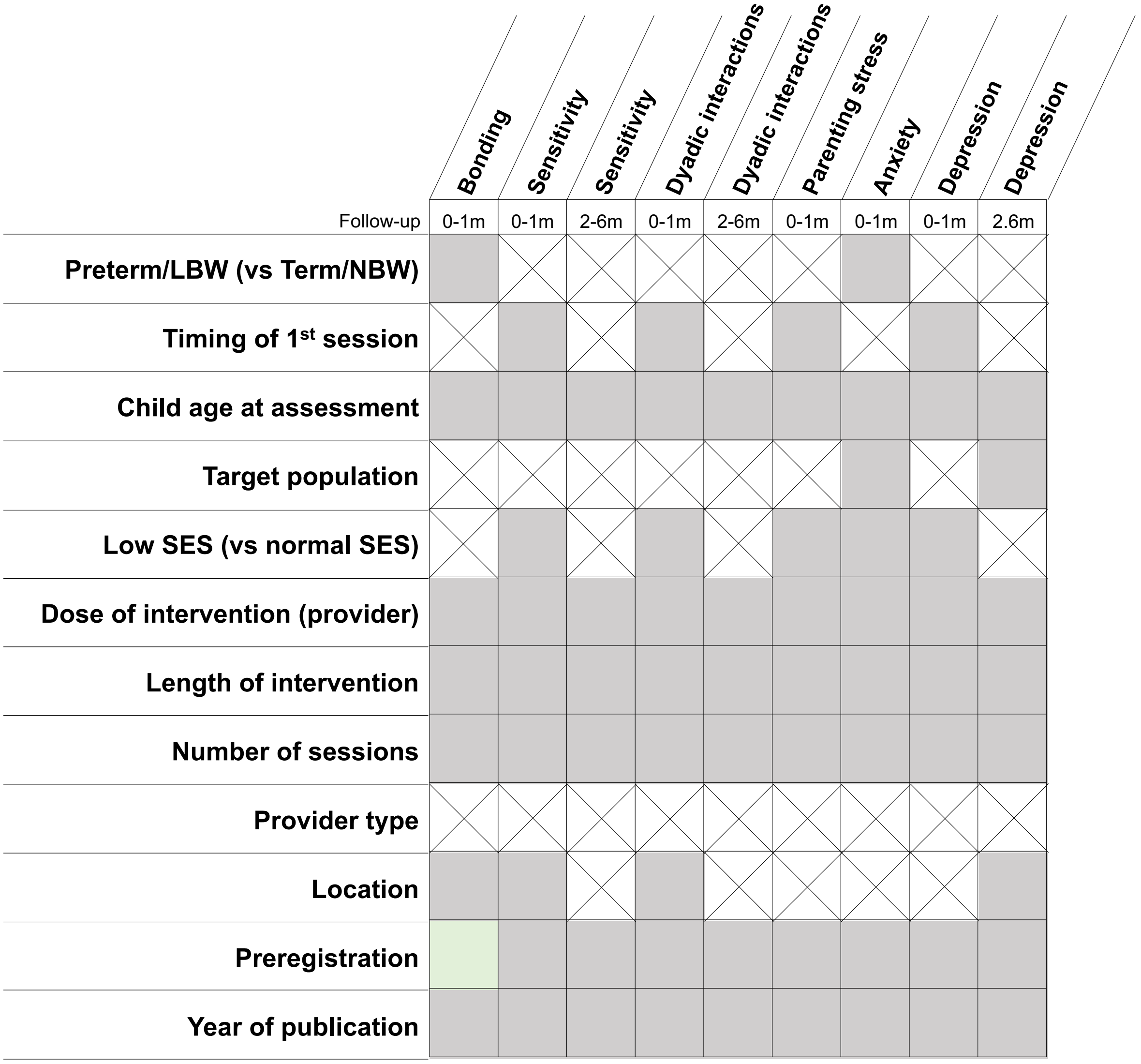


Note: Grey sharing indicate non-significant association. Green shading indicated signification association (p<.05). Crossed-out squared indicate associations that could not be tested.

### eFigure2. Bonding 0-1-month Follow-Up Forrest Plot

| | Experimental | | | Control | | | | Std. Mean Difference |
|---|---|---|---|---|---|---|---|---|
| **Study or Subgroup** | **Mean** | **SD** | **Total** | **Mean** | **SD** | **Total** | **Weight** | **IV, Random, 95% CI** |
| Akai 2008 | 0 | 0 | 0 | 0 | 0 | 0 | | Not estimable |
| Alarcao 2021 | 0 | 0 | 0 | 0 | 0 | 0 | | Not estimable |
| Alvarenga 2019 | 0 | 0 | 0 | 0 | 0 | 0 | | Not estimable |
| Badr 2006 | 0 | 0 | 0 | 0 | 0 | 0 | | Not estimable |
| Baggett 2010 | 0 | 0 | 0 | 0 | 0 | 0 | | Not estimable |
| Baggett 2017 | 0 | 0 | 0 | 0 | 0 | 0 | | Not estimable |
| Barlow 2016 | 0 | 0 | 0 | 0 | 0 | 0 | | Not estimable |
| Barnes 2022 | 89.4 | 3.7 | 17 | 85.7 | 6.2 | 17 | 6.5% | 0.71 [0.01, 1.40] |
| Berlin 2017 | 0 | 0 | 0 | 0 | 0 | 0 | | Not estimable |
| Berlin 2018 | 0 | 0 | 0 | 0 | 0 | 0 | | Not estimable |
| Bernard 2012 | 0 | 0 | 0 | 0 | 0 | 0 | | Not estimable |
| Betancourt 2020 | 0 | 0 | 0 | 0 | 0 | 0 | | Not estimable |
| Bick 2013 | 0 | 0 | 0 | 0 | 0 | 0 | | Not estimable |
| Borghini 2014 | 0 | 0 | 0 | 0 | 0 | 0 | | Not estimable |
| Brisch 2003 | 0 | 0 | 0 | 0 | 0 | 0 | | Not estimable |
| Browne 2005 | 0 | 0 | 0 | 0 | 0 | 0 | | Not estimable |
| Cates 2018 | 0 | 0 | 0 | 0 | 0 | 0 | | Not estimable |
| Cevasco 2008 | 0 | 0 | 0 | 0 | 0 | 0 | | Not estimable |
| Chiu 2009 | 0 | 0 | 0 | 0 | 0 | 0 | | Not estimable |
| Constantino 2001 | 0 | 0 | 0 | 0 | 0 | 0 | | Not estimable |
| Cooper 2009 | 0 | 0 | 0 | 0 | 0 | 0 | | Not estimable |
| Cooper 2015 | 0 | 0 | 0 | 0 | 0 | 0 | | Not estimable |
| Corrigan 2021 | -0.64 | 1.6 | 14 | -0.57 | 1.5 | 28 | 6.8% | -0.04 [-0.69, 0.60] |
| Dozier 2009 | 0 | 0 | 0 | 0 | 0 | 0 | | Not estimable |
| Eriksen 2018 | 0 | 0 | 0 | 0 | 0 | 0 | | Not estimable |
| Feil 2020 | 0 | 0 | 0 | 0 | 0 | 0 | | Not estimable |
| Feldman 2014 | 0 | 0 | 0 | 0 | 0 | 0 | | Not estimable |
| Firk 2021 | 0 | 0 | 0 | 0 | 0 | 0 | | Not estimable |
| Fonagy 2016 | 0 | 0 | 0 | 0 | 0 | 0 | | Not estimable |
| Gaden 2022 | -5.51 | 6.65 | 105 | -5.92 | 6.38 | 101 | 8.3% | 0.06 [-0.21, 0.34] |
| Georg 2021 | 0 | 0 | 0 | 0 | 0 | 0 | | Not estimable |
| Glazebrook 2007 | 0 | 0 | 0 | 0 | 0 | 0 | | Not estimable |
| Hane 2015 | 0 | 0 | 0 | 0 | 0 | 0 | | Not estimable |
| Heo 2019 | 3.88 | 0.01 | 30 | 3.65 | 0.02 | 32 | 1.5% | 14.22 [11.59, 16.86] |
| Herbers 2020 | 0 | 0 | 0 | 0 | 0 | 0 | | Not estimable |
| Hoffenkamp 2015 | -5.6 | 4.68 | 75 | -6.31 | 4.68 | 75 | 8.2% | 0.15 [-0.17, 0.47] |
| Hoivik 2015 | 0 | 0 | 0 | 0 | 0 | 0 | | Not estimable |
| Holditch-Davis 2014 | 0 | 0 | 0 | 0 | 0 | 0 | | Not estimable |
| Holt 2021 | -7.41 | 3.59 | 27 | -8.62 | 5.33 | 28 | 7.3% | 0.26 [-0.27, 0.79] |
| Horowitz 2001 | 0 | 0 | 0 | 0 | 0 | 0 | | Not estimable |
| Horowitz 2013 | 0 | 0 | 0 | 0 | 0 | 0 | | Not estimable |
| Juffer 2005 | 0 | 0 | 0 | 0 | 0 | 0 | | Not estimable |
| Klein Velderman 2006 | 0 | 0 | 0 | 0 | 0 | 0 | | Not estimable |
| Knoche 2012 | 0 | 0 | 0 | 0 | 0 | 0 | | Not estimable |
| Kristensen 2020 | 0 | 0 | 0 | 0 | 0 | 0 | | Not estimable |
| Landry 2006 | 0 | 0 | 0 | 0 | 0 | 0 | | Not estimable |
| Lavallee 2022 | 0 | 0 | 0 | 0 | 0 | 0 | | Not estimable |
| Magil-Evans 2007 | 0 | 0 | 0 | 0 | 0 | 0 | | Not estimable |
| Meijssen 2010 | 0 | 0 | 0 | 0 | 0 | 0 | | Not estimable |
| Melnyk 2006 | 0 | 0 | 0 | 0 | 0 | 0 | | Not estimable |
| Mendelsohn 2007 | 0 | 0 | 0 | 0 | 0 | 0 | | Not estimable |
| Milgrom 2013 | 0 | 0 | 0 | 0 | 0 | 0 | | Not estimable |
| Nelson 2001 | 0 | 0 | 0 | 0 | 0 | 0 | | Not estimable |
| Neu 2010 | 0 | 0 | 0 | 0 | 0 | 0 | | Not estimable |
| Newnham 2009 | 0 | 0 | 0 | 0 | 0 | 0 | | Not estimable |
| Nugent 2017 | 0 | 0 | 0 | 0 | 0 | 0 | | Not estimable |
| Onozawa 2001 | 0 | 0 | 0 | 0 | 0 | 0 | | Not estimable |
| Oxford 2021 | 0 | 0 | 0 | 0 | 0 | 0 | | Not estimable |
| Perrone 2021 | 0 | 0 | 0 | 0 | 0 | 0 | | Not estimable |
| Pontoppidan 2016 | 0 | 0 | 0 | 0 | 0 | 0 | | Not estimable |
| Porter 2015 | 3.88 | 0.27 | 56 | 3.51 | 0.64 | 30 | 7.6% | 0.84 [0.38, 1.30] |
| Ramsauer 2019 | 0 | 0 | 0 | 0 | 0 | 0 | | Not estimable |
| Ravn 2011 | 0 | 0 | 0 | 0 | 0 | 0 | | Not estimable |
| Rheinheimer 2022 | 0 | 0 | 0 | 0 | 0 | 0 | | Not estimable |
| Robertson 2019 | 0 | 0 | 0 | 0 | 0 | 0 | | Not estimable |
| Roby 2021 | 6.7 | 0.44 | 178 | 6.67 | 0.44 | 184 | 8.5% | 0.07 [-0.14, 0.27] |
| Sadler 2013 | 0 | 0 | 0 | 0 | 0 | 0 | | Not estimable |
| Sahlen Helmer 2020 | 0 | 0 | 0 | 0 | 0 | 0 | | Not estimable |
| Sajaniemi 2001 | 0 | 0 | 0 | 0 | 0 | 0 | | Not estimable |
| Salo 2019 | 0 | 0 | 0 | 0 | 0 | 0 | | Not estimable |
| Santelices 2010 | 0 | 0 | 0 | 0 | 0 | 0 | | Not estimable |
| Schuler 2000 | 0 | 0 | 0 | 0 | 0 | 0 | | Not estimable |
| Shoghi 2018 | 0 | 0 | 0 | 0 | 0 | 0 | | Not estimable |
| Sleed 2013 | 0 | 0 | 0 | 0 | 0 | 0 | | Not estimable |
| Steele 2019 | 0 | 0 | 0 | 0 | 0 | 0 | | Not estimable |
| Stein 2006 | 0 | 0 | 0 | 0 | 0 | 0 | | Not estimable |
| Stein 2018 | 0 | 0 | 0 | 0 | 0 | 0 | | Not estimable |
| Taneja 2020 | 87.33 | 5.2 | 254 | 86.44 | 5.5 | 262 | 8.6% | 0.17 [-0.01, 0.34] |
| Tereno 2017 | 0 | 0 | 0 | 0 | 0 | 0 | | Not estimable |
| Teti 2009 | 0 | 0 | 0 | 0 | 0 | 0 | | Not estimable |
| Tryphonopoulos 2020 | 0 | 0 | 0 | 0 | 0 | 0 | | Not estimable |
| Tsivos 2015 | -21.5 | 20.49 | 12 | -14.63 | 12.38 | 9 | 5.7% | -0.38 [-1.25, 0.50] |
| Twohig 2021 | 0 | 0 | 0 | 0 | 0 | 0 | | Not estimable |
| Valades 2021 | 0 | 0 | 0 | 0 | 0 | 0 | | Not estimable |
| van Doesum 2008 | 0 | 0 | 0 | 0 | 0 | 0 | | Not estimable |
| Vargas-Porras 2021 | 96 | 0.001 | 33 | 90.03 | 6.05 | 33 | 7.3% | 1.38 [0.84, 1.92] |
| Walter 2019 | 0 | 0 | 0 | 0 | 0 | 0 | | Not estimable |
| White-Traut 2013 | 0 | 0 | 0 | 0 | 0 | 0 | | Not estimable |
| Williams 2020 | 0 | 0 | 0 | 0 | 0 | 0 | | Not estimable |
| Wulff 2021 | -6.77 | 4.48 | 56 | -6.04 | 4.4 | 57 | 8.0% | -0.16 [-0.53, 0.21] |
| Youn 2021 | 100.1 | 4.3 | 69 | 101.3 | 5.1 | 67 | 8.1% | -0.25 [-0.59, 0.08] |
| Yu 2022 | 104.34 | 7.36 | 32 | 102.63 | 9.82 | 32 | 7.5% | 0.19 [-0.30, 0.69] |
| Zelkowitz 2011 | 0 | 0 | 0 | 0 | 0 | 0 | | Not estimable |
| **Total (95% CI)** | | | **958** | | | **955** | **100.0%** | **0.43 [0.08, 0.78]** |

Heterogeneity: Tau² = 0.36; Chi² = 151.77, df = 13 (P < 0.00001); I² = 91%

Test for overall effect: Z = 2.43 (P = 0.02)

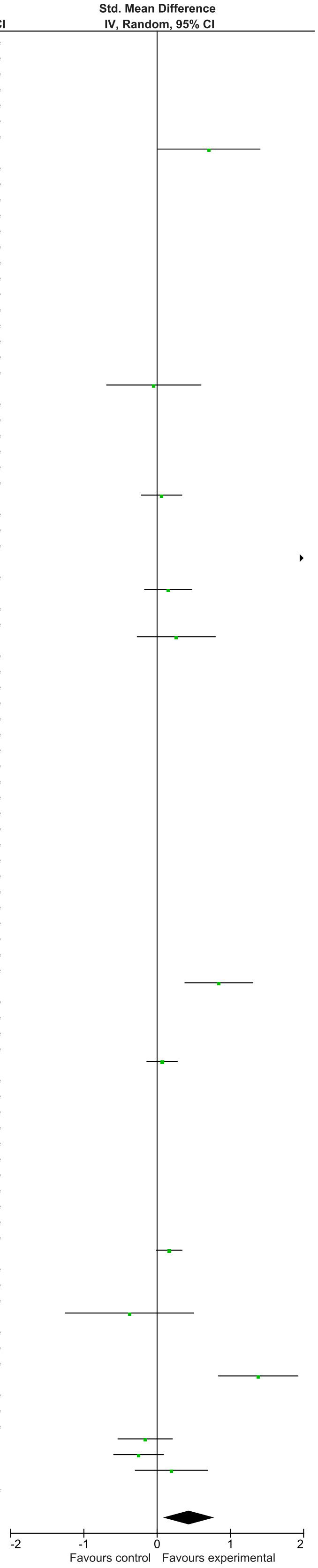

### eFigure 3. Bonding 2-6-month Follow-Up Forrest Plot

| Study or Subgroup | Experimental Mean | Experimental SD | Experimental Total | Control Mean | Control SD | Control Total | Weight | Std. Mean Difference IV, Random, 95% CI |
|---|---|---|---|---|---|---|---|---|
| Akai 2008 | 0 | 0 | 0 | 0 | 0 | 0 | | Not estimable |
| Alarcao 2021 | 0 | 0 | 0 | 0 | 0 | 0 | | Not estimable |
| Alvarenga 2019 | 0 | 0 | 0 | 0 | 0 | 0 | | Not estimable |
| Badr 2006 | 0 | 0 | 0 | 0 | 0 | 0 | | Not estimable |
| Baggett 2010 | 0 | 0 | 0 | 0 | 0 | 0 | | Not estimable |
| Baggett 2017 | 0 | 0 | 0 | 0 | 0 | 0 | | Not estimable |
| Barlow 2016 | 0 | 0 | 0 | 0 | 0 | 0 | | Not estimable |
| Barnes 2022 | 0 | 0 | 0 | 0 | 0 | 0 | | Not estimable |
| Berlin 2017 | 0 | 0 | 0 | 0 | 0 | 0 | | Not estimable |
| Berlin 2018 | 0 | 0 | 0 | 0 | 0 | 0 | | Not estimable |
| Bernard 2012 | 0 | 0 | 0 | 0 | 0 | 0 | | Not estimable |
| Betancourt 2020 | 0 | 0 | 0 | 0 | 0 | 0 | | Not estimable |
| Bick 2013 | 0 | 0 | 0 | 0 | 0 | 0 | | Not estimable |
| Borghini 2014 | 0 | 0 | 0 | 0 | 0 | 0 | | Not estimable |
| Brisch 2003 | 0 | 0 | 0 | 0 | 0 | 0 | | Not estimable |
| Browne 2005 | 0 | 0 | 0 | 0 | 0 | 0 | | Not estimable |
| Cates 2018 | 0 | 0 | 0 | 0 | 0 | 0 | | Not estimable |
| Cevasco 2008 | 0 | 0 | 0 | 0 | 0 | 0 | | Not estimable |
| Chiu 2009 | 0 | 0 | 0 | 0 | 0 | 0 | | Not estimable |
| Constantino 2001 | 0 | 0 | 0 | 0 | 0 | 0 | | Not estimable |
| Cooper 2009 | 0 | 0 | 0 | 0 | 0 | 0 | | Not estimable |
| Cooper 2015 | 0 | 0 | 0 | 0 | 0 | 0 | | Not estimable |
| Corrigan 2021 | 0 | 0 | 0 | 0 | 0 | 0 | | Not estimable |
| Dozier 2009 | 0 | 0 | 0 | 0 | 0 | 0 | | Not estimable |
| Eriksen 2018 | 0 | 0 | 0 | 0 | 0 | 0 | | Not estimable |
| Feil 2020 | 0 | 0 | 0 | 0 | 0 | 0 | | Not estimable |
| Feldman 2014 | 0 | 0 | 0 | 0 | 0 | 0 | | Not estimable |
| Firk 2021 | 0 | 0 | 0 | 0 | 0 | 0 | | Not estimable |
| Fonagy 2016 | 0 | 0 | 0 | 0 | 0 | 0 | | Not estimable |
| Gaden 2022 | 0 | 0 | 0 | 0 | 0 | 0 | | Not estimable |
| Georg 2021 | 0 | 0 | 0 | 0 | 0 | 0 | | Not estimable |
| Glazebrook 2007 | 0 | 0 | 0 | 0 | 0 | 0 | | Not estimable |
| Hane 2015 | 0 | 0 | 0 | 0 | 0 | 0 | | Not estimable |
| Heo 2019 | 0 | 0 | 0 | 0 | 0 | 0 | | Not estimable |
| Herbers 2020 | 0 | 0 | 0 | 0 | 0 | 0 | | Not estimable |
| Hoffenkamp 2015 | -4.53 | 3.81 | 75 | -4.04 | 3.81 | 75 | 32.0% | -0.13 [-0.45, 0.19] |
| Hoivik 2015 | 0 | 0 | 0 | 0 | 0 | 0 | | Not estimable |
| Holditch-Davis 2014 | 0 | 0 | 0 | 0 | 0 | 0 | | Not estimable |
| Holt 2021 | -6.55 | 3.63 | 22 | -8.79 | 5.24 | 28 | 24.0% | 0.48 [-0.09, 1.05] |
| Horowitz 2001 | 0 | 0 | 0 | 0 | 0 | 0 | | Not estimable |
| Horowitz 2013 | 0 | 0 | 0 | 0 | 0 | 0 | | Not estimable |
| Juffer 2005 | 0 | 0 | 0 | 0 | 0 | 0 | | Not estimable |
| Klein Velderman 2006 | 0 | 0 | 0 | 0 | 0 | 0 | | Not estimable |
| Knoche 2012 | 0 | 0 | 0 | 0 | 0 | 0 | | Not estimable |
| Kristensen 2020 | 0 | 0 | 0 | 0 | 0 | 0 | | Not estimable |
| Landry 2006 | 0 | 0 | 0 | 0 | 0 | 0 | | Not estimable |
| Lavallee 2022 | 0 | 0 | 0 | 0 | 0 | 0 | | Not estimable |
| Magil-Evans 2007 | 0 | 0 | 0 | 0 | 0 | 0 | | Not estimable |
| Meijssen 2010 | 0 | 0 | 0 | 0 | 0 | 0 | | Not estimable |
| Melnyk 2006 | 0 | 0 | 0 | 0 | 0 | 0 | | Not estimable |
| Mendelsohn 2007 | 0 | 0 | 0 | 0 | 0 | 0 | | Not estimable |
| Milgrom 2013 | 0 | 0 | 0 | 0 | 0 | 0 | | Not estimable |
| Nelson 2001 | 0 | 0 | 0 | 0 | 0 | 0 | | Not estimable |
| Neu 2010 | 0 | 0 | 0 | 0 | 0 | 0 | | Not estimable |
| Newnham 2009 | 0 | 0 | 0 | 0 | 0 | 0 | | Not estimable |
| Nugent 2017 | 0 | 0 | 0 | 0 | 0 | 0 | | Not estimable |
| Onozawa 2001 | 0 | 0 | 0 | 0 | 0 | 0 | | Not estimable |
| Oxford 2021 | 0 | 0 | 0 | 0 | 0 | 0 | | Not estimable |
| Perrone 2021 | 0 | 0 | 0 | 0 | 0 | 0 | | Not estimable |
| Pontoppidan 2016 | 0 | 0 | 0 | 0 | 0 | 0 | | Not estimable |
| Porter 2015 | 3.89 | 0.27 | 50 | 3.54 | 0.71 | 35 | 27.9% | 0.69 [0.25, 1.14] |
| Ramsauer 2019 | 0 | 0 | 0 | 0 | 0 | 0 | | Not estimable |
| Ravn 2011 | 0 | 0 | 0 | 0 | 0 | 0 | | Not estimable |
| Rheinheimer 2022 | 0 | 0 | 0 | 0 | 0 | 0 | | Not estimable |
| Robertson 2019 | 0 | 0 | 0 | 0 | 0 | 0 | | Not estimable |
| Roby 2021 | 0 | 0 | 0 | 0 | 0 | 0 | | Not estimable |
| Sadler 2013 | 0 | 0 | 0 | 0 | 0 | 0 | | Not estimable |
| Sahlen Helmer 2020 | 0 | 0 | 0 | 0 | 0 | 0 | | Not estimable |
| Sajaniemi 2001 | 0 | 0 | 0 | 0 | 0 | 0 | | Not estimable |
| Salo 2019 | 0 | 0 | 0 | 0 | 0 | 0 | | Not estimable |
| Santelices 2010 | 0 | 0 | 0 | 0 | 0 | 0 | | Not estimable |
| Schuler 2000 | 0 | 0 | 0 | 0 | 0 | 0 | | Not estimable |
| Shoghi 2018 | 0 | 0 | 0 | 0 | 0 | 0 | | Not estimable |
| Sleed 2013 | 0 | 0 | 0 | 0 | 0 | 0 | | Not estimable |
| Steele 2019 | 0 | 0 | 0 | 0 | 0 | 0 | | Not estimable |
| Stein 2006 | 0 | 0 | 0 | 0 | 0 | 0 | | Not estimable |
| Stein 2018 | 0 | 0 | 0 | 0 | 0 | 0 | | Not estimable |
| Taneja 2020 | 0 | 0 | 0 | 0 | 0 | 0 | | Not estimable |
| Tereno 2017 | 0 | 0 | 0 | 0 | 0 | 0 | | Not estimable |
| Teti 2009 | 0 | 0 | 0 | 0 | 0 | 0 | | Not estimable |
| Tryphonopoulos 2020 | 0 | 0 | 0 | 0 | 0 | 0 | | Not estimable |
| Tsivos 2015 | -15.5 | 14.69 | 12 | -15.75 | 10.37 | 9 | 16.1% | 0.02 [-0.85, 0.88] |
| Twohig 2021 | 0 | 0 | 0 | 0 | 0 | 0 | | Not estimable |
| Valades 2021 | 0 | 0 | 0 | 0 | 0 | 0 | | Not estimable |
| van Doesum 2008 | 0 | 0 | 0 | 0 | 0 | 0 | | Not estimable |
| Vargas-Porras 2021 | 0 | 0 | 0 | 0 | 0 | 0 | | Not estimable |
| Walter 2019 | 0 | 0 | 0 | 0 | 0 | 0 | | Not estimable |
| White-Traut 2013 | 0 | 0 | 0 | 0 | 0 | 0 | | Not estimable |
| Williams 2020 | 0 | 0 | 0 | 0 | 0 | 0 | | Not estimable |
| Wulff 2021 | 0 | 0 | 0 | 0 | 0 | 0 | | Not estimable |
| Youn 2021 | 0 | 0 | 0 | 0 | 0 | 0 | | Not estimable |
| Yu 2022 | 0 | 0 | 0 | 0 | 0 | 0 | | Not estimable |
| Zelkowitz 2011 | 0 | 0 | 0 | 0 | 0 | 0 | | Not estimable |
| **Total (95% CI)** | | | **159** | | | **147** | **100.0%** | **0.27 [-0.19, 0.73]** |

Heterogeneity: Tau² = 0.14; Chi² = 9.86, df = 3 (P = 0.02); I² = 70%

Test for overall effect: Z = 1.16 (P = 0.25)

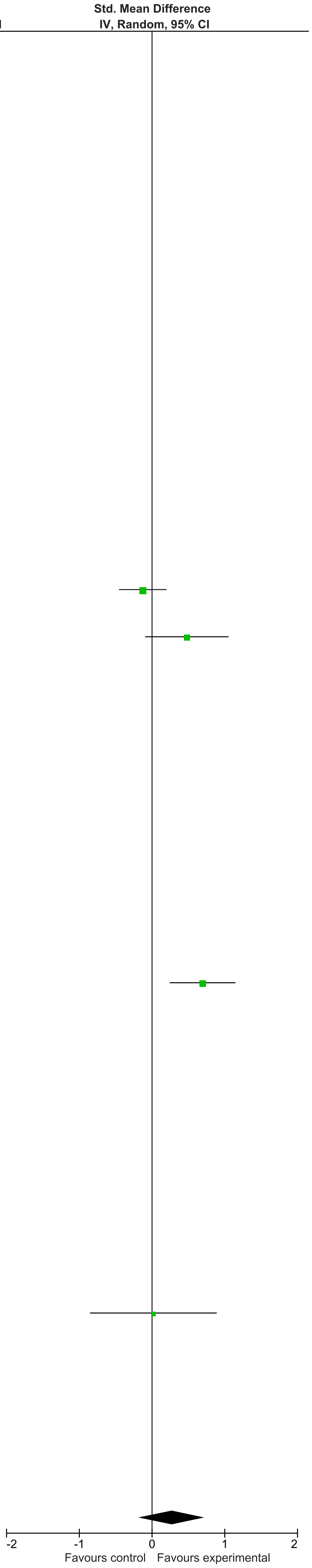

### eFigure 4. Sensitivity 0-1-month Follow-Up Forrest Plot

| | Experimental | | | Control | | | | Std. Mean Difference |
|---|---|---|---|---|---|---|---|---|
| **Study or Subgroup** | **Mean** | **SD** | **Total** | **Mean** | **SD** | **Total** | **Weight** | **IV, Random, 95% CI** |
| Akai 2008 | 3.16 | 1.16 | 23 | 2.62 | 1.02 | 25 | 2.0% | 0.49 [-0.09, 1.06] |
| Alarcao 2021 | 8.96 | 0.58 | 30 | 7.99 | 0.58 | 32 | 1.9% | 1.65 [1.07, 2.23] |
| Alvarenga 2019 | 4.57 | 5.23 | 22 | 1.82 | 2.98 | 22 | 1.8% | 0.63 [0.03, 1.24] |
| Badr 2006 | 0 | 0 | 0 | 0 | 0 | 0 | | Not estimable |
| Baggett 2010 | 0 | 0 | 0 | 0 | 0 | 0 | | Not estimable |
| Baggett 2017 | 0 | 0 | 0 | 0 | 0 | 0 | | Not estimable |
| Barlow 2016 | 9.29 | 2.61 | 14 | 7.08 | 2.53 | 13 | 1.3% | 0.83 [0.04, 1.63] |
| Barnes 2022 | 0 | 0 | 0 | 0 | 0 | 0 | | Not estimable |
| Berlin 2017 | 0 | 0 | 0 | 0 | 0 | 0 | | Not estimable |
| Berlin 2018 | 3.01 | 0.96 | 99 | 2.68 | 0.98 | 103 | 3.4% | 0.34 [0.06, 0.62] |
| Bernard 2012 | 0 | 0 | 0 | 0 | 0 | 0 | | Not estimable |
| Betancourt 2020 | 43.22 | 10.09 | 496 | 41.63 | 11.07 | 466 | 4.2% | 0.15 [0.02, 0.28] |
| Bick 2013 | 0 | 0 | 0 | 0 | 0 | 0 | | Not estimable |
| Borghini 2014 | 7.65 | 0.46 | 26 | 7.45 | 0.44 | 29 | 2.1% | 0.44 [-0.10, 0.97] |
| Brisch 2003 | 0 | 0 | 0 | 0 | 0 | 0 | | Not estimable |
| Browne 2005 | 0 | 0 | 0 | 0 | 0 | 0 | | Not estimable |
| Cates 2018 | 5.68 | 2.3 | 152 | 5.15 | 2.21 | 149 | 3.7% | 0.23 [0.01, 0.46] |
| Cevasco 2008 | 0 | 0 | 0 | 0 | 0 | 0 | | Not estimable |
| Chiu 2009 | 0 | 0 | 0 | 0 | 0 | 0 | | Not estimable |
| Constantino 2001 | 0 | 0 | 0 | 0 | 0 | 0 | | Not estimable |
| Cooper 2009 | 15.35 | 3.36 | 153 | 14.58 | 3.18 | 165 | 3.7% | 0.24 [0.01, 0.46] |
| Cooper 2015 | 3.33 | 0.62 | 79 | 3.3 | 0.67 | 77 | 3.2% | 0.05 [-0.27, 0.36] |
| Corrigan 2021 | 0 | 0 | 0 | 0 | 0 | 0 | | Not estimable |
| Dozier 2009 | 0 | 0 | 0 | 0 | 0 | 0 | | Not estimable |
| Eriksen 2018 | 0 | 0 | 0 | 0 | 0 | 0 | | Not estimable |
| Feil 2020 | 0.4 | 2.48 | 83 | -0.44 | 2.53 | 76 | 3.2% | 0.33 [0.02, 0.65] |
| Feldman 2014 | 0.53 | 0.23 | 73 | 0.44 | 0.21 | 73 | 3.1% | 0.41 [0.08, 0.73] |
| Firk 2021 | 3.59 | 0.9 | 27 | 3.6 | 1.2 | 20 | 1.9% | -0.01 [-0.59, 0.57] |
| Fonagy 2016 | 21.5 | 3.5 | 26 | 21.3 | 2.9 | 20 | 1.9% | 0.06 [-0.52, 0.64] |
| Gaden 2022 | 0 | 0 | 0 | 0 | 0 | 0 | | Not estimable |
| Georg 2021 | 23.09 | 2.87 | 81 | 22.24 | 3.34 | 73 | 3.2% | 0.27 [-0.05, 0.59] |
| Glazebrook 2007 | 36.6 | 5.1 | 94 | 37.4 | 4.9 | 102 | 3.4% | -0.16 [-0.44, 0.12] |
| Hane 2015 | 5.05 | 0.77 | 35 | 4.41 | 0.88 | 30 | 2.2% | 0.77 [0.26, 1.28] |
| Heo 2019 | 0 | 0 | 0 | 0 | 0 | 0 | | Not estimable |
| Herbers 2020 | 0 | 0 | 0 | 0 | 0 | 0 | | Not estimable |
| Hoffenkamp 2015 | 0 | 0 | 0 | 0 | 0 | 0 | | Not estimable |
| Hoivik 2015 | 0 | 0 | 0 | 0 | 0 | 0 | | Not estimable |
| Holditch-Davis 2014 | 0 | 0 | 0 | 0 | 0 | 0 | | Not estimable |
| Holt 2021 | 3.57 | 0.79 | 28 | 3.69 | 0.81 | 29 | 2.2% | -0.15 [-0.67, 0.37] |
| Horowitz 2001 | 0 | 0 | 0 | 0 | 0 | 0 | | Not estimable |
| Horowitz 2013 | 7.44 | 3.1 | 62 | 7.03 | 3.1 | 63 | 3.0% | 0.13 [-0.22, 0.48] |
| Juffer 2005 | 5.64 | 1.61 | 49 | 5.04 | 1.75 | 79 | 3.0% | 0.35 [-0.01, 0.71] |
| Klein Velderman 2006 | 0 | 0 | 0 | 0 | 0 | 0 | | Not estimable |
| Knoche 2012 | 3.92 | 0.56 | 37 | 3.63 | 0.78 | 24 | 2.2% | 0.44 [-0.08, 0.96] |
| Kristensen 2020 | 0 | 0 | 0 | 0 | 0 | 0 | | Not estimable |
| Landry 2006 | 0 | 0 | 0 | 0 | 0 | 0 | | Not estimable |
| Lavallee 2022 | 41.56 | 3.88 | 9 | 36.83 | 3.19 | 6 | 0.7% | 1.23 [0.07, 2.38] |
| Magil-Evans 2007 | 41.73 | 3.79 | 81 | 38.44 | 4.66 | 81 | 3.2% | 0.77 [0.45, 1.09] |
| Meijssen 2010 | 4.13 | 0.78 | 53 | 3.91 | 0.84 | 56 | 2.9% | 0.27 [-0.11, 0.65] |
| Melnyk 2006 | 9.1 | 3.54 | 116 | 8.38 | 3.55 | 95 | 3.4% | 0.20 [-0.07, 0.47] |
| Mendelsohn 2007 | 2.6 | 1.2 | 52 | 2.1 | 1.3 | 46 | 2.7% | 0.40 [-0.00, 0.80] |
| Milgrom 2013 | 2.08 | 1.97 | 54 | 1.7 | 1.93 | 50 | 2.8% | 0.19 [-0.19, 0.58] |
| Nelson 2001 | 0 | 0 | 0 | 0 | 0 | 0 | | Not estimable |
| Neu 2010 | 0 | 0 | 0 | 0 | 0 | 0 | | Not estimable |
| Newnham 2009 | 1.97 | 0.08 | 32 | 1.64 | 0.45 | 31 | 2.2% | 1.02 [0.49, 1.54] |
| Nugent 2017 | 0 | 0 | 0 | 0 | 0 | 0 | | Not estimable |
| Onozawa 2001 | 0 | 0 | 0 | 0 | 0 | 0 | | Not estimable |
| Oxford 2021 | 0 | 0 | 0 | 0 | 0 | 0 | | Not estimable |
| Perrone 2021 | 0 | 0 | 0 | 0 | 0 | 0 | | Not estimable |
| Pontoppidan 2016 | 0 | 0 | 0 | 0 | 0 | 0 | | Not estimable |
| Porter 2015 | 0 | 0 | 0 | 0 | 0 | 0 | | Not estimable |
| Ramsauer 2019 | 0 | 0 | 0 | 0 | 0 | 0 | | Not estimable |
| Ravn 2011 | 0 | 0 | 0 | 0 | 0 | 0 | | Not estimable |
| Rheinheimer 2022 | 6.19 | 1.84 | 56 | 5.95 | 2.07 | 60 | 2.9% | 0.12 [-0.24, 0.49] |
| Robertson 2019 | 0 | 0 | 0 | 0 | 0 | 0 | | Not estimable |
| Roby 2021 | 4.11 | 1.19 | 178 | 3.93 | 1.19 | 184 | 3.8% | 0.15 [-0.06, 0.36] |
| Sadler 2013 | 0 | 0 | 0 | 0 | 0 | 0 | | Not estimable |
| Sahlen Helmer 2020 | 0 | 0 | 0 | 0 | 0 | 0 | | Not estimable |
| Sajaniemi 2001 | 0 | 0 | 0 | 0 | 0 | 0 | | Not estimable |
| Salo 2019 | 0 | 0 | 0 | 0 | 0 | 0 | | Not estimable |
| Santelices 2010 | 0 | 0 | 0 | 0 | 0 | 0 | | Not estimable |
| Schuler 2000 | 3.6 | 0.4 | 84 | 3.3 | 0.4 | 87 | 3.2% | 0.75 [0.44, 1.06] |
| Shoghi 2018 | 57.76 | 4.2 | 20 | 46.23 | 4.35 | 20 | 1.1% | 2.64 [1.77, 3.51] |
| Sleed 2013 | 19.13 | 2.7 | 51 | 19.3 | 3.2 | 37 | 2.6% | -0.06 [-0.48, 0.37] |
| Steele 2019 | 0 | 0 | 0 | 0 | 0 | 0 | | Not estimable |
| Stein 2006 | 0 | 0 | 0 | 0 | 0 | 0 | | Not estimable |
| Stein 2018 | 0 | 0 | 0 | 0 | 0 | 0 | | Not estimable |
| Taneja 2020 | 0 | 0 | 0 | 0 | 0 | 0 | | Not estimable |
| Tereno 2017 | 0 | 0 | 0 | 0 | 0 | 0 | | Not estimable |
| Teti 2009 | 0 | 0 | 0 | 0 | 0 | 0 | | Not estimable |
| Tryphonopoulos 2020 | 0 | 0 | 0 | 0 | 0 | 0 | | Not estimable |
| Tsivos 2015 | 4.83 | 6.71 | 12 | 6.67 | 7.56 | 9 | 1.1% | -0.25 [-1.12, 0.62] |
| Twohig 2021 | 0 | 0 | 0 | 0 | 0 | 0 | | Not estimable |
| Valades 2021 | 3.42 | 0.99 | 31 | 2.7 | 0.95 | 30 | 2.2% | 0.73 [0.21, 1.25] |
| van Doesum 2008 | 4.82 | 1.78 | 35 | 3.79 | 1.86 | 36 | 2.4% | 0.56 [0.08, 1.03] |
| Vargas-Porras 2021 | 0 | 0 | 0 | 0 | 0 | 0 | | Not estimable |
| Walter 2019 | 0 | 0 | 0 | 0 | 0 | 0 | | Not estimable |
| White-Traut 2013 | 44.4 | 4.3 | 66 | 43.4 | 5.5 | 76 | 3.1% | 0.20 [-0.13, 0.53] |
| Williams 2020 | 0 | 0 | 0 | 0 | 0 | 0 | | Not estimable |
| Wulff 2021 | 0 | 0 | 0 | 0 | 0 | 0 | | Not estimable |
| Youn 2021 | 0 | 0 | 0 | 0 | 0 | 0 | | Not estimable |
| Yu 2022 | 0 | 0 | 0 | 0 | 0 | 0 | | Not estimable |
| Zelkowitz 2011 | 3.9 | 0.7 | 48 | 3.9 | 0.7 | 50 | 2.8% | 0.00 [-0.40, 0.40] |
| **Total (95% CI)** | | | **2667** | | | **2624** | **100.0%** | **0.36 [0.25, 0.47]** |

Heterogeneity: Tau² = 0.07; Chi² = 120.55, df = 37 (P < 0.00001); I² = 69%

Test for overall effect: Z = 6.47 (P < 0.00001)

Std. Mean Difference IV, Random, 95% CI

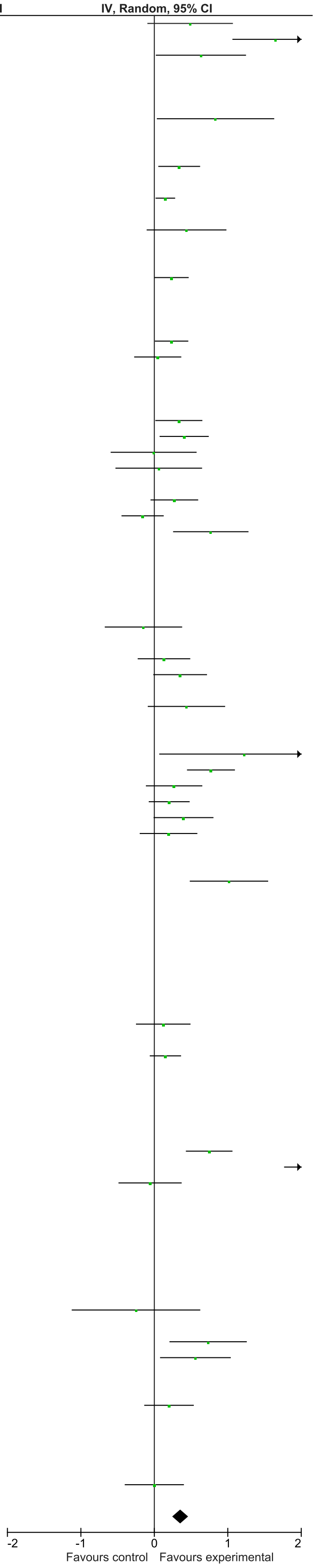

### eFigure 5. Sensitivity 2-6-month Follow-Up Forrest Plot

| | Early Dyadic Intervention | | | Standard Care | | | | Std. Mean Difference |
|---|---|---|---|---|---|---|---|---|
| Study or Subgroup | Mean | SD | Total | Mean | SD | Total | Weight | IV, Random, 95% CI |
| Akai 2008 | 0 | 0 | 0 | 0 | 0 | 0 | | Not estimable |
| Alarcao 2021 | 0 | 0 | 0 | 0 | 0 | 0 | | Not estimable |
| Alvarenga 2019 | 0 | 0 | 0 | 0 | 0 | 0 | | Not estimable |
| Badr 2006 | 0 | 0 | 0 | 0 | 0 | 0 | | Not estimable |
| Baggett 2010 | 0 | 0 | 0 | 0 | 0 | 0 | | Not estimable |
| Baggett 2017 | 0 | 0 | 0 | 0 | 0 | 0 | | Not estimable |
| Barlow 2016 | 0 | 0 | 0 | 0 | 0 | 0 | | Not estimable |
| Barnes 2022 | 0 | 0 | 0 | 0 | 0 | 0 | | Not estimable |
| Berlin 2017 | 0 | 0 | 0 | 0 | 0 | 0 | | Not estimable |
| Berlin 2018 | 0 | 0 | 0 | 0 | 0 | 0 | | Not estimable |
| Bernard 2012 | 0 | 0 | 0 | 0 | 0 | 0 | | Not estimable |
| Betancourt 2020 | 0 | 0 | 0 | 0 | 0 | 0 | | Not estimable |
| Bick 2013 | 0 | 0 | 0 | 0 | 0 | 0 | | Not estimable |
| Borghini 2014 | 0 | 0 | 0 | 0 | 0 | 0 | | Not estimable |
| Brisch 2003 | 0 | 0 | 0 | 0 | 0 | 0 | | Not estimable |
| Browne 2005 | 0 | 0 | 0 | 0 | 0 | 0 | | Not estimable |
| Cates 2018 | 0 | 0 | 0 | 0 | 0 | 0 | | Not estimable |
| Cevasco 2008 | 0 | 0 | 0 | 0 | 0 | 0 | | Not estimable |
| Chiu 2009 | 41.2 | 3.9 | 40 | 40.4 | 4.9 | 29 | 9.1% | 0.18 [-0.30, 0.66] |
| Constantino 2001 | 0 | 0 | 0 | 0 | 0 | 0 | | Not estimable |
| Cooper 2009 | 0 | 0 | 0 | 0 | 0 | 0 | | Not estimable |
| Cooper 2015 | 0 | 0 | 0 | 0 | 0 | 0 | | Not estimable |
| Corrigan 2021 | 0 | 0 | 0 | 0 | 0 | 0 | | Not estimable |
| Dozier 2009 | 0 | 0 | 0 | 0 | 0 | 0 | | Not estimable |
| Eriksen 2018 | 0 | 0 | 0 | 0 | 0 | 0 | | Not estimable |
| Feil 2020 | 0 | 0 | 0 | 0 | 0 | 0 | | Not estimable |
| Feldman 2014 | 0 | 0 | 0 | 0 | 0 | 0 | | Not estimable |
| Firk 2021 | 3.5 | 0.93 | 24 | 3.6 | 1.1 | 21 | 7.4% | -0.10 [-0.68, 0.49] |
| Fonagy 2016 | 0 | 0 | 0 | 0 | 0 | 0 | | Not estimable |
| Gaden 2022 | 0 | 0 | 0 | 0 | 0 | 0 | | Not estimable |
| Georg 2021 | 0 | 0 | 0 | 0 | 0 | 0 | | Not estimable |
| Glazebrook 2007 | 37.4 | 4.8 | 93 | 38.3 | 5.2 | 106 | 13.0% | -0.18 [-0.46, 0.10] |
| Hane 2015 | 0 | 0 | 0 | 0 | 0 | 0 | | Not estimable |
| Heo 2019 | 0 | 0 | 0 | 0 | 0 | 0 | | Not estimable |
| Herbers 2020 | 0 | 0 | 0 | 0 | 0 | 0 | | Not estimable |
| Hoffenkamp 2015 | 0 | 0 | 0 | 0 | 0 | 0 | | Not estimable |
| Hoivik 2015 | 0 | 0 | 0 | 0 | 0 | 0 | | Not estimable |
| Holditch-Davis 2014 | 0 | 0 | 0 | 0 | 0 | 0 | | Not estimable |
| Holt 2021 | 3.7 | 1.03 | 23 | 3.61 | 0.996 | 28 | 7.9% | 0.09 [-0.46, 0.64] |
| Horowitz 2001 | 0 | 0 | 0 | 0 | 0 | 0 | | Not estimable |
| Horowitz 2013 | 0 | 0 | 0 | 0 | 0 | 0 | | Not estimable |
| Juffer 2005 | 0 | 0 | 0 | 0 | 0 | 0 | | Not estimable |
| Klein Velderman 2006 | 0 | 0 | 0 | 0 | 0 | 0 | | Not estimable |
| Knoche 2012 | 0 | 0 | 0 | 0 | 0 | 0 | | Not estimable |
| Kristensen 2020 | 0 | 0 | 0 | 0 | 0 | 0 | | Not estimable |
| Landry 2006 | 0 | 0 | 0 | 0 | 0 | 0 | | Not estimable |
| Lavallee 2022 | 0 | 0 | 0 | 0 | 0 | 0 | | Not estimable |
| Magil-Evans 2007 | 0 | 0 | 0 | 0 | 0 | 0 | | Not estimable |
| Meijssen 2010 | 0 | 0 | 0 | 0 | 0 | 0 | | Not estimable |
| Melnyk 2006 | 0 | 0 | 0 | 0 | 0 | 0 | | Not estimable |
| Mendelsohn 2007 | 0 | 0 | 0 | 0 | 0 | 0 | | Not estimable |
| Milgrom 2013 | 0 | 0 | 0 | 0 | 0 | 0 | | Not estimable |
| Nelson 2001 | 43.7 | 4.37 | 10 | 45.75 | 4.95 | 9 | 4.1% | -0.42 [-1.33, 0.49] |
| Neu 2010 | 0 | 0 | 0 | 0 | 0 | 0 | | Not estimable |
| Newnham 2009 | 0 | 0 | 0 | 0 | 0 | 0 | | Not estimable |
| Nugent 2017 | 0 | 0 | 0 | 0 | 0 | 0 | | Not estimable |
| Onozawa 2001 | 0 | 0 | 0 | 0 | 0 | 0 | | Not estimable |
| Oxford 2021 | 33.86 | 5.22 | 127 | 32.38 | 4.74 | 125 | 13.6% | 0.30 [0.05, 0.54] |
| Perrone 2021 | 2.98 | 0.84 | 75 | 2.71 | 0.8 | 77 | 12.1% | 0.33 [0.01, 0.65] |
| Pontoppidan 2016 | 0 | 0 | 0 | 0 | 0 | 0 | | Not estimable |
| Porter 2015 | 0 | 0 | 0 | 0 | 0 | 0 | | Not estimable |
| Ramsauer 2019 | 0.81 | 0.16 | 31 | 0.83 | 0.14 | 30 | 8.7% | -0.13 [-0.63, 0.37] |
| Ravn 2011 | 0 | 0 | 0 | 0 | 0 | 0 | | Not estimable |
| Rheinheimer 2022 | 0 | 0 | 0 | 0 | 0 | 0 | | Not estimable |
| Robertson 2019 | 5.51 | 4.74 | 21 | 4.15 | 5.92 | 24 | 7.4% | 0.25 [-0.34, 0.84] |
| Roby 2021 | 0 | 0 | 0 | 0 | 0 | 0 | | Not estimable |
| Sadler 2013 | 0 | 0 | 0 | 0 | 0 | 0 | | Not estimable |
| Sahlen Helmer 2020 | 3 | 1.62 | 17 | 3.5 | 0.82 | 14 | 5.8% | -0.37 [-1.08, 0.35] |
| Sajaniemi 2001 | 0 | 0 | 0 | 0 | 0 | 0 | | Not estimable |
| Salo 2019 | 4.77 | 0.68 | 24 | 3.76 | 0.96 | 21 | 6.7% | 1.21 [0.57, 1.85] |
| Santelices 2010 | 0 | 0 | 0 | 0 | 0 | 0 | | Not estimable |
| Schuler 2000 | 0 | 0 | 0 | 0 | 0 | 0 | | Not estimable |
| Shoghi 2018 | 0 | 0 | 0 | 0 | 0 | 0 | | Not estimable |
| Sleed 2013 | 0 | 0 | 0 | 0 | 0 | 0 | | Not estimable |
| Steele 2019 | 0 | 0 | 0 | 0 | 0 | 0 | | Not estimable |
| Stein 2006 | 0 | 0 | 0 | 0 | 0 | 0 | | Not estimable |
| Stein 2018 | 0 | 0 | 0 | 0 | 0 | 0 | | Not estimable |
| Taneja 2020 | 0 | 0 | 0 | 0 | 0 | 0 | | Not estimable |
| Tereno 2017 | 0 | 0 | 0 | 0 | 0 | 0 | | Not estimable |
| Teti 2009 | 0 | 0 | 0 | 0 | 0 | 0 | | Not estimable |
| Tryphonopoulos 2020 | 0 | 0 | 0 | 0 | 0 | 0 | | Not estimable |
| Tsivos 2015 | 5 | 1.83 | 12 | 6.25 | 3.01 | 9 | 4.4% | -0.50 [-1.38, 0.38] |
| Twohig 2021 | 0 | 0 | 0 | 0 | 0 | 0 | | Not estimable |
| Valades 2021 | 0 | 0 | 0 | 0 | 0 | 0 | | Not estimable |
| van Doesum 2008 | 0 | 0 | 0 | 0 | 0 | 0 | | Not estimable |
| Vargas-Porras 2021 | 0 | 0 | 0 | 0 | 0 | 0 | | Not estimable |
| Walter 2019 | 0 | 0 | 0 | 0 | 0 | 0 | | Not estimable |
| White-Traut 2013 | 0 | 0 | 0 | 0 | 0 | 0 | | Not estimable |
| Williams 2020 | 0 | 0 | 0 | 0 | 0 | 0 | | Not estimable |
| Wulff 2021 | 0 | 0 | 0 | 0 | 0 | 0 | | Not estimable |
| Youn 2021 | 0 | 0 | 0 | 0 | 0 | 0 | | Not estimable |
| Yu 2022 | 0 | 0 | 0 | 0 | 0 | 0 | | Not estimable |
| Zelkowitz 2011 | 0 | 0 | 0 | 0 | 0 | 0 | | Not estimable |
| **Total (95% CI)** | | | **497** | | | **493** | **100.0%** | **0.10 [-0.11, 0.31]** |

Heterogeneity: Tau² = 0.07; Chi² = 25.77, df = 11 (P = 0.007); I² = 57%

Test for overall effect: Z = 0.91 (P = 0.36)

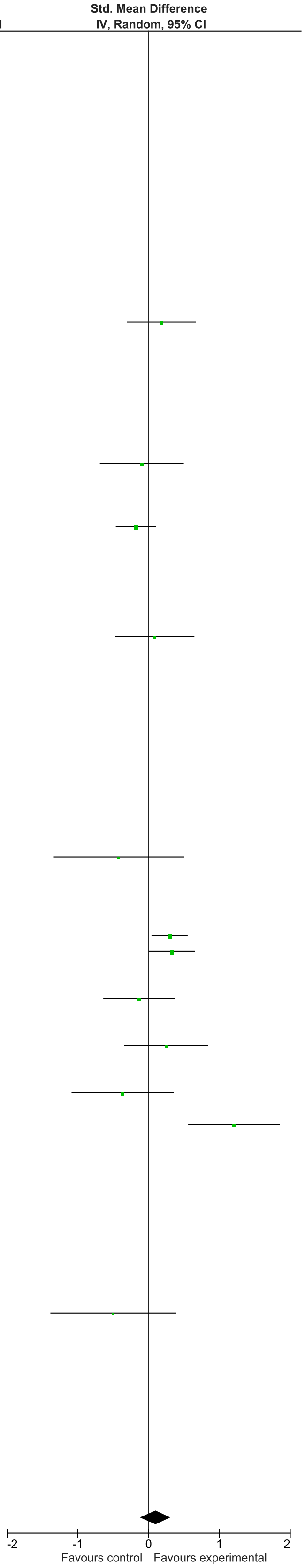

### eFigure 6. Sensitivity 7-12-month Follow-Up Forrest Plot

| | Early Dyadic Intervention | | | Standard Care | | | | Std. Mean Difference |
|---|---|---|---|---|---|---|---|---|
| Study or Subgroup | Mean | SD | Total | Mean | SD | Total | Weight | IV, Random, 95% CI |
| Akai 2008 | 0 | 0 | 0 | 0 | 0 | 0 | | Not estimable |
| Alarcao 2021 | 0 | 0 | 0 | 0 | 0 | 0 | | Not estimable |
| Alvarenga 2019 | 0 | 0 | 0 | 0 | 0 | 0 | | Not estimable |
| Badr 2006 | 0 | 0 | 0 | 0 | 0 | 0 | | Not estimable |
| Baggett 2010 | 0 | 0 | 0 | 0 | 0 | 0 | | Not estimable |
| Baggett 2017 | 0 | 0 | 0 | 0 | 0 | 0 | | Not estimable |
| Barlow 2016 | 0 | 0 | 0 | 0 | 0 | 0 | | Not estimable |
| Barnes 2022 | 0 | 0 | 0 | 0 | 0 | 0 | | Not estimable |
| Berlin 2017 | 0 | 0 | 0 | 0 | 0 | 0 | | Not estimable |
| Berlin 2018 | 0 | 0 | 0 | 0 | 0 | 0 | | Not estimable |
| Bernard 2012 | 0 | 0 | 0 | 0 | 0 | 0 | | Not estimable |
| Betancourt 2020 | 0 | 0 | 0 | 0 | 0 | 0 | | Not estimable |
| Bick 2013 | 0 | 0 | 0 | 0 | 0 | 0 | | Not estimable |
| Borghini 2014 | 0 | 0 | 0 | 0 | 0 | 0 | | Not estimable |
| Brisch 2003 | 0 | 0 | 0 | 0 | 0 | 0 | | Not estimable |
| Browne 2005 | 0 | 0 | 0 | 0 | 0 | 0 | | Not estimable |
| Cates 2018 | 0 | 0 | 0 | 0 | 0 | 0 | | Not estimable |
| Cevasco 2008 | 0 | 0 | 0 | 0 | 0 | 0 | | Not estimable |
| Chiu 2009 | 38.3 | 4.8 | 40 | 37.1 | 4.7 | 30 | 8.1% | 0.25 [-0.23, 0.72] |
| Constantino 2001 | 0 | 0 | 0 | 0 | 0 | 0 | | Not estimable |
| Cooper 2009 | 5.74 | 1.88 | 153 | 5.31 | 1.51 | 165 | 28.8% | 0.25 [0.03, 0.47] |
| Cooper 2015 | 0 | 0 | 0 | 0 | 0 | 0 | | Not estimable |
| Corrigan 2021 | 0 | 0 | 0 | 0 | 0 | 0 | | Not estimable |
| Dozier 2009 | 0 | 0 | 0 | 0 | 0 | 0 | | Not estimable |
| Eriksen 2018 | 0 | 0 | 0 | 0 | 0 | 0 | | Not estimable |
| Feil 2020 | 0 | 0 | 0 | 0 | 0 | 0 | | Not estimable |
| Feldman 2014 | 0 | 0 | 0 | 0 | 0 | 0 | | Not estimable |
| Firk 2021 | 0 | 0 | 0 | 0 | 0 | 0 | | Not estimable |
| Fonagy 2016 | 0 | 0 | 0 | 0 | 0 | 0 | | Not estimable |
| Gaden 2022 | 0 | 0 | 0 | 0 | 0 | 0 | | Not estimable |
| Georg 2021 | 0 | 0 | 0 | 0 | 0 | 0 | | Not estimable |
| Glazebrook 2007 | 0 | 0 | 0 | 0 | 0 | 0 | | Not estimable |
| Hane 2015 | 0 | 0 | 0 | 0 | 0 | 0 | | Not estimable |
| Heo 2019 | 0 | 0 | 0 | 0 | 0 | 0 | | Not estimable |
| Herbers 2020 | 0 | 0 | 0 | 0 | 0 | 0 | | Not estimable |
| Hoffenkamp 2015 | 0 | 0 | 0 | 0 | 0 | 0 | | Not estimable |
| Hoivik 2015 | 0 | 0 | 0 | 0 | 0 | 0 | | Not estimable |
| Holditch-Davis 2014 | 0 | 0 | 0 | 0 | 0 | 0 | | Not estimable |
| Holt 2021 | 0 | 0 | 0 | 0 | 0 | 0 | | Not estimable |
| Horowitz 2001 | 0 | 0 | 0 | 0 | 0 | 0 | | Not estimable |
| Horowitz 2013 | 0 | 0 | 0 | 0 | 0 | 0 | | Not estimable |
| Juffer 2005 | 0 | 0 | 0 | 0 | 0 | 0 | | Not estimable |
| Klein Velderman 2006 | 0 | 0 | 0 | 0 | 0 | 0 | | Not estimable |
| Knoche 2012 | 0 | 0 | 0 | 0 | 0 | 0 | | Not estimable |
| Kristensen 2020 | 0 | 0 | 0 | 0 | 0 | 0 | | Not estimable |
| Landry 2006 | 0 | 0 | 0 | 0 | 0 | 0 | | Not estimable |
| Lavallee 2022 | 0 | 0 | 0 | 0 | 0 | 0 | | Not estimable |
| Magil-Evans 2007 | 0 | 0 | 0 | 0 | 0 | 0 | | Not estimable |
| Meijssen 2010 | 0 | 0 | 0 | 0 | 0 | 0 | | Not estimable |
| Melnyk 2006 | 0 | 0 | 0 | 0 | 0 | 0 | | Not estimable |
| Mendelsohn 2007 | 0 | 0 | 0 | 0 | 0 | 0 | | Not estimable |
| Milgrom 2013 | 0 | 0 | 0 | 0 | 0 | 0 | | Not estimable |
| Nelson 2001 | 0 | 0 | 0 | 0 | 0 | 0 | | Not estimable |
| Neu 2010 | 0 | 0 | 0 | 0 | 0 | 0 | | Not estimable |
| Newnham 2009 | 0 | 0 | 0 | 0 | 0 | 0 | | Not estimable |
| Nugent 2017 | 0 | 0 | 0 | 0 | 0 | 0 | | Not estimable |
| Onozawa 2001 | 0 | 0 | 0 | 0 | 0 | 0 | | Not estimable |
| Oxford 2021 | 36.73 | 4.3 | 127 | 35.32 | 4.52 | 125 | 24.3% | 0.32 [0.07, 0.57] |
| Perrone 2021 | 0 | 0 | 0 | 0 | 0 | 0 | | Not estimable |
| Pontoppidan 2016 | 0 | 0 | 0 | 0 | 0 | 0 | | Not estimable |
| Porter 2015 | 0 | 0 | 0 | 0 | 0 | 0 | | Not estimable |
| Ramsauer 2019 | 0 | 0 | 0 | 0 | 0 | 0 | | Not estimable |
| Ravn 2011 | 0 | 0 | 0 | 0 | 0 | 0 | | Not estimable |
| Rheinheimer 2022 | 0 | 0 | 0 | 0 | 0 | 0 | | Not estimable |
| Robertson 2019 | 0 | 0 | 0 | 0 | 0 | 0 | | Not estimable |
| Roby 2021 | 0 | 0 | 0 | 0 | 0 | 0 | | Not estimable |
| Sadler 2013 | 0 | 0 | 0 | 0 | 0 | 0 | | Not estimable |
| Sahlen Helmer 2020 | 0 | 0 | 0 | 0 | 0 | 0 | | Not estimable |
| Sajaniemi 2001 | 0 | 0 | 0 | 0 | 0 | 0 | | Not estimable |
| Salo 2019 | 0 | 0 | 0 | 0 | 0 | 0 | | Not estimable |
| Santelices 2010 | 0 | 0 | 0 | 0 | 0 | 0 | | Not estimable |
| Schuler 2000 | 2.8 | 0.4 | 67 | 2.7 | 0.4 | 64 | 14.4% | 0.25 [-0.10, 0.59] |
| Shoghi 2018 | 0 | 0 | 0 | 0 | 0 | 0 | | Not estimable |
| Sleed 2013 | 0 | 0 | 0 | 0 | 0 | 0 | | Not estimable |
| Steele 2019 | 0 | 0 | 0 | 0 | 0 | 0 | | Not estimable |
| Stein 2006 | 142.65 | 70.08 | 38 | 141.85 | 67.79 | 39 | 9.1% | 0.01 [-0.44, 0.46] |
| Stein 2018 | 0 | 0 | 0 | 0 | 0 | 0 | | Not estimable |
| Taneja 2020 | 0 | 0 | 0 | 0 | 0 | 0 | | Not estimable |
| Tereno 2017 | 0 | 0 | 0 | 0 | 0 | 0 | | Not estimable |
| Teti 2009 | 0 | 0 | 0 | 0 | 0 | 0 | | Not estimable |
| Tryphonopoulos 2020 | 0 | 0 | 0 | 0 | 0 | 0 | | Not estimable |
| Tsivos 2015 | 0 | 0 | 0 | 0 | 0 | 0 | | Not estimable |
| Twohig 2021 | 8.35 | 3.54 | 29 | 8.19 | 3.5 | 32 | 7.4% | 0.04 [-0.46, 0.55] |
| Valades 2021 | 0 | 0 | 0 | 0 | 0 | 0 | | Not estimable |
| van Doesum 2008 | 5.18 | 2.01 | 35 | 3.63 | 1.76 | 36 | 7.8% | 0.81 [0.33, 1.30] |
| Vargas-Porras 2021 | 0 | 0 | 0 | 0 | 0 | 0 | | Not estimable |
| Walter 2019 | 0 | 0 | 0 | 0 | 0 | 0 | | Not estimable |
| White-Traut 2013 | 0 | 0 | 0 | 0 | 0 | 0 | | Not estimable |
| Williams 2020 | 0 | 0 | 0 | 0 | 0 | 0 | | Not estimable |
| Wulff 2021 | 0 | 0 | 0 | 0 | 0 | 0 | | Not estimable |
| Youn 2021 | 0 | 0 | 0 | 0 | 0 | 0 | | Not estimable |
| Yu 2022 | 0 | 0 | 0 | 0 | 0 | 0 | | Not estimable |
| Zelkowitz 2011 | 0 | 0 | 0 | 0 | 0 | 0 | | Not estimable |
| **Total (95% CI)** | | | **489** | | | **491** | **100.0%** | **0.27 [0.13, 0.42]** |

Heterogeneity: $Tau^2 = 0.01$; $Chi^2 = 7.05$, df = 6 (P = 0.32); $I^2 = 15\%$
Test for overall effect: Z = 3.79 (P = 0.0001)

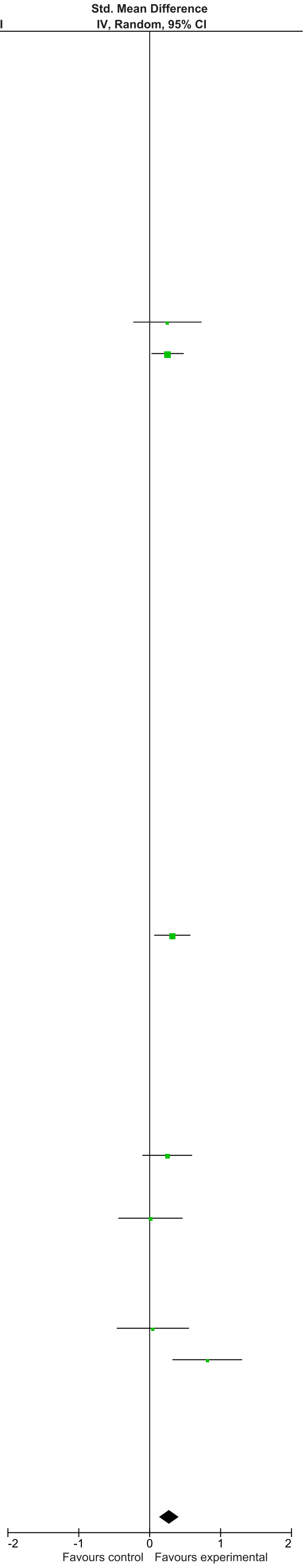

### eFigure 7. Sensitivity 15+ months Follow-Up Forrest Plot

| | Early Dyadic Intervention | | | Standard Care | | | | Std. Mean Difference |
|---|---|---|---|---|---|---|---|---|
| Study or Subgroup | Mean | SD | Total | Mean | SD | Total | Weight | IV, Random, 95% CI |
| Akai 2008 | 0 | 0 | 0 | 0 | 0 | 0 | | Not estimable |
| Alarcao 2021 | 0 | 0 | 0 | 0 | 0 | 0 | | Not estimable |
| Alvarenga 2019 | 0 | 0 | 0 | 0 | 0 | 0 | | Not estimable |
| Badr 2006 | 0 | 0 | 0 | 0 | 0 | 0 | | Not estimable |
| Baggett 2010 | 0 | 0 | 0 | 0 | 0 | 0 | | Not estimable |
| Baggett 2017 | 0 | 0 | 0 | 0 | 0 | 0 | | Not estimable |
| Barlow 2016 | 0 | 0 | 0 | 0 | 0 | 0 | | Not estimable |
| Barnes 2022 | 0 | 0 | 0 | 0 | 0 | 0 | | Not estimable |
| Berlin 2017 | 0 | 0 | 0 | 0 | 0 | 0 | | Not estimable |
| Berlin 2018 | 0 | 0 | 0 | 0 | 0 | 0 | | Not estimable |
| Bernard 2012 | 0 | 0 | 0 | 0 | 0 | 0 | | Not estimable |
| Betancourt 2020 | 0 | 0 | 0 | 0 | 0 | 0 | | Not estimable |
| Bick 2013 | 0 | 0 | 0 | 0 | 0 | 0 | | Not estimable |
| Borghini 2014 | 0 | 0 | 0 | 0 | 0 | 0 | | Not estimable |
| Brisch 2003 | 0 | 0 | 0 | 0 | 0 | 0 | | Not estimable |
| Browne 2005 | 0 | 0 | 0 | 0 | 0 | 0 | | Not estimable |
| Cates 2018 | 9.2 | 3.63 | 123 | 8.1 | 3.77 | 130 | 76.9% | 0.30 [0.05, 0.54] |
| Cevasco 2008 | 0 | 0 | 0 | 0 | 0 | 0 | | Not estimable |
| Chiu 2009 | 35.5 | 13.3 | 42 | 34.2 | 14.4 | 34 | 23.1% | 0.09 [-0.36, 0.55] |
| Constantino 2001 | 0 | 0 | 0 | 0 | 0 | 0 | | Not estimable |
| Cooper 2009 | 0 | 0 | 0 | 0 | 0 | 0 | | Not estimable |
| Cooper 2015 | 0 | 0 | 0 | 0 | 0 | 0 | | Not estimable |
| Corrigan 2021 | 0 | 0 | 0 | 0 | 0 | 0 | | Not estimable |
| Dozier 2009 | 0 | 0 | 0 | 0 | 0 | 0 | | Not estimable |
| Eriksen 2018 | 0 | 0 | 0 | 0 | 0 | 0 | | Not estimable |
| Feil 2020 | 0 | 0 | 0 | 0 | 0 | 0 | | Not estimable |
| Feldman 2014 | 0 | 0 | 0 | 0 | 0 | 0 | | Not estimable |
| Firk 2021 | 0 | 0 | 0 | 0 | 0 | 0 | | Not estimable |
| Fonagy 2016 | 0 | 0 | 0 | 0 | 0 | 0 | | Not estimable |
| Gaden 2022 | 0 | 0 | 0 | 0 | 0 | 0 | | Not estimable |
| Georg 2021 | 0 | 0 | 0 | 0 | 0 | 0 | | Not estimable |
| Glazebrook 2007 | 0 | 0 | 0 | 0 | 0 | 0 | | Not estimable |
| Hane 2015 | 0 | 0 | 0 | 0 | 0 | 0 | | Not estimable |
| Heo 2019 | 0 | 0 | 0 | 0 | 0 | 0 | | Not estimable |
| Herbers 2020 | 0 | 0 | 0 | 0 | 0 | 0 | | Not estimable |
| Hoffenkamp 2015 | 0 | 0 | 0 | 0 | 0 | 0 | | Not estimable |
| Hoivik 2015 | 0 | 0 | 0 | 0 | 0 | 0 | | Not estimable |
| Holditch-Davis 2014 | 0 | 0 | 0 | 0 | 0 | 0 | | Not estimable |
| Holt 2021 | 0 | 0 | 0 | 0 | 0 | 0 | | Not estimable |
| Horowitz 2001 | 0 | 0 | 0 | 0 | 0 | 0 | | Not estimable |
| Horowitz 2013 | 0 | 0 | 0 | 0 | 0 | 0 | | Not estimable |
| Juffer 2005 | 0 | 0 | 0 | 0 | 0 | 0 | | Not estimable |
| Klein Velderman 2006 | 0 | 0 | 0 | 0 | 0 | 0 | | Not estimable |
| Knoche 2012 | 0 | 0 | 0 | 0 | 0 | 0 | | Not estimable |
| Kristensen 2020 | 0 | 0 | 0 | 0 | 0 | 0 | | Not estimable |
| Landry 2006 | 0 | 0 | 0 | 0 | 0 | 0 | | Not estimable |
| Lavallee 2022 | 0 | 0 | 0 | 0 | 0 | 0 | | Not estimable |
| Magil-Evans 2007 | 0 | 0 | 0 | 0 | 0 | 0 | | Not estimable |
| Meijssen 2010 | 0 | 0 | 0 | 0 | 0 | 0 | | Not estimable |
| Melnyk 2006 | 0 | 0 | 0 | 0 | 0 | 0 | | Not estimable |
| Mendelsohn 2007 | 0 | 0 | 0 | 0 | 0 | 0 | | Not estimable |
| Milgrom 2013 | 0 | 0 | 0 | 0 | 0 | 0 | | Not estimable |
| Nelson 2001 | 0 | 0 | 0 | 0 | 0 | 0 | | Not estimable |
| Neu 2010 | 0 | 0 | 0 | 0 | 0 | 0 | | Not estimable |
| Newnham 2009 | 0 | 0 | 0 | 0 | 0 | 0 | | Not estimable |
| Nugent 2017 | 0 | 0 | 0 | 0 | 0 | 0 | | Not estimable |
| Onozawa 2001 | 0 | 0 | 0 | 0 | 0 | 0 | | Not estimable |
| Oxford 2021 | 0 | 0 | 0 | 0 | 0 | 0 | | Not estimable |
| Perrone 2021 | 0 | 0 | 0 | 0 | 0 | 0 | | Not estimable |
| Pontoppidan 2016 | 0 | 0 | 0 | 0 | 0 | 0 | | Not estimable |
| Porter 2015 | 0 | 0 | 0 | 0 | 0 | 0 | | Not estimable |
| Ramsauer 2019 | 0 | 0 | 0 | 0 | 0 | 0 | | Not estimable |
| Ravn 2011 | 0 | 0 | 0 | 0 | 0 | 0 | | Not estimable |
| Rheinheimer 2022 | 0 | 0 | 0 | 0 | 0 | 0 | | Not estimable |
| Robertson 2019 | 0 | 0 | 0 | 0 | 0 | 0 | | Not estimable |
| Roby 2021 | 0 | 0 | 0 | 0 | 0 | 0 | | Not estimable |
| Sadler 2013 | 0 | 0 | 0 | 0 | 0 | 0 | | Not estimable |
| Sahlen Helmer 2020 | 0 | 0 | 0 | 0 | 0 | 0 | | Not estimable |
| Sajaniemi 2001 | 0 | 0 | 0 | 0 | 0 | 0 | | Not estimable |
| Salo 2019 | 0 | 0 | 0 | 0 | 0 | 0 | | Not estimable |
| Santelices 2010 | 0 | 0 | 0 | 0 | 0 | 0 | | Not estimable |
| Schuler 2000 | 0 | 0 | 0 | 0 | 0 | 0 | | Not estimable |
| Shoghi 2018 | 0 | 0 | 0 | 0 | 0 | 0 | | Not estimable |
| Sleed 2013 | 0 | 0 | 0 | 0 | 0 | 0 | | Not estimable |
| Steele 2019 | 0 | 0 | 0 | 0 | 0 | 0 | | Not estimable |
| Stein 2006 | 0 | 0 | 0 | 0 | 0 | 0 | | Not estimable |
| Stein 2018 | 0 | 0 | 0 | 0 | 0 | 0 | | Not estimable |
| Taneja 2020 | 0 | 0 | 0 | 0 | 0 | 0 | | Not estimable |
| Tereno 2017 | 0 | 0 | 0 | 0 | 0 | 0 | | Not estimable |
| Teti 2009 | 0 | 0 | 0 | 0 | 0 | 0 | | Not estimable |
| Tryphonopoulos 2020 | 0 | 0 | 0 | 0 | 0 | 0 | | Not estimable |
| Tsivos 2015 | 0 | 0 | 0 | 0 | 0 | 0 | | Not estimable |
| Twohig 2021 | 0 | 0 | 0 | 0 | 0 | 0 | | Not estimable |
| Valades 2021 | 0 | 0 | 0 | 0 | 0 | 0 | | Not estimable |
| van Doesum 2008 | 0 | 0 | 0 | 0 | 0 | 0 | | Not estimable |
| Vargas-Porras 2021 | 0 | 0 | 0 | 0 | 0 | 0 | | Not estimable |
| Walter 2019 | 0 | 0 | 0 | 0 | 0 | 0 | | Not estimable |
| White-Traut 2013 | 0 | 0 | 0 | 0 | 0 | 0 | | Not estimable |
| Williams 2020 | 0 | 0 | 0 | 0 | 0 | 0 | | Not estimable |
| Wulff 2021 | 0 | 0 | 0 | 0 | 0 | 0 | | Not estimable |
| Youn 2021 | 0 | 0 | 0 | 0 | 0 | 0 | | Not estimable |
| Yu 2022 | 0 | 0 | 0 | 0 | 0 | 0 | | Not estimable |
| Zelkowitz 2011 | 0 | 0 | 0 | 0 | 0 | 0 | | Not estimable |
| **Total (95% CI)** | | | **165** | | | **164** | **100.0%** | **0.25 [0.03, 0.47]** |

Heterogeneity: Tau² = 0.00; Chi² = 0.59, df = 1 (P = 0.44); I² = 0%

Test for overall effect: Z = 2.25 (P = 0.02)

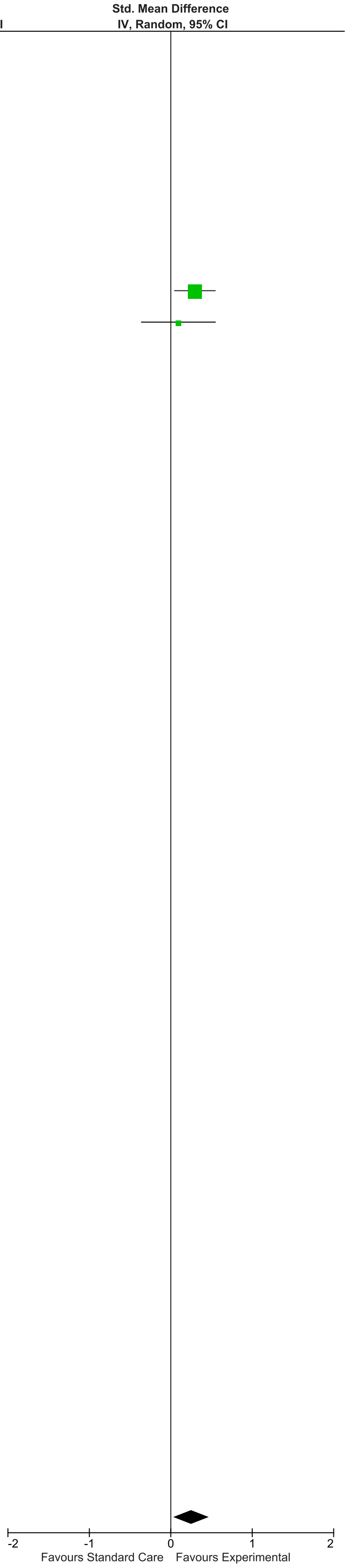

### eFigure 8. Secure Attachment 0-1-month Follow-Up Forrest Plot

| Study or Subgroup | Early Dyadic Intervention Events | Early Dyadic Intervention Total | Standard Care Events | Standard Care Total | Weight | Odds Ratio IV, Random, 95% CI |
|---|---|---|---|---|---|---|
| Akai 2008 | 0 | 0 | 0 | 0 | | Not estimable |
| Alarcao 2021 | 18 | 28 | 10 | 26 | 9.7% | 2.88 [0.95, 8.70] |
| Alvarenga 2019 | 0 | 0 | 0 | 0 | | Not estimable |
| Badr 2006 | 0 | 0 | 0 | 0 | | Not estimable |
| Baggett 2010 | 0 | 0 | 0 | 0 | | Not estimable |
| Baggett 2017 | 0 | 0 | 0 | 0 | | Not estimable |
| Barlow 2016 | 0 | 0 | 0 | 0 | | Not estimable |
| Barnes 2022 | 0 | 0 | 0 | 0 | | Not estimable |
| Berlin 2017 | 34 | 67 | 12 | 27 | 14.7% | 1.29 [0.52, 3.16] |
| Berlin 2018 | 0 | 0 | 0 | 0 | | Not estimable |
| Bernard 2012 | 31 | 60 | 20 | 60 | 21.7% | 2.14 [1.02, 4.47] |
| Betancourt 2020 | 0 | 0 | 0 | 0 | | Not estimable |
| Bick 2013 | 0 | 0 | 0 | 0 | | Not estimable |
| Borghini 2014 | 0 | 0 | 0 | 0 | | Not estimable |
| Brisch 2003 | 0 | 0 | 0 | 0 | | Not estimable |
| Browne 2005 | 0 | 0 | 0 | 0 | | Not estimable |
| Cates 2018 | 0 | 0 | 0 | 0 | | Not estimable |
| Cevasco 2008 | 0 | 0 | 0 | 0 | | Not estimable |
| Chiu 2009 | 0 | 0 | 0 | 0 | | Not estimable |
| Constantino 2001 | 0 | 0 | 0 | 0 | | Not estimable |
| Cooper 2009 | 0 | 0 | 0 | 0 | | Not estimable |
| Cooper 2015 | 0 | 0 | 0 | 0 | | Not estimable |
| Corrigan 2021 | 0 | 0 | 0 | 0 | | Not estimable |
| Dozier 2009 | 0 | 0 | 0 | 0 | | Not estimable |
| Eriksen 2018 | 0 | 0 | 0 | 0 | | Not estimable |
| Feil 2020 | 0 | 0 | 0 | 0 | | Not estimable |
| Feldman 2014 | 0 | 0 | 0 | 0 | | Not estimable |
| Firk 2021 | 10 | 25 | 10 | 19 | 8.1% | 0.60 [0.18, 2.00] |
| Fonagy 2016 | 22 | 28 | 17 | 25 | 7.8% | 1.73 [0.50, 5.92] |
| Gaden 2022 | 0 | 0 | 0 | 0 | | Not estimable |
| Georg 2021 | 0 | 0 | 0 | 0 | | Not estimable |
| Glazebrook 2007 | 0 | 0 | 0 | 0 | | Not estimable |
| Hane 2015 | 0 | 0 | 0 | 0 | | Not estimable |
| Heo 2019 | 0 | 0 | 0 | 0 | | Not estimable |
| Herbers 2020 | 0 | 0 | 0 | 0 | | Not estimable |
| Hoffenkamp 2015 | 0 | 0 | 0 | 0 | | Not estimable |
| Hoivik 2015 | 0 | 0 | 0 | 0 | | Not estimable |
| Holditch-Davis 2014 | 0 | 0 | 0 | 0 | | Not estimable |
| Holt 2021 | 0 | 0 | 0 | 0 | | Not estimable |
| Horowitz 2001 | 0 | 0 | 0 | 0 | | Not estimable |
| Horowitz 2013 | 0 | 0 | 0 | 0 | | Not estimable |
| Juffer 2005 | 0 | 0 | 0 | 0 | | Not estimable |
| Klein Velderman 2006 | 36 | 54 | 15 | 27 | 13.2% | 1.60 [0.62, 4.12] |
| Knoche 2012 | 0 | 0 | 0 | 0 | | Not estimable |
| Kristensen 2020 | 0 | 0 | 0 | 0 | | Not estimable |
| Landry 2006 | 0 | 0 | 0 | 0 | | Not estimable |
| Lavallee 2022 | 0 | 0 | 0 | 0 | | Not estimable |
| Magil-Evans 2007 | 0 | 0 | 0 | 0 | | Not estimable |
| Meijssen 2010 | 0 | 0 | 0 | 0 | | Not estimable |
| Melnyk 2006 | 0 | 0 | 0 | 0 | | Not estimable |
| Mendelsohn 2007 | 0 | 0 | 0 | 0 | | Not estimable |
| Milgrom 2013 | 0 | 0 | 0 | 0 | | Not estimable |
| Nelson 2001 | 0 | 0 | 0 | 0 | | Not estimable |
| Neu 2010 | 0 | 0 | 0 | 0 | | Not estimable |
| Newnham 2009 | 0 | 0 | 0 | 0 | | Not estimable |
| Nugent 2017 | 0 | 0 | 0 | 0 | | Not estimable |
| Onozawa 2001 | 0 | 0 | 0 | 0 | | Not estimable |
| Oxford 2021 | 0 | 0 | 0 | 0 | | Not estimable |
| Perrone 2021 | 0 | 0 | 0 | 0 | | Not estimable |
| Pontoppidan 2016 | 0 | 0 | 0 | 0 | | Not estimable |
| Porter 2015 | 0 | 0 | 0 | 0 | | Not estimable |
| Ramsauer 2019 | 0 | 0 | 0 | 0 | | Not estimable |
| Ravn 2011 | 0 | 0 | 0 | 0 | | Not estimable |
| Rheinheimer 2022 | 0 | 0 | 0 | 0 | | Not estimable |
| Robertson 2019 | 0 | 0 | 0 | 0 | | Not estimable |
| Roby 2021 | 0 | 0 | 0 | 0 | | Not estimable |
| Sadler 2013 | 26 | 41 | 15 | 30 | 12.9% | 1.73 [0.67, 4.51] |
| Sahlen Helmer 2020 | 0 | 0 | 0 | 0 | | Not estimable |
| Sajaniemi 2001 | 0 | 0 | 0 | 0 | | Not estimable |
| Salo 2019 | 0 | 0 | 0 | 0 | | Not estimable |
| Santelices 2010 | 31 | 43 | 16 | 29 | 12.1% | 2.10 [0.78, 5.65] |
| Schuler 2000 | 0 | 0 | 0 | 0 | | Not estimable |
| Shoghi 2018 | 0 | 0 | 0 | 0 | | Not estimable |
| Sleed 2013 | 0 | 0 | 0 | 0 | | Not estimable |
| Steele 2019 | 0 | 0 | 0 | 0 | | Not estimable |
| Stein 2006 | 0 | 0 | 0 | 0 | | Not estimable |
| Stein 2018 | 0 | 0 | 0 | 0 | | Not estimable |
| Taneja 2020 | 0 | 0 | 0 | 0 | | Not estimable |
| Tereno 2017 | 0 | 0 | 0 | 0 | | Not estimable |
| Teti 2009 | 0 | 0 | 0 | 0 | | Not estimable |
| Tryphonopoulos 2020 | 0 | 0 | 0 | 0 | | Not estimable |
| Tsivos 2015 | 0 | 0 | 0 | 0 | | Not estimable |
| Twohig 2021 | 0 | 0 | 0 | 0 | | Not estimable |
| Valades 2021 | 0 | 0 | 0 | 0 | | Not estimable |
| van Doesum 2008 | 0 | 0 | 0 | 0 | | Not estimable |
| Vargas-Porras 2021 | 0 | 0 | 0 | 0 | | Not estimable |
| Walter 2019 | 0 | 0 | 0 | 0 | | Not estimable |
| White-Traut 2013 | 0 | 0 | 0 | 0 | | Not estimable |
| Williams 2020 | 0 | 0 | 0 | 0 | | Not estimable |
| Wulff 2021 | 0 | 0 | 0 | 0 | | Not estimable |
| Youn 2021 | 0 | 0 | 0 | 0 | | Not estimable |
| Yu 2022 | 0 | 0 | 0 | 0 | | Not estimable |
| **Total (95% CI)** | | **346** | | **243** | **100.0%** | **1.69 [1.20, 2.39]** |
| Total events | 208 | | 115 | | | |

Heterogeneity: Tau² = 0.00; Chi² = 4.67, df = 7 (P = 0.70); I² = 0%

Test for overall effect: Z = 3.01 (P = 0.003)

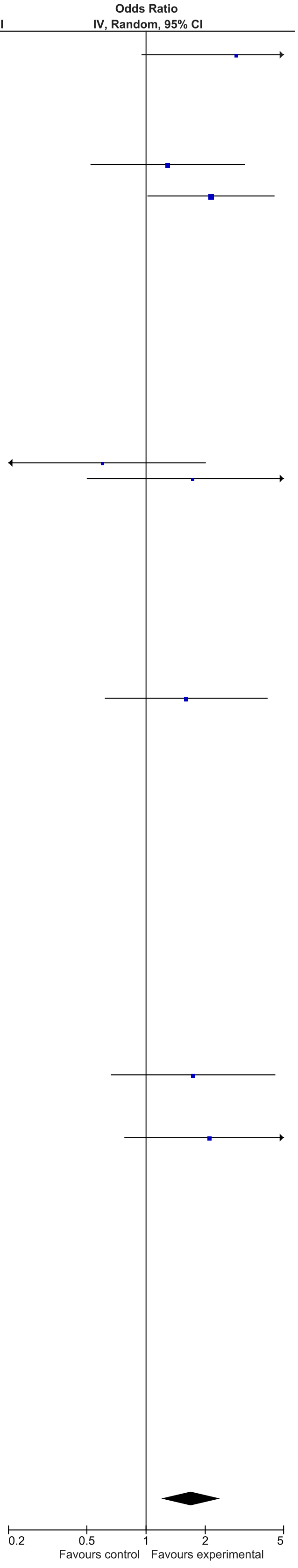

### eFigure 9. Secute Attachment 2-6-month Follow-Up Forrest Plot

| Study or Subgroup | Early Dyadic Intervention Events | Early Dyadic Intervention Total | Standard Care Events | Standard Care Total | Weight | Odds Ratio IV, Random, 95% CI |
|---|---|---|---|---|---|---|
| Akai 2008 | 0 | 0 | 0 | 0 | | Not estimable |
| Alarcao 2021 | 0 | 0 | 0 | 0 | | Not estimable |
| Alvarenga 2019 | 0 | 0 | 0 | 0 | | Not estimable |
| Badr 2006 | 0 | 0 | 0 | 0 | | Not estimable |
| Baggett 2010 | 0 | 0 | 0 | 0 | | Not estimable |
| Baggett 2017 | 0 | 0 | 0 | 0 | | Not estimable |
| Barlow 2016 | 0 | 0 | 0 | 0 | | Not estimable |
| Barnes 2022 | 0 | 0 | 0 | 0 | | Not estimable |
| Berlin 2017 | 0 | 0 | 0 | 0 | | Not estimable |
| Berlin 2018 | 0 | 0 | 0 | 0 | | Not estimable |
| Bernard 2012 | 0 | 0 | 0 | 0 | | Not estimable |
| Betancourt 2020 | 0 | 0 | 0 | 0 | | Not estimable |
| Bick 2013 | 0 | 0 | 0 | 0 | | Not estimable |
| Borghini 2014 | 0 | 0 | 0 | 0 | | Not estimable |
| Brisch 2003 | 0 | 0 | 0 | 0 | | Not estimable |
| Browne 2005 | 0 | 0 | 0 | 0 | | Not estimable |
| Cates 2018 | 0 | 0 | 0 | 0 | | Not estimable |
| Cevasco 2008 | 0 | 0 | 0 | 0 | | Not estimable |
| Chiu 2009 | 0 | 0 | 0 | 0 | | Not estimable |
| Constantino 2001 | 0 | 0 | 0 | 0 | | Not estimable |
| Cooper 2009 | 0 | 0 | 0 | 0 | | Not estimable |
| Cooper 2015 | 0 | 0 | 0 | 0 | | Not estimable |
| Corrigan 2021 | 0 | 0 | 0 | 0 | | Not estimable |
| Dozier 2009 | 0 | 0 | 0 | 0 | | Not estimable |
| Eriksen 2018 | 0 | 0 | 0 | 0 | | Not estimable |
| Feil 2020 | 0 | 0 | 0 | 0 | | Not estimable |
| Feldman 2014 | 0 | 0 | 0 | 0 | | Not estimable |
| Firk 2021 | 3 | 24 | 2 | 21 | 9.9% | 1.36 [0.20, 9.02] |
| Fonagy 2016 | 0 | 0 | 0 | 0 | | Not estimable |
| Gaden 2022 | 0 | 0 | 0 | 0 | | Not estimable |
| Georg 2021 | 0 | 0 | 0 | 0 | | Not estimable |
| Glazebrook 2007 | 0 | 0 | 0 | 0 | | Not estimable |
| Hane 2015 | 0 | 0 | 0 | 0 | | Not estimable |
| Heo 2019 | 0 | 0 | 0 | 0 | | Not estimable |
| Herbers 2020 | 0 | 0 | 0 | 0 | | Not estimable |
| Hoffenkamp 2015 | 0 | 0 | 0 | 0 | | Not estimable |
| Hoivik 2015 | 0 | 0 | 0 | 0 | | Not estimable |
| Holditch-Davis 2014 | 0 | 0 | 0 | 0 | | Not estimable |
| Holt 2021 | 0 | 0 | 0 | 0 | | Not estimable |
| Horowitz 2001 | 0 | 0 | 0 | 0 | | Not estimable |
| Horowitz 2013 | 0 | 0 | 0 | 0 | | Not estimable |
| Juffer 2005 | 0 | 0 | 0 | 0 | | Not estimable |
| Klein Velderman 2006 | 0 | 0 | 0 | 0 | | Not estimable |
| Knoche 2012 | 0 | 0 | 0 | 0 | | Not estimable |
| Kristensen 2020 | 0 | 0 | 0 | 0 | | Not estimable |
| Landry 2006 | 0 | 0 | 0 | 0 | | Not estimable |
| Lavallee 2022 | 0 | 0 | 0 | 0 | | Not estimable |
| Magil-Evans 2007 | 0 | 0 | 0 | 0 | | Not estimable |
| Meijssen 2010 | 0 | 0 | 0 | 0 | | Not estimable |
| Melnyk 2006 | 0 | 0 | 0 | 0 | | Not estimable |
| Mendelsohn 2007 | 0 | 0 | 0 | 0 | | Not estimable |
| Milgrom 2013 | 0 | 0 | 0 | 0 | | Not estimable |
| Nelson 2001 | 0 | 0 | 0 | 0 | | Not estimable |
| Neu 2010 | 0 | 0 | 0 | 0 | | Not estimable |
| Newnham 2009 | 0 | 0 | 0 | 0 | | Not estimable |
| Nugent 2017 | 0 | 0 | 0 | 0 | | Not estimable |
| Onozawa 2001 | 0 | 0 | 0 | 0 | | Not estimable |
| Oxford 2021 | 0 | 0 | 0 | 0 | | Not estimable |
| Perrone 2021 | 0 | 0 | 0 | 0 | | Not estimable |
| Pontoppidan 2016 | 0 | 0 | 0 | 0 | | Not estimable |
| Porter 2015 | 0 | 0 | 0 | 0 | | Not estimable |
| Ramsauer 2019 | 21 | 34 | 18 | 31 | 35.9% | 1.17 [0.43, 3.15] |
| Ravn 2011 | 0 | 0 | 0 | 0 | | Not estimable |
| Rheinheimer 2022 | 0 | 0 | 0 | 0 | | Not estimable |
| Robertson 2019 | 0 | 0 | 0 | 0 | | Not estimable |
| Roby 2021 | 0 | 0 | 0 | 0 | | Not estimable |
| Sadler 2013 | 0 | 0 | 0 | 0 | | Not estimable |
| Sahlen Helmer 2020 | 0 | 0 | 0 | 0 | | Not estimable |
| Sajaniemi 2001 | 0 | 0 | 0 | 0 | | Not estimable |
| Salo 2019 | 0 | 0 | 0 | 0 | | Not estimable |
| Santelices 2010 | 0 | 0 | 0 | 0 | | Not estimable |
| Schuler 2000 | 0 | 0 | 0 | 0 | | Not estimable |
| Shoghi 2018 | 0 | 0 | 0 | 0 | | Not estimable |
| Sleed 2013 | 0 | 0 | 0 | 0 | | Not estimable |
| Steele 2019 | 0 | 0 | 0 | 0 | | Not estimable |
| Stein 2006 | 0 | 0 | 0 | 0 | | Not estimable |
| Stein 2018 | 0 | 0 | 0 | 0 | | Not estimable |
| Taneja 2020 | 0 | 0 | 0 | 0 | | Not estimable |
| Tereno 2017 | 0 | 0 | 0 | 0 | | Not estimable |
| Teti 2009 | 0 | 0 | 0 | 0 | | Not estimable |
| Tryphonopoulos 2020 | 0 | 0 | 0 | 0 | | Not estimable |
| Tsivos 2015 | 0 | 0 | 0 | 0 | | Not estimable |
| Twohig 2021 | 0 | 0 | 0 | 0 | | Not estimable |
| Valades 2021 | 0 | 0 | 0 | 0 | | Not estimable |
| van Doesum 2008 | 0 | 0 | 0 | 0 | | Not estimable |
| Vargas-Porras 2021 | 0 | 0 | 0 | 0 | | Not estimable |
| Walter 2019 | 36 | 54 | 32 | 50 | 54.2% | 1.13 [0.50, 2.52] |
| White-Traut 2013 | 0 | 0 | 0 | 0 | | Not estimable |
| Williams 2020 | 0 | 0 | 0 | 0 | | Not estimable |
| Wulff 2021 | 0 | 0 | 0 | 0 | | Not estimable |
| Youn 2021 | 0 | 0 | 0 | 0 | | Not estimable |
| Yu 2022 | 0 | 0 | 0 | 0 | | Not estimable |
| **Total (95% CI)** | | **112** | | **102** | **100.0%** | **1.16 [0.64, 2.11]** |
| Total events | 60 | | 52 | | | |

Heterogeneity: Tau² = 0.00; Chi² = 0.03, df = 2 (P = 0.98); I² = 0%
Test for overall effect: Z = 0.49 (P = 0.62)

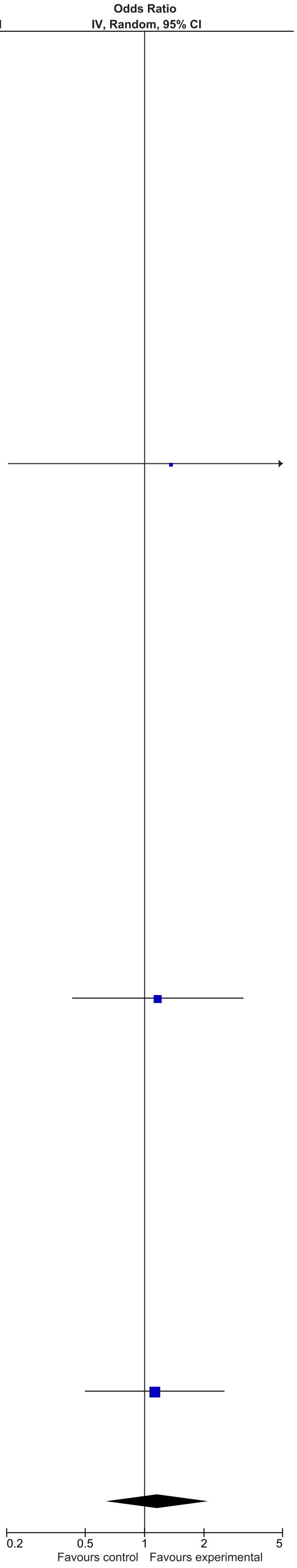

### eFigure 10. Secure Attachment 13-24-month Follow-Up Forrest Plot

| | Early Dyadic Intervention | | Standard Care | | | Odds Ratio |
|---|---|---|---|---|---|---|
| Study or Subgroup | Events | Total | Events | Total | Weight | IV, Random, 95% CI |
| Akai 2008 | 0 | 0 | 0 | 0 | | Not estimable |
| Alarcao 2021 | 0 | 0 | 0 | 0 | | Not estimable |
| Alvarenga 2019 | 0 | 0 | 0 | 0 | | Not estimable |
| Badr 2006 | 0 | 0 | 0 | 0 | | Not estimable |
| Baggett 2010 | 0 | 0 | 0 | 0 | | Not estimable |
| Baggett 2017 | 0 | 0 | 0 | 0 | | Not estimable |
| Barlow 2016 | 0 | 0 | 0 | 0 | | Not estimable |
| Barnes 2022 | 0 | 0 | 0 | 0 | | Not estimable |
| Berlin 2017 | 0 | 0 | 0 | 0 | | Not estimable |
| Berlin 2018 | 0 | 0 | 0 | 0 | | Not estimable |
| Bernard 2012 | 0 | 0 | 0 | 0 | | Not estimable |
| Betancourt 2020 | 0 | 0 | 0 | 0 | | Not estimable |
| Bick 2013 | 0 | 0 | 0 | 0 | | Not estimable |
| Borghini 2014 | 0 | 0 | 0 | 0 | | Not estimable |
| Brisch 2003 | 19 | 32 | 28 | 36 | 25.3% | 0.42 [0.15, 1.20] |
| Browne 2005 | 0 | 0 | 0 | 0 | | Not estimable |
| Cates 2018 | 0 | 0 | 0 | 0 | | Not estimable |
| Cevasco 2008 | 0 | 0 | 0 | 0 | | Not estimable |
| Chiu 2009 | 0 | 0 | 0 | 0 | | Not estimable |
| Constantino 2001 | 0 | 0 | 0 | 0 | | Not estimable |
| Cooper 2009 | 116 | 156 | 102 | 162 | 37.0% | 1.71 [1.06, 2.76] |
| Cooper 2015 | 34 | 52 | 39 | 59 | 30.8% | 0.97 [0.44, 2.12] |
| Corrigan 2021 | 0 | 0 | 0 | 0 | | Not estimable |
| Dozier 2009 | 0 | 0 | 0 | 0 | | Not estimable |
| Eriksen 2018 | 0 | 0 | 0 | 0 | | Not estimable |
| Feil 2020 | 0 | 0 | 0 | 0 | | Not estimable |
| Feldman 2014 | 0 | 0 | 0 | 0 | | Not estimable |
| Firk 2021 | 0 | 0 | 0 | 0 | | Not estimable |
| Fonagy 2016 | 0 | 0 | 0 | 0 | | Not estimable |
| Gaden 2022 | 0 | 0 | 0 | 0 | | Not estimable |
| Georg 2021 | 0 | 0 | 0 | 0 | | Not estimable |
| Glazebrook 2007 | 0 | 0 | 0 | 0 | | Not estimable |
| Hane 2015 | 0 | 0 | 0 | 0 | | Not estimable |
| Heo 2019 | 0 | 0 | 0 | 0 | | Not estimable |
| Herbers 2020 | 0 | 0 | 0 | 0 | | Not estimable |
| Hoffenkamp 2015 | 0 | 0 | 0 | 0 | | Not estimable |
| Hoivik 2015 | 0 | 0 | 0 | 0 | | Not estimable |
| Holditch-Davis 2014 | 0 | 0 | 0 | 0 | | Not estimable |
| Holt 2021 | 0 | 0 | 0 | 0 | | Not estimable |
| Horowitz 2001 | 0 | 0 | 0 | 0 | | Not estimable |
| Horowitz 2013 | 0 | 0 | 0 | 0 | | Not estimable |
| Juffer 2005 | 0 | 0 | 0 | 0 | | Not estimable |
| Klein Velderman 2006 | 0 | 0 | 0 | 0 | | Not estimable |
| Knoche 2012 | 0 | 0 | 0 | 0 | | Not estimable |
| Kristensen 2020 | 0 | 0 | 0 | 0 | | Not estimable |
| Landry 2006 | 0 | 0 | 0 | 0 | | Not estimable |
| Lavallee 2022 | 0 | 0 | 0 | 0 | | Not estimable |
| Magil-Evans 2007 | 0 | 0 | 0 | 0 | | Not estimable |
| Meijssen 2010 | 0 | 0 | 0 | 0 | | Not estimable |
| Melnyk 2006 | 0 | 0 | 0 | 0 | | Not estimable |
| Mendelsohn 2007 | 0 | 0 | 0 | 0 | | Not estimable |
| Milgrom 2013 | 0 | 0 | 0 | 0 | | Not estimable |
| Nelson 2001 | 0 | 0 | 0 | 0 | | Not estimable |
| Neu 2010 | 0 | 0 | 0 | 0 | | Not estimable |
| Newnham 2009 | 0 | 0 | 0 | 0 | | Not estimable |
| Nugent 2017 | 0 | 0 | 0 | 0 | | Not estimable |
| Onozawa 2001 | 0 | 0 | 0 | 0 | | Not estimable |
| Oxford 2021 | 0 | 0 | 0 | 0 | | Not estimable |
| Perrone 2021 | 0 | 0 | 0 | 0 | | Not estimable |
| Pontoppidan 2016 | 0 | 0 | 0 | 0 | | Not estimable |
| Porter 2015 | 0 | 0 | 0 | 0 | | Not estimable |
| Ramsauer 2019 | 0 | 0 | 0 | 0 | | Not estimable |
| Ravn 2011 | 0 | 0 | 0 | 0 | | Not estimable |
| Rheinheimer 2022 | 0 | 0 | 0 | 0 | | Not estimable |
| Robertson 2019 | 0 | 0 | 0 | 0 | | Not estimable |
| Roby 2021 | 0 | 0 | 0 | 0 | | Not estimable |
| Sadler 2013 | 0 | 0 | 0 | 0 | | Not estimable |
| Sahlen Helmer 2020 | 0 | 0 | 0 | 0 | | Not estimable |
| Sajaniemi 2001 | 6 | 23 | 0 | 25 | 6.9% | 18.94 [1.00, 358.35] |
| Salo 2019 | 0 | 0 | 0 | 0 | | Not estimable |
| Santelices 2010 | 0 | 0 | 0 | 0 | | Not estimable |
| Schuler 2000 | 0 | 0 | 0 | 0 | | Not estimable |
| Shoghi 2018 | 0 | 0 | 0 | 0 | | Not estimable |
| Sleed 2013 | 0 | 0 | 0 | 0 | | Not estimable |
| Steele 2019 | 0 | 0 | 0 | 0 | | Not estimable |
| Stein 2006 | 0 | 0 | 0 | 0 | | Not estimable |
| Stein 2018 | 0 | 0 | 0 | 0 | | Not estimable |
| Taneja 2020 | 0 | 0 | 0 | 0 | | Not estimable |
| Tereno 2017 | 0 | 0 | 0 | 0 | | Not estimable |
| Teti 2009 | 0 | 0 | 0 | 0 | | Not estimable |
| Tryphonopoulos 2020 | 0 | 0 | 0 | 0 | | Not estimable |
| Tsivos 2015 | 0 | 0 | 0 | 0 | | Not estimable |
| Twohig 2021 | 0 | 0 | 0 | 0 | | Not estimable |
| Valades 2021 | 0 | 0 | 0 | 0 | | Not estimable |
| van Doesum 2008 | 0 | 0 | 0 | 0 | | Not estimable |
| Vargas-Porras 2021 | 0 | 0 | 0 | 0 | | Not estimable |
| Walter 2019 | 0 | 0 | 0 | 0 | | Not estimable |
| White-Traut 2013 | 0 | 0 | 0 | 0 | | Not estimable |
| Williams 2020 | 0 | 0 | 0 | 0 | | Not estimable |
| Wulff 2021 | 0 | 0 | 0 | 0 | | Not estimable |
| Youn 2021 | 0 | 0 | 0 | 0 | | Not estimable |
| Yu 2022 | 0 | 0 | 0 | 0 | | Not estimable |
| Zelkowitz 2011 | 0 | 0 | 0 | 0 | | Not estimable |
| **Total (95% CI)** | | **263** | | **282** | **100.0%** | **1.18 [0.51, 2.75]** |
| Total events | 175 | | 169 | | | |

Heterogeneity: Tau² = 0.44; Chi² = 9.40, df = 3 (P = 0.02); I² = 68%
Test for overall effect: Z = 0.39 (P = 0.69)

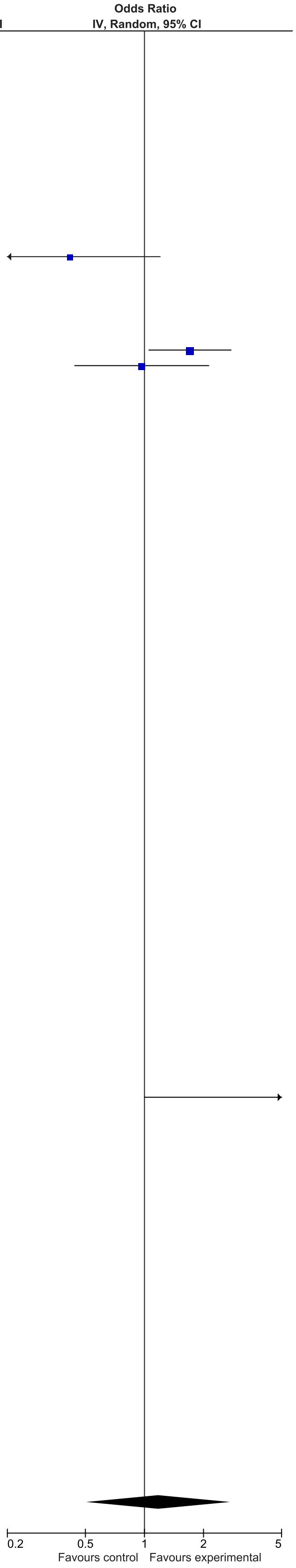

### eFigure 11. Organized Attachment 2-6-month Follow-Up Forrest Plot

| | Early Dyadic Intervention | | Standard Care | | | Odds Ratio |
|---|---|---|---|---|---|---|
| Study or Subgroup | Events | Total | Events | Total | Weight | IV, Random, 95% CI |
| Akai 2008 | 0 | 0 | 0 | 0 | | Not estimable |
| Alarcao 2021 | 0 | 0 | 0 | 0 | | Not estimable |
| Alvarenga 2019 | 0 | 0 | 0 | 0 | | Not estimable |
| Badr 2006 | 0 | 0 | 0 | 0 | | Not estimable |
| Baggett 2010 | 0 | 0 | 0 | 0 | | Not estimable |
| Baggett 2017 | 0 | 0 | 0 | 0 | | Not estimable |
| Barlow 2016 | 0 | 0 | 0 | 0 | | Not estimable |
| Barnes 2022 | 0 | 0 | 0 | 0 | | Not estimable |
| Berlin 2017 | 51 | 67 | 20 | 27 | 28.7% | 1.12 [0.40, 3.12] |
| Berlin 2018 | 0 | 0 | 0 | 0 | | Not estimable |
| Bernard 2012 | 41 | 60 | 26 | 60 | 46.7% | 2.82 [1.34, 5.95] |
| Betancourt 2020 | 0 | 0 | 0 | 0 | | Not estimable |
| Bick 2013 | 0 | 0 | 0 | 0 | | Not estimable |
| Borghini 2014 | 0 | 0 | 0 | 0 | | Not estimable |
| Brisch 2003 | 0 | 0 | 0 | 0 | | Not estimable |
| Browne 2005 | 0 | 0 | 0 | 0 | | Not estimable |
| Cates 2018 | 0 | 0 | 0 | 0 | | Not estimable |
| Cevasco 2008 | 0 | 0 | 0 | 0 | | Not estimable |
| Chiu 2009 | 0 | 0 | 0 | 0 | | Not estimable |
| Constantino 2001 | 0 | 0 | 0 | 0 | | Not estimable |
| Cooper 2009 | 0 | 0 | 0 | 0 | | Not estimable |
| Cooper 2015 | 0 | 0 | 0 | 0 | | Not estimable |
| Corrigan 2021 | 0 | 0 | 0 | 0 | | Not estimable |
| Dozier 2009 | 0 | 0 | 0 | 0 | | Not estimable |
| Eriksen 2018 | 0 | 0 | 0 | 0 | | Not estimable |
| Feil 2020 | 0 | 0 | 0 | 0 | | Not estimable |
| Feldman 2014 | 0 | 0 | 0 | 0 | | Not estimable |
| Firk 2021 | 0 | 0 | 0 | 0 | | Not estimable |
| Fonagy 2016 | 0 | 0 | 0 | 0 | | Not estimable |
| Gaden 2022 | 0 | 0 | 0 | 0 | | Not estimable |
| Georg 2021 | 0 | 0 | 0 | 0 | | Not estimable |
| Glazebrook 2007 | 0 | 0 | 0 | 0 | | Not estimable |
| Hane 2015 | 0 | 0 | 0 | 0 | | Not estimable |
| Heo 2019 | 0 | 0 | 0 | 0 | | Not estimable |
| Herbers 2020 | 0 | 0 | 0 | 0 | | Not estimable |
| Hoffenkamp 2015 | 0 | 0 | 0 | 0 | | Not estimable |
| Hoivik 2015 | 0 | 0 | 0 | 0 | | Not estimable |
| Holditch-Davis 2014 | 0 | 0 | 0 | 0 | | Not estimable |
| Holt 2021 | 0 | 0 | 0 | 0 | | Not estimable |
| Horowitz 2001 | 0 | 0 | 0 | 0 | | Not estimable |
| Horowitz 2013 | 0 | 0 | 0 | 0 | | Not estimable |
| Juffer 2005 | 0 | 0 | 0 | 0 | | Not estimable |
| Klein Velderman 2006 | 0 | 0 | 0 | 0 | | Not estimable |
| Knoche 2012 | 0 | 0 | 0 | 0 | | Not estimable |
| Kristensen 2020 | 0 | 0 | 0 | 0 | | Not estimable |
| Landry 2006 | 0 | 0 | 0 | 0 | | Not estimable |
| Lavallee 2022 | 0 | 0 | 0 | 0 | | Not estimable |
| Magil-Evans 2007 | 0 | 0 | 0 | 0 | | Not estimable |
| Meijssen 2010 | 0 | 0 | 0 | 0 | | Not estimable |
| Melnyk 2006 | 0 | 0 | 0 | 0 | | Not estimable |
| Mendelsohn 2007 | 0 | 0 | 0 | 0 | | Not estimable |
| Milgrom 2013 | 0 | 0 | 0 | 0 | | Not estimable |
| Nelson 2001 | 0 | 0 | 0 | 0 | | Not estimable |
| Neu 2010 | 0 | 0 | 0 | 0 | | Not estimable |
| Newnham 2009 | 0 | 0 | 0 | 0 | | Not estimable |
| Nugent 2017 | 0 | 0 | 0 | 0 | | Not estimable |
| Onozawa 2001 | 0 | 0 | 0 | 0 | | Not estimable |
| Oxford 2021 | 0 | 0 | 0 | 0 | | Not estimable |
| Perrone 2021 | 0 | 0 | 0 | 0 | | Not estimable |
| Pontoppidan 2016 | 0 | 0 | 0 | 0 | | Not estimable |
| Porter 2015 | 0 | 0 | 0 | 0 | | Not estimable |
| Ramsauer 2019 | 0 | 0 | 0 | 0 | | Not estimable |
| Ravn 2011 | 0 | 0 | 0 | 0 | | Not estimable |
| Rheinheimer 2022 | 0 | 0 | 0 | 0 | | Not estimable |
| Robertson 2019 | 0 | 0 | 0 | 0 | | Not estimable |
| Roby 2021 | 0 | 0 | 0 | 0 | | Not estimable |
| Sadler 2013 | 0 | 0 | 0 | 0 | | Not estimable |
| Sahlen Helmer 2020 | 0 | 0 | 0 | 0 | | Not estimable |
| Sajaniemi 2001 | 0 | 0 | 0 | 0 | | Not estimable |
| Salo 2019 | 0 | 0 | 0 | 0 | | Not estimable |
| Santelices 2010 | 0 | 0 | 0 | 0 | | Not estimable |
| Schuler 2000 | 0 | 0 | 0 | 0 | | Not estimable |
| Shoghi 2018 | 0 | 0 | 0 | 0 | | Not estimable |
| Sleed 2013 | 0 | 0 | 0 | 0 | | Not estimable |
| Steele 2019 | 0 | 0 | 0 | 0 | | Not estimable |
| Stein 2006 | 0 | 0 | 0 | 0 | | Not estimable |
| Stein 2018 | 0 | 0 | 0 | 0 | | Not estimable |
| Taneja 2020 | 0 | 0 | 0 | 0 | | Not estimable |
| Tereno 2017 | 60 | 65 | 41 | 52 | 24.6% | 3.22 [1.04, 9.96] |
| Teti 2009 | 0 | 0 | 0 | 0 | | Not estimable |
| Tryphonopoulos 2020 | 0 | 0 | 0 | 0 | | Not estimable |
| Tsivos 2015 | 0 | 0 | 0 | 0 | | Not estimable |
| Twohig 2021 | 0 | 0 | 0 | 0 | | Not estimable |
| Valades 2021 | 0 | 0 | 0 | 0 | | Not estimable |
| van Doesum 2008 | 0 | 0 | 0 | 0 | | Not estimable |
| Vargas-Porras 2021 | 0 | 0 | 0 | 0 | | Not estimable |
| Walter 2019 | 0 | 0 | 0 | 0 | | Not estimable |
| White-Traut 2013 | 0 | 0 | 0 | 0 | | Not estimable |
| Williams 2020 | 0 | 0 | 0 | 0 | | Not estimable |
| Wulff 2021 | 0 | 0 | 0 | 0 | | Not estimable |
| Youn 2021 | 0 | 0 | 0 | 0 | | Not estimable |
| Yu 2022 | 0 | 0 | 0 | 0 | | Not estimable |
| Zelkowitz 2011 | 0 | 0 | 0 | 0 | | Not estimable |
| **Total (95% CI)** | | **192** | | **139** | **100.0%** | **2.23 [1.21, 4.11]** |
| Total events | 152 | | 87 | | | |

Heterogeneity: Tau² = 0.06; Chi² = 2.53, df = 2 (P = 0.28); I² = 21%
Test for overall effect: Z = 2.58 (P = 0.010)

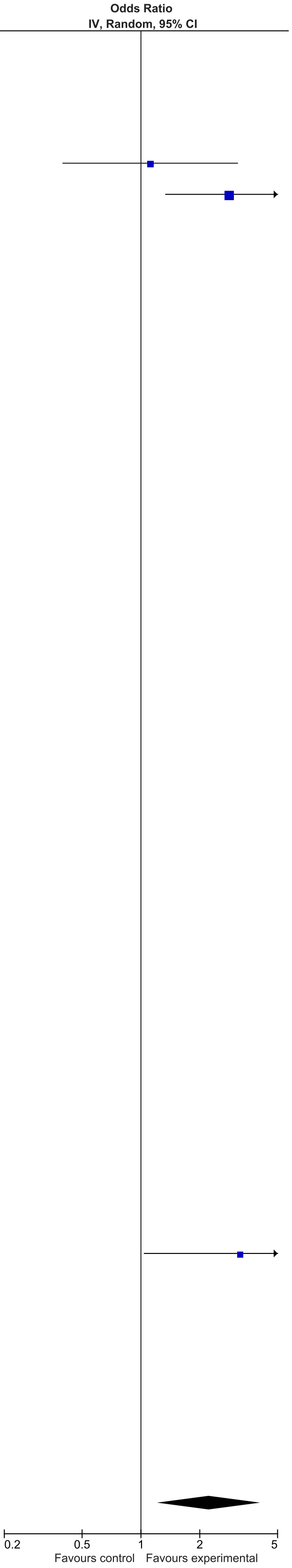

### eFigure 12. Insecure Attachment 0-1-month Follow-Up Forrest Plot

| | Early Dyadic Intervention | | Standard Care | | | Odds Ratio |
|---|---|---|---|---|---|---|
| Study or Subgroup | Events | Total | Events | Total | Weight | IV, Random, 95% CI |
| Akai 2008 | 0 | 0 | 0 | 0 | | Not estimable |
| Alarcao 2021 | 10 | 28 | 18 | 26 | 14.5% | 0.25 [0.08, 0.77] |
| Alvarenga 2019 | 0 | 0 | 0 | 0 | | Not estimable |
| Badr 2006 | 0 | 0 | 0 | 0 | | Not estimable |
| Baggett 2010 | 0 | 0 | 0 | 0 | | Not estimable |
| Baggett 2017 | 0 | 0 | 0 | 0 | | Not estimable |
| Barlow 2016 | 0 | 0 | 0 | 0 | | Not estimable |
| Barnes 2022 | 0 | 0 | 0 | 0 | | Not estimable |
| Berlin 2017 | 33 | 67 | 15 | 27 | 20.1% | 0.78 [0.32, 1.91] |
| Berlin 2018 | 0 | 0 | 0 | 0 | | Not estimable |
| Bernard 2012 | 29 | 60 | 40 | 60 | 25.4% | 0.47 [0.22, 0.98] |
| Betancourt 2020 | 0 | 0 | 0 | 0 | | Not estimable |
| Bick 2013 | 0 | 0 | 0 | 0 | | Not estimable |
| Borghini 2014 | 0 | 0 | 0 | 0 | | Not estimable |
| Brisch 2003 | 0 | 0 | 0 | 0 | | Not estimable |
| Browne 2005 | 0 | 0 | 0 | 0 | | Not estimable |
| Cates 2018 | 0 | 0 | 0 | 0 | | Not estimable |
| Cevasco 2008 | 0 | 0 | 0 | 0 | | Not estimable |
| Chiu 2009 | 0 | 0 | 0 | 0 | | Not estimable |
| Constantino 2001 | 0 | 0 | 0 | 0 | | Not estimable |
| Cooper 2009 | 0 | 0 | 0 | 0 | | Not estimable |
| Cooper 2015 | 0 | 0 | 0 | 0 | | Not estimable |
| Corrigan 2021 | 0 | 0 | 0 | 0 | | Not estimable |
| Dozier 2009 | 0 | 0 | 0 | 0 | | Not estimable |
| Eriksen 2018 | 0 | 0 | 0 | 0 | | Not estimable |
| Feil 2020 | 0 | 0 | 0 | 0 | | Not estimable |
| Feldman 2014 | 0 | 0 | 0 | 0 | | Not estimable |
| Firk 2021 | 8 | 25 | 3 | 19 | 9.4% | 2.51 [0.56, 11.16] |
| Fonagy 2016 | 7 | 28 | 7 | 25 | 13.0% | 0.86 [0.25, 2.91] |
| Gaden 2022 | 0 | 0 | 0 | 0 | | Not estimable |
| Georg 2021 | 0 | 0 | 0 | 0 | | Not estimable |
| Glazebrook 2007 | 0 | 0 | 0 | 0 | | Not estimable |
| Hane 2015 | 0 | 0 | 0 | 0 | | Not estimable |
| Heo 2019 | 0 | 0 | 0 | 0 | | Not estimable |
| Herbers 2020 | 0 | 0 | 0 | 0 | | Not estimable |
| Hoffenkamp 2015 | 0 | 0 | 0 | 0 | | Not estimable |
| Hoivik 2015 | 0 | 0 | 0 | 0 | | Not estimable |
| Holditch-Davis 2014 | 0 | 0 | 0 | 0 | | Not estimable |
| Holt 2021 | 0 | 0 | 0 | 0 | | Not estimable |
| Horowitz 2001 | 0 | 0 | 0 | 0 | | Not estimable |
| Horowitz 2013 | 0 | 0 | 0 | 0 | | Not estimable |
| Juffer 2005 | 0 | 0 | 0 | 0 | | Not estimable |
| Klein Velderman 2006 | 0 | 0 | 0 | 0 | | Not estimable |
| Knoche 2012 | 0 | 0 | 0 | 0 | | Not estimable |
| Kristensen 2020 | 0 | 0 | 0 | 0 | | Not estimable |
| Landry 2006 | 0 | 0 | 0 | 0 | | Not estimable |
| Lavallee 2022 | 0 | 0 | 0 | 0 | | Not estimable |
| Magil-Evans 2007 | 0 | 0 | 0 | 0 | | Not estimable |
| Meijssen 2010 | 0 | 0 | 0 | 0 | | Not estimable |
| Melnyk 2006 | 0 | 0 | 0 | 0 | | Not estimable |
| Mendelsohn 2007 | 0 | 0 | 0 | 0 | | Not estimable |
| Milgrom 2013 | 0 | 0 | 0 | 0 | | Not estimable |
| Nelson 2001 | 0 | 0 | 0 | 0 | | Not estimable |
| Neu 2010 | 0 | 0 | 0 | 0 | | Not estimable |
| Newnham 2009 | 0 | 0 | 0 | 0 | | Not estimable |
| Nugent 2017 | 0 | 0 | 0 | 0 | | Not estimable |
| Onozawa 2001 | 0 | 0 | 0 | 0 | | Not estimable |
| Oxford 2021 | 0 | 0 | 0 | 0 | | Not estimable |
| Perrone 2021 | 0 | 0 | 0 | 0 | | Not estimable |
| Pontoppidan 2016 | 0 | 0 | 0 | 0 | | Not estimable |
| Porter 2015 | 0 | 0 | 0 | 0 | | Not estimable |
| Ramsauer 2019 | 0 | 0 | 0 | 0 | | Not estimable |
| Ravn 2011 | 0 | 0 | 0 | 0 | | Not estimable |
| Rheinheimer 2022 | 0 | 0 | 0 | 0 | | Not estimable |
| Robertson 2019 | 0 | 0 | 0 | 0 | | Not estimable |
| Roby 2021 | 0 | 0 | 0 | 0 | | Not estimable |
| Sadler 2013 | 0 | 0 | 0 | 0 | | Not estimable |
| Sahlen Helmer 2020 | 0 | 0 | 0 | 0 | | Not estimable |
| Sajaniemi 2001 | 0 | 0 | 0 | 0 | | Not estimable |
| Salo 2019 | 0 | 0 | 0 | 0 | | Not estimable |
| Santelices 2010 | 12 | 43 | 13 | 29 | 17.6% | 0.48 [0.18, 1.28] |
| Schuler 2000 | 0 | 0 | 0 | 0 | | Not estimable |
| Shoghi 2018 | 0 | 0 | 0 | 0 | | Not estimable |
| Sleed 2013 | 0 | 0 | 0 | 0 | | Not estimable |
| Steele 2019 | 0 | 0 | 0 | 0 | | Not estimable |
| Stein 2006 | 0 | 0 | 0 | 0 | | Not estimable |
| Stein 2018 | 0 | 0 | 0 | 0 | | Not estimable |
| Taneja 2020 | 0 | 0 | 0 | 0 | | Not estimable |
| Tereno 2017 | 0 | 0 | 0 | 0 | | Not estimable |
| Teti 2009 | 0 | 0 | 0 | 0 | | Not estimable |
| Tryphonopoulos 2020 | 0 | 0 | 0 | 0 | | Not estimable |
| Tsivos 2015 | 0 | 0 | 0 | 0 | | Not estimable |
| Twohig 2021 | 0 | 0 | 0 | 0 | | Not estimable |
| Valades 2021 | 0 | 0 | 0 | 0 | | Not estimable |
| van Doesum 2008 | 0 | 0 | 0 | 0 | | Not estimable |
| Vargas-Porras 2021 | 0 | 0 | 0 | 0 | | Not estimable |
| Walter 2019 | 0 | 0 | 0 | 0 | | Not estimable |
| White-Traut 2013 | 0 | 0 | 0 | 0 | | Not estimable |
| Williams 2020 | 0 | 0 | 0 | 0 | | Not estimable |
| Wulff 2021 | 0 | 0 | 0 | 0 | | Not estimable |
| Youn 2021 | 0 | 0 | 0 | 0 | | Not estimable |
| Yu 2022 | 0 | 0 | 0 | 0 | | Not estimable |
| Zelkowitz 2011 | 0 | 0 | 0 | 0 | | Not estimable |
| **Total (95% CI)** | | **251** | | **186** | **100.0%** | **0.60 [0.36, 0.99]** |
| Total events | 99 | | 96 | | | |

Heterogeneity: Tau² = 0.12; Chi² = 7.15, df = 5 (P = 0.21); I² = 30%
Test for overall effect: Z = 1.99 (P = 0.05)

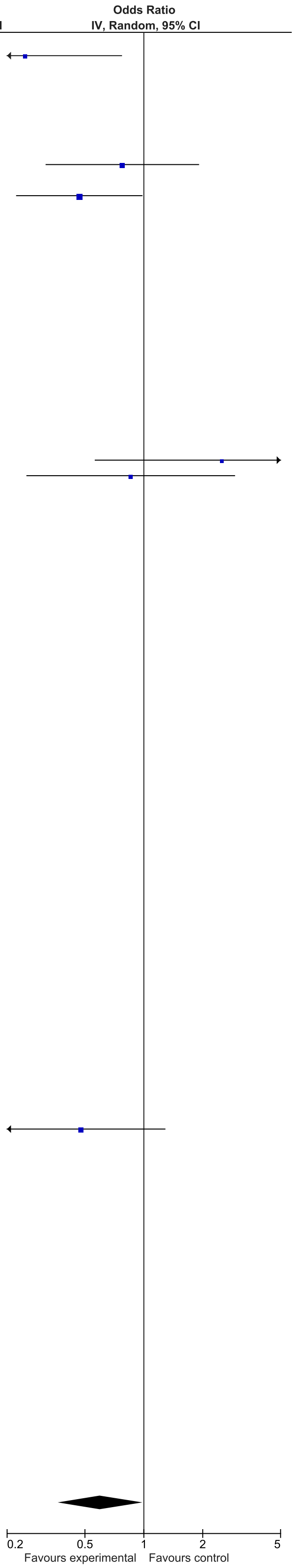

### eFigure 13. Insecure Attachment 2-6-month Follow-Up Forrest Plot

| Study or Subgroup | Early Dyadic Intervention Events | Early Dyadic Intervention Total | Standard Care Events | Standard Care Total | Weight | Odds Ratio IV, Random, 95% CI |
|---|---|---|---|---|---|---|
| Akai 2008 | 0 | 0 | 0 | 0 | | Not estimable |
| Alarcao 2021 | 0 | 0 | 0 | 0 | | Not estimable |
| Alvarenga 2019 | 0 | 0 | 0 | 0 | | Not estimable |
| Badr 2006 | 0 | 0 | 0 | 0 | | Not estimable |
| Baggett 2010 | 0 | 0 | 0 | 0 | | Not estimable |
| Baggett 2017 | 0 | 0 | 0 | 0 | | Not estimable |
| Barlow 2016 | 0 | 0 | 0 | 0 | | Not estimable |
| Barnes 2022 | 0 | 0 | 0 | 0 | | Not estimable |
| Berlin 2017 | 0 | 0 | 0 | 0 | | Not estimable |
| Berlin 2018 | 0 | 0 | 0 | 0 | | Not estimable |
| Bernard 2012 | 0 | 0 | 0 | 0 | | Not estimable |
| Betancourt 2020 | 0 | 0 | 0 | 0 | | Not estimable |
| Bick 2013 | 0 | 0 | 0 | 0 | | Not estimable |
| Borghini 2014 | 0 | 0 | 0 | 0 | | Not estimable |
| Brisch 2003 | 0 | 0 | 0 | 0 | | Not estimable |
| Browne 2005 | 0 | 0 | 0 | 0 | | Not estimable |
| Cates 2018 | 0 | 0 | 0 | 0 | | Not estimable |
| Cevasco 2008 | 0 | 0 | 0 | 0 | | Not estimable |
| Chiu 2009 | 0 | 0 | 0 | 0 | | Not estimable |
| Constantino 2001 | 0 | 0 | 0 | 0 | | Not estimable |
| Cooper 2009 | 0 | 0 | 0 | 0 | | Not estimable |
| Cooper 2015 | 0 | 0 | 0 | 0 | | Not estimable |
| Corrigan 2021 | 0 | 0 | 0 | 0 | | Not estimable |
| Dozier 2009 | 0 | 0 | 0 | 0 | | Not estimable |
| Eriksen 2018 | 0 | 0 | 0 | 0 | | Not estimable |
| Feil 2020 | 0 | 0 | 0 | 0 | | Not estimable |
| Feldman 2014 | 0 | 0 | 0 | 0 | | Not estimable |
| Firk 2021 | 13 | 24 | 13 | 21 | 24.7% | 0.73 [0.22, 2.39] |
| Fonagy 2016 | 0 | 0 | 0 | 0 | | Not estimable |
| Gaden 2022 | 0 | 0 | 0 | 0 | | Not estimable |
| Georg 2021 | 0 | 0 | 0 | 0 | | Not estimable |
| Glazebrook 2007 | 0 | 0 | 0 | 0 | | Not estimable |
| Hane 2015 | 0 | 0 | 0 | 0 | | Not estimable |
| Heo 2019 | 0 | 0 | 0 | 0 | | Not estimable |
| Herbers 2020 | 0 | 0 | 0 | 0 | | Not estimable |
| Hoffenkamp 2015 | 0 | 0 | 0 | 0 | | Not estimable |
| Hoivik 2015 | 0 | 0 | 0 | 0 | | Not estimable |
| Holditch-Davis 2014 | 0 | 0 | 0 | 0 | | Not estimable |
| Holt 2021 | 0 | 0 | 0 | 0 | | Not estimable |
| Horowitz 2001 | 0 | 0 | 0 | 0 | | Not estimable |
| Horowitz 2013 | 0 | 0 | 0 | 0 | | Not estimable |
| Juffer 2005 | 0 | 0 | 0 | 0 | | Not estimable |
| Klein Velderman 2006 | 0 | 0 | 0 | 0 | | Not estimable |
| Knoche 2012 | 0 | 0 | 0 | 0 | | Not estimable |
| Kristensen 2020 | 0 | 0 | 0 | 0 | | Not estimable |
| Landry 2006 | 0 | 0 | 0 | 0 | | Not estimable |
| Lavallee 2022 | 0 | 0 | 0 | 0 | | Not estimable |
| Magil-Evans 2007 | 0 | 0 | 0 | 0 | | Not estimable |
| Meijssen 2010 | 0 | 0 | 0 | 0 | | Not estimable |
| Melnyk 2006 | 0 | 0 | 0 | 0 | | Not estimable |
| Mendelsohn 2007 | 0 | 0 | 0 | 0 | | Not estimable |
| Milgrom 2013 | 0 | 0 | 0 | 0 | | Not estimable |
| Nelson 2001 | 0 | 0 | 0 | 0 | | Not estimable |
| Neu 2010 | 0 | 0 | 0 | 0 | | Not estimable |
| Newnham 2009 | 0 | 0 | 0 | 0 | | Not estimable |
| Nugent 2017 | 0 | 0 | 0 | 0 | | Not estimable |
| Onozawa 2001 | 0 | 0 | 0 | 0 | | Not estimable |
| Oxford 2021 | 0 | 0 | 0 | 0 | | Not estimable |
| Perrone 2021 | 0 | 0 | 0 | 0 | | Not estimable |
| Pontoppidan 2016 | 0 | 0 | 0 | 0 | | Not estimable |
| Porter 2015 | 0 | 0 | 0 | 0 | | Not estimable |
| Ramsauer 2019 | 5 | 34 | 7 | 31 | 21.8% | 0.59 [0.17, 2.10] |
| Ravn 2011 | 0 | 0 | 0 | 0 | | Not estimable |
| Rheinheimer 2022 | 0 | 0 | 0 | 0 | | Not estimable |
| Robertson 2019 | 0 | 0 | 0 | 0 | | Not estimable |
| Roby 2021 | 0 | 0 | 0 | 0 | | Not estimable |
| Sadler 2013 | 0 | 0 | 0 | 0 | | Not estimable |
| Sahlen Helmer 2020 | 0 | 0 | 0 | 0 | | Not estimable |
| Sajaniemi 2001 | 0 | 0 | 0 | 0 | | Not estimable |
| Salo 2019 | 0 | 0 | 0 | 0 | | Not estimable |
| Santelices 2010 | 0 | 0 | 0 | 0 | | Not estimable |
| Schuler 2000 | 0 | 0 | 0 | 0 | | Not estimable |
| Shoghi 2018 | 0 | 0 | 0 | 0 | | Not estimable |
| Sleed 2013 | 0 | 0 | 0 | 0 | | Not estimable |
| Steele 2019 | 0 | 0 | 0 | 0 | | Not estimable |
| Stein 2006 | 0 | 0 | 0 | 0 | | Not estimable |
| Stein 2018 | 0 | 0 | 0 | 0 | | Not estimable |
| Taneja 2020 | 0 | 0 | 0 | 0 | | Not estimable |
| Tereno 2017 | 0 | 0 | 0 | 0 | | Not estimable |
| Teti 2009 | 0 | 0 | 0 | 0 | | Not estimable |
| Tryphonopoulos 2020 | 0 | 0 | 0 | 0 | | Not estimable |
| Tsivos 2015 | 0 | 0 | 0 | 0 | | Not estimable |
| Twohig 2021 | 0 | 0 | 0 | 0 | | Not estimable |
| Valades 2021 | 0 | 0 | 0 | 0 | | Not estimable |
| van Doesum 2008 | 0 | 0 | 0 | 0 | | Not estimable |
| Vargas-Porras 2021 | 0 | 0 | 0 | 0 | | Not estimable |
| Walter 2019 | 18 | 54 | 18 | 50 | 53.6% | 0.89 [0.40, 2.00] |
| White-Traut 2013 | 0 | 0 | 0 | 0 | | Not estimable |
| Williams 2020 | 0 | 0 | 0 | 0 | | Not estimable |
| Wulff 2021 | 0 | 0 | 0 | 0 | | Not estimable |
| Youn 2021 | 0 | 0 | 0 | 0 | | Not estimable |
| Yu 2022 | 0 | 0 | 0 | 0 | | Not estimable |
| Zelkowitz 2011 | 0 | 0 | 0 | 0 | | Not estimable |
| **Total (95% CI)** | | **112** | | **102** | **100.0%** | **0.77 [0.43, 1.40]** |
| Total events | 36 | | 38 | | | |

Heterogeneity: Tau² = 0.00; Chi² = 0.30, df = 2 (P = 0.86); I² = 0%

Test for overall effect: Z = 0.85 (P = 0.40)

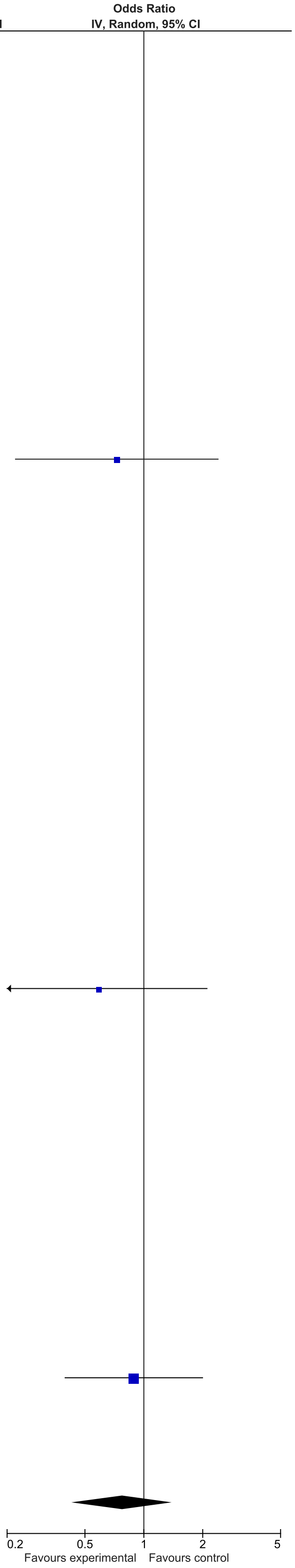

### eFigure 14. Bonding 13-24-month Follow-Up Forrest Plot

| | Early Dyadic Intervention | | Standard Care | | | Odds Ratio |
|---|---|---|---|---|---|---|
| **Study or Subgroup** | **Events** | **Total** | **Events** | **Total** | **Weight** | **IV, Random, 95% CI** |
| Akai 2008 | 0 | 0 | 0 | 0 | | Not estimable |
| Alarcao 2021 | 0 | 0 | 0 | 0 | | Not estimable |
| Alvarenga 2019 | 0 | 0 | 0 | 0 | | Not estimable |
| Badr 2006 | 0 | 0 | 0 | 0 | | Not estimable |
| Baggett 2010 | 0 | 0 | 0 | 0 | | Not estimable |
| Baggett 2017 | 0 | 0 | 0 | 0 | | Not estimable |
| Barlow 2016 | 0 | 0 | 0 | 0 | | Not estimable |
| Barnes 2022 | 0 | 0 | 0 | 0 | | Not estimable |
| Berlin 2017 | 0 | 0 | 0 | 0 | | Not estimable |
| Berlin 2018 | 0 | 0 | 0 | 0 | | Not estimable |
| Bernard 2012 | 0 | 0 | 0 | 0 | | Not estimable |
| Betancourt 2020 | 0 | 0 | 0 | 0 | | Not estimable |
| Bick 2013 | 0 | 0 | 0 | 0 | | Not estimable |
| Borghini 2014 | 0 | 0 | 0 | 0 | | Not estimable |
| Brisch 2003 | 13 | 32 | 8 | 36 | 43.8% | 2.39 [0.83, 6.88] |
| Browne 2005 | 0 | 0 | 0 | 0 | | Not estimable |
| Cates 2018 | 0 | 0 | 0 | 0 | | Not estimable |
| Cevasco 2008 | 0 | 0 | 0 | 0 | | Not estimable |
| Chiu 2009 | 0 | 0 | 0 | 0 | | Not estimable |
| Constantino 2001 | 0 | 0 | 0 | 0 | | Not estimable |
| Cooper 2009 | 30 | 156 | 44 | 162 | 56.2% | 0.64 [0.38, 1.08] |
| Cooper 2015 | 0 | 0 | 0 | 0 | | Not estimable |
| Corrigan 2021 | 0 | 0 | 0 | 0 | | Not estimable |
| Dozier 2009 | 0 | 0 | 0 | 0 | | Not estimable |
| Eriksen 2018 | 0 | 0 | 0 | 0 | | Not estimable |
| Feil 2020 | 0 | 0 | 0 | 0 | | Not estimable |
| Feldman 2014 | 0 | 0 | 0 | 0 | | Not estimable |
| Firk 2021 | 0 | 0 | 0 | 0 | | Not estimable |
| Fonagy 2016 | 0 | 0 | 0 | 0 | | Not estimable |
| Gaden 2022 | 0 | 0 | 0 | 0 | | Not estimable |
| Georg 2021 | 0 | 0 | 0 | 0 | | Not estimable |
| Glazebrook 2007 | 0 | 0 | 0 | 0 | | Not estimable |
| Hane 2015 | 0 | 0 | 0 | 0 | | Not estimable |
| Heo 2019 | 0 | 0 | 0 | 0 | | Not estimable |
| Herbers 2020 | 0 | 0 | 0 | 0 | | Not estimable |
| Hoffenkamp 2015 | 0 | 0 | 0 | 0 | | Not estimable |
| Hoivik 2015 | 0 | 0 | 0 | 0 | | Not estimable |
| Holditch-Davis 2014 | 0 | 0 | 0 | 0 | | Not estimable |
| Holt 2021 | 0 | 0 | 0 | 0 | | Not estimable |
| Horowitz 2001 | 0 | 0 | 0 | 0 | | Not estimable |
| Horowitz 2013 | 0 | 0 | 0 | 0 | | Not estimable |
| Juffer 2005 | 0 | 0 | 0 | 0 | | Not estimable |
| Klein Velderman 2006 | 0 | 0 | 0 | 0 | | Not estimable |
| Knoche 2012 | 0 | 0 | 0 | 0 | | Not estimable |
| Kristensen 2020 | 0 | 0 | 0 | 0 | | Not estimable |
| Landry 2006 | 0 | 0 | 0 | 0 | | Not estimable |
| Lavallee 2022 | 0 | 0 | 0 | 0 | | Not estimable |
| Magil-Evans 2007 | 0 | 0 | 0 | 0 | | Not estimable |
| Meijssen 2010 | 0 | 0 | 0 | 0 | | Not estimable |
| Melnyk 2006 | 0 | 0 | 0 | 0 | | Not estimable |
| Mendelsohn 2007 | 0 | 0 | 0 | 0 | | Not estimable |
| Milgrom 2013 | 0 | 0 | 0 | 0 | | Not estimable |
| Nelson 2001 | 0 | 0 | 0 | 0 | | Not estimable |
| Neu 2010 | 0 | 0 | 0 | 0 | | Not estimable |
| Newnham 2009 | 0 | 0 | 0 | 0 | | Not estimable |
| Nugent 2017 | 0 | 0 | 0 | 0 | | Not estimable |
| Onozawa 2001 | 0 | 0 | 0 | 0 | | Not estimable |
| Oxford 2021 | 0 | 0 | 0 | 0 | | Not estimable |
| Perrone 2021 | 0 | 0 | 0 | 0 | | Not estimable |
| Pontoppidan 2016 | 0 | 0 | 0 | 0 | | Not estimable |
| Porter 2015 | 0 | 0 | 0 | 0 | | Not estimable |
| Ramsauer 2019 | 0 | 0 | 0 | 0 | | Not estimable |
| Ravn 2011 | 0 | 0 | 0 | 0 | | Not estimable |
| Rheinheimer 2022 | 0 | 0 | 0 | 0 | | Not estimable |
| Robertson 2019 | 0 | 0 | 0 | 0 | | Not estimable |
| Roby 2021 | 0 | 0 | 0 | 0 | | Not estimable |
| Sadler 2013 | 0 | 0 | 0 | 0 | | Not estimable |
| Sahlen Helmer 2020 | 0 | 0 | 0 | 0 | | Not estimable |
| Sajaniemi 2001 | 0 | 0 | 0 | 0 | | Not estimable |
| Salo 2019 | 0 | 0 | 0 | 0 | | Not estimable |
| Santelices 2010 | 0 | 0 | 0 | 0 | | Not estimable |
| Schuler 2000 | 0 | 0 | 0 | 0 | | Not estimable |
| Shoghi 2018 | 0 | 0 | 0 | 0 | | Not estimable |
| Sleed 2013 | 0 | 0 | 0 | 0 | | Not estimable |
| Steele 2019 | 0 | 0 | 0 | 0 | | Not estimable |
| Stein 2006 | 0 | 0 | 0 | 0 | | Not estimable |
| Stein 2018 | 0 | 0 | 0 | 0 | | Not estimable |
| Taneja 2020 | 0 | 0 | 0 | 0 | | Not estimable |
| Tereno 2017 | 0 | 0 | 0 | 0 | | Not estimable |
| Teti 2009 | 0 | 0 | 0 | 0 | | Not estimable |
| Tryphonopoulos 2020 | 0 | 0 | 0 | 0 | | Not estimable |
| Tsivos 2015 | 0 | 0 | 0 | 0 | | Not estimable |
| Twohig 2021 | 0 | 0 | 0 | 0 | | Not estimable |
| Valades 2021 | 0 | 0 | 0 | 0 | | Not estimable |
| van Doesum 2008 | 0 | 0 | 0 | 0 | | Not estimable |
| Vargas-Porras 2021 | 0 | 0 | 0 | 0 | | Not estimable |
| Walter 2019 | 0 | 0 | 0 | 0 | | Not estimable |
| White-Traut 2013 | 0 | 0 | 0 | 0 | | Not estimable |
| Williams 2020 | 0 | 0 | 0 | 0 | | Not estimable |
| Wulff 2021 | 0 | 0 | 0 | 0 | | Not estimable |
| Youn 2021 | 0 | 0 | 0 | 0 | | Not estimable |
| Yu 2022 | 0 | 0 | 0 | 0 | | Not estimable |
| Zelkowitz 2011 | 0 | 0 | 0 | 0 | | Not estimable |
| **Total (95% CI)** | | **188** | | **198** | **100.0%** | **1.14 [0.31, 4.12]** |
| Total events | 43 | | 52 | | | |

Heterogeneity: Tau² = 0.69; Chi² = 4.82, df = 1 (P = 0.03); I² = 79%
Test for overall effect: Z = 0.20 (P = 0.84)

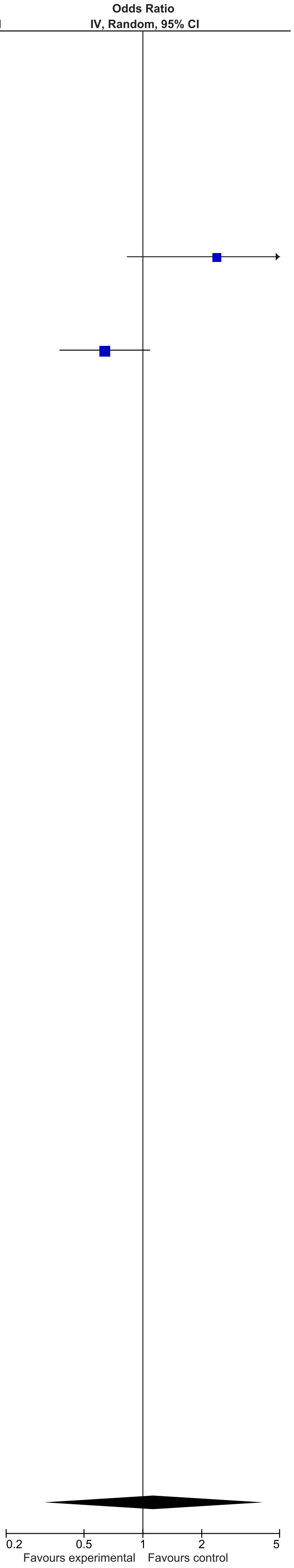

### eFigure 15. Disorganized Attachment 0-1-month Follow-Up Forrest Plot

| | Early Dyadic Intervention | | Standard Care | | | Odds Ratio |
|---|---|---|---|---|---|---|
| Study or Subgroup | Events | Total | Events | Total | Weight | IV, Random, 95% CI |
| Akai 2008 | 0 | 0 | 0 | 0 | | Not estimable |
| Alarcao 2021 | 0 | 0 | 0 | 0 | | Not estimable |
| Alvarenga 2019 | 0 | 0 | 0 | 0 | | Not estimable |
| Badr 2006 | 0 | 0 | 0 | 0 | | Not estimable |
| Baggett 2010 | 0 | 0 | 0 | 0 | | Not estimable |
| Baggett 2017 | 0 | 0 | 0 | 0 | | Not estimable |
| Barlow 2016 | 0 | 0 | 0 | 0 | | Not estimable |
| Barnes 2022 | 0 | 0 | 0 | 0 | | Not estimable |
| Berlin 2017 | 16 | 67 | 7 | 27 | 17.0% | 0.90 [0.32, 2.51] |
| Berlin 2018 | 0 | 0 | 0 | 0 | | Not estimable |
| Bernard 2012 | 19 | 60 | 34 | 60 | 32.3% | 0.35 [0.17, 0.75] |
| Betancourt 2020 | 0 | 0 | 0 | 0 | | Not estimable |
| Bick 2013 | 0 | 0 | 0 | 0 | | Not estimable |
| Borghini 2014 | 0 | 0 | 0 | 0 | | Not estimable |
| Brisch 2003 | 0 | 0 | 0 | 0 | | Not estimable |
| Browne 2005 | 0 | 0 | 0 | 0 | | Not estimable |
| Cates 2018 | 0 | 0 | 0 | 0 | | Not estimable |
| Cevasco 2008 | 0 | 0 | 0 | 0 | | Not estimable |
| Chiu 2009 | 0 | 0 | 0 | 0 | | Not estimable |
| Constantino 2001 | 0 | 0 | 0 | 0 | | Not estimable |
| Cooper 2009 | 0 | 0 | 0 | 0 | | Not estimable |
| Cooper 2015 | 0 | 0 | 0 | 0 | | Not estimable |
| Corrigan 2021 | 0 | 0 | 0 | 0 | | Not estimable |
| Dozier 2009 | 0 | 0 | 0 | 0 | | Not estimable |
| Eriksen 2018 | 0 | 0 | 0 | 0 | | Not estimable |
| Feil 2020 | 0 | 0 | 0 | 0 | | Not estimable |
| Feldman 2014 | 0 | 0 | 0 | 0 | | Not estimable |
| Firk 2021 | 7 | 25 | 6 | 19 | 10.6% | 0.84 [0.23, 3.10] |
| Fonagy 2016 | 4 | 28 | 4 | 25 | 7.9% | 0.88 [0.19, 3.94] |
| Gaden 2022 | 0 | 0 | 0 | 0 | | Not estimable |
| Georg 2021 | 0 | 0 | 0 | 0 | | Not estimable |
| Glazebrook 2007 | 0 | 0 | 0 | 0 | | Not estimable |
| Hane 2015 | 0 | 0 | 0 | 0 | | Not estimable |
| Heo 2019 | 0 | 0 | 0 | 0 | | Not estimable |
| Herbers 2020 | 0 | 0 | 0 | 0 | | Not estimable |
| Hoffenkamp 2015 | 0 | 0 | 0 | 0 | | Not estimable |
| Hoivik 2015 | 0 | 0 | 0 | 0 | | Not estimable |
| Holditch-Davis 2014 | 0 | 0 | 0 | 0 | | Not estimable |
| Holt 2021 | 0 | 0 | 0 | 0 | | Not estimable |
| Horowitz 2001 | 0 | 0 | 0 | 0 | | Not estimable |
| Horowitz 2013 | 0 | 0 | 0 | 0 | | Not estimable |
| Juffer 2005 | 0 | 0 | 0 | 0 | | Not estimable |
| Klein Velderman 2006 | 0 | 0 | 0 | 0 | | Not estimable |
| Knoche 2012 | 0 | 0 | 0 | 0 | | Not estimable |
| Kristensen 2020 | 0 | 0 | 0 | 0 | | Not estimable |
| Landry 2006 | 0 | 0 | 0 | 0 | | Not estimable |
| Lavallee 2022 | 0 | 0 | 0 | 0 | | Not estimable |
| Magil-Evans 2007 | 0 | 0 | 0 | 0 | | Not estimable |
| Meijssen 2010 | 0 | 0 | 0 | 0 | | Not estimable |
| Melnyk 2006 | 0 | 0 | 0 | 0 | | Not estimable |
| Mendelsohn 2007 | 0 | 0 | 0 | 0 | | Not estimable |
| Milgrom 2013 | 0 | 0 | 0 | 0 | | Not estimable |
| Nelson 2001 | 0 | 0 | 0 | 0 | | Not estimable |
| Neu 2010 | 0 | 0 | 0 | 0 | | Not estimable |
| Newnham 2009 | 0 | 0 | 0 | 0 | | Not estimable |
| Nugent 2017 | 0 | 0 | 0 | 0 | | Not estimable |
| Onozawa 2001 | 0 | 0 | 0 | 0 | | Not estimable |
| Oxford 2021 | 0 | 0 | 0 | 0 | | Not estimable |
| Perrone 2021 | 0 | 0 | 0 | 0 | | Not estimable |
| Pontoppidan 2016 | 0 | 0 | 0 | 0 | | Not estimable |
| Porter 2015 | 0 | 0 | 0 | 0 | | Not estimable |
| Ramsauer 2019 | 0 | 0 | 0 | 0 | | Not estimable |
| Ravn 2011 | 0 | 0 | 0 | 0 | | Not estimable |
| Rheinheimer 2022 | 0 | 0 | 0 | 0 | | Not estimable |
| Robertson 2019 | 0 | 0 | 0 | 0 | | Not estimable |
| Roby 2021 | 0 | 0 | 0 | 0 | | Not estimable |
| Sadler 2013 | 11 | 41 | 13 | 30 | 18.0% | 0.48 [0.18, 1.30] |
| Sahlen Helmer 2020 | 0 | 0 | 0 | 0 | | Not estimable |
| Sajaniemi 2001 | 0 | 0 | 0 | 0 | | Not estimable |
| Salo 2019 | 0 | 0 | 0 | 0 | | Not estimable |
| Santelices 2010 | 0 | 0 | 0 | 0 | | Not estimable |
| Schuler 2000 | 0 | 0 | 0 | 0 | | Not estimable |
| Shoghi 2018 | 0 | 0 | 0 | 0 | | Not estimable |
| Sleed 2013 | 0 | 0 | 0 | 0 | | Not estimable |
| Steele 2019 | 0 | 0 | 0 | 0 | | Not estimable |
| Stein 2006 | 0 | 0 | 0 | 0 | | Not estimable |
| Stein 2018 | 0 | 0 | 0 | 0 | | Not estimable |
| Taneja 2020 | 0 | 0 | 0 | 0 | | Not estimable |
| Tereno 2017 | 5 | 65 | 11 | 52 | 14.1% | 0.31 [0.10, 0.96] |
| Teti 2009 | 0 | 0 | 0 | 0 | | Not estimable |
| Tryphonopoulos 2020 | 0 | 0 | 0 | 0 | | Not estimable |
| Tsivos 2015 | 0 | 0 | 0 | 0 | | Not estimable |
| Twohig 2021 | 0 | 0 | 0 | 0 | | Not estimable |
| Valades 2021 | 0 | 0 | 0 | 0 | | Not estimable |
| van Doesum 2008 | 0 | 0 | 0 | 0 | | Not estimable |
| Vargas-Porras 2021 | 0 | 0 | 0 | 0 | | Not estimable |
| Walter 2019 | 0 | 0 | 0 | 0 | | Not estimable |
| White-Traut 2013 | 0 | 0 | 0 | 0 | | Not estimable |
| Williams 2020 | 0 | 0 | 0 | 0 | | Not estimable |
| Wulff 2021 | 0 | 0 | 0 | 0 | | Not estimable |
| Youn 2021 | 0 | 0 | 0 | 0 | | Not estimable |
| Yu 2022 | 0 | 0 | 0 | 0 | | Not estimable |
| Zelkowitz 2011 | 0 | 0 | 0 | 0 | | Not estimable |
| **Total (95% CI)** | | **286** | | **213** | **100.0%** | **0.51 [0.33, 0.77]** |
| Total events | 62 | | 75 | | | |

Heterogeneity: Tau² = 0.00; Chi² = 3.89, df = 5 (P = 0.57); I² = 0%

Test for overall effect: Z = 3.14 (P = 0.002)

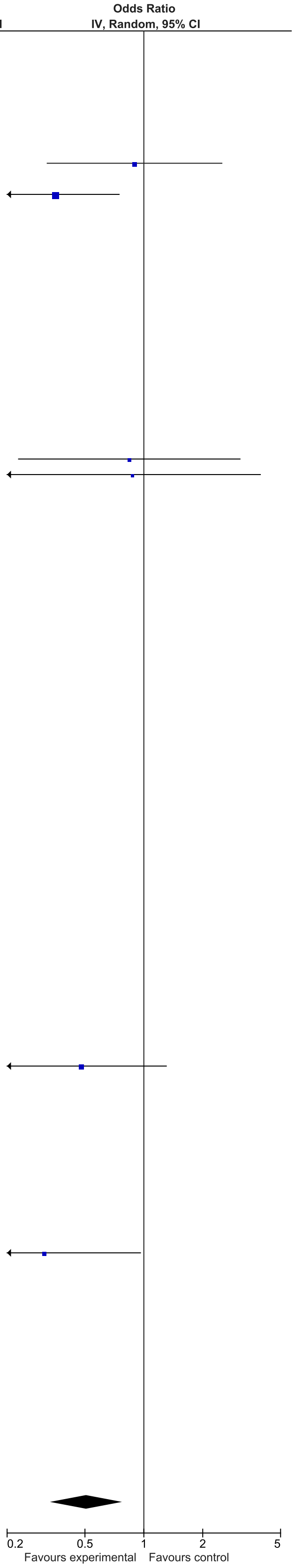

### eFigure 16. Disorganized Attachment 2-6-month Follow-Up Forrest Plot

| Study or Subgroup | Early Dyadic Intervention Events | Early Dyadic Intervention Total | Standard Care Events | Standard Care Total | Weight | Odds Ratio IV, Random, 95% CI |
|---|---|---|---|---|---|---|
| Akai 2008 | 0 | 0 | 0 | 0 | | Not estimable |
| Alarcao 2021 | 0 | 0 | 0 | 0 | | Not estimable |
| Alvarenga 2019 | 0 | 0 | 0 | 0 | | Not estimable |
| Badr 2006 | 0 | 0 | 0 | 0 | | Not estimable |
| Baggett 2010 | 0 | 0 | 0 | 0 | | Not estimable |
| Baggett 2017 | 0 | 0 | 0 | 0 | | Not estimable |
| Barlow 2016 | 0 | 0 | 0 | 0 | | Not estimable |
| Barnes 2022 | 0 | 0 | 0 | 0 | | Not estimable |
| Berlin 2017 | 0 | 0 | 0 | 0 | | Not estimable |
| Berlin 2018 | 0 | 0 | 0 | 0 | | Not estimable |
| Bernard 2012 | 0 | 0 | 0 | 0 | | Not estimable |
| Betancourt 2020 | 0 | 0 | 0 | 0 | | Not estimable |
| Bick 2013 | 0 | 0 | 0 | 0 | | Not estimable |
| Borghini 2014 | 0 | 0 | 0 | 0 | | Not estimable |
| Brisch 2003 | 0 | 0 | 0 | 0 | | Not estimable |
| Browne 2005 | 0 | 0 | 0 | 0 | | Not estimable |
| Cates 2018 | 0 | 0 | 0 | 0 | | Not estimable |
| Cevasco 2008 | 0 | 0 | 0 | 0 | | Not estimable |
| Chiu 2009 | 0 | 0 | 0 | 0 | | Not estimable |
| Constantino 2001 | 0 | 0 | 0 | 0 | | Not estimable |
| Cooper 2009 | 0 | 0 | 0 | 0 | | Not estimable |
| Cooper 2015 | 0 | 0 | 0 | 0 | | Not estimable |
| Corrigan 2021 | 0 | 0 | 0 | 0 | | Not estimable |
| Dozier 2009 | 0 | 0 | 0 | 0 | | Not estimable |
| Eriksen 2018 | 0 | 0 | 0 | 0 | | Not estimable |
| Feil 2020 | 0 | 0 | 0 | 0 | | Not estimable |
| Feldman 2014 | 0 | 0 | 0 | 0 | | Not estimable |
| Firk 2021 | 0 | 0 | 0 | 0 | | Not estimable |
| Fonagy 2016 | 0 | 0 | 0 | 0 | | Not estimable |
| Gaden 2022 | 0 | 0 | 0 | 0 | | Not estimable |
| Georg 2021 | 0 | 0 | 0 | 0 | | Not estimable |
| Glazebrook 2007 | 0 | 0 | 0 | 0 | | Not estimable |
| Hane 2015 | 0 | 0 | 0 | 0 | | Not estimable |
| Heo 2019 | 0 | 0 | 0 | 0 | | Not estimable |
| Herbers 2020 | 0 | 0 | 0 | 0 | | Not estimable |
| Hoffenkamp 2015 | 0 | 0 | 0 | 0 | | Not estimable |
| Hoivik 2015 | 0 | 0 | 0 | 0 | | Not estimable |
| Holditch-Davis 2014 | 0 | 0 | 0 | 0 | | Not estimable |
| Holt 2021 | 0 | 0 | 0 | 0 | | Not estimable |
| Horowitz 2001 | 0 | 0 | 0 | 0 | | Not estimable |
| Horowitz 2013 | 0 | 0 | 0 | 0 | | Not estimable |
| Juffer 2005 | 3 | 49 | 17 | 79 | 49.1% | 0.24 [0.07, 0.86] |
| Klein Velderman 2006 | 0 | 0 | 0 | 0 | | Not estimable |
| Knoche 2012 | 0 | 0 | 0 | 0 | | Not estimable |
| Kristensen 2020 | 0 | 0 | 0 | 0 | | Not estimable |
| Landry 2006 | 0 | 0 | 0 | 0 | | Not estimable |
| Lavallee 2022 | 0 | 0 | 0 | 0 | | Not estimable |
| Magil-Evans 2007 | 0 | 0 | 0 | 0 | | Not estimable |
| Meijssen 2010 | 0 | 0 | 0 | 0 | | Not estimable |
| Melnyk 2006 | 0 | 0 | 0 | 0 | | Not estimable |
| Mendelsohn 2007 | 0 | 0 | 0 | 0 | | Not estimable |
| Milgrom 2013 | 0 | 0 | 0 | 0 | | Not estimable |
| Nelson 2001 | 0 | 0 | 0 | 0 | | Not estimable |
| Neu 2010 | 0 | 0 | 0 | 0 | | Not estimable |
| Newnham 2009 | 0 | 0 | 0 | 0 | | Not estimable |
| Nugent 2017 | 0 | 0 | 0 | 0 | | Not estimable |
| Onozawa 2001 | 0 | 0 | 0 | 0 | | Not estimable |
| Oxford 2021 | 0 | 0 | 0 | 0 | | Not estimable |
| Perrone 2021 | 0 | 0 | 0 | 0 | | Not estimable |
| Pontoppidan 2016 | 0 | 0 | 0 | 0 | | Not estimable |
| Porter 2015 | 0 | 0 | 0 | 0 | | Not estimable |
| Ramsauer 2019 | 7 | 34 | 6 | 31 | 50.9% | 1.08 [0.32, 3.65] |
| Ravn 2011 | 0 | 0 | 0 | 0 | | Not estimable |
| Rheinheimer 2022 | 0 | 0 | 0 | 0 | | Not estimable |
| Robertson 2019 | 0 | 0 | 0 | 0 | | Not estimable |
| Roby 2021 | 0 | 0 | 0 | 0 | | Not estimable |
| Sadler 2013 | 0 | 0 | 0 | 0 | | Not estimable |
| Sahlen Helmer 2020 | 0 | 0 | 0 | 0 | | Not estimable |
| Sajaniemi 2001 | 0 | 0 | 0 | 0 | | Not estimable |
| Salo 2019 | 0 | 0 | 0 | 0 | | Not estimable |
| Santelices 2010 | 0 | 0 | 0 | 0 | | Not estimable |
| Schuler 2000 | 0 | 0 | 0 | 0 | | Not estimable |
| Shoghi 2018 | 0 | 0 | 0 | 0 | | Not estimable |
| Sleed 2013 | 0 | 0 | 0 | 0 | | Not estimable |
| Steele 2019 | 0 | 0 | 0 | 0 | | Not estimable |
| Stein 2006 | 0 | 0 | 0 | 0 | | Not estimable |
| Stein 2018 | 0 | 0 | 0 | 0 | | Not estimable |
| Taneja 2020 | 0 | 0 | 0 | 0 | | Not estimable |
| Tereno 2017 | 0 | 0 | 0 | 0 | | Not estimable |
| Teti 2009 | 0 | 0 | 0 | 0 | | Not estimable |
| Tryphonopoulos 2020 | 0 | 0 | 0 | 0 | | Not estimable |
| Tsivos 2015 | 0 | 0 | 0 | 0 | | Not estimable |
| Twohig 2021 | 0 | 0 | 0 | 0 | | Not estimable |
| Valades 2021 | 0 | 0 | 0 | 0 | | Not estimable |
| van Doesum 2008 | 0 | 0 | 0 | 0 | | Not estimable |
| Vargas-Porras 2021 | 0 | 0 | 0 | 0 | | Not estimable |
| Walter 2019 | 0 | 0 | 0 | 0 | | Not estimable |
| White-Traut 2013 | 0 | 0 | 0 | 0 | | Not estimable |
| Williams 2020 | 0 | 0 | 0 | 0 | | Not estimable |
| Wulff 2021 | 0 | 0 | 0 | 0 | | Not estimable |
| Youn 2021 | 0 | 0 | 0 | 0 | | Not estimable |
| Yu 2022 | 0 | 0 | 0 | 0 | | Not estimable |
| Zelkowitz 2011 | 0 | 0 | 0 | 0 | | Not estimable |
| **Total (95% CI)** | | **83** | | **110** | **100.0%** | **0.51 [0.12, 2.27]** |
| Total events | 10 | | 23 | | | |

Heterogeneity: Tau² = 0.74; Chi² = 2.80, df = 1 (P = 0.09); I² = 64%
Test for overall effect: Z = 0.88 (P = 0.38)

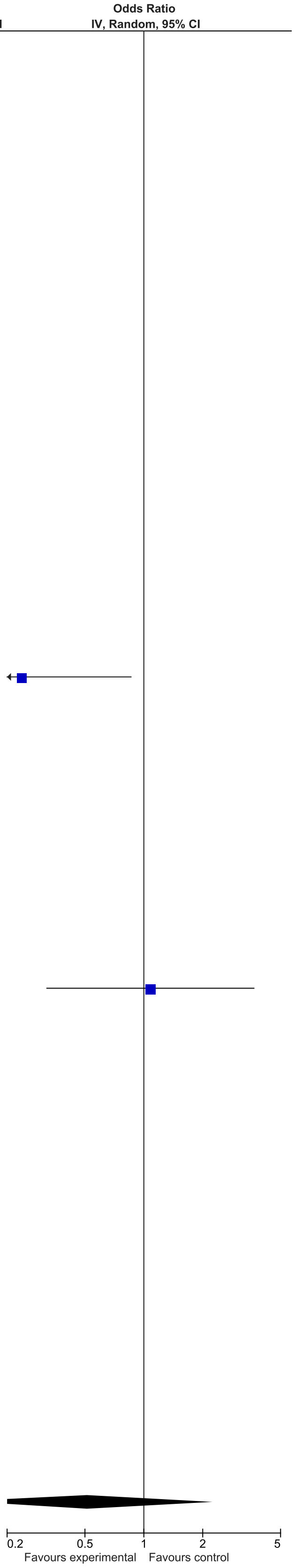

### eFigure 17. Dyadic Interactions 0-1-month Follow-Up Forrest Plot

| | Early Dyadic Intervention | | | Standard Care | | | | Std. Mean Difference |
|---|---|---|---|---|---|---|---|---|
| **Study or Subgroup** | **Mean** | **SD** | **Total** | **Mean** | **SD** | **Total** | **Weight** | **IV, Random, 95% CI** |
| Akai 2008 | 0 | 0 | 0 | 0 | 0 | 0 | | Not estimable |
| Alarcao 2021 | 4.9 | 3.65 | 28 | 4.58 | 3.43 | 26 | 3.3% | 0.09 [-0.45, 0.62] |
| Alvarenga 2019 | 0 | 0 | 0 | 0 | 0 | 0 | | Not estimable |
| Badr 2006 | 60.18 | 5.1 | 47 | 57.36 | 4.89 | 47 | 4.6% | 0.56 [0.15, 0.97] |
| Baggett 2010 | 0 | 0 | 0 | 0 | 0 | 0 | | Not estimable |
| Baggett 2017 | 0 | 0 | 0 | 0 | 0 | 0 | | Not estimable |
| Barlow 2016 | 0 | 0 | 0 | 0 | 0 | 0 | | Not estimable |
| Barnes 2022 | 0 | 0 | 0 | 0 | 0 | 0 | | Not estimable |
| Berlin 2017 | 0 | 0 | 0 | 0 | 0 | 0 | | Not estimable |
| Berlin 2018 | 0 | 0 | 0 | 0 | 0 | 0 | | Not estimable |
| Bernard 2012 | 0 | 0 | 0 | 0 | 0 | 0 | | Not estimable |
| Betancourt 2020 | 43.22 | 10.09 | 496 | 41.63 | 11.07 | 466 | 9.3% | 0.15 [0.02, 0.28] |
| Bick 2013 | 0 | 0 | 0 | 0 | 0 | 0 | | Not estimable |
| Borghini 2014 | 0 | 0 | 0 | 0 | 0 | 0 | | Not estimable |
| Brisch 2003 | 0 | 0 | 0 | 0 | 0 | 0 | | Not estimable |
| Browne 2005 | 46.65 | 6.2 | 28 | 48.08 | 7.22 | 56 | 4.1% | -0.21 [-0.66, 0.25] |
| Cates 2018 | 0 | 0 | 0 | 0 | 0 | 0 | | Not estimable |
| Cevasco 2008 | 0 | 0 | 0 | 0 | 0 | 0 | | Not estimable |
| Chiu 2009 | 0 | 0 | 0 | 0 | 0 | 0 | | Not estimable |
| Constantino 2001 | 0 | 0 | 0 | 0 | 0 | 0 | | Not estimable |
| Cooper 2009 | 0 | 0 | 0 | 0 | 0 | 0 | | Not estimable |
| Cooper 2015 | 2.965 | 1.01 | 79 | 3 | 1.05 | 77 | 6.0% | -0.03 [-0.35, 0.28] |
| Corrigan 2021 | 0 | 0 | 0 | 0 | 0 | 0 | | Not estimable |
| Dozier 2009 | 0 | 0 | 0 | 0 | 0 | 0 | | Not estimable |
| Eriksen 2018 | -6.21 | 3.72 | 7 | -6 | 1.58 | 5 | 1.0% | -0.06 [-1.21, 1.08] |
| Feil 2020 | 0 | 0 | 0 | 0 | 0 | 0 | | Not estimable |
| Feldman 2014 | 0 | 0 | 0 | 0 | 0 | 0 | | Not estimable |
| Firk 2021 | 4.06 | 2.23 | 27 | 4.15 | 2.96 | 20 | 3.0% | -0.03 [-0.61, 0.54] |
| Fonagy 2016 | 27.3 | 5.7 | 26 | 28.1 | 5.6 | 20 | 2.9% | -0.14 [-0.72, 0.44] |
| Gaden 2022 | 0 | 0 | 0 | 0 | 0 | 0 | | Not estimable |
| Georg 2021 | 18.62 | 3.76 | 81 | 18.11 | 3.95 | 73 | 5.9% | 0.13 [-0.18, 0.45] |
| Glazebrook 2007 | 0 | 0 | 0 | 0 | 0 | 0 | | Not estimable |
| Hane 2015 | 0 | 0 | 0 | 0 | 0 | 0 | | Not estimable |
| Heo 2019 | 0 | 0 | 0 | 0 | 0 | 0 | | Not estimable |
| Herbers 2020 | 3.07 | 0.98 | 20 | 2.46 | 0.92 | 18 | 2.5% | 0.63 [-0.03, 1.28] |
| Hoffenkamp 2015 | 0 | 0 | 0 | 0 | 0 | 0 | | Not estimable |
| Hoivik 2015 | 0 | 0 | 0 | 0 | 0 | 0 | | Not estimable |
| Holditch-Davis 2014 | 0 | 0 | 0 | 0 | 0 | 0 | | Not estimable |
| Holt 2021 | 0 | 0 | 0 | 0 | 0 | 0 | | Not estimable |
| Horowitz 2001 | 9.55 | 1.77 | 60 | 8.8 | 1.86 | 57 | 5.2% | 0.41 [0.04, 0.78] |
| Horowitz 2013 | 53.16 | 8.3 | 62 | 53.71 | 7 | 63 | 5.4% | -0.07 [-0.42, 0.28] |
| Juffer 2005 | 0 | 0 | 0 | 0 | 0 | 0 | | Not estimable |
| Klein Velderman 2006 | 0 | 0 | 0 | 0 | 0 | 0 | | Not estimable |
| Knoche 2012 | 0 | 0 | 0 | 0 | 0 | 0 | | Not estimable |
| Kristensen 2020 | 0 | 0 | 0 | 0 | 0 | 0 | | Not estimable |
| Landry 2006 | 0 | 0 | 0 | 0 | 0 | 0 | | Not estimable |
| Lavallee 2022 | 57.56 | 8.73 | 9 | 51.83 | 5.38 | 6 | 1.1% | 0.71 [-0.37, 1.78] |
| Magil-Evans 2007 | 0 | 0 | 0 | 0 | 0 | 0 | | Not estimable |
| Meijssen 2010 | 0 | 0 | 0 | 0 | 0 | 0 | | Not estimable |
| Melnyk 2006 | 0 | 0 | 0 | 0 | 0 | 0 | | Not estimable |
| Mendelsohn 2007 | 0 | 0 | 0 | 0 | 0 | 0 | | Not estimable |
| Milgrom 2013 | 2.67 | 1.42 | 54 | 2.45 | 1.49 | 50 | 4.9% | 0.15 [-0.24, 0.54] |
| Nelson 2001 | 0 | 0 | 0 | 0 | 0 | 0 | | Not estimable |
| Neu 2010 | 35.73 | 22.84 | 22 | 20.71 | 22.02 | 43 | 3.4% | 0.67 [0.14, 1.19] |
| Newnham 2009 | 0.29 | 0.09 | 32 | 0.23 | 0.11 | 31 | 3.6% | 0.59 [0.09, 1.10] |
| Nugent 2017 | 0 | 0 | 0 | 0 | 0 | 0 | | Not estimable |
| Onozawa 2001 | 3.2 | 1.13 | 10 | 2 | 0.97 | 12 | 1.4% | 1.10 [0.19, 2.02] |
| Oxford 2021 | 0 | 0 | 0 | 0 | 0 | 0 | | Not estimable |
| Perrone 2021 | 0 | 0 | 0 | 0 | 0 | 0 | | Not estimable |
| Pontoppidan 2016 | 4.35 | 0.38 | 64 | 4.19 | 0.47 | 33 | 4.4% | 0.38 [-0.04, 0.81] |
| Porter 2015 | 110.8 | 11.44 | 59 | 106.17 | 19.68 | 35 | 4.5% | 0.31 [-0.11, 0.73] |
| Ramsauer 2019 | 0 | 0 | 0 | 0 | 0 | 0 | | Not estimable |
| Ravn 2011 | 0 | 0 | 0 | 0 | 0 | 0 | | Not estimable |
| Rheinheimer 2022 | 0 | 0 | 0 | 0 | 0 | 0 | | Not estimable |
| Robertson 2019 | 0 | 0 | 0 | 0 | 0 | 0 | | Not estimable |
| Roby 2021 | 0 | 0 | 0 | 0 | 0 | 0 | | Not estimable |
| Sadler 2013 | 0 | 0 | 0 | 0 | 0 | 0 | | Not estimable |
| Sahlen Helmer 2020 | 0 | 0 | 0 | 0 | 0 | 0 | | Not estimable |
| Sajaniemi 2001 | 0 | 0 | 0 | 0 | 0 | 0 | | Not estimable |
| Salo 2019 | 0 | 0 | 0 | 0 | 0 | 0 | | Not estimable |
| Santelices 2010 | 0 | 0 | 0 | 0 | 0 | 0 | | Not estimable |
| Schuler 2000 | 0 | 0 | 0 | 0 | 0 | 0 | | Not estimable |
| Shoghi 2018 | 0 | 0 | 0 | 0 | 0 | 0 | | Not estimable |
| Sleed 2013 | 34.98 | 8.5 | 51 | 38.06 | 7.3 | 37 | 4.4% | -0.38 [-0.81, 0.05] |
| Steele 2019 | 3.15 | 0.85 | 43 | 2.57 | 0.75 | 35 | 4.0% | 0.71 [0.25, 1.17] |
| Stein 2006 | 0 | 0 | 0 | 0 | 0 | 0 | | Not estimable |
| Stein 2018 | 0 | 0 | 0 | 0 | 0 | 0 | | Not estimable |
| Taneja 2020 | 0 | 0 | 0 | 0 | 0 | 0 | | Not estimable |
| Tereno 2017 | 0 | 0 | 0 | 0 | 0 | 0 | | Not estimable |
| Teti 2009 | 0 | 0 | 0 | 0 | 0 | 0 | | Not estimable |
| Tryphonopoulos 2020 | 61 | 4.52 | 6 | 53.5 | 4.04 | 6 | 0.7% | 1.62 [0.24, 2.99] |
| Tsivos 2015 | 0 | 0 | 0 | 0 | 0 | 0 | | Not estimable |
| Twohig 2021 | 0 | 0 | 0 | 0 | 0 | 0 | | Not estimable |
| Valades 2021 | 0 | 0 | 0 | 0 | 0 | 0 | | Not estimable |
| van Doesum 2008 | 4.07 | 3.54 | 35 | 3.43 | 3.59 | 36 | 4.0% | 0.18 [-0.29, 0.64] |
| Vargas-Porras 2021 | 0 | 0 | 0 | 0 | 0 | 0 | | Not estimable |
| Walter 2019 | 0 | 0 | 0 | 0 | 0 | 0 | | Not estimable |
| White-Traut 2013 | 64.3 | 5.2 | 66 | 62.5 | 7 | 76 | 5.7% | 0.29 [-0.04, 0.62] |
| Williams 2020 | 0 | 0 | 0 | 0 | 0 | 0 | | Not estimable |
| Wulff 2021 | 0 | 0 | 0 | 0 | 0 | 0 | | Not estimable |
| Youn 2021 | 0 | 0 | 0 | 0 | 0 | 0 | | Not estimable |
| Yu 2022 | 0 | 0 | 0 | 0 | 0 | 0 | | Not estimable |
| Zelkowitz 2011 | 2.8 | 1 | 48 | 2.8 | 1 | 50 | 4.8% | 0.00 [-0.40, 0.40] |
| **Total (95% CI)** | | | **1460** | | | **1378** | **100.0%** | **0.22 [0.10, 0.34]** |

Heterogeneity: Tau² = 0.03; Chi² = 44.34, df = 24 (P = 0.007); I² = 46%
Test for overall effect: Z = 3.64 (P = 0.0003)

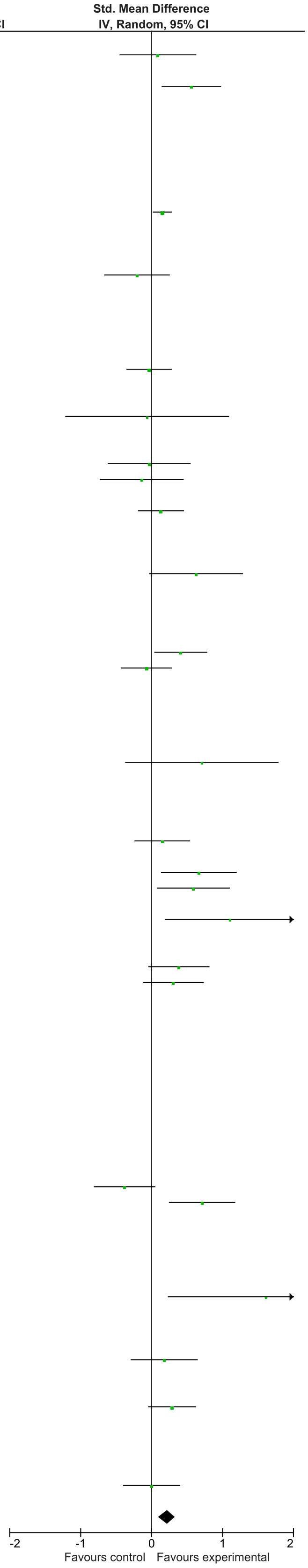

### eFigure 18. Dyadic Interactions 2-6-month Follow-Up Forrest Plot

| | Early Dyadic Intervention | | | Standard Care | | | | Std. Mean Difference |
|---|---|---|---|---|---|---|---|---|
| Study or Subgroup | Mean | SD | Total | Mean | SD | Total | Weight | IV, Random, 95% CI |
| Akai 2008 | 0 | 0 | 0 | 0 | 0 | 0 | | Not estimable |
| Alarcao 2021 | 0 | 0 | 0 | 0 | 0 | 0 | | Not estimable |
| Alvarenga 2019 | 0 | 0 | 0 | 0 | 0 | 0 | | Not estimable |
| Badr 2006 | 57.73 | 3.98 | 43 | 57.11 | 4.11 | 43 | 9.2% | 0.15 [-0.27, 0.58] |
| Baggett 2010 | 0 | 0 | 0 | 0 | 0 | 0 | | Not estimable |
| Baggett 2017 | 0 | 0 | 0 | 0 | 0 | 0 | | Not estimable |
| Barlow 2016 | 0 | 0 | 0 | 0 | 0 | 0 | | Not estimable |
| Barnes 2022 | 0 | 0 | 0 | 0 | 0 | 0 | | Not estimable |
| Berlin 2017 | 0 | 0 | 0 | 0 | 0 | 0 | | Not estimable |
| Berlin 2018 | 0 | 0 | 0 | 0 | 0 | 0 | | Not estimable |
| Bernard 2012 | 0 | 0 | 0 | 0 | 0 | 0 | | Not estimable |
| Betancourt 2020 | 0 | 0 | 0 | 0 | 0 | 0 | | Not estimable |
| Bick 2013 | 0 | 0 | 0 | 0 | 0 | 0 | | Not estimable |
| Borghini 2014 | 0 | 0 | 0 | 0 | 0 | 0 | | Not estimable |
| Brisch 2003 | 0 | 0 | 0 | 0 | 0 | 0 | | Not estimable |
| Browne 2005 | 0 | 0 | 0 | 0 | 0 | 0 | | Not estimable |
| Cates 2018 | 0 | 0 | 0 | 0 | 0 | 0 | | Not estimable |
| Cevasco 2008 | 0 | 0 | 0 | 0 | 0 | 0 | | Not estimable |
| Chiu 2009 | 61.4 | 6 | 40 | 60.9 | 6.9 | 29 | 8.6% | 0.08 [-0.40, 0.56] |
| Constantino 2001 | 0 | 0 | 0 | 0 | 0 | 0 | | Not estimable |
| Cooper 2009 | 0 | 0 | 0 | 0 | 0 | 0 | | Not estimable |
| Cooper 2015 | 0 | 0 | 0 | 0 | 0 | 0 | | Not estimable |
| Corrigan 2021 | 0 | 0 | 0 | 0 | 0 | 0 | | Not estimable |
| Dozier 2009 | 0 | 0 | 0 | 0 | 0 | 0 | | Not estimable |
| Eriksen 2018 | 0 | 0 | 0 | 0 | 0 | 0 | | Not estimable |
| Feil 2020 | 0 | 0 | 0 | 0 | 0 | 0 | | Not estimable |
| Feldman 2014 | 0 | 0 | 0 | 0 | 0 | 0 | | Not estimable |
| Firk 2021 | 4.2 | 1.03 | 24 | 4.24 | 1.23 | 21 | 7.6% | -0.03 [-0.62, 0.55] |
| Fonagy 2016 | 0 | 0 | 0 | 0 | 0 | 0 | | Not estimable |
| Gaden 2022 | 0 | 0 | 0 | 0 | 0 | 0 | | Not estimable |
| Georg 2021 | 0 | 0 | 0 | 0 | 0 | 0 | | Not estimable |
| Glazebrook 2007 | 0 | 0 | 0 | 0 | 0 | 0 | | Not estimable |
| Hane 2015 | 0.22 | 0.21 | 39 | 0.19 | 0.16 | 32 | 8.7% | 0.16 [-0.31, 0.63] |
| Heo 2019 | 0 | 0 | 0 | 0 | 0 | 0 | | Not estimable |
| Herbers 2020 | 0 | 0 | 0 | 0 | 0 | 0 | | Not estimable |
| Hoffenkamp 2015 | 0 | 0 | 0 | 0 | 0 | 0 | | Not estimable |
| Hoivik 2015 | 153.4 | 0.21 | 39 | 156.15 | 19.25 | 47 | 9.2% | -0.19 [-0.62, 0.23] |
| Holditch-Davis 2014 | 0 | 0 | 0 | 0 | 0 | 0 | | Not estimable |
| Holt 2021 | 0 | 0 | 0 | 0 | 0 | 0 | | Not estimable |
| Horowitz 2001 | 0 | 0 | 0 | 0 | 0 | 0 | | Not estimable |
| Horowitz 2013 | 0 | 0 | 0 | 0 | 0 | 0 | | Not estimable |
| Juffer 2005 | 0 | 0 | 0 | 0 | 0 | 0 | | Not estimable |
| Klein Velderman 2006 | 0 | 0 | 0 | 0 | 0 | 0 | | Not estimable |
| Knoche 2012 | 0 | 0 | 0 | 0 | 0 | 0 | | Not estimable |
| Kristensen 2020 | 0 | 0 | 0 | 0 | 0 | 0 | | Not estimable |
| Landry 2006 | 0 | 0 | 0 | 0 | 0 | 0 | | Not estimable |
| Lavallee 2022 | 0 | 0 | 0 | 0 | 0 | 0 | | Not estimable |
| Magil-Evans 2007 | 0 | 0 | 0 | 0 | 0 | 0 | | Not estimable |
| Meijssen 2010 | 0 | 0 | 0 | 0 | 0 | 0 | | Not estimable |
| Melnyk 2006 | 0 | 0 | 0 | 0 | 0 | 0 | | Not estimable |
| Mendelsohn 2007 | 0 | 0 | 0 | 0 | 0 | 0 | | Not estimable |
| Milgrom 2013 | 0 | 0 | 0 | 0 | 0 | 0 | | Not estimable |
| Nelson 2001 | 63.3 | 6.88 | 10 | 64.56 | 6.19 | 9 | 5.0% | -0.18 [-1.09, 0.72] |
| Neu 2010 | 0 | 0 | 0 | 0 | 0 | 0 | | Not estimable |
| Newnham 2009 | 0.45 | 0.08 | 32 | 0.24 | 0.13 | 31 | 7.4% | 1.93 [1.32, 2.53] |
| Nugent 2017 | 0 | 0 | 0 | 0 | 0 | 0 | | Not estimable |
| Onozawa 2001 | 0 | 0 | 0 | 0 | 0 | 0 | | Not estimable |
| Oxford 2021 | 5.43 | 1.95 | 127 | 5.31 | 2.57 | 125 | 10.8% | 0.05 [-0.19, 0.30] |
| Perrone 2021 | 0 | 0 | 0 | 0 | 0 | 0 | | Not estimable |
| Pontoppidan 2016 | 0 | 0 | 0 | 0 | 0 | 0 | | Not estimable |
| Porter 2015 | 119.41 | 11.5 | 51 | 112.74 | 9.95 | 35 | 9.0% | 0.61 [0.17, 1.05] |
| Ramsauer 2019 | 0 | 0 | 0 | 0 | 0 | 0 | | Not estimable |
| Ravn 2011 | 0 | 0 | 0 | 0 | 0 | 0 | | Not estimable |
| Rheinheimer 2022 | 0 | 0 | 0 | 0 | 0 | 0 | | Not estimable |
| Robertson 2019 | 5.196 | 5.19 | 21 | 3.782 | 6.49 | 24 | 7.5% | 0.23 [-0.35, 0.82] |
| Roby 2021 | 0 | 0 | 0 | 0 | 0 | 0 | | Not estimable |
| Sadler 2013 | 0 | 0 | 0 | 0 | 0 | 0 | | Not estimable |
| Sahlen Helmer 2020 | 0 | 0 | 0 | 0 | 0 | 0 | | Not estimable |
| Sajaniemi 2001 | 0 | 0 | 0 | 0 | 0 | 0 | | Not estimable |
| Salo 2019 | 4.33 | 2.33 | 24 | 4.13 | 2.68 | 21 | 7.6% | 0.08 [-0.51, 0.66] |
| Santelices 2010 | 0 | 0 | 0 | 0 | 0 | 0 | | Not estimable |
| Schuler 2000 | 0 | 0 | 0 | 0 | 0 | 0 | | Not estimable |
| Shoghi 2018 | 0 | 0 | 0 | 0 | 0 | 0 | | Not estimable |
| Sleed 2013 | 0 | 0 | 0 | 0 | 0 | 0 | | Not estimable |
| Steele 2019 | 0 | 0 | 0 | 0 | 0 | 0 | | Not estimable |
| Stein 2006 | 0 | 0 | 0 | 0 | 0 | 0 | | Not estimable |
| Stein 2018 | 0 | 0 | 0 | 0 | 0 | 0 | | Not estimable |
| Taneja 2020 | 0 | 0 | 0 | 0 | 0 | 0 | | Not estimable |
| Tereno 2017 | 0 | 0 | 0 | 0 | 0 | 0 | | Not estimable |
| Teti 2009 | 0 | 0 | 0 | 0 | 0 | 0 | | Not estimable |
| Tryphonopoulos 2020 | 0 | 0 | 0 | 0 | 0 | 0 | | Not estimable |
| Tsivos 2015 | 0 | 0 | 0 | 0 | 0 | 0 | | Not estimable |
| Twohig 2021 | 0 | 0 | 0 | 0 | 0 | 0 | | Not estimable |
| Valades 2021 | 0 | 0 | 0 | 0 | 0 | 0 | | Not estimable |
| van Doesum 2008 | 0 | 0 | 0 | 0 | 0 | 0 | | Not estimable |
| Vargas-Porras 2021 | 0 | 0 | 0 | 0 | 0 | 0 | | Not estimable |
| Walter 2019 | 0 | 0 | 0 | 0 | 0 | 0 | | Not estimable |
| White-Traut 2013 | 0 | 0 | 0 | 0 | 0 | 0 | | Not estimable |
| Williams 2020 | 0 | 0 | 0 | 0 | 0 | 0 | | Not estimable |
| Wulff 2021 | 0 | 0 | 0 | 0 | 0 | 0 | | Not estimable |
| Youn 2021 | 0 | 0 | 0 | 0 | 0 | 0 | | Not estimable |
| Yu 2022 | 0 | 0 | 0 | 0 | 0 | 0 | | Not estimable |
| Zelkowitz 2011 | 3.3 | 0.8 | 46 | 3.3 | 0.9 | 50 | 9.4% | 0.00 [-0.40, 0.40] |
| **Total (95% CI)** | | | **496** | | | **467** | **100.0%** | **0.23 [-0.03, 0.50]** |

Heterogeneity: Tau² = 0.15; Chi² = 42.01, df = 11 (P < 0.0001); I² = 74%

Test for overall effect: Z = 1.71 (P = 0.09)

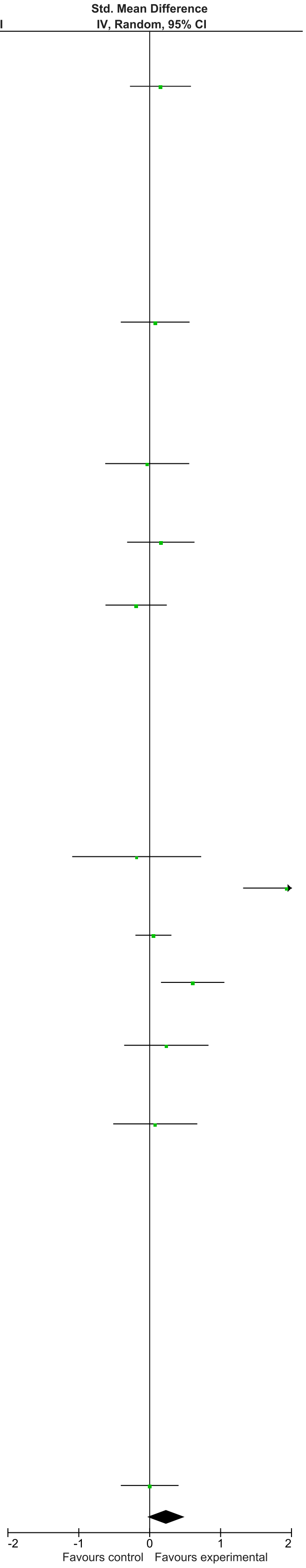

### eFigure 19. Dyadic Interactions 7-12-month Follow-Up Forrest Plot

| | Early Dyadic Intervention | | | Standard Care | | | | Std. Mean Difference |
|---|---|---|---|---|---|---|---|---|
| Study or Subgroup | Mean | SD | Total | Mean | SD | Total | Weight | IV, Random, 95% CI |
| Akai 2008 | 0 | 0 | 0 | 0 | 0 | 0 | | Not estimable |
| Alarcao 2021 | 0 | 0 | 0 | 0 | 0 | 0 | | Not estimable |
| Alvarenga 2019 | 0 | 0 | 0 | 0 | 0 | 0 | | Not estimable |
| Badr 2006 | 0 | 0 | 0 | 0 | 0 | 0 | | Not estimable |
| Baggett 2010 | 0 | 0 | 0 | 0 | 0 | 0 | | Not estimable |
| Baggett 2017 | 0 | 0 | 0 | 0 | 0 | 0 | | Not estimable |
| Barlow 2016 | 0 | 0 | 0 | 0 | 0 | 0 | | Not estimable |
| Barnes 2022 | 0 | 0 | 0 | 0 | 0 | 0 | | Not estimable |
| Berlin 2017 | 0 | 0 | 0 | 0 | 0 | 0 | | Not estimable |
| Berlin 2018 | 0 | 0 | 0 | 0 | 0 | 0 | | Not estimable |
| Bernard 2012 | 0 | 0 | 0 | 0 | 0 | 0 | | Not estimable |
| Betancourt 2020 | 0 | 0 | 0 | 0 | 0 | 0 | | Not estimable |
| Bick 2013 | 0 | 0 | 0 | 0 | 0 | 0 | | Not estimable |
| Borghini 2014 | 0 | 0 | 0 | 0 | 0 | 0 | | Not estimable |
| Brisch 2003 | 0 | 0 | 0 | 0 | 0 | 0 | | Not estimable |
| Browne 2005 | 0 | 0 | 0 | 0 | 0 | 0 | | Not estimable |
| Cates 2018 | 0 | 0 | 0 | 0 | 0 | 0 | | Not estimable |
| Cevasco 2008 | 0 | 0 | 0 | 0 | 0 | 0 | | Not estimable |
| Chiu 2009 | 56.5 | 5.1 | 40 | 55.2 | 5.7 | 30 | 13.1% | 0.24 [-0.24, 0.71] |
| Constantino 2001 | 0 | 0 | 0 | 0 | 0 | 0 | | Not estimable |
| Cooper 2009 | 0 | 0 | 0 | 0 | 0 | 0 | | Not estimable |
| Cooper 2015 | 0 | 0 | 0 | 0 | 0 | 0 | | Not estimable |
| Corrigan 2021 | 0 | 0 | 0 | 0 | 0 | 0 | | Not estimable |
| Dozier 2009 | 0 | 0 | 0 | 0 | 0 | 0 | | Not estimable |
| Eriksen 2018 | 0 | 0 | 0 | 0 | 0 | 0 | | Not estimable |
| Feil 2020 | 0 | 0 | 0 | 0 | 0 | 0 | | Not estimable |
| Feldman 2014 | 0 | 0 | 0 | 0 | 0 | 0 | | Not estimable |
| Firk 2021 | 0 | 0 | 0 | 0 | 0 | 0 | | Not estimable |
| Fonagy 2016 | 0 | 0 | 0 | 0 | 0 | 0 | | Not estimable |
| Gaden 2022 | 0 | 0 | 0 | 0 | 0 | 0 | | Not estimable |
| Georg 2021 | 0 | 0 | 0 | 0 | 0 | 0 | | Not estimable |
| Glazebrook 2007 | 0 | 0 | 0 | 0 | 0 | 0 | | Not estimable |
| Hane 2015 | 0 | 0 | 0 | 0 | 0 | 0 | | Not estimable |
| Heo 2019 | 0 | 0 | 0 | 0 | 0 | 0 | | Not estimable |
| Herbers 2020 | 0 | 0 | 0 | 0 | 0 | 0 | | Not estimable |
| Hoffenkamp 2015 | 0 | 0 | 0 | 0 | 0 | 0 | | Not estimable |
| Hoivik 2015 | 0 | 0 | 0 | 0 | 0 | 0 | | Not estimable |
| Holditch-Davis 2014 | 0 | 0 | 0 | 0 | 0 | 0 | | Not estimable |
| Holt 2021 | 0 | 0 | 0 | 0 | 0 | 0 | | Not estimable |
| Horowitz 2001 | 0 | 0 | 0 | 0 | 0 | 0 | | Not estimable |
| Horowitz 2013 | 0 | 0 | 0 | 0 | 0 | 0 | | Not estimable |
| Juffer 2005 | 0 | 0 | 0 | 0 | 0 | 0 | | Not estimable |
| Klein Velderman 2006 | 0 | 0 | 0 | 0 | 0 | 0 | | Not estimable |
| Knoche 2012 | 0 | 0 | 0 | 0 | 0 | 0 | | Not estimable |
| Kristensen 2020 | 0 | 0 | 0 | 0 | 0 | 0 | | Not estimable |
| Landry 2006 | 0 | 0 | 0 | 0 | 0 | 0 | | Not estimable |
| Lavallee 2022 | 0 | 0 | 0 | 0 | 0 | 0 | | Not estimable |
| Magil-Evans 2007 | 0 | 0 | 0 | 0 | 0 | 0 | | Not estimable |
| Meijssen 2010 | 0 | 0 | 0 | 0 | 0 | 0 | | Not estimable |
| Melnyk 2006 | 0 | 0 | 0 | 0 | 0 | 0 | | Not estimable |
| Mendelsohn 2007 | 0 | 0 | 0 | 0 | 0 | 0 | | Not estimable |
| Milgrom 2013 | 0 | 0 | 0 | 0 | 0 | 0 | | Not estimable |
| Nelson 2001 | 0 | 0 | 0 | 0 | 0 | 0 | | Not estimable |
| Neu 2010 | 0 | 0 | 0 | 0 | 0 | 0 | | Not estimable |
| Newnham 2009 | 0 | 0 | 0 | 0 | 0 | 0 | | Not estimable |
| Nugent 2017 | 0 | 0 | 0 | 0 | 0 | 0 | | Not estimable |
| Onozawa 2001 | 0 | 0 | 0 | 0 | 0 | 0 | | Not estimable |
| Oxford 2021 | 6.45 | 2.12 | 127 | 5.98 | 2.35 | 125 | 48.2% | 0.21 [-0.04, 0.46] |
| Perrone 2021 | 0 | 0 | 0 | 0 | 0 | 0 | | Not estimable |
| Pontoppidan 2016 | 0 | 0 | 0 | 0 | 0 | 0 | | Not estimable |
| Porter 2015 | 0 | 0 | 0 | 0 | 0 | 0 | | Not estimable |
| Ramsauer 2019 | 0 | 0 | 0 | 0 | 0 | 0 | | Not estimable |
| Ravn 2011 | 0 | 0 | 0 | 0 | 0 | 0 | | Not estimable |
| Rheinheimer 2022 | 0 | 0 | 0 | 0 | 0 | 0 | | Not estimable |
| Robertson 2019 | 0 | 0 | 0 | 0 | 0 | 0 | | Not estimable |
| Roby 2021 | 0 | 0 | 0 | 0 | 0 | 0 | | Not estimable |
| Sadler 2013 | 0 | 0 | 0 | 0 | 0 | 0 | | Not estimable |
| Sahlen Helmer 2020 | 0 | 0 | 0 | 0 | 0 | 0 | | Not estimable |
| Sajaniemi 2001 | 0 | 0 | 0 | 0 | 0 | 0 | | Not estimable |
| Salo 2019 | 0 | 0 | 0 | 0 | 0 | 0 | | Not estimable |
| Santelices 2010 | 0 | 0 | 0 | 0 | 0 | 0 | | Not estimable |
| Schuler 2000 | 2.95 | 0.5 | 67 | 2.9 | 0.5 | 64 | 25.2% | 0.10 [-0.24, 0.44] |
| Shoghi 2018 | 0 | 0 | 0 | 0 | 0 | 0 | | Not estimable |
| Sleed 2013 | 0 | 0 | 0 | 0 | 0 | 0 | | Not estimable |
| Steele 2019 | 0 | 0 | 0 | 0 | 0 | 0 | | Not estimable |
| Stein 2006 | 0 | 0 | 0 | 0 | 0 | 0 | | Not estimable |
| Stein 2018 | 0 | 0 | 0 | 0 | 0 | 0 | | Not estimable |
| Taneja 2020 | 0 | 0 | 0 | 0 | 0 | 0 | | Not estimable |
| Tereno 2017 | 0 | 0 | 0 | 0 | 0 | 0 | | Not estimable |
| Teti 2009 | 0 | 0 | 0 | 0 | 0 | 0 | | Not estimable |
| Tryphonopoulos 2020 | 0 | 0 | 0 | 0 | 0 | 0 | | Not estimable |
| Tsivos 2015 | 0 | 0 | 0 | 0 | 0 | 0 | | Not estimable |
| Twohig 2021 | 0 | 0 | 0 | 0 | 0 | 0 | | Not estimable |
| Valades 2021 | 0 | 0 | 0 | 0 | 0 | 0 | | Not estimable |
| van Doesum 2008 | 4.13 | 2.74 | 35 | 3.42 | 2.88 | 36 | 13.5% | 0.25 [-0.22, 0.72] |
| Vargas-Porras 2021 | 0 | 0 | 0 | 0 | 0 | 0 | | Not estimable |
| Walter 2019 | 0 | 0 | 0 | 0 | 0 | 0 | | Not estimable |
| White-Traut 2013 | 0 | 0 | 0 | 0 | 0 | 0 | | Not estimable |
| Williams 2020 | 0 | 0 | 0 | 0 | 0 | 0 | | Not estimable |
| Wulff 2021 | 0 | 0 | 0 | 0 | 0 | 0 | | Not estimable |
| Youn 2021 | 0 | 0 | 0 | 0 | 0 | 0 | | Not estimable |
| Yu 2022 | 0 | 0 | 0 | 0 | 0 | 0 | | Not estimable |
| Zelkowitz 2011 | 0 | 0 | 0 | 0 | 0 | 0 | | Not estimable |
| **Total (95% CI)** | | | **269** | | | **255** | **100.0%** | **0.19 [0.02, 0.36]** |

Heterogeneity: Tau² = 0.00; Chi² = 0.40, df = 3 (P = 0.94); I² = 0%
Test for overall effect: Z = 2.18 (P = 0.03)

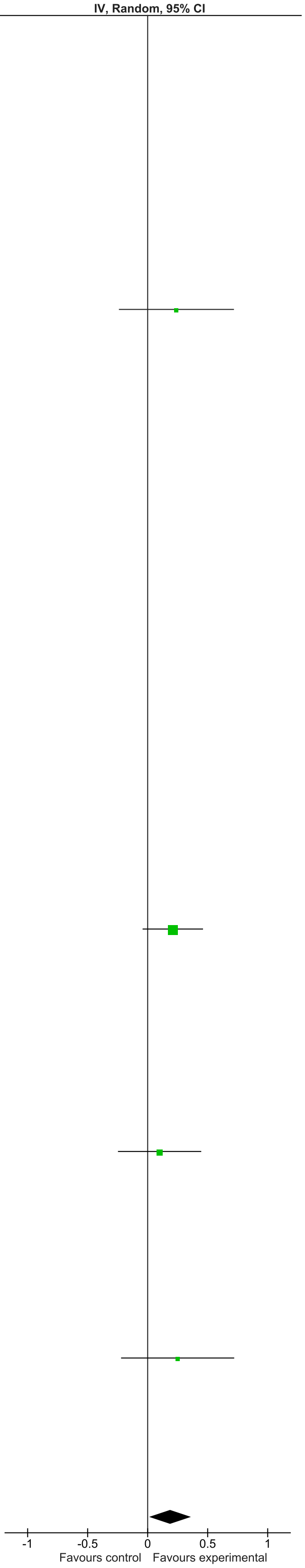

### eFigure 20. Cognitive Development 0-1-month Follow-Up Forrest Plot

| | Early Dyadic Intervention | | | Standard Care | | | | Std. Mean Difference |
|---|---|---|---|---|---|---|---|---|
| Study or Subgroup | Mean | SD | Total | Mean | SD | Total | Weight | IV, Random, 95% CI |
| Akai 2008 | 0 | 0 | 0 | 0 | 0 | 0 | | Not estimable |
| Alarcao 2021 | 0 | 0 | 0 | 0 | 0 | 0 | | Not estimable |
| Alvarenga 2019 | 0 | 0 | 0 | 0 | 0 | 0 | | Not estimable |
| Badr 2006 | 87.37 | 12.45 | 44 | 92.28 | 14.96 | 44 | 47.2% | -0.35 [-0.77, 0.07] |
| Baggett 2010 | 0 | 0 | 0 | 0 | 0 | 0 | | Not estimable |
| Baggett 2017 | 0 | 0 | 0 | 0 | 0 | 0 | | Not estimable |
| Barlow 2016 | 0 | 0 | 0 | 0 | 0 | 0 | | Not estimable |
| Barnes 2022 | 0 | 0 | 0 | 0 | 0 | 0 | | Not estimable |
| Berlin 2017 | 0 | 0 | 0 | 0 | 0 | 0 | | Not estimable |
| Berlin 2018 | 0 | 0 | 0 | 0 | 0 | 0 | | Not estimable |
| Bernard 2012 | 0 | 0 | 0 | 0 | 0 | 0 | | Not estimable |
| Betancourt 2020 | 0 | 0 | 0 | 0 | 0 | 0 | | Not estimable |
| Bick 2013 | 0 | 0 | 0 | 0 | 0 | 0 | | Not estimable |
| Borghini 2014 | 0 | 0 | 0 | 0 | 0 | 0 | | Not estimable |
| Brisch 2003 | 0 | 0 | 0 | 0 | 0 | 0 | | Not estimable |
| Browne 2005 | 0 | 0 | 0 | 0 | 0 | 0 | | Not estimable |
| Cates 2018 | 0 | 0 | 0 | 0 | 0 | 0 | | Not estimable |
| Cevasco 2008 | 0 | 0 | 0 | 0 | 0 | 0 | | Not estimable |
| Chiu 2009 | 0 | 0 | 0 | 0 | 0 | 0 | | Not estimable |
| Constantino 2001 | 0 | 0 | 0 | 0 | 0 | 0 | | Not estimable |
| Cooper 2009 | 0 | 0 | 0 | 0 | 0 | 0 | | Not estimable |
| Cooper 2015 | 0 | 0 | 0 | 0 | 0 | 0 | | Not estimable |
| Corrigan 2021 | 0 | 0 | 0 | 0 | 0 | 0 | | Not estimable |
| Dozier 2009 | 0 | 0 | 0 | 0 | 0 | 0 | | Not estimable |
| Eriksen 2018 | 0 | 0 | 0 | 0 | 0 | 0 | | Not estimable |
| Feil 2020 | 0 | 0 | 0 | 0 | 0 | 0 | | Not estimable |
| Feldman 2014 | 0 | 0 | 0 | 0 | 0 | 0 | | Not estimable |
| Firk 2021 | 9.67 | 1.7 | 27 | 10.15 | 1.4 | 20 | 24.8% | -0.30 [-0.88, 0.28] |
| Fonagy 2016 | 104.6 | 12.2 | 28 | 110.8 | 15 | 25 | 28.0% | -0.45 [-1.00, 0.10] |
| Gaden 2022 | 0 | 0 | 0 | 0 | 0 | 0 | | Not estimable |
| Georg 2021 | 0 | 0 | 0 | 0 | 0 | 0 | | Not estimable |
| Glazebrook 2007 | 0 | 0 | 0 | 0 | 0 | 0 | | Not estimable |
| Hane 2015 | 0 | 0 | 0 | 0 | 0 | 0 | | Not estimable |
| Heo 2019 | 0 | 0 | 0 | 0 | 0 | 0 | | Not estimable |
| Herbers 2020 | 0 | 0 | 0 | 0 | 0 | 0 | | Not estimable |
| Hoffenkamp 2015 | 0 | 0 | 0 | 0 | 0 | 0 | | Not estimable |
| Hoivik 2015 | 0 | 0 | 0 | 0 | 0 | 0 | | Not estimable |
| Holditch-Davis 2014 | 0 | 0 | 0 | 0 | 0 | 0 | | Not estimable |
| Holt 2021 | 0 | 0 | 0 | 0 | 0 | 0 | | Not estimable |
| Horowitz 2001 | 0 | 0 | 0 | 0 | 0 | 0 | | Not estimable |
| Horowitz 2013 | 0 | 0 | 0 | 0 | 0 | 0 | | Not estimable |
| Juffer 2005 | 0 | 0 | 0 | 0 | 0 | 0 | | Not estimable |
| Klein Velderman 2006 | 0 | 0 | 0 | 0 | 0 | 0 | | Not estimable |
| Knoche 2012 | 0 | 0 | 0 | 0 | 0 | 0 | | Not estimable |
| Kristensen 2020 | 0 | 0 | 0 | 0 | 0 | 0 | | Not estimable |
| Landry 2006 | 0 | 0 | 0 | 0 | 0 | 0 | | Not estimable |
| Lavallee 2022 | 0 | 0 | 0 | 0 | 0 | 0 | | Not estimable |
| Magil-Evans 2007 | 0 | 0 | 0 | 0 | 0 | 0 | | Not estimable |
| Meijssen 2010 | 0 | 0 | 0 | 0 | 0 | 0 | | Not estimable |
| Melnyk 2006 | 0 | 0 | 0 | 0 | 0 | 0 | | Not estimable |
| Mendelsohn 2007 | 0 | 0 | 0 | 0 | 0 | 0 | | Not estimable |
| Milgrom 2013 | 0 | 0 | 0 | 0 | 0 | 0 | | Not estimable |
| Nelson 2001 | 0 | 0 | 0 | 0 | 0 | 0 | | Not estimable |
| Neu 2010 | 0 | 0 | 0 | 0 | 0 | 0 | | Not estimable |
| Newnham 2009 | 0 | 0 | 0 | 0 | 0 | 0 | | Not estimable |
| Nugent 2017 | 0 | 0 | 0 | 0 | 0 | 0 | | Not estimable |
| Onozawa 2001 | 0 | 0 | 0 | 0 | 0 | 0 | | Not estimable |
| Oxford 2021 | 0 | 0 | 0 | 0 | 0 | 0 | | Not estimable |
| Perrone 2021 | 0 | 0 | 0 | 0 | 0 | 0 | | Not estimable |
| Pontoppidan 2016 | 0 | 0 | 0 | 0 | 0 | 0 | | Not estimable |
| Porter 2015 | 0 | 0 | 0 | 0 | 0 | 0 | | Not estimable |
| Ramsauer 2019 | 0 | 0 | 0 | 0 | 0 | 0 | | Not estimable |
| Ravn 2011 | 0 | 0 | 0 | 0 | 0 | 0 | | Not estimable |
| Rheinheimer 2022 | 0 | 0 | 0 | 0 | 0 | 0 | | Not estimable |
| Robertson 2019 | 0 | 0 | 0 | 0 | 0 | 0 | | Not estimable |
| Roby 2021 | 0 | 0 | 0 | 0 | 0 | 0 | | Not estimable |
| Sadler 2013 | 0 | 0 | 0 | 0 | 0 | 0 | | Not estimable |
| Sahlen Helmer 2020 | 0 | 0 | 0 | 0 | 0 | 0 | | Not estimable |
| Sajaniemi 2001 | 0 | 0 | 0 | 0 | 0 | 0 | | Not estimable |
| Salo 2019 | 0 | 0 | 0 | 0 | 0 | 0 | | Not estimable |
| Santelices 2010 | 0 | 0 | 0 | 0 | 0 | 0 | | Not estimable |
| Schuler 2000 | 0 | 0 | 0 | 0 | 0 | 0 | | Not estimable |
| Shoghi 2018 | 0 | 0 | 0 | 0 | 0 | 0 | | Not estimable |
| Sleed 2013 | 0 | 0 | 0 | 0 | 0 | 0 | | Not estimable |
| Steele 2019 | 0 | 0 | 0 | 0 | 0 | 0 | | Not estimable |
| Stein 2006 | 0 | 0 | 0 | 0 | 0 | 0 | | Not estimable |
| Stein 2018 | 0 | 0 | 0 | 0 | 0 | 0 | | Not estimable |
| Taneja 2020 | 0 | 0 | 0 | 0 | 0 | 0 | | Not estimable |
| Tereno 2017 | 0 | 0 | 0 | 0 | 0 | 0 | | Not estimable |
| Teti 2009 | 0 | 0 | 0 | 0 | 0 | 0 | | Not estimable |
| Tryphonopoulos 2020 | 0 | 0 | 0 | 0 | 0 | 0 | | Not estimable |
| Tsivos 2015 | 0 | 0 | 0 | 0 | 0 | 0 | | Not estimable |
| Twohig 2021 | 0 | 0 | 0 | 0 | 0 | 0 | | Not estimable |
| Valades 2021 | 0 | 0 | 0 | 0 | 0 | 0 | | Not estimable |
| van Doesum 2008 | 0 | 0 | 0 | 0 | 0 | 0 | | Not estimable |
| Vargas-Porras 2021 | 0 | 0 | 0 | 0 | 0 | 0 | | Not estimable |
| Walter 2019 | 0 | 0 | 0 | 0 | 0 | 0 | | Not estimable |
| White-Traut 2013 | 0 | 0 | 0 | 0 | 0 | 0 | | Not estimable |
| Williams 2020 | 0 | 0 | 0 | 0 | 0 | 0 | | Not estimable |
| Wulff 2021 | 0 | 0 | 0 | 0 | 0 | 0 | | Not estimable |
| Youn 2021 | 0 | 0 | 0 | 0 | 0 | 0 | | Not estimable |
| Yu 2022 | 0 | 0 | 0 | 0 | 0 | 0 | | Not estimable |
| Zelkowitz 2011 | 0 | 0 | 0 | 0 | 0 | 0 | | Not estimable |
| **Total (95% CI)** | | | **99** | | | **89** | **100.0%** | **-0.37 [-0.66, -0.08]** |

Heterogeneity: Tau² = 0.00; Chi² = 0.14, df = 2 (P = 0.93); I² = 0%

Test for overall effect: Z = 2.48 (P = 0.01)

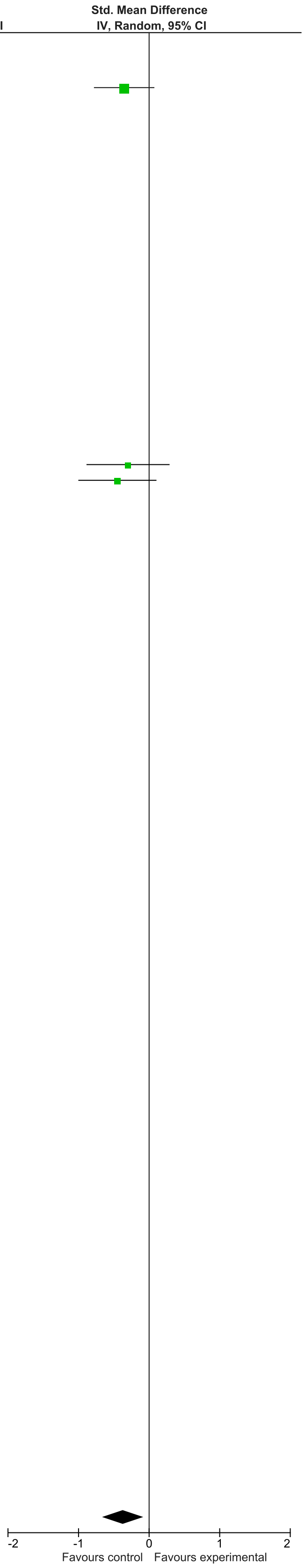

### eFigure 21. Cognitive Development 2-6-month Follow-Up Forrest Plot

| | Early Dyadic Intervention | | | Standard Care | | | | Std. Mean Difference |
|---|---|---|---|---|---|---|---|---|
| **Study or Subgroup** | **Mean** | **SD** | **Total** | **Mean** | **SD** | **Total** | **Weight** | **IV, Random, 95% CI** |
| Akai 2008 | 0 | 0 | 0 | 0 | 0 | 0 | | Not estimable |
| Alarcao 2021 | 0 | 0 | 0 | 0 | 0 | 0 | | Not estimable |
| Alvarenga 2019 | 0 | 0 | 0 | 0 | 0 | 0 | | Not estimable |
| Badr 2006 | 80.88 | 11.33 | 41 | 79.4 | 15.42 | 41 | 7.9% | 0.11 [-0.32, 0.54] |
| Baggett 2010 | 0 | 0 | 0 | 0 | 0 | 0 | | Not estimable |
| Baggett 2017 | 0 | 0 | 0 | 0 | 0 | 0 | | Not estimable |
| Barlow 2016 | 0 | 0 | 0 | 0 | 0 | 0 | | Not estimable |
| Barnes 2022 | 0 | 0 | 0 | 0 | 0 | 0 | | Not estimable |
| Berlin 2017 | 0 | 0 | 0 | 0 | 0 | 0 | | Not estimable |
| Berlin 2018 | 0 | 0 | 0 | 0 | 0 | 0 | | Not estimable |
| Bernard 2012 | 0 | 0 | 0 | 0 | 0 | 0 | | Not estimable |
| Betancourt 2020 | 0 | 0 | 0 | 0 | 0 | 0 | | Not estimable |
| Bick 2013 | 0 | 0 | 0 | 0 | 0 | 0 | | Not estimable |
| Borghini 2014 | 0 | 0 | 0 | 0 | 0 | 0 | | Not estimable |
| Brisch 2003 | 0 | 0 | 0 | 0 | 0 | 0 | | Not estimable |
| Browne 2005 | 0 | 0 | 0 | 0 | 0 | 0 | | Not estimable |
| Cates 2018 | 0 | 0 | 0 | 0 | 0 | 0 | | Not estimable |
| Cevasco 2008 | 0 | 0 | 0 | 0 | 0 | 0 | | Not estimable |
| Chiu 2009 | 0 | 0 | 0 | 0 | 0 | 0 | | Not estimable |
| Constantino 2001 | 0 | 0 | 0 | 0 | 0 | 0 | | Not estimable |
| Cooper 2009 | 0 | 0 | 0 | 0 | 0 | 0 | | Not estimable |
| Cooper 2015 | 0 | 0 | 0 | 0 | 0 | 0 | | Not estimable |
| Corrigan 2021 | 0 | 0 | 0 | 0 | 0 | 0 | | Not estimable |
| Dozier 2009 | 0 | 0 | 0 | 0 | 0 | 0 | | Not estimable |
| Eriksen 2018 | 0 | 0 | 0 | 0 | 0 | 0 | | Not estimable |
| Feil 2020 | 0 | 0 | 0 | 0 | 0 | 0 | | Not estimable |
| Feldman 2014 | 0 | 0 | 0 | 0 | 0 | 0 | | Not estimable |
| Firk 2021 | 9.57 | 1.4 | 24 | 9.48 | 1.7 | 21 | 4.3% | 0.06 [-0.53, 0.64] |
| Fonagy 2016 | 0 | 0 | 0 | 0 | 0 | 0 | | Not estimable |
| Gaden 2022 | 0 | 0 | 0 | 0 | 0 | 0 | | Not estimable |
| Georg 2021 | 0 | 0 | 0 | 0 | 0 | 0 | | Not estimable |
| Glazebrook 2007 | 0 | 0 | 0 | 0 | 0 | 0 | | Not estimable |
| Hane 2015 | 0 | 0 | 0 | 0 | 0 | 0 | | Not estimable |
| Heo 2019 | 0 | 0 | 0 | 0 | 0 | 0 | | Not estimable |
| Herbers 2020 | 0 | 0 | 0 | 0 | 0 | 0 | | Not estimable |
| Hoffenkamp 2015 | 0 | 0 | 0 | 0 | 0 | 0 | | Not estimable |
| Hoivik 2015 | 0 | 0 | 0 | 0 | 0 | 0 | | Not estimable |
| Holditch-Davis 2014 | 0 | 0 | 0 | 0 | 0 | 0 | | Not estimable |
| Holt 2021 | 0 | 0 | 0 | 0 | 0 | 0 | | Not estimable |
| Horowitz 2001 | 0 | 0 | 0 | 0 | 0 | 0 | | Not estimable |
| Horowitz 2013 | 0 | 0 | 0 | 0 | 0 | 0 | | Not estimable |
| Juffer 2005 | 0 | 0 | 0 | 0 | 0 | 0 | | Not estimable |
| Klein Velderman 2006 | 0 | 0 | 0 | 0 | 0 | 0 | | Not estimable |
| Knoche 2012 | 0 | 0 | 0 | 0 | 0 | 0 | | Not estimable |
| Kristensen 2020 | 0 | 0 | 0 | 0 | 0 | 0 | | Not estimable |
| Landry 2006 | 0 | 0 | 0 | 0 | 0 | 0 | | Not estimable |
| Lavallee 2022 | 0 | 0 | 0 | 0 | 0 | 0 | | Not estimable |
| Magil-Evans 2007 | 0 | 0 | 0 | 0 | 0 | 0 | | Not estimable |
| Meijssen 2010 | 0 | 0 | 0 | 0 | 0 | 0 | | Not estimable |
| Melnyk 2006 | 0 | 0 | 0 | 0 | 0 | 0 | | Not estimable |
| Mendelsohn 2007 | 0 | 0 | 0 | 0 | 0 | 0 | | Not estimable |
| Milgrom 2013 | 0 | 0 | 0 | 0 | 0 | 0 | | Not estimable |
| Nelson 2001 | 0 | 0 | 0 | 0 | 0 | 0 | | Not estimable |
| Neu 2010 | 0 | 0 | 0 | 0 | 0 | 0 | | Not estimable |
| Newnham 2009 | 0 | 0 | 0 | 0 | 0 | 0 | | Not estimable |
| Nugent 2017 | 0 | 0 | 0 | 0 | 0 | 0 | | Not estimable |
| Onozawa 2001 | 0 | 0 | 0 | 0 | 0 | 0 | | Not estimable |
| Oxford 2021 | 0 | 0 | 0 | 0 | 0 | 0 | | Not estimable |
| Perrone 2021 | 0 | 0 | 0 | 0 | 0 | 0 | | Not estimable |
| Pontoppidan 2016 | 0 | 0 | 0 | 0 | 0 | 0 | | Not estimable |
| Porter 2015 | 0 | 0 | 0 | 0 | 0 | 0 | | Not estimable |
| Ramsauer 2019 | 0 | 0 | 0 | 0 | 0 | 0 | | Not estimable |
| Ravn 2011 | 0 | 0 | 0 | 0 | 0 | 0 | | Not estimable |
| Rheinheimer 2022 | 0 | 0 | 0 | 0 | 0 | 0 | | Not estimable |
| Robertson 2019 | 0 | 0 | 0 | 0 | 0 | 0 | | Not estimable |
| Roby 2021 | 0 | 0 | 0 | 0 | 0 | 0 | | Not estimable |
| Sadler 2013 | 0 | 0 | 0 | 0 | 0 | 0 | | Not estimable |
| Sahlen Helmer 2020 | 0 | 0 | 0 | 0 | 0 | 0 | | Not estimable |
| Sajaniemi 2001 | 0 | 0 | 0 | 0 | 0 | 0 | | Not estimable |
| Salo 2019 | 0 | 0 | 0 | 0 | 0 | 0 | | Not estimable |
| Santelices 2010 | 0 | 0 | 0 | 0 | 0 | 0 | | Not estimable |
| Schuler 2000 | 0 | 0 | 0 | 0 | 0 | 0 | | Not estimable |
| Shoghi 2018 | 0 | 0 | 0 | 0 | 0 | 0 | | Not estimable |
| Sleed 2013 | 0 | 0 | 0 | 0 | 0 | 0 | | Not estimable |
| Steele 2019 | 0 | 0 | 0 | 0 | 0 | 0 | | Not estimable |
| Stein 2006 | 0 | 0 | 0 | 0 | 0 | 0 | | Not estimable |
| Stein 2018 | 0 | 0 | 0 | 0 | 0 | 0 | | Not estimable |
| Taneja 2020 | 96.47 | 12.7 | 276 | 95.49 | 13.8 | 276 | 53.3% | 0.07 [-0.09, 0.24] |
| Tereno 2017 | 0 | 0 | 0 | 0 | 0 | 0 | | Not estimable |
| Teti 2009 | 97.14 | 11.49 | 65 | 95.42 | 10.94 | 69 | 12.9% | 0.15 [-0.19, 0.49] |
| Tryphonopoulos 2020 | 0 | 0 | 0 | 0 | 0 | 0 | | Not estimable |
| Tsivos 2015 | 0 | 0 | 0 | 0 | 0 | 0 | | Not estimable |
| Twohig 2021 | 0 | 0 | 0 | 0 | 0 | 0 | | Not estimable |
| Valades 2021 | 0 | 0 | 0 | 0 | 0 | 0 | | Not estimable |
| van Doesum 2008 | 0 | 0 | 0 | 0 | 0 | 0 | | Not estimable |
| Vargas-Porras 2021 | 0 | 0 | 0 | 0 | 0 | 0 | | Not estimable |
| Walter 2019 | 0 | 0 | 0 | 0 | 0 | 0 | | Not estimable |
| White-Traut 2013 | 0 | 0 | 0 | 0 | 0 | 0 | | Not estimable |
| Williams 2020 | 0 | 0 | 0 | 0 | 0 | 0 | | Not estimable |
| Wulff 2021 | 0 | 0 | 0 | 0 | 0 | 0 | | Not estimable |
| Youn 2021 | 99.5 | 10.8 | 66 | 98.4 | 11.7 | 61 | 12.2% | 0.10 [-0.25, 0.45] |
| Yu 2022 | 0 | 0 | 0 | 0 | 0 | 0 | | Not estimable |
| Zelkowitz 2011 | 103.6 | 14.2 | 46 | 102.7 | 12.1 | 50 | 9.3% | 0.07 [-0.33, 0.47] |
| **Total (95% CI)** | | | **518** | | | **518** | **100.0%** | **0.09 [-0.03, 0.21]** |

Heterogeneity: Tau² = 0.00; Chi² = 0.20, df = 5 (P = 1.00); I² = 0%

Test for overall effect: Z = 1.42 (P = 0.16)

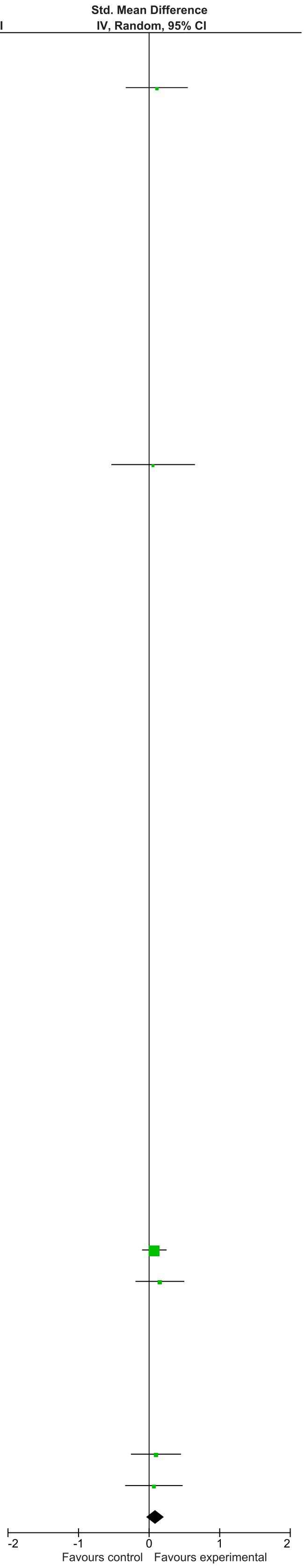

### eFigure 22. Cognitive Development 7-12-month Follow-Up Forrest Plot

| | Early Dyadic Intervention | | | Standard Care | | | | Std. Mean Difference |
|---|---|---|---|---|---|---|---|---|
| Study or Subgroup | Mean | SD | Total | Mean | SD | Total | Weight | IV, Random, 95% CI |
| Akai 2008 | 0 | 0 | 0 | 0 | 0 | 0 | | Not estimable |
| Alarcao 2021 | 0 | 0 | 0 | 0 | 0 | 0 | | Not estimable |
| Alvarenga 2019 | 0 | 0 | 0 | 0 | 0 | 0 | | Not estimable |
| Badr 2006 | 0 | 0 | 0 | 0 | 0 | 0 | | Not estimable |
| Baggett 2010 | 0 | 0 | 0 | 0 | 0 | 0 | | Not estimable |
| Baggett 2017 | 0 | 0 | 0 | 0 | 0 | 0 | | Not estimable |
| Barlow 2016 | 0 | 0 | 0 | 0 | 0 | 0 | | Not estimable |
| Barnes 2022 | 0 | 0 | 0 | 0 | 0 | 0 | | Not estimable |
| Berlin 2017 | 0 | 0 | 0 | 0 | 0 | 0 | | Not estimable |
| Berlin 2018 | 0 | 0 | 0 | 0 | 0 | 0 | | Not estimable |
| Bernard 2012 | 0 | 0 | 0 | 0 | 0 | 0 | | Not estimable |
| Betancourt 2020 | 0 | 0 | 0 | 0 | 0 | 0 | | Not estimable |
| Bick 2013 | 0 | 0 | 0 | 0 | 0 | 0 | | Not estimable |
| Borghini 2014 | 0 | 0 | 0 | 0 | 0 | 0 | | Not estimable |
| Brisch 2003 | 0 | 0 | 0 | 0 | 0 | 0 | | Not estimable |
| Browne 2005 | 0 | 0 | 0 | 0 | 0 | 0 | | Not estimable |
| Cates 2018 | 0 | 0 | 0 | 0 | 0 | 0 | | Not estimable |
| Cevasco 2008 | 0 | 0 | 0 | 0 | 0 | 0 | | Not estimable |
| Chiu 2009 | 0 | 0 | 0 | 0 | 0 | 0 | | Not estimable |
| Constantino 2001 | 0 | 0 | 0 | 0 | 0 | 0 | | Not estimable |
| Cooper 2009 | 0 | 0 | 0 | 0 | 0 | 0 | | Not estimable |
| Cooper 2015 | 0 | 0 | 0 | 0 | 0 | 0 | | Not estimable |
| Corrigan 2021 | 0 | 0 | 0 | 0 | 0 | 0 | | Not estimable |
| Dozier 2009 | 0 | 0 | 0 | 0 | 0 | 0 | | Not estimable |
| Eriksen 2018 | 0 | 0 | 0 | 0 | 0 | 0 | | Not estimable |
| Feil 2020 | 0 | 0 | 0 | 0 | 0 | 0 | | Not estimable |
| Feldman 2014 | 96.09 | 6.75 | 73 | 93.25 | 8.26 | 73 | 42.1% | 0.37 [0.05, 0.70] |
| Firk 2021 | 0 | 0 | 0 | 0 | 0 | 0 | | Not estimable |
| Fonagy 2016 | 0 | 0 | 0 | 0 | 0 | 0 | | Not estimable |
| Gaden 2022 | 0 | 0 | 0 | 0 | 0 | 0 | | Not estimable |
| Georg 2021 | 0 | 0 | 0 | 0 | 0 | 0 | | Not estimable |
| Glazebrook 2007 | 0 | 0 | 0 | 0 | 0 | 0 | | Not estimable |
| Hane 2015 | 0 | 0 | 0 | 0 | 0 | 0 | | Not estimable |
| Heo 2019 | 0 | 0 | 0 | 0 | 0 | 0 | | Not estimable |
| Herbers 2020 | 0 | 0 | 0 | 0 | 0 | 0 | | Not estimable |
| Hoffenkamp 2015 | 0 | 0 | 0 | 0 | 0 | 0 | | Not estimable |
| Hoivik 2015 | 0 | 0 | 0 | 0 | 0 | 0 | | Not estimable |
| Holditch-Davis 2014 | 0 | 0 | 0 | 0 | 0 | 0 | | Not estimable |
| Holt 2021 | 0 | 0 | 0 | 0 | 0 | 0 | | Not estimable |
| Horowitz 2001 | 0 | 0 | 0 | 0 | 0 | 0 | | Not estimable |
| Horowitz 2013 | 0 | 0 | 0 | 0 | 0 | 0 | | Not estimable |
| Juffer 2005 | 0 | 0 | 0 | 0 | 0 | 0 | | Not estimable |
| Klein Velderman 2006 | 0 | 0 | 0 | 0 | 0 | 0 | | Not estimable |
| Knoche 2012 | 0 | 0 | 0 | 0 | 0 | 0 | | Not estimable |
| Kristensen 2020 | 0 | 0 | 0 | 0 | 0 | 0 | | Not estimable |
| Landry 2006 | 0 | 0 | 0 | 0 | 0 | 0 | | Not estimable |
| Lavallee 2022 | 0 | 0 | 0 | 0 | 0 | 0 | | Not estimable |
| Magil-Evans 2007 | 0 | 0 | 0 | 0 | 0 | 0 | | Not estimable |
| Meijssen 2010 | 0 | 0 | 0 | 0 | 0 | 0 | | Not estimable |
| Melnyk 2006 | 0 | 0 | 0 | 0 | 0 | 0 | | Not estimable |
| Mendelsohn 2007 | 0 | 0 | 0 | 0 | 0 | 0 | | Not estimable |
| Milgrom 2013 | 0 | 0 | 0 | 0 | 0 | 0 | | Not estimable |
| Nelson 2001 | 0 | 0 | 0 | 0 | 0 | 0 | | Not estimable |
| Neu 2010 | 0 | 0 | 0 | 0 | 0 | 0 | | Not estimable |
| Newnham 2009 | 0 | 0 | 0 | 0 | 0 | 0 | | Not estimable |
| Nugent 2017 | 0 | 0 | 0 | 0 | 0 | 0 | | Not estimable |
| Onozawa 2001 | 0 | 0 | 0 | 0 | 0 | 0 | | Not estimable |
| Oxford 2021 | 0 | 0 | 0 | 0 | 0 | 0 | | Not estimable |
| Perrone 2021 | 0 | 0 | 0 | 0 | 0 | 0 | | Not estimable |
| Pontoppidan 2016 | 0 | 0 | 0 | 0 | 0 | 0 | | Not estimable |
| Porter 2015 | 0 | 0 | 0 | 0 | 0 | 0 | | Not estimable |
| Ramsauer 2019 | 0 | 0 | 0 | 0 | 0 | 0 | | Not estimable |
| Ravn 2011 | 0 | 0 | 0 | 0 | 0 | 0 | | Not estimable |
| Rheinheimer 2022 | 0 | 0 | 0 | 0 | 0 | 0 | | Not estimable |
| Robertson 2019 | 0 | 0 | 0 | 0 | 0 | 0 | | Not estimable |
| Roby 2021 | 0 | 0 | 0 | 0 | 0 | 0 | | Not estimable |
| Sadler 2013 | 0 | 0 | 0 | 0 | 0 | 0 | | Not estimable |
| Sahlen Helmer 2020 | 0 | 0 | 0 | 0 | 0 | 0 | | Not estimable |
| Sajaniemi 2001 | 0 | 0 | 0 | 0 | 0 | 0 | | Not estimable |
| Salo 2019 | 0 | 0 | 0 | 0 | 0 | 0 | | Not estimable |
| Santelices 2010 | 0 | 0 | 0 | 0 | 0 | 0 | | Not estimable |
| Schuler 2000 | 0 | 0 | 0 | 0 | 0 | 0 | | Not estimable |
| Shoghi 2018 | 0 | 0 | 0 | 0 | 0 | 0 | | Not estimable |
| Sleed 2013 | 0 | 0 | 0 | 0 | 0 | 0 | | Not estimable |
| Steele 2019 | 0 | 0 | 0 | 0 | 0 | 0 | | Not estimable |
| Stein 2006 | 0 | 0 | 0 | 0 | 0 | 0 | | Not estimable |
| Stein 2018 | 0 | 0 | 0 | 0 | 0 | 0 | | Not estimable |
| Taneja 2020 | 102.19 | 12.1 | 254 | 101.98 | 11.6 | 262 | 57.9% | 0.02 [-0.15, 0.19] |
| Tereno 2017 | 0 | 0 | 0 | 0 | 0 | 0 | | Not estimable |
| Teti 2009 | 0 | 0 | 0 | 0 | 0 | 0 | | Not estimable |
| Tryphonopoulos 2020 | 0 | 0 | 0 | 0 | 0 | 0 | | Not estimable |
| Tsivos 2015 | 0 | 0 | 0 | 0 | 0 | 0 | | Not estimable |
| Twohig 2021 | 0 | 0 | 0 | 0 | 0 | 0 | | Not estimable |
| Valades 2021 | 0 | 0 | 0 | 0 | 0 | 0 | | Not estimable |
| van Doesum 2008 | 0 | 0 | 0 | 0 | 0 | 0 | | Not estimable |
| Vargas-Porras 2021 | 0 | 0 | 0 | 0 | 0 | 0 | | Not estimable |
| Walter 2019 | 0 | 0 | 0 | 0 | 0 | 0 | | Not estimable |
| White-Traut 2013 | 0 | 0 | 0 | 0 | 0 | 0 | | Not estimable |
| Williams 2020 | 0 | 0 | 0 | 0 | 0 | 0 | | Not estimable |
| Wulff 2021 | 0 | 0 | 0 | 0 | 0 | 0 | | Not estimable |
| Youn 2021 | 0 | 0 | 0 | 0 | 0 | 0 | | Not estimable |
| Yu 2022 | 0 | 0 | 0 | 0 | 0 | 0 | | Not estimable |
| Zelkowitz 2011 | 0 | 0 | 0 | 0 | 0 | 0 | | Not estimable |
| **Total (95% CI)** | | | **327** | | | **335** | **100.0%** | **0.17 [-0.18, 0.51]** |

Heterogeneity: Tau² = 0.05; Chi² = 3.57, df = 1 (P = 0.06); I² = 72%
Test for overall effect: Z = 0.95 (P = 0.34)

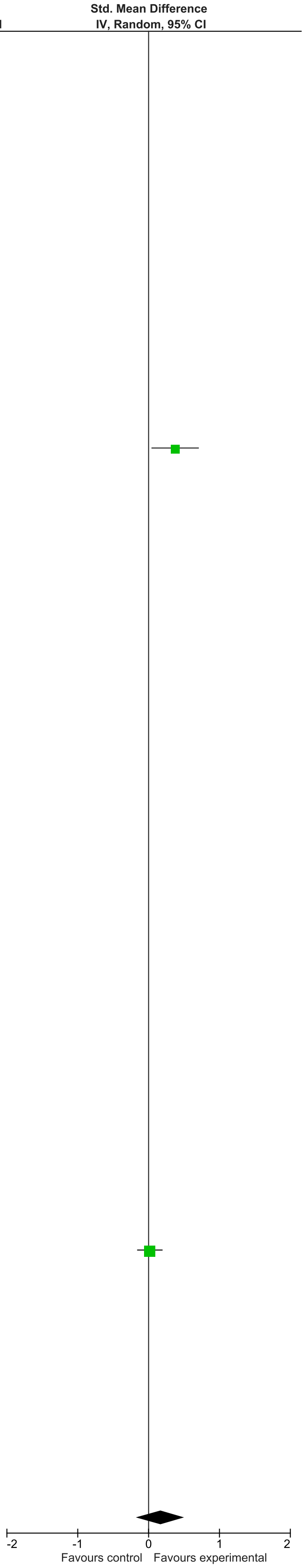

### eFigure 23. Cognitive Development 13-24-month Follow-Up Forrest Plot

| | Early Dyadic Intervention | | | Standard Care | | | | Std. Mean Difference |
|---|---|---|---|---|---|---|---|---|
| Study or Subgroup | Mean | SD | Total | Mean | SD | Total | Weight | IV, Random, 95% CI |
| Akai 2008 | 0 | 0 | 0 | 0 | 0 | 0 | | Not estimable |
| Alarcao 2021 | 0 | 0 | 0 | 0 | 0 | 0 | | Not estimable |
| Alvarenga 2019 | 0 | 0 | 0 | 0 | 0 | 0 | | Not estimable |
| Badr 2006 | 0 | 0 | 0 | 0 | 0 | 0 | | Not estimable |
| Baggett 2010 | 0 | 0 | 0 | 0 | 0 | 0 | | Not estimable |
| Baggett 2017 | 0 | 0 | 0 | 0 | 0 | 0 | | Not estimable |
| Barlow 2016 | 0 | 0 | 0 | 0 | 0 | 0 | | Not estimable |
| Barnes 2022 | 0 | 0 | 0 | 0 | 0 | 0 | | Not estimable |
| Berlin 2017 | 0 | 0 | 0 | 0 | 0 | 0 | | Not estimable |
| Berlin 2018 | 0 | 0 | 0 | 0 | 0 | 0 | | Not estimable |
| Bernard 2012 | 0 | 0 | 0 | 0 | 0 | 0 | | Not estimable |
| Betancourt 2020 | 0 | 0 | 0 | 0 | 0 | 0 | | Not estimable |
| Bick 2013 | 0 | 0 | 0 | 0 | 0 | 0 | | Not estimable |
| Borghini 2014 | 0 | 0 | 0 | 0 | 0 | 0 | | Not estimable |
| Brisch 2003 | 0 | 0 | 0 | 0 | 0 | 0 | | Not estimable |
| Browne 2005 | 0 | 0 | 0 | 0 | 0 | 0 | | Not estimable |
| Cates 2018 | 0 | 0 | 0 | 0 | 0 | 0 | | Not estimable |
| Cevasco 2008 | 0 | 0 | 0 | 0 | 0 | 0 | | Not estimable |
| Chiu 2009 | 0 | 0 | 0 | 0 | 0 | 0 | | Not estimable |
| Constantino 2001 | 0 | 0 | 0 | 0 | 0 | 0 | | Not estimable |
| Cooper 2009 | 0 | 0 | 0 | 0 | 0 | 0 | | Not estimable |
| Cooper 2015 | 0 | 0 | 0 | 0 | 0 | 0 | | Not estimable |
| Corrigan 2021 | 0 | 0 | 0 | 0 | 0 | 0 | | Not estimable |
| Dozier 2009 | 0 | 0 | 0 | 0 | 0 | 0 | | Not estimable |
| Eriksen 2018 | 0 | 0 | 0 | 0 | 0 | 0 | | Not estimable |
| Feil 2020 | 0 | 0 | 0 | 0 | 0 | 0 | | Not estimable |
| Feldman 2014 | 91.33 | 8.13 | 73 | 84.96 | 10.59 | 73 | 20.8% | 0.67 [0.34, 1.00] |
| Firk 2021 | 0 | 0 | 0 | 0 | 0 | 0 | | Not estimable |
| Fonagy 2016 | 0 | 0 | 0 | 0 | 0 | 0 | | Not estimable |
| Gaden 2022 | 0 | 0 | 0 | 0 | 0 | 0 | | Not estimable |
| Georg 2021 | 0 | 0 | 0 | 0 | 0 | 0 | | Not estimable |
| Glazebrook 2007 | 0 | 0 | 0 | 0 | 0 | 0 | | Not estimable |
| Hane 2015 | 98.73 | 17.98 | 45 | 93.66 | 13.36 | 31 | 15.6% | 0.31 [-0.15, 0.77] |
| Heo 2019 | 0 | 0 | 0 | 0 | 0 | 0 | | Not estimable |
| Herbers 2020 | 0 | 0 | 0 | 0 | 0 | 0 | | Not estimable |
| Hoffenkamp 2015 | 0 | 0 | 0 | 0 | 0 | 0 | | Not estimable |
| Hoivik 2015 | 0 | 0 | 0 | 0 | 0 | 0 | | Not estimable |
| Holditch-Davis 2014 | 0 | 0 | 0 | 0 | 0 | 0 | | Not estimable |
| Holt 2021 | 0 | 0 | 0 | 0 | 0 | 0 | | Not estimable |
| Horowitz 2001 | 0 | 0 | 0 | 0 | 0 | 0 | | Not estimable |
| Horowitz 2013 | 0 | 0 | 0 | 0 | 0 | 0 | | Not estimable |
| Juffer 2005 | 0 | 0 | 0 | 0 | 0 | 0 | | Not estimable |
| Klein Velderman 2006 | 0 | 0 | 0 | 0 | 0 | 0 | | Not estimable |
| Knoche 2012 | 0 | 0 | 0 | 0 | 0 | 0 | | Not estimable |
| Kristensen 2020 | 0 | 0 | 0 | 0 | 0 | 0 | | Not estimable |
| Landry 2006 | 0 | 0 | 0 | 0 | 0 | 0 | | Not estimable |
| Lavallee 2022 | 0 | 0 | 0 | 0 | 0 | 0 | | Not estimable |
| Magil-Evans 2007 | 0 | 0 | 0 | 0 | 0 | 0 | | Not estimable |
| Meijssen 2010 | 0 | 0 | 0 | 0 | 0 | 0 | | Not estimable |
| Melnyk 2006 | 0 | 0 | 0 | 0 | 0 | 0 | | Not estimable |
| Mendelsohn 2007 | 0 | 0 | 0 | 0 | 0 | 0 | | Not estimable |
| Milgrom 2013 | 107 | 16.52 | 55 | 105.87 | 17.98 | 52 | 18.8% | 0.07 [-0.31, 0.44] |
| Nelson 2001 | 82.67 | 23.31 | 15 | 74.67 | 17.32 | 12 | 8.1% | 0.37 [-0.40, 1.14] |
| Neu 2010 | 0 | 0 | 0 | 0 | 0 | 0 | | Not estimable |
| Newnham 2009 | 0 | 0 | 0 | 0 | 0 | 0 | | Not estimable |
| Nugent 2017 | 0 | 0 | 0 | 0 | 0 | 0 | | Not estimable |
| Onozawa 2001 | 0 | 0 | 0 | 0 | 0 | 0 | | Not estimable |
| Oxford 2021 | 0 | 0 | 0 | 0 | 0 | 0 | | Not estimable |
| Perrone 2021 | 0 | 0 | 0 | 0 | 0 | 0 | | Not estimable |
| Pontoppidan 2016 | 0 | 0 | 0 | 0 | 0 | 0 | | Not estimable |
| Porter 2015 | 0 | 0 | 0 | 0 | 0 | 0 | | Not estimable |
| Ramsauer 2019 | 0 | 0 | 0 | 0 | 0 | 0 | | Not estimable |
| Ravn 2011 | 0 | 0 | 0 | 0 | 0 | 0 | | Not estimable |
| Rheinheimer 2022 | 0 | 0 | 0 | 0 | 0 | 0 | | Not estimable |
| Robertson 2019 | 0 | 0 | 0 | 0 | 0 | 0 | | Not estimable |
| Roby 2021 | 0 | 0 | 0 | 0 | 0 | 0 | | Not estimable |
| Sadler 2013 | 0 | 0 | 0 | 0 | 0 | 0 | | Not estimable |
| Sahlen Helmer 2020 | 0 | 0 | 0 | 0 | 0 | 0 | | Not estimable |
| Sajaniemi 2001 | 0 | 0 | 0 | 0 | 0 | 0 | | Not estimable |
| Salo 2019 | 0 | 0 | 0 | 0 | 0 | 0 | | Not estimable |
| Santelices 2010 | 0 | 0 | 0 | 0 | 0 | 0 | | Not estimable |
| Schuler 2000 | 0 | 0 | 0 | 0 | 0 | 0 | | Not estimable |
| Shoghi 2018 | 0 | 0 | 0 | 0 | 0 | 0 | | Not estimable |
| Sleed 2013 | 0 | 0 | 0 | 0 | 0 | 0 | | Not estimable |
| Steele 2019 | 0 | 0 | 0 | 0 | 0 | 0 | | Not estimable |
| Stein 2006 | 0 | 0 | 0 | 0 | 0 | 0 | | Not estimable |
| Stein 2018 | 99.2 | 12 | 62 | 99.9 | 12.9 | 67 | 20.2% | -0.06 [-0.40, 0.29] |
| Taneja 2020 | 0 | 0 | 0 | 0 | 0 | 0 | | Not estimable |
| Tereno 2017 | 0 | 0 | 0 | 0 | 0 | 0 | | Not estimable |
| Teti 2009 | 0 | 0 | 0 | 0 | 0 | 0 | | Not estimable |
| Tryphonopoulos 2020 | 0 | 0 | 0 | 0 | 0 | 0 | | Not estimable |
| Tsivos 2015 | 0 | 0 | 0 | 0 | 0 | 0 | | Not estimable |
| Twohig 2021 | 0 | 0 | 0 | 0 | 0 | 0 | | Not estimable |
| Valades 2021 | 0 | 0 | 0 | 0 | 0 | 0 | | Not estimable |
| van Doesum 2008 | 0 | 0 | 0 | 0 | 0 | 0 | | Not estimable |
| Vargas-Porras 2021 | 0 | 0 | 0 | 0 | 0 | 0 | | Not estimable |
| Walter 2019 | 0 | 0 | 0 | 0 | 0 | 0 | | Not estimable |
| White-Traut 2013 | 0 | 0 | 0 | 0 | 0 | 0 | | Not estimable |
| Williams 2020 | 0 | 0 | 0 | 0 | 0 | 0 | | Not estimable |
| Wulff 2021 | 0 | 0 | 0 | 0 | 0 | 0 | | Not estimable |
| Youn 2021 | 100.3 | 17.1 | 43 | 98.3 | 15.4 | 39 | 16.6% | 0.12 [-0.31, 0.56] |
| Yu 2022 | 0 | 0 | 0 | 0 | 0 | 0 | | Not estimable |
| Zelkowitz 2011 | 0 | 0 | 0 | 0 | 0 | 0 | | Not estimable |
| **Total (95% CI)** | | | **293** | | | **274** | **100.0%** | **0.24 [-0.01, 0.49]** |

Heterogeneity: Tau² = 0.05; Chi² = 10.53, df = 5 (P = 0.06); I² = 53%
Test for overall effect: Z = 1.87 (P = 0.06)

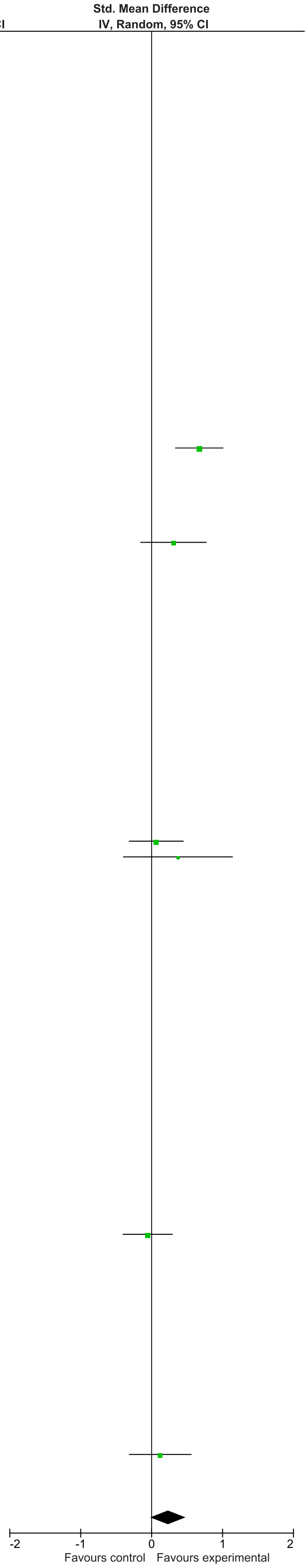

### eFigure 24. Cognitive Development 25+-month Follow-Up Forrest Plot

| | Early Dyadic Intervention | | | Standard Care | | | | Std. Mean Difference |
|---|---|---|---|---|---|---|---|---|
| Study or Subgroup | Mean | SD | Total | Mean | SD | Total | Weight | IV, Random, 95% CI |
| Akai 2008 | 0 | 0 | 0 | 0 | 0 | 0 | | Not estimable |
| Alarcao 2021 | 0 | 0 | 0 | 0 | 0 | 0 | | Not estimable |
| Alvarenga 2019 | 0 | 0 | 0 | 0 | 0 | 0 | | Not estimable |
| Badr 2006 | 0 | 0 | 0 | 0 | 0 | 0 | | Not estimable |
| Baggett 2010 | 0 | 0 | 0 | 0 | 0 | 0 | | Not estimable |
| Baggett 2017 | 0 | 0 | 0 | 0 | 0 | 0 | | Not estimable |
| Barlow 2016 | 0 | 0 | 0 | 0 | 0 | 0 | | Not estimable |
| Barnes 2022 | 0 | 0 | 0 | 0 | 0 | 0 | | Not estimable |
| Berlin 2017 | 0 | 0 | 0 | 0 | 0 | 0 | | Not estimable |
| Berlin 2018 | 0 | 0 | 0 | 0 | 0 | 0 | | Not estimable |
| Bernard 2012 | 0 | 0 | 0 | 0 | 0 | 0 | | Not estimable |
| Betancourt 2020 | 0 | 0 | 0 | 0 | 0 | 0 | | Not estimable |
| Bick 2013 | 0 | 0 | 0 | 0 | 0 | 0 | | Not estimable |
| Borghini 2014 | 0 | 0 | 0 | 0 | 0 | 0 | | Not estimable |
| Brisch 2003 | 0 | 0 | 0 | 0 | 0 | 0 | | Not estimable |
| Browne 2005 | 0 | 0 | 0 | 0 | 0 | 0 | | Not estimable |
| Cates 2018 | 0 | 0 | 0 | 0 | 0 | 0 | | Not estimable |
| Cevasco 2008 | 0 | 0 | 0 | 0 | 0 | 0 | | Not estimable |
| Chiu 2009 | 0 | 0 | 0 | 0 | 0 | 0 | | Not estimable |
| Constantino 2001 | 0 | 0 | 0 | 0 | 0 | 0 | | Not estimable |
| Cooper 2009 | 0 | 0 | 0 | 0 | 0 | 0 | | Not estimable |
| Cooper 2015 | 0 | 0 | 0 | 0 | 0 | 0 | | Not estimable |
| Corrigan 2021 | 0 | 0 | 0 | 0 | 0 | 0 | | Not estimable |
| Dozier 2009 | 0 | 0 | 0 | 0 | 0 | 0 | | Not estimable |
| Eriksen 2018 | 0 | 0 | 0 | 0 | 0 | 0 | | Not estimable |
| Feil 2020 | 0 | 0 | 0 | 0 | 0 | 0 | | Not estimable |
| Feldman 2014 | 95.62 | 12.94 | 73 | 89.3 | 12.29 | 73 | 52.1% | 0.50 [0.17, 0.83] |
| Firk 2021 | 0 | 0 | 0 | 0 | 0 | 0 | | Not estimable |
| Fonagy 2016 | 0 | 0 | 0 | 0 | 0 | 0 | | Not estimable |
| Gaden 2022 | 0 | 0 | 0 | 0 | 0 | 0 | | Not estimable |
| Georg 2021 | 0 | 0 | 0 | 0 | 0 | 0 | | Not estimable |
| Glazebrook 2007 | 0 | 0 | 0 | 0 | 0 | 0 | | Not estimable |
| Hane 2015 | 0 | 0 | 0 | 0 | 0 | 31 | | Not estimable |
| Heo 2019 | 0 | 0 | 0 | 0 | 0 | 0 | | Not estimable |
| Herbers 2020 | 0 | 0 | 0 | 0 | 0 | 0 | | Not estimable |
| Hoffenkamp 2015 | 0 | 0 | 0 | 0 | 0 | 0 | | Not estimable |
| Hoivik 2015 | 0 | 0 | 0 | 0 | 0 | 0 | | Not estimable |
| Holditch-Davis 2014 | 0 | 0 | 0 | 0 | 0 | 0 | | Not estimable |
| Holt 2021 | 0 | 0 | 0 | 0 | 0 | 0 | | Not estimable |
| Horowitz 2001 | 0 | 0 | 0 | 0 | 0 | 0 | | Not estimable |
| Horowitz 2013 | 0 | 0 | 0 | 0 | 0 | 0 | | Not estimable |
| Juffer 2005 | 0 | 0 | 0 | 0 | 0 | 0 | | Not estimable |
| Klein Velderman 2006 | 0 | 0 | 0 | 0 | 0 | 0 | | Not estimable |
| Knoche 2012 | 0 | 0 | 0 | 0 | 0 | 0 | | Not estimable |
| Kristensen 2020 | 0 | 0 | 0 | 0 | 0 | 0 | | Not estimable |
| Landry 2006 | 0 | 0 | 0 | 0 | 0 | 0 | | Not estimable |
| Lavallee 2022 | 0 | 0 | 0 | 0 | 0 | 0 | | Not estimable |
| Magil-Evans 2007 | 0 | 0 | 0 | 0 | 0 | 0 | | Not estimable |
| Meijssen 2010 | 0 | 0 | 0 | 0 | 0 | 0 | | Not estimable |
| Melnyk 2006 | 0 | 0 | 0 | 0 | 0 | 0 | | Not estimable |
| Mendelsohn 2007 | 0 | 0 | 0 | 0 | 0 | 0 | | Not estimable |
| Milgrom 2013 | 106.51 | 15.37 | 41 | 108.57 | 12.99 | 47 | 47.9% | -0.14 [-0.56, 0.28] |
| Nelson 2001 | 0 | 0 | 0 | 0 | 0 | 0 | | Not estimable |
| Neu 2010 | 0 | 0 | 0 | 0 | 0 | 0 | | Not estimable |
| Newnham 2009 | 0 | 0 | 0 | 0 | 0 | 0 | | Not estimable |
| Nugent 2017 | 0 | 0 | 0 | 0 | 0 | 0 | | Not estimable |
| Onozawa 2001 | 0 | 0 | 0 | 0 | 0 | 0 | | Not estimable |
| Oxford 2021 | 0 | 0 | 0 | 0 | 0 | 0 | | Not estimable |
| Perrone 2021 | 0 | 0 | 0 | 0 | 0 | 0 | | Not estimable |
| Pontoppidan 2016 | 0 | 0 | 0 | 0 | 0 | 0 | | Not estimable |
| Porter 2015 | 0 | 0 | 0 | 0 | 0 | 0 | | Not estimable |
| Ramsauer 2019 | 0 | 0 | 0 | 0 | 0 | 0 | | Not estimable |
| Ravn 2011 | 0 | 0 | 0 | 0 | 0 | 0 | | Not estimable |
| Rheinheimer 2022 | 0 | 0 | 0 | 0 | 0 | 0 | | Not estimable |
| Robertson 2019 | 0 | 0 | 0 | 0 | 0 | 0 | | Not estimable |
| Roby 2021 | 0 | 0 | 0 | 0 | 0 | 0 | | Not estimable |
| Sadler 2013 | 0 | 0 | 0 | 0 | 0 | 0 | | Not estimable |
| Sahlen Helmer 2020 | 0 | 0 | 0 | 0 | 0 | 0 | | Not estimable |
| Sajaniemi 2001 | 0 | 0 | 0 | 0 | 0 | 0 | | Not estimable |
| Salo 2019 | 0 | 0 | 0 | 0 | 0 | 0 | | Not estimable |
| Santelices 2010 | 0 | 0 | 0 | 0 | 0 | 0 | | Not estimable |
| Schuler 2000 | 0 | 0 | 0 | 0 | 0 | 0 | | Not estimable |
| Shoghi 2018 | 0 | 0 | 0 | 0 | 0 | 0 | | Not estimable |
| Sleed 2013 | 0 | 0 | 0 | 0 | 0 | 0 | | Not estimable |
| Steele 2019 | 0 | 0 | 0 | 0 | 0 | 0 | | Not estimable |
| Stein 2006 | 0 | 0 | 0 | 0 | 0 | 0 | | Not estimable |
| Stein 2018 | 0 | 0 | 0 | 0 | 0 | 0 | | Not estimable |
| Taneja 2020 | 0 | 0 | 0 | 0 | 0 | 0 | | Not estimable |
| Tereno 2017 | 0 | 0 | 0 | 0 | 0 | 0 | | Not estimable |
| Teti 2009 | 0 | 0 | 0 | 0 | 0 | 0 | | Not estimable |
| Tryphonopoulos 2020 | 0 | 0 | 0 | 0 | 0 | 0 | | Not estimable |
| Tsivos 2015 | 0 | 0 | 0 | 0 | 0 | 0 | | Not estimable |
| Twohig 2021 | 0 | 0 | 0 | 0 | 0 | 0 | | Not estimable |
| Valades 2021 | 0 | 0 | 0 | 0 | 0 | 0 | | Not estimable |
| van Doesum 2008 | 0 | 0 | 0 | 0 | 0 | 0 | | Not estimable |
| Vargas-Porras 2021 | 0 | 0 | 0 | 0 | 0 | 0 | | Not estimable |
| Walter 2019 | 0 | 0 | 0 | 0 | 0 | 0 | | Not estimable |
| White-Traut 2013 | 0 | 0 | 0 | 0 | 0 | 0 | | Not estimable |
| Williams 2020 | 0 | 0 | 0 | 0 | 0 | 0 | | Not estimable |
| Wulff 2021 | 0 | 0 | 0 | 0 | 0 | 0 | | Not estimable |
| Youn 2021 | 0 | 0 | 0 | 0 | 0 | 0 | | Not estimable |
| Yu 2022 | 0 | 0 | 0 | 0 | 0 | 0 | | Not estimable |
| Zelkowitz 2011 | 0 | 0 | 0 | 0 | 0 | 0 | | Not estimable |
| **Total (95% CI)** | | | **114** | | | **151** | **100.0%** | **0.19 [-0.44, 0.82]** |

Heterogeneity: Tau² = 0.17; Chi² = 5.57, df = 1 (P = 0.02); I² = 82%

Test for overall effect: Z = 0.59 (P = 0.55)

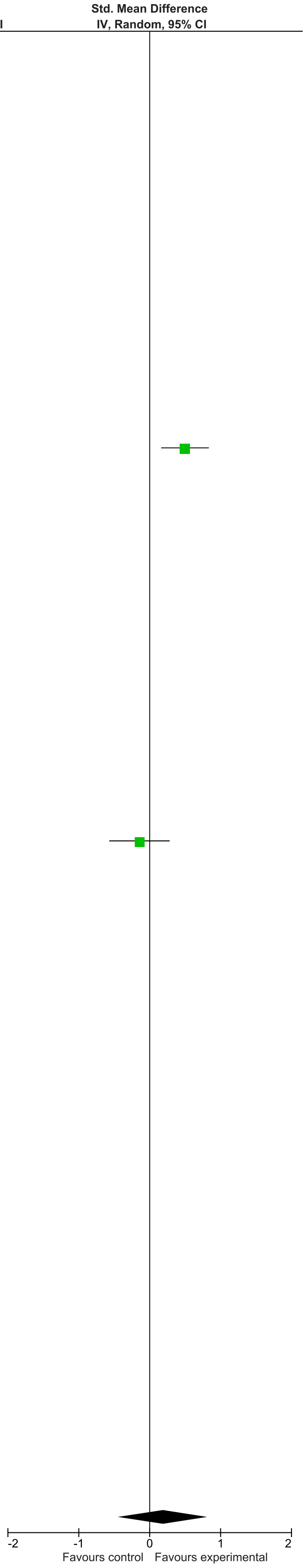

### eFigure 25. Language Development 0-1-month Follow-Up Forrest Plot

| | Early Dyadic Intervention | | | Standard Care | | | | Std. Mean Difference |
|---|---|---|---|---|---|---|---|---|
| Study or Subgroup | Mean | SD | Total | Mean | SD | Total | Weight | IV, Random, 95% CI |
| Akai 2008 | 0 | 0 | 0 | 0 | 0 | 0 | | Not estimable |
| Alarcao 2021 | 0 | 0 | 0 | 0 | 0 | 0 | | Not estimable |
| Alvarenga 2019 | 0 | 0 | 0 | 0 | 0 | 0 | | Not estimable |
| Badr 2006 | 0 | 0 | 0 | 0 | 0 | 0 | | Not estimable |
| Baggett 2010 | 0 | 0 | 0 | 0 | 0 | 0 | | Not estimable |
| Baggett 2017 | 0 | 0 | 0 | 0 | 0 | 0 | | Not estimable |
| Barlow 2016 | 0 | 0 | 0 | 0 | 0 | 0 | | Not estimable |
| Barnes 2022 | 0 | 0 | 0 | 0 | 0 | 0 | | Not estimable |
| Berlin 2017 | 0 | 0 | 0 | 0 | 0 | 0 | | Not estimable |
| Berlin 2018 | 0 | 0 | 0 | 0 | 0 | 0 | | Not estimable |
| Bernard 2012 | 0 | 0 | 0 | 0 | 0 | 0 | | Not estimable |
| Betancourt 2020 | 0 | 0 | 0 | 0 | 0 | 0 | | Not estimable |
| Bick 2013 | 0 | 0 | 0 | 0 | 0 | 0 | | Not estimable |
| Borghini 2014 | 0 | 0 | 0 | 0 | 0 | 0 | | Not estimable |
| Brisch 2003 | 0 | 0 | 0 | 0 | 0 | 0 | | Not estimable |
| Browne 2005 | 0 | 0 | 0 | 0 | 0 | 0 | | Not estimable |
| Cates 2018 | 0 | 0 | 0 | 0 | 0 | 0 | | Not estimable |
| Cevasco 2008 | 0 | 0 | 0 | 0 | 0 | 0 | | Not estimable |
| Chiu 2009 | 0 | 0 | 0 | 0 | 0 | 0 | | Not estimable |
| Constantino 2001 | 0 | 0 | 0 | 0 | 0 | 0 | | Not estimable |
| Cooper 2009 | 0 | 0 | 0 | 0 | 0 | 0 | | Not estimable |
| Cooper 2015 | 0 | 0 | 0 | 0 | 0 | 0 | | Not estimable |
| Corrigan 2021 | 0 | 0 | 0 | 0 | 0 | 0 | | Not estimable |
| Dozier 2009 | 0 | 0 | 0 | 0 | 0 | 0 | | Not estimable |
| Eriksen 2018 | 0 | 0 | 0 | 0 | 0 | 0 | | Not estimable |
| Feil 2020 | 0 | 0 | 0 | 0 | 0 | 0 | | Not estimable |
| Feldman 2014 | 0 | 0 | 0 | 0 | 0 | 0 | | Not estimable |
| Firk 2021 | 8.54 | 1.9 | 27 | 8.73 | 1.93 | 20 | 46.5% | -0.10 [-0.68, 0.48] |
| Fonagy 2016 | 92.6 | 1.9 | 28 | 93.4 | 16.8 | 25 | 53.5% | -0.07 [-0.61, 0.47] |
| Gaden 2022 | 0 | 0 | 0 | 0 | 0 | 0 | | Not estimable |
| Georg 2021 | 0 | 0 | 0 | 0 | 0 | 0 | | Not estimable |
| Glazebrook 2007 | 0 | 0 | 0 | 0 | 0 | 0 | | Not estimable |
| Hane 2015 | 0 | 0 | 0 | 0 | 0 | 0 | | Not estimable |
| Heo 2019 | 0 | 0 | 0 | 0 | 0 | 0 | | Not estimable |
| Herbers 2020 | 0 | 0 | 0 | 0 | 0 | 0 | | Not estimable |
| Hoffenkamp 2015 | 0 | 0 | 0 | 0 | 0 | 0 | | Not estimable |
| Hoivik 2015 | 0 | 0 | 0 | 0 | 0 | 0 | | Not estimable |
| Holditch-Davis 2014 | 0 | 0 | 0 | 0 | 0 | 0 | | Not estimable |
| Holt 2021 | 0 | 0 | 0 | 0 | 0 | 0 | | Not estimable |
| Horowitz 2001 | 0 | 0 | 0 | 0 | 0 | 0 | | Not estimable |
| Horowitz 2013 | 0 | 0 | 0 | 0 | 0 | 0 | | Not estimable |
| Juffer 2005 | 0 | 0 | 0 | 0 | 0 | 0 | | Not estimable |
| Klein Velderman 2006 | 0 | 0 | 0 | 0 | 0 | 0 | | Not estimable |
| Knoche 2012 | 0 | 0 | 0 | 0 | 0 | 0 | | Not estimable |
| Kristensen 2020 | 0 | 0 | 0 | 0 | 0 | 0 | | Not estimable |
| Landry 2006 | 0 | 0 | 0 | 0 | 0 | 0 | | Not estimable |
| Lavallee 2022 | 0 | 0 | 0 | 0 | 0 | 0 | | Not estimable |
| Magil-Evans 2007 | 0 | 0 | 0 | 0 | 0 | 0 | | Not estimable |
| Meijssen 2010 | 0 | 0 | 0 | 0 | 0 | 0 | | Not estimable |
| Melnyk 2006 | 0 | 0 | 0 | 0 | 0 | 0 | | Not estimable |
| Mendelsohn 2007 | 0 | 0 | 0 | 0 | 0 | 0 | | Not estimable |
| Milgrom 2013 | 0 | 0 | 0 | 0 | 0 | 0 | | Not estimable |
| Nelson 2001 | 0 | 0 | 0 | 0 | 0 | 0 | | Not estimable |
| Neu 2010 | 0 | 0 | 0 | 0 | 0 | 0 | | Not estimable |
| Newnham 2009 | 0 | 0 | 0 | 0 | 0 | 0 | | Not estimable |
| Nugent 2017 | 0 | 0 | 0 | 0 | 0 | 0 | | Not estimable |
| Onozawa 2001 | 0 | 0 | 0 | 0 | 0 | 0 | | Not estimable |
| Oxford 2021 | 0 | 0 | 0 | 0 | 0 | 0 | | Not estimable |
| Perrone 2021 | 0 | 0 | 0 | 0 | 0 | 0 | | Not estimable |
| Pontoppidan 2016 | 0 | 0 | 0 | 0 | 0 | 0 | | Not estimable |
| Porter 2015 | 0 | 0 | 0 | 0 | 0 | 0 | | Not estimable |
| Ramsauer 2019 | 0 | 0 | 0 | 0 | 0 | 0 | | Not estimable |
| Ravn 2011 | 0 | 0 | 0 | 0 | 0 | 0 | | Not estimable |
| Rheinheimer 2022 | 0 | 0 | 0 | 0 | 0 | 0 | | Not estimable |
| Robertson 2019 | 0 | 0 | 0 | 0 | 0 | 0 | | Not estimable |
| Roby 2021 | 0 | 0 | 0 | 0 | 0 | 0 | | Not estimable |
| Sadler 2013 | 0 | 0 | 0 | 0 | 0 | 0 | | Not estimable |
| Sahlen Helmer 2020 | 0 | 0 | 0 | 0 | 0 | 0 | | Not estimable |
| Sajaniemi 2001 | 0 | 0 | 0 | 0 | 0 | 0 | | Not estimable |
| Salo 2019 | 0 | 0 | 0 | 0 | 0 | 0 | | Not estimable |
| Santelices 2010 | 0 | 0 | 0 | 0 | 0 | 0 | | Not estimable |
| Schuler 2000 | 0 | 0 | 0 | 0 | 0 | 0 | | Not estimable |
| Shoghi 2018 | 0 | 0 | 0 | 0 | 0 | 0 | | Not estimable |
| Sleed 2013 | 0 | 0 | 0 | 0 | 0 | 0 | | Not estimable |
| Steele 2019 | 0 | 0 | 0 | 0 | 0 | 0 | | Not estimable |
| Stein 2006 | 0 | 0 | 0 | 0 | 0 | 0 | | Not estimable |
| Stein 2018 | 0 | 0 | 0 | 0 | 0 | 0 | | Not estimable |
| Taneja 2020 | 0 | 0 | 0 | 0 | 0 | 0 | | Not estimable |
| Tereno 2017 | 0 | 0 | 0 | 0 | 0 | 0 | | Not estimable |
| Teti 2009 | 0 | 0 | 0 | 0 | 0 | 0 | | Not estimable |
| Tryphonopoulos 2020 | 0 | 0 | 0 | 0 | 0 | 0 | | Not estimable |
| Tsivos 2015 | 0 | 0 | 0 | 0 | 0 | 0 | | Not estimable |
| Twohig 2021 | 0 | 0 | 0 | 0 | 0 | 0 | | Not estimable |
| Valades 2021 | 0 | 0 | 0 | 0 | 0 | 0 | | Not estimable |
| van Doesum 2008 | 0 | 0 | 0 | 0 | 0 | 0 | | Not estimable |
| Vargas-Porras 2021 | 0 | 0 | 0 | 0 | 0 | 0 | | Not estimable |
| Walter 2019 | 0 | 0 | 0 | 0 | 0 | 0 | | Not estimable |
| White-Traut 2013 | 0 | 0 | 0 | 0 | 0 | 0 | | Not estimable |
| Williams 2020 | 0 | 0 | 0 | 0 | 0 | 0 | | Not estimable |
| Wulff 2021 | 0 | 0 | 0 | 0 | 0 | 0 | | Not estimable |
| Youn 2021 | 0 | 0 | 0 | 0 | 0 | 0 | | Not estimable |
| Yu 2022 | 0 | 0 | 0 | 0 | 0 | 0 | | Not estimable |
| Zelkowitz 2011 | 0 | 0 | 0 | 0 | 0 | 0 | | Not estimable |
| **Total (95% CI)** | | | **55** | | | **45** | **100.0%** | **-0.08 [-0.48, 0.31]** |

Heterogeneity: Tau² = 0.00; Chi² = 0.01, df = 1 (P = 0.94); I² = 0%
Test for overall effect: Z = 0.41 (P = 0.68)

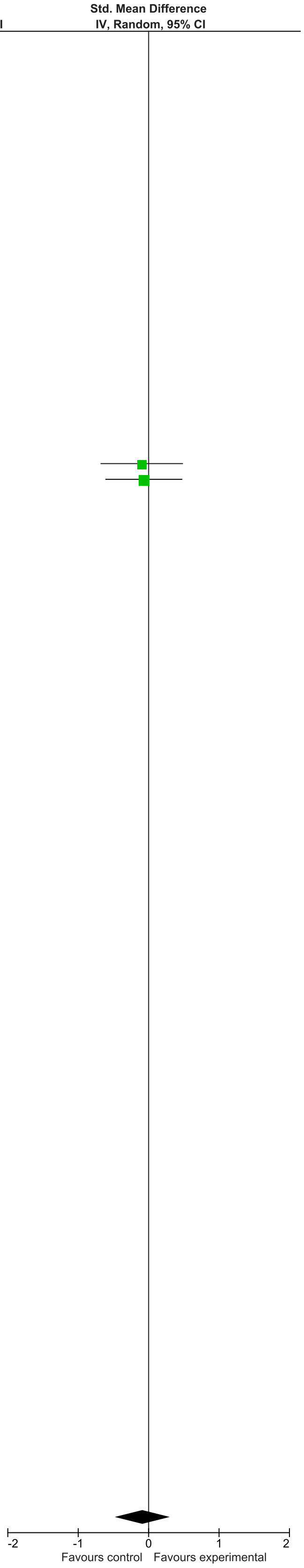

### eFigure 26. Language Development 2-6-month Follow-Up Forrest Plot

| Study or Subgroup | Early Dyadic Intervention Mean | SD | Total | Standard Care Mean | SD | Total | Weight | Std. Mean Difference IV, Random, 95% CI |
|---|---|---|---|---|---|---|---|---|
| Akai 2008 | 0 | 0 | 0 | 0 | 0 | 0 | | Not estimable |
| Alarcao 2021 | 0 | 0 | 0 | 0 | 0 | 0 | | Not estimable |
| Alvarenga 2019 | 0 | 0 | 0 | 0 | 0 | 0 | | Not estimable |
| Badr 2006 | 0 | 0 | 0 | 0 | 0 | 0 | | Not estimable |
| Baggett 2010 | 0 | 0 | 0 | 0 | 0 | 0 | | Not estimable |
| Baggett 2017 | 0 | 0 | 0 | 0 | 0 | 0 | | Not estimable |
| Barlow 2016 | 0 | 0 | 0 | 0 | 0 | 0 | | Not estimable |
| Barnes 2022 | 0 | 0 | 0 | 0 | 0 | 0 | | Not estimable |
| Berlin 2017 | 0 | 0 | 0 | 0 | 0 | 0 | | Not estimable |
| Berlin 2018 | 0 | 0 | 0 | 0 | 0 | 0 | | Not estimable |
| Bernard 2012 | 0 | 0 | 0 | 0 | 0 | 0 | | Not estimable |
| Betancourt 2020 | 0 | 0 | 0 | 0 | 0 | 0 | | Not estimable |
| Bick 2013 | 0 | 0 | 0 | 0 | 0 | 0 | | Not estimable |
| Borghini 2014 | 0 | 0 | 0 | 0 | 0 | 0 | | Not estimable |
| Brisch 2003 | 0 | 0 | 0 | 0 | 0 | 0 | | Not estimable |
| Browne 2005 | 0 | 0 | 0 | 0 | 0 | 0 | | Not estimable |
| Cates 2018 | 0 | 0 | 0 | 0 | 0 | 0 | | Not estimable |
| Cevasco 2008 | 0 | 0 | 0 | 0 | 0 | 0 | | Not estimable |
| Chiu 2009 | 0 | 0 | 0 | 0 | 0 | 0 | | Not estimable |
| Constantino 2001 | 0 | 0 | 0 | 0 | 0 | 0 | | Not estimable |
| Cooper 2009 | 0 | 0 | 0 | 0 | 0 | 0 | | Not estimable |
| Cooper 2015 | 0 | 0 | 0 | 0 | 0 | 0 | | Not estimable |
| Corrigan 2021 | 0 | 0 | 0 | 0 | 0 | 0 | | Not estimable |
| Dozier 2009 | 0 | 0 | 0 | 0 | 0 | 0 | | Not estimable |
| Eriksen 2018 | 0 | 0 | 0 | 0 | 0 | 0 | | Not estimable |
| Feil 2020 | 0 | 0 | 0 | 0 | 0 | 0 | | Not estimable |
| Feldman 2014 | 0 | 0 | 0 | 0 | 0 | 0 | | Not estimable |
| Firk 2021 | 8.9 | 2.9 | 24 | 9.45 | 2.62 | 21 | 5.8% | -0.19 [-0.78, 0.39] |
| Fonagy 2016 | 0 | 0 | 0 | 0 | 0 | 0 | | Not estimable |
| Gaden 2022 | 0 | 0 | 0 | 0 | 0 | 0 | | Not estimable |
| Georg 2021 | 0 | 0 | 0 | 0 | 0 | 0 | | Not estimable |
| Glazebrook 2007 | 0 | 0 | 0 | 0 | 0 | 0 | | Not estimable |
| Hane 2015 | 0 | 0 | 0 | 0 | 0 | 0 | | Not estimable |
| Heo 2019 | 0 | 0 | 0 | 0 | 0 | 0 | | Not estimable |
| Herbers 2020 | 0 | 0 | 0 | 0 | 0 | 0 | | Not estimable |
| Hoffenkamp 2015 | 0 | 0 | 0 | 0 | 0 | 0 | | Not estimable |
| Hoivik 2015 | 0 | 0 | 0 | 0 | 0 | 0 | | Not estimable |
| Holditch-Davis 2014 | 0 | 0 | 0 | 0 | 0 | 0 | | Not estimable |
| Holt 2021 | 0 | 0 | 0 | 0 | 0 | 0 | | Not estimable |
| Horowitz 2001 | 0 | 0 | 0 | 0 | 0 | 0 | | Not estimable |
| Horowitz 2013 | 0 | 0 | 0 | 0 | 0 | 0 | | Not estimable |
| Juffer 2005 | 0 | 0 | 0 | 0 | 0 | 0 | | Not estimable |
| Klein Velderman 2006 | 0 | 0 | 0 | 0 | 0 | 0 | | Not estimable |
| Knoche 2012 | 0 | 0 | 0 | 0 | 0 | 0 | | Not estimable |
| Kristensen 2020 | 0 | 0 | 0 | 0 | 0 | 0 | | Not estimable |
| Landry 2006 | 0 | 0 | 0 | 0 | 0 | 0 | | Not estimable |
| Lavallee 2022 | 0 | 0 | 0 | 0 | 0 | 0 | | Not estimable |
| Magil-Evans 2007 | 0 | 0 | 0 | 0 | 0 | 0 | | Not estimable |
| Meijssen 2010 | 0 | 0 | 0 | 0 | 0 | 0 | | Not estimable |
| Melnyk 2006 | 0 | 0 | 0 | 0 | 0 | 0 | | Not estimable |
| Mendelsohn 2007 | 0 | 0 | 0 | 0 | 0 | 0 | | Not estimable |
| Milgrom 2013 | 19.7 | 6.8 | 46 | 17.1 | 5.5 | 45 | 11.3% | 0.42 [0.00, 0.83] |
| Nelson 2001 | 0 | 0 | 0 | 0 | 0 | 0 | | Not estimable |
| Neu 2010 | 0 | 0 | 0 | 0 | 0 | 0 | | Not estimable |
| Newnham 2009 | 0 | 0 | 0 | 0 | 0 | 0 | | Not estimable |
| Nugent 2017 | 0 | 0 | 0 | 0 | 0 | 0 | | Not estimable |
| Onozawa 2001 | 0 | 0 | 0 | 0 | 0 | 0 | | Not estimable |
| Oxford 2021 | 0 | 0 | 0 | 0 | 0 | 0 | | Not estimable |
| Perrone 2021 | 0 | 0 | 0 | 0 | 0 | 0 | | Not estimable |
| Pontoppidan 2016 | 0 | 0 | 0 | 0 | 0 | 0 | | Not estimable |
| Porter 2015 | 0 | 0 | 0 | 0 | 0 | 0 | | Not estimable |
| Ramsauer 2019 | 0 | 0 | 0 | 0 | 0 | 0 | | Not estimable |
| Ravn 2011 | 0 | 0 | 0 | 0 | 0 | 0 | | Not estimable |
| Rheinheimer 2022 | 0 | 0 | 0 | 0 | 0 | 0 | | Not estimable |
| Robertson 2019 | 0 | 0 | 0 | 0 | 0 | 0 | | Not estimable |
| Roby 2021 | 0 | 0 | 0 | 0 | 0 | 0 | | Not estimable |
| Sadler 2013 | 0 | 0 | 0 | 0 | 0 | 0 | | Not estimable |
| Sahlen Helmer 2020 | 0 | 0 | 0 | 0 | 0 | 0 | | Not estimable |
| Sajaniemi 2001 | 0 | 0 | 0 | 0 | 0 | 0 | | Not estimable |
| Salo 2019 | 0 | 0 | 0 | 0 | 0 | 0 | | Not estimable |
| Santelices 2010 | 0 | 0 | 0 | 0 | 0 | 0 | | Not estimable |
| Schuler 2000 | 0 | 0 | 0 | 0 | 0 | 0 | | Not estimable |
| Shoghi 2018 | 0 | 0 | 0 | 0 | 0 | 0 | | Not estimable |
| Sleed 2013 | 0 | 0 | 0 | 0 | 0 | 0 | | Not estimable |
| Steele 2019 | 0 | 0 | 0 | 0 | 0 | 0 | | Not estimable |
| Stein 2006 | 0 | 0 | 0 | 0 | 0 | 0 | | Not estimable |
| Stein 2018 | 0 | 0 | 0 | 0 | 0 | 0 | | Not estimable |
| Taneja 2020 | 88.3 | 9.9 | 276 | 88.51 | 10.8 | 276 | 55.0% | -0.02 [-0.19, 0.15] |
| Tereno 2017 | 0 | 0 | 0 | 0 | 0 | 0 | | Not estimable |
| Teti 2009 | 0 | 0 | 0 | 0 | 0 | 0 | | Not estimable |
| Tryphonopoulos 2020 | 0 | 0 | 0 | 0 | 0 | 0 | | Not estimable |
| Tsivos 2015 | 0 | 0 | 0 | 0 | 0 | 0 | | Not estimable |
| Twohig 2021 | 0 | 0 | 0 | 0 | 0 | 0 | | Not estimable |
| Valades 2021 | 0 | 0 | 0 | 0 | 0 | 0 | | Not estimable |
| van Doesum 2008 | 0 | 0 | 0 | 0 | 0 | 0 | | Not estimable |
| Vargas-Porras 2021 | 0 | 0 | 0 | 0 | 0 | 0 | | Not estimable |
| Walter 2019 | 0 | 0 | 0 | 0 | 0 | 0 | | Not estimable |
| White-Traut 2013 | 0 | 0 | 0 | 0 | 0 | 0 | | Not estimable |
| Williams 2020 | 0 | 0 | 0 | 0 | 0 | 0 | | Not estimable |
| Wulff 2021 | 0 | 0 | 0 | 0 | 0 | 0 | | Not estimable |
| Youn 2021 | 92.8 | 8.3 | 66 | 92.7 | 10.1 | 61 | 15.8% | 0.01 [-0.34, 0.36] |
| Yu 2022 | 0 | 0 | 0 | 0 | 0 | 0 | | Not estimable |
| Zelkowitz 2011 | 93.6 | 10.7 | 46 | 94.1 | 11 | 50 | 12.1% | -0.05 [-0.45, 0.35] |
| **Total (95% CI)** | | | **458** | | | **453** | **100.0%** | **0.02 [-0.12, 0.16]** |

Heterogeneity: Tau² = 0.00; Chi² = 4.33, df = 4 (P = 0.36); I² = 8%
Test for overall effect: Z = 0.28 (P = 0.78)

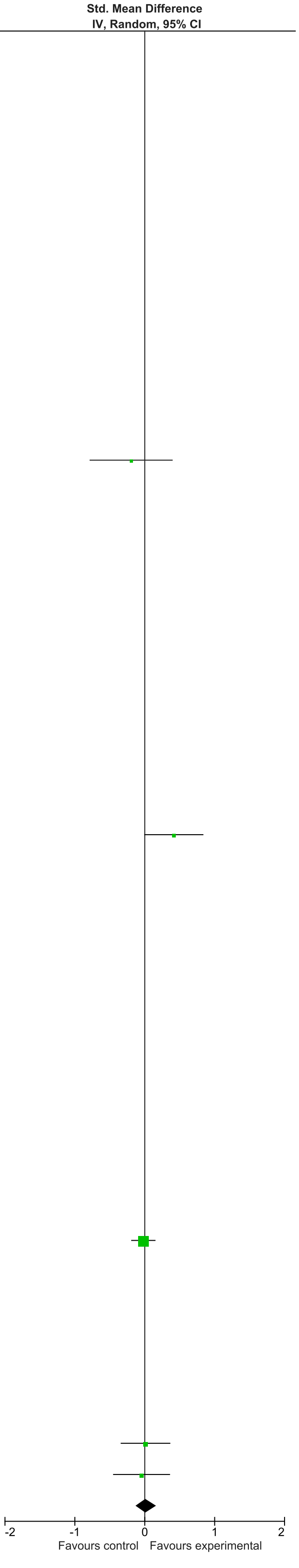

### eFigure 27. Language Development 7-12-month Follow-Up Forrest Plot

| | Early Dyadic Intervention | | | Standard Care | | | | Std. Mean Difference |
|---|---|---|---|---|---|---|---|---|
| Study or Subgroup | Mean | SD | Total | Mean | SD | Total | Weight | IV, Random, 95% CI |
| Akai 2008 | 0 | 0 | 0 | 0 | 0 | 0 | | Not estimable |
| Alarcao 2021 | 0 | 0 | 0 | 0 | 0 | 0 | | Not estimable |
| Alvarenga 2019 | 0 | 0 | 0 | 0 | 0 | 0 | | Not estimable |
| Badr 2006 | 0 | 0 | 0 | 0 | 0 | 0 | | Not estimable |
| Baggett 2010 | 0 | 0 | 0 | 0 | 0 | 0 | | Not estimable |
| Baggett 2017 | 0 | 0 | 0 | 0 | 0 | 0 | | Not estimable |
| Barlow 2016 | 0 | 0 | 0 | 0 | 0 | 0 | | Not estimable |
| Barnes 2022 | 0 | 0 | 0 | 0 | 0 | 0 | | Not estimable |
| Berlin 2017 | 0 | 0 | 0 | 0 | 0 | 0 | | Not estimable |
| Berlin 2018 | 0 | 0 | 0 | 0 | 0 | 0 | | Not estimable |
| Bernard 2012 | 0 | 0 | 0 | 0 | 0 | 0 | | Not estimable |
| Betancourt 2020 | 0 | 0 | 0 | 0 | 0 | 0 | | Not estimable |
| Bick 2013 | 0 | 0 | 0 | 0 | 0 | 0 | | Not estimable |
| Borghini 2014 | 0 | 0 | 0 | 0 | 0 | 0 | | Not estimable |
| Brisch 2003 | 0 | 0 | 0 | 0 | 0 | 0 | | Not estimable |
| Browne 2005 | 0 | 0 | 0 | 0 | 0 | 0 | | Not estimable |
| Cates 2018 | 0 | 0 | 0 | 0 | 0 | 0 | | Not estimable |
| Cevasco 2008 | 0 | 0 | 0 | 0 | 0 | 0 | | Not estimable |
| Chiu 2009 | 0 | 0 | 0 | 0 | 0 | 0 | | Not estimable |
| Constantino 2001 | 0 | 0 | 0 | 0 | 0 | 0 | | Not estimable |
| Cooper 2009 | 0 | 0 | 0 | 0 | 0 | 0 | | Not estimable |
| Cooper 2015 | 0 | 0 | 0 | 0 | 0 | 0 | | Not estimable |
| Corrigan 2021 | 0 | 0 | 0 | 0 | 0 | 0 | | Not estimable |
| Dozier 2009 | 0 | 0 | 0 | 0 | 0 | 0 | | Not estimable |
| Eriksen 2018 | 0 | 0 | 0 | 0 | 0 | 0 | | Not estimable |
| Feil 2020 | 0 | 0 | 0 | 0 | 0 | 0 | | Not estimable |
| Feldman 2014 | 0 | 0 | 0 | 0 | 0 | 0 | | Not estimable |
| Firk 2021 | 0 | 0 | 0 | 0 | 0 | 0 | | Not estimable |
| Fonagy 2016 | 0 | 0 | 0 | 0 | 0 | 0 | | Not estimable |
| Gaden 2022 | 0 | 0 | 0 | 0 | 0 | 0 | | Not estimable |
| Georg 2021 | 0 | 0 | 0 | 0 | 0 | 0 | | Not estimable |
| Glazebrook 2007 | 0 | 0 | 0 | 0 | 0 | 0 | | Not estimable |
| Hane 2015 | 0 | 0 | 0 | 0 | 0 | 0 | | Not estimable |
| Heo 2019 | 0 | 0 | 0 | 0 | 0 | 0 | | Not estimable |
| Herbers 2020 | 0 | 0 | 0 | 0 | 0 | 0 | | Not estimable |
| Hoffenkamp 2015 | 0 | 0 | 0 | 0 | 0 | 0 | | Not estimable |
| Hoivik 2015 | 0 | 0 | 0 | 0 | 0 | 0 | | Not estimable |
| Holditch-Davis 2014 | 0 | 0 | 0 | 0 | 0 | 0 | | Not estimable |
| Holt 2021 | 0 | 0 | 0 | 0 | 0 | 0 | | Not estimable |
| Horowitz 2001 | 0 | 0 | 0 | 0 | 0 | 0 | | Not estimable |
| Horowitz 2013 | 0 | 0 | 0 | 0 | 0 | 0 | | Not estimable |
| Juffer 2005 | 0 | 0 | 0 | 0 | 0 | 0 | | Not estimable |
| Klein Velderman 2006 | 0 | 0 | 0 | 0 | 0 | 0 | | Not estimable |
| Knoche 2012 | 0 | 0 | 0 | 0 | 0 | 0 | | Not estimable |
| Kristensen 2020 | 0 | 0 | 0 | 0 | 0 | 0 | | Not estimable |
| Landry 2006 | 0 | 0 | 0 | 0 | 0 | 0 | | Not estimable |
| Lavallee 2022 | 0 | 0 | 0 | 0 | 0 | 0 | | Not estimable |
| Magil-Evans 2007 | 0 | 0 | 0 | 0 | 0 | 0 | | Not estimable |
| Meijssen 2010 | 0 | 0 | 0 | 0 | 0 | 0 | | Not estimable |
| Melnyk 2006 | 0 | 0 | 0 | 0 | 0 | 0 | | Not estimable |
| Mendelsohn 2007 | 0 | 0 | 0 | 0 | 0 | 0 | | Not estimable |
| Milgrom 2013 | 0 | 0 | 0 | 0 | 0 | 0 | | Not estimable |
| Nelson 2001 | 0 | 0 | 0 | 0 | 0 | 0 | | Not estimable |
| Neu 2010 | 0 | 0 | 0 | 0 | 0 | 0 | | Not estimable |
| Newnham 2009 | 0 | 0 | 0 | 0 | 0 | 0 | | Not estimable |
| Nugent 2017 | 0 | 0 | 0 | 0 | 0 | 0 | | Not estimable |
| Onozawa 2001 | 0 | 0 | 0 | 0 | 0 | 0 | | Not estimable |
| Oxford 2021 | 0 | 0 | 0 | 0 | 0 | 0 | | Not estimable |
| Perrone 2021 | 0 | 0 | 0 | 0 | 0 | 0 | | Not estimable |
| Pontoppidan 2016 | 0 | 0 | 0 | 0 | 0 | 0 | | Not estimable |
| Porter 2015 | 0 | 0 | 0 | 0 | 0 | 0 | | Not estimable |
| Ramsauer 2019 | 0 | 0 | 0 | 0 | 0 | 0 | | Not estimable |
| Ravn 2011 | 0 | 0 | 0 | 0 | 0 | 0 | | Not estimable |
| Rheinheimer 2022 | 0 | 0 | 0 | 0 | 0 | 0 | | Not estimable |
| Robertson 2019 | 0 | 0 | 0 | 0 | 0 | 0 | | Not estimable |
| Roby 2021 | 0 | 0 | 0 | 0 | 0 | 0 | | Not estimable |
| Sadler 2013 | 0 | 0 | 0 | 0 | 0 | 0 | | Not estimable |
| Sahlen Helmer 2020 | 0 | 0 | 0 | 0 | 0 | 0 | | Not estimable |
| Sajaniemi 2001 | 0 | 0 | 0 | 0 | 0 | 0 | | Not estimable |
| Salo 2019 | 0 | 0 | 0 | 0 | 0 | 0 | | Not estimable |
| Santelices 2010 | 0 | 0 | 0 | 0 | 0 | 0 | | Not estimable |
| Schuler 2000 | 0 | 0 | 0 | 0 | 0 | 0 | | Not estimable |
| Shoghi 2018 | 0 | 0 | 0 | 0 | 0 | 0 | | Not estimable |
| Sleed 2013 | 0 | 0 | 0 | 0 | 0 | 0 | | Not estimable |
| Steele 2019 | 0 | 0 | 0 | 0 | 0 | 0 | | Not estimable |
| Stein 2006 | 0 | 0 | 0 | 0 | 0 | 0 | | Not estimable |
| Stein 2018 | 103.5 | 17.2 | 62 | 102 | 16.3 | 67 | 20.0% | 0.09 [-0.26, 0.43] |
| Taneja 2020 | 84.48 | 9.1 | 254 | 85.39 | 8.9 | 262 | 80.0% | -0.10 [-0.27, 0.07] |
| Tereno 2017 | 0 | 0 | 0 | 0 | 0 | 0 | | Not estimable |
| Teti 2009 | 0 | 0 | 0 | 0 | 0 | 0 | | Not estimable |
| Tryphonopoulos 2020 | 0 | 0 | 0 | 0 | 0 | 0 | | Not estimable |
| Tsivos 2015 | 0 | 0 | 0 | 0 | 0 | 0 | | Not estimable |
| Twohig 2021 | 0 | 0 | 0 | 0 | 0 | 0 | | Not estimable |
| Valades 2021 | 0 | 0 | 0 | 0 | 0 | 0 | | Not estimable |
| van Doesum 2008 | 0 | 0 | 0 | 0 | 0 | 0 | | Not estimable |
| Vargas-Porras 2021 | 0 | 0 | 0 | 0 | 0 | 0 | | Not estimable |
| Walter 2019 | 0 | 0 | 0 | 0 | 0 | 0 | | Not estimable |
| White-Traut 2013 | 0 | 0 | 0 | 0 | 0 | 0 | | Not estimable |
| Williams 2020 | 0 | 0 | 0 | 0 | 0 | 0 | | Not estimable |
| Wulff 2021 | 0 | 0 | 0 | 0 | 0 | 0 | | Not estimable |
| Youn 2021 | 0 | 0 | 0 | 0 | 0 | 0 | | Not estimable |
| Yu 2022 | 0 | 0 | 0 | 0 | 0 | 0 | | Not estimable |
| Zelkowitz 2011 | 0 | 0 | 0 | 0 | 0 | 0 | | Not estimable |
| **Total (95% CI)** | | | **316** | | | **329** | **100.0%** | **-0.06 [-0.22, 0.09]** |

Heterogeneity: Tau² = 0.00; Chi² = 0.93, df = 1 (P = 0.33); I² = 0%

Test for overall effect: Z = 0.80 (P = 0.42)

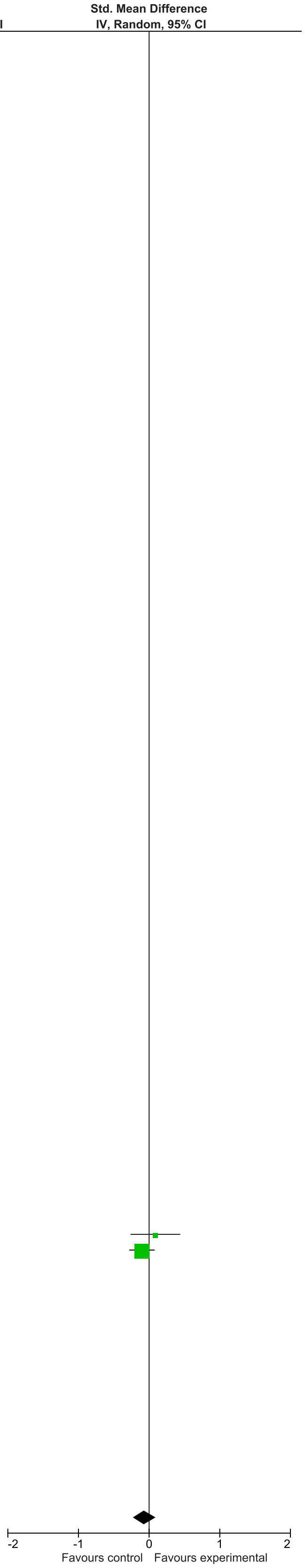

### eFigure 28. Language Development 13-24-month Follow-Up Forrest Plot

| Study or Subgroup | Early Dyadic Intervention Mean | SD | Total | Standard Care Mean | SD | Total | Weight | Std. Mean Difference IV, Random, 95% CI |
|---|---|---|---|---|---|---|---|---|
| Akai 2008 | 0 | 0 | 0 | 0 | 0 | 0 | | Not estimable |
| Alarcao 2021 | 0 | 0 | 0 | 0 | 0 | 0 | | Not estimable |
| Alvarenga 2019 | 0 | 0 | 0 | 0 | 0 | 0 | | Not estimable |
| Badr 2006 | 0 | 0 | 0 | 0 | 0 | 0 | | Not estimable |
| Baggett 2010 | 0 | 0 | 0 | 0 | 0 | 0 | | Not estimable |
| Baggett 2017 | 0 | 0 | 0 | 0 | 0 | 0 | | Not estimable |
| Barlow 2016 | 0 | 0 | 0 | 0 | 0 | 0 | | Not estimable |
| Barnes 2022 | 0 | 0 | 0 | 0 | 0 | 0 | | Not estimable |
| Berlin 2017 | 0 | 0 | 0 | 0 | 0 | 0 | | Not estimable |
| Berlin 2018 | 0 | 0 | 0 | 0 | 0 | 0 | | Not estimable |
| Bernard 2012 | 0 | 0 | 0 | 0 | 0 | 0 | | Not estimable |
| Betancourt 2020 | 0 | 0 | 0 | 0 | 0 | 0 | | Not estimable |
| Bick 2013 | 0 | 0 | 0 | 0 | 0 | 0 | | Not estimable |
| Borghini 2014 | 0 | 0 | 0 | 0 | 0 | 0 | | Not estimable |
| Brisch 2003 | 0 | 0 | 0 | 0 | 0 | 0 | | Not estimable |
| Browne 2005 | 0 | 0 | 0 | 0 | 0 | 0 | | Not estimable |
| Cates 2018 | 0 | 0 | 0 | 0 | 0 | 0 | | Not estimable |
| Cevasco 2008 | 0 | 0 | 0 | 0 | 0 | 0 | | Not estimable |
| Chiu 2009 | 0 | 0 | 0 | 0 | 0 | 0 | | Not estimable |
| Constantino 2001 | 0 | 0 | 0 | 0 | 0 | 0 | | Not estimable |
| Cooper 2009 | 0 | 0 | 0 | 0 | 0 | 0 | | Not estimable |
| Cooper 2015 | 0 | 0 | 0 | 0 | 0 | 0 | | Not estimable |
| Corrigan 2021 | 0 | 0 | 0 | 0 | 0 | 0 | | Not estimable |
| Dozier 2009 | 0 | 0 | 0 | 0 | 0 | 0 | | Not estimable |
| Eriksen 2018 | 0 | 0 | 0 | 0 | 0 | 0 | | Not estimable |
| Feil 2020 | 0 | 0 | 0 | 0 | 0 | 0 | | Not estimable |
| Feldman 2014 | 0 | 0 | 0 | 0 | 0 | 0 | | Not estimable |
| Firk 2021 | 0 | 0 | 0 | 0 | 0 | 0 | | Not estimable |
| Fonagy 2016 | 0 | 0 | 0 | 0 | 0 | 0 | | Not estimable |
| Gaden 2022 | 0 | 0 | 0 | 0 | 0 | 0 | | Not estimable |
| Georg 2021 | 0 | 0 | 0 | 0 | 0 | 0 | | Not estimable |
| Glazebrook 2007 | 0 | 0 | 0 | 0 | 0 | 0 | | Not estimable |
| Hane 2015 | 88.45 | 16.85 | 44 | 85.07 | 14.92 | 31 | 28.8% | 0.21 [-0.25, 0.67] |
| Heo 2019 | 0 | 0 | 0 | 0 | 0 | 0 | | Not estimable |
| Herbers 2020 | 0 | 0 | 0 | 0 | 0 | 0 | | Not estimable |
| Hoffenkamp 2015 | 0 | 0 | 0 | 0 | 0 | 0 | | Not estimable |
| Hoivik 2015 | 0 | 0 | 0 | 0 | 0 | 0 | | Not estimable |
| Holditch-Davis 2014 | 0 | 0 | 0 | 0 | 0 | 0 | | Not estimable |
| Holt 2021 | 0 | 0 | 0 | 0 | 0 | 0 | | Not estimable |
| Horowitz 2001 | 0 | 0 | 0 | 0 | 0 | 0 | | Not estimable |
| Horowitz 2013 | 0 | 0 | 0 | 0 | 0 | 0 | | Not estimable |
| Juffer 2005 | 0 | 0 | 0 | 0 | 0 | 0 | | Not estimable |
| Klein Velderman 2006 | 0 | 0 | 0 | 0 | 0 | 0 | | Not estimable |
| Knoche 2012 | 0 | 0 | 0 | 0 | 0 | 0 | | Not estimable |
| Kristensen 2020 | 0 | 0 | 0 | 0 | 0 | 0 | | Not estimable |
| Landry 2006 | 0 | 0 | 0 | 0 | 0 | 0 | | Not estimable |
| Lavallee 2022 | 0 | 0 | 0 | 0 | 0 | 0 | | Not estimable |
| Magil-Evans 2007 | 0 | 0 | 0 | 0 | 0 | 0 | | Not estimable |
| Meijssen 2010 | 0 | 0 | 0 | 0 | 0 | 0 | | Not estimable |
| Melnyk 2006 | 0 | 0 | 0 | 0 | 0 | 0 | | Not estimable |
| Mendelsohn 2007 | 0 | 0 | 0 | 0 | 0 | 0 | | Not estimable |
| Milgrom 2013 | 96.9 | 18.74 | 52 | 101.26 | 17.85 | 50 | 39.0% | -0.24 [-0.63, 0.15] |
| Nelson 2001 | 0 | 0 | 0 | 0 | 0 | 0 | | Not estimable |
| Neu 2010 | 0 | 0 | 0 | 0 | 0 | 0 | | Not estimable |
| Newnham 2009 | 0 | 0 | 0 | 0 | 0 | 0 | | Not estimable |
| Nugent 2017 | 0 | 0 | 0 | 0 | 0 | 0 | | Not estimable |
| Onozawa 2001 | 0 | 0 | 0 | 0 | 0 | 0 | | Not estimable |
| Oxford 2021 | 0 | 0 | 0 | 0 | 0 | 0 | | Not estimable |
| Perrone 2021 | 0 | 0 | 0 | 0 | 0 | 0 | | Not estimable |
| Pontoppidan 2016 | 0 | 0 | 0 | 0 | 0 | 0 | | Not estimable |
| Porter 2015 | 0 | 0 | 0 | 0 | 0 | 0 | | Not estimable |
| Ramsauer 2019 | 0 | 0 | 0 | 0 | 0 | 0 | | Not estimable |
| Ravn 2011 | 0 | 0 | 0 | 0 | 0 | 0 | | Not estimable |
| Rheinheimer 2022 | 0 | 0 | 0 | 0 | 0 | 0 | | Not estimable |
| Robertson 2019 | 0 | 0 | 0 | 0 | 0 | 0 | | Not estimable |
| Roby 2021 | 0 | 0 | 0 | 0 | 0 | 0 | | Not estimable |
| Sadler 2013 | 0 | 0 | 0 | 0 | 0 | 0 | | Not estimable |
| Sahlen Helmer 2020 | 0 | 0 | 0 | 0 | 0 | 0 | | Not estimable |
| Sajaniemi 2001 | 0 | 0 | 0 | 0 | 0 | 0 | | Not estimable |
| Salo 2019 | 0 | 0 | 0 | 0 | 0 | 0 | | Not estimable |
| Santelices 2010 | 0 | 0 | 0 | 0 | 0 | 0 | | Not estimable |
| Schuler 2000 | 0 | 0 | 0 | 0 | 0 | 0 | | Not estimable |
| Shoghi 2018 | 0 | 0 | 0 | 0 | 0 | 0 | | Not estimable |
| Sleed 2013 | 0 | 0 | 0 | 0 | 0 | 0 | | Not estimable |
| Steele 2019 | 0 | 0 | 0 | 0 | 0 | 0 | | Not estimable |
| Stein 2006 | 0 | 0 | 0 | 0 | 0 | 0 | | Not estimable |
| Stein 2018 | 0 | 0 | 0 | 0 | 0 | 0 | | Not estimable |
| Taneja 2020 | 0 | 0 | 0 | 0 | 0 | 0 | | Not estimable |
| Tereno 2017 | 0 | 0 | 0 | 0 | 0 | 0 | | Not estimable |
| Teti 2009 | 0 | 0 | 0 | 0 | 0 | 0 | | Not estimable |
| Tryphonopoulos 2020 | 0 | 0 | 0 | 0 | 0 | 0 | | Not estimable |
| Tsivos 2015 | 0 | 0 | 0 | 0 | 0 | 0 | | Not estimable |
| Twohig 2021 | 0 | 0 | 0 | 0 | 0 | 0 | | Not estimable |
| Valades 2021 | 0 | 0 | 0 | 0 | 0 | 0 | | Not estimable |
| van Doesum 2008 | 0 | 0 | 0 | 0 | 0 | 0 | | Not estimable |
| Vargas-Porras 2021 | 0 | 0 | 0 | 0 | 0 | 0 | | Not estimable |
| Walter 2019 | 0 | 0 | 0 | 0 | 0 | 0 | | Not estimable |
| White-Traut 2013 | 0 | 0 | 0 | 0 | 0 | 0 | | Not estimable |
| Williams 2020 | 0 | 0 | 0 | 0 | 0 | 0 | | Not estimable |
| Wulff 2021 | 0 | 0 | 0 | 0 | 0 | 0 | | Not estimable |
| Youn 2021 | 95.3 | 17.1 | 43 | 94.6 | 17.2 | 39 | 32.2% | 0.04 [-0.39, 0.47] |
| Yu 2022 | 0 | 0 | 0 | 0 | 0 | 0 | | Not estimable |
| Zelkowitz 2011 | 0 | 0 | 0 | 0 | 0 | 0 | | Not estimable |
| **Total (95% CI)** | | | **139** | | | **120** | **100.0%** | **-0.02 [-0.28, 0.24]** |

Heterogeneity: Tau² = 0.00; Chi² = 2.20, df = 2 (P = 0.33); I² = 9%
Test for overall effect: Z = 0.15 (P = 0.88)

Std. Mean Difference IV, Random, 95% CI

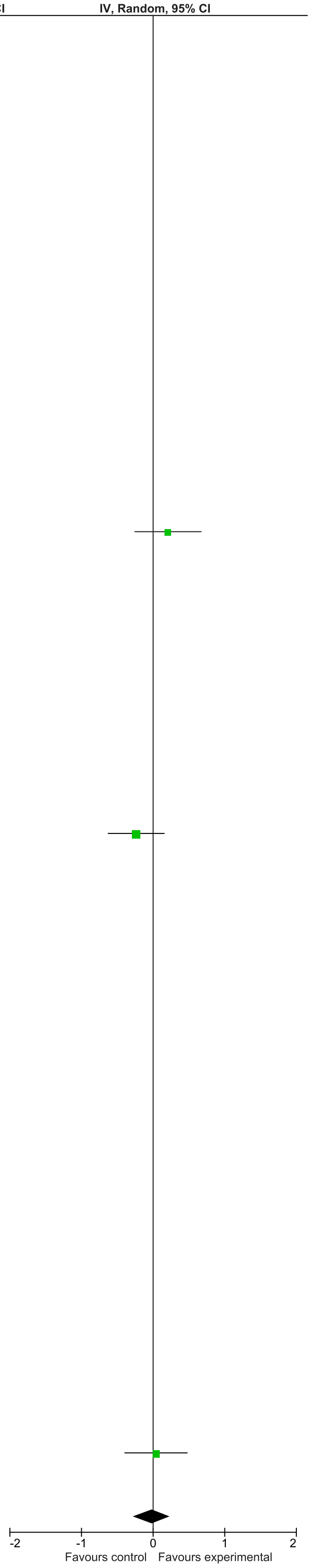

### eFigure 29. Montor Development 0-1-month Follow-Up Forrest Plot

| | Early Dyadic Intervention | | | Standard Care | | | | Std. Mean Difference |
|---|---|---|---|---|---|---|---|---|
| Study or Subgroup | Mean | SD | Total | Mean | SD | Total | Weight | IV, Random, 95% CI |
| Akai 2008 | 0 | 0 | 0 | 0 | 0 | 0 | | Not estimable |
| Alarcao 2021 | 0 | 0 | 0 | 0 | 0 | 0 | | Not estimable |
| Alvarenga 2019 | 0 | 0 | 0 | 0 | 0 | 0 | | Not estimable |
| Badr 2006 | 81.24 | 3.77 | 44 | 82.98 | 4.01 | 44 | 46.5% | -0.44 [-0.87, -0.02] |
| Baggett 2010 | 0 | 0 | 0 | 0 | 0 | 0 | | Not estimable |
| Baggett 2017 | 0 | 0 | 0 | 0 | 0 | 0 | | Not estimable |
| Barlow 2016 | 0 | 0 | 0 | 0 | 0 | 0 | | Not estimable |
| Barnes 2022 | 0 | 0 | 0 | 0 | 0 | 0 | | Not estimable |
| Berlin 2017 | 0 | 0 | 0 | 0 | 0 | 0 | | Not estimable |
| Berlin 2018 | 0 | 0 | 0 | 0 | 0 | 0 | | Not estimable |
| Bernard 2012 | 0 | 0 | 0 | 0 | 0 | 0 | | Not estimable |
| Betancourt 2020 | 0 | 0 | 0 | 0 | 0 | 0 | | Not estimable |
| Bick 2013 | 0 | 0 | 0 | 0 | 0 | 0 | | Not estimable |
| Borghini 2014 | 0 | 0 | 0 | 0 | 0 | 0 | | Not estimable |
| Brisch 2003 | 0 | 0 | 0 | 0 | 0 | 0 | | Not estimable |
| Browne 2005 | 0 | 0 | 0 | 0 | 0 | 0 | | Not estimable |
| Cates 2018 | 0 | 0 | 0 | 0 | 0 | 0 | | Not estimable |
| Cevasco 2008 | 0 | 0 | 0 | 0 | 0 | 0 | | Not estimable |
| Chiu 2009 | 0 | 0 | 0 | 0 | 0 | 0 | | Not estimable |
| Constantino 2001 | 0 | 0 | 0 | 0 | 0 | 0 | | Not estimable |
| Cooper 2009 | 0 | 0 | 0 | 0 | 0 | 0 | | Not estimable |
| Cooper 2015 | 0 | 0 | 0 | 0 | 0 | 0 | | Not estimable |
| Corrigan 2021 | 0 | 0 | 0 | 0 | 0 | 0 | | Not estimable |
| Dozier 2009 | 0 | 0 | 0 | 0 | 0 | 0 | | Not estimable |
| Eriksen 2018 | 0 | 0 | 0 | 0 | 0 | 0 | | Not estimable |
| Feil 2020 | 0 | 0 | 0 | 0 | 0 | 0 | | Not estimable |
| Feldman 2014 | 0 | 0 | 0 | 0 | 0 | 0 | | Not estimable |
| Firk 2021 | 10.02 | 2.51 | 27 | 10.23 | 1.97 | 20 | 24.9% | -0.09 [-0.67, 0.49] |
| Fonagy 2016 | 107 | 14.2 | 28 | 106.9 | 15.8 | 25 | 28.6% | 0.01 [-0.53, 0.55] |
| Gaden 2022 | 0 | 0 | 0 | 0 | 0 | 0 | | Not estimable |
| Georg 2021 | 0 | 0 | 0 | 0 | 0 | 0 | | Not estimable |
| Glazebrook 2007 | 0 | 0 | 0 | 0 | 0 | 0 | | Not estimable |
| Hane 2015 | 0 | 0 | 0 | 0 | 0 | 0 | | Not estimable |
| Heo 2019 | 0 | 0 | 0 | 0 | 0 | 0 | | Not estimable |
| Herbers 2020 | 0 | 0 | 0 | 0 | 0 | 0 | | Not estimable |
| Hoffenkamp 2015 | 0 | 0 | 0 | 0 | 0 | 0 | | Not estimable |
| Hoivik 2015 | 0 | 0 | 0 | 0 | 0 | 0 | | Not estimable |
| Holditch-Davis 2014 | 0 | 0 | 0 | 0 | 0 | 0 | | Not estimable |
| Holt 2021 | 0 | 0 | 0 | 0 | 0 | 0 | | Not estimable |
| Horowitz 2001 | 0 | 0 | 0 | 0 | 0 | 0 | | Not estimable |
| Horowitz 2013 | 0 | 0 | 0 | 0 | 0 | 0 | | Not estimable |
| Juffer 2005 | 0 | 0 | 0 | 0 | 0 | 0 | | Not estimable |
| Klein Velderman 2006 | 0 | 0 | 0 | 0 | 0 | 0 | | Not estimable |
| Knoche 2012 | 0 | 0 | 0 | 0 | 0 | 0 | | Not estimable |
| Kristensen 2020 | 0 | 0 | 0 | 0 | 0 | 0 | | Not estimable |
| Landry 2006 | 0 | 0 | 0 | 0 | 0 | 0 | | Not estimable |
| Lavallee 2022 | 0 | 0 | 0 | 0 | 0 | 0 | | Not estimable |
| Magil-Evans 2007 | 0 | 0 | 0 | 0 | 0 | 0 | | Not estimable |
| Meijssen 2010 | 0 | 0 | 0 | 0 | 0 | 0 | | Not estimable |
| Melnyk 2006 | 0 | 0 | 0 | 0 | 0 | 0 | | Not estimable |
| Mendelsohn 2007 | 0 | 0 | 0 | 0 | 0 | 0 | | Not estimable |
| Milgrom 2013 | 0 | 0 | 0 | 0 | 0 | 0 | | Not estimable |
| Nelson 2001 | 0 | 0 | 0 | 0 | 0 | 0 | | Not estimable |
| Neu 2010 | 0 | 0 | 0 | 0 | 0 | 0 | | Not estimable |
| Newnham 2009 | 0 | 0 | 0 | 0 | 0 | 0 | | Not estimable |
| Nugent 2017 | 0 | 0 | 0 | 0 | 0 | 0 | | Not estimable |
| Onozawa 2001 | 0 | 0 | 0 | 0 | 0 | 0 | | Not estimable |
| Oxford 2021 | 0 | 0 | 0 | 0 | 0 | 0 | | Not estimable |
| Perrone 2021 | 0 | 0 | 0 | 0 | 0 | 0 | | Not estimable |
| Pontoppidan 2016 | 0 | 0 | 0 | 0 | 0 | 0 | | Not estimable |
| Porter 2015 | 0 | 0 | 0 | 0 | 0 | 0 | | Not estimable |
| Ramsauer 2019 | 0 | 0 | 0 | 0 | 0 | 0 | | Not estimable |
| Ravn 2011 | 0 | 0 | 0 | 0 | 0 | 0 | | Not estimable |
| Rheinheimer 2022 | 0 | 0 | 0 | 0 | 0 | 0 | | Not estimable |
| Robertson 2019 | 0 | 0 | 0 | 0 | 0 | 0 | | Not estimable |
| Roby 2021 | 0 | 0 | 0 | 0 | 0 | 0 | | Not estimable |
| Sadler 2013 | 0 | 0 | 0 | 0 | 0 | 0 | | Not estimable |
| Sahlen Helmer 2020 | 0 | 0 | 0 | 0 | 0 | 0 | | Not estimable |
| Sajaniemi 2001 | 0 | 0 | 0 | 0 | 0 | 0 | | Not estimable |
| Salo 2019 | 0 | 0 | 0 | 0 | 0 | 0 | | Not estimable |
| Santelices 2010 | 0 | 0 | 0 | 0 | 0 | 0 | | Not estimable |
| Schuler 2000 | 0 | 0 | 0 | 0 | 0 | 0 | | Not estimable |
| Shoghi 2018 | 0 | 0 | 0 | 0 | 0 | 0 | | Not estimable |
| Sleed 2013 | 0 | 0 | 0 | 0 | 0 | 0 | | Not estimable |
| Steele 2019 | 0 | 0 | 0 | 0 | 0 | 0 | | Not estimable |
| Stein 2006 | 0 | 0 | 0 | 0 | 0 | 0 | | Not estimable |
| Stein 2018 | 0 | 0 | 0 | 0 | 0 | 0 | | Not estimable |
| Taneja 2020 | 0 | 0 | 0 | 0 | 0 | 0 | | Not estimable |
| Tereno 2017 | 0 | 0 | 0 | 0 | 0 | 0 | | Not estimable |
| Teti 2009 | 0 | 0 | 0 | 0 | 0 | 0 | | Not estimable |
| Tryphonopoulos 2020 | 0 | 0 | 0 | 0 | 0 | 0 | | Not estimable |
| Tsivos 2015 | 0 | 0 | 0 | 0 | 0 | 0 | | Not estimable |
| Twohig 2021 | 0 | 0 | 0 | 0 | 0 | 0 | | Not estimable |
| Valades 2021 | 0 | 0 | 0 | 0 | 0 | 0 | | Not estimable |
| van Doesum 2008 | 0 | 0 | 0 | 0 | 0 | 0 | | Not estimable |
| Vargas-Porras 2021 | 0 | 0 | 0 | 0 | 0 | 0 | | Not estimable |
| Walter 2019 | 0 | 0 | 0 | 0 | 0 | 0 | | Not estimable |
| White-Traut 2013 | 0 | 0 | 0 | 0 | 0 | 0 | | Not estimable |
| Williams 2020 | 0 | 0 | 0 | 0 | 0 | 0 | | Not estimable |
| Wulff 2021 | 0 | 0 | 0 | 0 | 0 | 0 | | Not estimable |
| Youn 2021 | 0 | 0 | 0 | 0 | 0 | 0 | | Not estimable |
| Yu 2022 | 0 | 0 | 0 | 0 | 0 | 0 | | Not estimable |
| Zelkowitz 2011 | 0 | 0 | 0 | 0 | 0 | 0 | | Not estimable |
| **Total (95% CI)** | | | **99** | | | **89** | **100.0%** | **-0.23 [-0.52, 0.06]** |

Heterogeneity: Tau² = 0.00; Chi² = 1.94, df = 2 (P = 0.38); I² = 0%

Test for overall effect: Z = 1.54 (P = 0.12)

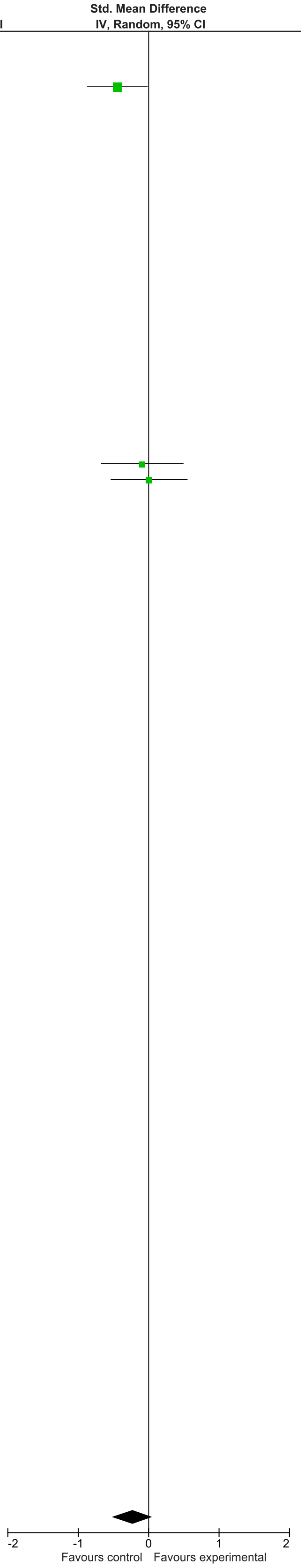

### eFigure 30. Motor Development 2-6-month Follow-Up Forrest Plot

| | Early Dyadic Intervention | | | Standard Care | | | | Std. Mean Difference |
|---|---|---|---|---|---|---|---|---|
| Study or Subgroup | Mean | SD | Total | Mean | SD | Total | Weight | IV, Random, 95% CI |
| Akai 2008 | 0 | 0 | 0 | 0 | 0 | 0 | | Not estimable |
| Alarcao 2021 | 0 | 0 | 0 | 0 | 0 | 0 | | Not estimable |
| Alvarenga 2019 | 0 | 0 | 0 | 0 | 0 | 0 | | Not estimable |
| Badr 2006 | 81.56 | 2.56 | 41 | 85.71 | 3.55 | 41 | 18.6% | -1.33 [-1.81, -0.85] |
| Baggett 2010 | 0 | 0 | 0 | 0 | 0 | 0 | | Not estimable |
| Baggett 2017 | 0 | 0 | 0 | 0 | 0 | 0 | | Not estimable |
| Barlow 2016 | 0 | 0 | 0 | 0 | 0 | 0 | | Not estimable |
| Barnes 2022 | 0 | 0 | 0 | 0 | 0 | 0 | | Not estimable |
| Berlin 2017 | 0 | 0 | 0 | 0 | 0 | 0 | | Not estimable |
| Berlin 2018 | 0 | 0 | 0 | 0 | 0 | 0 | | Not estimable |
| Bernard 2012 | 0 | 0 | 0 | 0 | 0 | 0 | | Not estimable |
| Betancourt 2020 | 0 | 0 | 0 | 0 | 0 | 0 | | Not estimable |
| Bick 2013 | 0 | 0 | 0 | 0 | 0 | 0 | | Not estimable |
| Borghini 2014 | 0 | 0 | 0 | 0 | 0 | 0 | | Not estimable |
| Brisch 2003 | 0 | 0 | 0 | 0 | 0 | 0 | | Not estimable |
| Browne 2005 | 0 | 0 | 0 | 0 | 0 | 0 | | Not estimable |
| Cates 2018 | 0 | 0 | 0 | 0 | 0 | 0 | | Not estimable |
| Cevasco 2008 | 0 | 0 | 0 | 0 | 0 | 0 | | Not estimable |
| Chiu 2009 | 0 | 0 | 0 | 0 | 0 | 0 | | Not estimable |
| Constantino 2001 | 0 | 0 | 0 | 0 | 0 | 0 | | Not estimable |
| Cooper 2009 | 0 | 0 | 0 | 0 | 0 | 0 | | Not estimable |
| Cooper 2015 | 0 | 0 | 0 | 0 | 0 | 0 | | Not estimable |
| Corrigan 2021 | 0 | 0 | 0 | 0 | 0 | 0 | | Not estimable |
| Dozier 2009 | 0 | 0 | 0 | 0 | 0 | 0 | | Not estimable |
| Eriksen 2018 | 0 | 0 | 0 | 0 | 0 | 0 | | Not estimable |
| Feil 2020 | 0 | 0 | 0 | 0 | 0 | 0 | | Not estimable |
| Feldman 2014 | 0 | 0 | 0 | 0 | 0 | 0 | | Not estimable |
| Firk 2021 | 9.84 | 2.36 | 24 | 9.86 | 2.42 | 21 | 16.7% | -0.01 [-0.59, 0.58] |
| Fonagy 2016 | 0 | 0 | 0 | 0 | 0 | 0 | | Not estimable |
| Gaden 2022 | 0 | 0 | 0 | 0 | 0 | 0 | | Not estimable |
| Georg 2021 | 0 | 0 | 0 | 0 | 0 | 0 | | Not estimable |
| Glazebrook 2007 | 0 | 0 | 0 | 0 | 0 | 0 | | Not estimable |
| Hane 2015 | 0 | 0 | 0 | 0 | 0 | 0 | | Not estimable |
| Heo 2019 | 0 | 0 | 0 | 0 | 0 | 0 | | Not estimable |
| Herbers 2020 | 0 | 0 | 0 | 0 | 0 | 0 | | Not estimable |
| Hoffenkamp 2015 | 0 | 0 | 0 | 0 | 0 | 0 | | Not estimable |
| Hoivik 2015 | 0 | 0 | 0 | 0 | 0 | 0 | | Not estimable |
| Holditch-Davis 2014 | 0 | 0 | 0 | 0 | 0 | 0 | | Not estimable |
| Holt 2021 | 0 | 0 | 0 | 0 | 0 | 0 | | Not estimable |
| Horowitz 2001 | 0 | 0 | 0 | 0 | 0 | 0 | | Not estimable |
| Horowitz 2013 | 0 | 0 | 0 | 0 | 0 | 0 | | Not estimable |
| Juffer 2005 | 0 | 0 | 0 | 0 | 0 | 0 | | Not estimable |
| Klein Velderman 2006 | 0 | 0 | 0 | 0 | 0 | 0 | | Not estimable |
| Knoche 2012 | 0 | 0 | 0 | 0 | 0 | 0 | | Not estimable |
| Kristensen 2020 | 0 | 0 | 0 | 0 | 0 | 0 | | Not estimable |
| Landry 2006 | 0 | 0 | 0 | 0 | 0 | 0 | | Not estimable |
| Lavallee 2022 | 0 | 0 | 0 | 0 | 0 | 0 | | Not estimable |
| Magil-Evans 2007 | 0 | 0 | 0 | 0 | 0 | 0 | | Not estimable |
| Meijssen 2010 | 0 | 0 | 0 | 0 | 0 | 0 | | Not estimable |
| Melnyk 2006 | 0 | 0 | 0 | 0 | 0 | 0 | | Not estimable |
| Mendelsohn 2007 | 0 | 0 | 0 | 0 | 0 | 0 | | Not estimable |
| Milgrom 2013 | 0 | 0 | 0 | 0 | 0 | 0 | | Not estimable |
| Nelson 2001 | 0 | 0 | 0 | 0 | 0 | 0 | | Not estimable |
| Neu 2010 | 0 | 0 | 0 | 0 | 0 | 0 | | Not estimable |
| Newnham 2009 | 0 | 0 | 0 | 0 | 0 | 0 | | Not estimable |
| Nugent 2017 | 0 | 0 | 0 | 0 | 0 | 0 | | Not estimable |
| Onozawa 2001 | 0 | 0 | 0 | 0 | 0 | 0 | | Not estimable |
| Oxford 2021 | 0 | 0 | 0 | 0 | 0 | 0 | | Not estimable |
| Perrone 2021 | 0 | 0 | 0 | 0 | 0 | 0 | | Not estimable |
| Pontoppidan 2016 | 0 | 0 | 0 | 0 | 0 | 0 | | Not estimable |
| Porter 2015 | 0 | 0 | 0 | 0 | 0 | 0 | | Not estimable |
| Ramsauer 2019 | 0 | 0 | 0 | 0 | 0 | 0 | | Not estimable |
| Ravn 2011 | 0 | 0 | 0 | 0 | 0 | 0 | | Not estimable |
| Rheinheimer 2022 | 0 | 0 | 0 | 0 | 0 | 0 | | Not estimable |
| Robertson 2019 | 0 | 0 | 0 | 0 | 0 | 0 | | Not estimable |
| Roby 2021 | 0 | 0 | 0 | 0 | 0 | 0 | | Not estimable |
| Sadler 2013 | 0 | 0 | 0 | 0 | 0 | 0 | | Not estimable |
| Sahlen Helmer 2020 | 0 | 0 | 0 | 0 | 0 | 0 | | Not estimable |
| Sajaniemi 2001 | 0 | 0 | 0 | 0 | 0 | 0 | | Not estimable |
| Salo 2019 | 0 | 0 | 0 | 0 | 0 | 0 | | Not estimable |
| Santelices 2010 | 0 | 0 | 0 | 0 | 0 | 0 | | Not estimable |
| Schuler 2000 | 0 | 0 | 0 | 0 | 0 | 0 | | Not estimable |
| Shoghi 2018 | 0 | 0 | 0 | 0 | 0 | 0 | | Not estimable |
| Sleed 2013 | 0 | 0 | 0 | 0 | 0 | 0 | | Not estimable |
| Steele 2019 | 0 | 0 | 0 | 0 | 0 | 0 | | Not estimable |
| Stein 2006 | 0 | 0 | 0 | 0 | 0 | 0 | | Not estimable |
| Stein 2018 | 0 | 0 | 0 | 0 | 0 | 0 | | Not estimable |
| Taneja 2020 | 96.74 | 16.2 | 276 | 95.91 | 15.6 | 276 | 23.5% | 0.05 [-0.11, 0.22] |
| Tereno 2017 | 0 | 0 | 0 | 0 | 0 | 0 | | Not estimable |
| Teti 2009 | 0 | 0 | 0 | 0 | 0 | 0 | | Not estimable |
| Tryphonopoulos 2020 | 0 | 0 | 0 | 0 | 0 | 0 | | Not estimable |
| Tsivos 2015 | 0 | 0 | 0 | 0 | 0 | 0 | | Not estimable |
| Twohig 2021 | 0 | 0 | 0 | 0 | 0 | 0 | | Not estimable |
| Valades 2021 | 0 | 0 | 0 | 0 | 0 | 0 | | Not estimable |
| van Doesum 2008 | 0 | 0 | 0 | 0 | 0 | 0 | | Not estimable |
| Vargas-Porras 2021 | 0 | 0 | 0 | 0 | 0 | 0 | | Not estimable |
| Walter 2019 | 0 | 0 | 0 | 0 | 0 | 0 | | Not estimable |
| White-Traut 2013 | 0 | 0 | 0 | 0 | 0 | 0 | | Not estimable |
| Williams 2020 | 0 | 0 | 0 | 0 | 0 | 0 | | Not estimable |
| Wulff 2021 | 0 | 0 | 0 | 0 | 0 | 0 | | Not estimable |
| Youn 2021 | 93.7 | 12.6 | 66 | 93.9 | 12.8 | 61 | 21.0% | -0.02 [-0.36, 0.33] |
| Yu 2022 | 0 | 0 | 0 | 0 | 0 | 0 | | Not estimable |
| Zelkowitz 2011 | 96 | 16 | 46 | 97.9 | 15.7 | 50 | 20.1% | -0.12 [-0.52, 0.28] |
| **Total (95% CI)** | | | **453** | | | **449** | **100.0%** | **-0.26 [-0.69, 0.16]** |

Heterogeneity: Tau² = 0.19; Chi² = 28.57, df = 4 (P < 0.00001); I² = 86%
Test for overall effect: Z = 1.21 (P = 0.23)

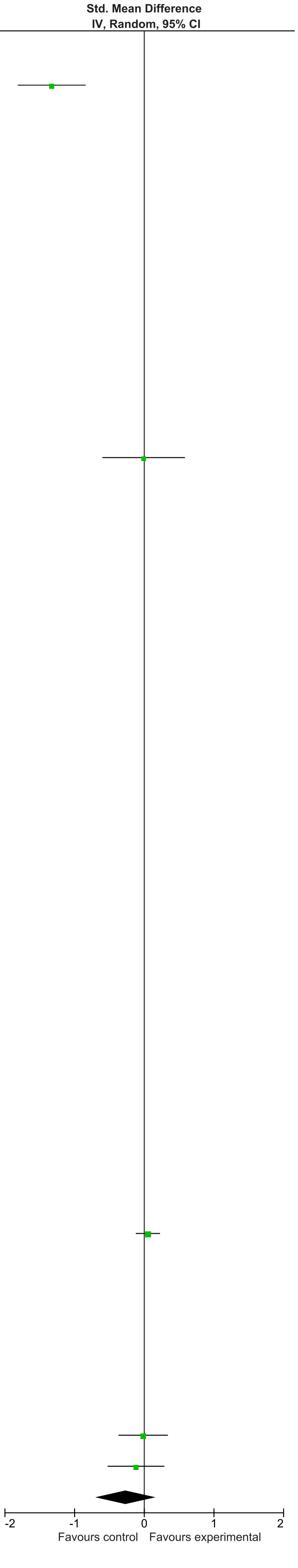

### eFigure 31. Motor Development 7-12-month Follow-Up Forrest Plot

| | Early Dyadic Intervention | | | Standard Care | | | | Std. Mean Difference |
|---|---|---|---|---|---|---|---|---|
| Study or Subgroup | Mean | SD | Total | Mean | SD | Total | Weight | IV, Random, 95% CI |
| Akai 2008 | 0 | 0 | 0 | 0 | 0 | 0 | | Not estimable |
| Alarcao 2021 | 0 | 0 | 0 | 0 | 0 | 0 | | Not estimable |
| Alvarenga 2019 | 0 | 0 | 0 | 0 | 0 | 0 | | Not estimable |
| Badr 2006 | 0 | 0 | 0 | 0 | 0 | 0 | | Not estimable |
| Baggett 2010 | 0 | 0 | 0 | 0 | 0 | 0 | | Not estimable |
| Baggett 2017 | 0 | 0 | 0 | 0 | 0 | 0 | | Not estimable |
| Barlow 2016 | 0 | 0 | 0 | 0 | 0 | 0 | | Not estimable |
| Barnes 2022 | 0 | 0 | 0 | 0 | 0 | 0 | | Not estimable |
| Berlin 2017 | 0 | 0 | 0 | 0 | 0 | 0 | | Not estimable |
| Berlin 2018 | 0 | 0 | 0 | 0 | 0 | 0 | | Not estimable |
| Bernard 2012 | 0 | 0 | 0 | 0 | 0 | 0 | | Not estimable |
| Betancourt 2020 | 0 | 0 | 0 | 0 | 0 | 0 | | Not estimable |
| Bick 2013 | 0 | 0 | 0 | 0 | 0 | 0 | | Not estimable |
| Borghini 2014 | 0 | 0 | 0 | 0 | 0 | 0 | | Not estimable |
| Brisch 2003 | 0 | 0 | 0 | 0 | 0 | 0 | | Not estimable |
| Browne 2005 | 0 | 0 | 0 | 0 | 0 | 0 | | Not estimable |
| Cates 2018 | 0 | 0 | 0 | 0 | 0 | 0 | | Not estimable |
| Cevasco 2008 | 0 | 0 | 0 | 0 | 0 | 0 | | Not estimable |
| Chiu 2009 | 0 | 0 | 0 | 0 | 0 | 0 | | Not estimable |
| Constantino 2001 | 0 | 0 | 0 | 0 | 0 | 0 | | Not estimable |
| Cooper 2009 | 0 | 0 | 0 | 0 | 0 | 0 | | Not estimable |
| Cooper 2015 | 0 | 0 | 0 | 0 | 0 | 0 | | Not estimable |
| Corrigan 2021 | 0 | 0 | 0 | 0 | 0 | 0 | | Not estimable |
| Dozier 2009 | 0 | 0 | 0 | 0 | 0 | 0 | | Not estimable |
| Eriksen 2018 | 0 | 0 | 0 | 0 | 0 | 0 | | Not estimable |
| Feil 2020 | 0 | 0 | 0 | 0 | 0 | 0 | | Not estimable |
| Feldman 2014 | 86.48 | 9.93 | 73 | 87.57 | 10.29 | 73 | 21.2% | -0.11 [-0.43, 0.22] |
| Firk 2021 | 0 | 0 | 0 | 0 | 0 | 0 | | Not estimable |
| Fonagy 2016 | 0 | 0 | 0 | 0 | 0 | 0 | | Not estimable |
| Gaden 2022 | 0 | 0 | 0 | 0 | 0 | 0 | | Not estimable |
| Georg 2021 | 0 | 0 | 0 | 0 | 0 | 0 | | Not estimable |
| Glazebrook 2007 | 0 | 0 | 0 | 0 | 0 | 0 | | Not estimable |
| Hane 2015 | 0 | 0 | 0 | 0 | 0 | 0 | | Not estimable |
| Heo 2019 | 0 | 0 | 0 | 0 | 0 | 0 | | Not estimable |
| Herbers 2020 | 0 | 0 | 0 | 0 | 0 | 0 | | Not estimable |
| Hoffenkamp 2015 | 0 | 0 | 0 | 0 | 0 | 0 | | Not estimable |
| Hoivik 2015 | 0 | 0 | 0 | 0 | 0 | 0 | | Not estimable |
| Holditch-Davis 2014 | 0 | 0 | 0 | 0 | 0 | 0 | | Not estimable |
| Holt 2021 | 0 | 0 | 0 | 0 | 0 | 0 | | Not estimable |
| Horowitz 2001 | 0 | 0 | 0 | 0 | 0 | 0 | | Not estimable |
| Horowitz 2013 | 0 | 0 | 0 | 0 | 0 | 0 | | Not estimable |
| Juffer 2005 | 0 | 0 | 0 | 0 | 0 | 0 | | Not estimable |
| Klein Velderman 2006 | 0 | 0 | 0 | 0 | 0 | 0 | | Not estimable |
| Knoche 2012 | 0 | 0 | 0 | 0 | 0 | 0 | | Not estimable |
| Kristensen 2020 | 0 | 0 | 0 | 0 | 0 | 0 | | Not estimable |
| Landry 2006 | 0 | 0 | 0 | 0 | 0 | 0 | | Not estimable |
| Lavallee 2022 | 0 | 0 | 0 | 0 | 0 | 0 | | Not estimable |
| Magil-Evans 2007 | 0 | 0 | 0 | 0 | 0 | 0 | | Not estimable |
| Meijssen 2010 | 0 | 0 | 0 | 0 | 0 | 0 | | Not estimable |
| Melnyk 2006 | 0 | 0 | 0 | 0 | 0 | 0 | | Not estimable |
| Mendelsohn 2007 | 0 | 0 | 0 | 0 | 0 | 0 | | Not estimable |
| Milgrom 2013 | 0 | 0 | 0 | 0 | 0 | 0 | | Not estimable |
| Nelson 2001 | 72.27 | 20.21 | 15 | 66.17 | 16.32 | 12 | 3.8% | 0.32 [-0.45, 1.08] |
| Neu 2010 | 0 | 0 | 0 | 0 | 0 | 0 | | Not estimable |
| Newnham 2009 | 0 | 0 | 0 | 0 | 0 | 0 | | Not estimable |
| Nugent 2017 | 0 | 0 | 0 | 0 | 0 | 0 | | Not estimable |
| Onozawa 2001 | 0 | 0 | 0 | 0 | 0 | 0 | | Not estimable |
| Oxford 2021 | 0 | 0 | 0 | 0 | 0 | 0 | | Not estimable |
| Perrone 2021 | 0 | 0 | 0 | 0 | 0 | 0 | | Not estimable |
| Pontoppidan 2016 | 0 | 0 | 0 | 0 | 0 | 0 | | Not estimable |
| Porter 2015 | 0 | 0 | 0 | 0 | 0 | 0 | | Not estimable |
| Ramsauer 2019 | 0 | 0 | 0 | 0 | 0 | 0 | | Not estimable |
| Ravn 2011 | 0 | 0 | 0 | 0 | 0 | 0 | | Not estimable |
| Rheinheimer 2022 | 0 | 0 | 0 | 0 | 0 | 0 | | Not estimable |
| Robertson 2019 | 0 | 0 | 0 | 0 | 0 | 0 | | Not estimable |
| Roby 2021 | 0 | 0 | 0 | 0 | 0 | 0 | | Not estimable |
| Sadler 2013 | 0 | 0 | 0 | 0 | 0 | 0 | | Not estimable |
| Sahlen Helmer 2020 | 0 | 0 | 0 | 0 | 0 | 0 | | Not estimable |
| Sajaniemi 2001 | 0 | 0 | 0 | 0 | 0 | 0 | | Not estimable |
| Salo 2019 | 0 | 0 | 0 | 0 | 0 | 0 | | Not estimable |
| Santelices 2010 | 0 | 0 | 0 | 0 | 0 | 0 | | Not estimable |
| Schuler 2000 | 0 | 0 | 0 | 0 | 0 | 0 | | Not estimable |
| Shoghi 2018 | 0 | 0 | 0 | 0 | 0 | 0 | | Not estimable |
| Sleed 2013 | 0 | 0 | 0 | 0 | 0 | 0 | | Not estimable |
| Steele 2019 | 0 | 0 | 0 | 0 | 0 | 0 | | Not estimable |
| Stein 2006 | 0 | 0 | 0 | 0 | 0 | 0 | | Not estimable |
| Stein 2018 | 0 | 0 | 0 | 0 | 0 | 0 | | Not estimable |
| Taneja 2020 | 89.79 | 10.2 | 254 | 90.64 | 16.32 | 262 | 75.0% | -0.06 [-0.23, 0.11] |
| Tereno 2017 | 0 | 0 | 0 | 0 | 0 | 0 | | Not estimable |
| Teti 2009 | 0 | 0 | 0 | 0 | 0 | 0 | | Not estimable |
| Tryphonopoulos 2020 | 0 | 0 | 0 | 0 | 0 | 0 | | Not estimable |
| Tsivos 2015 | 0 | 0 | 0 | 0 | 0 | 0 | | Not estimable |
| Twohig 2021 | 0 | 0 | 0 | 0 | 0 | 0 | | Not estimable |
| Valades 2021 | 0 | 0 | 0 | 0 | 0 | 0 | | Not estimable |
| van Doesum 2008 | 0 | 0 | 0 | 0 | 0 | 0 | | Not estimable |
| Vargas-Porras 2021 | 0 | 0 | 0 | 0 | 0 | 0 | | Not estimable |
| Walter 2019 | 0 | 0 | 0 | 0 | 0 | 0 | | Not estimable |
| White-Traut 2013 | 0 | 0 | 0 | 0 | 0 | 0 | | Not estimable |
| Williams 2020 | 0 | 0 | 0 | 0 | 0 | 0 | | Not estimable |
| Wulff 2021 | 0 | 0 | 0 | 0 | 0 | 0 | | Not estimable |
| Youn 2021 | 0 | 0 | 0 | 0 | 0 | 0 | | Not estimable |
| Yu 2022 | 0 | 0 | 0 | 0 | 0 | 0 | | Not estimable |
| Zelkowitz 2011 | 0 | 0 | 0 | 0 | 0 | 0 | | Not estimable |
| **Total (95% CI)** | | | **342** | | | **347** | **100.0%** | **-0.06 [-0.21, 0.09]** |

Heterogeneity: Tau² = 0.00; Chi² = 1.02, df = 2 (P = 0.60); I² = 0%

Test for overall effect: Z = 0.75 (P = 0.45)

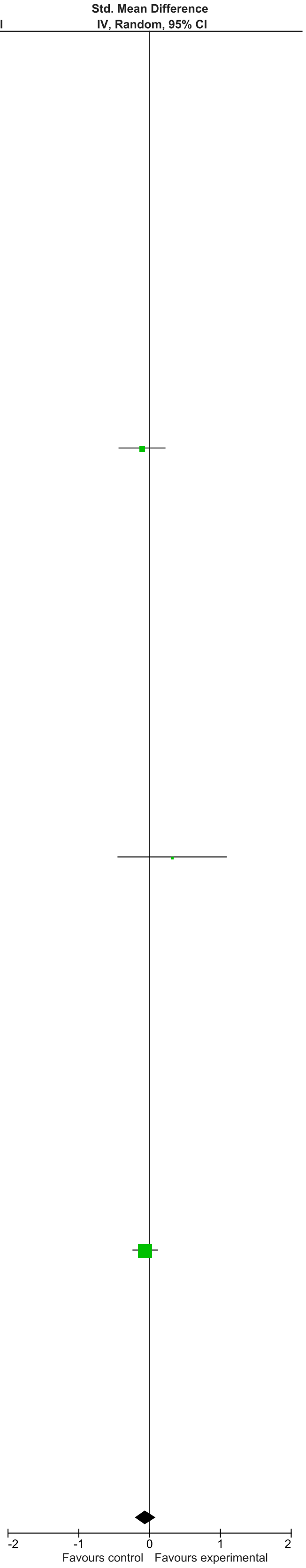

### eFigure 32. Motor Development 13-24-month Follow-Up Forrest Plot

| | Early Dyadic Intervention | | | Standard Care | | | | Std. Mean Difference |
|---|---|---|---|---|---|---|---|---|
| Study or Subgroup | Mean | SD | Total | Mean | SD | Total | Weight | IV, Random, 95% CI |
| Akai 2008 | 0 | 0 | 0 | 0 | 0 | 0 | | Not estimable |
| Alarcao 2021 | 0 | 0 | 0 | 0 | 0 | 0 | | Not estimable |
| Alvarenga 2019 | 0 | 0 | 0 | 0 | 0 | 0 | | Not estimable |
| Badr 2006 | 0 | 0 | 0 | 0 | 0 | 0 | | Not estimable |
| Baggett 2010 | 0 | 0 | 0 | 0 | 0 | 0 | | Not estimable |
| Baggett 2017 | 0 | 0 | 0 | 0 | 0 | 0 | | Not estimable |
| Barlow 2016 | 0 | 0 | 0 | 0 | 0 | 0 | | Not estimable |
| Barnes 2022 | 0 | 0 | 0 | 0 | 0 | 0 | | Not estimable |
| Berlin 2017 | 0 | 0 | 0 | 0 | 0 | 0 | | Not estimable |
| Berlin 2018 | 0 | 0 | 0 | 0 | 0 | 0 | | Not estimable |
| Bernard 2012 | 0 | 0 | 0 | 0 | 0 | 0 | | Not estimable |
| Betancourt 2020 | 0 | 0 | 0 | 0 | 0 | 0 | | Not estimable |
| Bick 2013 | 0 | 0 | 0 | 0 | 0 | 0 | | Not estimable |
| Borghini 2014 | 0 | 0 | 0 | 0 | 0 | 0 | | Not estimable |
| Brisch 2003 | 0 | 0 | 0 | 0 | 0 | 0 | | Not estimable |
| Browne 2005 | 0 | 0 | 0 | 0 | 0 | 0 | | Not estimable |
| Cates 2018 | 0 | 0 | 0 | 0 | 0 | 0 | | Not estimable |
| Cevasco 2008 | 0 | 0 | 0 | 0 | 0 | 0 | | Not estimable |
| Chiu 2009 | 0 | 0 | 0 | 0 | 0 | 0 | | Not estimable |
| Constantino 2001 | 0 | 0 | 0 | 0 | 0 | 0 | | Not estimable |
| Cooper 2009 | 0 | 0 | 0 | 0 | 0 | 0 | | Not estimable |
| Cooper 2015 | 0 | 0 | 0 | 0 | 0 | 0 | | Not estimable |
| Corrigan 2021 | 0 | 0 | 0 | 0 | 0 | 0 | | Not estimable |
| Dozier 2009 | 0 | 0 | 0 | 0 | 0 | 0 | | Not estimable |
| Eriksen 2018 | 0 | 0 | 0 | 0 | 0 | 0 | | Not estimable |
| Feil 2020 | 0 | 0 | 0 | 0 | 0 | 0 | | Not estimable |
| Feldman 2014 | 87.29 | 9.59 | 73 | 88.46 | 10.53 | 73 | 36.1% | -0.12 [-0.44, 0.21] |
| Firk 2021 | 0 | 0 | 0 | 0 | 0 | 0 | | Not estimable |
| Fonagy 2016 | 0 | 0 | 0 | 0 | 0 | 0 | | Not estimable |
| Gaden 2022 | 0 | 0 | 0 | 0 | 0 | 0 | | Not estimable |
| Georg 2021 | 0 | 0 | 0 | 0 | 0 | 0 | | Not estimable |
| Glazebrook 2007 | 0 | 0 | 0 | 0 | 0 | 0 | | Not estimable |
| Hane 2015 | 94.37 | 9.39 | 45 | 92.25 | 12.58 | 31 | 18.1% | 0.19 [-0.26, 0.65] |
| Heo 2019 | 0 | 0 | 0 | 0 | 0 | 0 | | Not estimable |
| Herbers 2020 | 0 | 0 | 0 | 0 | 0 | 0 | | Not estimable |
| Hoffenkamp 2015 | 0 | 0 | 0 | 0 | 0 | 0 | | Not estimable |
| Hoivik 2015 | 0 | 0 | 0 | 0 | 0 | 0 | | Not estimable |
| Holditch-Davis 2014 | 0 | 0 | 0 | 0 | 0 | 0 | | Not estimable |
| Holt 2021 | 0 | 0 | 0 | 0 | 0 | 0 | | Not estimable |
| Horowitz 2001 | 0 | 0 | 0 | 0 | 0 | 0 | | Not estimable |
| Horowitz 2013 | 0 | 0 | 0 | 0 | 0 | 0 | | Not estimable |
| Juffer 2005 | 0 | 0 | 0 | 0 | 0 | 0 | | Not estimable |
| Klein Velderman 2006 | 0 | 0 | 0 | 0 | 0 | 0 | | Not estimable |
| Knoche 2012 | 0 | 0 | 0 | 0 | 0 | 0 | | Not estimable |
| Kristensen 2020 | 0 | 0 | 0 | 0 | 0 | 0 | | Not estimable |
| Landry 2006 | 0 | 0 | 0 | 0 | 0 | 0 | | Not estimable |
| Lavallee 2022 | 0 | 0 | 0 | 0 | 0 | 0 | | Not estimable |
| Magil-Evans 2007 | 0 | 0 | 0 | 0 | 0 | 0 | | Not estimable |
| Meijssen 2010 | 0 | 0 | 0 | 0 | 0 | 0 | | Not estimable |
| Melnyk 2006 | 0 | 0 | 0 | 0 | 0 | 0 | | Not estimable |
| Mendelsohn 2007 | 0 | 0 | 0 | 0 | 0 | 0 | | Not estimable |
| Milgrom 2013 | 103.25 | 17.16 | 52 | 106.08 | 20.13 | 52 | 25.7% | -0.15 [-0.54, 0.23] |
| Nelson 2001 | 0 | 0 | 0 | 0 | 0 | 0 | | Not estimable |
| Neu 2010 | 0 | 0 | 0 | 0 | 0 | 0 | | Not estimable |
| Newnham 2009 | 0 | 0 | 0 | 0 | 0 | 0 | | Not estimable |
| Nugent 2017 | 0 | 0 | 0 | 0 | 0 | 0 | | Not estimable |
| Onozawa 2001 | 0 | 0 | 0 | 0 | 0 | 0 | | Not estimable |
| Oxford 2021 | 0 | 0 | 0 | 0 | 0 | 0 | | Not estimable |
| Perrone 2021 | 0 | 0 | 0 | 0 | 0 | 0 | | Not estimable |
| Pontoppidan 2016 | 0 | 0 | 0 | 0 | 0 | 0 | | Not estimable |
| Porter 2015 | 0 | 0 | 0 | 0 | 0 | 0 | | Not estimable |
| Ramsauer 2019 | 0 | 0 | 0 | 0 | 0 | 0 | | Not estimable |
| Ravn 2011 | 0 | 0 | 0 | 0 | 0 | 0 | | Not estimable |
| Rheinheimer 2022 | 0 | 0 | 0 | 0 | 0 | 0 | | Not estimable |
| Robertson 2019 | 0 | 0 | 0 | 0 | 0 | 0 | | Not estimable |
| Roby 2021 | 0 | 0 | 0 | 0 | 0 | 0 | | Not estimable |
| Sadler 2013 | 0 | 0 | 0 | 0 | 0 | 0 | | Not estimable |
| Sahlen Helmer 2020 | 0 | 0 | 0 | 0 | 0 | 0 | | Not estimable |
| Sajaniemi 2001 | 0 | 0 | 0 | 0 | 0 | 0 | | Not estimable |
| Salo 2019 | 0 | 0 | 0 | 0 | 0 | 0 | | Not estimable |
| Santelices 2010 | 0 | 0 | 0 | 0 | 0 | 0 | | Not estimable |
| Schuler 2000 | 0 | 0 | 0 | 0 | 0 | 0 | | Not estimable |
| Shoghi 2018 | 0 | 0 | 0 | 0 | 0 | 0 | | Not estimable |
| Sleed 2013 | 0 | 0 | 0 | 0 | 0 | 0 | | Not estimable |
| Steele 2019 | 0 | 0 | 0 | 0 | 0 | 0 | | Not estimable |
| Stein 2006 | 0 | 0 | 0 | 0 | 0 | 0 | | Not estimable |
| Stein 2018 | 0 | 0 | 0 | 0 | 0 | 0 | | Not estimable |
| Taneja 2020 | 0 | 0 | 0 | 0 | 0 | 0 | | Not estimable |
| Tereno 2017 | 0 | 0 | 0 | 0 | 0 | 0 | | Not estimable |
| Teti 2009 | 0 | 0 | 0 | 0 | 0 | 0 | | Not estimable |
| Tryphonopoulos 2020 | 0 | 0 | 0 | 0 | 0 | 0 | | Not estimable |
| Tsivos 2015 | 0 | 0 | 0 | 0 | 0 | 0 | | Not estimable |
| Twohig 2021 | 0 | 0 | 0 | 0 | 0 | 0 | | Not estimable |
| Valades 2021 | 0 | 0 | 0 | 0 | 0 | 0 | | Not estimable |
| van Doesum 2008 | 0 | 0 | 0 | 0 | 0 | 0 | | Not estimable |
| Vargas-Porras 2021 | 0 | 0 | 0 | 0 | 0 | 0 | | Not estimable |
| Walter 2019 | 0 | 0 | 0 | 0 | 0 | 0 | | Not estimable |
| White-Traut 2013 | 0 | 0 | 0 | 0 | 0 | 0 | | Not estimable |
| Williams 2020 | 0 | 0 | 0 | 0 | 0 | 0 | | Not estimable |
| Wulff 2021 | 0 | 0 | 0 | 0 | 0 | 0 | | Not estimable |
| Youn 2021 | 95 | 16.9 | 43 | 96.3 | 19.1 | 39 | 20.2% | -0.07 [-0.51, 0.36] |
| Yu 2022 | 0 | 0 | 0 | 0 | 0 | 0 | | Not estimable |
| Zelkowitz 2011 | 0 | 0 | 0 | 0 | 0 | 0 | | Not estimable |
| **Total (95% CI)** | | | **213** | | | **195** | **100.0%** | **-0.06 [-0.25, 0.14]** |

Heterogeneity: Tau² = 0.00; Chi² = 1.51, df = 3 (P = 0.68); I² = 0%
Test for overall effect: Z = 0.60 (P = 0.55)

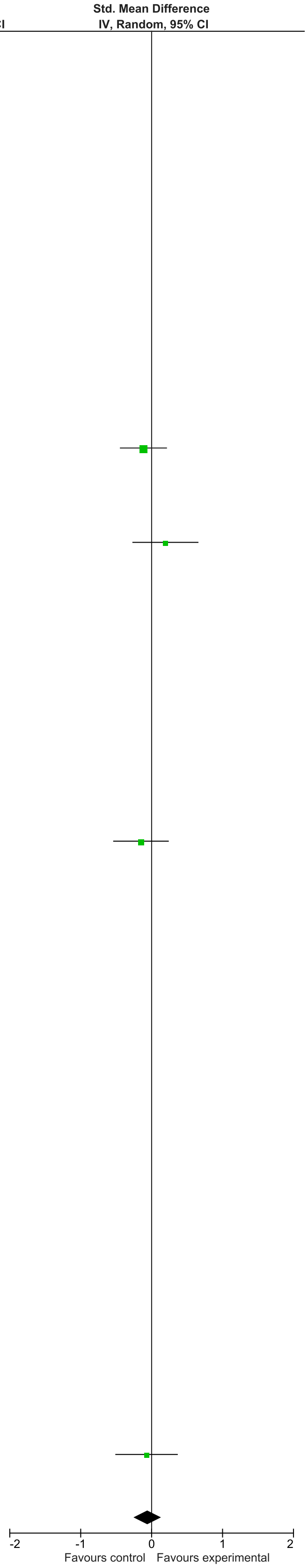

### eFigure 33. Socio-Emotional Functioning 2-6-month Follow-Up Forrest Plot

| | Early Dyadic Intervention | | | Standard Care | | | | Std. Mean Difference |
|---|---|---|---|---|---|---|---|---|
| Study or Subgroup | Mean | SD | Total | Mean | SD | Total | Weight | IV, Random, 95% CI |
| Akai 2008 | 0 | 0 | 0 | 0 | 0 | 0 | | Not estimable |
| Alarcao 2021 | 0 | 0 | 0 | 0 | 0 | 0 | | Not estimable |
| Alvarenga 2019 | 0 | 0 | 0 | 0 | 0 | 0 | | Not estimable |
| Badr 2006 | 0 | 0 | 0 | 0 | 0 | 0 | | Not estimable |
| Baggett 2010 | 0 | 0 | 0 | 0 | 0 | 0 | | Not estimable |
| Baggett 2017 | 0 | 0 | 0 | 0 | 0 | 0 | | Not estimable |
| Barlow 2016 | 0 | 0 | 0 | 0 | 0 | 0 | | Not estimable |
| Barnes 2022 | 0 | 0 | 0 | 0 | 0 | 0 | | Not estimable |
| Berlin 2017 | 0 | 0 | 0 | 0 | 0 | 0 | | Not estimable |
| Berlin 2018 | 0 | 0 | 0 | 0 | 0 | 0 | | Not estimable |
| Bernard 2012 | 0 | 0 | 0 | 0 | 0 | 0 | | Not estimable |
| Betancourt 2020 | 0 | 0 | 0 | 0 | 0 | 0 | | Not estimable |
| Bick 2013 | 0 | 0 | 0 | 0 | 0 | 0 | | Not estimable |
| Borghini 2014 | 0 | 0 | 0 | 0 | 0 | 0 | | Not estimable |
| Brisch 2003 | 0 | 0 | 0 | 0 | 0 | 0 | | Not estimable |
| Browne 2005 | 0 | 0 | 0 | 0 | 0 | 0 | | Not estimable |
| Cates 2018 | 0 | 0 | 0 | 0 | 0 | 0 | | Not estimable |
| Cevasco 2008 | 0 | 0 | 0 | 0 | 0 | 0 | | Not estimable |
| Chiu 2009 | 0 | 0 | 0 | 0 | 0 | 0 | | Not estimable |
| Constantino 2001 | 0 | 0 | 0 | 0 | 0 | 0 | | Not estimable |
| Cooper 2009 | 0 | 0 | 0 | 0 | 0 | 0 | | Not estimable |
| Cooper 2015 | 0 | 0 | 0 | 0 | 0 | 0 | | Not estimable |
| Corrigan 2021 | 0 | 0 | 0 | 0 | 0 | 0 | | Not estimable |
| Dozier 2009 | 0 | 0 | 0 | 0 | 0 | 0 | | Not estimable |
| Eriksen 2018 | 0 | 0 | 0 | 0 | 0 | 0 | | Not estimable |
| Feil 2020 | 0 | 0 | 0 | 0 | 0 | 0 | | Not estimable |
| Feldman 2014 | 0 | 0 | 0 | 0 | 0 | 0 | | Not estimable |
| Firk 2021 | 0 | 0 | 0 | 0 | 0 | 0 | | Not estimable |
| Fonagy 2016 | 0 | 0 | 0 | 0 | 0 | 0 | | Not estimable |
| Gaden 2022 | 0 | 0 | 0 | 0 | 0 | 0 | | Not estimable |
| Georg 2021 | 0 | 0 | 0 | 0 | 0 | 0 | | Not estimable |
| Glazebrook 2007 | 0 | 0 | 0 | 0 | 0 | 0 | | Not estimable |
| Hane 2015 | 0 | 0 | 0 | 0 | 0 | 0 | | Not estimable |
| Heo 2019 | 0 | 0 | 0 | 0 | 0 | 0 | | Not estimable |
| Herbers 2020 | 0 | 0 | 0 | 0 | 0 | 0 | | Not estimable |
| Hoffenkamp 2015 | 0 | 0 | 0 | 0 | 0 | 0 | | Not estimable |
| Hoivik 2015 | -20.44 | 13.45 | 26 | -25 | 16.53 | 27 | 18.6% | 0.30 [-0.24, 0.84] |
| Holditch-Davis 2014 | 0 | 0 | 0 | 0 | 0 | 0 | | Not estimable |
| Holt 2021 | 0.82 | 1.47 | 21 | 0.3 | 1.22 | 27 | 17.0% | 0.38 [-0.19, 0.96] |
| Horowitz 2001 | 0 | 0 | 0 | 0 | 0 | 0 | | Not estimable |
| Horowitz 2013 | 0 | 0 | 0 | 0 | 0 | 0 | | Not estimable |
| Juffer 2005 | 0 | 0 | 0 | 0 | 0 | 0 | | Not estimable |
| Klein Velderman 2006 | 0 | 0 | 0 | 0 | 0 | 0 | | Not estimable |
| Knoche 2012 | 0 | 0 | 0 | 0 | 0 | 0 | | Not estimable |
| Kristensen 2020 | 1.23 | 1.11 | 715 | 1.27 | 1.11 | 612 | 64.4% | -0.04 [-0.14, 0.07] |
| Landry 2006 | 0 | 0 | 0 | 0 | 0 | 0 | | Not estimable |
| Lavallee 2022 | 0 | 0 | 0 | 0 | 0 | 0 | | Not estimable |
| Magil-Evans 2007 | 0 | 0 | 0 | 0 | 0 | 0 | | Not estimable |
| Meijssen 2010 | 0 | 0 | 0 | 0 | 0 | 0 | | Not estimable |
| Melnyk 2006 | 0 | 0 | 0 | 0 | 0 | 0 | | Not estimable |
| Mendelsohn 2007 | 0 | 0 | 0 | 0 | 0 | 0 | | Not estimable |
| Milgrom 2013 | 0 | 0 | 0 | 0 | 0 | 0 | | Not estimable |
| Nelson 2001 | 0 | 0 | 0 | 0 | 0 | 0 | | Not estimable |
| Neu 2010 | 0 | 0 | 0 | 0 | 0 | 0 | | Not estimable |
| Newnham 2009 | 0 | 0 | 0 | 0 | 0 | 0 | | Not estimable |
| Nugent 2017 | 0 | 0 | 0 | 0 | 0 | 0 | | Not estimable |
| Onozawa 2001 | 0 | 0 | 0 | 0 | 0 | 0 | | Not estimable |
| Oxford 2021 | 0 | 0 | 0 | 0 | 0 | 0 | | Not estimable |
| Perrone 2021 | 0 | 0 | 0 | 0 | 0 | 0 | | Not estimable |
| Pontoppidan 2016 | 0 | 0 | 0 | 0 | 0 | 0 | | Not estimable |
| Porter 2015 | 0 | 0 | 0 | 0 | 0 | 0 | | Not estimable |
| Ramsauer 2019 | 0 | 0 | 0 | 0 | 0 | 0 | | Not estimable |
| Ravn 2011 | 0 | 0 | 0 | 0 | 0 | 0 | | Not estimable |
| Rheinheimer 2022 | 0 | 0 | 0 | 0 | 0 | 0 | | Not estimable |
| Robertson 2019 | 0 | 0 | 0 | 0 | 0 | 0 | | Not estimable |
| Roby 2021 | 0 | 0 | 0 | 0 | 0 | 0 | | Not estimable |
| Sadler 2013 | 0 | 0 | 0 | 0 | 0 | 0 | | Not estimable |
| Sahlen Helmer 2020 | 0 | 0 | 0 | 0 | 0 | 0 | | Not estimable |
| Sajaniemi 2001 | 0 | 0 | 0 | 0 | 0 | 0 | | Not estimable |
| Salo 2019 | 0 | 0 | 0 | 0 | 0 | 0 | | Not estimable |
| Santelices 2010 | 0 | 0 | 0 | 0 | 0 | 0 | | Not estimable |
| Schuler 2000 | 0 | 0 | 0 | 0 | 0 | 0 | | Not estimable |
| Shoghi 2018 | 0 | 0 | 0 | 0 | 0 | 0 | | Not estimable |
| Sleed 2013 | 0 | 0 | 0 | 0 | 0 | 0 | | Not estimable |
| Steele 2019 | 0 | 0 | 0 | 0 | 0 | 0 | | Not estimable |
| Stein 2006 | 0 | 0 | 0 | 0 | 0 | 0 | | Not estimable |
| Stein 2018 | 0 | 0 | 0 | 0 | 0 | 0 | | Not estimable |
| Taneja 2020 | 0 | 0 | 0 | 0 | 0 | 0 | | Not estimable |
| Tereno 2017 | 0 | 0 | 0 | 0 | 0 | 0 | | Not estimable |
| Teti 2009 | 0 | 0 | 0 | 0 | 0 | 0 | | Not estimable |
| Tryphonopoulos 2020 | 0 | 0 | 0 | 0 | 0 | 0 | | Not estimable |
| Tsivos 2015 | 0 | 0 | 0 | 0 | 0 | 0 | | Not estimable |
| Twohig 2021 | 0 | 0 | 0 | 0 | 0 | 0 | | Not estimable |
| Valades 2021 | 0 | 0 | 0 | 0 | 0 | 0 | | Not estimable |
| van Doesum 2008 | 0 | 0 | 0 | 0 | 0 | 0 | | Not estimable |
| Vargas-Porras 2021 | 0 | 0 | 0 | 0 | 0 | 0 | | Not estimable |
| Walter 2019 | 0 | 0 | 0 | 0 | 0 | 0 | | Not estimable |
| White-Traut 2013 | 0 | 0 | 0 | 0 | 0 | 0 | | Not estimable |
| Williams 2020 | 0 | 0 | 0 | 0 | 0 | 0 | | Not estimable |
| Wulff 2021 | 0 | 0 | 0 | 0 | 0 | 0 | | Not estimable |
| Youn 2021 | 0 | 0 | 0 | 0 | 0 | 0 | | Not estimable |
| Yu 2022 | 0 | 0 | 0 | 0 | 0 | 0 | | Not estimable |
| Zelkowitz 2011 | 0 | 0 | 0 | 0 | 0 | 0 | | Not estimable |
| **Total (95% CI)** | | | **762** | | | **666** | **100.0%** | **0.10 [-0.17, 0.37]** |

Heterogeneity: Tau² = 0.03; Chi² = 3.25, df = 2 (P = 0.20); I² = 39%

Test for overall effect: Z = 0.70 (P = 0.48)

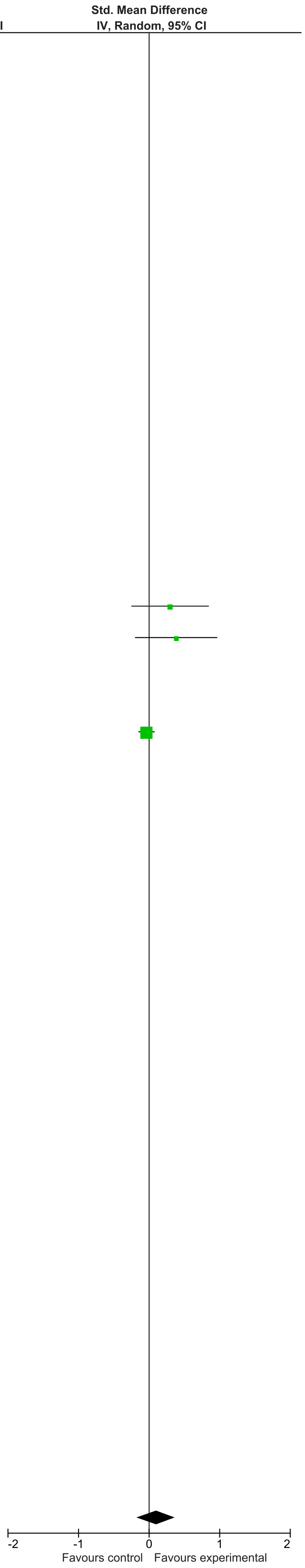

### eFigure 34. Child Behavior 0-1-month Follow-Up Forrest Plot

| | Early Dyadic Intervention | | | Standard Care | | | | Std. Mean Difference |
|---|---|---|---|---|---|---|---|---|
| Study or Subgroup | Mean | SD | Total | Mean | SD | Total | Weight | IV, Random, 95% CI |
| Akai 2008 | 0 | 0 | 0 | 0 | 0 | 0 | | Not estimable |
| Alarcao 2021 | 0 | 0 | 0 | 0 | 0 | 0 | | Not estimable |
| Alvarenga 2019 | 0 | 0 | 0 | 0 | 0 | 0 | | Not estimable |
| Badr 2006 | 0 | 0 | 0 | 0 | 0 | 0 | | Not estimable |
| Baggett 2010 | 0 | 0 | 0 | 0 | 0 | 0 | | Not estimable |
| Baggett 2017 | 0 | 0 | 0 | 0 | 0 | 0 | | Not estimable |
| Barlow 2016 | 0 | 0 | 0 | 0 | 0 | 0 | | Not estimable |
| Barnes 2022 | 0 | 0 | 0 | 0 | 0 | 0 | | Not estimable |
| Berlin 2017 | 0 | 0 | 0 | 0 | 0 | 0 | | Not estimable |
| Berlin 2018 | 0 | 0 | 0 | 0 | 0 | 0 | | Not estimable |
| Bernard 2012 | 0 | 0 | 0 | 0 | 0 | 0 | | Not estimable |
| Betancourt 2020 | 0 | 0 | 0 | 0 | 0 | 0 | | Not estimable |
| Bick 2013 | 0 | 0 | 0 | 0 | 0 | 0 | | Not estimable |
| Borghini 2014 | 0 | 0 | 0 | 0 | 0 | 0 | | Not estimable |
| Brisch 2003 | 0 | 0 | 0 | 0 | 0 | 0 | | Not estimable |
| Browne 2005 | 0 | 0 | 0 | 0 | 0 | 0 | | Not estimable |
| Cates 2018 | 0 | 0 | 0 | 0 | 0 | 0 | | Not estimable |
| Cevasco 2008 | 0 | 0 | 0 | 0 | 0 | 0 | | Not estimable |
| Chiu 2009 | 0 | 0 | 0 | 0 | 0 | 0 | | Not estimable |
| Constantino 2001 | 0 | 0 | 0 | 0 | 0 | 0 | | Not estimable |
| Cooper 2009 | 0 | 0 | 0 | 0 | 0 | 0 | | Not estimable |
| Cooper 2015 | 0 | 0 | 0 | 0 | 0 | 0 | | Not estimable |
| Corrigan 2021 | 0 | 0 | 0 | 0 | 0 | 0 | | Not estimable |
| Dozier 2009 | 0 | 0 | 0 | 0 | 0 | 0 | | Not estimable |
| Eriksen 2018 | 0 | 0 | 0 | 0 | 0 | 0 | | Not estimable |
| Feil 2020 | 0 | 0 | 0 | 0 | 0 | 0 | | Not estimable |
| Feldman 2014 | 0 | 0 | 0 | 0 | 0 | 0 | | Not estimable |
| Firk 2021 | -13.9 | 6.4 | 27 | -13.5 | 5.5 | 20 | 32.9% | -0.07 [-0.64, 0.51] |
| Fonagy 2016 | 0 | 0 | 0 | 0 | 0 | 0 | | Not estimable |
| Gaden 2022 | 0 | 0 | 0 | 0 | 0 | 0 | | Not estimable |
| Georg 2021 | 0 | 0 | 0 | 0 | 0 | 0 | | Not estimable |
| Glazebrook 2007 | 0 | 0 | 0 | 0 | 0 | 0 | | Not estimable |
| Hane 2015 | 0 | 0 | 0 | 0 | 0 | 0 | | Not estimable |
| Heo 2019 | 0 | 0 | 0 | 0 | 0 | 0 | | Not estimable |
| Herbers 2020 | 0 | 0 | 0 | 0 | 0 | 0 | | Not estimable |
| Hoffenkamp 2015 | 0 | 0 | 0 | 0 | 0 | 0 | | Not estimable |
| Hoivik 2015 | 0 | 0 | 0 | 0 | 0 | 0 | | Not estimable |
| Holditch-Davis 2014 | 0 | 0 | 0 | 0 | 0 | 0 | | Not estimable |
| Holt 2021 | 0 | 0 | 0 | 0 | 0 | 0 | | Not estimable |
| Horowitz 2001 | 0 | 0 | 0 | 0 | 0 | 0 | | Not estimable |
| Horowitz 2013 | 0 | 0 | 0 | 0 | 0 | 0 | | Not estimable |
| Juffer 2005 | 0 | 0 | 0 | 0 | 0 | 0 | | Not estimable |
| Klein Velderman 2006 | 0 | 0 | 0 | 0 | 0 | 0 | | Not estimable |
| Knoche 2012 | 0 | 0 | 0 | 0 | 0 | 0 | | Not estimable |
| Kristensen 2020 | 0 | 0 | 0 | 0 | 0 | 0 | | Not estimable |
| Landry 2006 | 0 | 0 | 0 | 0 | 0 | 0 | | Not estimable |
| Lavallee 2022 | 0 | 0 | 0 | 0 | 0 | 0 | | Not estimable |
| Magil-Evans 2007 | 0 | 0 | 0 | 0 | 0 | 0 | | Not estimable |
| Meijssen 2010 | 0 | 0 | 0 | 0 | 0 | 0 | | Not estimable |
| Melnyk 2006 | 0 | 0 | 0 | 0 | 0 | 0 | | Not estimable |
| Mendelsohn 2007 | -50.2 | 10 | 52 | -53.2 | 9.7 | 47 | 67.1% | 0.30 [-0.09, 0.70] |
| Milgrom 2013 | 0 | 0 | 0 | 0 | 0 | 0 | | Not estimable |
| Nelson 2001 | 0 | 0 | 0 | 0 | 0 | 0 | | Not estimable |
| Neu 2010 | 0 | 0 | 0 | 0 | 0 | 0 | | Not estimable |
| Newnham 2009 | 0 | 0 | 0 | 0 | 0 | 0 | | Not estimable |
| Nugent 2017 | 0 | 0 | 0 | 0 | 0 | 0 | | Not estimable |
| Onozawa 2001 | 0 | 0 | 0 | 0 | 0 | 0 | | Not estimable |
| Oxford 2021 | 0 | 0 | 0 | 0 | 0 | 0 | | Not estimable |
| Perrone 2021 | 0 | 0 | 0 | 0 | 0 | 0 | | Not estimable |
| Pontoppidan 2016 | 0 | 0 | 0 | 0 | 0 | 0 | | Not estimable |
| Porter 2015 | 0 | 0 | 0 | 0 | 0 | 0 | | Not estimable |
| Ramsauer 2019 | 0 | 0 | 0 | 0 | 0 | 0 | | Not estimable |
| Ravn 2011 | 0 | 0 | 0 | 0 | 0 | 0 | | Not estimable |
| Rheinheimer 2022 | 0 | 0 | 0 | 0 | 0 | 0 | | Not estimable |
| Robertson 2019 | 0 | 0 | 0 | 0 | 0 | 0 | | Not estimable |
| Roby 2021 | 0 | 0 | 0 | 0 | 0 | 0 | | Not estimable |
| Sadler 2013 | 0 | 0 | 0 | 0 | 0 | 0 | | Not estimable |
| Sahlen Helmer 2020 | 0 | 0 | 0 | 0 | 0 | 0 | | Not estimable |
| Sajaniemi 2001 | 0 | 0 | 0 | 0 | 0 | 0 | | Not estimable |
| Salo 2019 | 0 | 0 | 0 | 0 | 0 | 0 | | Not estimable |
| Santelices 2010 | 0 | 0 | 0 | 0 | 0 | 0 | | Not estimable |
| Schuler 2000 | 0 | 0 | 0 | 0 | 0 | 0 | | Not estimable |
| Shoghi 2018 | 0 | 0 | 0 | 0 | 0 | 0 | | Not estimable |
| Sleed 2013 | 0 | 0 | 0 | 0 | 0 | 0 | | Not estimable |
| Steele 2019 | 0 | 0 | 0 | 0 | 0 | 0 | | Not estimable |
| Stein 2006 | 0 | 0 | 0 | 0 | 0 | 0 | | Not estimable |
| Stein 2018 | 0 | 0 | 0 | 0 | 0 | 0 | | Not estimable |
| Taneja 2020 | 0 | 0 | 0 | 0 | 0 | 0 | | Not estimable |
| Tereno 2017 | 0 | 0 | 0 | 0 | 0 | 0 | | Not estimable |
| Teti 2009 | 0 | 0 | 0 | 0 | 0 | 0 | | Not estimable |
| Tryphonopoulos 2020 | 0 | 0 | 0 | 0 | 0 | 0 | | Not estimable |
| Tsivos 2015 | 0 | 0 | 0 | 0 | 0 | 0 | | Not estimable |
| Twohig 2021 | 0 | 0 | 0 | 0 | 0 | 0 | | Not estimable |
| Valades 2021 | 0 | 0 | 0 | 0 | 0 | 0 | | Not estimable |
| van Doesum 2008 | 0 | 0 | 0 | 0 | 0 | 0 | | Not estimable |
| Vargas-Porras 2021 | 0 | 0 | 0 | 0 | 0 | 0 | | Not estimable |
| Walter 2019 | 0 | 0 | 0 | 0 | 0 | 0 | | Not estimable |
| White-Traut 2013 | 0 | 0 | 0 | 0 | 0 | 0 | | Not estimable |
| Williams 2020 | 0 | 0 | 0 | 0 | 0 | 0 | | Not estimable |
| Wulff 2021 | 0 | 0 | 0 | 0 | 0 | 0 | | Not estimable |
| Youn 2021 | 0 | 0 | 0 | 0 | 0 | 0 | | Not estimable |
| Yu 2022 | 0 | 0 | 0 | 0 | 0 | 0 | | Not estimable |
| Zelkowitz 2011 | 0 | 0 | 0 | 0 | 0 | 0 | | Not estimable |
| **Total (95% CI)** | | | **79** | | | **67** | **100.0%** | **0.18 [-0.16, 0.52]** |

Heterogeneity: Tau² = 0.00; Chi² = 1.05, df = 1 (P = 0.31); I² = 5%

Test for overall effect: Z = 1.05 (P = 0.29)

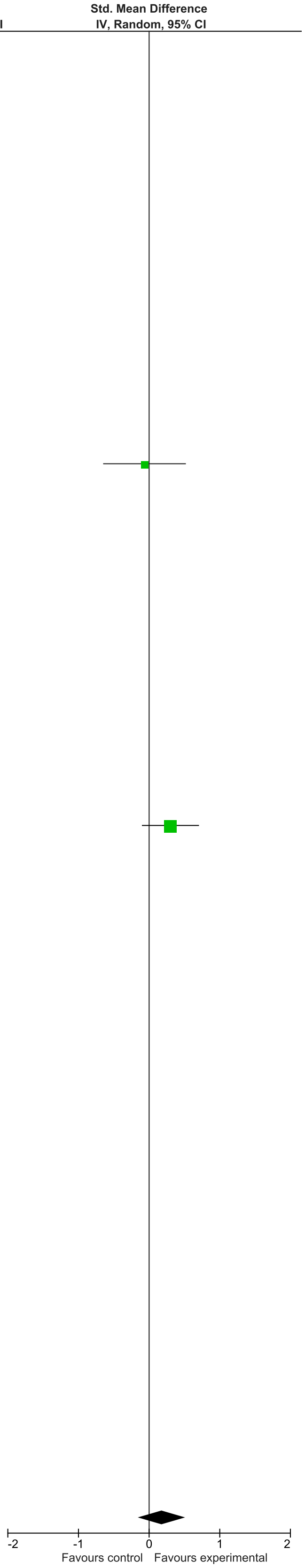

### eFigure 35. Child Behavior 2-6-month Follow-Up Forrest Plot

| | Early Dyadic Intervention | | | Standard Care | | | | Std. Mean Difference |
|---|---|---|---|---|---|---|---|---|
| Study or Subgroup | Mean | SD | Total | Mean | SD | Total | Weight | IV, Random, 95% CI |
| Akai 2008 | 0 | 0 | 0 | 0 | 0 | 0 | | Not estimable |
| Alarcao 2021 | 0 | 0 | 0 | 0 | 0 | 0 | | Not estimable |
| Alvarenga 2019 | 0 | 0 | 0 | 0 | 0 | 0 | | Not estimable |
| Badr 2006 | 0 | 0 | 0 | 0 | 0 | 0 | | Not estimable |
| Baggett 2010 | 0 | 0 | 0 | 0 | 0 | 0 | | Not estimable |
| Baggett 2017 | 0 | 0 | 0 | 0 | 0 | 0 | | Not estimable |
| Barlow 2016 | 0 | 0 | 0 | 0 | 0 | 0 | | Not estimable |
| Barnes 2022 | 0 | 0 | 0 | 0 | 0 | 0 | | Not estimable |
| Berlin 2017 | 0 | 0 | 0 | 0 | 0 | 0 | | Not estimable |
| Berlin 2018 | 0 | 0 | 0 | 0 | 0 | 0 | | Not estimable |
| Bernard 2012 | 0 | 0 | 0 | 0 | 0 | 0 | | Not estimable |
| Betancourt 2020 | 0 | 0 | 0 | 0 | 0 | 0 | | Not estimable |
| Bick 2013 | 0 | 0 | 0 | 0 | 0 | 0 | | Not estimable |
| Borghini 2014 | 0 | 0 | 0 | 0 | 0 | 0 | | Not estimable |
| Brisch 2003 | 0 | 0 | 0 | 0 | 0 | 0 | | Not estimable |
| Browne 2005 | 0 | 0 | 0 | 0 | 0 | 0 | | Not estimable |
| Cates 2018 | 0 | 0 | 0 | 0 | 0 | 0 | | Not estimable |
| Cevasco 2008 | 0 | 0 | 0 | 0 | 0 | 0 | | Not estimable |
| Chiu 2009 | 0 | 0 | 0 | 0 | 0 | 0 | | Not estimable |
| Constantino 2001 | 0 | 0 | 0 | 0 | 0 | 0 | | Not estimable |
| Cooper 2009 | 0 | 0 | 0 | 0 | 0 | 0 | | Not estimable |
| Cooper 2015 | 0 | 0 | 0 | 0 | 0 | 0 | | Not estimable |
| Corrigan 2021 | 0 | 0 | 0 | 0 | 0 | 0 | | Not estimable |
| Dozier 2009 | 0 | 0 | 0 | 0 | 0 | 0 | | Not estimable |
| Eriksen 2018 | 0 | 0 | 0 | 0 | 0 | 0 | | Not estimable |
| Feil 2020 | 0 | 0 | 0 | 0 | 0 | 0 | | Not estimable |
| Feldman 2014 | 0 | 0 | 0 | 0 | 0 | 0 | | Not estimable |
| Firk 2021 | -13.38 | 7.6 | 24 | -13.43 | 6 | 21 | 38.7% | 0.01 [-0.58, 0.59] |
| Fonagy 2016 | 0 | 0 | 0 | 0 | 0 | 0 | | Not estimable |
| Gaden 2022 | 0 | 0 | 0 | 0 | 0 | 0 | | Not estimable |
| Georg 2021 | 0 | 0 | 0 | 0 | 0 | 0 | | Not estimable |
| Glazebrook 2007 | 0 | 0 | 0 | 0 | 0 | 0 | | Not estimable |
| Hane 2015 | 0 | 0 | 0 | 0 | 0 | 0 | | Not estimable |
| Heo 2019 | 0 | 0 | 0 | 0 | 0 | 0 | | Not estimable |
| Herbers 2020 | 0 | 0 | 0 | 0 | 0 | 0 | | Not estimable |
| Hoffenkamp 2015 | 0 | 0 | 0 | 0 | 0 | 0 | | Not estimable |
| Hoivik 2015 | 0 | 0 | 0 | 0 | 0 | 0 | | Not estimable |
| Holditch-Davis 2014 | 0 | 0 | 0 | 0 | 0 | 0 | | Not estimable |
| Holt 2021 | 0 | 0 | 0 | 0 | 0 | 0 | | Not estimable |
| Horowitz 2001 | 0 | 0 | 0 | 0 | 0 | 0 | | Not estimable |
| Horowitz 2013 | 0 | 0 | 0 | 0 | 0 | 0 | | Not estimable |
| Juffer 2005 | 0 | 0 | 0 | 0 | 0 | 0 | | Not estimable |
| Klein Velderman 2006 | 0 | 0 | 0 | 0 | 0 | 0 | | Not estimable |
| Knoche 2012 | 0 | 0 | 0 | 0 | 0 | 0 | | Not estimable |
| Kristensen 2020 | 0 | 0 | 0 | 0 | 0 | 0 | | Not estimable |
| Landry 2006 | 0 | 0 | 0 | 0 | 0 | 0 | | Not estimable |
| Lavallee 2022 | 0 | 0 | 0 | 0 | 0 | 0 | | Not estimable |
| Magil-Evans 2007 | 0 | 0 | 0 | 0 | 0 | 0 | | Not estimable |
| Meijssen 2010 | 0 | 0 | 0 | 0 | 0 | 0 | | Not estimable |
| Melnyk 2006 | 0 | 0 | 0 | 0 | 0 | 0 | | Not estimable |
| Mendelsohn 2007 | 0 | 0 | 0 | 0 | 0 | 0 | | Not estimable |
| Milgrom 2013 | 0 | 0 | 0 | 0 | 0 | 0 | | Not estimable |
| Nelson 2001 | 0 | 0 | 0 | 0 | 0 | 0 | | Not estimable |
| Neu 2010 | 0 | 0 | 0 | 0 | 0 | 0 | | Not estimable |
| Newnham 2009 | 0 | 0 | 0 | 0 | 0 | 0 | | Not estimable |
| Nugent 2017 | 0 | 0 | 0 | 0 | 0 | 0 | | Not estimable |
| Onozawa 2001 | 0 | 0 | 0 | 0 | 0 | 0 | | Not estimable |
| Oxford 2021 | 0 | 0 | 0 | 0 | 0 | 0 | | Not estimable |
| Perrone 2021 | 0 | 0 | 0 | 0 | 0 | 0 | | Not estimable |
| Pontoppidan 2016 | 0 | 0 | 0 | 0 | 0 | 0 | | Not estimable |
| Porter 2015 | 0 | 0 | 0 | 0 | 0 | 0 | | Not estimable |
| Ramsauer 2019 | 0 | 0 | 0 | 0 | 0 | 0 | | Not estimable |
| Ravn 2011 | 0 | 0 | 0 | 0 | 0 | 0 | | Not estimable |
| Rheinheimer 2022 | 0 | 0 | 0 | 0 | 0 | 0 | | Not estimable |
| Robertson 2019 | 0 | 0 | 0 | 0 | 0 | 0 | | Not estimable |
| Roby 2021 | 0 | 0 | 0 | 0 | 0 | 0 | | Not estimable |
| Sadler 2013 | 0 | 0 | 0 | 0 | 0 | 0 | | Not estimable |
| Sahlen Helmer 2020 | 0 | 0 | 0 | 0 | 0 | 0 | | Not estimable |
| Sajaniemi 2001 | 0 | 0 | 0 | 0 | 0 | 0 | | Not estimable |
| Salo 2019 | 0 | 0 | 0 | 0 | 0 | 0 | | Not estimable |
| Santelices 2010 | 0 | 0 | 0 | 0 | 0 | 0 | | Not estimable |
| Schuler 2000 | 0 | 0 | 0 | 0 | 0 | 0 | | Not estimable |
| Shoghi 2018 | 0 | 0 | 0 | 0 | 0 | 0 | | Not estimable |
| Sleed 2013 | 0 | 0 | 0 | 0 | 0 | 0 | | Not estimable |
| Steele 2019 | 0 | 0 | 0 | 0 | 0 | 0 | | Not estimable |
| Stein 2006 | 0 | 0 | 0 | 0 | 0 | 0 | | Not estimable |
| Stein 2018 | 0 | 0 | 0 | 0 | 0 | 0 | | Not estimable |
| Taneja 2020 | 0 | 0 | 0 | 0 | 0 | 0 | | Not estimable |
| Tereno 2017 | 0 | 0 | 0 | 0 | 0 | 0 | | Not estimable |
| Teti 2009 | 0 | 0 | 0 | 0 | 0 | 0 | | Not estimable |
| Tryphonopoulos 2020 | 0 | 0 | 0 | 0 | 0 | 0 | | Not estimable |
| Tsivos 2015 | 0 | 0 | 0 | 0 | 0 | 0 | | Not estimable |
| Twohig 2021 | 0 | 0 | 0 | 0 | 0 | 0 | | Not estimable |
| Valades 2021 | 0 | 0 | 0 | 0 | 0 | 0 | | Not estimable |
| van Doesum 2008 | -0.5 | 0.52 | 35 | -0.48 | 0.43 | 36 | 61.3% | -0.04 [-0.51, 0.42] |
| Vargas-Porras 2021 | 0 | 0 | 0 | 0 | 0 | 0 | | Not estimable |
| Walter 2019 | 0 | 0 | 0 | 0 | 0 | 0 | | Not estimable |
| White-Traut 2013 | 0 | 0 | 0 | 0 | 0 | 0 | | Not estimable |
| Williams 2020 | 0 | 0 | 0 | 0 | 0 | 0 | | Not estimable |
| Wulff 2021 | 0 | 0 | 0 | 0 | 0 | 0 | | Not estimable |
| Youn 2021 | 0 | 0 | 0 | 0 | 0 | 0 | | Not estimable |
| Yu 2022 | 0 | 0 | 0 | 0 | 0 | 0 | | Not estimable |
| Zelkowitz 2011 | 0 | 0 | 0 | 0 | 0 | 0 | | Not estimable |
| **Total (95% CI)** | | | **59** | | | **57** | **100.0%** | **-0.02 [-0.39, 0.34]** |

Heterogeneity: Tau² = 0.00; Chi² = 0.02, df = 1 (P = 0.90); I² = 0%
Test for overall effect: Z = 0.12 (P = 0.90)

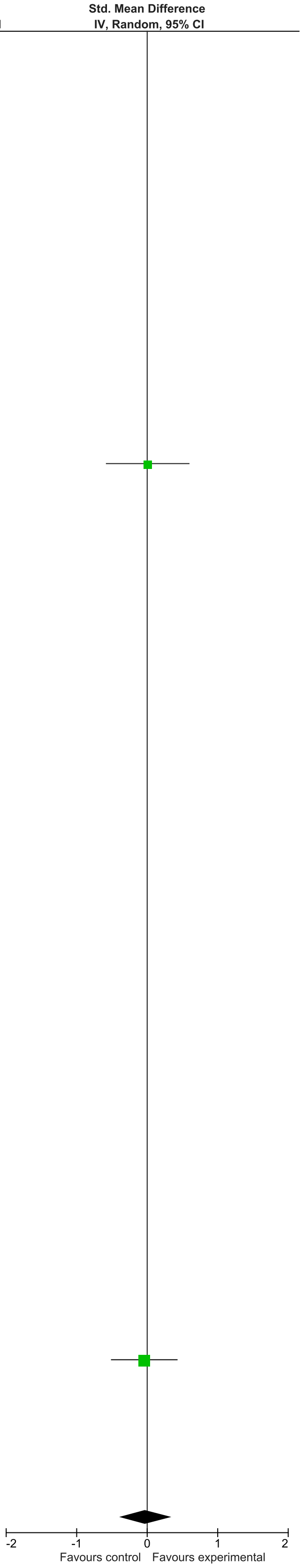

### eFigure 36. Child Behavior 13-24-month Follow-Up Forrest Plot

| | Early Dyadic Intervention | | | Standard Care | | | | Std. Mean Difference |
|---|---|---|---|---|---|---|---|---|
| Study or Subgroup | Mean | SD | Total | Mean | SD | Total | Weight | IV, Random, 95% CI |
| Akai 2008 | 0 | 0 | 0 | 0 | 0 | 0 | | Not estimable |
| Alarcao 2021 | 0 | 0 | 0 | 0 | 0 | 0 | | Not estimable |
| Alvarenga 2019 | 0 | 0 | 0 | 0 | 0 | 0 | | Not estimable |
| Badr 2006 | 0 | 0 | 0 | 0 | 0 | 0 | | Not estimable |
| Baggett 2010 | 0 | 0 | 0 | 0 | 0 | 0 | | Not estimable |
| Baggett 2017 | 0 | 0 | 0 | 0 | 0 | 0 | | Not estimable |
| Barlow 2016 | 0 | 0 | 0 | 0 | 0 | 0 | | Not estimable |
| Barnes 2022 | 0 | 0 | 0 | 0 | 0 | 0 | | Not estimable |
| Berlin 2017 | 0 | 0 | 0 | 0 | 0 | 0 | | Not estimable |
| Berlin 2018 | 0 | 0 | 0 | 0 | 0 | 0 | | Not estimable |
| Bernard 2012 | 0 | 0 | 0 | 0 | 0 | 0 | | Not estimable |
| Betancourt 2020 | 0 | 0 | 0 | 0 | 0 | 0 | | Not estimable |
| Bick 2013 | 0 | 0 | 0 | 0 | 0 | 0 | | Not estimable |
| Borghini 2014 | 0 | 0 | 0 | 0 | 0 | 0 | | Not estimable |
| Brisch 2003 | 0 | 0 | 0 | 0 | 0 | 0 | | Not estimable |
| Browne 2005 | 0 | 0 | 0 | 0 | 0 | 0 | | Not estimable |
| Cates 2018 | 0 | 0 | 0 | 0 | 0 | 0 | | Not estimable |
| Cevasco 2008 | 0 | 0 | 0 | 0 | 0 | 0 | | Not estimable |
| Chiu 2009 | 0 | 0 | 0 | 0 | 0 | 0 | | Not estimable |
| Constantino 2001 | 0 | 0 | 0 | 0 | 0 | 0 | | Not estimable |
| Cooper 2009 | 0 | 0 | 0 | 0 | 0 | 0 | | Not estimable |
| Cooper 2015 | -3.83 | 3.08 | 52 | -3.9 | 3.26 | 59 | 17.5% | 0.02 [-0.35, 0.39] |
| Corrigan 2021 | 0 | 0 | 0 | 0 | 0 | 0 | | Not estimable |
| Dozier 2009 | 0 | 0 | 0 | 0 | 0 | 0 | | Not estimable |
| Eriksen 2018 | 0 | 0 | 0 | 0 | 0 | 0 | | Not estimable |
| Feil 2020 | 0 | 0 | 0 | 0 | 0 | 0 | | Not estimable |
| Feldman 2014 | 0 | 0 | 0 | 0 | 0 | 0 | | Not estimable |
| Firk 2021 | 0 | 0 | 0 | 0 | 0 | 0 | | Not estimable |
| Fonagy 2016 | 0 | 0 | 0 | 0 | 0 | 0 | | Not estimable |
| Gaden 2022 | 0 | 0 | 0 | 0 | 0 | 0 | | Not estimable |
| Georg 2021 | 0 | 0 | 0 | 0 | 0 | 0 | | Not estimable |
| Glazebrook 2007 | 0 | 0 | 0 | 0 | 0 | 0 | | Not estimable |
| Hane 2015 | -17.7 | 2.7 | 26 | -22.4 | 2.5 | 31 | 13.5% | 1.79 [1.17, 2.41] |
| Heo 2019 | 0 | 0 | 0 | 0 | 0 | 0 | | Not estimable |
| Herbers 2020 | 0 | 0 | 0 | 0 | 0 | 0 | | Not estimable |
| Hoffenkamp 2015 | 0 | 0 | 0 | 0 | 0 | 0 | | Not estimable |
| Hoivik 2015 | 0 | 0 | 0 | 0 | 0 | 0 | | Not estimable |
| Holditch-Davis 2014 | 0 | 0 | 0 | 0 | 0 | 0 | | Not estimable |
| Holt 2021 | 0 | 0 | 0 | 0 | 0 | 0 | | Not estimable |
| Horowitz 2001 | 0 | 0 | 0 | 0 | 0 | 0 | | Not estimable |
| Horowitz 2013 | 0 | 0 | 0 | 0 | 0 | 0 | | Not estimable |
| Juffer 2005 | 0 | 0 | 0 | 0 | 0 | 0 | | Not estimable |
| Klein Velderman 2006 | -40.9 | 17.6 | 51 | -40.35 | 19.77 | 26 | 15.9% | -0.03 [-0.50, 0.44] |
| Knoche 2012 | 0 | 0 | 0 | 0 | 0 | 0 | | Not estimable |
| Kristensen 2020 | 0 | 0 | 0 | 0 | 0 | 0 | | Not estimable |
| Landry 2006 | 0 | 0 | 0 | 0 | 0 | 0 | | Not estimable |
| Lavallee 2022 | 0 | 0 | 0 | 0 | 0 | 0 | | Not estimable |
| Magil-Evans 2007 | 0 | 0 | 0 | 0 | 0 | 0 | | Not estimable |
| Meijssen 2010 | 0 | 0 | 0 | 0 | 0 | 0 | | Not estimable |
| Melnyk 2006 | 0 | 0 | 0 | 0 | 0 | 0 | | Not estimable |
| Mendelsohn 2007 | 0 | 0 | 0 | 0 | 0 | 0 | | Not estimable |
| Milgrom 2013 | -10.48 | 6.31 | 50 | -12.53 | 9.5 | 51 | 17.2% | 0.25 [-0.14, 0.64] |
| Nelson 2001 | 0 | 0 | 0 | 0 | 0 | 0 | | Not estimable |
| Neu 2010 | 0 | 0 | 0 | 0 | 0 | 0 | | Not estimable |
| Newnham 2009 | 0 | 0 | 0 | 0 | 0 | 0 | | Not estimable |
| Nugent 2017 | 0 | 0 | 0 | 0 | 0 | 0 | | Not estimable |
| Onozawa 2001 | 0 | 0 | 0 | 0 | 0 | 0 | | Not estimable |
| Oxford 2021 | 0 | 0 | 0 | 0 | 0 | 0 | | Not estimable |
| Perrone 2021 | 0 | 0 | 0 | 0 | 0 | 0 | | Not estimable |
| Pontoppidan 2016 | 0 | 0 | 0 | 0 | 0 | 0 | | Not estimable |
| Porter 2015 | 0 | 0 | 0 | 0 | 0 | 0 | | Not estimable |
| Ramsauer 2019 | 0 | 0 | 0 | 0 | 0 | 0 | | Not estimable |
| Ravn 2011 | 0 | 0 | 0 | 0 | 0 | 0 | | Not estimable |
| Rheinheimer 2022 | 0 | 0 | 0 | 0 | 0 | 0 | | Not estimable |
| Robertson 2019 | 0 | 0 | 0 | 0 | 0 | 0 | | Not estimable |
| Roby 2021 | 0 | 0 | 0 | 0 | 0 | 0 | | Not estimable |
| Sadler 2013 | 0 | 0 | 0 | 0 | 0 | 0 | | Not estimable |
| Sahlen Helmer 2020 | 0 | 0 | 0 | 0 | 0 | 0 | | Not estimable |
| Sajaniemi 2001 | 0 | 0 | 0 | 0 | 0 | 0 | | Not estimable |
| Salo 2019 | 0 | 0 | 0 | 0 | 0 | 0 | | Not estimable |
| Santelices 2010 | 0 | 0 | 0 | 0 | 0 | 0 | | Not estimable |
| Schuler 2000 | 0 | 0 | 0 | 0 | 0 | 0 | | Not estimable |
| Shoghi 2018 | 0 | 0 | 0 | 0 | 0 | 0 | | Not estimable |
| Sleed 2013 | 0 | 0 | 0 | 0 | 0 | 0 | | Not estimable |
| Steele 2019 | 0 | 0 | 0 | 0 | 0 | 0 | | Not estimable |
| Stein 2006 | 0 | 0 | 0 | 0 | 0 | 0 | | Not estimable |
| Stein 2018 | -12.5 | 8.8 | 64 | -14.1 | 7.2 | 68 | 18.0% | 0.20 [-0.14, 0.54] |
| Taneja 2020 | 0 | 0 | 0 | 0 | 0 | 0 | | Not estimable |
| Tereno 2017 | 0 | 0 | 0 | 0 | 0 | 0 | | Not estimable |
| Teti 2009 | 0 | 0 | 0 | 0 | 0 | 0 | | Not estimable |
| Tryphonopoulos 2020 | 0 | 0 | 0 | 0 | 0 | 0 | | Not estimable |
| Tsivos 2015 | 0 | 0 | 0 | 0 | 0 | 0 | | Not estimable |
| Twohig 2021 | 0 | 0 | 0 | 0 | 0 | 0 | | Not estimable |
| Valades 2021 | 0 | 0 | 0 | 0 | 0 | 0 | | Not estimable |
| van Doesum 2008 | 0 | 0 | 0 | 0 | 0 | 0 | | Not estimable |
| Vargas-Porras 2021 | 0 | 0 | 0 | 0 | 0 | 0 | | Not estimable |
| Walter 2019 | 0 | 0 | 0 | 0 | 0 | 0 | | Not estimable |
| White-Traut 2013 | 0 | 0 | 0 | 0 | 0 | 0 | | Not estimable |
| Williams 2020 | 0 | 0 | 0 | 0 | 0 | 0 | | Not estimable |
| Wulff 2021 | 0 | 0 | 0 | 0 | 0 | 0 | | Not estimable |
| Youn 2021 | -12.5 | 8.8 | 64 | -14.1 | 7.2 | 68 | 18.0% | 0.20 [-0.14, 0.54] |
| Yu 2022 | 0 | 0 | 0 | 0 | 0 | 0 | | Not estimable |
| Zelkowitz 2011 | 0 | 0 | 0 | 0 | 0 | 0 | | Not estimable |
| **Total (95% CI)** | | | **307** | | | **303** | **100.0%** | **0.35 [-0.03, 0.74]** |

Heterogeneity: Tau² = 0.18; Chi² = 26.41, df = 5 (P < 0.0001); I² = 81%
Test for overall effect: Z = 1.82 (P = 0.07)

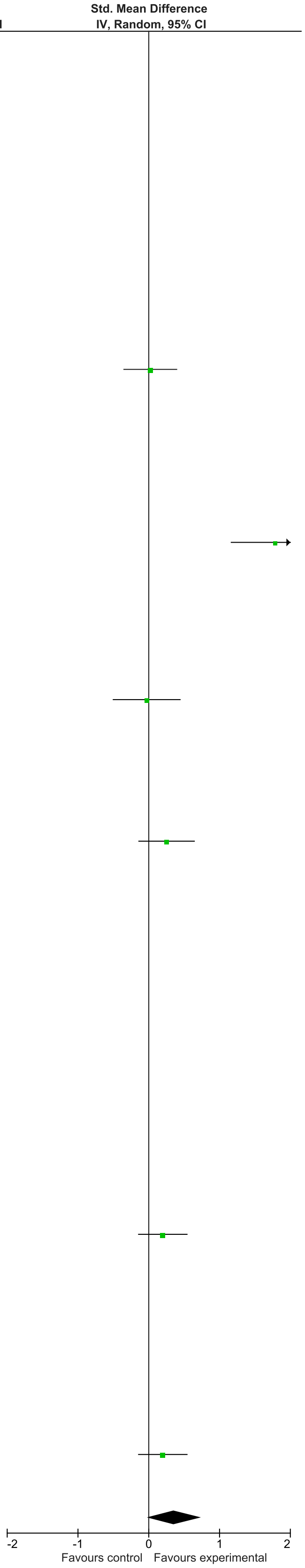

### eFigure 37. Child Behavior 25+-month Follow-Up Forrest Plot

| | Early Dyadic Intervention | | | Standard Care | | | | Std. Mean Difference |
|---|---|---|---|---|---|---|---|---|
| Study or Subgroup | Mean | SD | Total | Mean | SD | Total | Weight | IV, Random, 95% CI |
| Akai 2008 | 0 | 0 | 0 | 0 | 0 | 0 | | Not estimable |
| Alarcao 2021 | 0 | 0 | 0 | 0 | 0 | 0 | | Not estimable |
| Alvarenga 2019 | 0 | 0 | 0 | 0 | 0 | 0 | | Not estimable |
| Badr 2006 | 0 | 0 | 0 | 0 | 0 | 0 | | Not estimable |
| Baggett 2010 | 0 | 0 | 0 | 0 | 0 | 0 | | Not estimable |
| Baggett 2017 | 0 | 0 | 0 | 0 | 0 | 0 | | Not estimable |
| Barlow 2016 | 0 | 0 | 0 | 0 | 0 | 0 | | Not estimable |
| Barnes 2022 | 0 | 0 | 0 | 0 | 0 | 0 | | Not estimable |
| Berlin 2017 | 0 | 0 | 0 | 0 | 0 | 0 | | Not estimable |
| Berlin 2018 | 0 | 0 | 0 | 0 | 0 | 0 | | Not estimable |
| Bernard 2012 | 0 | 0 | 0 | 0 | 0 | 0 | | Not estimable |
| Betancourt 2020 | 0 | 0 | 0 | 0 | 0 | 0 | | Not estimable |
| Bick 2013 | 0 | 0 | 0 | 0 | 0 | 0 | | Not estimable |
| Borghini 2014 | 0 | 0 | 0 | 0 | 0 | 0 | | Not estimable |
| Brisch 2003 | 0 | 0 | 0 | 0 | 0 | 0 | | Not estimable |
| Browne 2005 | 0 | 0 | 0 | 0 | 0 | 0 | | Not estimable |
| Cates 2018 | 0 | 0 | 0 | 0 | 0 | 0 | | Not estimable |
| Cevasco 2008 | 0 | 0 | 0 | 0 | 0 | 0 | | Not estimable |
| Chiu 2009 | 0 | 0 | 0 | 0 | 0 | 0 | | Not estimable |
| Constantino 2001 | 0 | 0 | 0 | 0 | 0 | 0 | | Not estimable |
| Cooper 2009 | 0 | 0 | 0 | 0 | 0 | 0 | | Not estimable |
| Cooper 2015 | 0 | 0 | 0 | 0 | 0 | 0 | | Not estimable |
| Corrigan 2021 | 0 | 0 | 0 | 0 | 0 | 0 | | Not estimable |
| Dozier 2009 | 0 | 0 | 0 | 0 | 0 | 0 | | Not estimable |
| Eriksen 2018 | 0 | 0 | 0 | 0 | 0 | 0 | | Not estimable |
| Feil 2020 | 0 | 0 | 0 | 0 | 0 | 0 | | Not estimable |
| Feldman 2014 | 0 | 0 | 0 | 0 | 0 | 0 | | Not estimable |
| Firk 2021 | 0 | 0 | 0 | 0 | 0 | 0 | | Not estimable |
| Fonagy 2016 | 0 | 0 | 0 | 0 | 0 | 0 | | Not estimable |
| Gaden 2022 | 0 | 0 | 0 | 0 | 0 | 0 | | Not estimable |
| Georg 2021 | 0 | 0 | 0 | 0 | 0 | 0 | | Not estimable |
| Glazebrook 2007 | 0 | 0 | 0 | 0 | 0 | 0 | | Not estimable |
| Hane 2015 | 0 | 0 | 0 | 0 | 0 | 0 | | Not estimable |
| Heo 2019 | 0 | 0 | 0 | 0 | 0 | 0 | | Not estimable |
| Herbers 2020 | 0 | 0 | 0 | 0 | 0 | 0 | | Not estimable |
| Hoffenkamp 2015 | 0 | 0 | 0 | 0 | 0 | 0 | | Not estimable |
| Hoivik 2015 | 0 | 0 | 0 | 0 | 0 | 0 | | Not estimable |
| Holditch-Davis 2014 | 0 | 0 | 0 | 0 | 0 | 0 | | Not estimable |
| Holt 2021 | 0 | 0 | 0 | 0 | 0 | 0 | | Not estimable |
| Horowitz 2001 | 0 | 0 | 0 | 0 | 0 | 0 | | Not estimable |
| Horowitz 2013 | 0 | 0 | 0 | 0 | 0 | 0 | | Not estimable |
| Juffer 2005 | 0 | 0 | 0 | 0 | 0 | 0 | | Not estimable |
| Klein Velderman 2006 | 0 | 0 | 0 | 0 | 0 | 0 | | Not estimable |
| Knoche 2012 | 0 | 0 | 0 | 0 | 0 | 0 | | Not estimable |
| Kristensen 2020 | 0 | 0 | 0 | 0 | 0 | 0 | | Not estimable |
| Landry 2006 | 0 | 0 | 0 | 0 | 0 | 0 | | Not estimable |
| Lavallee 2022 | 0 | 0 | 0 | 0 | 0 | 0 | | Not estimable |
| Magil-Evans 2007 | 0 | 0 | 0 | 0 | 0 | 0 | | Not estimable |
| Meijssen 2010 | 0 | 0 | 0 | 0 | 0 | 0 | | Not estimable |
| Melnyk 2006 | 0 | 0 | 0 | 0 | 0 | 0 | | Not estimable |
| Mendelsohn 2007 | 0 | 0 | 0 | 0 | 0 | 0 | | Not estimable |
| Milgrom 2013 | -27.68 | 17.22 | 42 | -26.13 | 15.22 | 40 | 34.1% | -0.09 [-0.53, 0.34] |
| Nelson 2001 | 0 | 0 | 0 | 0 | 0 | 0 | | Not estimable |
| Neu 2010 | 0 | 0 | 0 | 0 | 0 | 0 | | Not estimable |
| Newnham 2009 | 0 | 0 | 0 | 0 | 0 | 0 | | Not estimable |
| Nugent 2017 | 0 | 0 | 0 | 0 | 0 | 0 | | Not estimable |
| Onozawa 2001 | 0 | 0 | 0 | 0 | 0 | 0 | | Not estimable |
| Oxford 2021 | 0 | 0 | 0 | 0 | 0 | 0 | | Not estimable |
| Perrone 2021 | 0 | 0 | 0 | 0 | 0 | 0 | | Not estimable |
| Pontoppidan 2016 | 0 | 0 | 0 | 0 | 0 | 0 | | Not estimable |
| Porter 2015 | 0 | 0 | 0 | 0 | 0 | 0 | | Not estimable |
| Ramsauer 2019 | 0 | 0 | 0 | 0 | 0 | 0 | | Not estimable |
| Ravn 2011 | 0 | 0 | 0 | 0 | 0 | 0 | | Not estimable |
| Rheinheimer 2022 | 0 | 0 | 0 | 0 | 0 | 0 | | Not estimable |
| Robertson 2019 | 0 | 0 | 0 | 0 | 0 | 0 | | Not estimable |
| Roby 2021 | 0 | 0 | 0 | 0 | 0 | 0 | | Not estimable |
| Sadler 2013 | -48.4 | 9.5 | 43 | -52.4 | 11 | 54 | 35.0% | 0.38 [-0.02, 0.79] |
| Sahlen Helmer 2020 | 0 | 0 | 0 | 0 | 0 | 0 | | Not estimable |
| Sajaniemi 2001 | 0 | 0 | 0 | 0 | 0 | 0 | | Not estimable |
| Salo 2019 | 0 | 0 | 0 | 0 | 0 | 0 | | Not estimable |
| Santelices 2010 | 0 | 0 | 0 | 0 | 0 | 0 | | Not estimable |
| Schuler 2000 | 0 | 0 | 0 | 0 | 0 | 0 | | Not estimable |
| Shoghi 2018 | 0 | 0 | 0 | 0 | 0 | 0 | | Not estimable |
| Sleed 2013 | 0 | 0 | 0 | 0 | 0 | 0 | | Not estimable |
| Steele 2019 | 0 | 0 | 0 | 0 | 0 | 0 | | Not estimable |
| Stein 2006 | 0 | 0 | 0 | 0 | 0 | 0 | | Not estimable |
| Stein 2018 | 0 | 0 | 0 | 0 | 0 | 0 | | Not estimable |
| Taneja 2020 | 0 | 0 | 0 | 0 | 0 | 0 | | Not estimable |
| Tereno 2017 | 0 | 0 | 0 | 0 | 0 | 0 | | Not estimable |
| Teti 2009 | 0 | 0 | 0 | 0 | 0 | 0 | | Not estimable |
| Tryphonopoulos 2020 | 0 | 0 | 0 | 0 | 0 | 0 | | Not estimable |
| Tsivos 2015 | 0 | 0 | 0 | 0 | 0 | 0 | | Not estimable |
| Twohig 2021 | 0 | 0 | 0 | 0 | 0 | 0 | | Not estimable |
| Valades 2021 | 0 | 0 | 0 | 0 | 0 | 0 | | Not estimable |
| van Doesum 2008 | -51.96 | 2.92 | 29 | -50.09 | 2.91 | 29 | 31.0% | -0.63 [-1.16, -0.10] |
| Vargas-Porras 2021 | 0 | 0 | 0 | 0 | 0 | 0 | | Not estimable |
| Walter 2019 | 0 | 0 | 0 | 0 | 0 | 0 | | Not estimable |
| White-Traut 2013 | 0 | 0 | 0 | 0 | 0 | 0 | | Not estimable |
| Williams 2020 | 0 | 0 | 0 | 0 | 0 | 0 | | Not estimable |
| Wulff 2021 | 0 | 0 | 0 | 0 | 0 | 0 | | Not estimable |
| Youn 2021 | 0 | 0 | 0 | 0 | 0 | 0 | | Not estimable |
| Yu 2022 | 0 | 0 | 0 | 0 | 0 | 0 | | Not estimable |
| Zelkowitz 2011 | 0 | 0 | 0 | 0 | 0 | 0 | | Not estimable |
| **Total (95% CI)** | | | **114** | | | **123** | **100.0%** | **-0.09 [-0.65, 0.46]** |

Heterogeneity: $Tau^2 = 0.19$; $Chi^2 = 9.09$, df = 2 (P = 0.01); $I^2 = 78\%$

Test for overall effect: Z = 0.33 (P = 0.74)

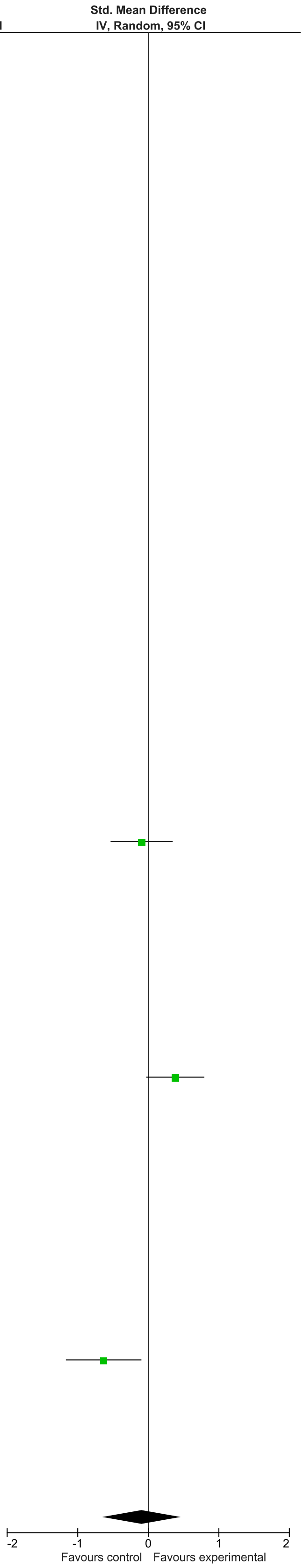

### eFigure 38. Parenting Stress 0-1-month Follow-Up Forrest Plot

| | Early Dyadic Intervention | | | Standard Care | | | | Std. Mean Difference |
|---|---|---|---|---|---|---|---|---|
| **Study or Subgroup** | **Mean** | **SD** | **Total** | **Mean** | **SD** | **Total** | **Weight** | **IV, Random, 95% CI** |
| Akai 2008 | 0 | 0 | 0 | 0 | 0 | 0 | | Not estimable |
| Alarcao 2021 | 0 | 0 | 0 | 0 | 0 | 0 | | Not estimable |
| Alvarenga 2019 | 0 | 0 | 0 | 0 | 0 | 0 | | Not estimable |
| Badr 2006 | 0 | 0 | 0 | 0 | 0 | 0 | | Not estimable |
| Baggett 2010 | 0 | 0 | 0 | 0 | 0 | 0 | | Not estimable |
| Baggett 2017 | 0 | 0 | 0 | 0 | 0 | 0 | | Not estimable |
| Barlow 2016 | -41.7 | 5.19 | 10 | -48.5 | 9.69 | 10 | 3.3% | 0.84 [-0.09, 1.76] |
| Barnes 2022 | 0 | 0 | 0 | 0 | 0 | 0 | | Not estimable |
| Berlin 2017 | 0 | 0 | 0 | 0 | 0 | 0 | | Not estimable |
| Berlin 2018 | 0 | 0 | 0 | 0 | 0 | 0 | | Not estimable |
| Bernard 2012 | 0 | 0 | 0 | 0 | 0 | 0 | | Not estimable |
| Betancourt 2020 | 0 | 0 | 0 | 0 | 0 | 0 | | Not estimable |
| Bick 2013 | 0 | 0 | 0 | 0 | 0 | 0 | | Not estimable |
| Borghini 2014 | 0 | 0 | 0 | 0 | 0 | 0 | | Not estimable |
| Brisch 2003 | 0 | 0 | 0 | 0 | 0 | 0 | | Not estimable |
| Browne 2005 | 0 | 0 | 0 | 0 | 0 | 0 | | Not estimable |
| Cates 2018 | 0 | 0 | 0 | 0 | 0 | 0 | | Not estimable |
| Cevasco 2008 | 0 | 0 | 0 | 0 | 0 | 0 | | Not estimable |
| Chiu 2009 | 0 | 0 | 0 | 0 | 0 | 0 | | Not estimable |
| Constantino 2001 | 0 | 0 | 0 | 0 | 0 | 0 | | Not estimable |
| Cooper 2009 | 0 | 0 | 0 | 0 | 0 | 0 | | Not estimable |
| Cooper 2015 | 0 | 0 | 0 | 0 | 0 | 0 | | Not estimable |
| Corrigan 2021 | 0 | 0 | 0 | 0 | 0 | 0 | | Not estimable |
| Dozier 2009 | 0 | 0 | 0 | 0 | 0 | 0 | | Not estimable |
| Eriksen 2018 | -81.75 | 12.83 | 13 | -74.17 | 16 | 12 | 4.2% | -0.51 [-1.31, 0.29] |
| Feil 2020 | 0 | 0 | 0 | 0 | 0 | 0 | | Not estimable |
| Feldman 2014 | 0 | 0 | 0 | 0 | 0 | 0 | | Not estimable |
| Firk 2021 | -104.07 | 29.7 | 27 | -121.25 | 34 | 20 | 6.7% | 0.53 [-0.05, 1.12] |
| Fonagy 2016 | -88.2 | 25.5 | 28 | -79.1 | 18.9 | 23 | 7.3% | -0.39 [-0.95, 0.16] |
| Gaden 2022 | 0 | 0 | 0 | 0 | 0 | 0 | | Not estimable |
| Georg 2021 | -128.23 | 32.08 | 81 | -130.41 | 34.32 | 73 | 13.7% | 0.07 [-0.25, 0.38] |
| Glazebrook 2007 | 0 | 0 | 0 | 0 | 0 | 0 | | Not estimable |
| Hane 2015 | 0 | 0 | 0 | 0 | 0 | 0 | | Not estimable |
| Heo 2019 | 0 | 0 | 0 | 0 | 0 | 0 | | Not estimable |
| Herbers 2020 | -45.38 | 5.37 | 20 | -45.21 | 7.55 | 18 | 6.0% | -0.03 [-0.66, 0.61] |
| Hoffenkamp 2015 | 0 | 0 | 0 | 0 | 0 | 0 | | Not estimable |
| Hoivik 2015 | 0 | 0 | 0 | 0 | 0 | 0 | | Not estimable |
| Holditch-Davis 2014 | 0 | 0 | 0 | 0 | 0 | 0 | | Not estimable |
| Holt 2021 | -257.68 | 44.5 | 27 | -244.93 | 34.53 | 28 | 7.8% | -0.32 [-0.85, 0.22] |
| Horowitz 2001 | 0 | 0 | 0 | 0 | 0 | 0 | | Not estimable |
| Horowitz 2013 | 0 | 0 | 0 | 0 | 0 | 0 | | Not estimable |
| Juffer 2005 | 0 | 0 | 0 | 0 | 0 | 0 | | Not estimable |
| Klein Velderman 2006 | 0 | 0 | 0 | 0 | 0 | 0 | | Not estimable |
| Knoche 2012 | 0 | 0 | 0 | 0 | 0 | 0 | | Not estimable |
| Kristensen 2020 | 0 | 0 | 0 | 0 | 0 | 0 | | Not estimable |
| Landry 2006 | 0 | 0 | 0 | 0 | 0 | 0 | | Not estimable |
| Lavallee 2022 | -62.6 | 21.74 | 10 | -78.75 | 27.47 | 10 | 3.4% | 0.62 [-0.28, 1.53] |
| Magil-Evans 2007 | 0 | 0 | 0 | 0 | 0 | 0 | | Not estimable |
| Meijssen 2010 | 0 | 0 | 0 | 0 | 0 | 0 | | Not estimable |
| Melnyk 2006 | 0 | 0 | 0 | 0 | 0 | 0 | | Not estimable |
| Mendelsohn 2007 | -71.8 | 23.8 | 51 | -80.6 | 18.8 | 46 | 10.9% | 0.40 [0.00, 0.81] |
| Milgrom 2013 | 0 | 0 | 0 | 0 | 0 | 0 | | Not estimable |
| Nelson 2001 | 0 | 0 | 0 | 0 | 0 | 0 | | Not estimable |
| Neu 2010 | 0 | 0 | 0 | 0 | 0 | 0 | | Not estimable |
| Newnham 2009 | -91.34 | 14.68 | 32 | -94.6 | 12.24 | 31 | 8.5% | 0.24 [-0.26, 0.73] |
| Nugent 2017 | 0 | 0 | 0 | 0 | 0 | 0 | | Not estimable |
| Onozawa 2001 | 0 | 0 | 0 | 0 | 0 | 0 | | Not estimable |
| Oxford 2021 | 0 | 0 | 0 | 0 | 0 | 0 | | Not estimable |
| Perrone 2021 | 0 | 0 | 0 | 0 | 0 | 0 | | Not estimable |
| Pontoppidan 2016 | 0 | 0 | 0 | 0 | 0 | 0 | | Not estimable |
| Porter 2015 | -151.73 | 21.93 | 41 | -147.58 | 25 | 24 | 8.3% | -0.18 [-0.68, 0.33] |
| Ramsauer 2019 | 0 | 0 | 0 | 0 | 0 | 0 | | Not estimable |
| Ravn 2011 | 0 | 0 | 0 | 0 | 0 | 0 | | Not estimable |
| Rheinheimer 2022 | 0 | 0 | 0 | 0 | 0 | 0 | | Not estimable |
| Robertson 2019 | 0 | 0 | 0 | 0 | 0 | 0 | | Not estimable |
| Roby 2021 | 0 | 0 | 0 | 0 | 0 | 0 | | Not estimable |
| Sadler 2013 | 0 | 0 | 0 | 0 | 0 | 0 | | Not estimable |
| Sahlen Helmer 2020 | 0 | 0 | 0 | 0 | 0 | 0 | | Not estimable |
| Sajaniemi 2001 | 0 | 0 | 0 | 0 | 0 | 0 | | Not estimable |
| Salo 2019 | 0 | 0 | 0 | 0 | 0 | 0 | | Not estimable |
| Santelices 2010 | 0 | 0 | 0 | 0 | 0 | 0 | | Not estimable |
| Schuler 2000 | 0 | 0 | 0 | 0 | 0 | 0 | | Not estimable |
| Shoghi 2018 | 0 | 0 | 0 | 0 | 0 | 0 | | Not estimable |
| Sleed 2013 | 0 | 0 | 0 | 0 | 0 | 0 | | Not estimable |
| Steele 2019 | 0 | 0 | 0 | 0 | 0 | 0 | | Not estimable |
| Stein 2006 | 0 | 0 | 0 | 0 | 0 | 0 | | Not estimable |
| Stein 2018 | 0 | 0 | 0 | 0 | 0 | 0 | | Not estimable |
| Taneja 2020 | 0 | 0 | 0 | 0 | 0 | 0 | | Not estimable |
| Tereno 2017 | 0 | 0 | 0 | 0 | 0 | 0 | | Not estimable |
| Teti 2009 | 0 | 0 | 0 | 0 | 0 | 0 | | Not estimable |
| Tryphonopoulos 2020 | 0 | 0 | 0 | 0 | 0 | 0 | | Not estimable |
| Tsivos 2015 | 0 | 0 | 0 | 0 | 0 | 0 | | Not estimable |
| Twohig 2021 | 0 | 0 | 0 | 0 | 0 | 0 | | Not estimable |
| Valades 2021 | 0 | 0 | 0 | 0 | 0 | 0 | | Not estimable |
| van Doesum 2008 | 0 | 0 | 0 | 0 | 0 | 0 | | Not estimable |
| Vargas-Porras 2021 | -3.21 | 2.7 | 33 | -2.79 | 2.07 | 33 | 8.8% | -0.17 [-0.66, 0.31] |
| Walter 2019 | 0 | 0 | 0 | 0 | 0 | 0 | | Not estimable |
| White-Traut 2013 | 0 | 0 | 0 | 0 | 0 | 0 | | Not estimable |
| Williams 2020 | 0 | 0 | 0 | 0 | 0 | 0 | | Not estimable |
| Wulff 2021 | 0 | 0 | 0 | 0 | 0 | 0 | | Not estimable |
| Youn 2021 | 0 | 0 | 0 | 0 | 0 | 0 | | Not estimable |
| Yu 2022 | 0 | 0 | 0 | 0 | 0 | 0 | | Not estimable |
| Zelkowitz 2011 | -1.85 | 0.86 | 48 | -1.95 | 1.45 | 50 | 11.1% | 0.08 [-0.31, 0.48] |
| **Total (95% CI)** | | | **421** | | | **378** | **100.0%** | **0.06 [-0.12, 0.24]** |

Heterogeneity: Tau² = 0.04; Chi² = 18.25, df = 12 (P = 0.11); I² = 34%

Test for overall effect: Z = 0.67 (P = 0.50)

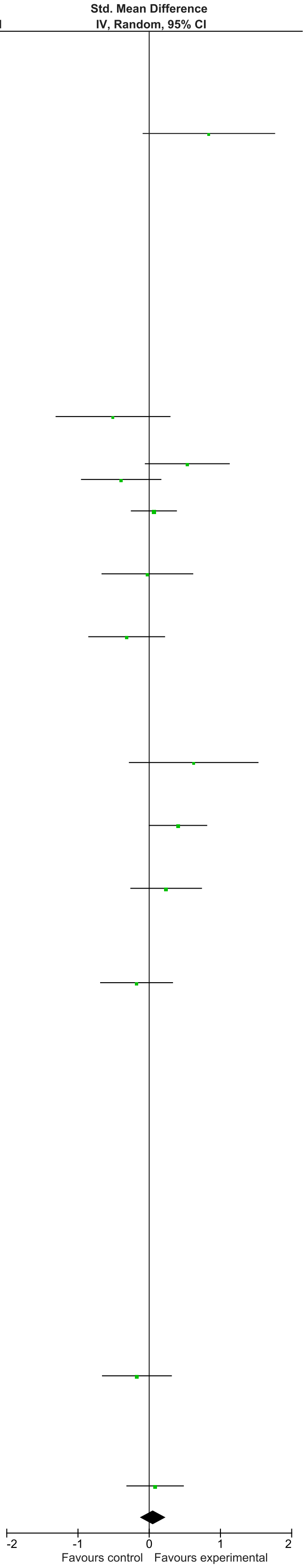

### eFigure 39. Parenting Stress 2-6-month Follow-Up Forrest Plot

| | Early Dyadic Intervention | | | Standard Care | | | | Std. Mean Difference |
|---|---|---|---|---|---|---|---|---|
| Study or Subgroup | Mean | SD | Total | Mean | SD | Total | Weight | IV, Random, 95% CI |
| Akai 2008 | 0 | 0 | 0 | 0 | 0 | 0 | | Not estimable |
| Alarcao 2021 | 0 | 0 | 0 | 0 | 0 | 0 | | Not estimable |
| Alvarenga 2019 | 0 | 0 | 0 | 0 | 0 | 0 | | Not estimable |
| Badr 2006 | -71.8 | 7.76 | 42 | -75.35 | 8.22 | 42 | 21.0% | 0.44 [0.01, 0.87] |
| Baggett 2010 | 0 | 0 | 0 | 0 | 0 | 0 | | Not estimable |
| Baggett 2017 | 0 | 0 | 0 | 0 | 0 | 0 | | Not estimable |
| Barlow 2016 | 0 | 0 | 0 | 0 | 0 | 10 | | Not estimable |
| Barnes 2022 | 0 | 0 | 0 | 0 | 0 | 0 | | Not estimable |
| Berlin 2017 | 0 | 0 | 0 | 0 | 0 | 0 | | Not estimable |
| Berlin 2018 | 0 | 0 | 0 | 0 | 0 | 0 | | Not estimable |
| Bernard 2012 | 0 | 0 | 0 | 0 | 0 | 0 | | Not estimable |
| Betancourt 2020 | 0 | 0 | 0 | 0 | 0 | 0 | | Not estimable |
| Bick 2013 | 0 | 0 | 0 | 0 | 0 | 0 | | Not estimable |
| Borghini 2014 | 0 | 0 | 0 | 0 | 0 | 0 | | Not estimable |
| Brisch 2003 | 0 | 0 | 0 | 0 | 0 | 0 | | Not estimable |
| Browne 2005 | 0 | 0 | 0 | 0 | 0 | 0 | | Not estimable |
| Cates 2018 | 0 | 0 | 0 | 0 | 0 | 0 | | Not estimable |
| Cevasco 2008 | 0 | 0 | 0 | 0 | 0 | 0 | | Not estimable |
| Chiu 2009 | 0 | 0 | 0 | 0 | 0 | 0 | | Not estimable |
| Constantino 2001 | 0 | 0 | 0 | 0 | 0 | 0 | | Not estimable |
| Cooper 2009 | 0 | 0 | 0 | 0 | 0 | 0 | | Not estimable |
| Cooper 2015 | 0 | 0 | 0 | 0 | 0 | 0 | | Not estimable |
| Corrigan 2021 | 0 | 0 | 0 | 0 | 0 | 0 | | Not estimable |
| Dozier 2009 | 0 | 0 | 0 | 0 | 0 | 0 | | Not estimable |
| Eriksen 2018 | 0 | 0 | 0 | 0 | 0 | 12 | | Not estimable |
| Feil 2020 | 0 | 0 | 0 | 0 | 0 | 0 | | Not estimable |
| Feldman 2014 | 0 | 0 | 0 | 0 | 0 | 0 | | Not estimable |
| Firk 2021 | -109.33 | 29.4 | 24 | -114 | 33.1 | 21 | 15.6% | 0.15 [-0.44, 0.73] |
| Fonagy 2016 | 0 | 0 | 0 | 0 | 0 | 0 | | Not estimable |
| Gaden 2022 | 0 | 0 | 0 | 0 | 0 | 0 | | Not estimable |
| Georg 2021 | 0 | 0 | 0 | 0 | 0 | 0 | | Not estimable |
| Glazebrook 2007 | -71.9 | 18.9 | 91 | -67.1 | 19.6 | 108 | 27.9% | -0.25 [-0.53, 0.03] |
| Hane 2015 | 0 | 0 | 0 | 0 | 0 | 0 | | Not estimable |
| Heo 2019 | 0 | 0 | 0 | 0 | 0 | 0 | | Not estimable |
| Herbers 2020 | 0 | 0 | 0 | 0 | 0 | 0 | | Not estimable |
| Hoffenkamp 2015 | 0 | 0 | 0 | 0 | 0 | 0 | | Not estimable |
| Hoivik 2015 | 0 | 0 | 0 | 0 | 0 | 0 | | Not estimable |
| Holditch-Davis 2014 | 0 | 0 | 0 | 0 | 0 | 0 | | Not estimable |
| Holt 2021 | -255.43 | 37.59 | 22 | -245.19 | 32.61 | 27 | 16.2% | -0.29 [-0.85, 0.28] |
| Horowitz 2001 | 0 | 0 | 0 | 0 | 0 | 0 | | Not estimable |
| Horowitz 2013 | 0 | 0 | 0 | 0 | 0 | 0 | | Not estimable |
| Juffer 2005 | 0 | 0 | 0 | 0 | 0 | 0 | | Not estimable |
| Klein Velderman 2006 | 0 | 0 | 0 | 0 | 0 | 0 | | Not estimable |
| Knoche 2012 | 0 | 0 | 0 | 0 | 0 | 0 | | Not estimable |
| Kristensen 2020 | 0 | 0 | 0 | 0 | 0 | 0 | | Not estimable |
| Landry 2006 | 0 | 0 | 0 | 0 | 0 | 0 | | Not estimable |
| Lavallee 2022 | 0 | 0 | 0 | 0 | 0 | 0 | | Not estimable |
| Magil-Evans 2007 | 0 | 0 | 0 | 0 | 0 | 0 | | Not estimable |
| Meijssen 2010 | 0 | 0 | 0 | 0 | 0 | 0 | | Not estimable |
| Melnyk 2006 | 0 | 0 | 0 | 0 | 0 | 0 | | Not estimable |
| Mendelsohn 2007 | 0 | 0 | 0 | 0 | 0 | 0 | | Not estimable |
| Milgrom 2013 | 0 | 0 | 0 | 0 | 0 | 0 | | Not estimable |
| Nelson 2001 | 0 | 0 | 0 | 0 | 0 | 0 | | Not estimable |
| Neu 2010 | 0 | 0 | 0 | 0 | 0 | 0 | | Not estimable |
| Newnham 2009 | 0 | 0 | 0 | 0 | 0 | 0 | | Not estimable |
| Nugent 2017 | 0 | 0 | 0 | 0 | 0 | 0 | | Not estimable |
| Onozawa 2001 | 0 | 0 | 0 | 0 | 0 | 0 | | Not estimable |
| Oxford 2021 | 0 | 0 | 0 | 0 | 0 | 0 | | Not estimable |
| Perrone 2021 | 0 | 0 | 0 | 0 | 0 | 0 | | Not estimable |
| Pontoppidan 2016 | 0 | 0 | 0 | 0 | 0 | 0 | | Not estimable |
| Porter 2015 | -158 | 32.27 | 40 | -147.67 | 23.5 | 30 | 19.3% | -0.35 [-0.83, 0.12] |
| Ramsauer 2019 | 0 | 0 | 0 | 0 | 0 | 0 | | Not estimable |
| Ravn 2011 | 0 | 0 | 0 | 0 | 0 | 0 | | Not estimable |
| Rheinheimer 2022 | 0 | 0 | 0 | 0 | 0 | 0 | | Not estimable |
| Robertson 2019 | 0 | 0 | 0 | 0 | 0 | 0 | | Not estimable |
| Roby 2021 | 0 | 0 | 0 | 0 | 0 | 0 | | Not estimable |
| Sadler 2013 | 0 | 0 | 0 | 0 | 0 | 0 | | Not estimable |
| Sahlen Helmer 2020 | 0 | 0 | 0 | 0 | 0 | 0 | | Not estimable |
| Sajaniemi 2001 | 0 | 0 | 0 | 0 | 0 | 0 | | Not estimable |
| Salo 2019 | 0 | 0 | 0 | 0 | 0 | 0 | | Not estimable |
| Santelices 2010 | 0 | 0 | 0 | 0 | 0 | 0 | | Not estimable |
| Schuler 2000 | 0 | 0 | 0 | 0 | 0 | 0 | | Not estimable |
| Shoghi 2018 | 0 | 0 | 0 | 0 | 0 | 0 | | Not estimable |
| Sleed 2013 | 0 | 0 | 0 | 0 | 0 | 0 | | Not estimable |
| Steele 2019 | 0 | 0 | 0 | 0 | 0 | 0 | | Not estimable |
| Stein 2006 | 0 | 0 | 0 | 0 | 0 | 0 | | Not estimable |
| Stein 2018 | 0 | 0 | 0 | 0 | 0 | 0 | | Not estimable |
| Taneja 2020 | 0 | 0 | 0 | 0 | 0 | 0 | | Not estimable |
| Tereno 2017 | 0 | 0 | 0 | 0 | 0 | 0 | | Not estimable |
| Teti 2009 | 0 | 0 | 0 | 0 | 0 | 0 | | Not estimable |
| Tryphonopoulos 2020 | 0 | 0 | 0 | 0 | 0 | 0 | | Not estimable |
| Tsivos 2015 | 0 | 0 | 0 | 0 | 0 | 0 | | Not estimable |
| Twohig 2021 | 0 | 0 | 0 | 0 | 0 | 0 | | Not estimable |
| Valades 2021 | 0 | 0 | 0 | 0 | 0 | 0 | | Not estimable |
| van Doesum 2008 | 0 | 0 | 0 | 0 | 0 | 0 | | Not estimable |
| Vargas-Porras 2021 | 0 | 0 | 0 | 0 | 0 | 0 | | Not estimable |
| Walter 2019 | 0 | 0 | 0 | 0 | 0 | 0 | | Not estimable |
| White-Traut 2013 | 0 | 0 | 0 | 0 | 0 | 0 | | Not estimable |
| Williams 2020 | 0 | 0 | 0 | 0 | 0 | 0 | | Not estimable |
| Wulff 2021 | 0 | 0 | 0 | 0 | 0 | 0 | | Not estimable |
| Youn 2021 | 0 | 0 | 0 | 0 | 0 | 0 | | Not estimable |
| Yu 2022 | 0 | 0 | 0 | 0 | 0 | 0 | | Not estimable |
| Zelkowitz 2011 | 0 | 0 | 0 | 0 | 0 | 0 | | Not estimable |
| **Total (95% CI)** | | | **219** | | | **250** | **100.0%** | **-0.07 [-0.37, 0.24]** |

Heterogeneity: Tau² = 0.07; Chi² = 9.24, df = 4 (P = 0.06); I² = 57%

Test for overall effect: Z = 0.44 (P = 0.66)

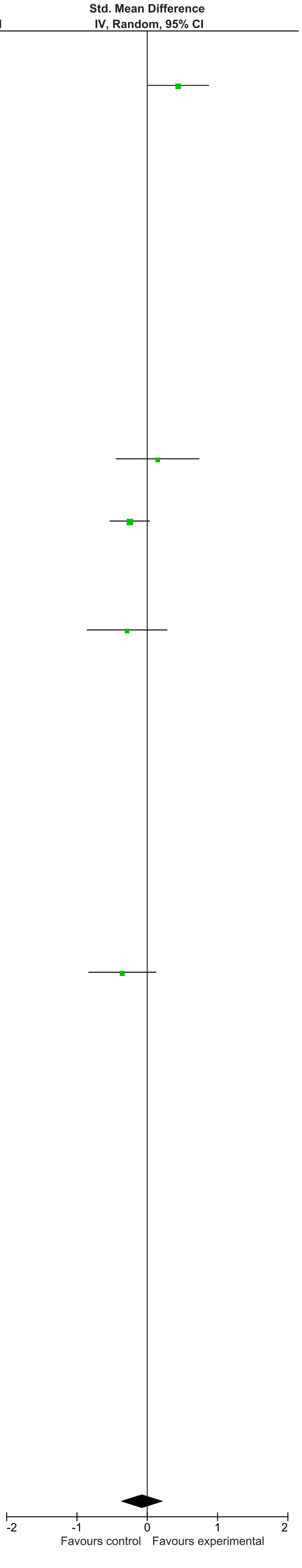

### eFigure 40. Parent/Caregiver Anxiety 0-1-month Follow-Up Forrest Plot

| | Early Dyadic Intervention | | | Standard Care | | | | Std. Mean Difference |
|---|---|---|---|---|---|---|---|---|
| **Study or Subgroup** | **Mean** | **SD** | **Total** | **Mean** | **SD** | **Total** | **Weight** | **IV, Random, 95% CI** |
| Akai 2008 | 0 | 0 | 0 | 0 | 0 | 0 | | Not estimable |
| Alarcao 2021 | 0 | 0 | 0 | 0 | 0 | 0 | | Not estimable |
| Alvarenga 2019 | 0 | 0 | 0 | 0 | 0 | 0 | | Not estimable |
| Badr 2006 | 0 | 0 | 0 | 0 | 0 | 0 | | Not estimable |
| Baggett 2010 | 0 | 0 | 0 | 0 | 0 | 0 | | Not estimable |
| Baggett 2017 | 0 | 0 | 0 | 0 | 0 | 0 | | Not estimable |
| Barlow 2016 | -5.38 | 3.91 | 16 | -6.73 | 3.08 | 15 | 3.6% | 0.37 [-0.34, 1.08] |
| Barnes 2022 | 0 | 0 | 0 | 0 | 0 | 0 | | Not estimable |
| Berlin 2017 | 0 | 0 | 0 | 0 | 0 | 0 | | Not estimable |
| Berlin 2018 | 0 | 0 | 0 | 0 | 0 | 0 | | Not estimable |
| Bernard 2012 | 0 | 0 | 0 | 0 | 0 | 0 | | Not estimable |
| Betancourt 2020 | 0 | 0 | 0 | 0 | 0 | 0 | | Not estimable |
| Bick 2013 | 0 | 0 | 0 | 0 | 0 | 0 | | Not estimable |
| Borghini 2014 | 0 | 0 | 0 | 0 | 0 | 0 | | Not estimable |
| Brisch 2003 | 0 | 0 | 0 | 0 | 0 | 0 | | Not estimable |
| Browne 2005 | 0 | 0 | 0 | 0 | 0 | 0 | | Not estimable |
| Cates 2018 | 0 | 0 | 0 | 0 | 0 | 0 | | Not estimable |
| Cevasco 2008 | 0 | 0 | 0 | 0 | 0 | 0 | | Not estimable |
| Chiu 2009 | 0 | 0 | 0 | 0 | 0 | 0 | | Not estimable |
| Constantino 2001 | 0 | 0 | 0 | 0 | 0 | 0 | | Not estimable |
| Cooper 2009 | 0 | 0 | 0 | 0 | 0 | 0 | | Not estimable |
| Cooper 2015 | 0 | 0 | 0 | 0 | 0 | 0 | | Not estimable |
| Corrigan 2021 | 0 | 0 | 0 | 0 | 0 | 0 | | Not estimable |
| Dozier 2009 | 0 | 0 | 0 | 0 | 0 | 0 | | Not estimable |
| Eriksen 2018 | -5.69 | 4.66 | 13 | -10.54 | 5.36 | 13 | 2.8% | 0.94 [0.12, 1.75] |
| Feil 2020 | 0 | 0 | 0 | 0 | 0 | 0 | | Not estimable |
| Feldman 2014 | -31.47 | 6.22 | 73 | -34.95 | 8.64 | 73 | 12.3% | 0.46 [0.13, 0.79] |
| Firk 2021 | 0 | 0 | 0 | 0 | 0 | 0 | | Not estimable |
| Fonagy 2016 | 0 | 0 | 0 | 0 | 0 | 0 | | Not estimable |
| Gaden 2022 | -4.74 | 3.78 | 105 | -4.6 | 4.67 | 99 | 15.2% | -0.03 [-0.31, 0.24] |
| Georg 2021 | 0 | 0 | 0 | 0 | 0 | 0 | | Not estimable |
| Glazebrook 2007 | 0 | 0 | 0 | 0 | 0 | 0 | | Not estimable |
| Hane 2015 | -31.02 | 7.69 | 44 | -34.08 | 8.45 | 41 | 8.4% | 0.38 [-0.05, 0.81] |
| Heo 2019 | 0 | 0 | 0 | 0 | 0 | 0 | | Not estimable |
| Herbers 2020 | 0 | 0 | 0 | 0 | 0 | 0 | | Not estimable |
| Hoffenkamp 2015 | -32 | 10.57 | 75 | -31.8 | 10.91 | 75 | 12.7% | -0.02 [-0.34, 0.30] |
| Hoivik 2015 | 0 | 0 | 0 | 0 | 0 | 0 | | Not estimable |
| Holditch-Davis 2014 | 0 | 0 | 0 | 0 | 0 | 0 | | Not estimable |
| Holt 2021 | -8.06 | 6.03 | 27 | -7.38 | 7.25 | 29 | 6.1% | -0.10 [-0.62, 0.42] |
| Horowitz 2001 | 0 | 0 | 0 | 0 | 0 | 0 | | Not estimable |
| Horowitz 2013 | 0 | 0 | 0 | 0 | 0 | 0 | | Not estimable |
| Juffer 2005 | 0 | 0 | 0 | 0 | 0 | 0 | | Not estimable |
| Klein Velderman 2006 | 0 | 0 | 0 | 0 | 0 | 0 | | Not estimable |
| Knoche 2012 | 0 | 0 | 0 | 0 | 0 | 0 | | Not estimable |
| Kristensen 2020 | 0 | 0 | 0 | 0 | 0 | 0 | | Not estimable |
| Landry 2006 | 0 | 0 | 0 | 0 | 0 | 0 | | Not estimable |
| Lavallee 2022 | -35.1 | 9.11 | 10 | -42.87 | 15.2 | 10 | 2.4% | 0.59 [-0.31, 1.49] |
| Magil-Evans 2007 | 0 | 0 | 0 | 0 | 0 | 0 | | Not estimable |
| Meijssen 2010 | 0 | 0 | 0 | 0 | 0 | 0 | | Not estimable |
| Melnyk 2006 | -31.08 | 8.68 | 134 | -31.63 | 9.18 | 105 | 16.4% | 0.06 [-0.19, 0.32] |
| Mendelsohn 2007 | 0 | 0 | 0 | 0 | 0 | 0 | | Not estimable |
| Milgrom 2013 | 0 | 0 | 0 | 0 | 0 | 0 | | Not estimable |
| Nelson 2001 | 0 | 0 | 0 | 0 | 0 | 0 | | Not estimable |
| Neu 2010 | 0 | 0 | 0 | 0 | 0 | 0 | | Not estimable |
| Newnham 2009 | 0 | 0 | 0 | 0 | 0 | 0 | | Not estimable |
| Nugent 2017 | 0 | 0 | 0 | 0 | 0 | 0 | | Not estimable |
| Onozawa 2001 | 0 | 0 | 0 | 0 | 0 | 0 | | Not estimable |
| Oxford 2021 | 0 | 0 | 0 | 0 | 0 | 0 | | Not estimable |
| Perrone 2021 | 0 | 0 | 0 | 0 | 0 | 0 | | Not estimable |
| Pontoppidan 2016 | 0 | 0 | 0 | 0 | 0 | 0 | | Not estimable |
| Porter 2015 | 0 | 0 | 0 | 0 | 0 | 0 | | Not estimable |
| Ramsauer 2019 | 0 | 0 | 0 | 0 | 0 | 0 | | Not estimable |
| Ravn 2011 | 0 | 0 | 0 | 0 | 0 | 0 | | Not estimable |
| Rheinheimer 2022 | 0 | 0 | 0 | 0 | 0 | 0 | | Not estimable |
| Robertson 2019 | 0 | 0 | 0 | 0 | 0 | 0 | | Not estimable |
| Roby 2021 | 0 | 0 | 0 | 0 | 0 | 0 | | Not estimable |
| Sadler 2013 | 0 | 0 | 0 | 0 | 0 | 0 | | Not estimable |
| Sahlen Helmer 2020 | 0 | 0 | 0 | 0 | 0 | 0 | | Not estimable |
| Sajaniemi 2001 | 0 | 0 | 0 | 0 | 0 | 0 | | Not estimable |
| Salo 2019 | 0 | 0 | 0 | 0 | 0 | 0 | | Not estimable |
| Santelices 2010 | 0 | 0 | 0 | 0 | 0 | 0 | | Not estimable |
| Schuler 2000 | 0 | 0 | 0 | 0 | 0 | 0 | | Not estimable |
| Shoghi 2018 | 0 | 0 | 0 | 0 | 0 | 0 | | Not estimable |
| Sleed 2013 | 0 | 0 | 0 | 0 | 0 | 0 | | Not estimable |
| Steele 2019 | 0 | 0 | 0 | 0 | 0 | 0 | | Not estimable |
| Stein 2006 | 0 | 0 | 0 | 0 | 0 | 0 | | Not estimable |
| Stein 2018 | 0 | 0 | 0 | 0 | 0 | 0 | | Not estimable |
| Taneja 2020 | 0 | 0 | 0 | 0 | 0 | 0 | | Not estimable |
| Tereno 2017 | 0 | 0 | 0 | 0 | 0 | 0 | | Not estimable |
| Teti 2009 | 0 | 0 | 0 | 0 | 0 | 0 | | Not estimable |
| Tryphonopoulos 2020 | 0 | 0 | 0 | 0 | 0 | 0 | | Not estimable |
| Tsivos 2015 | 0 | 0 | 0 | 0 | 0 | 0 | | Not estimable |
| Twohig 2021 | 0 | 0 | 0 | 0 | 0 | 0 | | Not estimable |
| Valades 2021 | 0 | 0 | 0 | 0 | 0 | 0 | | Not estimable |
| van Doesum 2008 | 0 | 0 | 0 | 0 | 0 | 0 | | Not estimable |
| Vargas-Porras 2021 | 0 | 0 | 0 | 0 | 0 | 0 | | Not estimable |
| Walter 2019 | 0 | 0 | 0 | 0 | 0 | 0 | | Not estimable |
| White-Traut 2013 | 0 | 0 | 0 | 0 | 0 | 0 | | Not estimable |
| Williams 2020 | 0 | 0 | 0 | 0 | 0 | 0 | | Not estimable |
| Wulff 2021 | -33.36 | 6.71 | 56 | -33.44 | 5.92 | 57 | 10.5% | 0.01 [-0.36, 0.38] |
| Youn 2021 | 0 | 0 | 0 | 0 | 0 | 0 | | Not estimable |
| Yu 2022 | 0 | 0 | 0 | 0 | 0 | 0 | | Not estimable |
| Zelkowitz 2011 | -26.6 | 6.8 | 48 | -27.8 | 8.2 | 50 | 9.5% | 0.16 [-0.24, 0.55] |
| **Total (95% CI)** | | | **601** | | | **567** | **100.0%** | **0.16 [0.01, 0.30]** |

Heterogeneity: Tau² = 0.02; Chi² = 13.92, df = 10 (P = 0.18); I² = 28%
Test for overall effect: Z = 2.12 (P = 0.03)

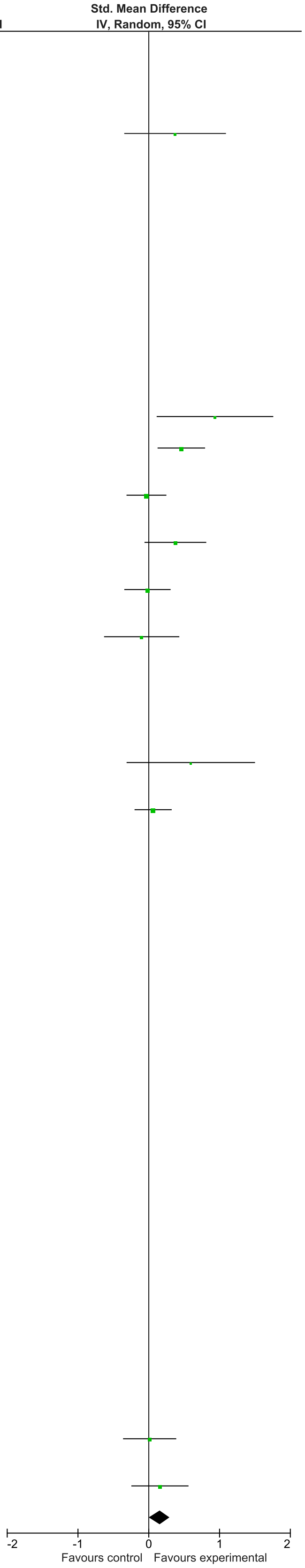

### eFigure 41. Parent/Caregiver Anxiety 2-6-month Follow-Up Forrest Plot

| | Early Dyadic Intervention | | | Standard Care | | | | Std. Mean Difference |
|---|---|---|---|---|---|---|---|---|
| Study or Subgroup | Mean | SD | Total | Mean | SD | Total | Weight | IV, Random, 95% CI |
| Akai 2008 | 0 | 0 | 0 | 0 | 0 | 0 | | Not estimable |
| Alarcao 2021 | 0 | 0 | 0 | 0 | 0 | 0 | | Not estimable |
| Alvarenga 2019 | 0 | 0 | 0 | 0 | 0 | 0 | | Not estimable |
| Badr 2006 | 0 | 0 | 0 | 0 | 0 | 0 | | Not estimable |
| Baggett 2010 | 0 | 0 | 0 | 0 | 0 | 0 | | Not estimable |
| Baggett 2017 | 0 | 0 | 0 | 0 | 0 | 0 | | Not estimable |
| Barlow 2016 | 0 | 0 | 0 | 0 | 0 | 0 | | Not estimable |
| Barnes 2022 | 0 | 0 | 0 | 0 | 0 | 0 | | Not estimable |
| Berlin 2017 | 0 | 0 | 0 | 0 | 0 | 0 | | Not estimable |
| Berlin 2018 | 0 | 0 | 0 | 0 | 0 | 0 | | Not estimable |
| Bernard 2012 | 0 | 0 | 0 | 0 | 0 | 0 | | Not estimable |
| Betancourt 2020 | 0 | 0 | 0 | 0 | 0 | 0 | | Not estimable |
| Bick 2013 | 0 | 0 | 0 | 0 | 0 | 0 | | Not estimable |
| Borghini 2014 | 0 | 0 | 0 | 0 | 0 | 0 | | Not estimable |
| Brisch 2003 | 0 | 0 | 0 | 0 | 0 | 0 | | Not estimable |
| Browne 2005 | 0 | 0 | 0 | 0 | 0 | 0 | | Not estimable |
| Cates 2018 | 0 | 0 | 0 | 0 | 0 | 0 | | Not estimable |
| Cevasco 2008 | 0 | 0 | 0 | 0 | 0 | 0 | | Not estimable |
| Chiu 2009 | 0 | 0 | 0 | 0 | 0 | 0 | | Not estimable |
| Constantino 2001 | 0 | 0 | 0 | 0 | 0 | 0 | | Not estimable |
| Cooper 2009 | 0 | 0 | 0 | 0 | 0 | 0 | | Not estimable |
| Cooper 2015 | 0 | 0 | 0 | 0 | 0 | 0 | | Not estimable |
| Corrigan 2021 | 0 | 0 | 0 | 0 | 0 | 0 | | Not estimable |
| Dozier 2009 | 0 | 0 | 0 | 0 | 0 | 0 | | Not estimable |
| Eriksen 2018 | 0 | 0 | 0 | 0 | 0 | 0 | | Not estimable |
| Feil 2020 | 0 | 0 | 0 | 0 | 0 | 0 | | Not estimable |
| Feldman 2014 | 0 | 0 | 0 | 0 | 0 | 0 | | Not estimable |
| Firk 2021 | 0 | 0 | 0 | 0 | 0 | 0 | | Not estimable |
| Fonagy 2016 | 0 | 0 | 0 | 0 | 0 | 0 | | Not estimable |
| Gaden 2022 | 0 | 0 | 0 | 0 | 0 | 0 | | Not estimable |
| Georg 2021 | 0 | 0 | 0 | 0 | 0 | 0 | | Not estimable |
| Glazebrook 2007 | 0 | 0 | 0 | 0 | 0 | 0 | | Not estimable |
| Hane 2015 | -26.18 | 4.8 | 42 | -32.18 | 6.26 | 40 | 24.0% | 1.07 [0.60, 1.53] |
| Heo 2019 | 0 | 0 | 0 | 0 | 0 | 0 | | Not estimable |
| Herbers 2020 | 0 | 0 | 0 | 0 | 0 | 0 | | Not estimable |
| Hoffenkamp 2015 | 0 | 0 | 0 | 0 | 0 | 0 | | Not estimable |
| Hoivik 2015 | 0 | 0 | 0 | 0 | 0 | 0 | | Not estimable |
| Holditch-Davis 2014 | 0 | 0 | 0 | 0 | 0 | 0 | | Not estimable |
| Holt 2021 | -10.09 | 8.71 | 22 | -7.59 | 7.47 | 27 | 21.4% | -0.31 [-0.87, 0.26] |
| Horowitz 2001 | 0 | 0 | 0 | 0 | 0 | 0 | | Not estimable |
| Horowitz 2013 | 0 | 0 | 0 | 0 | 0 | 0 | | Not estimable |
| Juffer 2005 | 0 | 0 | 0 | 0 | 0 | 0 | | Not estimable |
| Klein Velderman 2006 | 0 | 0 | 0 | 0 | 0 | 0 | | Not estimable |
| Knoche 2012 | 0 | 0 | 0 | 0 | 0 | 0 | | Not estimable |
| Kristensen 2020 | 0 | 0 | 0 | 0 | 0 | 0 | | Not estimable |
| Landry 2006 | 0 | 0 | 0 | 0 | 0 | 0 | | Not estimable |
| Lavallee 2022 | 0 | 0 | 0 | 0 | 0 | 0 | | Not estimable |
| Magil-Evans 2007 | 0 | 0 | 0 | 0 | 0 | 0 | | Not estimable |
| Meijssen 2010 | 0 | 0 | 0 | 0 | 0 | 0 | | Not estimable |
| Melnyk 2006 | -28.72 | 8.08 | 127 | -30.83 | 8.06 | 98 | 29.0% | 0.26 [-0.00, 0.53] |
| Mendelsohn 2007 | 0 | 0 | 0 | 0 | 0 | 0 | | Not estimable |
| Milgrom 2013 | 0 | 0 | 0 | 0 | 0 | 0 | | Not estimable |
| Nelson 2001 | 0 | 0 | 0 | 0 | 0 | 0 | | Not estimable |
| Neu 2010 | 0 | 0 | 0 | 0 | 0 | 0 | | Not estimable |
| Newnham 2009 | 0 | 0 | 0 | 0 | 0 | 0 | | Not estimable |
| Nugent 2017 | 0 | 0 | 0 | 0 | 0 | 0 | | Not estimable |
| Onozawa 2001 | 0 | 0 | 0 | 0 | 0 | 0 | | Not estimable |
| Oxford 2021 | 0 | 0 | 0 | 0 | 0 | 0 | | Not estimable |
| Perrone 2021 | 0 | 0 | 0 | 0 | 0 | 0 | | Not estimable |
| Pontoppidan 2016 | 0 | 0 | 0 | 0 | 0 | 0 | | Not estimable |
| Porter 2015 | 0 | 0 | 0 | 0 | 0 | 0 | | Not estimable |
| Ramsauer 2019 | 0 | 0 | 0 | 0 | 0 | 0 | | Not estimable |
| Ravn 2011 | 0 | 0 | 0 | 0 | 0 | 0 | | Not estimable |
| Rheinheimer 2022 | 0 | 0 | 0 | 0 | 0 | 0 | | Not estimable |
| Robertson 2019 | 0 | 0 | 0 | 0 | 0 | 0 | | Not estimable |
| Roby 2021 | 0 | 0 | 0 | 0 | 0 | 0 | | Not estimable |
| Sadler 2013 | 0 | 0 | 0 | 0 | 0 | 0 | | Not estimable |
| Sahlen Helmer 2020 | 0 | 0 | 0 | 0 | 0 | 0 | | Not estimable |
| Sajaniemi 2001 | 0 | 0 | 0 | 0 | 0 | 0 | | Not estimable |
| Salo 2019 | 0 | 0 | 0 | 0 | 0 | 0 | | Not estimable |
| Santelices 2010 | 0 | 0 | 0 | 0 | 0 | 0 | | Not estimable |
| Schuler 2000 | 0 | 0 | 0 | 0 | 0 | 0 | | Not estimable |
| Shoghi 2018 | 0 | 0 | 0 | 0 | 0 | 0 | | Not estimable |
| Sleed 2013 | 0 | 0 | 0 | 0 | 0 | 0 | | Not estimable |
| Steele 2019 | 0 | 0 | 0 | 0 | 0 | 0 | | Not estimable |
| Stein 2006 | 0 | 0 | 0 | 0 | 0 | 0 | | Not estimable |
| Stein 2018 | 0 | 0 | 0 | 0 | 0 | 0 | | Not estimable |
| Taneja 2020 | 0 | 0 | 0 | 0 | 0 | 0 | | Not estimable |
| Tereno 2017 | 0 | 0 | 0 | 0 | 0 | 0 | | Not estimable |
| Teti 2009 | 0 | 0 | 0 | 0 | 0 | 0 | | Not estimable |
| Tryphonopoulos 2020 | 0 | 0 | 0 | 0 | 0 | 0 | | Not estimable |
| Tsivos 2015 | 0 | 0 | 0 | 0 | 0 | 0 | | Not estimable |
| Twohig 2021 | 0 | 0 | 0 | 0 | 0 | 0 | | Not estimable |
| Valades 2021 | 0 | 0 | 0 | 0 | 0 | 0 | | Not estimable |
| van Doesum 2008 | 0 | 0 | 0 | 0 | 0 | 0 | | Not estimable |
| Vargas-Porras 2021 | 0 | 0 | 0 | 0 | 0 | 0 | | Not estimable |
| Walter 2019 | 0 | 0 | 0 | 0 | 0 | 0 | | Not estimable |
| White-Traut 2013 | 0 | 0 | 0 | 0 | 0 | 0 | | Not estimable |
| Williams 2020 | 0 | 0 | 0 | 0 | 0 | 0 | | Not estimable |
| Wulff 2021 | 0 | 0 | 0 | 0 | 0 | 0 | | Not estimable |
| Youn 2021 | 0 | 0 | 0 | 0 | 0 | 0 | | Not estimable |
| Yu 2022 | 0 | 0 | 0 | 0 | 0 | 0 | | Not estimable |
| Zelkowitz 2011 | -27.8 | 7.9 | 46 | -30.5 | 10 | 50 | 25.6% | 0.30 [-0.11, 0.70] |
| **Total (95% CI)** | | | **237** | | | **215** | **100.0%** | **0.34 [-0.11, 0.80]** |

Heterogeneity: Tau² = 0.17; Chi² = 14.85, df = 3 (P = 0.002); I² = 80%
Test for overall effect: Z = 1.48 (P = 0.14)

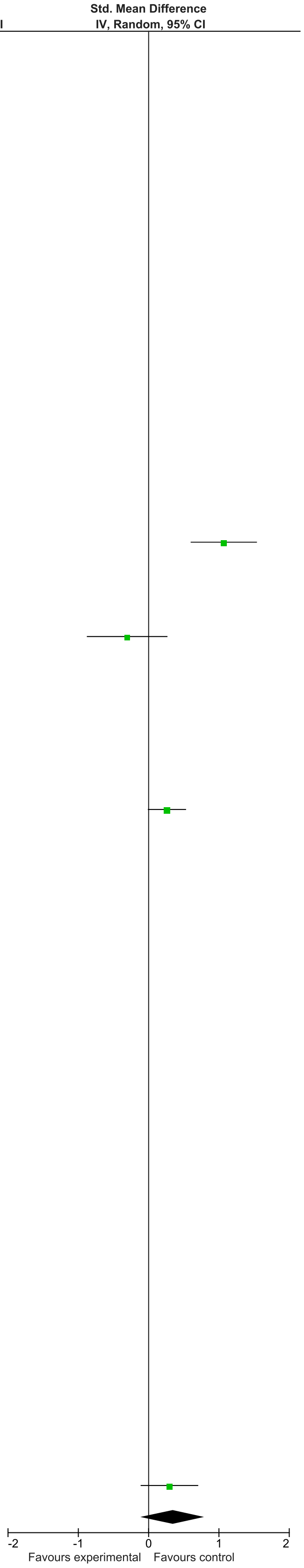

### eFigure 42. Parent/Caregiver Depression 0-1-month Follow-Up Forrest Plot

| | Early Dyadic Intervention | | | Standard Care | | | | Std. Mean Difference |
|---|---|---|---|---|---|---|---|---|
| Study or Subgroup | Mean | SD | Total | Mean | SD | Total | Weight | IV, Random, 95% CI |
| Akai 2008 | 0 | 0 | 0 | 0 | 0 | 0 | | Not estimable |
| Alarcao 2021 | 0 | 0 | 0 | 0 | 0 | 0 | | Not estimable |
| Alvarenga 2019 | 0 | 0 | 0 | 0 | 0 | 0 | | Not estimable |
| Badr 2006 | 0 | 0 | 0 | 0 | 0 | 0 | | Not estimable |
| Baggett 2010 | 0 | 0 | 0 | 0 | 0 | 0 | | Not estimable |
| Baggett 2017 | 0 | 0 | 0 | 0 | 0 | 0 | | Not estimable |
| Barlow 2016 | -3.13 | 3.03 | 16 | -4.07 | 2.69 | 15 | 2.4% | 0.32 [-0.39, 1.03] |
| Barnes 2022 | 0 | 0 | 0 | 0 | 0 | 0 | | Not estimable |
| Berlin 2017 | 0 | 0 | 0 | 0 | 0 | 0 | | Not estimable |
| Berlin 2018 | 0 | 0 | 0 | 0 | 0 | 0 | | Not estimable |
| Bernard 2012 | 0 | 0 | 0 | 0 | 0 | 0 | | Not estimable |
| Betancourt 2020 | 0 | 0 | 0 | 0 | 0 | 0 | | Not estimable |
| Bick 2013 | 0 | 0 | 0 | 0 | 0 | 0 | | Not estimable |
| Borghini 2014 | 0 | 0 | 0 | 0 | 0 | 0 | | Not estimable |
| Brisch 2003 | 0 | 0 | 0 | 0 | 0 | 0 | | Not estimable |
| Browne 2005 | 0 | 0 | 0 | 0 | 0 | 0 | | Not estimable |
| Cates 2018 | 0 | 0 | 0 | 0 | 0 | 0 | | Not estimable |
| Cevasco 2008 | 0 | 0 | 0 | 0 | 0 | 0 | | Not estimable |
| Chiu 2009 | 0 | 0 | 0 | 0 | 0 | 0 | | Not estimable |
| Constantino 2001 | 0 | 0 | 0 | 0 | 0 | 0 | | Not estimable |
| Cooper 2009 | -2.78 | 4.54 | 170 | -3.91 | 5.8 | 184 | 5.0% | 0.22 [0.01, 0.42] |
| Cooper 2015 | -6.9 | 4.97 | 80 | -6.7 | 4.53 | 79 | 4.5% | -0.04 [-0.35, 0.27] |
| Corrigan 2021 | 0 | 0 | 0 | 0 | 0 | 0 | | Not estimable |
| Dozier 2009 | 0 | 0 | 0 | 0 | 0 | 0 | | Not estimable |
| Eriksen 2018 | -7.69 | 6.69 | 13 | -15.23 | 11.04 | 13 | 2.1% | 0.80 [-0.00, 1.60] |
| Feil 2020 | 0 | 0 | 0 | 0 | 0 | 0 | | Not estimable |
| Feldman 2014 | -6.14 | 4.44 | 73 | -7.06 | 4.48 | 73 | 4.4% | 0.21 [-0.12, 0.53] |
| Firk 2021 | -10.11 | 8.5 | 27 | -11.05 | 9.4 | 20 | 3.0% | 0.10 [-0.47, 0.68] |
| Fonagy 2016 | -15.1 | 8.5 | 28 | -22.4 | 15 | 24 | 3.1% | 0.60 [0.04, 1.16] |
| Gaden 2022 | -6.02 | 4.45 | 104 | -6.2 | 5.21 | 101 | 4.7% | 0.04 [-0.24, 0.31] |
| Georg 2021 | -8.33 | 8.22 | 81 | -10.37 | 8.51 | 73 | 4.4% | 0.24 [-0.07, 0.56] |
| Glazebrook 2007 | 0 | 0 | 0 | 0 | 0 | 0 | | Not estimable |
| Hane 2015 | -7.7 | 5.84 | 43 | -9.63 | 6.21 | 41 | 3.8% | 0.32 [-0.11, 0.75] |
| Heo 2019 | 0 | 0 | 0 | 0 | 0 | 0 | | Not estimable |
| Herbers 2020 | 0 | 0 | 0 | 0 | 0 | 0 | | Not estimable |
| Hoffenkamp 2015 | -6.71 | 5.2 | 75 | -7.34 | 5.37 | 75 | 4.4% | 0.12 [-0.20, 0.44] |
| Hoivik 2015 | 0 | 0 | 0 | 0 | 0 | 0 | | Not estimable |
| Holditch-Davis 2014 | 0 | 0 | 0 | 0 | 0 | 0 | | Not estimable |
| Holt 2021 | -14.89 | 9.74 | 27 | 10.34 | 6.99 | 29 | 2.2% | -2.95 [-3.72, -2.18] |
| Horowitz 2001 | -10.27 | 0.99 | 60 | -9.51 | 0.77 | 57 | 4.1% | -0.85 [-1.23, -0.47] |
| Horowitz 2013 | -7.19 | 3.8 | 62 | -6.4 | 3.8 | 63 | 4.2% | -0.21 [-0.56, 0.14] |
| Juffer 2005 | 0 | 0 | 0 | 0 | 0 | 0 | | Not estimable |
| Klein Velderman 2006 | 0 | 0 | 0 | 0 | 0 | 0 | | Not estimable |
| Knoche 2012 | 0 | 0 | 0 | 0 | 0 | 0 | | Not estimable |
| Kristensen 2020 | 0 | 0 | 0 | 0 | 0 | 0 | | Not estimable |
| Landry 2006 | 0 | 0 | 0 | 0 | 0 | 0 | | Not estimable |
| Lavallee 2022 | 0 | 0 | 0 | 0 | 0 | 0 | | Not estimable |
| Magil-Evans 2007 | 0 | 0 | 0 | 0 | 0 | 0 | | Not estimable |
| Meijssen 2010 | 0 | 0 | 0 | 0 | 0 | 0 | | Not estimable |
| Melnyk 2006 | -7.05 | 5.77 | 132 | -7.51 | 5.78 | 105 | 4.8% | 0.08 [-0.18, 0.34] |
| Mendelsohn 2007 | -10.6 | 12 | 51 | -12.4 | 9.5 | 46 | 4.0% | 0.16 [-0.24, 0.56] |
| Milgrom 2013 | 0 | 0 | 0 | 0 | 0 | 0 | | Not estimable |
| Nelson 2001 | 0 | 0 | 0 | 0 | 0 | 0 | | Not estimable |
| Neu 2010 | 0 | 0 | 0 | 0 | 0 | 0 | | Not estimable |
| Newnham 2009 | -7.93 | 4.56 | 32 | -8.99 | 4.99 | 31 | 3.4% | 0.22 [-0.28, 0.71] |
| Nugent 2017 | 0 | 0 | 0 | 0 | 0 | 0 | | Not estimable |
| Onozawa 2001 | 0 | 0 | 0 | 0 | 0 | 0 | | Not estimable |
| Oxford 2021 | 0 | 0 | 0 | 0 | 0 | 0 | | Not estimable |
| Perrone 2021 | 0 | 0 | 0 | 0 | 0 | 0 | | Not estimable |
| Pontoppidan 2016 | 0 | 0 | 0 | 0 | 0 | 0 | | Not estimable |
| Porter 2015 | -6.14 | 8.2 | 59 | -8.5 | 7.89 | 34 | 3.8% | 0.29 [-0.13, 0.71] |
| Ramsauer 2019 | 0 | 0 | 0 | 0 | 0 | 0 | | Not estimable |
| Ravn 2011 | 0 | 0 | 0 | 0 | 0 | 0 | | Not estimable |
| Rheinheimer 2022 | 0 | 0 | 0 | 0 | 0 | 0 | | Not estimable |
| Robertson 2019 | 0 | 0 | 0 | 0 | 0 | 0 | | Not estimable |
| Roby 2021 | 0 | 0 | 0 | 0 | 0 | 0 | | Not estimable |
| Sadler 2013 | -12.8 | 10.4 | 44 | -10.8 | 8.9 | 31 | 3.6% | -0.20 [-0.66, 0.26] |
| Sahlen Helmer 2020 | 0 | 0 | 0 | 0 | 0 | 0 | | Not estimable |
| Sajaniemi 2001 | 0 | 0 | 0 | 0 | 0 | 0 | | Not estimable |
| Salo 2019 | 0 | 0 | 0 | 0 | 0 | 0 | | Not estimable |
| Santelices 2010 | 0 | 0 | 0 | 0 | 0 | 0 | | Not estimable |
| Schuler 2000 | 0 | 0 | 0 | 0 | 0 | 0 | | Not estimable |
| Shoghi 2018 | 0 | 0 | 0 | 0 | 0 | 0 | | Not estimable |
| Sleed 2013 | -13.6 | 9.4 | 62 | -15.3 | 11.8 | 53 | 4.1% | 0.16 [-0.21, 0.53] |
| Steele 2019 | 0 | 0 | 0 | 0 | 0 | 0 | | Not estimable |
| Stein 2006 | 0 | 0 | 0 | 0 | 0 | 0 | | Not estimable |
| Stein 2018 | 0 | 0 | 0 | 0 | 0 | 0 | | Not estimable |
| Taneja 2020 | -1.25 | 0.53 | 272 | -1.25 | 0.53 | 272 | 5.2% | 0.00 [-0.17, 0.17] |
| Tereno 2017 | 0 | 0 | 0 | 0 | 0 | 0 | | Not estimable |
| Teti 2009 | 0 | 0 | 0 | 0 | 0 | 0 | | Not estimable |
| Tryphonopoulos 2020 | -14 | 2.83 | 6 | -12.33 | 2.66 | 6 | 1.2% | -0.56 [-1.73, 0.60] |
| Tsivos 2015 | -9.39 | 9.35 | 12 | -11.23 | 10.71 | 9 | 1.9% | 0.18 [-0.69, 1.04] |
| Twohig 2021 | 0 | 0 | 0 | 0 | 0 | 0 | | Not estimable |
| Valades 2021 | 0 | 0 | 0 | 0 | 0 | 0 | | Not estimable |
| van Doesum 2008 | -18.9 | 11 | 35 | -17.2 | 9.3 | 36 | 3.6% | -0.17 [-0.63, 0.30] |
| Vargas-Porras 2021 | -6 | 3.03 | 33 | -5.64 | 2.54 | 33 | 3.5% | -0.13 [-0.61, 0.36] |
| Walter 2019 | 0 | 0 | 0 | 0 | 0 | 0 | | Not estimable |
| White-Traut 2013 | 0 | 0 | 0 | 0 | 0 | 0 | | Not estimable |
| Williams 2020 | 0 | 0 | 0 | 0 | 0 | 0 | | Not estimable |
| Wulff 2021 | -3.72 | 2.95 | 54 | -4.25 | 3.38 | 56 | 4.1% | 0.17 [-0.21, 0.54] |
| Youn 2021 | -7.5 | 6.3 | 69 | -8.8 | 6.8 | 67 | 4.3% | 0.20 [-0.14, 0.53] |
| Yu 2022 | 0 | 0 | 0 | 0 | 0 | 0 | | Not estimable |
| Zelkowitz 2011 | 0 | 0 | 0 | 0 | 0 | 0 | | Not estimable |
| **Total (95% CI)** | | | **1720** | | | **1626** | **100.0%** | **0.02 [-0.13, 0.16]** |

Heterogeneity: Tau² = 0.10; Chi² = 103.53, df = 26 (P < 0.00001); I² = 75%
Test for overall effect: Z = 0.20 (P = 0.84)

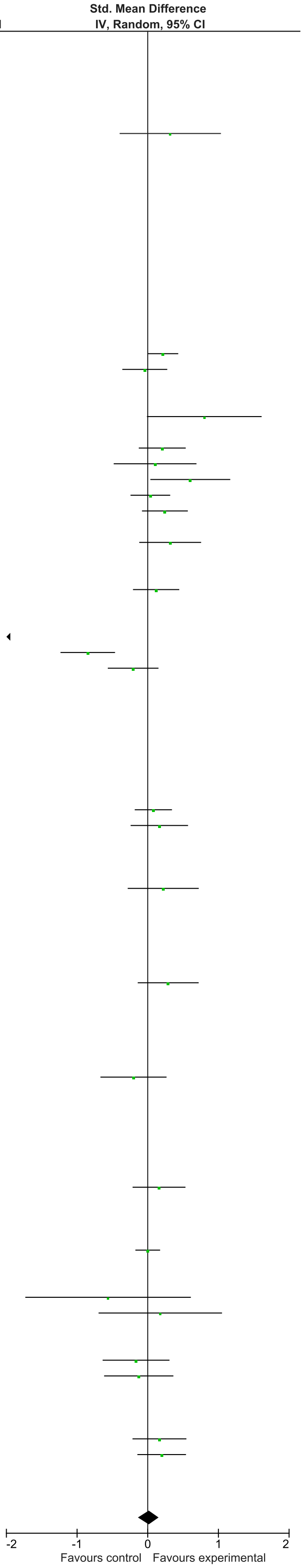

### eFigure 43. Parent/Caregiver Depression 2-6-month Follow-Up Forrest Plot

| Study or Subgroup | Early Dyadic Intervention Mean | SD | Total | Standard Care Mean | SD | Total | Weight | Std. Mean Difference IV, Random, 95% CI |
|---|---|---|---|---|---|---|---|---|
| Akai 2008 | 0 | 0 | 0 | 0 | 0 | 0 | | Not estimable |
| Alarcao 2021 | 0 | 0 | 0 | 0 | 0 | 0 | | Not estimable |
| Alvarenga 2019 | 0 | 0 | 0 | 0 | 0 | 0 | | Not estimable |
| Badr 2006 | 0 | 0 | 0 | 0 | 0 | 0 | | Not estimable |
| Baggett 2010 | 0 | 0 | 0 | 0 | 0 | 0 | | Not estimable |
| Baggett 2017 | 0 | 0 | 0 | 0 | 0 | 0 | | Not estimable |
| Barlow 2016 | 0 | 0 | 0 | 0 | 0 | 0 | | Not estimable |
| Barnes 2022 | 0 | 0 | 0 | 0 | 0 | 0 | | Not estimable |
| Berlin 2017 | 0 | 0 | 0 | 0 | 0 | 0 | | Not estimable |
| Berlin 2018 | 0 | 0 | 0 | 0 | 0 | 0 | | Not estimable |
| Bernard 2012 | 0 | 0 | 0 | 0 | 0 | 0 | | Not estimable |
| Betancourt 2020 | 0 | 0 | 0 | 0 | 0 | 0 | | Not estimable |
| Bick 2013 | 0 | 0 | 0 | 0 | 0 | 0 | | Not estimable |
| Borghini 2014 | 0 | 0 | 0 | 0 | 0 | 0 | | Not estimable |
| Brisch 2003 | 0 | 0 | 0 | 0 | 0 | 0 | | Not estimable |
| Browne 2005 | 0 | 0 | 0 | 0 | 0 | 0 | | Not estimable |
| Cates 2018 | 0 | 0 | 0 | 0 | 0 | 0 | | Not estimable |
| Cevasco 2008 | 0 | 0 | 0 | 0 | 0 | 0 | | Not estimable |
| Chiu 2009 | 0 | 0 | 0 | 0 | 0 | 0 | | Not estimable |
| Constantino 2001 | 0 | 0 | 0 | 0 | 0 | 0 | | Not estimable |
| Cooper 2009 | 0 | 0 | 0 | 0 | 0 | 0 | | Not estimable |
| Cooper 2015 | 0 | 0 | 0 | 0 | 0 | 0 | | Not estimable |
| Corrigan 2021 | 0 | 0 | 0 | 0 | 0 | 0 | | Not estimable |
| Dozier 2009 | 0 | 0 | 0 | 0 | 0 | 0 | | Not estimable |
| Eriksen 2018 | 0 | 0 | 0 | 0 | 0 | 0 | | Not estimable |
| Feil 2020 | 0 | 0 | 0 | 0 | 0 | 0 | | Not estimable |
| Feldman 2014 | 0 | 0 | 0 | 0 | 0 | 0 | | Not estimable |
| Firk 2021 | 0 | 0 | 0 | 0 | 0 | 0 | | Not estimable |
| Fonagy 2016 | 0 | 0 | 0 | 0 | 0 | 0 | | Not estimable |
| Gaden 2022 | 0 | 0 | 0 | 0 | 0 | 0 | | Not estimable |
| Georg 2021 | 0 | 0 | 0 | 0 | 0 | 0 | | Not estimable |
| Glazebrook 2007 | 0 | 0 | 0 | 0 | 0 | 0 | | Not estimable |
| Hane 2015 | -5.73 | 5.63 | 40 | -8.81 | 7.53 | 40 | 8.9% | 0.46 [0.01, 0.90] |
| Heo 2019 | 0 | 0 | 0 | 0 | 0 | 0 | | Not estimable |
| Herbers 2020 | 0 | 0 | 0 | 0 | 0 | 0 | | Not estimable |
| Hoffenkamp 2015 | 0 | 0 | 0 | 0 | 0 | 0 | | Not estimable |
| Hoivik 2015 | -15.36 | 8.66 | 22 | -12.19 | 7.79 | 27 | 7.0% | -0.38 [-0.95, 0.19] |
| Holditch-Davis 2014 | 0 | 0 | 0 | 0 | 0 | 0 | | Not estimable |
| Holt 2021 | -15.36 | 8.66 | 22 | -12.19 | 7.79 | 27 | 7.0% | -0.38 [-0.95, 0.19] |
| Horowitz 2001 | 0 | 0 | 0 | 0 | 0 | 0 | | Not estimable |
| Horowitz 2013 | 0 | 0 | 0 | 0 | 0 | 0 | | Not estimable |
| Juffer 2005 | 0 | 0 | 0 | 0 | 0 | 0 | | Not estimable |
| Klein Velderman 2006 | 0 | 0 | 0 | 0 | 0 | 0 | | Not estimable |
| Knoche 2012 | 0 | 0 | 0 | 0 | 0 | 0 | | Not estimable |
| Kristensen 2020 | 0 | 0 | 0 | 0 | 0 | 0 | | Not estimable |
| Landry 2006 | 0 | 0 | 0 | 0 | 0 | 0 | | Not estimable |
| Lavallee 2022 | 0 | 0 | 0 | 0 | 0 | 0 | | Not estimable |
| Magil-Evans 2007 | 0 | 0 | 0 | 0 | 0 | 0 | | Not estimable |
| Meijssen 2010 | 0 | 0 | 0 | 0 | 0 | 0 | | Not estimable |
| Melnyk 2006 | -5.56 | 5.13 | 126 | -7.21 | 5.13 | 98 | 12.2% | 0.32 [0.05, 0.59] |
| Mendelsohn 2007 | 0 | 0 | 0 | 0 | 0 | 0 | | Not estimable |
| Milgrom 2013 | 0 | 0 | 0 | 0 | 0 | 0 | | Not estimable |
| Nelson 2001 | 0 | 0 | 0 | 0 | 0 | 0 | | Not estimable |
| Neu 2010 | 0 | 0 | 0 | 0 | 0 | 0 | | Not estimable |
| Newnham 2009 | -8.1 | 5.4 | 32 | -9.7 | 4.99 | 31 | 8.0% | 0.30 [-0.19, 0.80] |
| Nugent 2017 | 0 | 0 | 0 | 0 | 0 | 0 | | Not estimable |
| Onozawa 2001 | 0 | 0 | 0 | 0 | 0 | 0 | | Not estimable |
| Oxford 2021 | 0 | 0 | 0 | 0 | 0 | 0 | | Not estimable |
| Perrone 2021 | -12.85 | 11.06 | 74 | -16.63 | 11.306 | 73 | 11.0% | 0.34 [0.01, 0.66] |
| Pontoppidan 2016 | 0 | 0 | 0 | 0 | 0 | 0 | | Not estimable |
| Porter 2015 | -3.16 | 5.31 | 51 | -9.74 | 9.71 | 35 | 8.8% | 0.88 [0.43, 1.33] |
| Ramsauer 2019 | -18.5 | 9.62 | 34 | -21.04 | 11.13 | 31 | 8.2% | 0.24 [-0.25, 0.73] |
| Ravn 2011 | 0 | 0 | 0 | 0 | 0 | 0 | | Not estimable |
| Rheinheimer 2022 | 0 | 0 | 0 | 0 | 0 | 0 | | Not estimable |
| Robertson 2019 | 0 | 0 | 0 | 0 | 0 | 0 | | Not estimable |
| Roby 2021 | 0 | 0 | 0 | 0 | 0 | 0 | | Not estimable |
| Sadler 2013 | 0 | 0 | 0 | 0 | 0 | 0 | | Not estimable |
| Sahlen Helmer 2020 | 0 | 0 | 0 | 0 | 0 | 0 | | Not estimable |
| Sajaniemi 2001 | 0 | 0 | 0 | 0 | 0 | 0 | | Not estimable |
| Salo 2019 | -8.16 | 2.99 | 24 | -9.65 | 3.92 | 21 | 6.7% | 0.42 [-0.17, 1.02] |
| Santelices 2010 | 0 | 0 | 0 | 0 | 0 | 0 | | Not estimable |
| Schuler 2000 | 0 | 0 | 0 | 0 | 0 | 0 | | Not estimable |
| Shoghi 2018 | 0 | 0 | 0 | 0 | 0 | 0 | | Not estimable |
| Sleed 2013 | 0 | 0 | 0 | 0 | 0 | 0 | | Not estimable |
| Steele 2019 | 0 | 0 | 0 | 0 | 0 | 0 | | Not estimable |
| Stein 2006 | 0 | 0 | 0 | 0 | 0 | 0 | | Not estimable |
| Stein 2018 | 0 | 0 | 0 | 0 | 0 | 0 | | Not estimable |
| Taneja 2020 | 0 | 0 | 0 | 0 | 0 | 0 | | Not estimable |
| Tereno 2017 | 0 | 0 | 0 | 0 | 0 | 0 | | Not estimable |
| Teti 2009 | 0 | 0 | 0 | 0 | 0 | 0 | | Not estimable |
| Tryphonopoulos 2020 | 0 | 0 | 0 | 0 | 0 | 0 | | Not estimable |
| Tsivos 2015 | -10.42 | 3.76 | 12 | -8.74 | 3.88 | 9 | 4.0% | -0.42 [-1.30, 0.45] |
| Twohig 2021 | 0 | 0 | 0 | 0 | 0 | 0 | | Not estimable |
| Valades 2021 | 0 | 0 | 0 | 0 | 0 | 0 | | Not estimable |
| van Doesum 2008 | -17.2 | 11.9 | 35 | -15.2 | 9.8 | 36 | 8.5% | -0.18 [-0.65, 0.28] |
| Vargas-Porras 2021 | 0 | 0 | 0 | 0 | 0 | 0 | | Not estimable |
| Walter 2019 | 0 | 0 | 0 | 0 | 0 | 0 | | Not estimable |
| White-Traut 2013 | 0 | 0 | 0 | 0 | 0 | 0 | | Not estimable |
| Williams 2020 | 0 | 0 | 0 | 0 | 0 | 0 | | Not estimable |
| Wulff 2021 | 0 | 0 | 0 | 0 | 0 | 0 | | Not estimable |
| Youn 2021 | 0 | 0 | 0 | 0 | 0 | 0 | | Not estimable |
| Yu 2022 | 0 | 0 | 0 | 0 | 0 | 0 | | Not estimable |
| Zelkowitz 2011 | -5.2 | 4.9 | 46 | -5 | 4.6 | 50 | 9.7% | -0.04 [-0.44, 0.36] |
| **Total (95% CI)** | | | **518** | | | **478** | **100.0%** | **0.18 [-0.03, 0.38]** |

Heterogeneity: Tau² = 0.07; Chi² = 26.10, df = 11 (P = 0.006); I² = 58%
Test for overall effect: Z = 1.70 (P = 0.09)

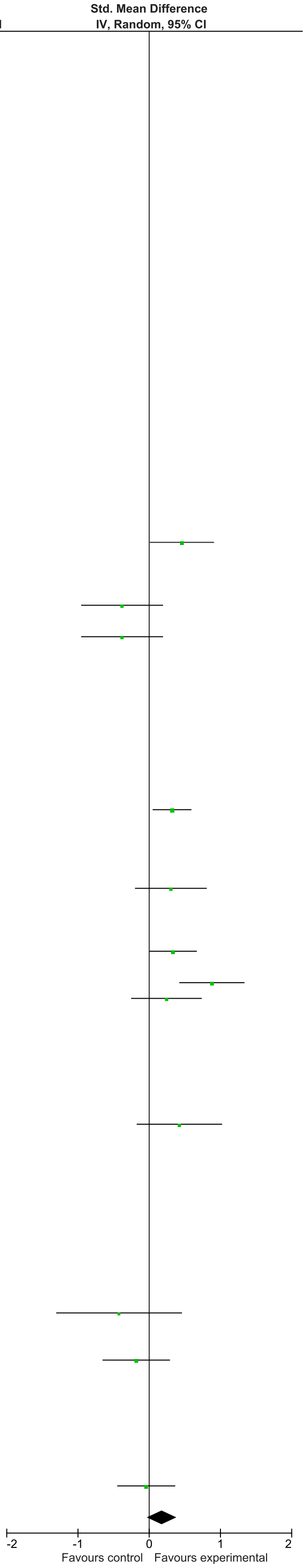

### eFigure 44. Parent/Caregiver Depression 7-12-month Follow-Up Forrest Plot

| | Early Dyadic Intervention | | | Standard Care | | | | Std. Mean Difference |
|---|---|---|---|---|---|---|---|---|
| Study or Subgroup | Mean | SD | Total | Mean | SD | Total | Weight | IV, Random, 95% CI |
| Akai 2008 | 0 | 0 | 0 | 0 | 0 | 0 | | Not estimable |
| Alarcao 2021 | 0 | 0 | 0 | 0 | 0 | 0 | | Not estimable |
| Alvarenga 2019 | 0 | 0 | 0 | 0 | 0 | 0 | | Not estimable |
| Badr 2006 | 0 | 0 | 0 | 0 | 0 | 0 | | Not estimable |
| Baggett 2010 | 0 | 0 | 0 | 0 | 0 | 0 | | Not estimable |
| Baggett 2017 | 0 | 0 | 0 | 0 | 0 | 0 | | Not estimable |
| Barlow 2016 | 0 | 0 | 0 | 0 | 0 | 0 | | Not estimable |
| Barnes 2022 | 0 | 0 | 0 | 0 | 0 | 0 | | Not estimable |
| Berlin 2017 | 0 | 0 | 0 | 0 | 0 | 0 | | Not estimable |
| Berlin 2018 | 0 | 0 | 0 | 0 | 0 | 0 | | Not estimable |
| Bernard 2012 | 0 | 0 | 0 | 0 | 0 | 0 | | Not estimable |
| Betancourt 2020 | 0 | 0 | 0 | 0 | 0 | 0 | | Not estimable |
| Bick 2013 | 0 | 0 | 0 | 0 | 0 | 0 | | Not estimable |
| Borghini 2014 | 0 | 0 | 0 | 0 | 0 | 0 | | Not estimable |
| Brisch 2003 | 0 | 0 | 0 | 0 | 0 | 0 | | Not estimable |
| Browne 2005 | 0 | 0 | 0 | 0 | 0 | 0 | | Not estimable |
| Cates 2018 | 0 | 0 | 0 | 0 | 0 | 0 | | Not estimable |
| Cevasco 2008 | 0 | 0 | 0 | 0 | 0 | 0 | | Not estimable |
| Chiu 2009 | 0 | 0 | 0 | 0 | 0 | 0 | | Not estimable |
| Constantino 2001 | 0 | 0 | 0 | 0 | 0 | 0 | | Not estimable |
| Cooper 2009 | -1.93 | 4.54 | 170 | -2.69 | 5.86 | 184 | 40.6% | 0.14 [-0.06, 0.35] |
| Cooper 2015 | -6.3 | 4.8 | 75 | -6.4 | 4.55 | 76 | 30.7% | 0.02 [-0.30, 0.34] |
| Corrigan 2021 | 0 | 0 | 0 | 0 | 0 | 0 | | Not estimable |
| Dozier 2009 | 0 | 0 | 0 | 0 | 0 | 0 | | Not estimable |
| Eriksen 2018 | 0 | 0 | 0 | 0 | 0 | 0 | | Not estimable |
| Feil 2020 | 0 | 0 | 0 | 0 | 0 | 0 | | Not estimable |
| Feldman 2014 | 0 | 0 | 0 | 0 | 0 | 0 | | Not estimable |
| Firk 2021 | 0 | 0 | 0 | 0 | 0 | 0 | | Not estimable |
| Fonagy 2016 | 0 | 0 | 0 | 0 | 0 | 0 | | Not estimable |
| Gaden 2022 | 0 | 0 | 0 | 0 | 0 | 0 | | Not estimable |
| Georg 2021 | 0 | 0 | 0 | 0 | 0 | 0 | | Not estimable |
| Glazebrook 2007 | 0 | 0 | 0 | 0 | 0 | 0 | | Not estimable |
| Hane 2015 | 0 | 0 | 0 | 0 | 0 | 0 | | Not estimable |
| Heo 2019 | 0 | 0 | 0 | 0 | 0 | 0 | | Not estimable |
| Herbers 2020 | 0 | 0 | 0 | 0 | 0 | 0 | | Not estimable |
| Hoffenkamp 2015 | 0 | 0 | 0 | 0 | 0 | 0 | | Not estimable |
| Hoivik 2015 | 0 | 0 | 0 | 0 | 0 | 0 | | Not estimable |
| Holditch-Davis 2014 | 0 | 0 | 0 | 0 | 0 | 0 | | Not estimable |
| Holt 2021 | 0 | 0 | 0 | 0 | 0 | 0 | | Not estimable |
| Horowitz 2001 | 0 | 0 | 0 | 0 | 0 | 0 | | Not estimable |
| Horowitz 2013 | 0 | 0 | 0 | 0 | 0 | 0 | | Not estimable |
| Juffer 2005 | 0 | 0 | 0 | 0 | 0 | 0 | | Not estimable |
| Klein Velderman 2006 | 0 | 0 | 0 | 0 | 0 | 0 | | Not estimable |
| Knoche 2012 | 0 | 0 | 0 | 0 | 0 | 0 | | Not estimable |
| Kristensen 2020 | 0 | 0 | 0 | 0 | 0 | 0 | | Not estimable |
| Landry 2006 | 0 | 0 | 0 | 0 | 0 | 0 | | Not estimable |
| Lavallee 2022 | 0 | 0 | 0 | 0 | 0 | 0 | | Not estimable |
| Magil-Evans 2007 | 0 | 0 | 0 | 0 | 0 | 0 | | Not estimable |
| Meijssen 2010 | 0 | 0 | 0 | 0 | 0 | 0 | | Not estimable |
| Melnyk 2006 | 0 | 0 | 0 | 0 | 0 | 0 | | Not estimable |
| Mendelsohn 2007 | 0 | 0 | 0 | 0 | 0 | 0 | | Not estimable |
| Milgrom 2013 | 0 | 0 | 0 | 0 | 0 | 0 | | Not estimable |
| Nelson 2001 | 0 | 0 | 0 | 0 | 0 | 0 | | Not estimable |
| Neu 2010 | 0 | 0 | 0 | 0 | 0 | 0 | | Not estimable |
| Newnham 2009 | 0 | 0 | 0 | 0 | 0 | 0 | | Not estimable |
| Nugent 2017 | 0 | 0 | 0 | 0 | 0 | 0 | | Not estimable |
| Onozawa 2001 | 0 | 0 | 0 | 0 | 0 | 0 | | Not estimable |
| Oxford 2021 | 0 | 0 | 0 | 0 | 0 | 0 | | Not estimable |
| Perrone 2021 | 0 | 0 | 0 | 0 | 0 | 0 | | Not estimable |
| Pontoppidan 2016 | 0 | 0 | 0 | 0 | 0 | 0 | | Not estimable |
| Porter 2015 | 0 | 0 | 0 | 0 | 0 | 0 | | Not estimable |
| Ramsauer 2019 | 0 | 0 | 0 | 0 | 0 | 0 | | Not estimable |
| Ravn 2011 | 0 | 0 | 0 | 0 | 0 | 0 | | Not estimable |
| Rheinheimer 2022 | 0 | 0 | 0 | 0 | 0 | 0 | | Not estimable |
| Robertson 2019 | 0 | 0 | 0 | 0 | 0 | 0 | | Not estimable |
| Roby 2021 | 0 | 0 | 0 | 0 | 0 | 0 | | Not estimable |
| Sadler 2013 | 0 | 0 | 0 | 0 | 0 | 0 | | Not estimable |
| Sahlen Helmer 2020 | 0 | 0 | 0 | 0 | 0 | 0 | | Not estimable |
| Sajaniemi 2001 | 0 | 0 | 0 | 0 | 0 | 0 | | Not estimable |
| Salo 2019 | 0 | 0 | 0 | 0 | 0 | 0 | | Not estimable |
| Santelices 2010 | 0 | 0 | 0 | 0 | 0 | 0 | | Not estimable |
| Schuler 2000 | 0 | 0 | 0 | 0 | 0 | 0 | | Not estimable |
| Shoghi 2018 | 0 | 0 | 0 | 0 | 0 | 0 | | Not estimable |
| Sleed 2013 | 0 | 0 | 0 | 0 | 0 | 0 | | Not estimable |
| Steele 2019 | 0 | 0 | 0 | 0 | 0 | 0 | | Not estimable |
| Stein 2006 | 0 | 0 | 0 | 0 | 0 | 0 | | Not estimable |
| Stein 2018 | -9.5 | 6.2 | 63 | -7.7 | 4.5 | 68 | 28.7% | -0.33 [-0.68, 0.01] |
| Taneja 2020 | 0 | 0 | 0 | 0 | 0 | 0 | | Not estimable |
| Tereno 2017 | 0 | 0 | 0 | 0 | 0 | 0 | | Not estimable |
| Teti 2009 | 0 | 0 | 0 | 0 | 0 | 0 | | Not estimable |
| Tryphonopoulos 2020 | 0 | 0 | 0 | 0 | 0 | 0 | | Not estimable |
| Tsivos 2015 | 0 | 0 | 0 | 0 | 0 | 0 | | Not estimable |
| Twohig 2021 | 0 | 0 | 0 | 0 | 0 | 0 | | Not estimable |
| Valades 2021 | 0 | 0 | 0 | 0 | 0 | 0 | | Not estimable |
| van Doesum 2008 | 0 | 0 | 0 | 0 | 0 | 0 | | Not estimable |
| Vargas-Porras 2021 | 0 | 0 | 0 | 0 | 0 | 0 | | Not estimable |
| Walter 2019 | 0 | 0 | 0 | 0 | 0 | 0 | | Not estimable |
| White-Traut 2013 | 0 | 0 | 0 | 0 | 0 | 0 | | Not estimable |
| Williams 2020 | 0 | 0 | 0 | 0 | 0 | 0 | | Not estimable |
| Wulff 2021 | 0 | 0 | 0 | 0 | 0 | 0 | | Not estimable |
| Youn 2021 | 0 | 0 | 0 | 0 | 0 | 0 | | Not estimable |
| Yu 2022 | 0 | 0 | 0 | 0 | 0 | 0 | | Not estimable |
| Zelkowitz 2011 | 0 | 0 | 0 | 0 | 0 | 0 | | Not estimable |
| **Total (95% CI)** | | | **308** | | | **328** | **100.0%** | **-0.03 [-0.30, 0.24]** |

Heterogeneity: Tau² = 0.04; Chi² = 5.36, df = 2 (P = 0.07); I² = 63%
Test for overall effect: Z = 0.22 (P = 0.83)

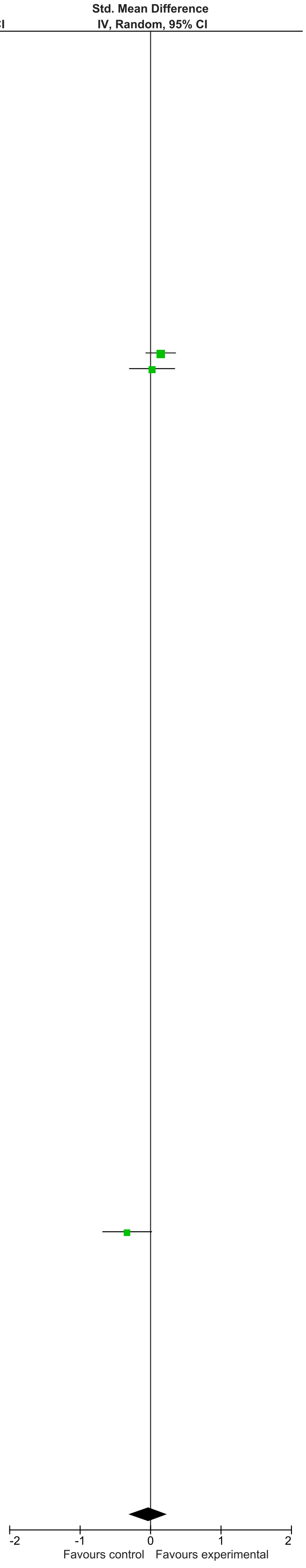

### eFigure 45. Parent/Caregiver Depression 13-24-month Follow-Up Forrest Plot

| | Early Dyadic Intervention | | | Standard Care | | | | Std. Mean Difference |
|---|---|---|---|---|---|---|---|---|
| Study or Subgroup | Mean | SD | Total | Mean | SD | Total | Weight | IV, Random, 95% CI |
| Akai 2008 | 0 | 0 | 0 | 0 | 0 | 0 | | Not estimable |
| Alarcao 2021 | 0 | 0 | 0 | 0 | 0 | 0 | | Not estimable |
| Alvarenga 2019 | 0 | 0 | 0 | 0 | 0 | 0 | | Not estimable |
| Badr 2006 | 0 | 0 | 0 | 0 | 0 | 0 | | Not estimable |
| Baggett 2010 | 0 | 0 | 0 | 0 | 0 | 0 | | Not estimable |
| Baggett 2017 | 0 | 0 | 0 | 0 | 0 | 0 | | Not estimable |
| Barlow 2016 | 0 | 0 | 0 | 0 | 0 | 0 | | Not estimable |
| Barnes 2022 | 0 | 0 | 0 | 0 | 0 | 0 | | Not estimable |
| Berlin 2017 | 0 | 0 | 0 | 0 | 0 | 0 | | Not estimable |
| Berlin 2018 | 0 | 0 | 0 | 0 | 0 | 0 | | Not estimable |
| Bernard 2012 | 0 | 0 | 0 | 0 | 0 | 0 | | Not estimable |
| Betancourt 2020 | 0 | 0 | 0 | 0 | 0 | 0 | | Not estimable |
| Bick 2013 | 0 | 0 | 0 | 0 | 0 | 0 | | Not estimable |
| Borghini 2014 | 0 | 0 | 0 | 0 | 0 | 0 | | Not estimable |
| Brisch 2003 | 0 | 0 | 0 | 0 | 0 | 0 | | Not estimable |
| Browne 2005 | 0 | 0 | 0 | 0 | 0 | 0 | | Not estimable |
| Cates 2018 | 0 | 0 | 0 | 0 | 0 | 0 | | Not estimable |
| Cevasco 2008 | 0 | 0 | 0 | 0 | 0 | 0 | | Not estimable |
| Chiu 2009 | 0 | 0 | 0 | 0 | 0 | 0 | | Not estimable |
| Constantino 2001 | 0 | 0 | 0 | 0 | 0 | 0 | | Not estimable |
| Cooper 2009 | 0 | 0 | 0 | 0 | 0 | 0 | | Not estimable |
| Cooper 2015 | -5.9 | 4.45 | 73 | -6.1 | 4.35 | 74 | 52.0% | 0.05 [-0.28, 0.37] |
| Corrigan 2021 | 0 | 0 | 0 | 0 | 0 | 0 | | Not estimable |
| Dozier 2009 | 0 | 0 | 0 | 0 | 0 | 0 | | Not estimable |
| Eriksen 2018 | 0 | 0 | 0 | 0 | 0 | 0 | | Not estimable |
| Feil 2020 | 0 | 0 | 0 | 0 | 0 | 0 | | Not estimable |
| Feldman 2014 | 0 | 0 | 0 | 0 | 0 | 0 | | Not estimable |
| Firk 2021 | 0 | 0 | 0 | 0 | 0 | 0 | | Not estimable |
| Fonagy 2016 | 0 | 0 | 0 | 0 | 0 | 0 | | Not estimable |
| Gaden 2022 | 0 | 0 | 0 | 0 | 0 | 0 | | Not estimable |
| Georg 2021 | 0 | 0 | 0 | 0 | 0 | 0 | | Not estimable |
| Glazebrook 2007 | 0 | 0 | 0 | 0 | 0 | 0 | | Not estimable |
| Hane 2015 | 0 | 0 | 0 | 0 | 0 | 0 | | Not estimable |
| Heo 2019 | 0 | 0 | 0 | 0 | 0 | 0 | | Not estimable |
| Herbers 2020 | 0 | 0 | 0 | 0 | 0 | 0 | | Not estimable |
| Hoffenkamp 2015 | 0 | 0 | 0 | 0 | 0 | 0 | | Not estimable |
| Hoivik 2015 | 0 | 0 | 0 | 0 | 0 | 0 | | Not estimable |
| Holditch-Davis 2014 | 0 | 0 | 0 | 0 | 0 | 0 | | Not estimable |
| Holt 2021 | 0 | 0 | 0 | 0 | 0 | 0 | | Not estimable |
| Horowitz 2001 | 0 | 0 | 0 | 0 | 0 | 0 | | Not estimable |
| Horowitz 2013 | 0 | 0 | 0 | 0 | 0 | 0 | | Not estimable |
| Juffer 2005 | 0 | 0 | 0 | 0 | 0 | 0 | | Not estimable |
| Klein Velderman 2006 | 0 | 0 | 0 | 0 | 0 | 0 | | Not estimable |
| Knoche 2012 | 0 | 0 | 0 | 0 | 0 | 0 | | Not estimable |
| Kristensen 2020 | 0 | 0 | 0 | 0 | 0 | 0 | | Not estimable |
| Landry 2006 | 0 | 0 | 0 | 0 | 0 | 0 | | Not estimable |
| Lavallee 2022 | 0 | 0 | 0 | 0 | 0 | 0 | | Not estimable |
| Magil-Evans 2007 | 0 | 0 | 0 | 0 | 0 | 0 | | Not estimable |
| Meijssen 2010 | 0 | 0 | 0 | 0 | 0 | 0 | | Not estimable |
| Melnyk 2006 | 0 | 0 | 0 | 0 | 0 | 0 | | Not estimable |
| Mendelsohn 2007 | 0 | 0 | 0 | 0 | 0 | 0 | | Not estimable |
| Milgrom 2013 | 0 | 0 | 0 | 0 | 0 | 0 | | Not estimable |
| Nelson 2001 | 0 | 0 | 0 | 0 | 0 | 0 | | Not estimable |
| Neu 2010 | 0 | 0 | 0 | 0 | 0 | 0 | | Not estimable |
| Newnham 2009 | 0 | 0 | 0 | 0 | 0 | 0 | | Not estimable |
| Nugent 2017 | 0 | 0 | 0 | 0 | 0 | 0 | | Not estimable |
| Onozawa 2001 | 0 | 0 | 0 | 0 | 0 | 0 | | Not estimable |
| Oxford 2021 | 0 | 0 | 0 | 0 | 0 | 0 | | Not estimable |
| Perrone 2021 | 0 | 0 | 0 | 0 | 0 | 0 | | Not estimable |
| Pontoppidan 2016 | 0 | 0 | 0 | 0 | 0 | 0 | | Not estimable |
| Porter 2015 | 0 | 0 | 0 | 0 | 0 | 0 | | Not estimable |
| Ramsauer 2019 | 0 | 0 | 0 | 0 | 0 | 0 | | Not estimable |
| Ravn 2011 | 0 | 0 | 0 | 0 | 0 | 0 | | Not estimable |
| Rheinheimer 2022 | 0 | 0 | 0 | 0 | 0 | 0 | | Not estimable |
| Robertson 2019 | 0 | 0 | 0 | 0 | 0 | 0 | | Not estimable |
| Roby 2021 | 0 | 0 | 0 | 0 | 0 | 0 | | Not estimable |
| Sadler 2013 | 0 | 0 | 0 | 0 | 0 | 0 | | Not estimable |
| Sahlen Helmer 2020 | 0 | 0 | 0 | 0 | 0 | 0 | | Not estimable |
| Sajaniemi 2001 | 0 | 0 | 0 | 0 | 0 | 0 | | Not estimable |
| Salo 2019 | 0 | 0 | 0 | 0 | 0 | 0 | | Not estimable |
| Santelices 2010 | 0 | 0 | 0 | 0 | 0 | 0 | | Not estimable |
| Schuler 2000 | 0 | 0 | 0 | 0 | 0 | 0 | | Not estimable |
| Shoghi 2018 | 0 | 0 | 0 | 0 | 0 | 0 | | Not estimable |
| Sleed 2013 | 0 | 0 | 0 | 0 | 0 | 0 | | Not estimable |
| Steele 2019 | 0 | 0 | 0 | 0 | 0 | 0 | | Not estimable |
| Stein 2006 | 0 | 0 | 0 | 0 | 0 | 0 | | Not estimable |
| Stein 2018 | 0 | 0 | 0 | 0 | 0 | 0 | | Not estimable |
| Taneja 2020 | 0 | 0 | 0 | 0 | 0 | 0 | | Not estimable |
| Tereno 2017 | 0 | 0 | 0 | 0 | 0 | 0 | | Not estimable |
| Teti 2009 | 0 | 0 | 0 | 0 | 0 | 0 | | Not estimable |
| Tryphonopoulos 2020 | 0 | 0 | 0 | 0 | 0 | 0 | | Not estimable |
| Tsivos 2015 | 0 | 0 | 0 | 0 | 0 | 0 | | Not estimable |
| Twohig 2021 | 0 | 0 | 0 | 0 | 0 | 0 | | Not estimable |
| Valades 2021 | 0 | 0 | 0 | 0 | 0 | 0 | | Not estimable |
| van Doesum 2008 | 0 | 0 | 0 | 0 | 0 | 0 | | Not estimable |
| Vargas-Porras 2021 | 0 | 0 | 0 | 0 | 0 | 0 | | Not estimable |
| Walter 2019 | 0 | 0 | 0 | 0 | 0 | 0 | | Not estimable |
| White-Traut 2013 | 0 | 0 | 0 | 0 | 0 | 0 | | Not estimable |
| Williams 2020 | 0 | 0 | 0 | 0 | 0 | 0 | | Not estimable |
| Wulff 2021 | 0 | 0 | 0 | 0 | 0 | 0 | | Not estimable |
| Youn 2021 | -9.3 | 7.9 | 69 | -8.1 | 7.1 | 67 | 48.0% | -0.16 [-0.50, 0.18] |
| Yu 2022 | 0 | 0 | 0 | 0 | 0 | 0 | | Not estimable |
| Zelkowitz 2011 | 0 | 0 | 0 | 0 | 0 | 0 | | Not estimable |
| **Total (95% CI)** | | | **142** | | | **141** | **100.0%** | **-0.05 [-0.29, 0.18]** |

Heterogeneity: Tau² = 0.00; Chi² = 0.73, df = 1 (P = 0.39); I² = 0%

Test for overall effect: Z = 0.44 (P = 0.66)

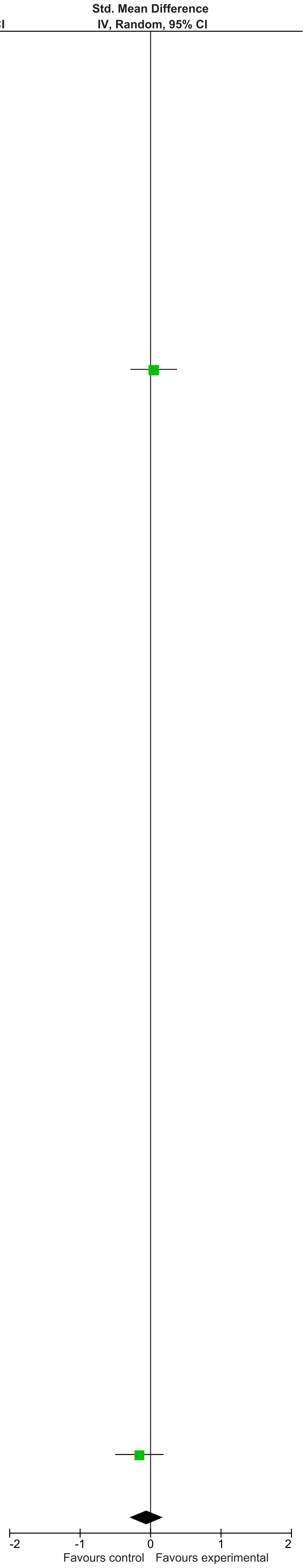

#### eFigure 46. Overview of Meta-Analysis Results of Subset of 20 Studies

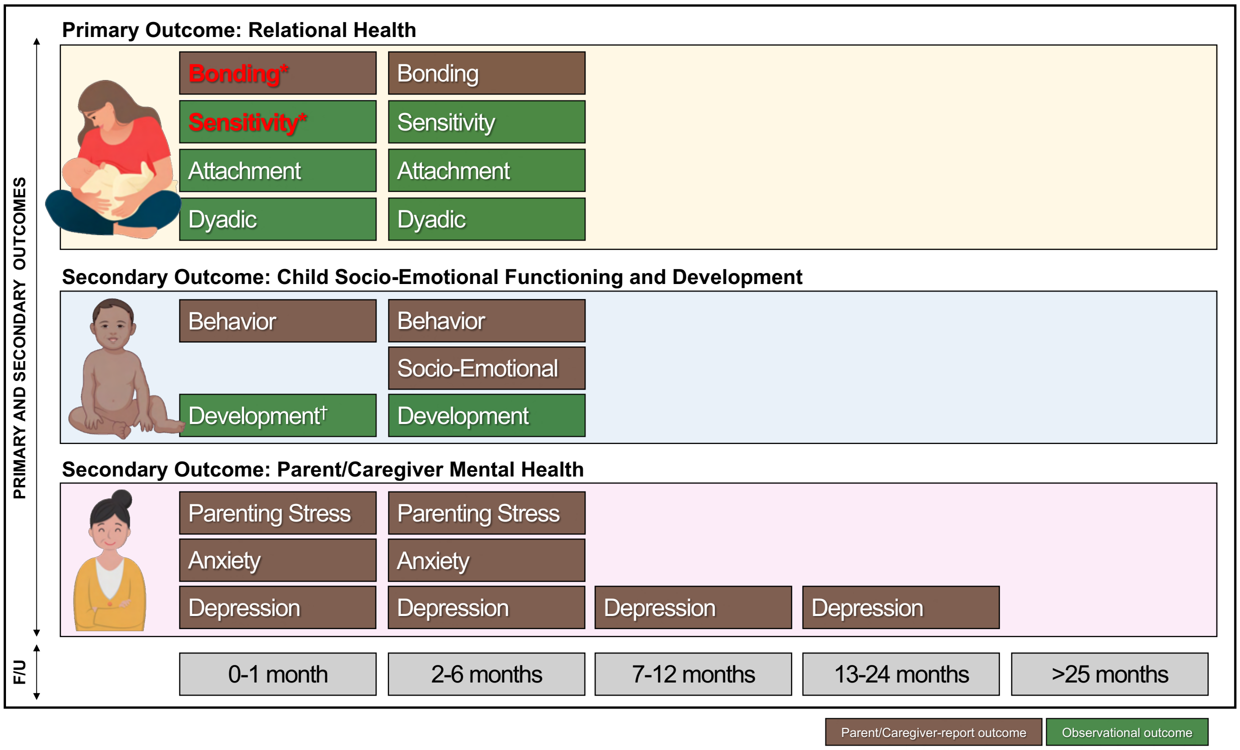


Top: Meta-analytic results of subset of 20 studies. Bottom: Timeline indicating follow-up (F/U) timing; **Bold/Red text**=significant effect; *Small effect size.

**eFigure 47. RoB (RCT) – Bonding (0-1 month F/U)**

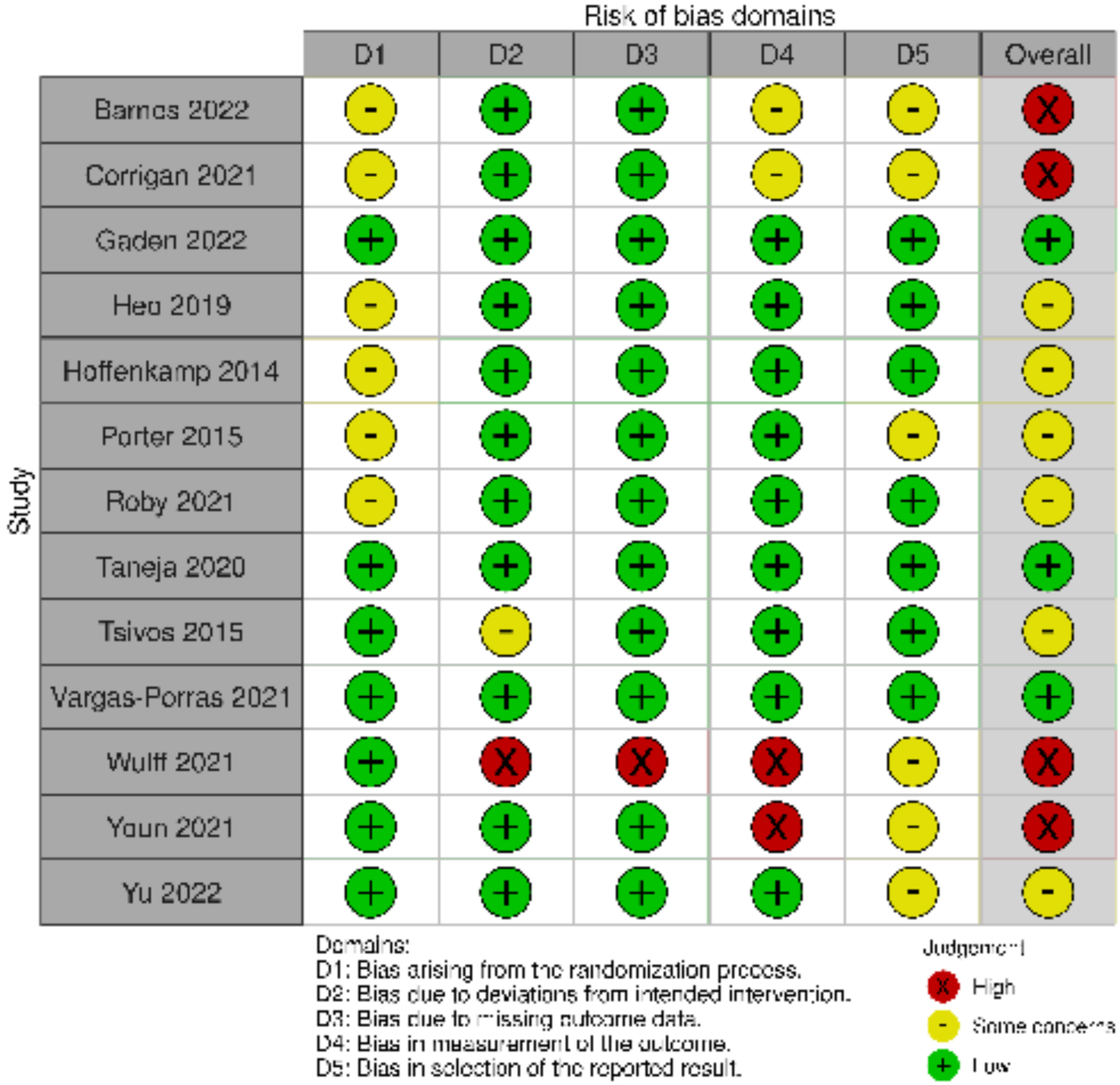

**eFigure 48. RoB (RCT) – Bonding (2-6 months F/U)**

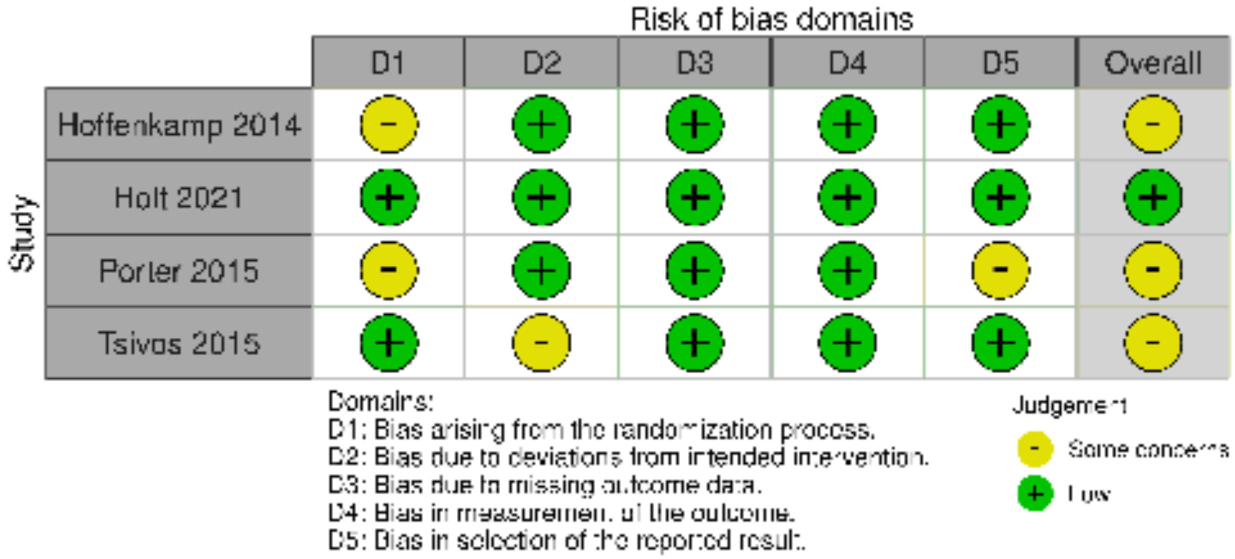

#### eFigure 49. RoB (RCT) – Sensitivity (0-1 months F/U)

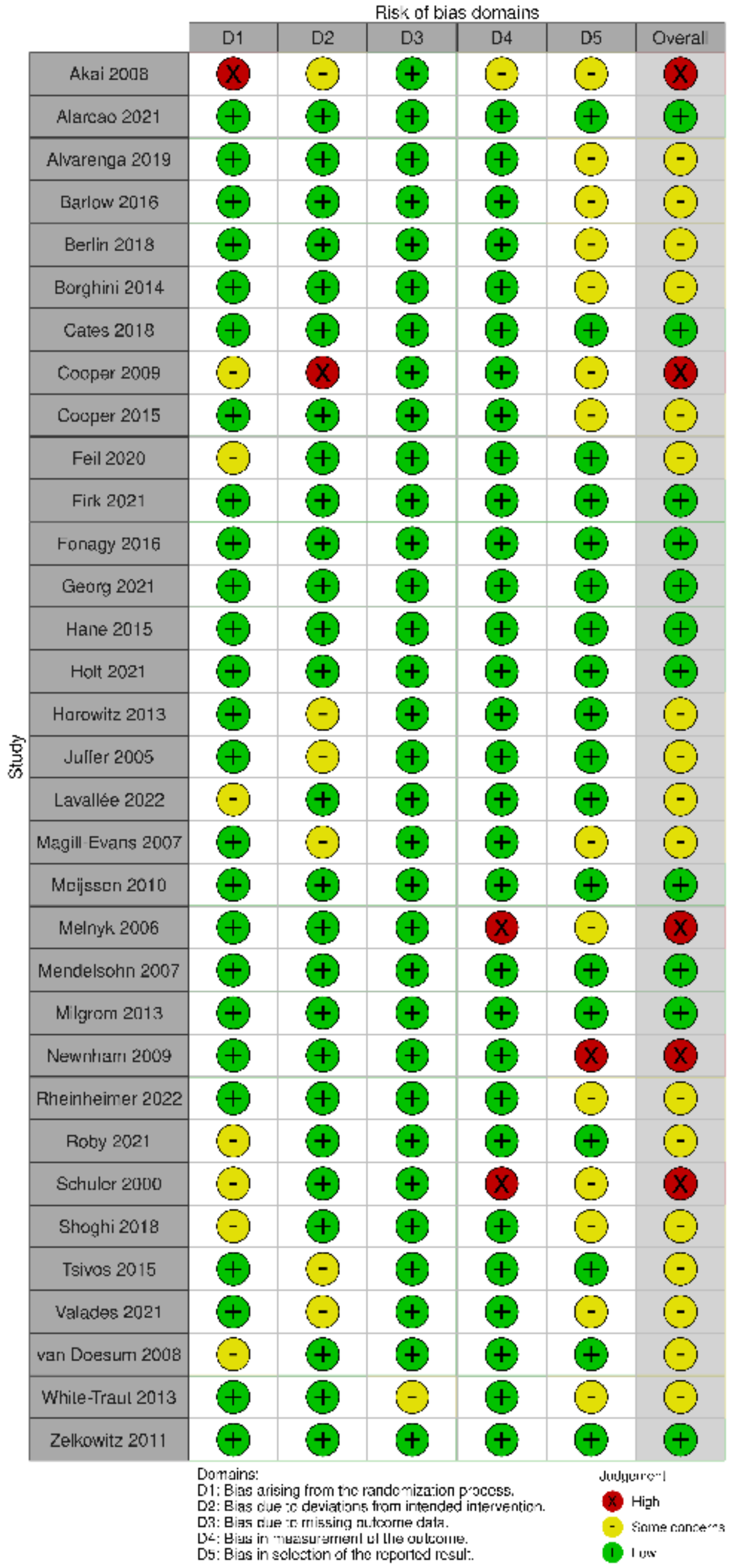

**eFigure 50. RoB (RCT) – Sensitivity (2-6 months F/U)**

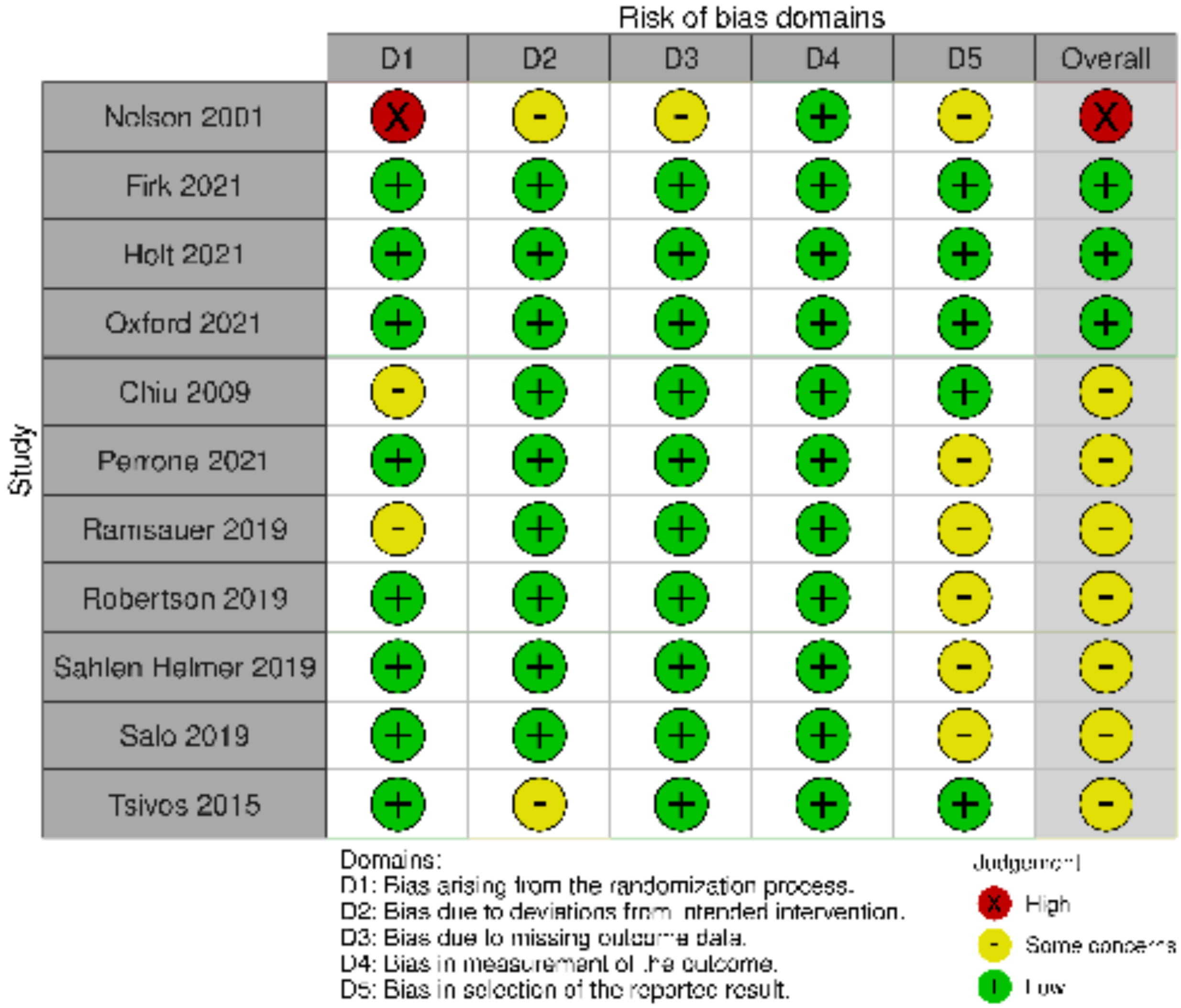


**eFigure 51. RoB (cluster RCT) – Sensitivity (2-6 months F/U)**

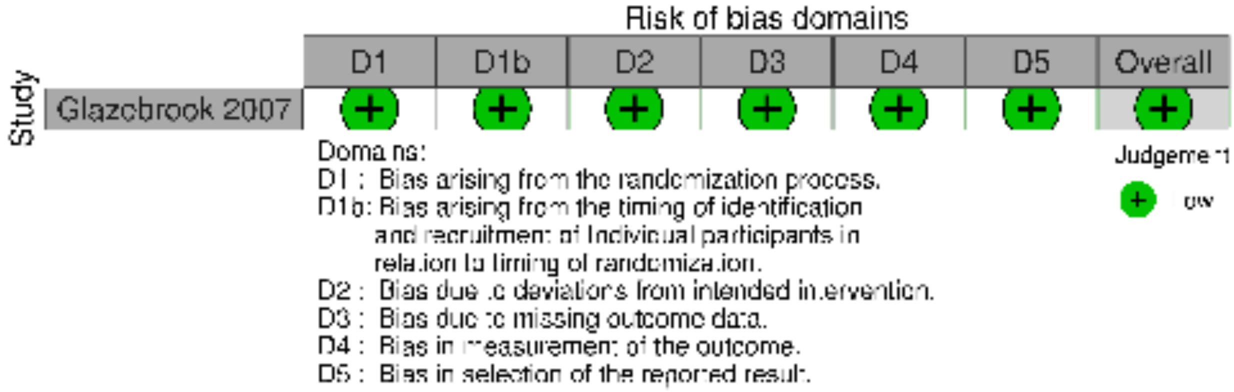

**eFigure 52. RoB (RCT) – Sensitivity (7-12 months F/U)**

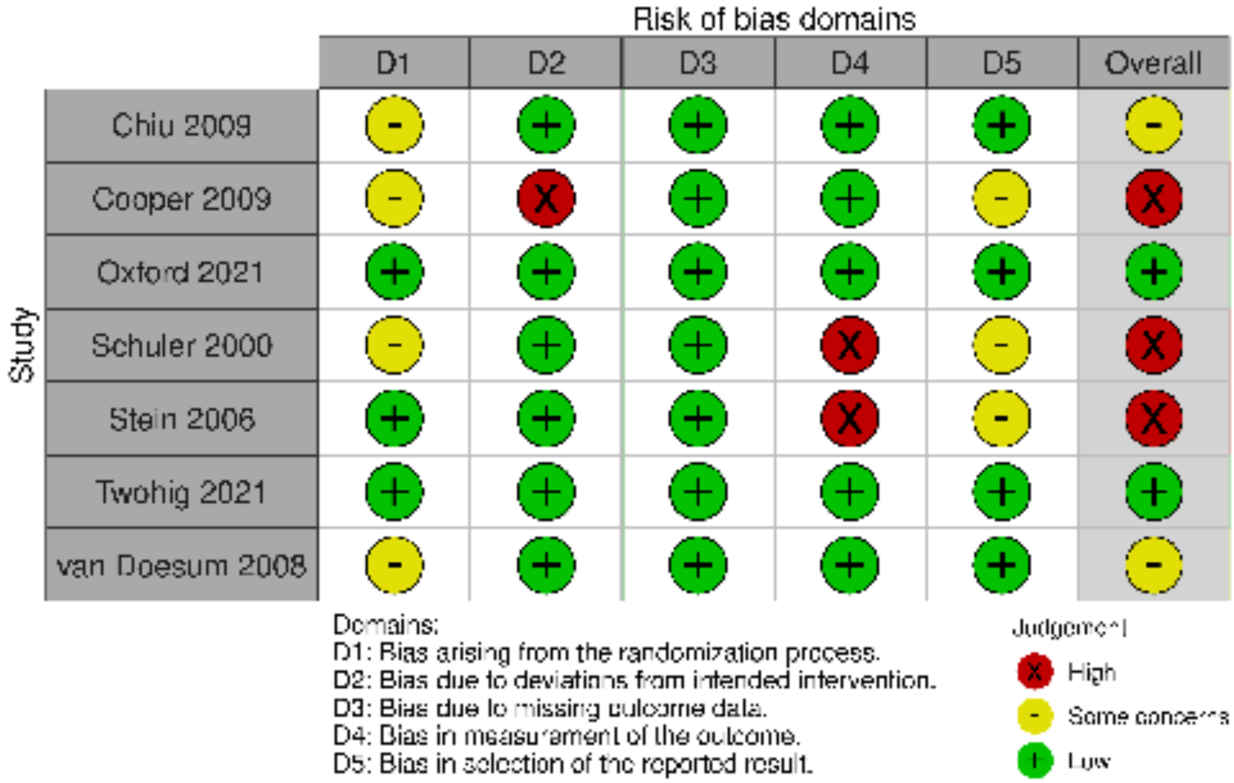


**eFigure 53. RoB (RCT) – Sensitivity (13-24 months F/U)**

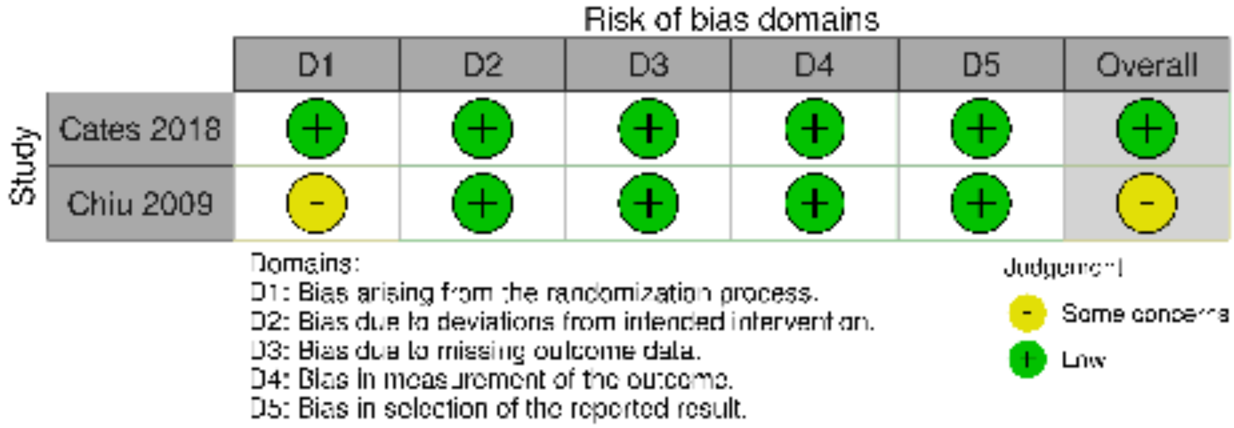

#### eFigure 54. RoB (RCT) – Secure Attachment (0-1-month F/U)

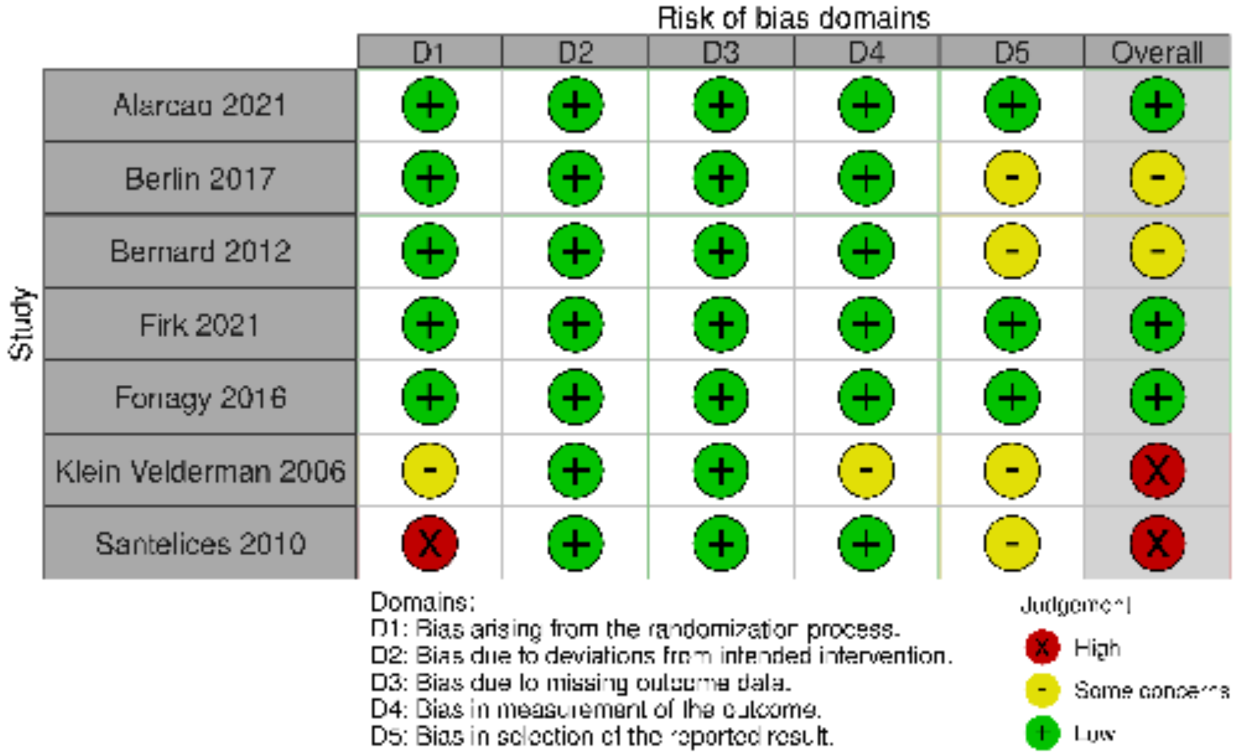


#### eFigure 55. RoB (cluster RCT) – Secure Attachment (0-1-month F/U)

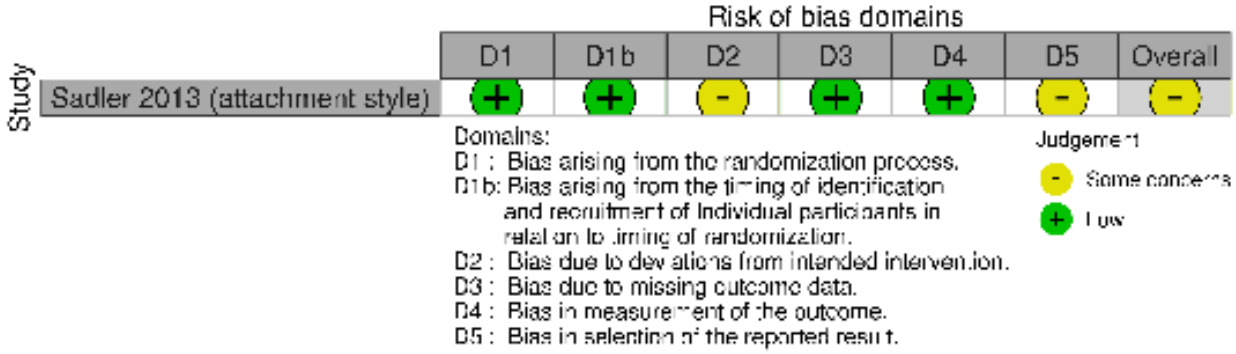

**eFigure 56. RoB (RCT) – Secure Attachment (2-6 months F/U)**

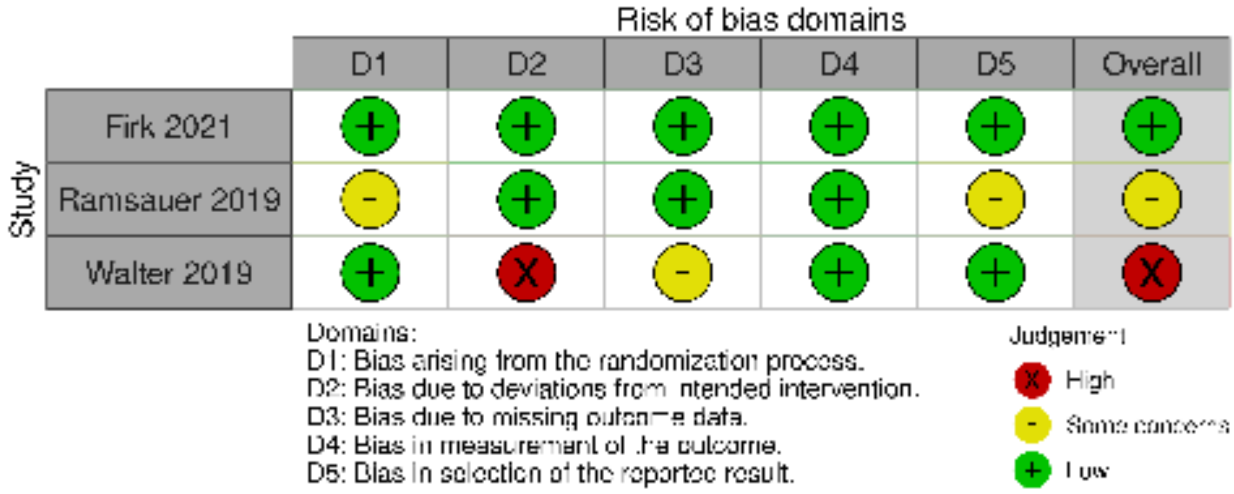


**eFigure 57. RoB (RCT) – Secure Attachment (13-24-month F/U)**

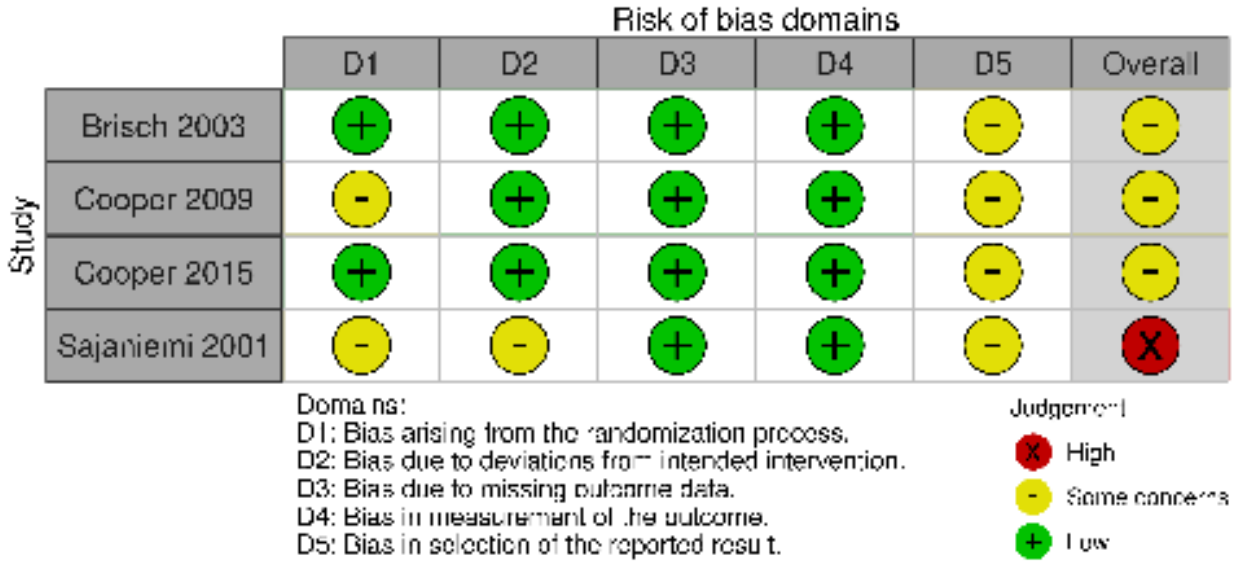

#### eFigure 58. RoB (RCT) – Organized Attachment (0-1-month F/U)

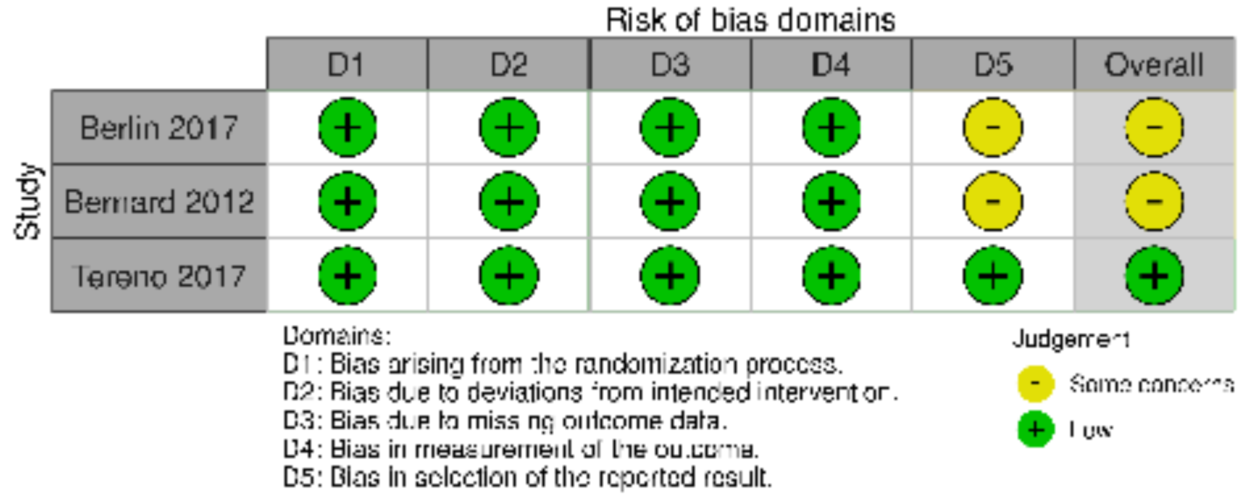


#### eFigure 59. RoB (RCT) – Insecure Attachment (0-1-month F/U)

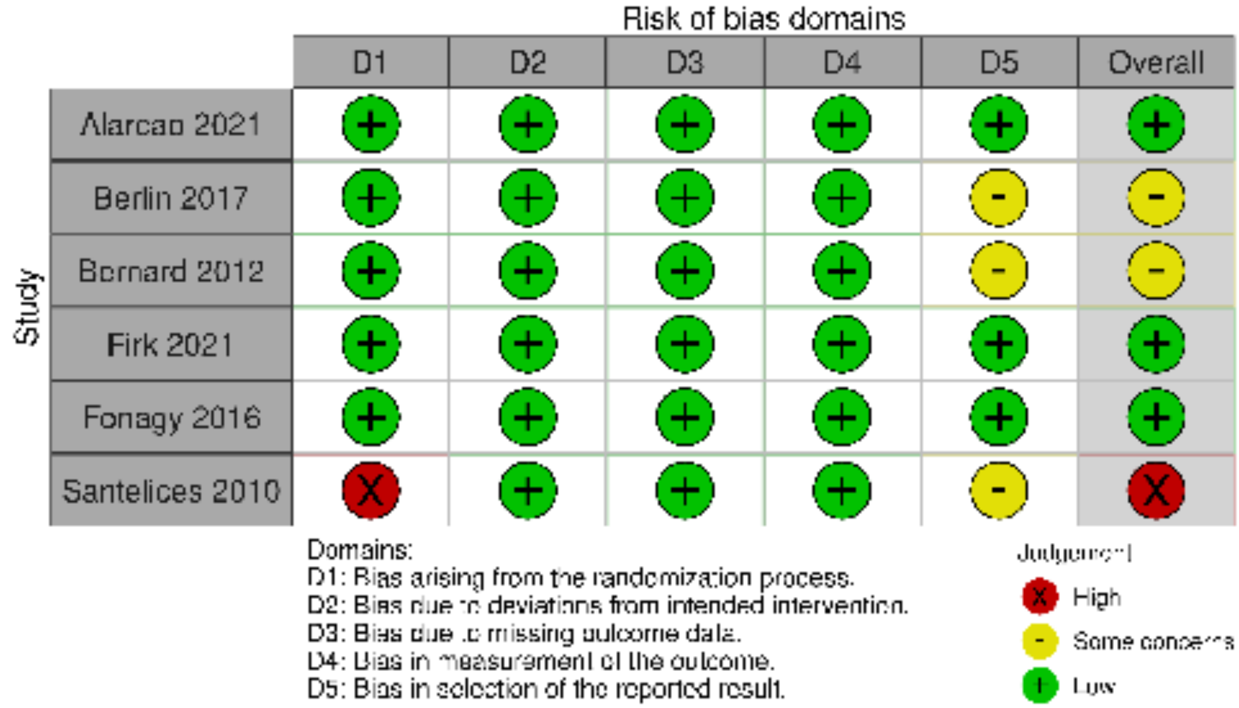

#### eFigure 60. RoB (RCT) – Insecure Attachment (2-6 months F/U)

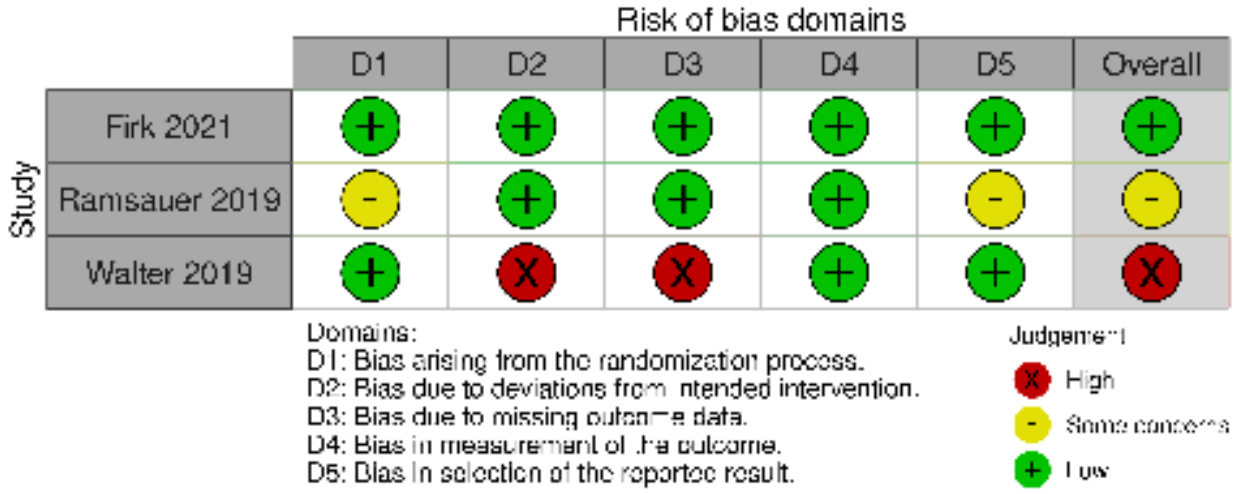


#### eFigure 61. RoB (RCT) – Insecure Attachment (13-24 months F/U)

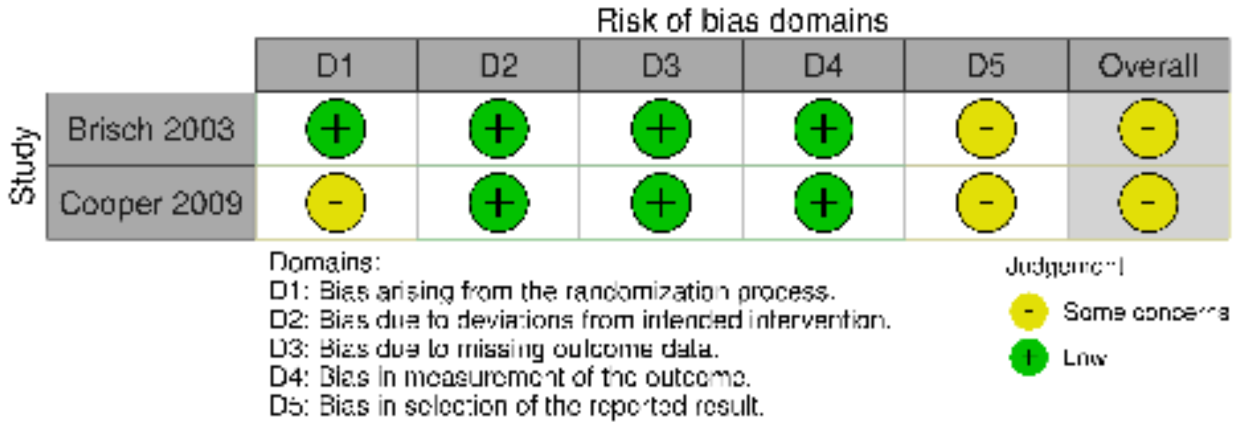

**eFigure 62. RoB (RCT) – Disorganized Attachment (0-1-month F/U)**

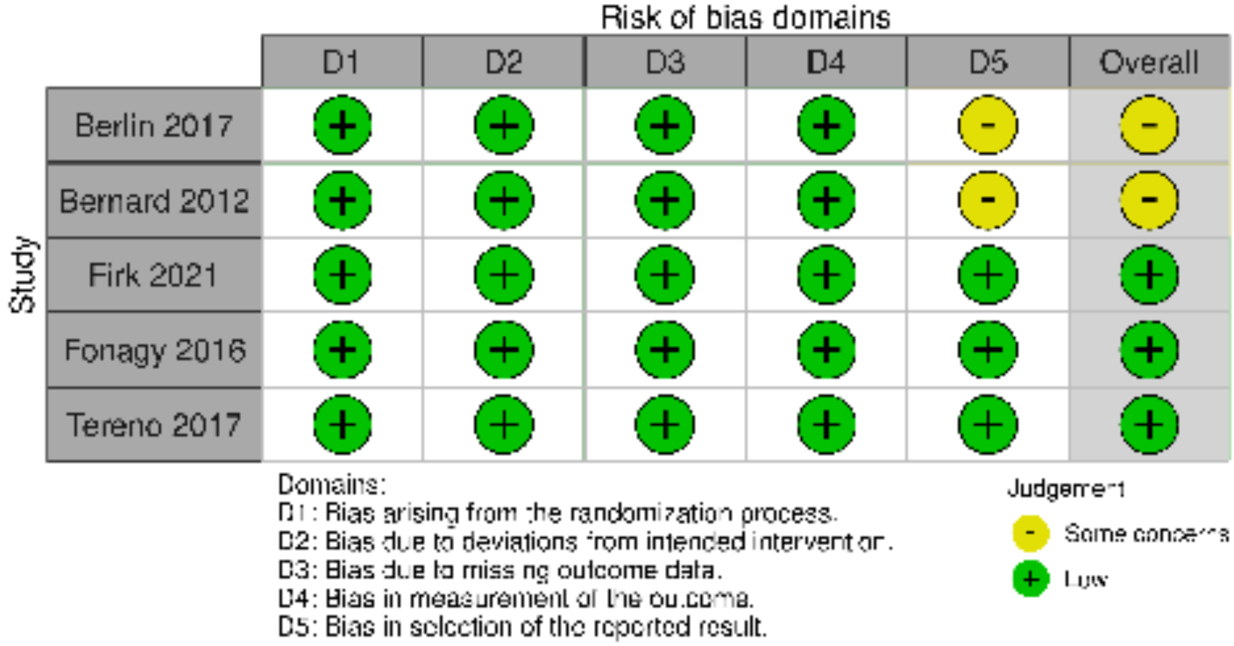


**eFigure 63. RoB (cluster RCT) – Disorganized Attachment (0-1 month F/U)**

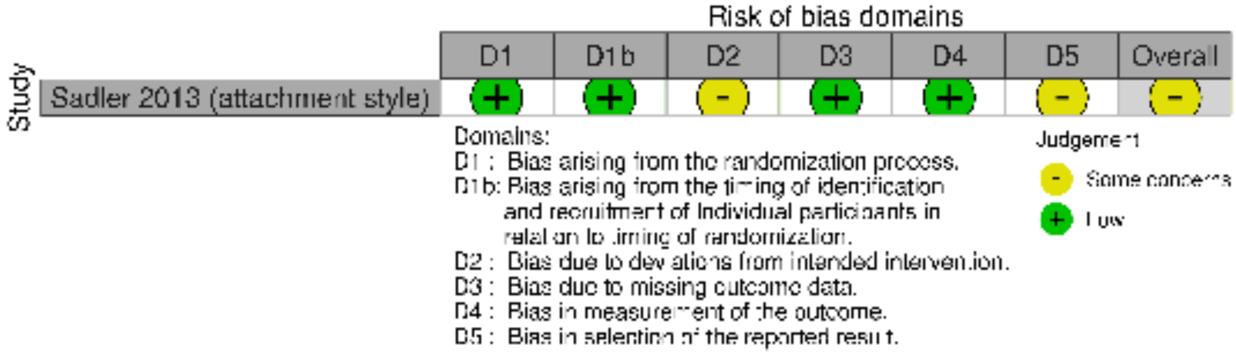

**eFigure 64. RoB (RCT) – Disorganized Attachment (2-6 months F/U)**

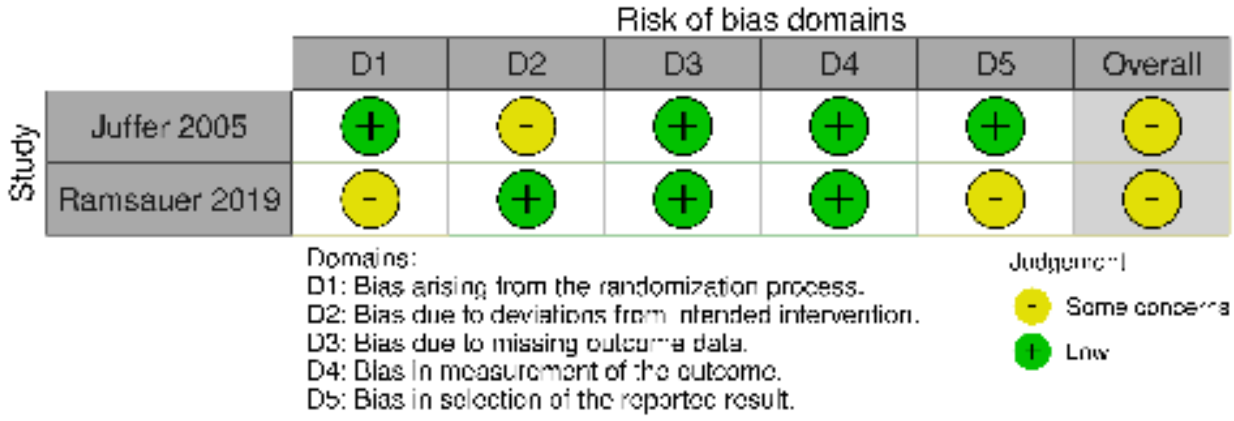

#### eFigure 65. RoB (RCT) – Dyadic Interactions (0-1 month F/U)

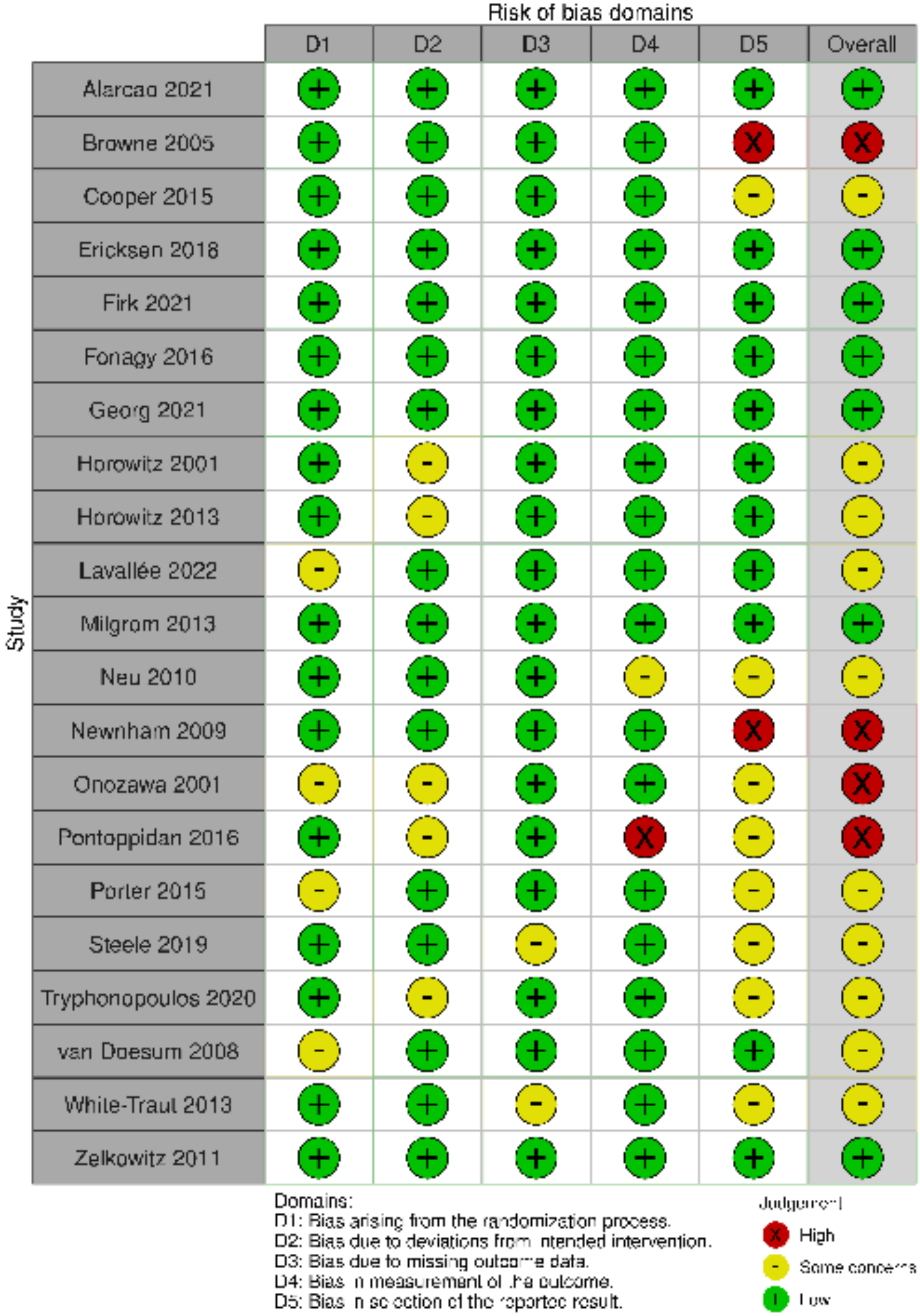

**eFigure 66. RoB (RCT) – Dyadic Interactions (2-6 months F/U)**

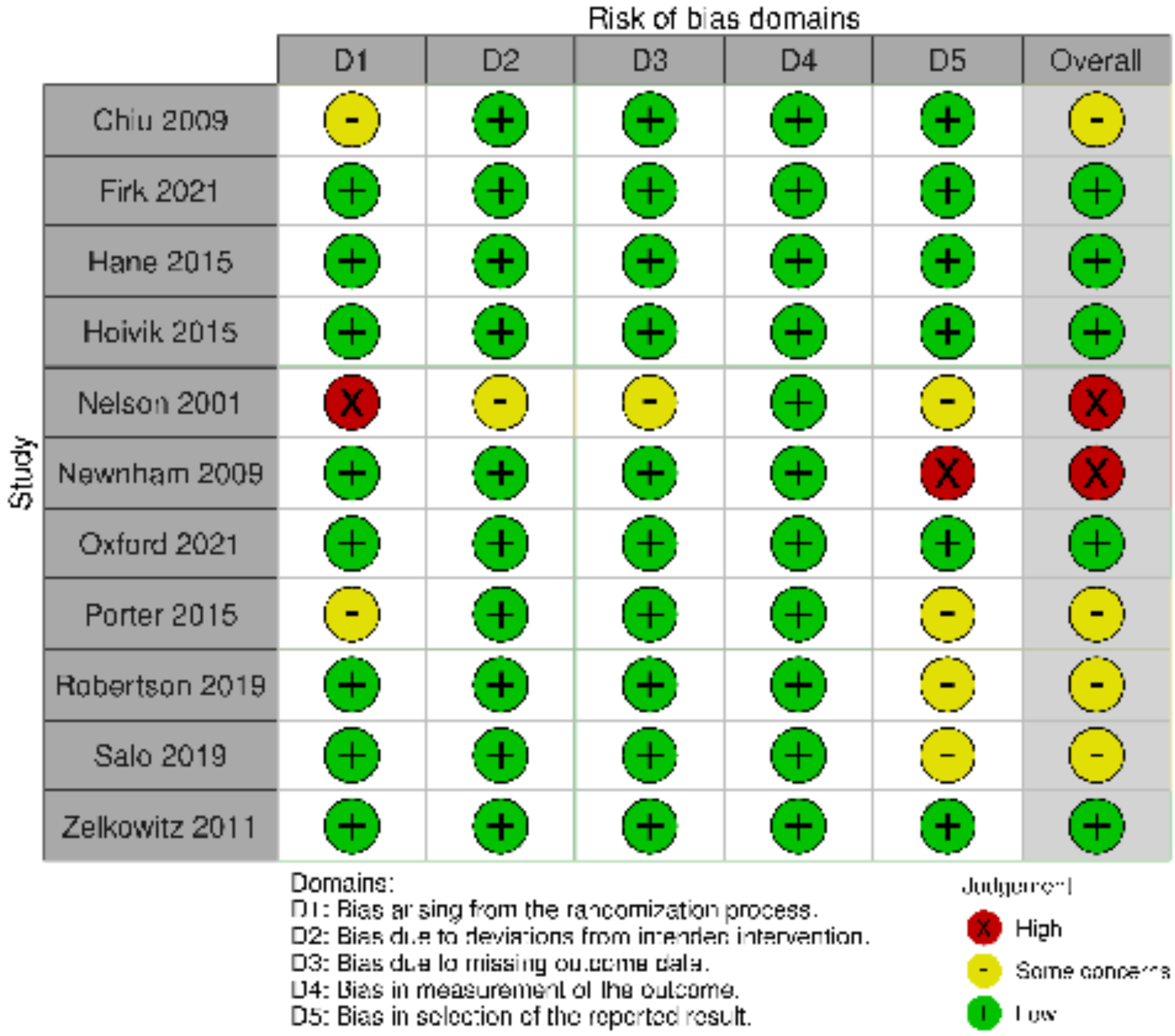

**eFigure 67. RoB (RCT) – Dyadic Interactions (7-12 months F/U)**

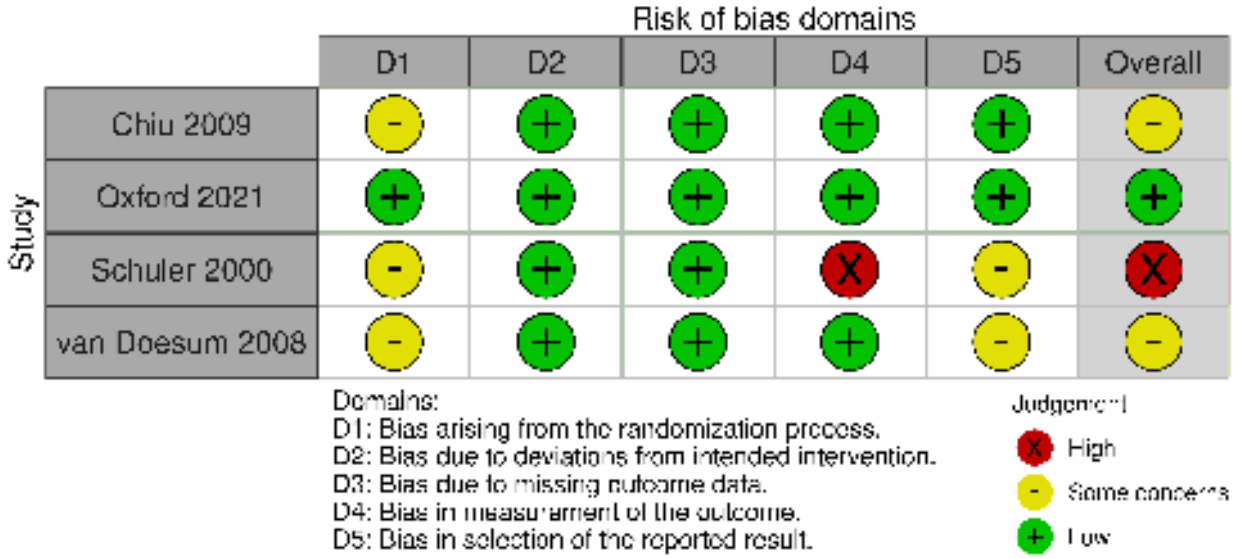


**eFigure 68. RoB (RCT) – Cognitive Development (0-1 month F/U)**

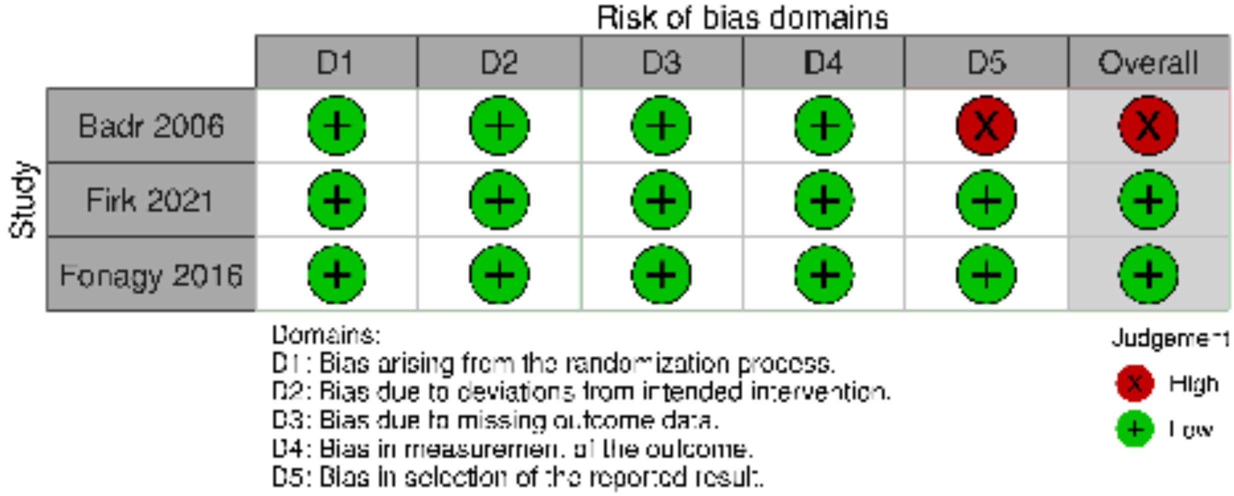

**eFigure 69. RoB (RCT) – Cognitive Development (2-6 months F/U)**

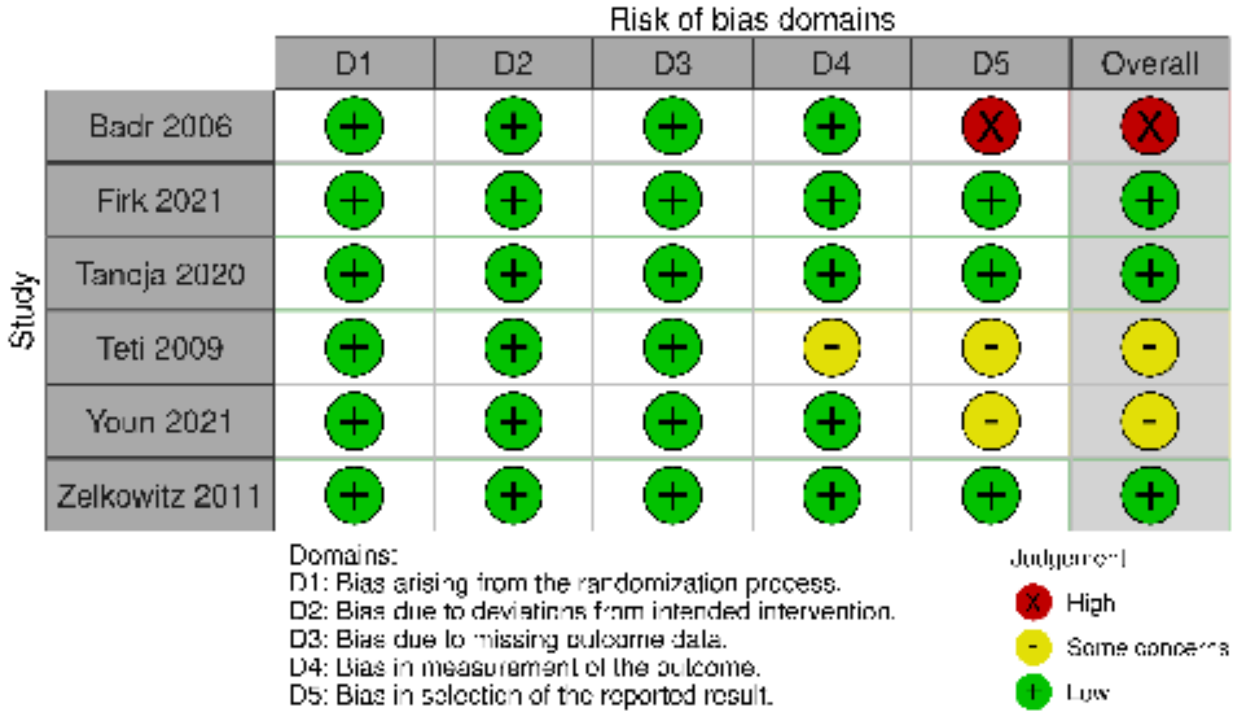


**eFigure 70. RoB (RCT) – Cognitive Development (7-12 months F/U)**

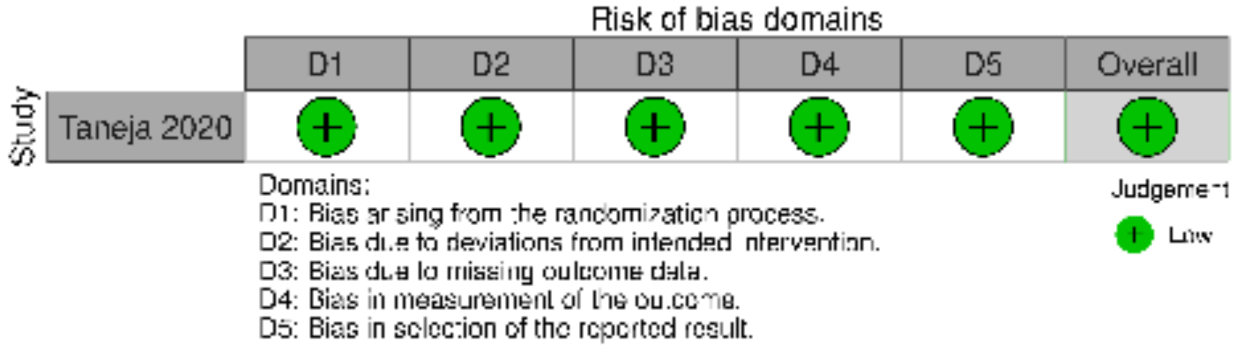

**eFigure 71. RoB (cluster RCT) – Cognitive Development (7-12 months F/U)**

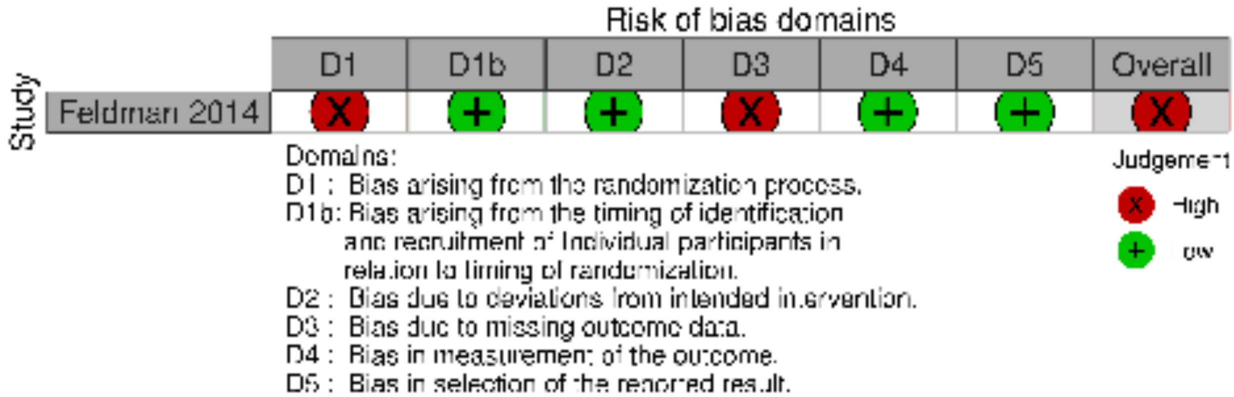


**eFigure 72. RoB (RCT) – Cognitive Development (13-24 months F/U)**

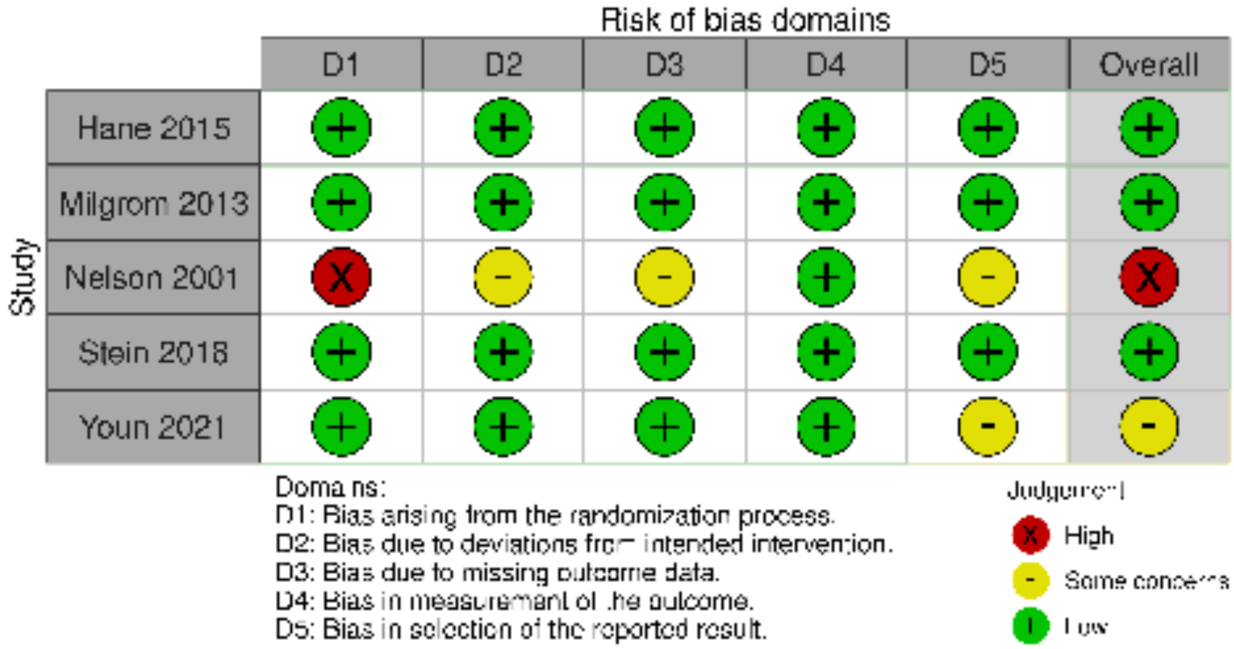

**eFigure 73. RoB (RCT) – Cognitive Development (25+ months F/U)**

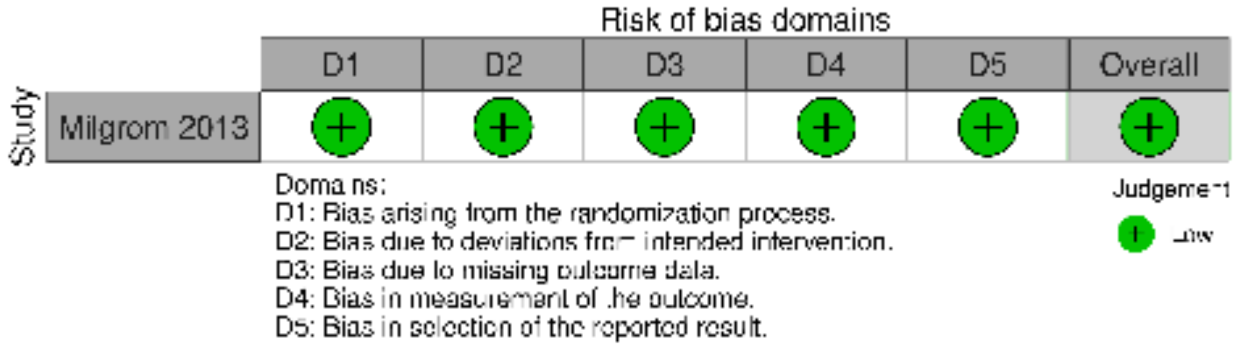


**eFigure 74. RoB (cluster RCT) – Cognitive Development (25+ months F/U)**

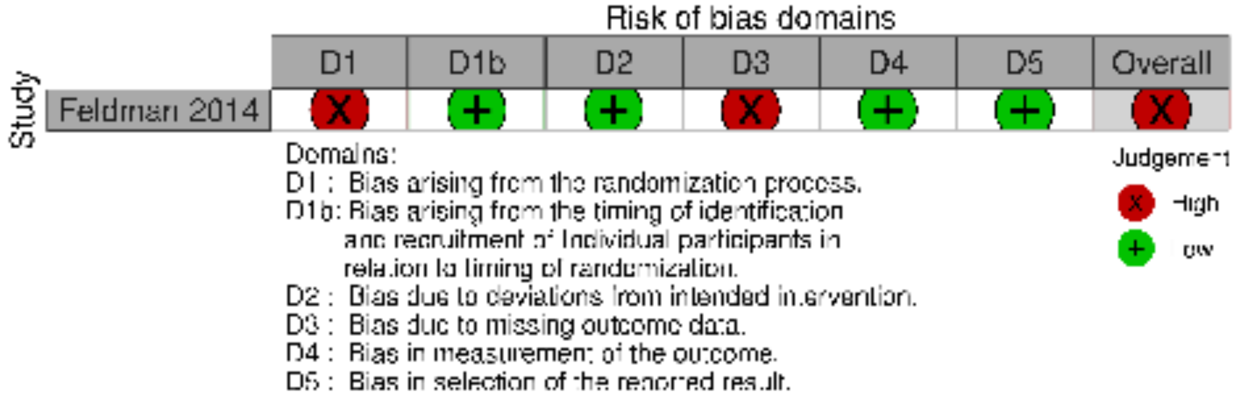


**eFigure 75. RoB (RCT) – Language Development (0-1 month F/U)**

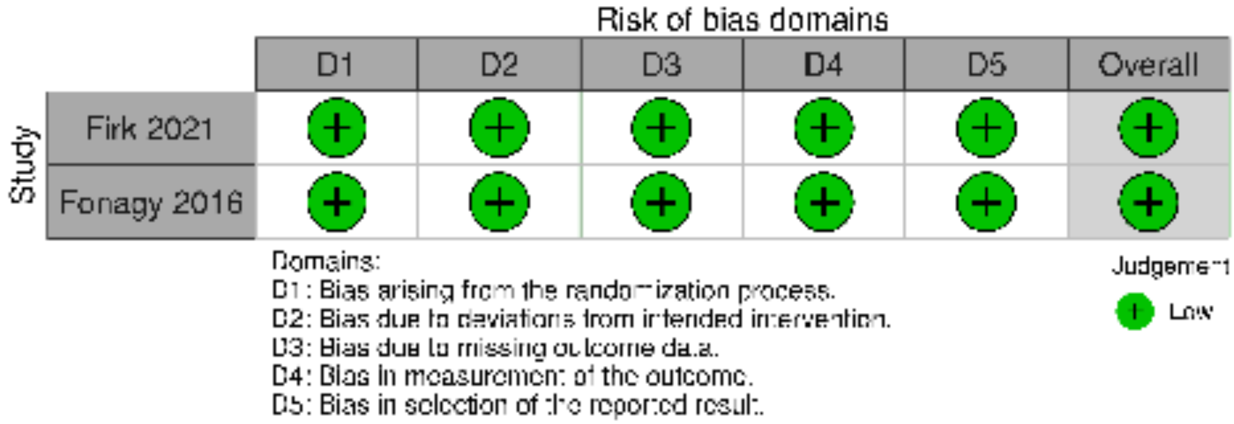

**eFigure 76. RoB (RCT) – Language Development (2-6 months F/U)**

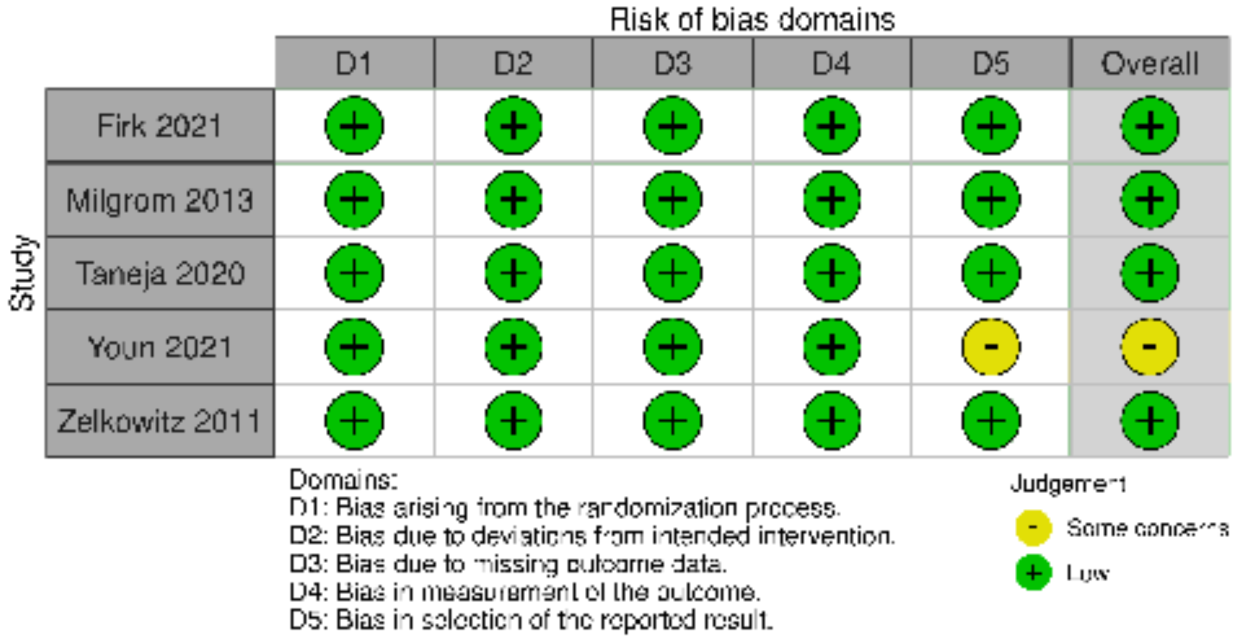


**eFigure 77. RoB (RCT) – Language Development (7-12 months F/U)**

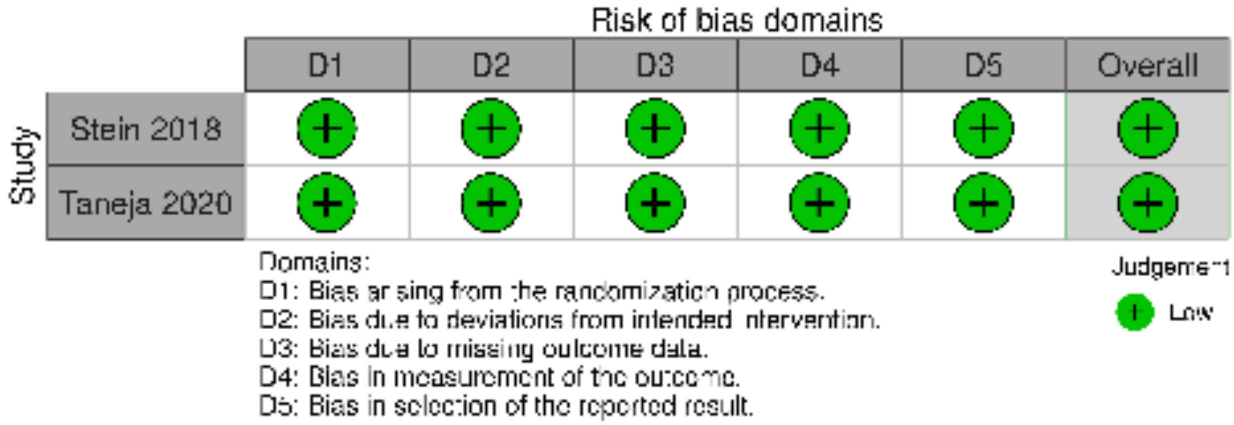

**eFigure 78. RoB (RCT) – Language Development (13-24 months F/U)**

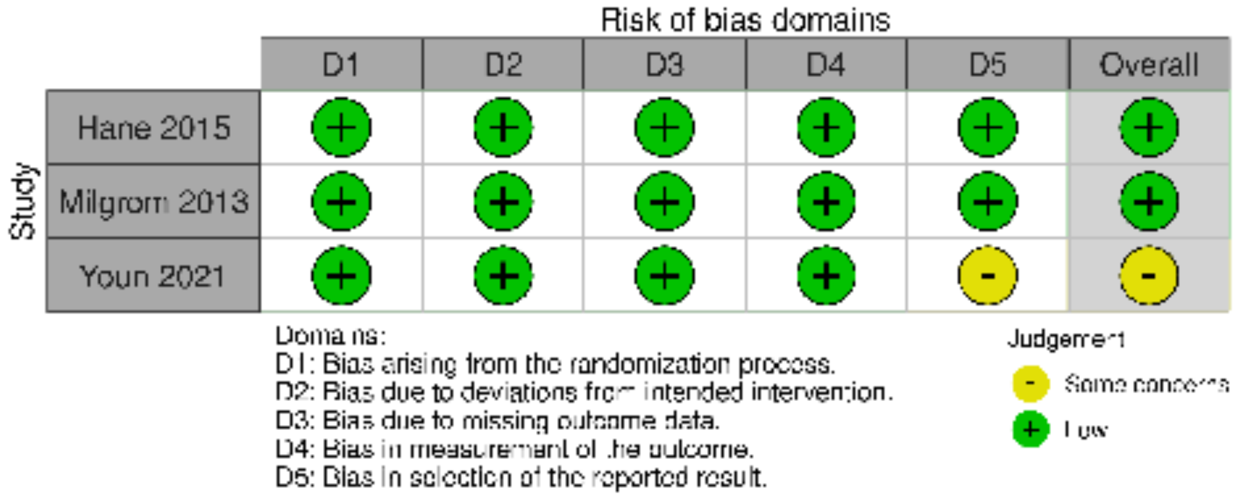


**eFigure 79. RoB (RCT) – Motor Development (0-1 month F/U)**

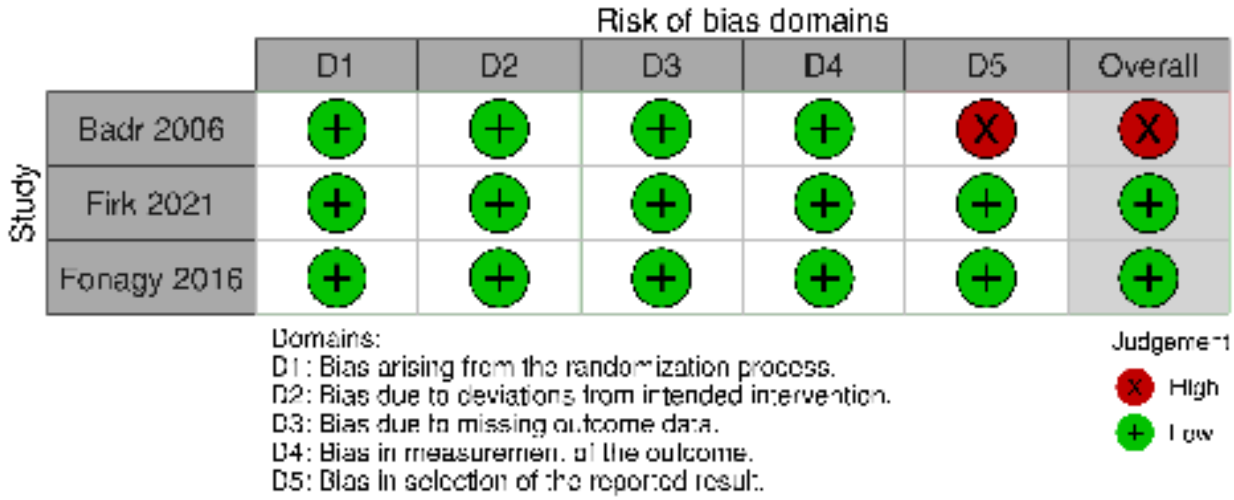

#### eFigure 80. RoB (RCT) – Motor Development (2-6 months F/U)

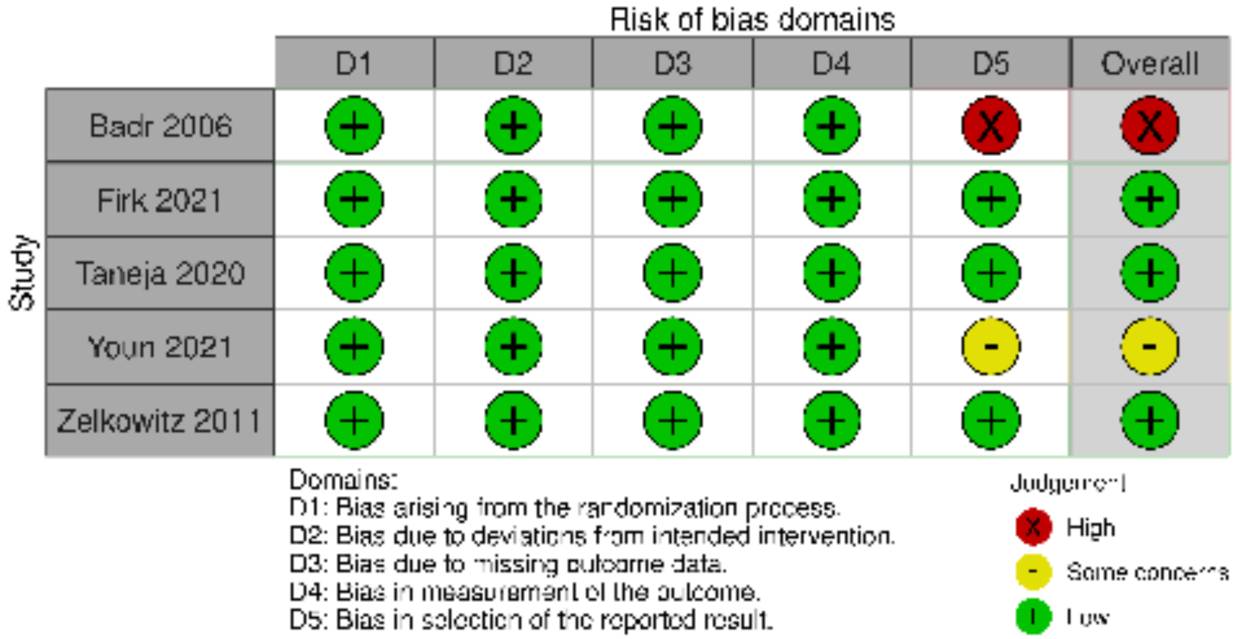


#### eFigure 81. RoB (RCT) – Motor Development (7-12 months F/U)

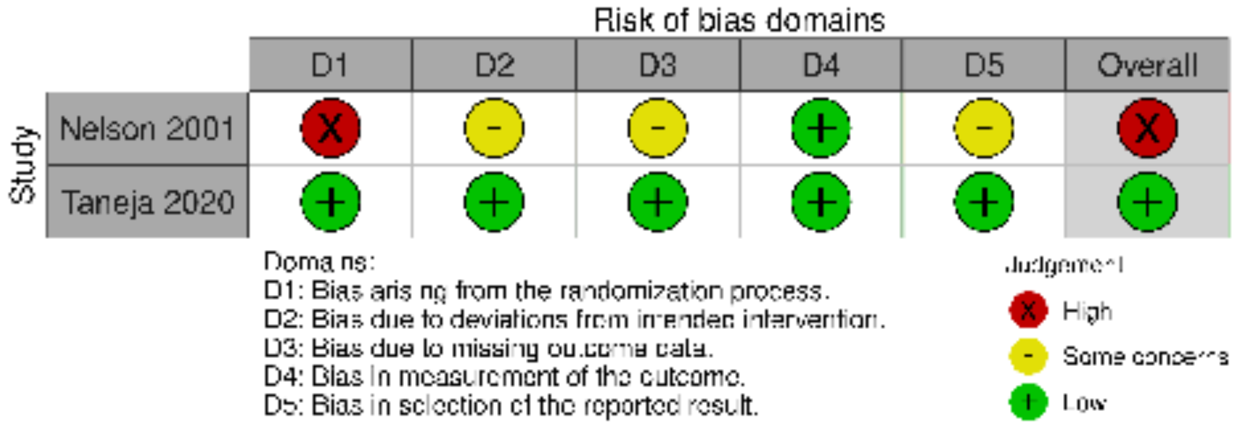

**eFigure 82. RoB (cluster RCT) – Motor Development (7-12 months F/U)**

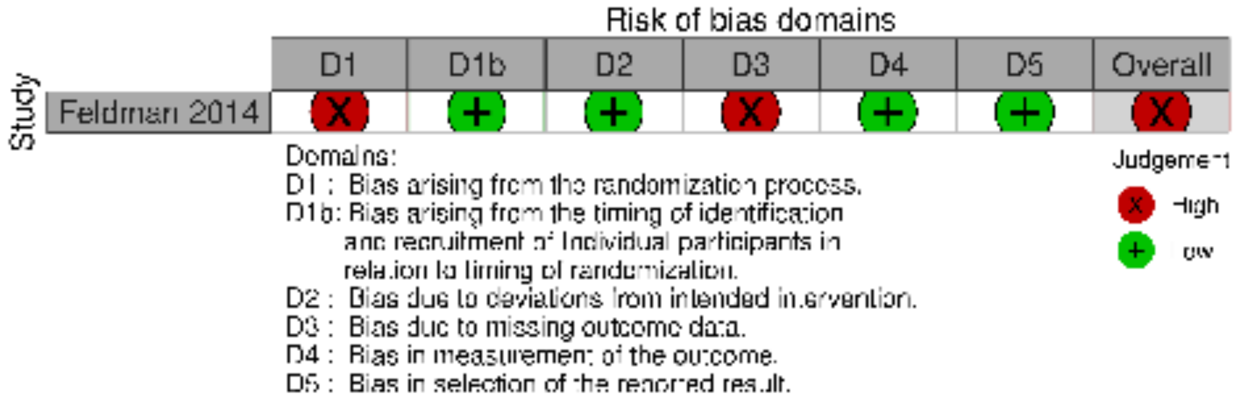


**eFigure 83. RoB (RCT) – Motor Development (13-24 months F/U)**

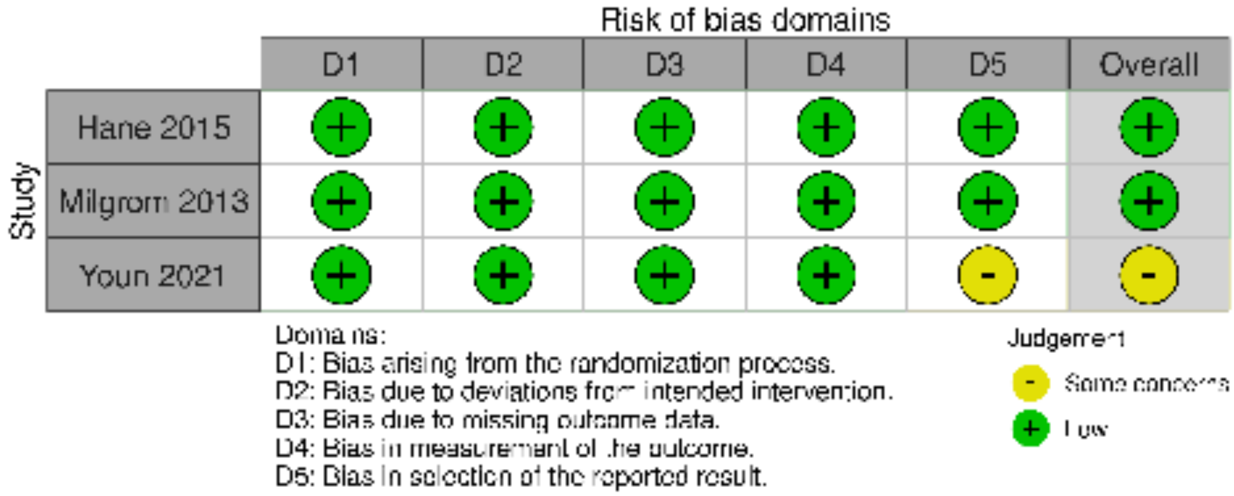


**eFigure 84. RoB (cluster RCT) – Motor Development (13-24 months F/U)**

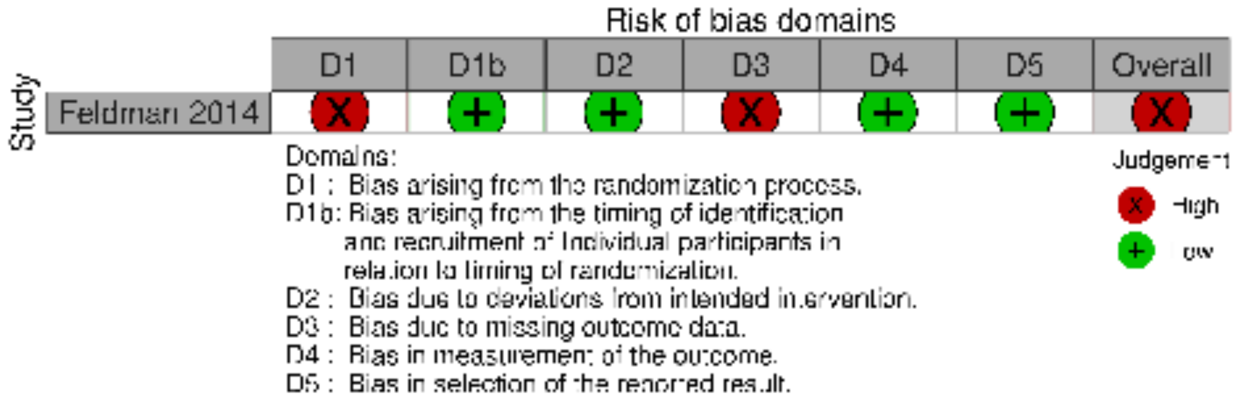

**eFigure 85. RoB (RCT) – Socio-Emotional Functioning (2-6 months F/U)**

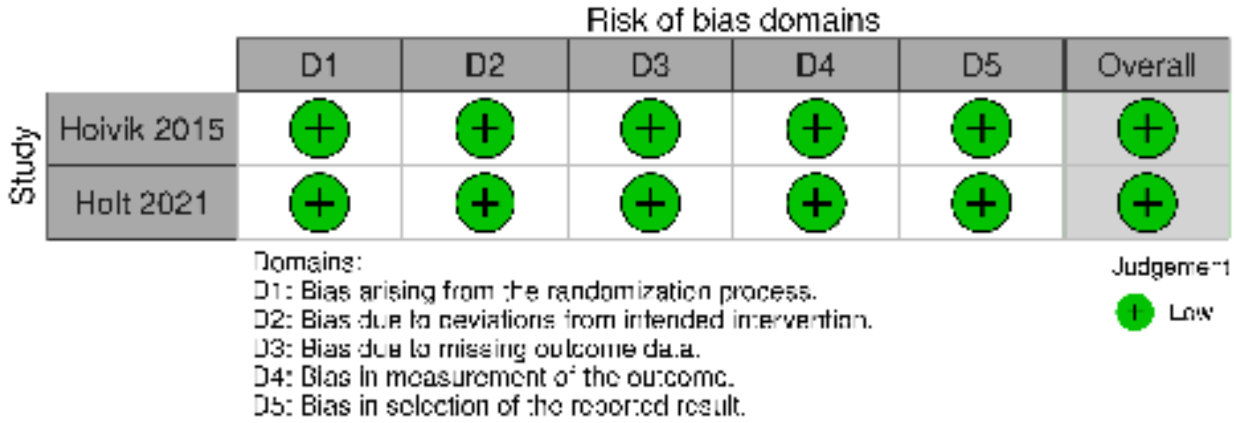


**eFigure 86. RoB (cluster RCT) – Socio-Emotional Functioning (2-6 months F/U)**

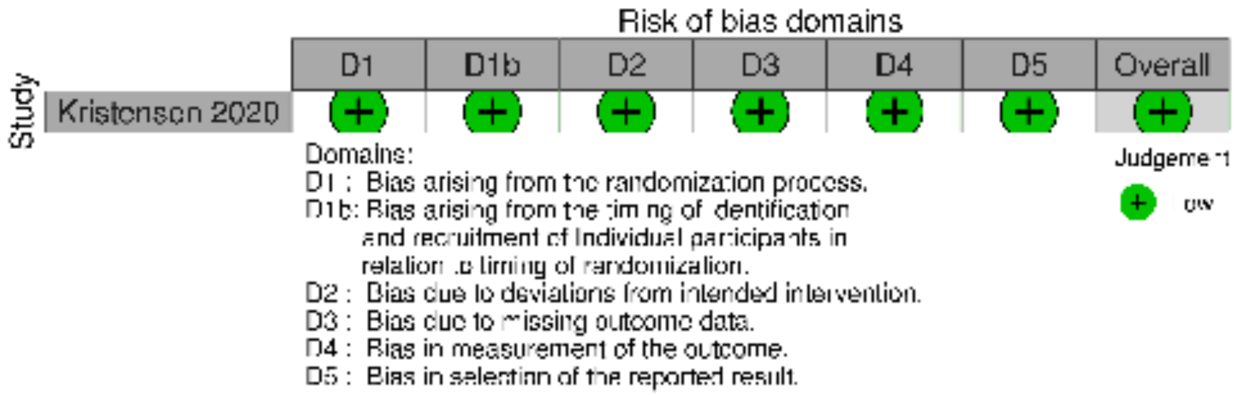


**eFigure 87. RoB (RCT) – Child Behavior (0-1 month F/U)**

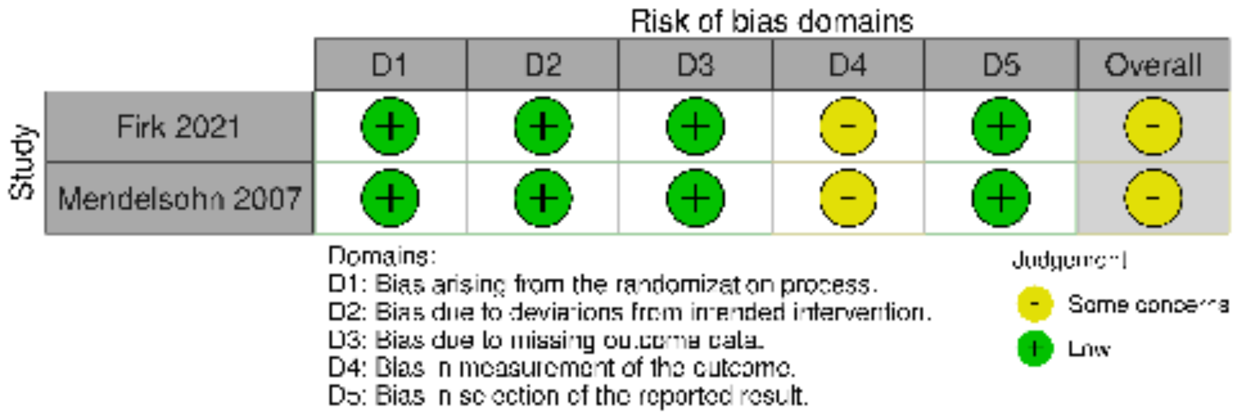

#### eFigure 88. RoB (RCT) – Child Behavior (2-6 months F/U)

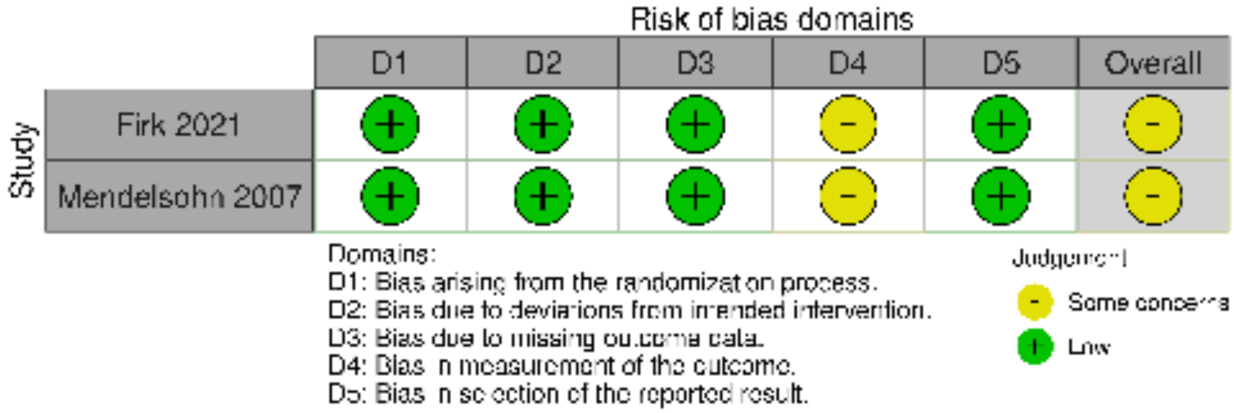


#### eFigure 89. RoB (RCT) – Child Behavior (13-24 months F/U)

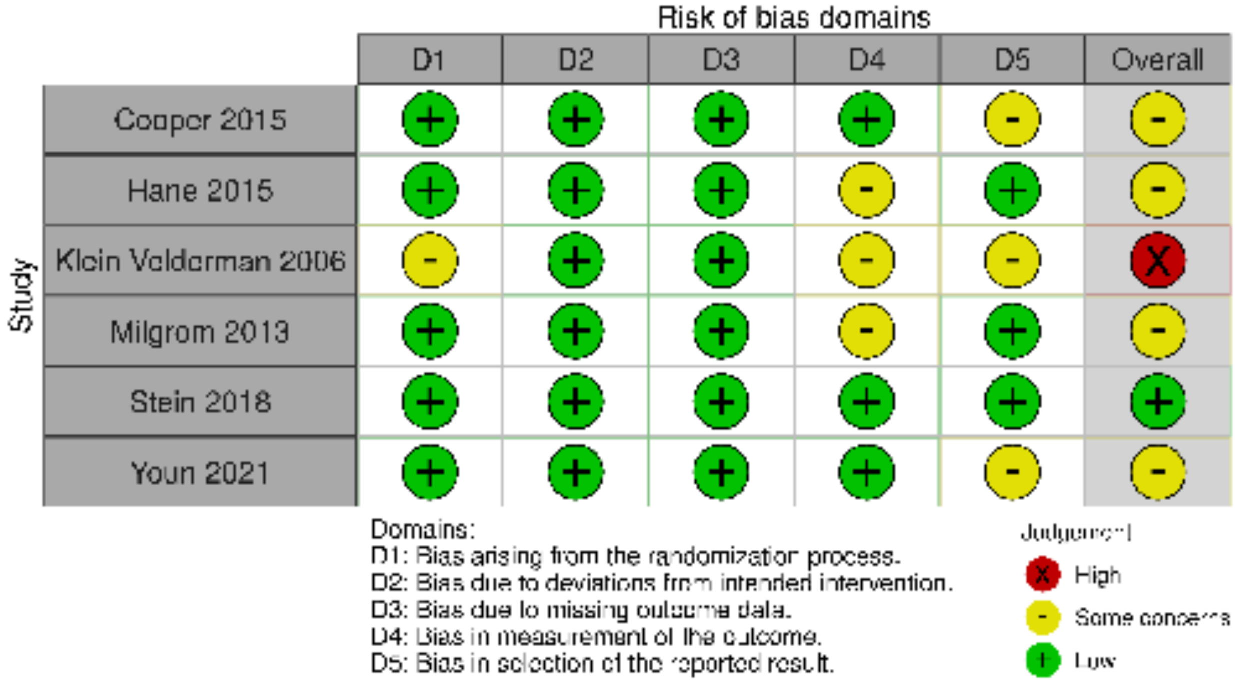

**eFigure 90. RoB (RCT) – Child Behavior (25+ months F/U)**

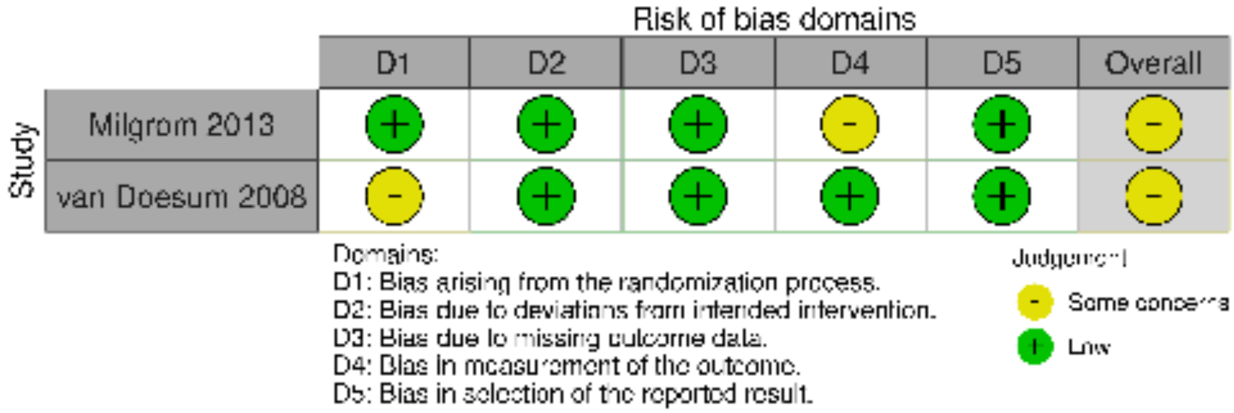


**eFigure 91. RoB (cluster RCT) – Child Behavior (25+ months F/U)**

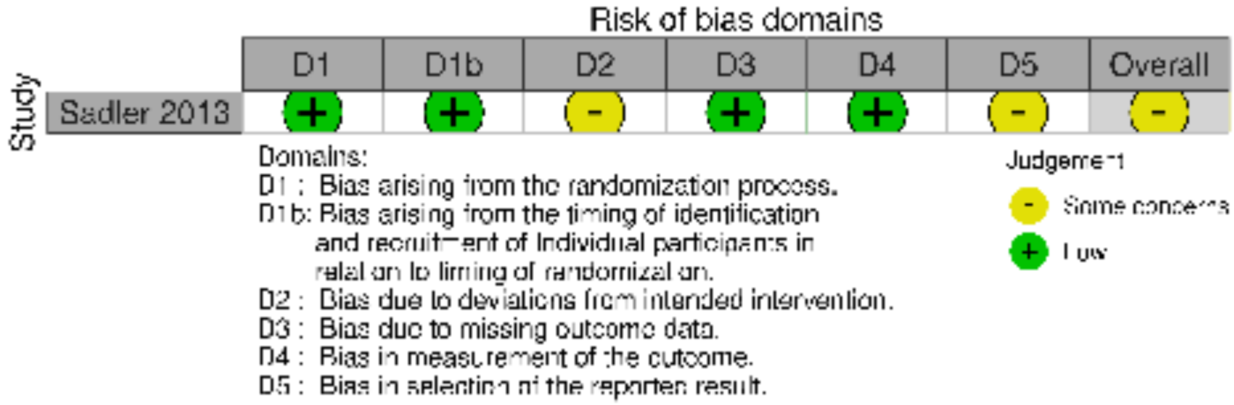

**eFigure 92. RoB (RCT) – Parenting Stress (0-1 month F/U)**

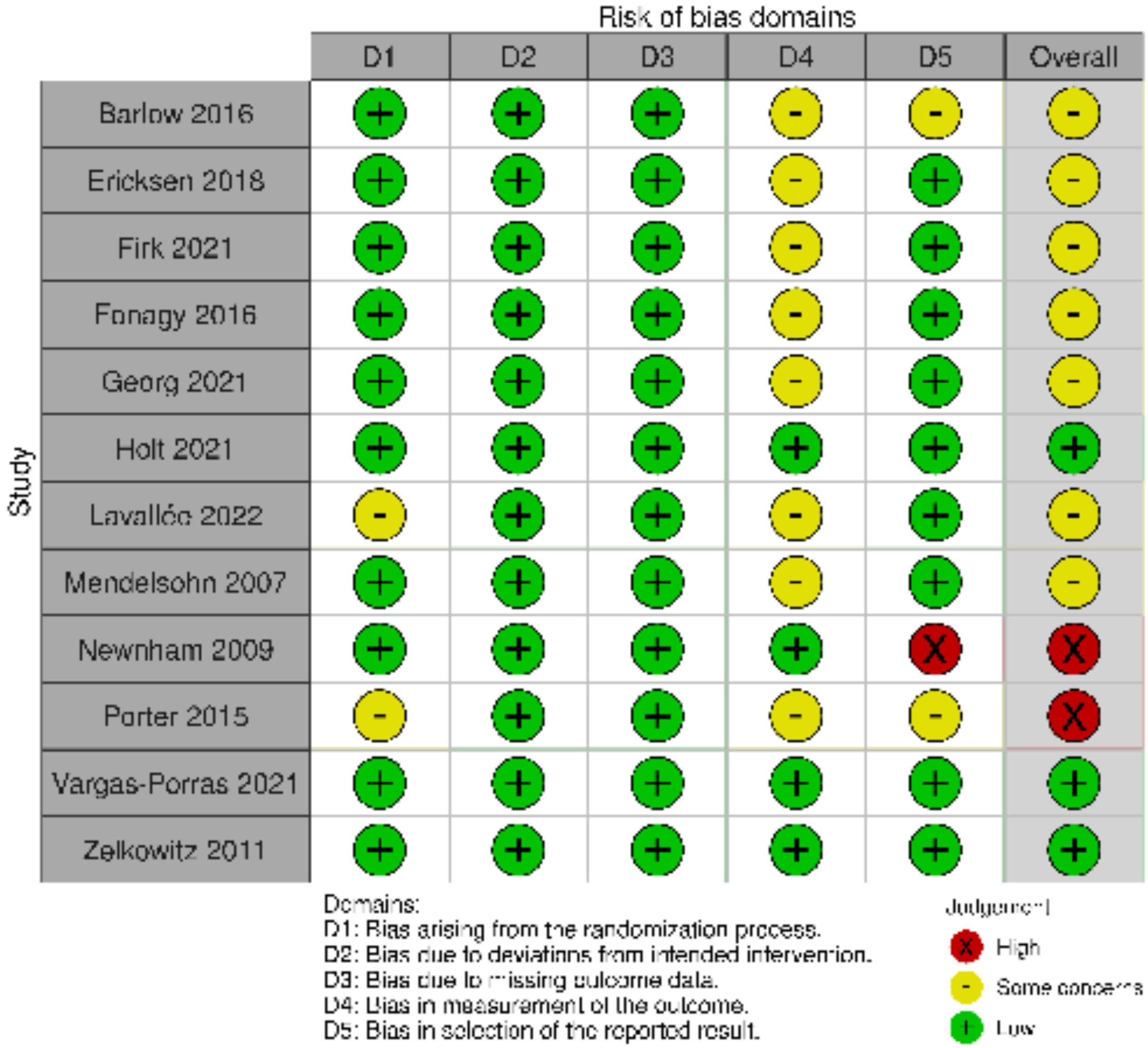

**eFigure 93. RoB (cluster RCT) – Parenting Stress (0-1 month F/U)**

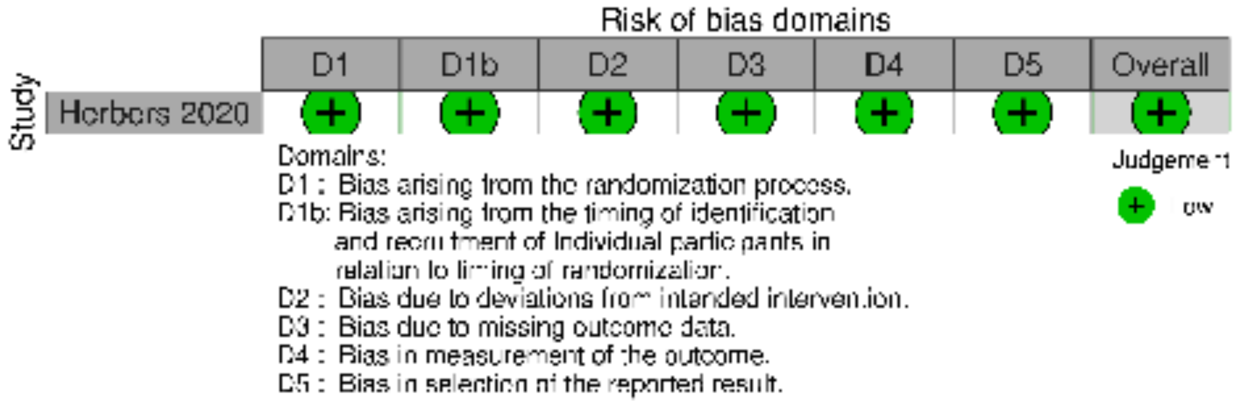


**eFigure 94. RoB (RCT) – Parenting Stress (2-6 months F/U)**

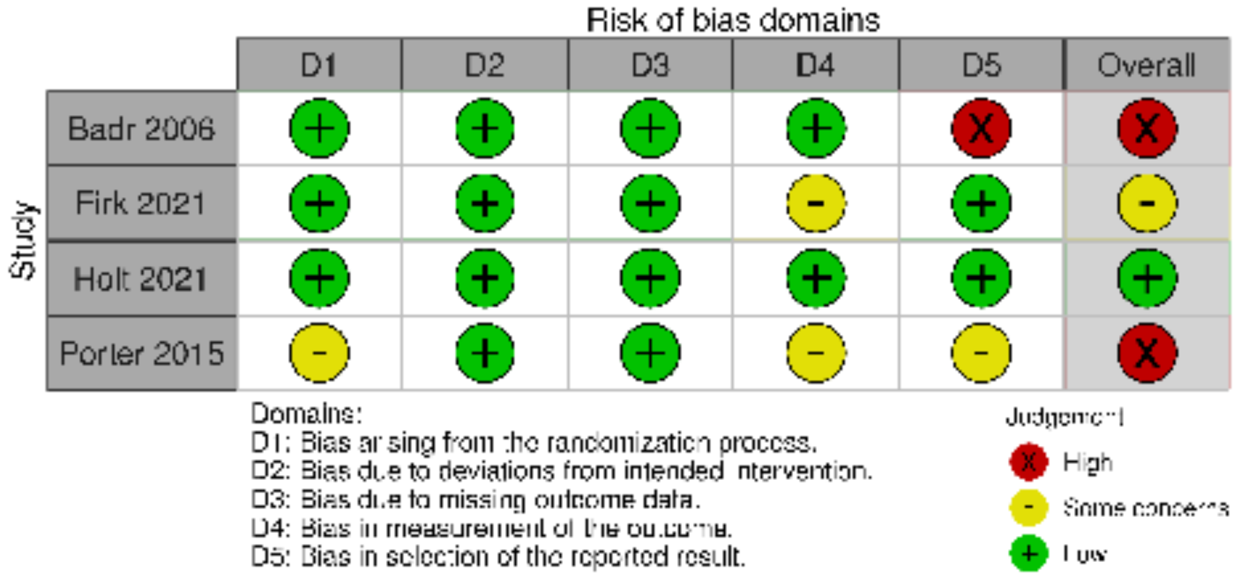

**eFigure 95. RoB (cluster RCT) – Parenting Stress (2-6 months F/U)**

Risk of bias domains

| Study | D1 | D1b | D2 | D3 | D4 | D5 | Overall |
|---|---|---|---|---|---|---|---|
| Glazebrook 2007 | + | + | + | + | + | + | + |

Domains:
D1 : Bias arising from the randomization process.
D1b: Bias arising from the timing of identification and recruitment of Individual participants in relation to timing of randomization.
D2 : Bias due to deviations from intended intervention.
D3 : Bias due to missing outcome data.
D4 : Bias in measurement of the outcome.
D5 : Bias in selection of the reported result.

Judgement
+ Low

**eFigure 96. RoB (RCT) – Parent/Caregiver Anxiety (0-1 month F/U)**

Risk of bias domains

| Study | D1 | D2 | D3 | D4 | D5 | Overall |
|---|---|---|---|---|---|---|
| Barlow 2016 | + | + | + | - | - | - |
| Ericksen 2018 | + | + | + | - | + | - |
| Gaden 2022 | + | + | + | - | + | - |
| Hane 2015 | + | + | + | - | + | - |
| Hoffenkamp 2015 | - | + | + | - | + | - |
| Holt 2021 | + | + | + | + | + | + |
| Lavallée 2022 | - | + | + | - | + | - |
| Melnyk 2006 | + | + | + | + | - | - |
| Wulff 2021 | + | X | X | X | - | X |
| Zelkowitz 2011 | + | + | + | + | + | + |

Domains:
D1: Bias arising from the randomization process.
D2: Bias due to deviations from intended intervention.
D3: Bias due to missing outcome data.
D4: Bias in measurement of the outcome.
D5: Bias in selection of the reported result.

Judgement
X High
- Some concerns
+ Low

**eFigure 97. RoB (cluster RCT) – Parent/Caregiver Anxiety (0-1 month F/U)**

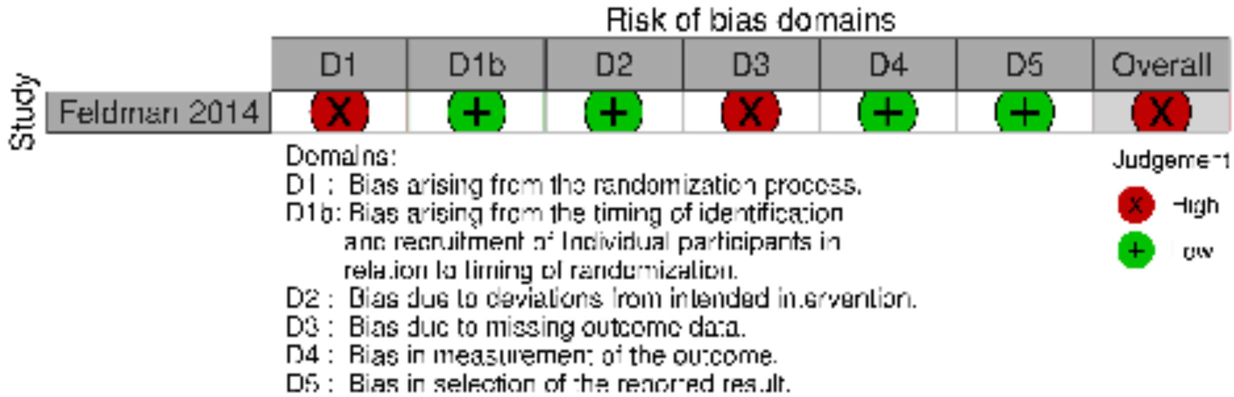


**eFigure 98. RoB (RCT) – Parent/Caregiver Anxiety (2-6 months F/U)**

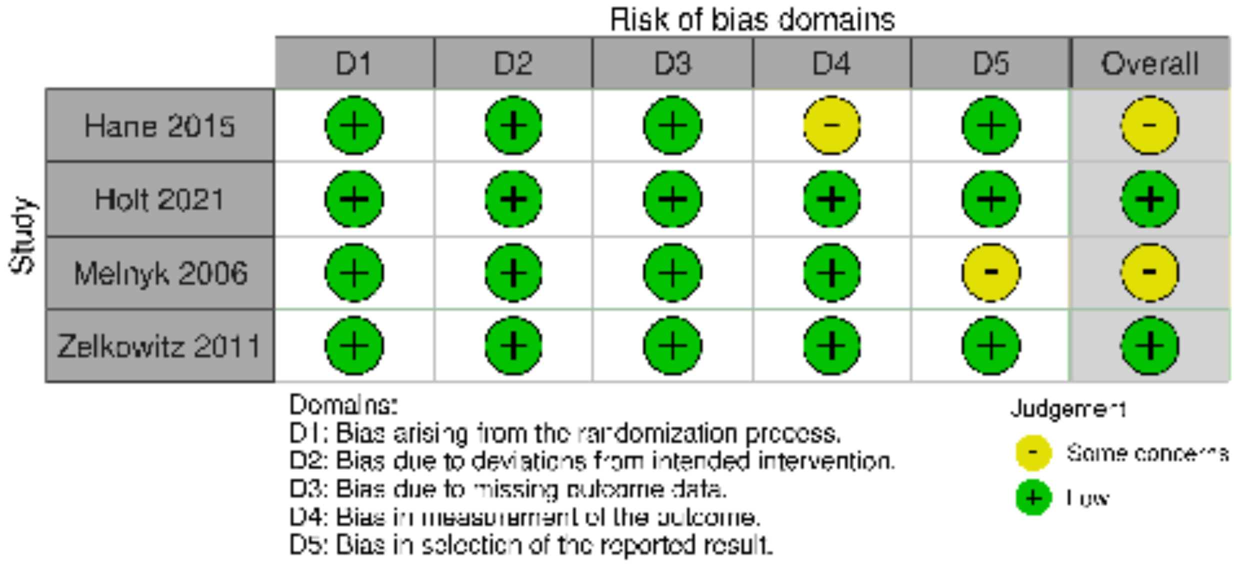

#### eFigure 99. RoB (RCT) – Parent/Caregiver Depression (0-1 month F/U)

Risk of bias domains

| Study | D1 | D2 | D3 | D4 | D5 | Overall |
|---|---|---|---|---|---|---|
| Barlow 2016 | + | + | + | - | - | - |
| Cooper 2009 | - | + | + | - | - | X |
| Cooper 2015 | + | + | + | + | - | - |
| Ericksen 2018 | + | + | + | - | + | - |
| Firk 2021 | + | + | + | - | + | - |
| Fonagy 2016 | + | + | + | - | + | - |
| Gaden 2022 | + | + | + | - | + | - |
| Georg 2021 | + | + | + | - | + | - |
| Hane 2015 | + | + | + | - | + | - |
| Hoffenkamp 2015 | - | + | + | - | + | - |
| Holt 2021 | + | + | + | + | + | + |
| Horowitz 2001 | + | - | + | + | + | - |
| Horowitz 2013 | + | - | + | + | + | - |
| Melnyk 2006 | + | + | + | + | - | - |
| Mendelsohn 2007 | + | + | + | - | + | - |
| Newnham 2009 | + | + | + | + | X | X |
| Porter 2015 | - | + | + | - | - | X |
| Taneja 2020 | + | + | + | + | + | + |
| Tryphonopoulos 2020 | + | - | + | + | - | - |
| Tsivos 2015 | + | - | + | + | + | - |
| van Doesum 2008 | - | + | + | + | + | - |
| Vargas-Porras 2021 | + | + | + | + | + | + |
| Wulff 2021 | + | X | X | X | - | X |
| Youn 2021 | + | + | + | + | - | - |

Domains:
D1: Bias arising from the randomization process.
D2: Bias due to deviations from intended intervention.
D3: Bias due to missing outcome data.
D4: Bias in measurement of the outcome.
D5: Bias in selection of the reported result.

Judgement
X High
- Some concerns
+ Low

**eFigure 100. RoB (cluster RCT) – Parent/Caregiver Depression (0-1 months F/U)**

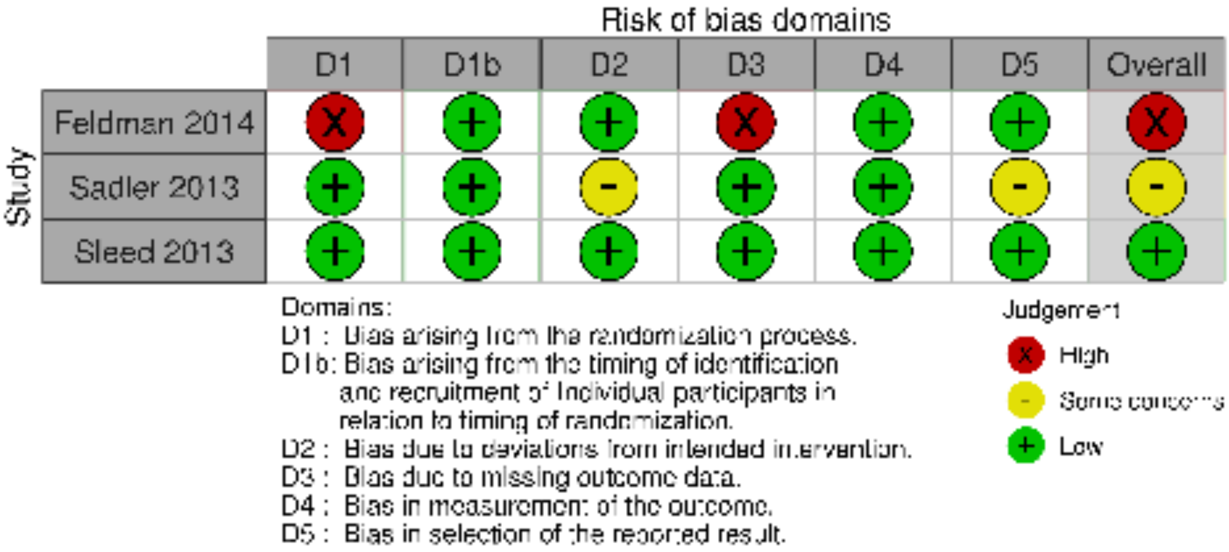

**eFigure 101. RoB (RCT) – Parent/Caregiver Depression (2-6 months F/U)**

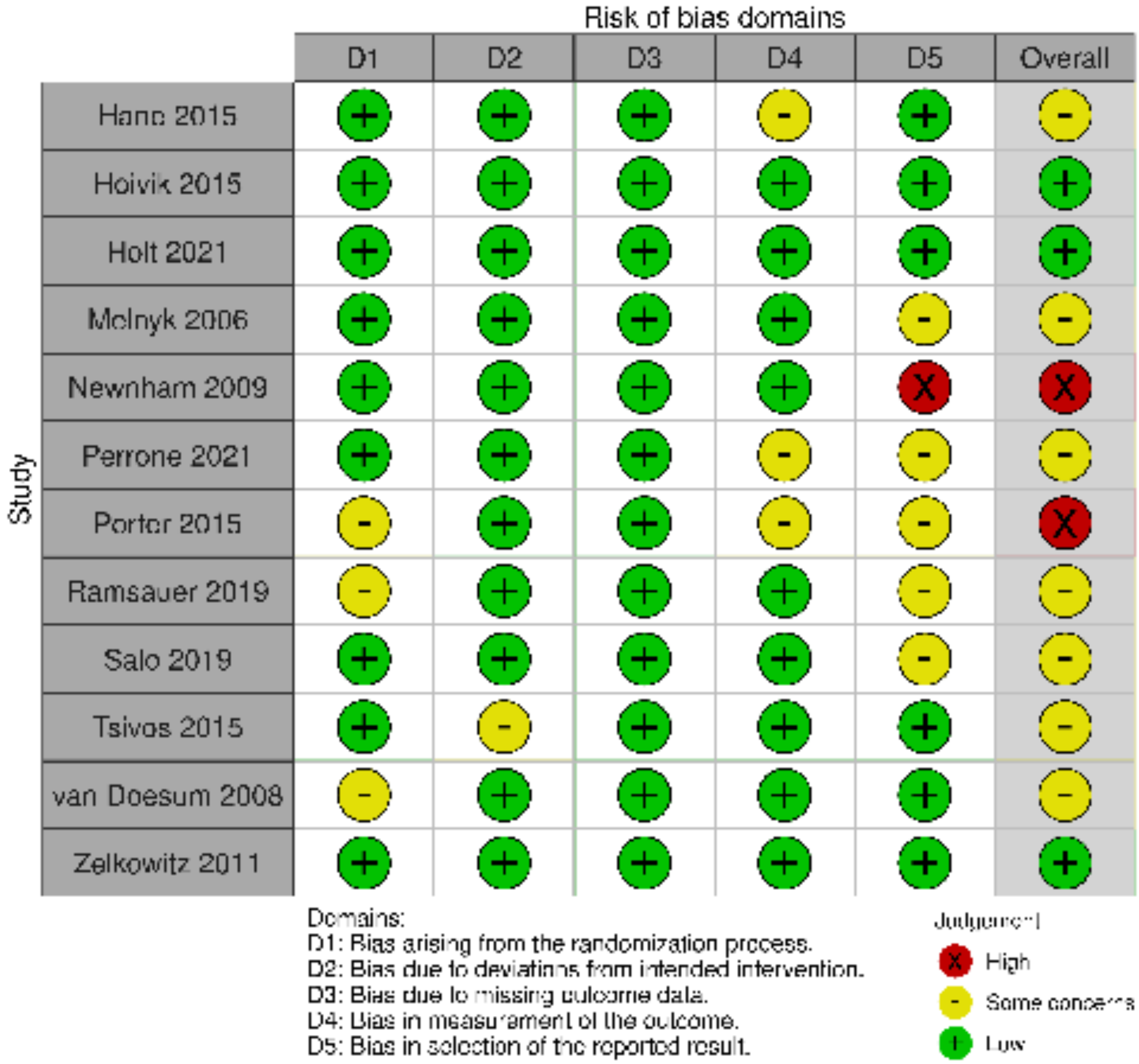

**eFigure 102. RoB (RCT) – Parent/Caregiver Depression (7-12 months F/U)**

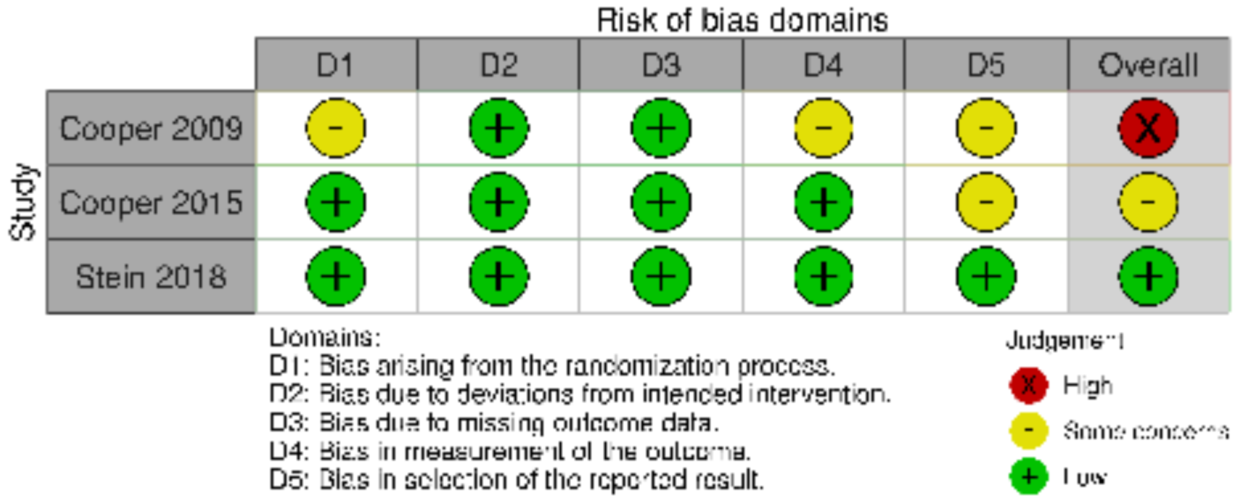


**eFigure 103. RoB (RCT) – Parent/Caregiver Depression (13-24 months F/U)**

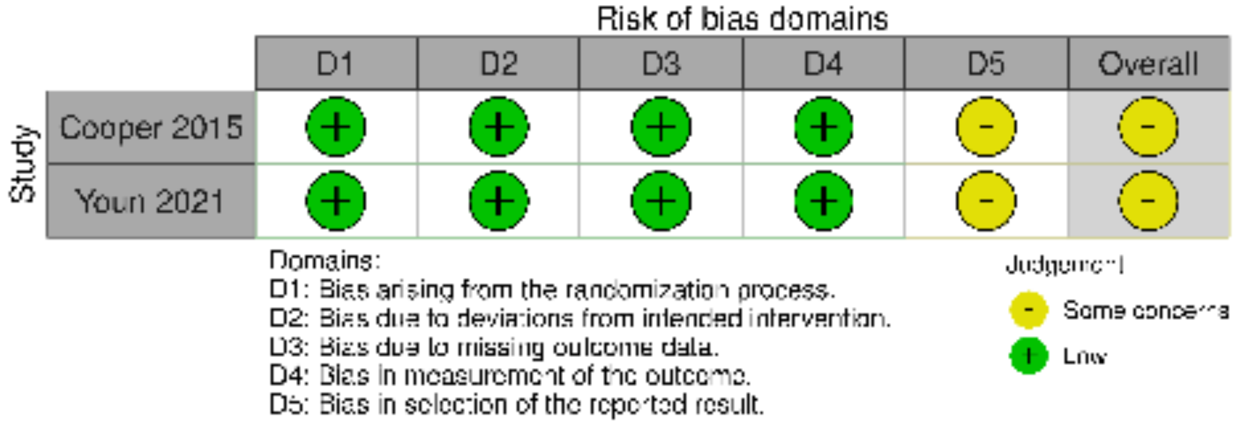

#### References

1. Chiu SH, Anderson GC. Effect of early skin-to-skin contact on mother-preterm infant interaction through 18 months: randomized controlled trial. *Int J Nurs Stud*. Sep 2009;46(9):1168-80. doi:10.1016/j.ijnurstu.2009.03.005
2. Feldman R, Rosenthal Z, Eidelman AI. Maternal-Preterm Skin-to-Skin Contact Enhances Child Physiologic Organization and Cognitive Control Across the First 10 Years of Life. *Biological Psychiatry*. 2014/01/01/ 2014;75(1):56-64. doi:https://doi.org/10.1016/j.biopsych.2013.08.012
3. Neu M, Robinson J. Maternal holding of preterm infants during the early weeks after birth and dyad interaction at six months. *J Obstet Gynecol Neonatal Nurs*. Jul-Aug 2010;39(4):401-14. doi:10.1111/j.1552-6909.2010.01152.x
4. Rheinheimer N, Beijers R, Cooijmans KHM, Brett BE, de Weerth C. Effects of skin-to-skin contact on full-term infants' stress reactivity and quality of mother–infant interactions. *Developmental Psychobiology*. 2022;64(7)doi:10.1002/dev.22308
5. Sahlen Helmer C, Birberg Thornberg U, Frostell A, Ortenstrand A, Morelius E. A Randomized Trial of Continuous Versus Intermittent Skin-to-Skin Contact After Premature Birth and the Effects on Mother-Infant Interaction. *Adv Neonatal Care*. Jun 2020;20(3):E48-E56. doi:10.1097/ANC.0000000000000675
6. Taneja S, Sinha B, Upadhyay RP, et al. Community initiated kangaroo mother care and early child development in low birth weight infants in India-a randomized controlled trial. *BMC Pediatr*. Apr 4 2020;20(1):150. doi:10.1186/s12887-020-02046-4
7. Williams LR, Turner PR. Infant carrying as a tool to promote secure attachments in young mothers: Comparing intervention and control infants during the still-face paradigm. *Infant Behav Dev*. Feb 2020;58:101413. doi:10.1016/j.infbeh.2019.101413
8. Williams LR, Turner PR. Experiences with "Babywearing": Trendy parenting gear or a developmentally attuned parenting tool? *Children and Youth Services Review*. 2020;112doi:10.1016/j.childyouth.2020.104918
9. Cevasco AM. The Effects of Mothers' Singing on Full-term and Preterm Infants and Maternal Emotional Responses. *Journal of Music Therapy*. 2008;45(3):273-306.
10. Corrigan M, Keeler J, Miller H, Naylor C, Diaz A. Music Therapy and Family-Integrated Care in the NICU: Using Heartbeat-Music Interventions to Promote Mother-Infant Bonding. *Adv Neonatal Care*. Oct 1 2022;22(5):E159-E168. doi:10.1097/ANC.0000000000000910
11. Gaden TS, Ghetti C, Kvestad I, et al. Short-term Music Therapy for Families With Preterm Infants: A Randomized Trial. *Pediatrics*. Feb 1 2022;149(2)doi:10.1542/peds.2021-052797
12. Robertson AM, Detmer MR. The Effects of Contingent Lullaby Music on Parent-Infant Interaction and Amount of Infant Crying in the First Six Weeks of Life. *J Pediatr Nurs*. May - Jun 2019;46:33-38. doi:10.1016/j.pedn.2019.02.025
13. Wulff V, Hepp P, Wolf OT, Fehm T, Schaal NK. The influence of maternal singing on well-being, postpartum depression and bonding - a randomised, controlled trial. *BMC Pregnancy Childbirth*. Jul 12 2021;21(1):501. doi:10.1186/s12884-021-03933-z
14. Yu WC, Chiang MC, Lin KC, Chang CC, Lin KH, Chen CW. Effects of maternal voice on pain and mother-Infant bonding in premature infants in Taiwan: A randomized controlled trial. *J Pediatr Nurs*. Mar-Apr 2022;63:e136-e142. doi:10.1016/j.pedn.2021.09.022
15. Akai CE, Guttentag CL, Baggett KM, Noria CC, Centers for the Prevention of Child N. Enhancing parenting practices of at-risk mothers. *J Prim Prev*. May 2008;29(3):223-42. doi:10.1007/s10935-008-0134-z
16. Baggett KM, Davis B, Feil EG, et al. Technologies for expanding the reach of evidence-based interventions: Preliminary results for promoting social-emotional development in early childhood. *Topics Early Child Spec Educ*. Feb 1 2010;29(4):226-238. doi:10.1177/0271121409354782
17. Baggett K, Davis B, Feil E, et al. A Randomized Controlled Trial Examination of a Remote Parenting Intervention: Engagement and Effects on Parenting Behavior and Child Abuse Potential. *Child Maltreat*. Nov 2017;22(4):315-323. doi:10.1177/1077559517712000
18. Feil EG, Baggett K, Davis B, et al. Randomized control trial of an internet-based parenting intervention for mothers of infants. *Early Child Res Q*. 1st Quarter 2020;50(Pt 1):36-44. doi:10.1016/j.ecresq.2018.11.003
19. Landry SH, Smith KE, Swank PR. Responsive parenting: establishing early foundations for social, communication, and independent problem-solving skills. *Dev Psychol*. Jul 2006;42(4):627-42. doi:10.1037/0012-1649.42.4.627
20. Barnes C, N. Adamson-Macedo E. Understanding the impact of newborn touch upon mothers of hospitalized preterm neonates. *Journal of Human Growth and Development*. 2022;32(2):294-301. doi:10.36311/jhgd.v32.13322

21. Onozawaa K, Gloverb V, Adamsb D, Modib N, Kumara RC. Infant massage improves mother–infant interaction for mothers with postnatal depression. *Journal of Affective Disorders*. 2001;63:201-207.
22. Porter LS, Porter BO, McCoy V, et al. Blended Infant Massage-Parenting Enhancement Program on Recovering Substance-Abusing Mothers' Parenting Stress, Self-Esteem, Depression, Maternal Attachment, and Mother-Infant Interaction. *Asian Nurs Res (Korean Soc Nurs Sci)*. Dec 2015;9(4):318-27. doi:10.1016/j.anr.2015.09.002
23. Sajaniemi N, Makela J, Salokorpi T, von Wendt L, Hamalainen T, Hakamies-Blomqvist L. Cognitive performance and attachment patterns at four years of age in extremely low birth weight infants after early intervention. *Eur Child Adolesc Psychiatry*. Jun 2001;10(2):122-9. doi:10.1007/s007870170035
24. Shoghi M, Sohrabi S, Rasouli M. The Effects of Massage by Mothers on Mother-Infant Attachment. *Alternative therapies in health and medicine*. 2018;24(3):34–39.
25. Teti DM, Black MM, Viscardi R, et al. Intervention With African American Premature Infants: Four-Month Results of an Early Intervention Program. *Journal of Early Intervention*. 2009;31(2):146-166. doi:10.1177/1053815109331864
26. Alvarenga P, Cerezo MA, Wiese E, Piccinini CA. Effects of a short video feedback intervention on enhancing maternal sensitivity and infant development in low-income families. *Attach Hum Dev*. Oct 2020;22(5):534-554. doi:10.1080/14616734.2019.1602660
27. Barlow J, Sembi S, Underdown A. Pilot RCT of the use of video interactive guidance with preterm babies. *Journal of Reproductive and Infant Psychology*. 2016;34(5):511-524. doi:10.1080/02646838.2016.1217404
28. Borghini A, Habersaat S, Forcada-Guex M, et al. Effects of an early intervention on maternal post-traumatic stress symptoms and the quality of mother-infant interaction: the case of preterm birth. *Infant Behav Dev*. Nov 2014;37(4):624-31. doi:10.1016/j.infbeh.2014.08.003
29. Brisch KH, Bechinger D, Betzler S, Heinemann H. Early preventive attachment-oriented psychotherapeutic intervention program with parents of a very low birthweight premature infant: results of attachment and neurological development. *Attach Hum Dev*. Jun 2003;5(2):120-35. doi:10.1080/1461673031000108504
30. Cates CB, Weisleder A, Berkule Johnson S, et al. Enhancing Parent Talk, Reading, and Play in Primary Care: Sustained Impacts of the Video Interaction Project. *J Pediatr*. Aug 2018;199:49-56 e1. doi:10.1016/j.jpeds.2018.03.002
31. Firk C, Dahmen B, Dempfle A, et al. A mother-child intervention program for adolescent mothers: Results from a randomized controlled trial (the TeeMo study). *Dev Psychopathol*. Aug 2021;33(3):992-1005. doi:10.1017/S0954579420000280
32. Hoffenkamp HN, Tooten A, Hall RA, et al. Effectiveness of hospital-based video interaction guidance on parental interactive behavior, bonding, and stress after preterm birth: A randomized controlled trial. *J Consult Clin Psychol*. Apr 2015;83(2):416-29. doi:10.1037/a0038401
33. Hoivik MS, Lydersen S, Drugli MB, Onsoien R, Hansen MB, Nielsen TS. Video feedback compared to treatment as usual in families with parent-child interactions problems: a randomized controlled trial. *Child Adolesc Psychiatry Ment Health*. 2015;9:3. doi:10.1186/s13034-015-0036-9
34. Juffer F, Bakermans-Kranenburg MJ, van Ijzendoorn MH. The importance of parenting in the development of disorganized attachment: Evidence from a preventive intervention study in adoptive families. *Journal of Child Psychology and Psychiatry*. 2005;46(3):263-274. doi:10.1111/j.1469-7610.2004.00353.x
35. Klein Velderman M, Bakermans-Kranenburg MJ, Juffer F, Van IMH, Mangelsdorf SC, Zevalkink J. Preventing preschool externalizing behavior problems through video-feedback intervention in infancy. *Infant Ment Health J*. Sep 2006;27(5):466-493. doi:10.1002/imhj.20104
36. Klein Velderman M, Bakermans-Kranenburg MJ, Juffer F, van IMH. Effects of attachment-based interventions on maternal sensitivity and infant attachment: differential susceptibility of highly reactive infants. *J Fam Psychol*. Jun 2006;20(2):266-274. doi:10.1037/0893-3200.20.2.266
37. Magill-Evans J, Harrison M, Benzies K, Gierl M, Kimak C. Effects of Parenting Education on First-Time Fathers' Skills in Interactions with Their Infants. *Fathering: A Journal of Theory, Research, and Practice about Men as Fathers*. 2007;5(1):42-57. doi:10.3149/fth.0501.42
38. Mendelsohn AL, Valdez PT, Flynn V, et al. Use of videotaped interactions during pediatric well-child care: impact at 33 months on parenting and on child development. *J Dev Behav Pediatr*. Jun 2007;28(3):206-12. doi:10.1097/DBP.0b013e3180324d87
39. Oxford ML, Hash JB, Lohr MJ, et al. Randomized trial of promoting first relationships for new mothers who received community mental health services in pregnancy. *Dev Psychol*. Aug 2021;57(8):1228-1241. doi:10.1037/dev0001219

40. Ramsauer B, Muhlhan C, Lotzin A, et al. Randomized controlled trial of the Circle of Security-Intensive intervention for mothers with postpartum depression: maternal unresolved attachment moderates changes in sensitivity. *Attach Hum Dev*. Dec 2020;22(6):705-726. doi:10.1080/14616734.2019.1689406
41. Roby E, Miller EB, Shaw DS, et al. Improving Parent-Child Interactions in Pediatric Health Care: A Two-Site Randomized Controlled Trial. *Pediatrics*. 2021;147(3):e20201799.
42. Canfield CF, Miller EB, Shaw DS, Morris P, Alonso A, Mendelsohn AL. Beyond language: Impacts of shared reading on parenting stress and early parent-child relational health. *Dev Psychol*. Jul 2020;56(7):1305-1315. doi:10.1037/dev0000940
43. Stein A, Woolley H, Senior R, et al. Treating Disturbances in the Relationship Between Mothers With Bulimic Eating Disorders and Their Infants: A Randomized, Controlled Trial of Video Feedback. *American Journal of Psychiatry*. 2006;163:899–906.
44. Stein A, Netsi E, Lawrence PJ, et al. Mitigating the effect of persistent postnatal depression on child outcomes through an intervention to treat depression and improve parenting: a randomised controlled trial. *The Lancet Psychiatry*. 2018;5(2):134-144. doi:10.1016/s2215-0366(18)30006-3
45. Tereno S, Madigan S, Lyons-Ruth K, et al. Assessing a change mechanism in a randomized home-visiting trial: Reducing disrupted maternal communication decreases infant disorganization. *Development and Psychopathology*. 2017;29(2):637-649. doi:10.1017/S0954579417000232
46. Tryphonopoulos PD, Letourneau N. Promising Results From a Video-Feedback Interaction Guidance Intervention for Improving Maternal-Infant Interaction Quality of Depressed Mothers: A Feasibility Pilot Study. *Can J Nurs Res*. Jun 2020;52(2):74-87. doi:10.1177/0844562119892769
47. Twohig A, Murphy JF, McCarthy A, et al. The preterm infant-parent programme for attachment-PIPPA Study: a randomised controlled trial. *Pediatr Res*. Sep 2021;90(3):617-624. doi:10.1038/s41390-020-01262-z
48. van Doesum KTM, Riksen-Walraven JM, Hosman CMH, Hoefnagels C. A Randomized Controlled Trial of a Home-Visiting Intervention Aimed at PreventingRelationship Problems in Depressed Mothers and Their Infants. *Child Development*. 2008;79(3):547-561.
49. Kersten-Alvarez LE, Hosman CM, Riksen-Walraven JM, Van Doesum KT, Hoefnagels C. Long-term effects of a home-visiting intervention for depressed mothers and their infants. *J Child Psychol Psychiatry*. Oct 2010;51(10):1160-70. doi:10.1111/j.1469-7610.2010.02268.x
50. Hane AA, Myers MM, Hofer MA, et al. Family Nurture Intervention Improves the Quality of Maternal Caregiving in the Neonatal Intensive Care Unit: Evidence from a Randomized Controlled Trial. *Journal of Developmental and Behavioral Pediatrics*. 2015;36:188–196.
51. Beebe B, Myers MM, Lee SH, et al. Family nurture intervention for preterm infants facilitates positive mother-infant face-to-face engagement at 4 months. *Dev Psychol*. Nov 2018;54(11):2016-2031. doi:10.1037/dev0000557
52. Welch MG, Firestein MR, Austin J, et al. Family Nurture Intervention in the Neonatal Intensive Care Unit improves social-relatedness, attention, and neurodevelopment of preterm infants at 18 months in a randomized controlled trial. *J Child Psychol Psychiatry*. Nov 2015;56(11):1202-11. doi:10.1111/jcpp.12405
53. Welch MG, Halperin MS, Austin J, et al. Depression and anxiety symptoms of mothers of preterm infants are decreased at 4 months corrected age with Family Nurture Intervention in the NICU. *Arch Womens Ment Health*. Feb 2016;19(1):51-61. doi:10.1007/s00737-015-0502-7
54. Betancourt TS, Jensen SKG, Barnhart DA, et al. Promoting parent-child relationships and preventing violence via home-visiting: a pre-post cluster randomised trial among Rwandan families linked to social protection programmes. *BMC Public Health*. May 6 2020;20(1):621. doi:10.1186/s12889-020-08693-7
55. Browne JV, Talmi A. Family-based intervention to enhance infant-parent relationships in the neonatal intensive care unit. *J Pediatr Psychol*. Dec 2005;30(8):667-77. doi:10.1093/jpepsy/jsi053
56. Constantino JN, Hashemib N, Solisb E, et al. Supplementation of urban home visitation with a series of group meetings for parents and infants: results of a "realworld" randomized, controlled trial. *Child Abuse & Neglect*. 2001;25:1571–1581.
57. Cooper PJ, Tomlinson M, Swartz L, et al. Improving quality of mother-infant relationship and infant attachment in socioeconomically deprived community in South Africa: randomised controlled trial. *BMJ*. Apr 14 2009;338:b974. doi:10.1136/bmj.b974
58. Tomlinson M, Rabie S, Skeen S, Hunt X, Murray L, Cooper PJ. Improving mother-infant interaction during infant feeding: A randomised controlled trial in a low-income community in South Africa. *Infant Ment Health J*. Nov 2020;41(6):850-858. doi:10.1002/imhj.21881

59. Cooper PJ, De Pascalis L, Woolgar M, Romaniuk H, Murray L. Attempting to prevent postnatal depression by targeting the mother-infant relationship: a randomised controlled trial. *Prim Health Care Res Dev*. Jul 2015;16(4):383-97. doi:10.1017/S1463423614000401
60. Glazebrook C, Marlow N, Israel C, et al. Randomised trial of a parenting intervention during neonatal intensive care. *Arch Dis Child Fetal Neonatal Ed*. Nov 2007;92(6):F438-43. doi:10.1136/adc.2006.103135
61. Heo YJ, Oh WO. The effectiveness of a parent participation improvement program for parents on partnership, attachment infant growth in a neonatal intensive care unit: A randomized controlled trial. *Int J Nurs Stud*. Jul 2019;95:19-27. doi:10.1016/j.ijnurstu.2019.03.018
62. Herbers JE, Cutuli JJ, Fugo PB, Nordeen ER, Hartman MJ. Promoting parent-infant responsiveness in families experiencing homelessness. *Infant Ment Health J*. Nov 2020;41(6):811-820. doi:10.1002/imhj.21868
63. Horowitz JA, Bell M, Trybulski J, et al. Promoting Responsiveness between Mothers with Depressive Symptoms and Their Infants. *Journal of Nursing Scholarship*. 2001;33(4):323-329.
64. Horowitz JA, Murphy CA, Gregory K, Wojcik J, Pulcini J, Solon L. Nurse home visits improve maternal/infant interaction and decrease severity of postpartum depression. *J Obstet Gynecol Neonatal Nurs*. May-Jun 2013;42(3):287-300. doi:10.1111/1552-6909.12038
65. Knoche L, Sheridan SM, Clark BL, et al. GETTING READY: RESULTS OF A RANDOMIZED TRIAL OF A RELATIONSHIP-FOCUSED INTERVENTION ON THE PARENT–INFANT RELATIONSHIP IN RURAL EARLY HEAD START. *Educational Psychology Papers and Publications*. 2012;207
66. Kristensen IH, Juul S, Kronborg H. What are the effects of supporting early parenting by newborn behavioral observations (NBO)? A cluster randomised trial. *BMC Psychol*. Oct 16 2020;8(1):107. doi:10.1186/s40359-020-00467-5
67. Lavallée A, Côté J, Luu TM, et al. Acceptability and feasibility of a nursing intervention to promote sensitive mother-infant interactions in the NICU. *Journal of Neonatal Nursing*. 2022;doi:10.1016/j.jnn.2022.07.011
68. Meijssen D, Wolf MJ, Koldewijn K, et al. The effect of the Infant Behavioral Assessment and Intervention Program on mother-infant interaction after very preterm birth. *J Child Psychol Psychiatry*. Nov 2010;51(11):1287-95. doi:10.1111/j.1469-7610.2010.02237.x
69. Meijssen D, Wolf MJ, van Bakel H, Koldewijn K, Kok J, van Baar A. Maternal attachment representations after very preterm birth and the effect of early intervention. *Infant Behav Dev*. Feb 2011;34(1):72-80. doi:10.1016/j.infbeh.2010.09.009
70. Meijssen DE, Wolf MJ, Koldewijn K, van Wassenaer AG, Kok JH, van Baar AL. Parenting stress in mothers after very preterm birth and the effect of the Infant Behavioural Assessment and Intervention Program. *Child Care Health Dev*. Mar 2011;37(2):195-202. doi:10.1111/j.1365-2214.2010.01119.x
71. Melnyk BM, Feinstein NF, Alpert-Gillis L, et al. Reducing premature infants' length of stay and improving parents' mental health outcomes with the Creating Opportunities for Parent Empowerment (COPE) neonatal intensive care unit program: a randomized, controlled trial. *Pediatrics*. Nov 2006;118(5):e1414-27. doi:10.1542/peds.2005-2580
72. Nugent JK, Bartlett JD, Von Ende A, Valim C. The Effects of the Newborn Behavioral Observations (NBO) System on Sensitivity in Mother–Infant Interactions. *Infants & Young Children*. 2017;30(4):257-268. doi:10.1097/iyc.0000000000000103
73. Pontoppidan M, Klest SK, Sandoy TM. The Incredible Years Parents and Babies Program: A Pilot Randomized Controlled Trial. *PLoS One*. 2016;11(12):e0167592. doi:10.1371/journal.pone.0167592
74. Pontoppidan M, Thorsager M, Steenhoff T. Parent-child Relationship Outcomes of the Incredible Years Parents and Babies Program: A Pilot Randomized Controlled Trial. *Scand J Child Adolesc Psychiatr Psychol*. Jan 2022;10(1):40-52. doi:10.2478/sjcapp-2022-0004
75. Salo SJ, Flykt M, Makela J, et al. The effectiveness of Nurture and Play: a mentalisation-based parenting group intervention for prenatally depressed mothers. *Prim Health Care Res Dev*. Dec 16 2019;20:e157. doi:10.1017/S1463423619000914
76. Schuler ME, Nair P, Black MM, Kettinger L. Mother-infant interaction: effects of a home intervention and ongoing maternal drug use. *J Clin Child Psychol*. Sep 2000;29(3):424-31. doi:10.1207/S15374424JCCP2903_13
77. Schuler ME, Nair P, Black MM. Ongoing Maternal Drug Use, Parenting Attitudes, and a Home Intervention: Effects on Mother-Child Interaction at 18 Months. *Developmental and Behavioral Pediatrics*. 2002;23(2)doi:0196-206X/00/2302-0087
78. Tsivos ZL, Calam R, Sanders MR, Wittkowski A. A pilot randomised controlled trial to evaluate the feasibility and acceptability of the Baby Triple P Positive Parenting Programme in mothers with postnatal depression. *Clin Child Psychol Psychiatry*. Oct 2015;20(4):532-54. doi:10.1177/1359104514531589

79. Valades J, Murray L, Bozicevic L, et al. The impact of a mother-infant intervention on parenting and infant response to challenge: A pilot randomized controlled trial with adolescent mothers in El Salvador. *Infant Ment Health J*. May 2021;42(3):400-412. doi:10.1002/imhj.21917
80. Vargas-Porras C, Roa-Diaz ZM, Hernandez-Hincapie HG, Ferre-Grau C, de Molina-Fernandez MI. Efficacy of a multimodal nursing intervention strategy in the process of becoming a mother: A randomized controlled trial. *Res Nurs Health*. Jun 2021;44(3):424-437. doi:10.1002/nur.22123
81. Zelkowitz P, Feeley N, Shrier I, et al. The Cues and Care Randomized Controlled Trial of a Neonatal Intensive Care Unit Intervention: Effects on Maternal Psychological Distress and Mother-Infant Interaction. *Journal of Developmental and Behavioral Pediatrics*. 2011;32:591–599.
82. Feeley N, Zelkowitz P, Shrier I, et al. Follow-Up of the Cues and Care Trial: Mother and Infant Outcomes at 6 Months. *Journal of Early Intervention*. 2012;34(2)doi:10.1177/1053815112453767
83. Alarcao FSP, Shephard E, Fatori D, et al. Promoting mother-infant relationships and underlying neural correlates: Results from a randomized controlled trial of a home-visiting program for adolescent mothers in Brazil. *Dev Sci*. Nov 2021;24(6):e13113. doi:10.1111/desc.13113
84. Fatori D, Fonseca Zuccolo P, Shephard E, et al. A randomized controlled trial testing the efficacy of a Nurse Home Visiting Program for Pregnant Adolescents. *Sci Rep*. Jul 13 2021;11(1):14432. doi:10.1038/s41598-021-93938-7
85. Berlin LJ, Martoccio TL, Appleyard Carmody K, et al. Can typical US home visits affect infant attachment? Preliminary findings from a randomized trial of Healthy Families Durham. *Attach Hum Dev*. Dec 2017;19(6):559-579. doi:10.1080/14616734.2017.1339359
86. Berlin LJ, Martoccio TL, Jones Harden B. Improving early head start's impacts on parenting through attachment-based intervention: A randomized controlled trial. *Dev Psychol*. Dec 2018;54(12):2316-2327. doi:10.1037/dev0000592
87. Bernard K, Dozier M, Bick J, Lewis-Morrarty E, Lindhiem O, Carlson E. Enhancing attachment organization among maltreated children: results of a randomized clinical trial. *Child Dev*. Mar-Apr 2012;83(2):623-36. doi:10.1111/j.1467-8624.2011.01712.x
88. Bick J, Dozier M. The Effectiveness of an Attachment-Based Intervention in Promoting Foster Mothers' Sensitivity toward Foster Infants. *Infant Ment Health J*. Mar 1 2013;34(2):95-103. doi:10.1002/imhj.21373
89. Dozier M, Lindhiem O, Lewis E, Bick J, Bernard K, Peloso E. Effects of a Foster Parent Training Program on Young Children's Attachment Behaviors: Preliminary Evidence from a Randomized Clinical Trial. *Child Adolesc Social Work J*. Aug 2009;26(4):321-332. doi:10.1007/s10560-009-0165-1
90. Fonagy P, Sleed M, Baradon T. Randomized Controlled Trial of Parent-Infant Psychotherapy for Parents with Mental Health Problems and Young Infants. *Infant Ment Health J*. Mar-Apr 2016;37(2):97-114. doi:10.1002/imhj.21553
91. Georg AK, Cierpka M, Schroder-Pfeifer P, Kress S, Taubner S. The Efficacy of Brief Parent-Infant Psychotherapy for Treating Early Regulatory Disorders: A Randomized Controlled Trial. *J Am Acad Child Adolesc Psychiatry*. Jun 2021;60(6):723-733. doi:10.1016/j.jaac.2020.06.016
92. Perrone L, Imrisek SD, Dash A, Rodriguez M, Monticciolo E, Bernard K. Changing parental depression and sensitivity: Randomized clinical trial of ABC's effectiveness in the community. *Dev Psychopathol*. Aug 2021;33(3):1026-1040. doi:10.1017/S0954579420000310
93. Sadler LS, Slade A, Close N, et al. Minding the Baby: Enhancing reflectiveness to improve early health and relationship outcomes in an interdisciplinary home visiting program. *Infant Ment Health J*. Sep 1 2013;34(5):391-405. doi:10.1002/imhj.21406
94. Londono Tobon A, Condon E, Sadler LS, Holland ML, Mayes LC, Slade A. School age effects of Minding the Baby-An attachment-based home-visiting intervention-On parenting and child behaviors. *Dev Psychopathol*. Feb 2022;34(1):55-67. doi:10.1017/S0954579420000905
95. Slade A, Holland ML, Ordway MR, et al. Minding the Baby(R): Enhancing parental reflective functioning and infant attachment in an attachment-based, interdisciplinary home visiting program. *Dev Psychopathol*. Jan 14 2019:1-15. doi:10.1017/s0954579418001463
96. Santelices MP, Guzman GM, Aracena M, et al. Promoting secure attachment: evaluation of the effectiveness of an early intervention pilot programme with mother-infant dyads in Santiago, Chile. *Child Care Health Dev*. Mar 2011;37(2):203-10. doi:10.1111/j.1365-2214.2010.01161.x
97. Sleed M, Baradon T, Fonagy P. New beginnings for mothers and babies in prison: A cluster randomized controlled trial. *Attachment & Human Development*. 2013;15(4):349-367. doi:10.1080/14616734.2013.782651

98. Steele H, Murphy A, Bonuck K, Meissner P, Steele M. Randomized control trial report on the effectiveness of Group Attachment-Based Intervention (GABI(c)): Improvements in the parent-child relationship not seen in the control group. *Dev Psychopathol*. Feb 2019;31(1):203-217. doi:10.1017/s0954579418001621
99. Walter I, Landers S, Quehenberger J, Carlson E, Brisch KH. *The efficacy of the attachment-based SAFE(R) prevention program: a randomized control trial including mothers and fathers. *Attach Hum Dev*. Oct 2019;21(5):510-531. doi:10.1080/14616734.2019.1582599
100. Badr LK, Garg M, Kamath M. Intervention for infants with brain injury: results of a randomized controlled study. *Infant Behav Dev*. Jan 2006;29(1):80-90. doi:10.1016/j.infbeh.2005.08.003
101. Milgrom J, Newnham C, Martin PR, et al. Early communication in preterm infants following intervention in the NICU. *Early Hum Dev*. Sep 2013;89(9):755-62. doi:10.1016/j.earlhumdev.2013.06.001
102. Milgrom J, Martin PR, Newnham C, et al. Behavioural and cognitive outcomes following an early stress-reduction intervention for very preterm and extremely preterm infants. *Pediatr Res*. Jul 2019;86(1):92-99. doi:10.1038/s41390-019-0385-9
103. Newnham CA, Milgrom J, Skouteris H. Effectiveness of a modified Mother-Infant Transaction Program on outcomes for preterm infants from 3 to 24 months of age. *Infant Behav Dev*. Jan 2009;32(1):17-26. doi:10.1016/j.infbeh.2008.09.004
104. Ravn IH, Smith L, Lindemann R, et al. Effect of early intervention on social interaction between mothers and preterm infants at 12 months of age: a randomized controlled trial. *Infant Behav Dev*. Apr 2011;34(2):215-25. doi:10.1016/j.infbeh.2010.11.004
105. Youn YA, Shin SH, Kim EK, et al. Preventive Intervention Program on the Outcomes of Very Preterm Infants and Caregivers: A Multicenter Randomized Controlled Trial. *Brain Sci*. Apr 29 2021;11(5)doi:10.3390/brainsci11050575
106. Holditch-Davis D, White-Traut RC, Levy JA, O'Shea TM, Geraldo V, David RJ. Maternally administered interventions for preterm infants in the NICU: effects on maternal psychological distress and mother-infant relationship. *Infant Behav Dev*. Nov 2014;37(4):695-710. doi:10.1016/j.infbeh.2014.08.005
107. Nelson MN, White-Traut RC, Vasan U, et al. One-Year Outcome of Auditory-Tactile-Visual-Vestibular Intervention in the Neonatal Intensive Care Unit: Effects of Severe Prematurity and Central Nervous System Injury. *Journal of Child Neurology*. 2001;16:493-498).
108. White-Traut R, Norr KF, Fabiyi C, Rankin KM, Li Z, Liu L. Mother-infant interaction improves with a developmental intervention for mother-preterm infant dyads. *Infant Behav Dev*. Dec 2013;36(4):694-706. doi:10.1016/j.infbeh.2013.07.004
109. Ericksen J, Loughlin E, Holt C, et al. A Therapeutic Playgroup for Depressed Mothers and Their Infants: Feasibility Study and Pilot Randomized Trial of Community Hugs. *Infant Ment Health J*. Jul 2018;39(4):396-409. doi:10.1002/imhj.21723
110. Holt C, Gentilleau C, Gemmill AW, Milgrom J. Improving the mother-infant relationship following postnatal depression: a randomised controlled trial of a brief intervention (HUGS). *Arch Womens Ment Health*. Dec 2021;24(6):913-923. doi:10.1007/s00737-021-01116-5
111. *EndNote*. Version EndNote X9. Clarivate; 2013.
112. *Covidence systematic review software*. Veritas Health Innovation; 2022. www.covidence.org
113. Hoffmann TC, Glasziou PP, Boutron I, et al. Better reporting of interventions: template for intervention description and replication (TIDieR) checklist and guide. *BMJ*. Mar 7 2014;348:g1687. doi:10.1136/bmj.g1687
114. Tereno S, Guedeney N, Dugravier R, et al. Implementation and assessment of an early home-based intervention on infant attachment organisation: the CAPEDP attachment study in France. *Global Health Promotion*. 2013;20(2_suppl):71-75. doi:10.1177/1757975913483337
115. Higgins JPT, Li T, Deeks JJ. Chapter 6: Choosing effect measures and computing estimates of effect. In: Higgins JPT, Thomas J, Chandler J, et al, eds. *Cochrane Handbook for Systematic Reviews of Interventions*. Cochrane; 2022.
116. *Engauge Digitizer*. Version 12.1. 2020. https://apps.apple.com/us/app/engauge-digitizer/id1107356098?mt=12
117. *PlotDigitizer*. 2022. https://plotdigitizer.com/app
118. Higgins JPT, Eldridge S, Li T. Chapter 23: Including variants on randomized trials. In: Higgins JPT, Thomas J, Chandler J, et al, eds. *Cochrane Handbook for Systematic Reviews of Interventions version 63*. Cochrane; 2022. www.training.cochrane.org/handbook
119. *SPSS Meta-Analysis Macro*. 2021. http://mason.gmu.edu/~dwilsonb/MetaAnal.html
120. *Review Manager (RevMan)*. Version 5.4. 2020.
121. *IBM SPSS Statistics for Macintosh*. Version 28. IBM Corp; 2021.

122. Higgins JPT, Savović J, Page MJ, Elbers RG, Sterne JAC. Chapter 8: Assessing risk of bias in a randomized trial. In: Higgins J, Thomas J, Chandler J, et al, eds. *Cochrane Handbook for Systematic Reviews of Interventions*. 6.3 ed. Cochrane; 2022.
123. Guyatt G, Oxman AD, Akl EA, et al. GRADE guidelines: 1. Introduction-GRADE evidence profiles and summary of findings tables. *J Clin Epidemiol*. Apr 2011;64(4):383-94. doi:10.1016/j.jclinepi.2010.04.026
124. Stovall KC, Dozier M. The development of attachment in new relationships: single subject analyses for 10 foster infants. *Dev Psychopathol*. Spring 2000;12(2):133-56. doi:10.1017/s0954579400002029
125. Müller ME. A questionnaire to measure mother-to-infant attachment. *Journal of nursing measurement*. 1994;2(2):129–141.
126. Shieh PL, Ying TH, Li RH, Hsieh PC. The development of mother-infant bonding inventory. *Bullentin of Educational Psychology*. 2015;47(1):133–158. doi:10.6251/bep.20141208.
127. Furman L, O'Riordan MA. How Do Mothers Feel About Their Very Low Birth Weight Infants? Development of a New Measure. *Infant Ment Health J*. Mar 2006;27(2):152-172. doi:10.1002/imhj.20086
128. Brockington I, Oates J, George S. A Screening Questionnaire for mother-infant bonding disorders. *Arch Womens Ment Health*. 2001;3:133–140. doi:10.1007/s007370170010
129. Brockington IF, Fraser C, Wilson D. The Postpartum Bonding Questionnaire: a validation. *Arch Womens Ment Health*. Sep 2006;9(5):233-42. doi:10.1007/s00737-006-0132-1
130. Zeanah CH, Benoit D. Clinical Applications of a Parent Perception Interview in Infant Mental Health. *Child and Adolescent Psychiatric Clinics of North America*. 1995/07/01/ 1995;4(3):539-554. doi:https://doi.org/10.1016/S1056-4993(18)30418-8
131. Feldman R, Weller A, Leckman JF, Kuint J, Eidelman AI. The nature of the mother's tie to her infant: maternal bonding under conditions of proximity, separation, and potential loss. *Journal of child psychology and psychiatry, and allied disciplines*. 1999;40(6):929–939.
132. Shin H, Kim YH. Maternal Attachment Inventory: psychometric evaluation of the Korean version. *J Adv Nurs*. Aug 2007;59(3):299-307. doi:10.1111/j.1365-2648.2007.04322.x
133. Vargas Vásquez AN, Pardo Torres MP. Validez y consistencia interna del instrumento Vínculo entre padres e hijos neonatos. *Enfermería Global*. 2020;19(59):255-285.
134. Taylor A, Atkins R, Kumar R, Adams D, Glover V. A new Mother-to-Infant Bonding Scale: links with early maternal mood. *Archives of women's mental health*. 2005;8(1):45–51. doi:10.1007/s00737-005-0074-z
135. Mathews TL, Emerson MR, Moore TA, Fial A, Hanna KM. Systematic Review: Feasibility, Reliability, and Validity of Maternal/Caregiver Attachment and Bonding Screening Tools for Clinical Use. *J Pediatr Health Care*. Nov - Dec 2019;33(6):663-674. doi:10.1016/j.pedhc.2019.04.018
136. Condon JT, Corkindale CJ. The assessment of parent-to-infant attachment: development of a self-report questionnaire instrument. *Journal of Reproductive and Infant Psychology*. 1998;16(1):57-76.
137. McEachern AD, Dishion TJ, Weaver CM, Shaw DS, Wilson MN, Gardner F. Parenting Young Children (PARYC): Validation of a self-report parenting measure. *Journal of child and family studies*. 2012;21(3):498-511.
138. Wittkowski A, Williams J, Wieck A. An examination of the psychometric properties and factor structure of the Post-partum Bonding Questionnaire in a clinical inpatient sample. *Br J Clin Psychol*. Jun 2010;49(Pt 2):163-72. doi:10.1348/014466509X445589
139. Reck C, Klier CM, Pabst K, et al. The German version of the Postpartum Bonding Instrument: psychometric properties and association with postpartum depression. *Arch Womens Ment Health*. Sep 2006;9(5):265-71. doi:10.1007/s00737-006-0144-x
140. Kaneko H, Honjo S. The Psychometric Properties and Factor Structure of the Postpartum Bonding Questionnaire in Japanese Mothers. *Psychology*. 2014;05(09):1135-1142. doi:10.4236/psych.2014.59126
141. Ohashi Y, Kitamura T, Sakanashi K, Tanaka T. Postpartum Bonding Disorder: Factor Structure, Validity, Reliability and a Model Comparison of the Postnatal Bonding Questionnaire in Japanese Mothers of Infants. *Healthcare (Basel)*. Aug 2 2016;4(3)doi:10.3390/healthcare4030050
142. Garcia-Esteve L, Torres A, Lasheras G, et al. Assessment of psychometric properties of the Postpartum Bonding Questionnaire (PBQ) in Spanish mothers. *Arch Womens Ment Health*. Apr 2016;19(2):385-94. doi:10.1007/s00737-015-0589-x
143. Busonera A, Cataudella S, Lampis J, Tommasi M, Zavattini GC. Psychometric properties of the Postpartum Bonding Questionnaire and correlates of mother-infant bonding impairment in Italian new mothers. *Midwifery*. Dec 2017;55:15-22. doi:10.1016/j.midw.2017.08.011
144. Vengadavaradan A, Bharadwaj B, Sathynarayanan G, Durairaj J, Rajaa S. Translation, validation and factor structure of the Tamil version of the Postpartum Bonding Questionnaire (PBQ-T). *Asian J Psychiatr*. Feb 2019;40:62-67. doi:10.1016/j.ajp.2019.01.018

145. Demanche A, Michel L, Chabbert M, Wendland J. Version française du Postpartum Bonding Questionnaire. *Devenir*. 2022;Vol. 33(4):287-310. doi:10.3917/dev.214.0287
146. Lotzin A, Lu X, Kriston L, et al. Observational tools for measuring parent-infant interaction: a systematic review. *Clin Child Fam Psychol Rev*. Jun 2015;18(2):99-132. doi:10.1007/s10567-015-0180-z
147. Howes C, Stewart P. Child's play with adults, toys, and peers: An examination of family and child-care influences. *Developmental Psychology*. 1987;23(3):423–430. doi:10.1037/0012-1649.23.3.423
148. Ainsworth MD, Bell SM, Stayton DJ. Individual differences in strange-situation behaviour of one-year-olds. In: Schaffer HR, ed. *The origins of human social relations*. Academic Press; 1971.
149. Ainsworth MDS, Bell SM, Stayton DF. Infant-mother attachment and social development: Socialization as a product of reciprocal responsiveness to signals. *The integration of a child into a social world*. Cambridge University Press; 1974:99-135.
150. Waters E. Appendix A: The attachment Q-set (version 3.0). *Monographs of the society for research in child development*. 1995:234-246.
151. Verschueren K, Marcoen A. Attachment story completion task classification system Unpublished Manual. Belgium: University of Louvain; 1994.
152. Bretherton I, Ridgeway D, Cassidy J. Assessing internal working models of the attachment relationship: An attachment story completion task for 3-year-olds. In: Greenberg MT, Cicchetti D, Cummings EM, eds. *Attachment in the preschool years: Theory, research, and intervention*. The University of Chicago Press; 1990:273–308.
153. Bronfman E, Madigan S, Lyons-Ruth K. *The Atypical Behavior Instrument for Assessment and Classification*. Harvard Medical School; 1992–2009.
154. Bronfman ET, Parsons E, Lyons-Ruth K. *Disrupted Maternal Behavior Instrument for Assessment and Classification (AMBIANCE): Manual for coding disrupted affective communication*. 1st ed. Harvard University Medical School; 1992-2008.
155. Avant PKC. *Maternal attachment and anxiety: An exploratory study*. Graduate School of the Texas Woman's University; 1978.
156. Goldsmith H, Rothbart M. The laboratory temperament assessment battery (LAB-TAB). University of Wisconsin1993.
157. Crittenden PM. CARE-Index: Coding Manual. Unpublished manuscript. Miami, FL1979-2004.
158. Crittenden PM. CARE-index: Infant Coding Manual. Miami FL: Family Relations Institute; 1979-2010.
159. Feldman R. Coding interactive behavior manual. Bar-Ilan University, Israel1998.
160. Piccinini C, Alvarenga P, Frizzo G. A responsividade como foco da análise da interação mãe-bebê e pai-bebê. *Observando a interação pais-bebê-criança*. 2007:131-153.
161. Beebe B, Jaffe J, Markese S, et al. The origins of 12-month attachment: a microanalysis of 4-month mother-infant interaction. *Attach Hum Dev*. Jan 2010;12(1-2):3-141. doi:10.1080/14616730903338985
162. Murray L, Fiori-Cowley A, Hooper R, Cooper P. The impact of postnatal depression and associated adversity on early mother-infant interactions and later infant outcome. *Child Dev*. Oct 1996;67(5):2512-26.
163. Holditch-Davis D, Schwartz T, Black B, Scher M. Correlates of mother-premature infant interactions. *Res Nurs Health*. Jun 2007;30(3):333-46. doi:10.1002/nur.20190
164. Holditch-Davis D, Miles MS, Burchinal MR, Goldman BD. Maternal role attainment with medically fragile infants: Part 2. Relationship to the quality of parenting. *Research in nursing & health*. 2011;34(1):35-48.
165. Caldwell B, Bradley R. Home observation for measurement of the environment. Little Rock, AK: University of Arkansas; 1978.
166. Melnyk B, Feinstein NF, Fairbanks E, Small L. The Index of Parental Behavior in the NICU. 1998
167. Weinberg MK, Tronick EZ. Infant and Caregiver Engagement Phases (ICEP). 1999
168. Landry SH, Smith KE. Playing and Learning Strategies. 1996
169. Landry SH, Smith KE, Miller-Loncar CL, Swank PR. The relation of change in maternal interactive styles to the developing social competence of full-term and preterm children. *Child Dev*. Feb 1998;69(1):105-23.
170. Beatty JR, Stacks AM, Partridge T, Tzilos GK, Loree A, Ondersma SJ. LoTTS parent-infant interaction coding scale: East of use and reliability in a sample of high-risk mothers and their infants. *Children and Youth Services Review*. 2011;33(1):86-90.
171. Pederson DR, Moran G. Appendix B: Maternal Behavior Q-set. In: Waters E, Vaughn BE, Posada G, Kondo-Ikemura KK, eds. *Caregiving, cultural, and cognitive perspectives on secure-base behavior and working models: New growing points of attachment theory and research*. 1995:247-254. *Monographs of the Society for Research in Child Development*.
172. Hane AA, Fox NA. Ordinary variations in maternal caregiving influence human infants' stress reactivity. *Psychol Sci*. Jun 2006;17(6):550-6. doi:10.1111/j.1467-9280.2006.01742.x

173. Hane AA, Philbrook LE. Beyond Licking and Grooming: Maternal Regulation of Infant Stress in the Context of Routine Care. *Parent Sci Pract*. 2012;12(2-3):144-153. doi:10.1080/15295192.2012.683341
174. Erickson MF, Sroufe LA, Egeland B. The relationship between quality of attachment and behavior problems in preschool in a high-risk sample. *Monogr Soc Res Child Dev*. 1985;50(1-2):147-66.
175. Smeekens S, Riksen-Walraven JM, Van Bakel HJA. Profiles of competence and adaptation in preschoolers as related to the quality of parent-child interaction. *Journal Of Research in Personality*. 2008;42:1490-1499.
176. Cenciotti F, Tronick EZ, Reck C. Maternal sensitivity and responsivity Scales-R. . 2004
177. Moran G. Mini-MBQS-V Revised Mini-MBQS 25 items for Video Coding. Ontario, CA: Western University; 2009.
178. Ramsauer B, Muhlhan C, Lotzin A, et al. Randomized controlled trial of the Circle of Security-Intensive intervention for mothers with postpartum depression: maternal unresolved attachment moderates changes in sensitivity. *Attach Hum Dev*. Dec 2019;22(6):705-726. doi:10.1080/14616734.2019.1689406
179. Cowan P, Cowan C. School children and their families project: Description of child-style ratings. Berkeley University of California, Department of Psychology; 1992.
180. Cowan P, Cowan C. Schoolchildren and their families project: Description o parent-style ratings. Berkeley, CA: University of California, Department of Psychology; 1992.
181. Schuler ME, Nair P, Black MM. Ongoing Maternal Drug Use, Parenting Attitudes and a Home Intervention: Effects on Mother-Child Interaction at 18 Months. *Developmental and Behavioral Pediatrics*. 2002;23(2):87-94.
182. Kochanska G. Mutually responsive orientation between mothers and their young children: A context for the early development of conscience. *Current Directions in Psychological Science*. 2002;11:191-195.
183. Child care and mother-child interaction in the first 3 years of life. NICHD Early Child Care Research Network. *Dev Psychol*. Nov 1999;35(6):1399-413.
184. Network. NNIoCHaHDECCR. Early child care and mother–child interaction from 36 months through first grade. . *Infant Behavior and Development*. 2003;26:345-370. doi:doi:10.1016/s0163-6383(03)00035-3
185. Sumner GA, Spietz A. NCAST caregiver/parent-child interaction feeding manual. NCAST Publications; 1995.
186. Ukeje I, Bendersky M, Lewis M. Mother-infant interaction at 12 months in prenatally cocaine-exposed children. *Am J Drug Alcohol Abuse*. May 2001;27(2):203-24. doi:10.1081/ada-100103706
187. Clark R. The parent-child early relational assessment: Instrument and manual. 2015
188. Clark R. The parent-child early relational assessment: A factorial validity study. . *Educational and Psychological Measurement*. 1999;59:821-846.
189. Fiese B, Poehlmann J, Irwin M, Gordon M, Curry-Bleggi E. A pediatric screening instrument to detect problematic infant-parent interactions: Initial reliability and validity in a sample of high- and low-risk infants. *Infant Mental Health Journal*. 2001;22:463-478.
190. Ainsworth MDS, Blehar MC, Waters E, Wall S. *Patterns of attachment: A psychological study of the Strange Situation*. Erlbaum; 1978.
191. Dreyer BP, Mendelsohn AL, Samis-LeMonda CS. Assessing the child's cognitive home environment through parental report: Reliability and validity. Early Developmental Parenting; 1996. p. 271-287.
192. Mendelsohn AL, Cates CB, Tamis-LeMonda CS, et al. Assessment of the cognitive home environment through parent report: Reliability and validity of the StimQ (revised). 2011:
193. Milgrom J, Meitz A. Synchrony coding scale in maternal perceptions of infant temperament, maternal compentency, mothering style, and later mother-infant synchrony. Melbourne, Australia: Melbourne College of Advanced Education; 1988.
194. Song M-K, Lin F-C, Ward SE, Fine JP. Composite variables: when and how. *Nursing research*. 2013;62(1):45.
195. Waters E, Deane KE. Defining and assessing individual differences in attachment relationships: Q-methodology and the organization of behavior in infancy and early childhood. *Monographs of the Society for Research in Child Development*. 1985;50(1-2):41-65. doi:10.2307/3333826
196. Van Ijzendoorn MH, Vereijken CM, Bakermans-Kranenburg MJ, Marianne Riksen-Walraven J. Assessing attachment security with the attachment Q sort: Meta-analytic evidence for the validity of the observer AQS. *Child development*. 2004;75(4):1188-1213.
197. Cassidy J. Child-mother attachment and the self in six-year-olds. *Child development*. 1988:121-134.
198. Verschueren K, Marcoen A, Schoefs V. The internal working model of the self, attachment, and competence in five-year-olds. *Child development*. 1996;67(5):2493-2511.

199. Lyons-Ruth K, Bronfman E, Parsons E. Maternal frightened, frightening, or atypical behavior and disorganized infant attachment patterns. *Monographs of the society for research in child development*. 1999:67-96.
200. Kelly K, Slade A, Lyons-Ruth K. Atypical Maternal Behavior Instrument for Assessment and Classification (AMBIANCE): An adaptation for four month old babies and their mothers. *City University of New York*. 2003;
201. Bronfman E, Parsons E, Lyons-Ruth K. *Atypical Maternal Behavior Instrument for Assessment and Classification (AMBIANCE): Manual for coding disrupted affective communication, version 2.* Harvard University School of Medicine; 1999.
202. Lyons-Ruth K, Bronfman E, Atwood G. A relational diathesis model of hostile-helpless states of mind: Expressions in mother–infant interaction. 1999;
203. Madigan S, Bakermans-Kranenburg MJ, Van Ijzendoorn MH, Moran G, Pederson DR, Benoit D. Unresolved states of mind, anomalous parental behavior, and disorganized attachment: A review and meta-analysis of a transmission gap. *Attachment & human development*. 2006;8(2):89-111.
204. Dutra L, Bureau J-F, Holmes B, Lyubchik A, Lyons-Ruth K. Quality of early care and childhood trauma: A prospective study of developmental pathways to dissociation. *The Journal of nervous and mental disease*. 2009;197(6):383.
205. Madigan S, Moran G, Schuengel C, Pederson DR, Otten R. Unresolved maternal attachment representations, disrupted maternal behavior and disorganized attachment in infancy: Links to toddler behavior problems. *Journal of child psychology and psychiatry*. 2007;48(10):1042-1050.
206. Shi Z, Bureau JF, Easterbrooks MA, Zhao X, Lyons-Ruth K. Childhood maltreatment and prospectively observed quality of early care as predictors of antisocial personality disorder features. *Infant Mental Health Journal*. 2012;33(1):55-69.
207. Avant KC. Anxiety as a potential factor affecting maternal attachment. *Journal of Obstetric, Gynecologic, & Neonatal Nursing*. 1981;10(6):416-419.
208. Bozicevic L, De Pascalis L, Schuitmaker N, Tomlinson M, Cooper PJ, Murray L. Longitudinal association between child emotion regulation and aggression, and the role of parenting: a comparison of three cultures. *Psychopathology*. 2016;49(4):228-235.
209. Crittenden PM. Distorted patterns of relationship in maltreating families: The role of internal representation models. *Journal of Reproductive and Infant Psychology*. 1988;6(3):183-199.
210. Mullick M, Miller LJ, Jacobsen T. Insight into mental illness and child maltreatment risk among mothers with major psychiatric disorders. *Psychiatric services*. 2001;52(4):488-492.
211. Crittenden PM, Bonvillian JD. The relationship between maternal risk status and maternal sensitivity. *American Journal of Orthopsychiatry*. 1984;54(2):250.
212. Feldman R. Coding Interactive Behaviour Manual. Unpublished manuscript. 1998
213. Feldman R, Eidelman AI. Direct and indirect effects of breast milk on the neurobehavioral and cognitive development of premature infants. *Developmental Psychobiology: The Journal of the International Society for Developmental Psychobiology*. 2003;43(2):109-119.
214. Feldman R, Eidelman AI, Rotenberg N. Parenting stress, infant emotion regulation, maternal sensitivity, and the cognitive development of triplets: A model for parent and child influences in a unique ecology. *Child development*. 2004;75(6):1774-1791.
215. Ferber SG, Feldman R. Delivery pain and the development of mother–infant interaction. *Infancy*. 2005;8(1):43-62.
216. Ferber SG, Feldman R, Kohelet D, et al. Massage therapy facilitates mother–infant interaction in premature infants. *Infant Behavior and Development*. 2005;28(1):74-81.
217. Mayes LC, Feldman R, Granger RH, Haynes OM, Bornstein MH, Schottenfeld R. The effects of polydrug use with and without cocaine on mother-infant interaction at 3 and 6 months. *Infant Behavior and Development*. 1997;20(4):489-502.
218. Censullo M. Dyadic mutuality code manual. *Wellesley, MA: Wellesley College Center for Research on Women*. 1991;
219. Censullo M, Bowler R, Lester B, Brazelton TB. An instrument for the measurement of infant-adult synchrony. *Nursing Research*. 1987;36(4):244–248. doi:10.1097/00006199-198707000-00015
220. Coffman S, Levitt MJ, Deets C, Quigley KL. Close relationships in mothers of distressed and normal newborns: Support, expectancy confirmation, and maternal well-being. *Journal of Family Psychology*. 1991;5(1):93.
221. Lester BM, Hoffman J, Brazelton TB. The rhythmic structure of mother-infant interaction in term and preterm infants. *Child development*. 1985:15-27.

222. Tronick E, Als H, Brazelton TB. Monadic phases: A structural descriptive analysis of infant-mother face to face interaction. *Merrill-Palmer Quarterly of Behavior and Development*. 1980;26(1):3-24.
223. Biringen Z. The Emotional Availability (EA) Scales and EA Zones Evaluation (4th ed.). Boulder, Colorado2008.
224. Wurster HE, Sarche M, Trucksess C, Morse B, Biringen Z. Parents' adverse childhood experiences and parent-child emotional availability in an American Indian community: Relations with young children's social-emotional development. *Dev Psychopathol*. May 2020;32(2):425-436. doi:10.1017/S095457941900018X
225. Biringen Z, Robinson JL, Emde RN. *The Emotional Availability Scale*. 3 ed. Fort Collins; 1998.
226. Salo S, Booth P. The MIM Handbook. Chicago: Theraplay Institute2019.
227. Salo S, Flykt M, Biringen Z. Pregnancy version of the Emotional Availability Scales (Pre-EA). 2016
228. Saunders H, Kraus A, Barone L, Biringen Z. Emotional availability: theory, research, and intervention. *Front Psychol*. 2015;6:1069. doi:10.3389/fpsyg.2015.01069
229. Easterbrooks MA, Bureau JF, Lyons-Ruth K. Developmental correlates and predictors of emotional availability in mother-child interaction: a longitudinal study from infancy to middle childhood. *Dev Psychopathol*. Feb 2012;24(1):65-78. doi:10.1017/s0954579411000666
230. Ziv Y, Aviezer O, Gini M, Sagi A, Koren-Karie N. Emotional availability in the mother-infant dyad as related to the quality of infant-mother attachment relationship. *Attach Hum Dev*. Sep 2000;2(2):149-69. doi:10.1080/14616730050085536
231. Gunning M, Conroy S, Valoriani V, et al. Measurement of mother-infant interactions and the home environment in a European setting: preliminary results from a cross-cultural study. *Br J Psychiatry Suppl*. Feb 2004;46:s38-44. doi:10.1192/bjp.184.46.s38
232. Abbott K. *Attachment during the first year of life: Validity and longitudinal assocaitions to 14 months of attachment classification at 7 months*. University of Liverpool; 2016.
233. Lee TY, Holditch-Davis D, Miles MS. The influence of maternal and child characteristics and paternal support on interactions of mothers and their medically fragile infants. *Res Nurs Health*. Feb 2007;30(1):17-30. doi:10.1002/nur.20184
234. Bradley RH, Mundfrom DJ, Whiteside L, et al. A reexamination of the association between HOME scores and income. *Nurs Res*. Sep-Oct 1994;43(5):260-6.
235. Holditch-Davis D, Tesh EM, Goldman BD, Miles MS, D'Auria J. Use of the HOME Inventory with medically fragile infants. *Children's Health Care*. 2000;29(4):257-277.
236. Tesh EM, Holditch-Davis D. HOME inventory and NCATS: relation to mother and child behaviors during naturalistic observations. Home Observation for Measurement of the Environment. Nursing Child Assessment Teaching Scale. *Res Nurs Health*. Aug 1997;20(4):295-307. doi:10.1002/(sici)1098-240x(199708)20:4<295::aid-nur3>3.0.co;2-b
237. Landry SH, Smith KE, Swank PR, Assel MA, Vellet S. Does early responsive parenting have a special importance for children's development or is consistency across early childhood necessary? *Dev Psychol*. May 2001;37(3):387-403. doi:10.1037//0012-1649.37.3.387
238. Landry SH, Smith KE, Swank PR, Guttentag C. A responsive parenting intervention: the optimal timing across early childhood for impacting maternal behaviors and child outcomes. *Dev Psychol*. Sep 2008;44(5):1335-53. doi:10.1037/a0013030
239. Pederson DR, Moran G. A categorical description of infant-mother relationships in the home and its relation to Q-sort measures of infant-mother interaction. *Monographs of the Society for Research in Child Development*. 1995;60(2-3):111-132.
240. Teti DM, Huang KY. Developmental perspectives on parenting competence. *Handbook of research methods in developmental science*. 2005:161-182.
241. van Bakel HJ, Riksen-Walraven JM. Parenting and development of one-year-olds: links with parental, contextual, and child characteristics. *Child Dev*. Jan-Feb 2002;73(1):256-73. doi:10.1111/1467-8624.00404
242. Aksan N, Kochanska G, Ortmann MR. Mutually responsive orientation between parents and their young children: toward methodological advances in the science of relationships. *Dev Psychol*. Sep 2006;42(5):833-48. doi:10.1037/0012-1649.42.5.833
243. Ayoub C, Vallotton CD, Mastergeorge AM. Developmental pathways to integrated social skills: the roles of parenting and early intervention. *Child Dev*. Mar-Apr 2011;82(2):583-600. doi:10.1111/j.1467-8624.2010.01549.x
244. Owen MT. The NICHD Study of Early Child Care Mother-Infant Interaction Scales. 1992
245. Cox M, Crnic K. Qualitative ratings for parent-child interaction at 3-15 months. 2003

246. Nelson MN, White-Traut RC, Vasan U, et al. One-Year OUtcome of Auditory-Tactile-Visual-Vestibular Intervention in the Neonatal Intensive Care Unit: Effects on Severe Prematurity and Central Nervous System Injury. *Journal of Child Neurology*. 2001;16(7)
247. Rasheed MA, Yousafzai AK. The development and reliability of an observational tool for assessing mother-child interactions in field studies- experience from Pakistan. *Child Care Health Dev*. Nov 2015;41(6):1161-71. doi:10.1111/cch.12287
248. Yousafzai AK, Rasheed MA, Rizvi A, Armstrong R, Bhutta ZA. Effect of integrated responsive stimulation and nutrition interventions in the Lady Health Worker programme in Pakistan on child development, growth, and health outcomes: a cluster-randomised factorial effectiveness trial. *Lancet*. Oct 4 2014;384(9950):1282-93. doi:10.1016/s0140-6736(14)60455-4
249. Yousafzai AK, Rasheed MA, Rizvi A, Armstrong R, Bhutta ZA. Parenting Skills and Emotional Availability: An RCT. *Pediatrics*. May 2015;135(5):e1247-57. doi:10.1542/peds.2014-2335
250. Knoche L, Sheridan SM, Clark BL, Edwards CP, Marvin CA. Getting Ready: Results of a Randomized Trial of a Relationship-focused Intervention on the Parent-Infant Relationship in Rural Early Head Start. *Infant Mental Health*. 2012;33(5):439-458.
251. Farran DC, Kasari C, Comfort M, Jay S. Parent/caregiver involvement scale. Greensboro, NC: University of North Carolina at Greensboro, Child Development-Family Relations, School of Human Environmental Sciences; 1986.
252. Tluczek A, Clark R, McKechnie AC, Orland KM, Brown RL. Task-oriented and bottle feeding adversely affect the quality of mother-infant interactions after abnormal newborn screens. *J Dev Behav Pediatr*. Jun 2010;31(5):414-26. doi:10.1097/DBP.0b013e3181dd5049
253. Mesman J, van Ijzendoorn MH, Bakermans-Kranenburg MJ. THe many faces of the still-face paradigm: A review and meta-analysis. *Developmental Review*. 2009;29(2):120-162. doi:https://doi.org/10.1016/j.dr.2009.02.001
254. Gunning M, Halligan SL, Murray L. Contributions of maternal and infant factors to infant responding to the still face paradigm: a longitudinal study. *Infant Behav Dev*. Jun 2013;36(3):319-28. doi:10.1016/j.infbeh.2013.02.003
255. Hill J, Pickles A, Wright N, De Pascalis L, Bozicevic L, Sharp H. Attachment security assessed in the still face and the strange situation, vagal reactivity and child irritability at age 7 years. 2018;
256. Dreyer BP, Mendelsohn AL, Tamis-LeMonda CS. Stim-Q: The cognitive home environment. http://pediatrics.med.nyu.edu/patient-care/for-healthcare-providers/stimq-cognitive-home-environment
257. Main M, Solomon J. Procedures for identifying infants as disorganized/disoriented during the Ainsworth Strange Situation. *Attachment in the preschool years: Theory, research, and intervention*. University of Chicago Press; 1990:121-160. *The John D. and Catherine T. MacArthur Foundation series on mental health and development.*
258. Censullo M, Bowler R, Lester B, Brazelton TB. An instrument for the measurement of infant-adult synchrony. *Nursing Research*. 1987;
259. Gunning M, Fiori-Cowley A, Murray L. Global rating scales of mother-infant interaction. Reading, England: Winnicott Research Unit; 1996.
260. Milgrom J, Westley DT, Gemmill AW. The mediating role of maternal responsiveness in some longer term effects of postnatal depression on infant development. *Infant Behavior and Development*. 2004/12/01/ 2004;27(4):443-454. doi:https://doi.org/10.1016/j.infbeh.2004.03.003
261. Feldman R. Bio-behavioral Synchrony: A Model for Integrating Biological and Microsocial Behavioral Processes in the Study of Parenting. *Parenting*. 2012/04/01 2012;12(2-3):154-164. doi:10.1080/15295192.2012.683342
262. Taneja S, Sinha B, Upadhyay RP, et al. Community initiated kangaroo mother care and early child development in low birth weight infants in India-a randomized controlled trial. *BMC Pediatr*. Apr 4 2020;20(1):150. doi:10.1186/s12887-020-02046-4
263. Peng JY. *Xiaoqi's yellow persimmon*. JHAOYU Independent Issue. 2012.
264. Landry SH, Smith KE, Glenn SM, Vellet NS. Joint-attention treatment training manual: the "PALS" Project- 1998 revised version for "My Baby and Me" study Houston, TX: University of Texas Health Science Center, Department of Pediatrics; 2002.
265. Kameenui EJ, Carnine DW. *Effective teaching strategies that accommodate diverse learners*. ERIC; 1998.
266. Clark MS. We should focus on interpersonal as well as intrapersonal processes in our search for how affect influences judgments and behavior. *Psychological Inquiry*. 2002;13(1):32-37.
267. Brazelton TB, Nugent JK. *Neonatal behavioral assessment scale*. Cambridge University Press; 1995.

268. Field T, Diego MA, Hernandez-Reif M, Deeds O, Figuereido B. Moderate versus light pressure massage therapy leads to greater weight gain in preterm infants. *Infant Behavior and Development*. 2006;29(4):574-578.
269. Cusson R, DeWeese M. Premie-talk: Understanding your premature baby's behavior [video-tape]. *University of Maryland at Baltimore School of Nursing*. 1992;
270. Blair C, Ramey C. Early intervention for low birth wight infants and the path to second-generation research. *The effectiveness early intervention*. Brookes Publishing; 1997:77-97.
271. Sumner G. NCAST caregiver/parent-child interaction teaching manual. Seattle. *NCAST Publications*. 1994;
272. Bader E. Lessons learned from practice: Focus on fathers. *IMPrint*. 2002;34:24.
273. Hastings PD, Kahle S, Fleming C, Lohr MJ, Katz LF, Oxford ML. An intervention that increases parental sensitivity in families referred to Child Protective Services also changes toddlers' parasympathetic regulation. *Developmental science*. 2019;22(1):e12725.
274. Aldridge VK, Dovey TM, El Hawi N, Martiniuc A, Martin CI, Meyer C. Observation and comparison of mealtime behaviors in a sample of children with avoidant/restrictive food intake disorders and a control sample of children with typical development. *Infant Mental Health Journal*. 2018;39(4):410-422.
275. Oxford ML, Fleming CB, Nelson EM, Kelly JF, Spieker SJ. Randomized trial of Promoting First Relationships: Effects on maltreated toddlers' separation distress and sleep regulation after reunification. *Children and youth services review*. 2013;35(12):1988-1992.
276. Hash JB, Oxford ML, Fleming CB, Ward TM, Spieker SJ, Lohr MJ. Impact of a home visiting program on sleep problems among young children experiencing adversity. *Child abuse & neglect*. 2019;89:143-154.
277. Koniak-Griffin D, Verzemnieks I, Cahill D. Using videotape instruction and feedback to improve adolescents' mothering behaviors. *Journal of Adolescent Health*. 1992;13(7):570-575.
278. Barnard KE, Eyres SJ. Child Health Assessment. Part 2: The First Year of Life. 1979;
279. Holden JM, Sagovsky R, Cox JL. Counselling in a general practice setting: controlled study of health visitor intervention in treatment of postnatal depression. *British Medical Journal*. 1989;298(6668):223-226.
280. Murray La, L. The social baby. 2000;London:Constable & Robinson.
281. McCollum JA, Yates T. Dyad as focus, triad as means: A family-centered approach to supporting parent-child interactions. *Infants & Young Children*. 1994;6(4):54-63.
282. Girolametto L, Verbey M, Tannock R. Improving joint engagement in parent-child interaction: An intervention study. *Journal of Early Intervention*. 1994;18(2):155-167.
283. Sheridan SM, Kratochwill TR. *Conjoint behavioral consultation: Promoting family-school connections and interventions*. Springer Science & Business Media; 2007.
284. Kratochwill TR, Bergan JR. *Conjoint behavioral consultation: A procedural manual*. Springer; 1996.
285. Brazelton T. Clinics in Developmental Medicine No. 50. *Phillidelphia: JP Lippincott*. 1973;
286. Schuler ME, Nair P, Black MM. Ongoing maternal drug use, parenting attitudes, and a home intervention: effects on mother-child interaction at 18 months. *Journal of Developmental & Behavioral Pediatrics*. 2002;23(2):87-94.
287. Furuno S, O'Reilly K, Hosaka C, Inatsuka T, Allman T, Zeisloft B. Hawaii early learning profile. *Palo Alto, CA: VORT Corporation*. 1979;
288. Gross RT. Enhancing the outcomes of low-birth-weight premature infants: a multisite randomized trial. *JAMA*. 1990;263:3035-3042.
289. Sanders MR. Triple P–Positive Parenting Program: A population approach to promoting competent parenting. *Australian e-journal for the Advancement of Mental Health*. 2003;2(3):127-143.
290. Sanders MR, Markie-Dadds C, Tully LA, Bor W. The triple P-positive parenting program: a comparison of enhanced, standard, and self-directed behavioral family intervention for parents of children with early onset conduct problems. *Journal of consulting and clinical psychology*. 2000;68(4):624.
291. Spry C, Morawska, A., & Sanders, M. R. . Baby Triple P. Brisbane, Queensland, Australia: Triple P International. 2009;
292. Cooper PJ, Landman M, Tomlinson M, Molteno C, Swartz L, Murray L. Impact of a mother–infant intervention in an indigent peri-urban South African context: pilot study. *The British journal of psychiatry*. 2002;180(1):76-81.
293. (WHO) WHO. Improving the psychosocial development of chidren. *WHO*. 1995;
294. Mercer RT. Becoming a Mother Versus Maternal Role Attainment. https://doi.org/10.1111/j.1547-5069.2004.04042.x. *Journal of Nursing Scholarship*. 2004/09/01 2004;36(3):226-232. doi:https://doi.org/10.1111/j.1547-5069.2004.04042.x
295. Urie Bronfenbrenner PAM. The ecology of developmental processes. 1998;

296. Mercer RT, Walker LO. A review of nursing interventions to foster becoming a mother. *Journal of Obstetric, Gynecologic & Neonatal Nursing*. 2006;35(5):568-582.
297. BOWLBY J. Attachment and Loss, I, Nueva York. Basic Books; 1969.
298. Bandura A. Self-efficacy: Toward a unifying theory of behavioral change. *Psychological Review*. 1977;84(191)(2)
299. Wagner M, Spiker D. Experiences and Outcomes for Children and Families: Multisite Parents as Teachers Evaluation. 2001;
300. Center PaTN. An evidence-based home visiting model consistent with the criteria and requirements for the Ma-ternal, Infant, and Early Childhood Home Visiting Program Enacted as part of the Patient Protec-tion and Affordable Care 2010;
301. Baradon T, Broughton C, Biseo M, et al. *The practice of psychoanalytic parent-infant psychotherapy: Claiming the baby*. Routledge; 2005.
302. Fraiberg S. Clinical studies in infant mental health. 1980;
303. Kaplan-Sarnoff M, Zuckerman B. Healthy steps manual. *Boston University School of Medicine*. 2007;
304. Reddemann L. Imagination als heilsame Kraft. Zur Behandlung von Traumafolgen mit ressourceorientierten Verfahren. *Aufl Stuttgart: Klett Cotta Verlag*. 2001;
305. Rauh VA, Nurcombe B, Achenbach T, Howell C. The Mother-Infant Transaction Program. The content and implications of an intervention for the mothers of low-birthweight infants. *Clinics in perinatology*. 1990;17(1):31-45.
306. Bromwich RM. Focus on maternal behavior in infant intervention. *American journal of Orthopsychiatry*. 1976;46(3):439.
307. Als H, Tronick E, Adamson L, Brazelton TB. The behavior of the full-term but underweight newborn infant. *Developmental Medicine & Child Neurology*. 1976;18(5):590-602.
308. Burns K, Cunningham N, White-Traut R, Silvestri J, Nelson MN. Infant Stimulation: Modification of an Intervention Based on Physiologic and Behavioral Cues. *Journal of Obstetric, Gynecologic & Neonatal Nursing*. 1994/09/01/ 1994;23(7):581-589. doi:https://doi.org/10.1111/j.1552-6909.1994.tb01924.x
309. Stroemple R. Medical Foster Parent Handbook. *National Center for Clinical Infant Programs: Zero to Three*. 1992;
310. Bakermans-Kranenburg MJ, Van Ijzendoorn MH, Juffer F. Less is more: meta-analyses of sensitivity and attachment interventions in early childhood. *Psychological bulletin*. 2003;129(2):195.
311. Martin PR, Negri LM. *Treating postnatal depression: a psychological approach for health care practitioners*. John Wiley; 1999.
312. Field T. Early interventions for infants of depressed mothers. *Pediatrics*. 1998;102(5 Suppl E):1305-1310.
